# Association of a child’s mental disorder with parental income and employment: Analysis of nationwide register-based cohorts in Finland and Denmark

**DOI:** 10.1101/2025.03.21.25324381

**Authors:** Kaisla Komulainen, Ripsa Niemi, Mai Gutvilig, Natalie C. Momen, Petri Böckerman, Marko Elovainio, Oleguer Plana-Ripoll, Christian Hakulinen

**Author notes:** Contributed equally (Oleguer Plana-Ripoll, Christian Hakulinen). **Corresponding author** Kaisla Komulainen, Department of Psychology and Research Program Unit, Faculty of Medicine, University of Helsinki, Finland, PO Box 21, 00014 University of Helsinki, Helsinki, Finland.

## Abstract

**Background:** The onset of a severe physical illness of a child has been associated with earnings and employment losses among parents, but less is known in the context of children’s mental disorders.

**Objectives:** We estimated parental income and employment trajectories associated with a child’s mental disorder diagnosis in nationwide register-based cohorts from Finland and Denmark.

**Methods:** All parents whose child was diagnosed with a mental disorder (F00–F99 in ICD-10) at ages 1–25 in Finland and in Denmark during 1994–2019 were matched 1:1 to parents with a child without a mental disorder on psychiatric and sociodemographic characteristics. Generalized estimating equations were used to estimate the associations of a child’s mental disorder with parental annual income and employment outcomes from five years before to five years after the child’s diagnosis.

**Findings:** In 1994–2019, over one million parents in Finland and Denmark had at least one child diagnosed with a mental disorder at age 1–25. Parents exposed to a child’s mental disorder had consistently lower income and were more often unemployed compared to the matched unexposed parents, already five years before the child’s diagnosis. These differences became slightly larger over time, especially in analyses on strata involving parents whose child was diagnosed at a younger age. However, there was no consistent evidence of a change in parental annual income or employment around the time of their child’s diagnosis.

**Conclusions:** Our analysis shows that even in countries with strong welfare systems, a younger child’s mental disorder may contribute to widening socioeconomic inequity among families. However, the inequity in children’s mental health appears to primarily exist prior to, rather than in response to, a child’s mental disorder.

**Clinical implications:** Clinical and policy efforts should prioritize addressing pre-existing socioeconomic vulnerabilities for effective primary prevention of children’s mental disorders.

**What is already known on this topic:**

- Previous studies have shown earnings and employment losses among parents of a child diagnosed with a severe physical illness, but less is known in the context of mental disorders.

**What this study adds:**

- In this analysis of Finnish and Danish nationwide register-based cohorts, a child’s mental disorder was consistently associated with lower annual income and a lower annual probability of employment among parents, already several years before the child’s diagnosis.
- There was no consistent evidence of a change in parental annual income or employment around the time of their child’s diagnosis.

**How this study might affect research, practice or policy:**

- While a child’s mental disorder can disrupt family life, it does not necessarily translate into immediate socioeconomic consequences for parents in welfare states.
- Further studies are needed in populations with different welfare systems and less universal access to healthcare.

## BACKGROUND

A child’s mental disorder may disrupt family life, interfere with family roles and dynamics, and increase vulnerability to psychological distress in all family members.(1–5) For parents, adapting to a child’s mental disorder may not only involve psychological strain, but also changes in financial situation or position in the labor market. Taking care of a child with a mental disorder may limit parents’ capacity to engage in work or find and maintain full-time employment. On the other hand, in welfare states, social welfare mechanisms available to parents (e.g., unemployment benefits, child care allowance) may compensate for economic hardships due to a child’s mental disorder.(6,7)

While the onset of a severe physical illness of a child has been linked with labor market losses, i.e., a temporary decline in income and employment among parents,(8–11) much less is known in the context of mental disorders. Previous studies report lower income and employment outcomes among parents exposed to a child’s mental or neurodevelopmental disorder,(12–18) but most focus on only a limited set of disorders in children within a wide age range, are based on cross-sectional data or small samples, or rely on self-reported measures of socioeconomic outcomes, which are susceptible to response bias. In a Swedish register study, parents of individuals diagnosed with schizophrenia were more likely to be unemployed and receive more family-related social welfare benefits than parents of healthy controls, while no differences in low-income or disability pension status were observed.(19) Due to lack of information on parental income and employment status prior to their child’s diagnosis, it is yet unclear whether these findings merely reflect the socioeconomic gradient in children’s mental disorders,(20,21) rather than a change in parents’ financial or labor market situation in response to the child’s disorder. Furthermore, the association between a child’s mental disorder and parental socioeconomic outcomes may vary according to several factors, such as the child’s specific disorder, or age at the disorder onset.

## OBJECTIVE

We conducted a matched cohort study in Finnish and Danish nationwide administrative and health registers to provide comprehensive estimates of the socioeconomic trajectories among parents of a child with a mental disorder compared to parents of children without mental disorders. We followed the parents’ annual income and employment before and after a child’s mental disorder diagnosis classified under the major diagnostic categories included in the International Classification of Diseases, 10^th^ revision (ICD-10). We hypothesized that parents of a child with a mental disorder experience a reduction in annual income and employment at the time of their child’s diagnosis. Additionally, we evaluated whether the associations of a child’s mental disorder with parental income and employment outcomes differ depending on the child’s age at diagnosis. Interactive visualizations of the results are available at https://mentalnet.shinyapps.io/app_ses/.

## METHODS

### Study Design and Data Sources

This study was a pair-matched register-based cohort study using population-based administrative and health records of eligible individuals and their children in Finland and in Denmark. The details of the registers used in the present study are provided in **eMethods in Supplement 1**. The registers were linked using unique personal identity numbers issued to all Finnish and Danish residents. The study was approved by the Ethics Committee of the Finnish Institute for Health and Welfare (THL/184/6.02.01/2023§933) and registered with the Danish Data Protection Agency at Aarhus University (No 2016-051-000001-2587). Data were linked with the approval of Statistics Finland (TK-53-1696-16), Statistics Denmark, the Finnish Institute of Health and Welfare, and the Danish Health Data Authority. According to Finnish and Danish law, informed consent is not required for register-based studies.

### Study Population and Exposure

The analyses were performed separately in Finland and in Denmark. All individuals living in Finland and Denmark whose child was diagnosed with a mental disorder in outpatient or inpatient secondary health care at age 1–25 between January 1^st^, 1994 and December 31^st^, 2019 were each matched to one control with a child without a mental disorder diagnosis and without siblings with a mental disorder diagnosis. Exposure to a child’s mental disorder was defined using diagnostic codes included in the subchapter F of the ICD-10 (F00–F99) (**eTable 1 in Supplement 1**). The first diagnosis among children of each family (i.e., same two parents) was considered (for details, see **eMethods in Supplement 1**).

In addition to any mental disorder, we examined exposure to specific disorders of the child. These disorders were classified into nine higher-order categories of mental disorders primarily based on the subchapter F categories of the ICD-10 (e.g., substance use disorders (F10–F19), psychotic disorders (F20–F29), mood disorders (F30–F39) etc.) (**eTable 1 in Supplement 1**). Exposure to a child’s specific mental disorder was restricted to diagnoses recorded in outpatient or inpatient secondary health care when the child was considered sufficiently old to receive a diagnosis, i.e., 5–25 for ICD-10 codes F10–F60, and 1–25 years old for ICD-10 codes F70–F98. Each individual whose child was diagnosed with a given mental disorder in Finland or in Denmark was matched to one control with a child without a diagnosis from the same diagnostic category and without siblings with a diagnosis from the same category.

Parents exposed to a mental disorder of a child were matched to unexposed parents on the child’s birth year and birth month (+/- 2 months), the child’s place of birth categorized into major geographical regions (four regions in Finland, five regions in Denmark; Finnish data from year 1987 was used if the child was born before 1987), both parents’ history of a mental disorder (any disorder/no disorder) prior to their child’s diagnosis, and both parents’ educational attainment (primary/secondary/tertiary) at the time of their child’s birth (Finnish data from year 1987 was used if the child was born before 1987, Danish data from year 1985 was used if the child was born before 1985). Women and men were matched separately.

### Outcomes

The outcomes were the individuals’ records of annual income and employment during a period that started five years before and ended five years after their child’s diagnosis. For the unexposed individuals, the diagnosis date of the matched parent’s child was used as the reference time point. We used records of annual earnings (in euros [EUR] when using Finnish data and Danish krone [DKK] when using Danish data), annual total disposable income (EUR/DKK), annual amount of received social welfare benefits, such as unemployment benefits, disability pension or child benefits (EUR/DKK), as well as annual records of employment status (yes/no) (for details, see **eMethods in Supplement 1**). All income and employment measures were recorded on the last day of each year. Earnings, disposable income and social welfare benefits were adjusted for inflation using the Finnish and Danish consumer price index, respectively, with 2015 as the base year. Earnings and disposable income were top coded at the 99^th^ percentile. To facilitate reporting of the results, DKK was converted to EUR using the average exchange rate for 2015 (1 DKK=0.1341 EUR).

### Statistical Analysis

Generalized estimating equations (GEE) were used to estimate the associations of a child’s mental disorder with parental annual income and employment status during the observation period from five years before to five years after the child’s diagnosis. All income and employment measures were analyzed in separate models. We conducted Poisson GEE models with a log link function for annual earnings, disposable income and social welfare benefits, and binomial GEE models with a logit link for employment status. All GEE models assumed an exchangeable correlation structure, and robust standard errors were used to allow for model misspecification. We used all available data on parental income and employment outcomes, i.e., individuals contributed data in the analysis if both the exposed individual and the matched control had non-missing outcome data from at least one year during the observation period. Women’s and men’s income and employment measures were analyzed in separate models.

In addition, we repeated all analyses stratified by the child’s age at diagnosis (5–9 years, 10–14 years, 15–19 years, 20–25 years for F10–F60, and 1–4 years, 5–9 years, 10–14 years, 15–19 years, 20–25 years for F70–F90 and F00–F99; categories were collapsed if there were too few diagnoses within a stratum). Statistical analyses were conducted in Stata 17 and R 4.2.2/4.4.1 in Finland, and Stata 18 and R 4.4.1 in Denmark.

## FINDINGS

Descriptive characteristics of parents exposed to any mental disorder of a child and the unexposed parents in the year of the affected child’s diagnosis are shown in **Table 1**. Between January 1^st^, 1994, and December 31^st^, 2019, 662 770 individuals (331 385 women and 331 385 men) in Finland and 431 022 individuals (215 511 women and 215 511 men) in Denmark had at least one child diagnosed with a mental disorder at age 1–25. The most common diagnoses among children were anxiety disorders, followed by mood disorders and childhood-onset behavioral and emotional disorders. The child’s average age at diagnosis was 13.3 (SD, 6.8) in Finland and 15.4 (SD, 6.0) in Denmark. The characteristics of parents included in each disorder-specific analysis are reported in **eTables 2–10 in Supplement 1.** Average and total amount of follow-up years during the observation period in each analysis are reported in **eTable 11 in Supplement 1**.

**Table 1.**
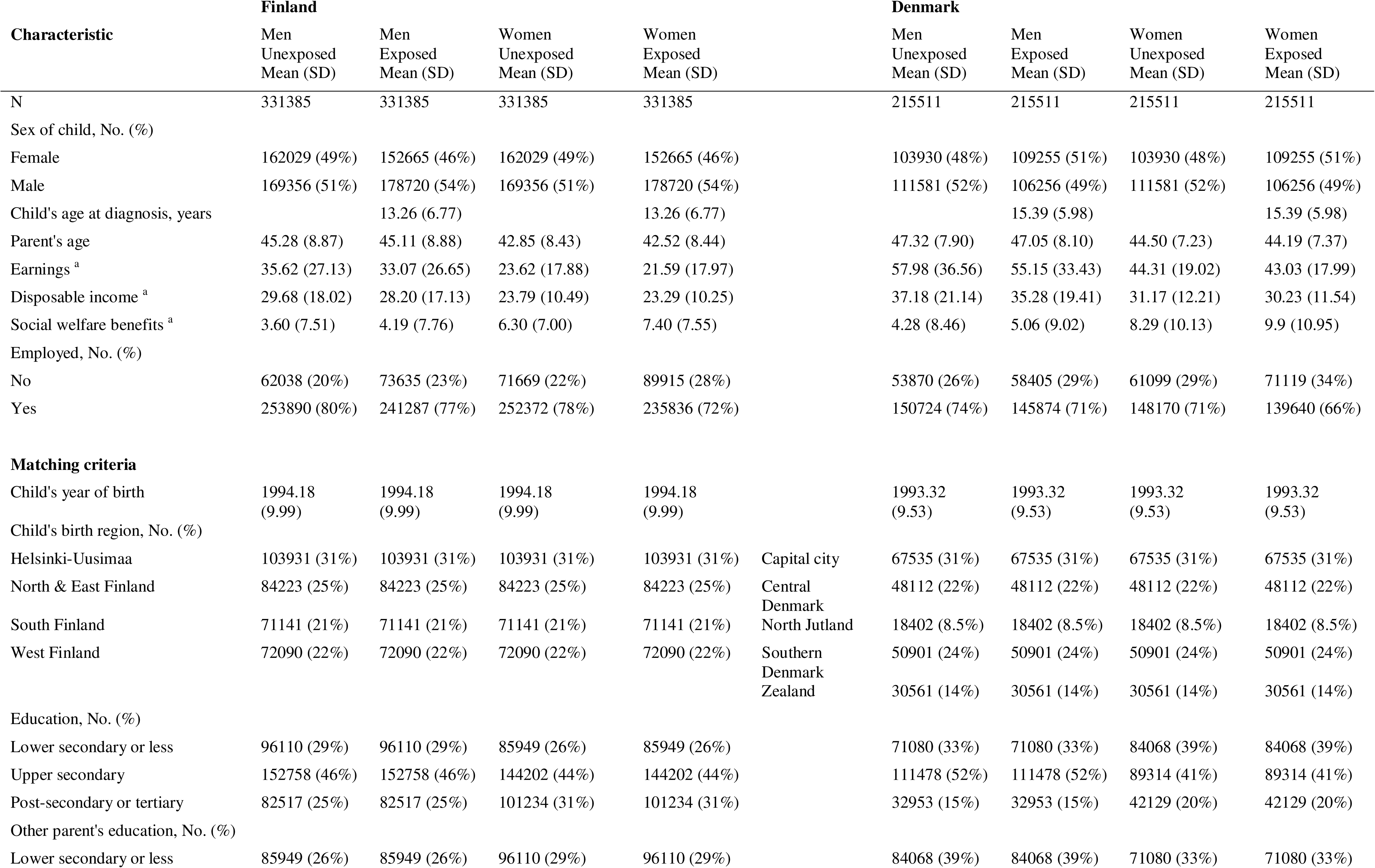
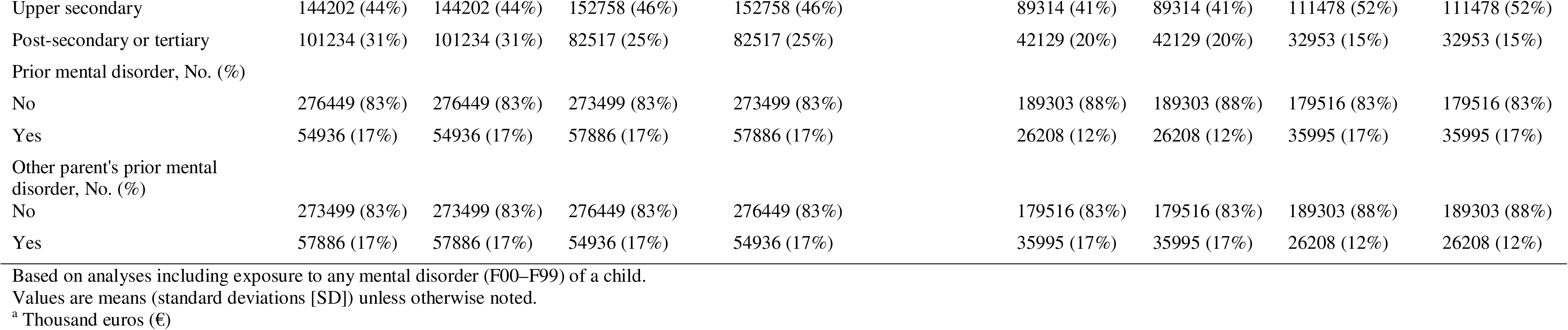
Descriptive characteristics of the Finnish and Danish cohorts.

**Figure 1** shows marginal predictions for the income and employment outcomes among parents exposed to any mental disorder of a child and the unexposed matched parents in Finland and in Denmark (for exact values, see **eTable 12 in Supplement 1**). Across both women and men, exposure to a child’s mental disorder was consistently associated with lower annual earnings, lower annual disposable income, a greater annual amount of received social welfare benefits, and a lower annual probability of employment throughout the observation period. Similar findings were consistently observed when examining exposure to each specific disorder of a child, except for eating disorders; during the observation period, the differences in income and employment between parents exposed to a child’s eating disorder and the unexposed parents were smaller and less consistent, and a child’s eating disorder was associated with greater annual earnings and disposable income among parents both in Finland and Denmark (**Figure 2, eFigures 1–3, eTables 13–21 in Supplement 1)**.

**Figure 1.**
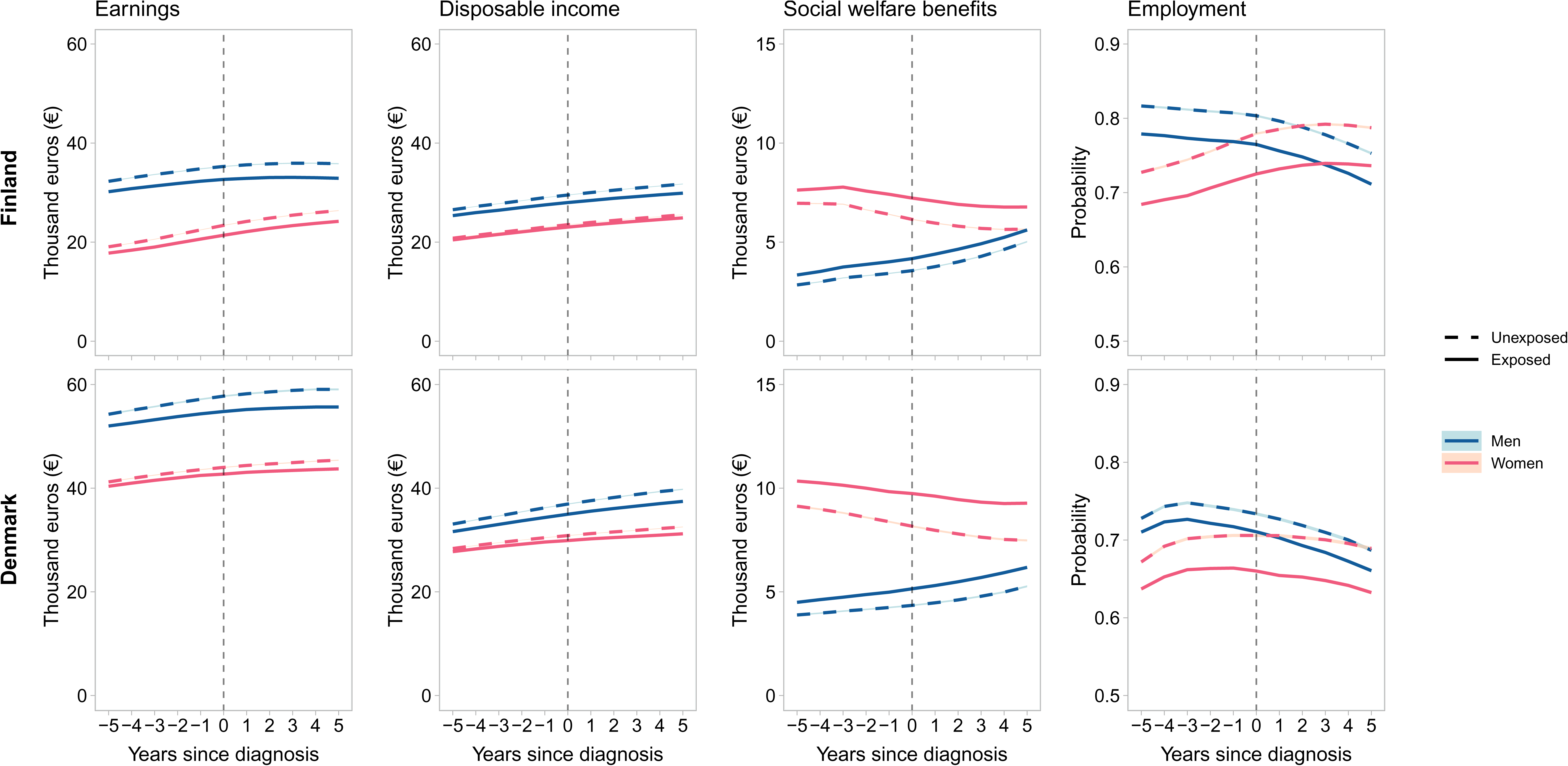
Marginal predictions of annual income and employment trajectories among parents exposed to child’s mental disorder and unexposed parents in Finland and Denmark Shaded areas represent the 95% confidence intervals.

**Figure 2.**
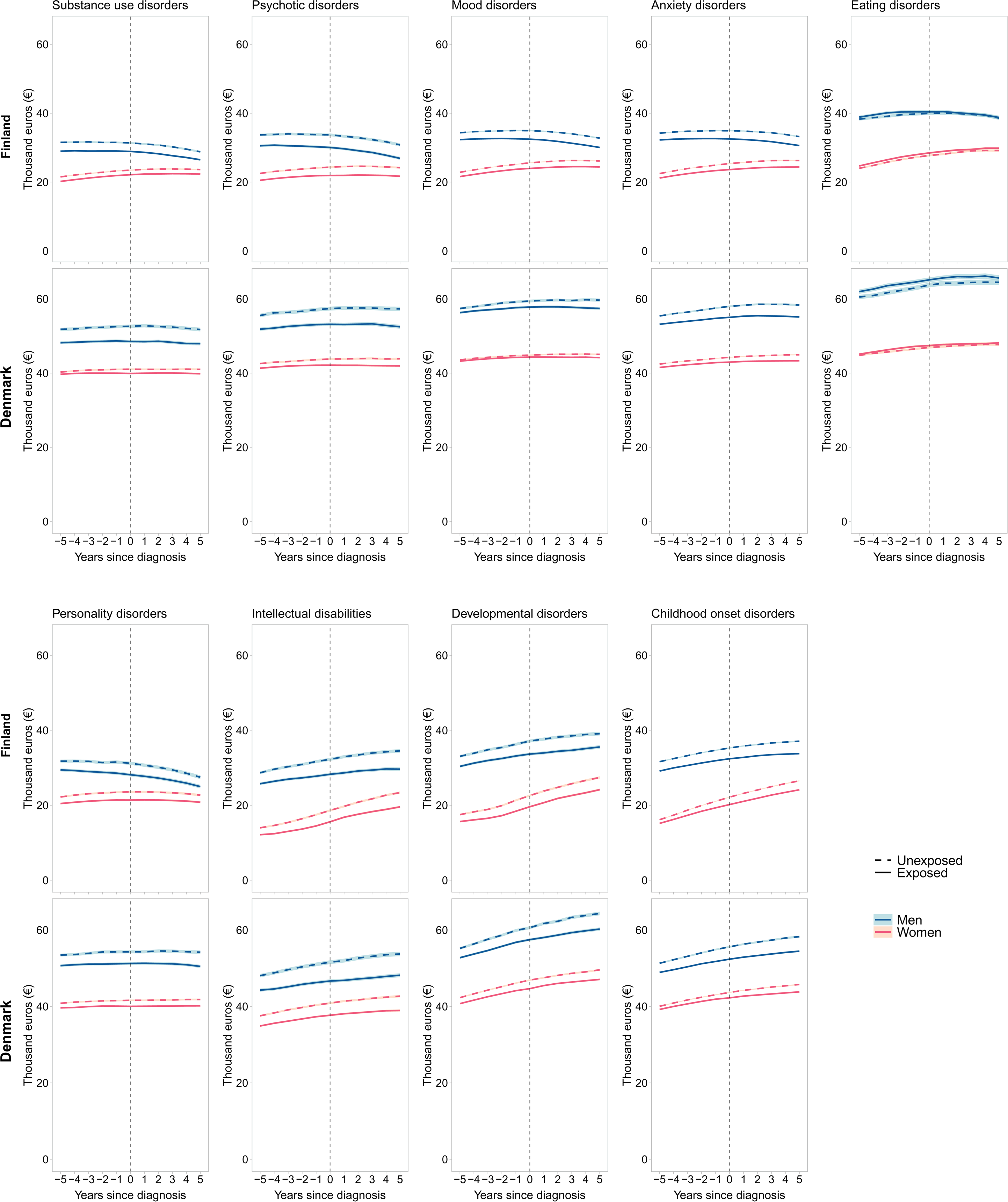
Marginal predictions of annual earnings among parents exposed to child’s specific mental disorders and unexposed parents in Finland and Denmark Shaded areas represent the 95% confidence intervals.

Across both women and men, the differences in income and employment between parents exposed to any mental disorder of a child and the unexposed parents remained relatively stable throughout the observation period. Based on marginal predictions, women whose child was diagnosed with a mental disorder earned 7% less than the unexposed women five years before and 8% less five years after the child’s diagnosis in Finland. In Denmark, they earned 2% and 4% less, respectively (**Figure 1, eTable 12 in Supplement 1**). There was no evidence of an immediate change in any of the outcomes among the exposed parents at the time of their child’s diagnosis. The results from the disorder-specific analyses were similar; the income and employment trajectories diverged slightly during the observation period between exposed and unexposed parents, but there was no evidence of a change at the time of the child’s diagnosis (**Figure 2, eFigures 1–3, eTables 13–21 in Supplement 1)**.

In analyses stratified by the child’s age at diagnosis, the differences in earnings, social welfare benefits, employment, and to a lesser extent disposable income, became larger during the period of observation between parents exposed to a younger child’s mental disorder and the matched unexposed parents. In contrast, this trend leveled off when the child was diagnosed in late adolescence or early adulthood. For instance, based on marginal predictions, women with a child diagnosed before age 5 in Finland earned 8% less than the unexposed women five years before the child’s diagnosis and 15% less five years after the diagnosis, while women with a child diagnosed at age 20–25 earned 6% less than the unexposed women five years before and 5% less five years after the child’s diagnosis (**Figure 3, eFigures 4–6, eTables 22–25 in Supplement 1**). Overall, similar patterns were observed in the disorder-specific analyses, although there was variation across different diagnostic categories. For instance, both in Finland and Denmark, the trajectories of receipt of social welfare benefits and employment clearly diverged among women exposed to a younger child’s anxiety disorder, while for substance use disorders and psychotic disorders, differences across strata were less pronounced (**eFigures 7–42, eTables 26–61**). After no initial difference between exposed and unexposed women during the 5 years leading to a child’s diagnosis, women with a child diagnosed with an eating disorder at age 5–9 in Denmark received markedly more social welfare benefits and were less likely employed in the 1–2 years following the child’s diagnosis. However, due to the low number of eating disorder cases in this stratum, the confidence intervals were wide, and no comparable trend was evident in Finland (**eFigure 25–26, eTables 44–45 in Supplement 1**).

**Figure 3.**
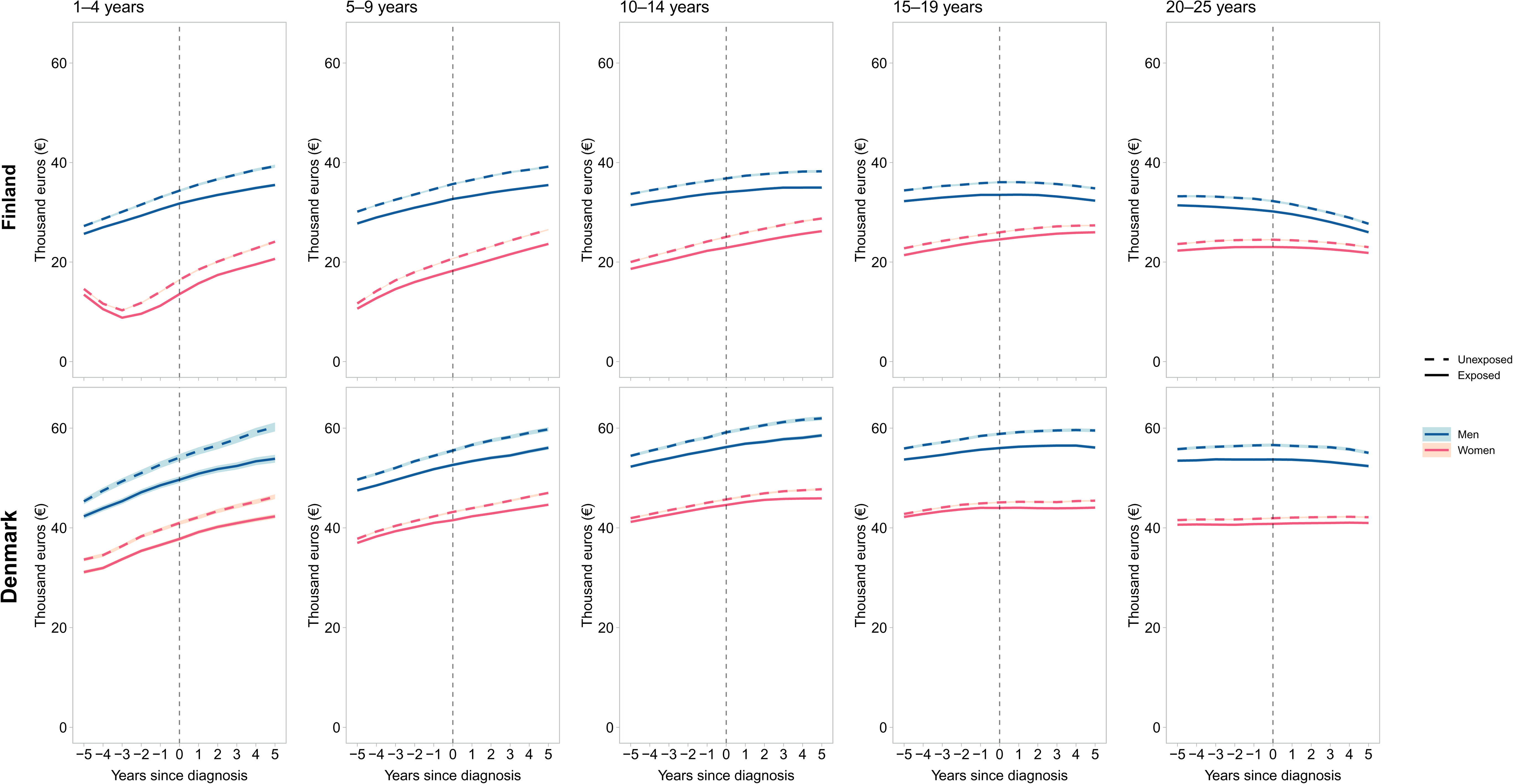
Marginal predictions of annual earnings among parents exposed to child’s mental disorder and unexposed parents stratified by the child’s age at diagnosis in Finland and Denmark Shaded areas represent the 95% confidence intervals.

## DISCUSSION

In this analysis of two nationwide cohorts covering over a million parents of children with mental disorders, we examined how a child’s mental disorder is associated with parental income and employment outcomes across a broad spectrum of mental disorders. During a period from five years before to five years after a child’s diagnosis, parents whose child was diagnosed with a mental disorder had consistently lower annual earnings and disposable income, received a greater annual amount of social welfare benefits, and had higher annual unemployment than unexposed parents. While these differences became slightly larger during the observation period, especially among parents whose children were diagnosed young, there was no evidence of a change in parental annual income or employment status among the exposed group at the time of their child’s mental disorder diagnosis. Similar findings were observed when examining exposure to specific disorders of the child.

These findings align with evidence on socioeconomic disparities in children’s mental disorders;(20,21) in our study, parents of children diagnosed with a mental disorder were on a lower income and employment trajectory already several years before the child’s diagnosis. The only exception to this overall pattern was the inverse association observed for exposure to child’s eating disorders. In line with previous evidence,(22) parents of children diagnosed with an eating disorder were generally on a higher income and employment trajectory throughout the follow-up, which has been hypothesized to reflect genetic factors, environmental expectations, or a socioeconomic gradient in seeking or receiving care for an eating disorder, for instance.(22,23)

Distinct from the pre-existing differences in parental income and employment, our data are not consistent with the hypothesis that parents of children with a mental disorder would experience an immediate reduction in their income or employment upon their child’s diagnosis. In a Swedish register-based study, parents of individuals diagnosed with schizophrenia were more likely unemployed and received more social welfare benefits than parents of healthy controls, while no differences in low-income status or disability pension were observed.(19) Given the lack of control for parental socioeconomic situation prior to their child’s diagnosis, these previous results may yet reflect an overall socioeconomic gradient in children’s mental health rather than a change in parents’ socioeconomic situation since the onset of their child’s disorder. Our study indicates that differences in parental income and employment status already existed five years before the child’s diagnosis, with parents exposed to a child’s mental disorder remaining on a lower socioeconomic trajectory throughout the period of observation.

In contrast to Nordic register-based studies reporting an immediate reduction in parental earnings and employment at the time of a child’s cancer diagnosis,(8–11) we observed no evidence for comparable labor market losses at the time of the child’s mental disorder diagnosis. The different findings likely reflect higher demands of caring for a child with cancer (e.g., more intensive treatment courses, severe side effects), greater perceived unpredictability, immediacy, or threat associated with a severe physical illness, or stigma related to mental illness, which may explain why parents of children with mental disorders could be more likely to maintain employment and their normal work routine. Indeed, the only instance of an immediate change in parental socioeconomic outcomes was observed among women in Denmark whose child was diagnosed with an eating disorder at age 5–9; these women received markedly more social welfare benefits and were less likely to be employed in the 1–2 years following the child’s diagnosis. This observation requires cautious interpretation, as the estimates were imprecise and not replicated in Finland. However, it is possible that eating disorders in young children mirror physical illnesses in some respects, such as requiring acute care initially, which could underlie the observed trajectory among women in Denmark.

The nature of mental disorders is often complex and varied, and diagnostic delays with several psychiatric conditions have been reported to be considerably longer than with severe physical conditions in pediatric populations.(24–26) Thus, the time of diagnosis may not be an unambiguous time zero for a mental disorder onset, but families may cope with their child’s psychiatric symptoms over extended time periods of varying lengths, even years.(27) While the pre-existing socioeconomic disadvantage among parents is likely a risk factor for children’s mental disorders, it can also represent an early consequence of managing an undiagnosed child's difficulties, in part explaining why no immediate change in parental socioeconomic outcomes was observed at the time of the child’s mental disorder diagnosis. In addition, the diagnostic delay could underlie the widening differences in income and employment between parents exposed to a younger child’s mental disorder compared to the unexposed parents during the observation period. Even subclinical symptoms a child experiences prior to the diagnosis can require parents to allocate resources to providing care for their child, potentially impeding parents’ career progression or compromising their capacity to work full-time, especially with children diagnosed at younger ages. Also, as expected, parents exposed to a younger child’s mental disorder were younger on average. Disruptions at earlier career stages have been associated with diminished long-term labor market prospects,(28–30) which can partially underlie the less progressive socioeconomic trajectory we observed among parents exposed to a younger child’s mental disorder.

There are limitations to our study. First, this is an observational register-based study, and causal interpretations of our findings have to be taken with caution. Although we controlled for a set of confounders with known or hypothesized associations with the exposures and outcomes through matching, there could be additional confounders, and we cannot rule out the possibility of unmeasured or residual confounding. Second, we used clinical diagnoses received in secondary health care to evaluate exposure to a child’s mental disorder. Although the diagnostic validity of many register-based mental disorders is considered good,(31,32) parents with children whose mental disorders were untreated, or treated solely in the primary health care were misclassified as unexposed in our analysis.

In addition, we had no information on the timing of the child’s symptom onset, remission or recovery, which likely affect parents’ capacity to engage in work. Finally, our data are from two Nordic welfare states, which limits generalizability. However, the similar findings in both Finland and Denmark strengthen the robustness of our conclusions in this context.

In this study, a child’s mental disorder was consistently associated with lower annual income and employment among their parents before and after the child’s diagnosis. While parents with younger children diagnosed with mental disorders followed a less progressive income and employment trajectory, there was no consistent evidence of an immediate decline in parental income or employment around the time of the child’s diagnosis. These findings suggest that in countries with robust welfare systems, a younger child’s mental disorder may contribute to widening socioeconomic inequity among families. However, the inequity in children’s mental health appears to primarily exist prior to, rather than in response to, a child’s mental disorder.

## Clinical implications

Although a child’s mental disorder can disrupt family life, it does not necessarily lead to immediate socioeconomic consequences for parents in welfare states. Clinical and policy efforts should prioritize addressing pre-existing socioeconomic vulnerabilities for effective primary prevention.

## Author Contributions

KK: conceptualization, methodology, visualization, writing – original draft. RN: methodology, formal analysis, visualization, writing – reviewing and editing. MG: methodology, formal analysis, visualization, writing – reviewing and editing. NCM: formal analysis, writing – reviewing and editing. PB: conceptualization, methodology, writing – reviewing and editing. ME: project administration, supervision, writing – reviewing and editing. OP-R: conceptualization, methodology, formal analysis, funding acquisition, project administration, supervision, writing – reviewing and editing. CH: conceptualization, methodology, visualization, funding acquisition, project administration, supervision, writing – reviewing and editing. KK, RN, MG and CH accessed and verified the Finnish data. NCM and OP-R accessed and verified the Danish data. All authors accept responsibility for the decision to submit for publication. OP-R and CH are co-senior authors. CH is the guarantor.

## Disclaimer

Views and opinions expressed are however those of the author(s) only and do not necessarily reflect those of the European Union or the European Research Council. Neither the European Union nor the granting authority can be held responsible for them. The funders had no role in the design and conduct of the study; collection, management, analysis, and interpretation of the data; preparation, review, or approval of the manuscript; or decision to submit the manuscript for publication.

## Patient consent for publication

According to Finnish and Danish law, informed consent is not required for register-based studies.

## Ethics approval

The study was approved by the Ethics Committee of the Finnish Institute for Health and Welfare (THL/184/6.02.01/2023§933) and registered with the Danish Data Protection Agency at Aarhus University (No 2016-051-000001-2587). Data were linked with the approval of Statistics Finland (TK-53-1696-16), Statistics Denmark, the Finnish Institute of Health and Welfare, and the Danish Health Data Authority.

## Data availability

The person-level data that support the findings of this study are available from the National Institute of Health and Welfare (http://www.thl.fi) and Statistics Finland (www.stat.fi) for Finnish data, and Statistics Denmark (https://www.dst.dk) and the Danish Health Data Authority (https://sundhedsdatastyrelsen.dk) for Danish data. Restrictions apply to the availability of these data, which were used under license for this study. Inquiries about secure access to Finnish data should be directed to data permit authority Findata (https://findata.fi/en/). Researchers who fulfill the requirements set by the Danish data providers could gain access to Danish data through Statistics Denmark (https://www.dst.dk) and/or the Danish Health Data Authority (https://sundhedsdatastyrelsen.dk).

## References

1. Jayasinghe A, Wrobel A, Filia K, Byrne LK, Melvin G, Murrihy S, et al. Distress, burden, and wellbeing in siblings of people with mental illness: a mixed studies systematic review and meta-analysis. Psychol Med. 2023 Nov;53(15):6945–64.

2. Schulze B, Rössler W. Caregiver burden in mental illness: review of measurement, findings and interventions in 2004–2005. Curr Opin Psychiatry. 2005 Nov;18(6):684–91.

3. Caqueo-Urízar A, Rus-Calafell M, Craig TKJ, Irarrazaval M, Urzúa A, Boyer L, et al. Schizophrenia: Impact on Family Dynamics. Curr Psychiatry Rep. 2017 Jan;19(1):2.

4. Hakulinen C, Gutvilig M, Niemi R, Momen NC, Pulkki-Råback L, Böckerman P, et al. Associations of mental disorders in children with parents’ subsequent mental disorders: nationwide cohort study from Finland and Denmark. Br J Psychiatry. 2025 Jan 15;1–8.

5. Ademosu T, Ebuenyi I, Hoekstra RA, Prince M, Salisbury T. Burden, impact, and needs of caregivers of children living with mental health or neurodevelopmental conditions in low-income and middle-income countries: a scoping review. Lancet Psychiatry. 2021 Oct;8(10):919–28.

6. O’Campo P, Molnar A, Ng E, Renahy E, Mitchell C, Shankardass K, et al. Social welfare matters: A realist review of when, how, and why unemployment insurance impacts poverty and health. Soc Sci Med. 2015 May;132:88–94.

7. Keskimäki I, Tynkkynen LK, Reissell E, Koivusalo M, Syrjä V, Vuorenkoski L, et al. Finland: Health Systems Review. Health Syst Transit. 2019;21(2):1–166.

8. Lindahl Norberg A, Montgomery SM, Bottai M, Heyman M, Hovén EI. Short-term and long-term effects of childhood cancer on income from employment and employment status: A national cohort study in Sweden: Economic Effects of Childhood Cancer. Cancer. 2017 Apr 1;123(7):1238–48.

9. Öhman M, Woodford J, Essen L. Socioeconomic consequences of parenting a child with cancer for fathers and mothers in Sweden: A population based difference in difference study. Int J Cancer. 2021 May 15;148(10):2535–41.

10. Vaalavuo M, Salokangas H, Tahvonen O. Gender Inequality Reinforced: The Impact of a Child’s Health Shock on Parents’ Labor Market Trajectories. Demography. 2023;1(60(4)):1005–29.

11. Mader L, Hargreave M, Bidstrup PE, Kjær SK, Nielsen TT, Krøyer A, et al. The impact of childhood cancer on parental working status and income in Denmark: Patterns over time and determinants of adverse changes. Int J Cancer. 2020 Aug 15;147(4):1006–17.

12. Busch SH, Barry CL. Mental Health Disorders In Childhood: Assessing The Burden On Families. Health Aff (Millwood). 2007 Jul;26(4):1088–95.

13. Cidav Z, Marcus SC, Mandell DS. Implications of Childhood Autism for Parental Employment and Earnings. Pediatrics. 2012 Apr 1;129(4):617–23.

14. Richard P. Children’s Mental Disorders and Their Mothers’ Earnings: Implications for the Affordable Care Act of 2010. J Fam Econ Issues. 2016 Jun;37(2):156–71.

15. Parish SL, Seltzer MM, Greenberg JS, Floyd F. Economic Implications of Caregiving at Midlife: Comparing Parents With and Without Children Who Have Developmental Disabilities. Taylor SJ, editor. Ment Retard. 2004 Dec;42(6):413–26.

16. Kvist AP, Nielsen HS, Simonsen M. The importance of children’s ADHD for parents’ relationship stability and labor supply. Soc Sci Med. 2013 Jul;88:30–8.

17. McEvilly M, Wicks S, Dalman C. Sick Leave and Work Participation Among Parents of Children with Autism Spectrum Disorder in the Stockholm Youth Cohort: A Register Linkage Study in Stockholm, Sweden. J Autism Dev Disord. 2015 Jul;45(7):2157–67.

18. McCall BP, Starr EM. Effects of autism spectrum disorder on parental employment in the United States: evidence from the National Health Interview Survey. Community Work Fam. 2018;21(4):367–92.

19. Mittendorfer-Rutz E, Rahman S, Tanskanen A, Majak M, Mehtälä J, Hoti F, et al. Burden for Parents of Patients With Schizophrenia—A Nationwide Comparative Study of Parents of Offspring With Rheumatoid Arthritis, Multiple Sclerosis, Epilepsy, and Healthy Controls. Schizophr Bull. 2019 Jun 18;45(4):794–803.

20. Reiss F. Socioeconomic inequalities and mental health problems in children and adolescents: A systematic review. Soc Sci Med. 2013 Aug;90:24–31.

21. Suokas K, Koivisto AM, Hakulinen C, Kaltiala R, Sund R, Lumme S, et al. Association of Income With the Incidence Rates of First Psychiatric Hospital Admissions in Finland, 1996-2014. JAMA Psychiatry. 2020 Mar 1;77(3):274.

22. Koch SV, Larsen JT, Plessen KJ, Thornton LM, Bulik CM, Petersen LV. Associations between parental socioeconomic, family, and sibling status and risk of eating disorders in offspring in a Danish national female cohort. Int J Eat Disord. 2022 Aug;55(8):1130–42.

23. Huryk KM, Drury CR, Loeb KL. Diseases of affluence? A systematic review of the literature on socioeconomic diversity in eating disorders. Eat Behav. 2021 Dec;43:101548.

24. Dang-Tan T, Franco EL. Diagnosis delays in childhood cancer: A review. Cancer. 2007 Aug 15;110(4):703–13.

25. Keramatian K, Pinto JV, Schaffer A, Sharma V, Beaulieu S, Parikh SV, et al. Clinical and demographic factors associated with delayed diagnosis of bipolar disorder: Data from Health Outcomes and Patient Evaluations in Bipolar Disorder (HOPE-BD) study. J Affect Disord. 2022 Jan;296:506–13.

26. Boulton KA, Hodge MA, Jewell A, Ong N, Silove N, Guastella AJ. Diagnostic delay in children with neurodevelopmental conditions attending a publicly funded developmental assessment service: findings from the Sydney Child Neurodevelopment Research Registry. BMJ Open. 2023 Feb;13(2):e069500.

27. Hansen AS, Kjaersdam Telléus G, Færk E, Mohr-Jensen C, Lauritsen MB. Help-seeking pathways prior to referral to outpatient child and adolescent mental health services. Clin Child Psychol Psychiatry. 2021 Apr;26(2):569–85.

28. Schmillen A, Umkehrer M. The scars of youth: Effects of early-career unemployment on future unemployment experience. Int Labour Rev. 2017 Dec;156(3–4):465–94.

29. Möller J, Umkehrer M. Are there Long-Term Earnings Scars from Youth Unemployment in Germany? Jahrb F Natl U Stat. 2015;235(4+5):474–98.

30. Strandh M, Winefield A, Nilsson K, Hammarstrom A. Unemployment and mental health scarring during the life course. Eur J Public Health. 2014 Jun 1;24(3):440–5.

31. Sund R. Quality of the Finnish Hospital Discharge Register: A systematic review. Scand J Public Health. 2012 Aug;40(6):505–15.

32. Mors O, Perto GP, Mortensen PB. The Danish Psychiatric Central Research Register. Scand J Public Health. 2011 Jul;39(7_suppl):54–7.

