## Supplement 1 for "Association of a child’s mental disorder with parental income and employment: Analysis of nationwide register-based cohorts in Finland and Denmark"

### eMethods

#### 1.1 Data sources

##### Finland

We linked individual-level records across the full Finnish Population Register (FOLK) maintained by Statistics Finland, and the Care Register for Health Care (CRHC) and the Hospital Discharge Register (HDR) maintained by the National Institute for Health and Welfare. FOLK contains annually updated demographic information of the whole Finnish population as well as records of income, employment, and education. The CRHC and the HDR include day-to-day data on mental disorder diagnoses based on records of contact with public and private secondary health care in Finland. We used FOLK data from 1987–2020, HRD data from 1970–1993, and CRHC data from 1994–2019. ICD-8 (used in HDR during 1970–1986) and ICD-9 codes (used in HDR and CRHC during 1987–1995) were converted to the corresponding ICD-10 codes (used from 1996 onwards).

##### Denmark

We linked individual-level records across the full Danish Population Register (BEF registers) maintained by Statistics Denmark. The BEF registers contain annually updated demographic information of the whole population living in Denmark, and we used data from 1986 to 2021. We linked this population to other data sources from Statistics Denmark on, e.g., income, employment, and education, available from 1985 onwards. These data were further linked to mental disorder diagnoses based on records of contact with public and private secondary health care in Denmark, which are available since 1968. ICD-8 codes (used during 1968–1993) were converted to the corresponding ICD-10 codes (used from 1994 onwards).

#### 1.2 Exposure to a child's mental disorder

Exposure to a child's mental disorder was restricted to diagnoses received in outpatient or inpatient secondary health care when the child was considered sufficiently old to receive a diagnosis, i.e., 5–25 for ICD-10 codes F10–F60, and 1–25 years old for ICD-10 codes F70–F98 and F00–F99. The first diagnosis among children of each family (i.e., same two parents) was considered. A parent could be considered "exposed" to several mental disorders if they had a child with diagnoses belonging to more than one diagnostic category. Parents who had diagnosed children with more than one partner or more than one child diagnosed on the same day could be included as exposed more than once in each analysis. Parents with multiple children who all were undiagnosed could be included as matched unexposed parents more than once in each analysis.

#### 1.3 Socioeconomic outcomes

##### Finland

Earnings comprised the sum of individuals' earned income and entrepreneurial income. Earned income included cash salary, compensation for employment-related costs, and employee benefits. Entrepreneurial income included the entrepreneur's entrepreneurial income, business groups' entrepreneurial income, and other entrepreneurial income (copyrights, patents etc.). Any negative sum values were imputed as 0 euros.

Disposable income consisted of the individuals' gross income (incl. earned income, entrepreneurial income, capital income, and social welfare benefits) after taxes and other mandatory tax-like payments.

Social welfare benefits included government transfer payments such as social security benefits (e.g., unemployment benefits, rehabilitation benefits), social allowances (e.g., child benefits, study and research grants), and other received transfers.

Employment was a binary variable where the employed individuals were classified as employed and all others (e.g., unemployed, students, pensioners) were categorized as unemployed.

Each income variable was measured on the last day of each statistical year. Employment was categorized based on the labor market status during the last week of the year.

**Denmark**

Earnings comprised the sum of individuals' annual income based on Statistics Denmark's variable perindkialt_13. This variable includes the sum of all work income, transfer of public benefits, capital income, and any other income that can be directly attributed to one individual. This value is the total value before any taxes, labor market contributions or interest expenses have been subtracted. For self-employed individuals, this variable can take negative values as a result of losses in the company.

Disposable income comprised the individuals' annual disposable income based on Statistics Denmark's variable dispon_13. It is the total annual income (i.e., the earnings described above) with the added rental value of an owned home and after subtracting taxes, interest expenses, children maintenance or alimony, and similar. The remaining value is a measure of disposable annual income, i.e., how much there is left for consumption and spending.

Social welfare benefits included annual public transfers based on Statistics Denmark's variable off_overforsel_13. The variable includes a sum of all transfers that are primarily financed by the public sector.

Employment was a binary variable where the employed individuals were classified as employed and all others (e.g., unemployed, students, pensioners) were categorized as unemployed.

Each income variable was measured based on tax reports with information from the entire year. Employment was categorized based on an algorithm created by Statistics Denmark to determine the socioeconomic position category over most of the year.

### Diagnostic categories of mental disorders in Finland and Denmark

**eTable 1. Diagnostic categories of mental disorders in Finland and Denmark**

| | ICD-10 | Abbreviation | Finnish Conversions | | Danish Conversions |
|---|---|---|---|---|---|
| | | | ICD-8 Equivalents | ICD-9 Equivalents | ICD-8 Equivalents |
| Mental and behavioral disorders due to psychoactive substance use | F10–F19 | Substance use disorders | 291, 303, 304 | 291, 292, 303, 304, 305 | 291.x9, 294.39, 303.x9, 303.20, 303.28, 303.90, 304.x9 |
| Schizophrenia, schizotypal and delusional disorders | F20–F29 | Psychotic disorders | 295, 297, 298.1, 298.2, 298.3, 298.9, 299 | 295, 297, 298 | 295.x9, 296.89, 297.x9, 298.29–298.99, 299.04, 299.05, 299.09, 301.83 |
| Mood (affective) disorders | F30–F39 | Mood disorders | 296, 298.0, 300.4, 301.1 | 296, 311 | 296.x9 (Excluding 296.89), 298.09, 298.19, 300.49, 301.19 |
| Neurotic, stress-related and somatoform disorders | F40–F48 | Anxiety disorders | 300.0, 300.1, 300.2, 300.3 300.5, 300.6, 300.7, 300.8, 300.9, 305, 306.8, 307, 308.4 | 300, 307.8, 309 | 300.x9 (Excluding 300.49), 305.x9, 305.68, 307.99 |
| Eating Disorders | F50 | Eating disorders | 305.60, 306.50, 306.58, 306.59 | 307.1, 307.5 | 305.60, 306.50, 306.58, 306.59 |
| Specific Personality Disorders | F60 | Personality disorders | 301.x9 (excluding 301.19), 301.80, 301.81, 301.82, 301.84 | 301 | 301.x9 (Excluding 301.19), 301.80, 301.81, 301.82, 301.84 |
| Intellectual disability | F70–F79 | Intellectual disabilities | 310, 311, 312, 313, 314, 315 | 317, 318, 319 | 311.xx, 312.xx, 313.xx, 314.xx, 315.xx |
| Pervasive Developmental Disorders | F84 | Developmental disorders | 299.00, 299.01, 299.02, 299.03 | 299 | 299.00, 299.01, 299.02, 299.03 |
| Behavioral and emotional disorders with onset in childhood and adolescence | F90–F98 | Childhood onset disorders | 306.2, 306.6, 306.7, 306.9, 308.1, 308.2 308.3 | 308.0, 307.2, 307.3, 307.6, 307.7, 307.9, 313, 314 | 306.x9, 308.0x |
| Any mental disorder | F00–F99 | Any mental disorder | All of the above + 290, 292, 293, 294, rest of 301, 302, 3060 3061, 3063, 3064, 3080, 309 | All of the above + 290, 293, 294, 302, 3074, 312, 316, 310, 315 | All of the above |

#### Descriptive characteristics of the Finnish and Danish cohorts

**eTable 2. Descriptive characteristics in analyses including exposure to child's substance use disorder (F10–F19)**

| | Finland | | | | Denmark | | | |
|---|---|---|---|---|---|---|---|---|
| Characteristic | Men Unexposed | Men Exposed | Women Unexposed | Women Exposed | Men Unexposed | Men Exposed | Women Unexposed | Women Exposed |
| N | 52618 | 52618 | 52618 | 52618 | 23877 | 23877 | 23877 | 23877 |
| | Mean (SD) | Mean (SD) | Mean (SD) | Mean (SD) | Mean (SD) | Mean (SD) | Mean (SD) | Mean (SD) |
| Sex of child, N (%) | | | | | | | | |
| Female | 25836 (49%) | 21376 (41%) | 25836 (49%) | 21376 (41%) | 11555 (48%) | 7749 (32%) | 11555 (48%) | 7749 (32%) |
| Male | 26782 (51%) | 31242 (59%) | 26782 (51%) | 31242 (59%) | 12322 (52%) | 16128 (68%) | 12322 (52%) | 16128 (68%) |
| Child's age at diagnosis | | 19.50 (3.41) | | 19.50 (3.41) | | 20.27 (2.95) | | 20.27 (2.95) |
| Parent's age | 51.05 (6.91) | 50.28 (6.95) | 48.53 (6.35) | 47.66 (6.44) | 51.31 (6.53) | 50.76 (6.75) | 48.35 (5.68) | 47.71 (5.78) |
| Earnings* | 31.87 (27.23) | 29.58 (26.80) | 24.03 (17.81) | 22.67 (17.94) | 52.97 (32.58) | 49.11 (29.52) | 41.44 (17.74) | 40.33 (16.83) |
| Disposable income* | 28.47 (17.73) | 27.05 (17.07) | 23.36 (10.36) | 23.05 (10.10) | 34.67 (19.11) | 31.98 (17.27) | 28.98 (11.36) | 27.96 (10.64) |
| Social welfare benefits* | 5.09 (8.72) | 5.60 (8.75) | 5.49 (7.24) | 6.31 (7.58) | 5.84 (9.69) | 7.39 (10.53) | 8.41 (10.84) | 10.31 (11.67) |
| Employed, N (%) | | | | | | | | |
| No | 13435 (28%) | 15260 (32%) | 12061 (24%) | 14039 (28%) | 7230 (33%) | 8243 (38%) | 7927 (34%) | 9184 (40%) |
| Yes | 35183 (72%) | 32365 (68%) | 38959 (76%) | 36896 (72%) | 14924 (67%) | 13424 (62%) | 15108 (66%) | 13717 (60%) |
| Matching criteria | | | | | | | | |
| | N (%) | N (%) | N (%) | N (%) | N (%) | N (%) | N (%) | N (%) |
| Child's year of birth, Mean (SD) | 1989.11 (7.89) | 1989.11 (7.89) | 1989.11 (7.89) | 1989.11 (7.89) | 1988.27 (7.66) | 1988.27 (7.66) | 1988.27 (7.66) | 1988.27 (7.66) |

**eTable 2. Descriptive characteristics in analyses including exposure to child's substance use disorder (F10–F19)**

| | Finland | | | | | Denmark | | | |
|---|---|---|---|---|---|---|---|---|---|
| Characteristic | Men Unexposed | Men Exposed | Women Unexposed | Women Exposed | | Men Unexposed | Men Exposed | Women Unexposed | Women Exposed |
| N | 52618 | 52618 | 52618 | 52618 | | 23877 | 23877 | 23877 | 23877 |
| Child's birth region | | | | | | | | | |
| Helsinki-Uusimaa | 14868 (28%) | 14868 (28%) | 14868 (28%) | 14868 (28%) | Capital city | 7204 (30%) | 7204 (30%) | 7204 (30%) | 7204 (30%) |
| North & East Finland | 14007 (27%) | 14007 (27%) | 14007 (27%) | 14007 (27%) | Central Denmark | 5201 (22%) | 5201 (22%) | 5201 (22%) | 5201 (22%) |
| South Finland | 12000 (23%) | 12000 (23%) | 12000 (23%) | 12000 (23%) | North Jutland | 2035 (8.5%) | 2035 (8.5%) | 2035 (8.5%) | 2035 (8.5%) |
| West Finland | 11743 (22%) | 11743 (22%) | 11743 (22%) | 11743 (22%) | Southern Denmark | 6253 (26%) | 6253 (26%) | 6253 (26%) | 6253 (26%) |
| | | | | | Zealand | 3184 (13%) | 3184 (13%) | 3184 (13%) | 3184 (13%) |
| Education | | | | | | | | | |
| Lower secondary or less | 18799 (36%) | 18799 (36%) | 17871 (34%) | 17871 (34%) | | 10280 (43%) | 10280 (43%) | 12846 (54%) | 12846 (54%) |
| Upper secondary | 23697 (45%) | 23697 (45%) | 22819 (43%) | 22819 (43%) | | 11332 (47%) | 11332 (47%) | 8187 (34%) | 8187 (34%) |
| Post-secondary or tertiary | 10122 (19%) | 10122 (19%) | 11928 (23%) | 11928 (23%) | | 2265 (9.5%) | 2265 (9.5%) | 2844 (12%) | 2844 (12%) |
| Other parent's education | | | | | | | | | |
| Lower secondary or less | 17871 (34%) | 17871 (34%) | 18799 (36%) | 18799 (36%) | | 12846 (54%) | 12846 (54%) | 10280 (43%) | 10280 (43%) |
| Upper secondary | 22819 (43%) | 22819 (43%) | 23697 (45%) | 23697 (45%) | | 8187 (34%) | 8187 (34%) | 11332 (47%) | 11332 (47%) |
| Post-secondary or tertiary | 11928 (23%) | 11928 (23%) | 10122 (19%) | 10122 (19%) | | 2844 (12%) | 2844 (12%) | 2265 (9.5%) | 2265 (9.5%) |
| Prior mental disorder | | | | | | | | | |

**eTable 2. Descriptive characteristics in analyses including exposure to child's substance use disorder (F10–F19)**

| | Finland | | | | Denmark | | | |
|---|---|---|---|---|---|---|---|---|
| Characteristic | Men Unexposed | Men Exposed | Women Unexposed | Women Exposed | Men Unexposed | Men Exposed | Women Unexposed | Women Exposed |
| N | 52618 | 52618 | 52618 | 52618 | 23877 | 23877 | 23877 | 23877 |
| No | 39775 (76%) | 39775 (76%) | 40450 (77%) | 40450 (77%) | 19589 (82%) | 19589 (82%) | 18595 (78%) | 18595 (78%) |
| Yes | 12843 (24%) | 12843 (24%) | 12168 (23%) | 12168 (23%) | 4288 (18%) | 4288 (18%) | 5282 (22%) | 5282 (22%) |
| Other parent's prior mental disorder | | | | | | | | |
| No | 40450 (77%) | 40450 (77%) | 39775 (76%) | 39775 (76%) | 18595 (78%) | 18595 (78%) | 19589 (82%) | 19589 (82%) |
| Yes | 12168 (23%) | 12168 (23%) | 12843 (24%) | 12843 (24%) | 5282 (22%) | 5282 (22%) | 4288 (18%) | 4288 (18%) |

* Thousand euros (€)

**eTable 3. Descriptive characteristics in analyses including exposure to child's psychotic disorder (F20–F29)**

| | Finland | | | | Denmark | | | |
|---|---|---|---|---|---|---|---|---|
| Characteristic | Men Unexposed | Men Exposed | Women Unexposed | Women Exposed | Men Unexposed | Men Exposed | Women Unexposed | Women Exposed |
| N | 24233 | 24233 | 24233 | 24233 | 22812 | 22812 | 22812 | 22812 |
| | Mean (SD) | Mean (SD) | Mean (SD) | Mean (SD) | Mean (SD) | Mean (SD) | Mean (SD) | Mean (SD) |
| Sex of child, N (%) | | | | | | | | |
| Female | 11758 (49%) | 10258 (42%) | 11758 (49%) | 10258 (42%) | 11086 (49%) | 10775 (47%) | 11086 (49%) | 10775 (47%) |
| Male | 12475 (51%) | 13975 (58%) | 12475 (51%) | 13975 (58%) | 11726 (51%) | 12037 (53%) | 11726 (51%) | 12037 (53%) |
| Child's age at diagnosis | | 19.79 (3.61) | | 19.79 (3.61) | | 19.49 (3.52) | | 19.49 (3.52) |
| Parent's age | 51.22 (6.84) | 51.48 (7.08) | 48.79 (6.33) | 48.96 (6.51) | 51.03 (6.64) | 51.08 (6.95) | 48.19 (5.77) | 48.03 (5.91) |
| Earnings* | 34.02 (29.50) | 30.51 (28.28) | 24.73 (18.68) | 22.30 (18.67) | 57.94 (37.69) | 53.69 (33.74) | 44.37 (19.92) | 42.69 (19.14) |
| Disposable income* | 29.75 (19.40) | 27.85 (18.08) | 23.62 (11.05) | 22.58 (10.71) | 37.68 (21.98) | 34.98 (19.69) | 31.07 (12.88) | 29.85 (12.18) |
| Social welfare benefits* | 4.98 (9.35) | 5.79 (9.12) | 5.29 (7.24) | 6.15 (7.48) | 5.07 (9.18) | 6.28 (9.98) | 7.72 (10.45) | 9.44 (11.24) |
| Employed, N (%) | | | | | | | | |
| No | 5957 (27%) | 7070 (32%) | 5550 (24%) | 7026 (30%) | 6234 (29%) | 7041 (33%) | 6822 (31%) | 8063 (37%) |
| Yes | 16516 (73%) | 15110 (68%) | 17971 (76%) | 16435 (70%) | 14933 (71%) | 14022 (67%) | 15182 (69%) | 13993 (63%) |
| Matching criteria | | | | | | | | |
| | N (%) | N (%) | N (%) | N (%) | N (%) | N (%) | N (%) | N (%) |
| Child's year of birth, Mean (SD) | 1987.61 (8.02) | 1987.61 (8.02) | 1987.61 (8.02) | 1987.61 (8.02) | 1989.82 (8.16) | 1989.82 (8.16) | 1989.82 (8.16) | 1989.82 (8.16) |
| Child's birth region | | | | | | | | |

**eTable 3. Descriptive characteristics in analyses including exposure to child's psychotic disorder (F20–F29)**

| Characteristic | Finland Men Unexposed | Finland Men Exposed | Finland Women Unexposed | Finland Women Exposed | | Denmark Men Unexposed | Denmark Men Exposed | Denmark Women Unexposed | Denmark Women Exposed |
|---|---|---|---|---|---|---|---|---|---|
| N | 24233 | 24233 | 24233 | 24233 | | 22812 | 22812 | 22812 | 22812 |
| Helsinki-Uusimaa | 8017 (33%) | 8017 (33%) | 8017 (33%) | 8017 (33%) | Capital city | 9014 (40%) | 9014 (40%) | 9014 (40%) | 9014 (40%) |
| North & East Finland | 6407 (26%) | 6407 (26%) | 6407 (26%) | 6407 (26%) | Central Denmark | 4087 (18%) | 4087 (18%) | 4087 (18%) | 4087 (18%) |
| South Finland | 4856 (20%) | 4856 (20%) | 4856 (20%) | 4856 (20%) | North Jutland | 1626 (7.1%) | 1626 (7.1%) | 1626 (7.1%) | 1626 (7.1%) |
| West Finland | 4953 (20%) | 4953 (20%) | 4953 (20%) | 4953 (20%) | Southern Denmark | 4609 (20%) | 4609 (20%) | 4609 (20%) | 4609 (20%) |
| | | | | | Zealand | 3476 (15%) | 3476 (15%) | 3476 (15%) | 3476 (15%) |
| Education | | | | | | | | | |
| Lower secondary or less | 7644 (32%) | 7644 (32%) | 7142 (29%) | 7142 (29%) | | 8156 (36%) | 8156 (36%) | 9821 (43%) | 9821 (43%) |
| Upper secondary | 10022 (41%) | 10022 (41%) | 10244 (42%) | 10244 (42%) | | 10967 (48%) | 10967 (48%) | 8745 (38%) | 8745 (38%) |
| Post-secondary or tertiary | 6567 (27%) | 6567 (27%) | 6847 (28%) | 6847 (28%) | | 3689 (16%) | 3689 (16%) | 4246 (19%) | 4246 (19%) |
| Other parent's education | | | | | | | | | |
| Lower secondary or less | 7142 (29%) | 7142 (29%) | 7644 (32%) | 7644 (32%) | | 9821 (43%) | 9821 (43%) | 8156 (36%) | 8156 (36%) |
| Upper secondary | 10244 (42%) | 10244 (42%) | 10022 (41%) | 10022 (41%) | | 8745 (38%) | 8745 (38%) | 10967 (48%) | 10967 (48%) |
| Post-secondary or tertiary | 6847 (28%) | 6847 (28%) | 6567 (27%) | 6567 (27%) | | 4246 (19%) | 4246 (19%) | 3689 (16%) | 3689 (16%) |
| Prior mental disorder | | | | | | | | | |
| No | 18786 (78%) | 18786 (78%) | 18556 (77%) | 18556 (77%) | | 19251 (84%) | 19251 (84%) | 18198 (80%) | 18198 (80%) |

**eTable 3. Descriptive characteristics in analyses including exposure to child's psychotic disorder (F20–F29)**

| | Finland | | | | Denmark | | | |
|---|---|---|---|---|---|---|---|---|
| Characteristic | Men Unexposed | Men Exposed | Women Unexposed | Women Exposed | Men Unexposed | Men Exposed | Women Unexposed | Women Exposed |
| N | 24233 | 24233 | 24233 | 24233 | 22812 | 22812 | 22812 | 22812 |
| Yes | 5447 (22%) | 5447 (22%) | 5677 (23%) | 5677 (23%) | 3561 (16%) | 3561 (16%) | 4614 (20%) | 4614 (20%) |
| Other parent's prior mental disorder | | | | | | | | |
| No | 18556 (77%) | 18556 (77%) | 18786 (78%) | 18786 (78%) | 18198 (80%) | 18198 (80%) | 19251 (84%) | 19251 (84%) |
| Yes | 5677 (23%) | 5677 (23%) | 5447 (22%) | 5447 (22%) | 4614 (20%) | 4614 (20%) | 3561 (16%) | 3561 (16%) |

* Thousand euros (€)

**eTable 4. Descriptive characteristics in analyses including exposure to child's mood disorder (F30–F39)**

| | Finland | | | | Denmark | | | |
|---|---|---|---|---|---|---|---|---|
| Characteristic | Men Unexposed | Men Exposed | Women Unexposed | Women Exposed | Men Unexposed | Men Exposed | Women Unexposed | Women Exposed |
| N | 118811 | 118811 | 118811 | 118811 | 50235 | 50235 | 50235 | 50235 |
| | Mean (SD) | Mean (SD) | Mean (SD) | Mean (SD) | Mean (SD) | Mean (SD) | Mean (SD) | Mean (SD) |
| Sex of child, N (%) | | | | | | | | |
| Female | 55924 (47%) | 75797 (64%) | 55924 (47%) | 75797 (64%) | 24184 (48%) | 33741 (67%) | 24184 (48%) | 33741 (67%) |
| Male | 62887 (53%) | 43014 (36%) | 62887 (53%) | 43014 (36%) | 26051 (52%) | 16494 (33%) | 26051 (52%) | 16494 (33%) |
| Child's age at diagnosis | | 18.20 (4.04) | | 18.20 (4.04) | | 19.37 (3.63) | | 19.37 (3.63) |
| Parent's age | 50.06 (7.13) | 49.84 (7.40) | 47.59 (6.59) | 47.29 (6.84) | 51.07 (6.55) | 51.03 (6.73) | 48.23 (5.70) | 48.17 (5.81) |
| Earnings* | 35.35 (29.16) | 33.04 (28.29) | 26.01 (18.84) | 24.36 (18.80) | 59.94 (40.03) | 58.46 (36.98) | 45.36 (20.17) | 44.88 (19.48) |
| Disposable income* | 30.52 (19.48) | 29.03 (18.07) | 24.81 (11.04) | 24.44 (10.70) | 38.99 (23.4) | 37.87 (21.64) | 31.83 (12.98) | 31.3 (12.42) |
| Social welfare benefits* | 4.56 (8.57) | 5.15 (8.89) | 5.51 (7.20) | 6.48 (7.69) | 4.58 (8.81) | 4.93 (9.05) | 7.23 (10.05) | 8.17 (10.55) |
| Employed, N (%) | | | | | | | | |
| No | 26589 (24%) | 29759 (27%) | 24117 (21%) | 28706 (25%) | 12885 (27%) | 13365 (29%) | 13912 (29%) | 15103 (31%) |
| Yes | 84619 (76%) | 80305 (73%) | 91447 (79%) | 86942 (75%) | 34111 (73%) | 33410 (71%) | 34642 (71%) | 33576 (69%) |
| Matching criteria | | | | | | | | |
| | N (%) | N (%) | N (%) | N (%) | N (%) | N (%) | N (%) | N (%) |
| Child's year of birth, Mean (SD) | 1991.17 (7.78) | 1991.17 (7.78) | 1991.17 (7.78) | 1991.17 (7.78) | 1990.57 (7.40) | 1990.57 (7.40) | 1990.57 (7.40) | 1990.57 (7.40) |
| Child's birth region | | | | | | | | |

**eTable 4. Descriptive characteristics in analyses including exposure to child's mood disorder (F30–F39)**

| | Finland | | | | | Denmark | | | |
|---|---|---|---|---|---|---|---|---|---|
| Characteristic | Men Unexposed | Men Exposed | Women Unexposed | Women Exposed | | Men Unexposed | Men Exposed | Women Unexposed | Women Exposed |
| N | 118811 | 118811 | 118811 | 118811 | | 50235 | 50235 | 50235 | 50235 |
| Helsinki-Uusimaa | 38434 (32%) | 38434 (32%) | 38434 (32%) | 38434 (32%) | Capital city | 15636 (31%) | 15636 (31%) | 15636 (31%) | 15636 (31%) |
| North & East Finland | 29992 (25%) | 29992 (25%) | 29992 (25%) | 29992 (25%) | Central Denmark | 12469 (25%) | 12469 (25%) | 12469 (25%) | 12469 (25%) |
| South Finland | 24534 (21%) | 24534 (21%) | 24534 (21%) | 24534 (21%) | North Jutland | 4073 (8.1%) | 4073 (8.1%) | 4073 (8.1%) | 4073 (8.1%) |
| West Finland | 25851 (22%) | 25851 (22%) | 25851 (22%) | 25851 (22%) | Southern Denmark | 11477 (23%) | 11477 (23%) | 11477 (23%) | 11477 (23%) |
| | | | | | Zealand | 6580 (13%) | 6580 (13%) | 6580 (13%) | 6580 (13%) |
| Education | | | | | | | | | |
| Lower secondary or less | 35258 (30%) | 35258 (30%) | 32104 (27%) | 32104 (27%) | | 15454 (31%) | 15454 (31%) | 18978 (38%) | 18978 (38%) |
| Upper secondary | 54235 (46%) | 54235 (46%) | 52194 (44%) | 52194 (44%) | | 26280 (52%) | 26280 (52%) | 20829 (41%) | 20829 (41%) |
| Post-secondary or tertiary | 29318 (25%) | 29318 (25%) | 34513 (29%) | 34513 (29%) | | 8501 (17%) | 8501 (17%) | 10428 (21%) | 10428 (21%) |
| Other parent's education | | | | | | | | | |
| Lower secondary or less | 32104 (27%) | 32104 (27%) | 35258 (30%) | 35258 (30%) | | 18978 (38%) | 18978 (38%) | 15454 (31%) | 15454 (31%) |
| Upper secondary | 52194 (44%) | 52194 (44%) | 54235 (46%) | 54235 (46%) | | 20829 (41%) | 20829 (41%) | 26280 (52%) | 26280 (52%) |
| Post-secondary or tertiary | 34513 (29%) | 34513 (29%) | 29318 (25%) | 29318 (25%) | | 10428 (21%) | 10428 (21%) | 8501 (17%) | 8501 (17%) |
| Prior mental disorder | | | | | | | | | |
| No | 93517 (79%) | 93517 (79%) | 91645 (77%) | 91645 (77%) | | 43747 (87%) | 43747 (87%) | 41265 (82%) | 41265 (82%) |

**eTable 4. Descriptive characteristics in analyses including exposure to child's mood disorder (F30–F39)**

| | Finland | | | | Denmark | | | |
|---|---|---|---|---|---|---|---|---|
| Characteristic | Men Unexposed | Men Exposed | Women Unexposed | Women Exposed | Men Unexposed | Men Exposed | Women Unexposed | Women Exposed |
| N | 118811 | 118811 | 118811 | 118811 | 50235 | 50235 | 50235 | 50235 |
| Yes | 25294 (21%) | 25294 (21%) | 27166 (23%) | 27166 (23%) | 6488 (13%) | 6488 (13%) | 8970 (18%) | 8970 (18%) |
| Other parent's prior mental disorder | | | | | | | | |
| No | 91645 (77%) | 91645 (77%) | 93517 (79%) | 93517 (79%) | 41265 (82%) | 41265 (82%) | 43747 (87%) | 43747 (87%) |
| Yes | 27166 (23%) | 27166 (23%) | 25294 (21%) | 25294 (21%) | 8970 (18%) | 8970 (18%) | 6488 (13%) | 6488 (13%) |

* Thousand euros (€)

**eTable 5. Descriptive characteristics in analyses including exposure to child's anxiety disorder (F40–F48)**

| | Finland | | | | Denmark | | | |
|---|---|---|---|---|---|---|---|---|
| Characteristic | Men Unexposed | Men Exposed | Women Unexposed | Women Exposed | Men Unexposed | Men Exposed | Women Unexposed | Women Exposed |
| N | 131386 | 131386 | 131386 | 131386 | 104772 | 104772 | 104772 | 104772 |
| | Mean (SD) | Mean (SD) | Mean (SD) | Mean (SD) | Mean (SD) | Mean (SD) | Mean (SD) | Mean (SD) |
| Sex of child, N (%) | | | | | | | | |
| Female | 62555 (48%) | 77752 (59%) | 62555 (48%) | 77752 (59%) | 50013 (48%) | 65468 (62%) | 50013 (48%) | 65468 (62%) |
| Male | 68831 (52%) | 53634 (41%) | 68831 (52%) | 53634 (41%) | 54759 (52%) | 39304 (38%) | 54759 (52%) | 39304 (38%) |
| Child's age at diagnosis | | 17.63 (4.54) | | 17.63 (4.54) | | 17.66 (4.55) | | 17.66 (4.55) |
| Parent's age | 49.49 (7.45) | 49.37 (7.56) | 47.01 (6.92) | 46.82 (7.01) | 49.43 (7.05) | 49.28 (7.27) | 46.58 (6.22) | 46.40 (6.39) |
| Earnings* | 35.40 (29.09) | 33.10 (28.28) | 25.75 (18.73) | 23.99 (18.62) | 58.5 (37.73) | 55.62 (33.98) | 44.71 (19.45) | 43.46 (18.27) |
| Disposable income* | 30.47 (19.33) | 29.00 (18.11) | 24.77 (11.06) | 24.30 (10.64) | 37.92 (22.03) | 35.94 (19.83) | 31.49 (12.5) | 30.53 (11.77) |
| Social welfare benefits* | 4.47 (8.44) | 5.05 (8.71) | 5.61 (7.28) | 6.58 (7.77) | 4.67 (8.85) | 5.37 (9.35) | 8.01 (10.33) | 9.56 (11.11) |
| Employed, N (%) | | | | | | | | |
| No | 28869 (23%) | 32726 (27%) | 26939 (21%) | 32177 (25%) | 27487 (28%) | 29316 (30%) | 29939 (30%) | 34592 (34%) |
| Yes | 94480 (77%) | 89336 (73%) | 100963 (79%) | 95781 (75%) | 70961 (72%) | 68648 (70%) | 71468 (70%) | 67202 (66%) |
| Matching criteria | | | | | | | | |
| | N (%) | N (%) | N (%) | N (%) | N (%) | N (%) | N (%) | N (%) |
| Child's year of birth, Mean (SD) | 1991.65 (8.44) | 1991.65 (8.44) | 1991.65 (8.44) | 1991.65 (8.44) | 1991.99 (8.49) | 1991.99 (8.49) | 1991.99 (8.49) | 1991.99 (8.49) |
| Child's birth region | | | | | | | | |

**eTable 5. Descriptive characteristics in analyses including exposure to child's anxiety disorder (F40–F48)**

| | Finland | | | | | Denmark | | | |
|---|---|---|---|---|---|---|---|---|---|
| Characteristic | Men Unexposed | Men Exposed | Women Unexposed | Women Exposed | | Men Unexposed | Men Exposed | Women Unexposed | Women Exposed |
| N | 131386 | 131386 | 131386 | 131386 | | 104772 | 104772 | 104772 | 104772 |
| Helsinki-Uusimaa | 41842 (32%) | 41842 (32%) | 41842 (32%) | 41842 (32%) | Capital city | 33856 (32%) | 33856 (32%) | 33856 (32%) | 33856 (32%) |
| North & East Finland | 32345 (25%) | 32345 (25%) | 32345 (25%) | 32345 (25%) | Central Denmark | 21421 (20%) | 21421 (20%) | 21421 (20%) | 21421 (20%) |
| South Finland | 28746 (22%) | 28746 (22%) | 28746 (22%) | 28746 (22%) | North Jutland | 8620 (8.2%) | 8620 (8.2%) | 8620 (8.2%) | 8620 (8.2%) |
| West Finland | 28453 (22%) | 28453 (22%) | 28453 (22%) | 28453 (22%) | Southern Denmark | 25634 (24%) | 25634 (24%) | 25634 (24%) | 25634 (24%) |
| | | | | | Zealand | 15241 (15%) | 15241 (15%) | 15241 (15%) | 15241 (15%) |
| Education | | | | | | | | | |
| Lower secondary or less | 39707 (30%) | 39707 (30%) | 36011 (27%) | 36011 (27%) | | 34915 (33%) | 34915 (33%) | 42283 (40%) | 42283 (40%) |
| Upper secondary | 59302 (45%) | 59302 (45%) | 56936 (43%) | 56936 (43%) | | 54456 (52%) | 54456 (52%) | 43289 (41%) | 43289 (41%) |
| Post-secondary or tertiary | 32377 (25%) | 32377 (25%) | 38439 (29%) | 38439 (29%) | | 15401 (15%) | 15401 (15%) | 19200 (18%) | 19200 (18%) |
| Other parent's education | | | | | | | | | |
| Lower secondary or less | 36011 (27%) | 36011 (27%) | 39707 (30%) | 39707 (30%) | | 42283 (40%) | 42283 (40%) | 34915 (33%) | 34915 (33%) |
| Upper secondary | 56936 (43%) | 56936 (43%) | 59302 (45%) | 59302 (45%) | | 43289 (41%) | 43289 (41%) | 54456 (52%) | 54456 (52%) |
| Post-secondary or tertiary | 38439 (29%) | 38439 (29%) | 32377 (25%) | 32377 (25%) | | 19200 (18%) | 19200 (18%) | 15401 (15%) | 15401 (15%) |
| Prior mental disorder | | | | | | | | | |
| No | 104092 (79%) | 104092 (79%) | 101865 (78%) | 101865 (78%) | | 90451 (86%) | 90451 (86%) | 85233 (81%) | 85233 (81%) |

**eTable 5. Descriptive characteristics in analyses including exposure to child's anxiety disorder (F40–F48)**

| | Finland | | | | Denmark | | | |
|---|---|---|---|---|---|---|---|---|
| Characteristic | Men Unexposed | Men Exposed | Women Unexposed | Women Exposed | Men Unexposed | Men Exposed | Women Unexposed | Women Exposed |
| N | 131386 | 131386 | 131386 | 131386 | 104772 | 104772 | 104772 | 104772 |
| Yes | 27294 (21%) | 27294 (21%) | 29521 (22%) | 29521 (22%) | 14321 (14%) | 14321 (14%) | 19539 (19%) | 19539 (19%) |
| Other parent's prior mental disorder | | | | | | | | |
| No | 101865 (78%) | 101865 (78%) | 104092 (79%) | 104092 (79%) | 85233 (81%) | 85233 (81%) | 90451 (86%) | 90451 (86%) |
| Yes | 29521 (22%) | 29521 (22%) | 27294 (21%) | 27294 (21%) | 19539 (19%) | 19539 (19%) | 14321 (14%) | 14321 (14%) |

* Thousand euros (€)

**eTable 6. Descriptive characteristics in analyses including exposure to child's eating disorder (F50)**

| Characteristic | Finland Men Unexposed | Finland Men Exposed | Finland Women Unexposed | Finland Women Exposed | Denmark Men Unexposed | Denmark Men Exposed | Denmark Women Unexposed | Denmark Women Exposed |
|---|---|---|---|---|---|---|---|---|
| N | 20738 | 20738 | 20738 | 20738 | 19465 | 19465 | 19465 | 19465 |
| | Mean (SD) | Mean (SD) | Mean (SD) | Mean (SD) | Mean (SD) | Mean (SD) | Mean (SD) | Mean (SD) |
| Sex of child, N (%) | | | | | | | | |
| Female | 9999 (48%) | 18862 (91%) | 9999 (48%) | 18862 (91%) | 9373 (48%) | 18235 (94%) | 9373 (48%) | 18235 (94%) |
| Male | 10739 (52%) | 1876 (9.0%) | 10739 (52%) | 1876 (9.0%) | 10092 (52%) | 1230 (6.3%) | 10092 (52%) | 1230 (6.3%) |
| Child's age at diagnosis | | 16.64 (3.78) | | 16.64 (3.78) | | 17.73 (3.85) | | 17.73 (3.85) |
| Parent's age | 48.85 (6.75) | 49.07 (6.89) | 46.53 (6.20) | 46.71 (6.24) | 49.80 (6.60) | 49.80 (6.54) | 47.07 (5.75) | 47.10 (5.68) |
| Earnings* | 40.37 (31.90) | 40.88 (32.93) | 28.11 (19.82) | 28.81 (20.63) | 64.32 (43.73) | 65.86 (44.75) | 47.64 (21.5) | 48.11 (21.27) |
| Disposable income* | 33.25 (21.79) | 33.72 (22.50) | 26.33 (11.82) | 26.81 (12.16) | 41.04 (25.24) | 41.7 (25.5) | 33.22 (13.75) | 33.52 (13.61) |
| Social welfare benefits* | 3.80 (8.34) | 3.98 (8.56) | 5.54 (6.98) | 5.64 (6.99) | 3.54 (7.8) | 3.5 (7.71) | 6.61 (9.21) | 6.72 (9.24) |
| Employed, N (%) | | | | | | | | |
| No | 3737 (19%) | 3804 (19%) | 3675 (18%) | 3681 (18%) | 4169 (23%) | 4083 (22%) | 4484 (24%) | 4481 (23%) |
| Yes | 15996 (81%) | 15943 (81%) | 16612 (82%) | 16732 (82%) | 14261 (77%) | 14528 (78%) | 14430 (76%) | 14594 (77%) |
| Matching criteria | | | | | | | | |
| | N (%) | N (%) | N (%) | N (%) | N (%) | N (%) | N (%) | N (%) |
| Child's year of birth, Mean (SD) | 1992.11 (7.61) | 1992.11 (7.61) | 1992.11 (7.61) | 1992.11 (7.61) | 1990.89 (8.40) | 1990.89 (8.40) | 1990.89 (8.40) | 1990.89 (8.40) |
| Child's birth region | | | | | | | | |

**eTable 6. Descriptive characteristics in analyses including exposure to child's eating disorder (F50)**

| | Finland | | | | Denmark | | | | |
|---|---|---|---|---|---|---|---|---|---|
| Characteristic | Men Unexposed | Men Exposed | Women Unexposed | Women Exposed | | Men Unexposed | Men Exposed | Women Unexposed | Women Exposed |
| N | 20738 | 20738 | 20738 | 20738 | | 19465 | 19465 | 19465 | 19465 |
| Helsinki-Uusimaa | 7035 (34%) | 7035 (34%) | 7035 (34%) | 7035 (34%) | Capital city | 6457 (33%) | 6457 (33%) | 6457 (33%) | 6457 (33%) |
| North & East Finland | 4744 (23%) | 4744 (23%) | 4744 (23%) | 4744 (23%) | Central Denmark | 4392 (23%) | 4392 (23%) | 4392 (23%) | 4392 (23%) |
| South Finland | 4122 (20%) | 4122 (20%) | 4122 (20%) | 4122 (20%) | North Jutland | 1717 (8.8%) | 1717 (8.8%) | 1717 (8.8%) | 1717 (8.8%) |
| West Finland | 4837 (23%) | 4837 (23%) | 4837 (23%) | 4837 (23%) | Southern Denmark | 4394 (23%) | 4394 (23%) | 4394 (23%) | 4394 (23%) |
| | | | | | Zealand | 2505 (13%) | 2505 (13%) | 2505 (13%) | 2505 (13%) |
| Education | | | | | | | | | |
| Lower secondary or less | 4601 (22%) | 4601 (22%) | 3777 (18%) | 3777 (18%) | | 4381 (23%) | 4381 (23%) | 5443 (28%) | 5443 (28%) |
| Upper secondary | 8689 (42%) | 8689 (42%) | 8219 (40%) | 8219 (40%) | | 10493 (54%) | 10493 (54%) | 8605 (44%) | 8605 (44%) |
| Post-secondary or tertiary | 7448 (36%) | 7448 (36%) | 8742 (42%) | 8742 (42%) | | 4591 (24%) | 4591 (24%) | 5417 (28%) | 5417 (28%) |
| Other parent's education | | | | | | | | | |
| Lower secondary or less | 3777 (18%) | 3777 (18%) | 4601 (22%) | 4601 (22%) | | 5443 (28%) | 5443 (28%) | 4381 (23%) | 4381 (23%) |
| Upper secondary | 8219 (40%) | 8219 (40%) | 8689 (42%) | 8689 (42%) | | 8605 (44%) | 8605 (44%) | 10493 (54%) | 10493 (54%) |
| Post-secondary or tertiary | 8742 (42%) | 8742 (42%) | 7448 (36%) | 7448 (36%) | | 5417 (28%) | 5417 (28%) | 4591 (24%) | 4591 (24%) |
| Prior mental disorder | | | | | | | | | |
| No | 17589 (85%) | 17589 (85%) | 17109 (83%) | 17109 (83%) | | 17776 (91%) | 17776 (91%) | 17027 (87%) | 17027 (87%) |

**eTable 6. Descriptive characteristics in analyses including exposure to child's eating disorder (F50)**

| | Finland | | | | Denmark | | | |
|---|---|---|---|---|---|---|---|---|
| Characteristic | Men Unexposed | Men Exposed | Women Unexposed | Women Exposed | Men Unexposed | Men Exposed | Women Unexposed | Women Exposed |
| N | 20738 | 20738 | 20738 | 20738 | 19465 | 19465 | 19465 | 19465 |
| Yes | 3149 (15%) | 3149 (15%) | 3629 (17%) | 3629 (17%) | 1689 (8.7%) | 1689 (8.7%) | 2438 (13%) | 2438 (13%) |
| Other parent's prior mental disorder | | | | | | | | |
| No | 17109 (83%) | 17109 (83%) | 17589 (85%) | 17589 (85%) | 17027 (87%) | 17027 (87%) | 17776 (91%) | 17776 (91%) |
| Yes | 3629 (17%) | 3629 (17%) | 3149 (15%) | 3149 (15%) | 2438 (13%) | 2438 (13%) | 1689 (8.7%) | 1689 (8.7%) |

* Thousand euros (€)

**eTable 7. Descriptive characteristics in analyses including exposure to child's personality disorder (F60)**

| | Finland | | | | Denmark | | | |
|---|---|---|---|---|---|---|---|---|
| Characteristic | Men Unexposed | Men Exposed | Women Unexposed | Women Exposed | Men Unexposed | Men Exposed | Women Unexposed | Women Exposed |
| N | 18998 | 18998 | 18998 | 18998 | 32170 | 32170 | 32170 | 32170 |
| | Mean (SD) | Mean (SD) | Mean (SD) | Mean (SD) | Mean (SD) | Mean (SD) | Mean (SD) | Mean (SD) |
| Sex of child, N (%) | | | | | | | | |
| Female | 9201 (48%) | 11905 (63%) | 9201 (48%) | 11905 (63%) | 15451 (48%) | 23526 (73%) | 15451 (48%) | 23526 (73%) |
| Male | 9797 (52%) | 7093 (37%) | 9797 (52%) | 7093 (37%) | 16719 (52%) | 8644 (27%) | 16719 (52%) | 8644 (27%) |
| Child's age at diagnosis | | 21.05 (2.63) | | 21.05 (2.63) | | 20.26 (3.05) | | 20.26 (3.05) |
| Parent's age | 52.31 (6.45) | 52.14 (6.81) | 49.86 (5.90) | 49.52 (6.19) | 51.26 (6.53) | 50.97 (6.73) | 48.33 (5.69) | 47.95 (5.82) |
| Earnings* | 31.81 (28.32) | 28.91 (27.70) | 24.24 (18.28) | 22.05 (18.37) | 54.65 (33.89) | 51.68 (31.16) | 41.99 (18.33) | 40.46 (17.13) |
| Disposable income* | 28.72 (18.30) | 26.91 (17.06) | 23.03 (10.62) | 22.27 (10.25) | 35.24 (19.23) | 33.22 (17.73) | 29.11 (11.58) | 27.94 (10.72) |
| Social welfare benefits* | 5.39 (9.17) | 6.17 (9.32) | 4.98 (7.06) | 6.03 (7.44) | 5.26 (9.27) | 6.32 (9.88) | 7.73 (10.39) | 9.6 (11.28) |
| Employed, N (%) | | | | | | | | |
| No | 5021 (29%) | 5842 (34%) | 4498 (25%) | 5559 (30%) | 8903 (30%) | 9968 (34%) | 10052 (32%) | 11823 (38%) |
| Yes | 12348 (71%) | 11211 (66%) | 13814 (75%) | 12762 (70%) | 21026 (70%) | 19681 (66%) | 20961 (68%) | 19214 (62%) |
| Matching criteria | | | | | | | | |
| | N (%) | N (%) | N (%) | N (%) | N (%) | N (%) | N (%) | N (%) |
| Child's year of birth, Mean (SD) | 1986.82 (7.58) | 1986.82 (7.58) | 1986.82 (7.58) | 1986.82 (7.58) | 1987.22 (7.77) | 1987.22 (7.77) | 1987.22 (7.77) | 1987.22 (7.77) |
| Child's birth region | | | | | | | | |

**eTable 7. Descriptive characteristics in analyses including exposure to child's personality disorder (F60)**

| | Finland | | | | | Denmark | | | |
|---|---|---|---|---|---|---|---|---|---|
| Characteristic | Men Unexposed | Men Exposed | Women Unexposed | Women Exposed | | Men Unexposed | Men Exposed | Women Unexposed | Women Exposed |
| N | 18998 | 18998 | 18998 | 18998 | | 32170 | 32170 | 32170 | 32170 |
| Helsinki-Uusimaa | 6737 (35%) | 6737 (35%) | 6737 (35%) | 6737 (35%) | Capital city | 10186 (32%) | 10186 (32%) | 10186 (32%) | 10186 (32%) |
| North & East Finland | 4301 (23%) | 4301 (23%) | 4301 (23%) | 4301 (23%) | Central Denmark | 5666 (18%) | 5666 (18%) | 5666 (18%) | 5666 (18%) |
| South Finland | 4297 (23%) | 4297 (23%) | 4297 (23%) | 4297 (23%) | North Jutland | 2605 (8.1%) | 2605 (8.1%) | 2605 (8.1%) | 2605 (8.1%) |
| West Finland | 3663 (19%) | 3663 (19%) | 3663 (19%) | 3663 (19%) | Southern Denmark | 7513 (23%) | 7513 (23%) | 7513 (23%) | 7513 (23%) |
| | | | | | Zealand | 6200 (19%) | 6200 (19%) | 6200 (19%) | 6200 (19%) |
| Education | | | | | | | | | |
| Lower secondary or less | 6663 (35%) | 6663 (35%) | 6448 (34%) | 6448 (34%) | | 12469 (39%) | 12469 (39%) | 15982 (50%) | 15982 (50%) |
| Upper secondary | 8233 (43%) | 8233 (43%) | 8079 (43%) | 8079 (43%) | | 15729 (49%) | 15729 (49%) | 11580 (36%) | 11580 (36%) |
| Post-secondary or tertiary | 4102 (22%) | 4102 (22%) | 4471 (24%) | 4471 (24%) | | 3972 (12%) | 3972 (12%) | 4608 (14%) | 4608 (14%) |
| Other parent's education | | | | | | | | | |
| Lower secondary or less | 6448 (34%) | 6448 (34%) | 6663 (35%) | 6663 (35%) | | 15982 (50%) | 15982 (50%) | 12469 (39%) | 12469 (39%) |
| Upper secondary | 8079 (43%) | 8079 (43%) | 8233 (43%) | 8233 (43%) | | 11580 (36%) | 11580 (36%) | 15729 (49%) | 15729 (49%) |
| Post-secondary or tertiary | 4471 (24%) | 4471 (24%) | 4102 (22%) | 4102 (22%) | | 4608 (14%) | 4608 (14%) | 3972 (12%) | 3972 (12%) |
| Prior mental disorder | | | | | | | | | |
| No | 14290 (75%) | 14290 (75%) | 14070 (74%) | 14070 (74%) | | 27231 (85%) | 27231 (85%) | 25670 (80%) | 25670 (80%) |

**eTable 7. Descriptive characteristics in analyses including exposure to child's personality disorder (F60)**

| | Finland | | | | Denmark | | | |
|---|---|---|---|---|---|---|---|---|
| Characteristic | Men Unexposed | Men Exposed | Women Unexposed | Women Exposed | Men Unexposed | Men Exposed | Women Unexposed | Women Exposed |
| N | 18998 | 18998 | 18998 | 18998 | 32170 | 32170 | 32170 | 32170 |
| Yes | 4708 (25%) | 4708 (25%) | 4928 (26%) | 4928 (26%) | 4939 (15%) | 4939 (15%) | 6500 (20%) | 6500 (20%) |
| Other parent's prior mental disorder | | | | | | | | |
| No | 14070 (74%) | 14070 (74%) | 14290 (75%) | 14290 (75%) | 25670 (80%) | 25670 (80%) | 27231 (85%) | 27231 (85%) |
| Yes | 4928 (26%) | 4928 (26%) | 4708 (25%) | 4708 (25%) | 6500 (20%) | 6500 (20%) | 4939 (15%) | 4939 (15%) |

* Thousand euros (€)

**eTable 8. Descriptive characteristics in analyses including exposure to child's intellectual disability (F70–F79)**

| | Finland | | | | Denmark | | | |
|---|---|---|---|---|---|---|---|---|
| Characteristic | Men Unexposed | Men Exposed | Women Unexposed | Women Exposed | Men Unexposed | Men Exposed | Women Unexposed | Women Exposed |
| N | 14576 | 14576 | 14576 | 14576 | 14189 | 14189 | 14189 | 14189 |
| | Mean (SD) | Mean (SD) | Mean (SD) | Mean (SD) | Mean (SD) | Mean (SD) | Mean (SD) | Mean (SD) |
| Sex of child, N (%) | | | | | | | | |
| Female | 7103 (49%) | 5631 (39%) | 7103 (49%) | 5631 (39%) | 6937 (49%) | 5011 (35%) | 6937 (49%) | 5011 (35%) |
| Male | 7473 (51%) | 8945 (61%) | 7473 (51%) | 8945 (61%) | 7252 (51%) | 9178 (65%) | 7252 (51%) | 9178 (65%) |
| Child's age at diagnosis | | 9.06 (5.97) | | 9.06 (5.97) | | 12.12 (6.30) | | 12.12 (6.30) |
| Parent's age | 41.11 (8.47) | 42.24 (8.69) | 38.51 (7.95) | 39.20 (8.02) | 43.91 (8.31) | 44.40 (8.41) | 41.00 (7.58) | 41.19 (7.61) |
| Earnings* | 32.68 (24.08) | 28.64 (23.87) | 18.86 (16.50) | 15.81 (16.05) | 51.78 (29.22) | 47.04 (25.84) | 40.96 (16) | 37.89 (15.09) |
| Disposable income* | 27.52 (15.27) | 25.61 (14.58) | 22.72 (9.63) | 21.29 (9.62) | 33.04 (16.86) | 30.36 (15.01) | 29.11 (10.28) | 27.14 (9.76) |
| Social welfare benefits* | 3.57 (6.89) | 4.73 (7.88) | 8.69 (7.67) | 9.68 (7.82) | 5.23 (9.17) | 6.99 (10.21) | 11.01 (11.06) | 13.44 (11.74) |
| Employed, N (%) | | | | | | | | |
| No | 2836 (20%) | 3855 (27%) | 4403 (31%) | 5900 (41%) | 4084 (30%) | 4894 (36%) | 5042 (36%) | 6553 (47%) |
| Yes | 11245 (80%) | 10261 (73%) | 9973 (69%) | 8492 (59%) | 9480 (70%) | 8662 (64%) | 8809 (64%) | 7393 (53%) |
| Matching criteria | | | | | | | | |
| | N (%) | N (%) | N (%) | N (%) | N (%) | N (%) | N (%) | N (%) |
| Child's year of birth, Mean (SD) | 1998.49 (8.98) | 1998.49 (8.98) | 1998.49 (8.98) | 1998.49 (8.98) | 1996.56 (8.55) | 1996.56 (8.55) | 1996.56 (8.55) | 1996.56 (8.55) |
| Child's birth region | | | | | | | | |

**eTable 8. Descriptive characteristics in analyses including exposure to child's intellectual disability (F70–F79)**

| | Finland | | | | | Denmark | | | |
|---|---|---|---|---|---|---|---|---|---|
| Characteristic | Men Unexposed | Men Exposed | Women Unexposed | Women Exposed | | Men Unexposed | Men Exposed | Women Unexposed | Women Exposed |
| N | 14576 | 14576 | 14576 | 14576 | | 14189 | 14189 | 14189 | 14189 |
| Helsinki-Uusimaa | 3583 (25%) | 3583 (25%) | 3583 (25%) | 3583 (25%) | Capital city | 4129 (29%) | 4129 (29%) | 4129 (29%) | 4129 (29%) |
| North & East Finland | 4578 (31%) | 4578 (31%) | 4578 (31%) | 4578 (31%) | Central Denmark | 3151 (22%) | 3151 (22%) | 3151 (22%) | 3151 (22%) |
| South Finland | 3166 (22%) | 3166 (22%) | 3166 (22%) | 3166 (22%) | North Jutland | 1253 (8.8%) | 1253 (8.8%) | 1253 (8.8%) | 1253 (8.8%) |
| West Finland | 3249 (22%) | 3249 (22%) | 3249 (22%) | 3249 (22%) | Southern Denmark | 3566 (25%) | 3566 (25%) | 3566 (25%) | 3566 (25%) |
| | | | | | Zealand | 2090 (15%) | 2090 (15%) | 2090 (15%) | 2090 (15%) |
| Education | | | | | | | | | |
| Lower secondary or less | 4437 (30%) | 4437 (30%) | 4079 (28%) | 4079 (28%) | | 6320 (45%) | 6320 (45%) | 7222 (51%) | 7222 (51%) |
| Upper secondary | 7150 (49%) | 7150 (49%) | 6700 (46%) | 6700 (46%) | | 6508 (46%) | 6508 (46%) | 5119 (36%) | 5119 (36%) |
| Post-secondary or tertiary | 2989 (21%) | 2989 (21%) | 3797 (26%) | 3797 (26%) | | 1361 (9.6%) | 1361 (9.6%) | 1848 (13%) | 1848 (13%) |
| Other parent's education | | | | | | | | | |
| Lower secondary or less | 4079 (28%) | 4079 (28%) | 4437 (30%) | 4437 (30%) | | 7222 (51%) | 7222 (51%) | 6320 (45%) | 6320 (45%) |
| Upper secondary | 6700 (46%) | 6700 (46%) | 7150 (49%) | 7150 (49%) | | 5119 (36%) | 5119 (36%) | 6508 (46%) | 6508 (46%) |
| Post-secondary or tertiary | 3797 (26%) | 3797 (26%) | 2989 (21%) | 2989 (21%) | | 1848 (13%) | 1848 (13%) | 1361 (9.6%) | 1361 (9.6%) |
| Prior mental disorder | | | | | | | | | |
| No | 12040 (83%) | 12040 (83%) | 11753 (81%) | 11753 (81%) | | 12132 (86%) | 12132 (86%) | 11546 (81%) | 11546 (81%) |

**eTable 8. Descriptive characteristics in analyses including exposure to child's intellectual disability (F70–F79)**

| | Finland | | | | Denmark | | | |
|---|---|---|---|---|---|---|---|---|
| Characteristic | Men Unexposed | Men Exposed | Women Unexposed | Women Exposed | Men Unexposed | Men Exposed | Women Unexposed | Women Exposed |
| N | 14576 | 14576 | 14576 | 14576 | 14189 | 14189 | 14189 | 14189 |
| Yes | 2536 (17%) | 2536 (17%) | 2823 (19%) | 2823 (19%) | 2057 (14%) | 2057 (14%) | 2643 (19%) | 2643 (19%) |
| Other parent's prior mental disorder | | | | | | | | |
| No | 11753 (81%) | 11753 (81%) | 12040 (83%) | 12040 (83%) | 11546 (81%) | 11546 (81%) | 12132 (86%) | 12132 (86%) |
| Yes | 2823 (19%) | 2823 (19%) | 2536 (17%) | 2536 (17%) | 2643 (19%) | 2643 (19%) | 2057 (14%) | 2057 (14%) |

* Thousand euros (€)

**eTable 9. Descriptive characteristics in analyses including exposure to child's developmental disorder (F84)**

| | Finland | | | | Denmark | | | |
|---|---|---|---|---|---|---|---|---|
| Characteristic | Men Unexposed | Men Exposed | Women Unexposed | Women Exposed | Men Unexposed | Men Exposed | Women Unexposed | Women Exposed |
| N | 18978 | 18978 | 18978 | 18978 | 38065 | 38065 | 38065 | 38065 |
| | Mean (SD) | Mean (SD) | Mean (SD) | Mean (SD) | Mean (SD) | Mean (SD) | Mean (SD) | Mean (SD) |
| Sex of child, N (%) | | | | | | | | |
| Female | 9352 (49%) | 4526 (24%) | 9352 (49%) | 4526 (24%) | 18647 (49%) | 10258 (27%) | 18647 (49%) | 10258 (27%) |
| Male | 9626 (51%) | 14452 (76%) | 9626 (51%) | 14452 (76%) | 19418 (51%) | 27807 (73%) | 19418 (51%) | 27807 (73%) |
| Child's age at diagnosis | | 9.64 (5.61) | | 9.64 (5.61) | | 10.85 (5.41) | | 10.85 (5.41) |
| Parent's age | 42.23 (8.25) | 42.76 (8.48) | 39.70 (7.69) | 39.98 (7.83) | 43.79 (7.66) | 43.91 (7.86) | 41.08 (6.93) | 41.11 (7.07) |
| Earnings* | 37.37 (27.36) | 33.86 (26.19) | 22.64 (18.66) | 19.63 (18.05) | 60.44 (37.59) | 57.5 (33.12) | 46.86 (20.31) | 44.65 (18.84) |
| Disposable income* | 30.54 (17.65) | 28.43 (16.19) | 24.92 (10.80) | 23.21 (10.48) | 38.87 (21.97) | 37 (19.47) | 33.49 (13.06) | 31.98 (12.21) |
| Social welfare benefits* | 3.34 (6.87) | 4.02 (7.31) | 8.15 (7.72) | 8.74 (7.63) | 3.81 (8.03) | 4.14 (8.27) | 9.39 (10.07) | 10.51 (10.6) |
| Employed, N (%) | | | | | | | | |
| No | 3238 (18%) | 4052 (22%) | 4707 (25%) | 6038 (32%) | 8880 (24%) | 9029 (24%) | 10445 (28%) | 12234 (32%) |
| Yes | 15154 (82%) | 14355 (78%) | 13994 (75%) | 12778 (68%) | 27725 (76%) | 27990 (76%) | 26643 (72%) | 25485 (68%) |
| Matching criteria | | | | | | | | |
| | N (%) | N (%) | N (%) | N (%) | N (%) | N (%) | N (%) | N (%) |
| Child's year of birth, Mean (SD) | 2000.00 (7.87) | 2000.00 (7.87) | 2000.00 (7.87) | 2000.00 (7.87) | 2000.43 (7.24) | 2000.43 (7.24) | 2000.43 (7.24) | 2000.43 (7.24) |
| Child's birth region | | | | | | | | |

**eTable 9. Descriptive characteristics in analyses including exposure to child's developmental disorder (F84)**

| | Finland | | | | | Denmark | | | |
|---|---|---|---|---|---|---|---|---|---|
| Characteristic | Men Unexposed | Men Exposed | Women Unexposed | Women Exposed | | Men Unexposed | Men Exposed | Women Unexposed | Women Exposed |
| N | 18978 | 18978 | 18978 | 18978 | | 38065 | 38065 | 38065 | 38065 |
| Helsinki-Uusimaa | 5963 (31%) | 5963 (31%) | 5963 (31%) | 5963 (31%) | Capital city | 14482 (38%) | 14482 (38%) | 14482 (38%) | 14482 (38%) |
| North & East Finland | 4333 (23%) | 4333 (23%) | 4333 (23%) | 4333 (23%) | Central Denmark | 8664 (23%) | 8664 (23%) | 8664 (23%) | 8664 (23%) |
| South Finland | 4042 (21%) | 4042 (21%) | 4042 (21%) | 4042 (21%) | North Jutland | 3509 (9.2%) | 3509 (9.2%) | 3509 (9.2%) | 3509 (9.2%) |
| West Finland | 4640 (24%) | 4640 (24%) | 4640 (24%) | 4640 (24%) | Southern Denmark | 5664 (15%) | 5664 (15%) | 5664 (15%) | 5664 (15%) |
| | | | | | Zealand | 5746 (15%) | 5746 (15%) | 5746 (15%) | 5746 (15%) |
| Education | | | | | | | | | |
| Lower secondary or less | 4455 (23%) | 4455 (23%) | 3873 (20%) | 3873 (20%) | | 10044 (26%) | 10044 (26%) | 10419 (27%) | 10419 (27%) |
| Upper secondary | 8823 (46%) | 8823 (46%) | 8069 (43%) | 8069 (43%) | | 20137 (53%) | 20137 (53%) | 17219 (45%) | 17219 (45%) |
| Post-secondary or tertiary | 5700 (30%) | 5700 (30%) | 7036 (37%) | 7036 (37%) | | 7884 (21%) | 7884 (21%) | 10427 (27%) | 10427 (27%) |
| Other parent's education | | | | | | | | | |
| Lower secondary or less | 3873 (20%) | 3873 (20%) | 4455 (23%) | 4455 (23%) | | 10419 (27%) | 10419 (27%) | 10044 (26%) | 10044 (26%) |
| Upper secondary | 8069 (43%) | 8069 (43%) | 8823 (46%) | 8823 (46%) | | 17219 (45%) | 17219 (45%) | 20137 (53%) | 20137 (53%) |
| Post-secondary or tertiary | 7036 (37%) | 7036 (37%) | 5700 (30%) | 5700 (30%) | | 10427 (27%) | 10427 (27%) | 7884 (21%) | 7884 (21%) |
| Prior mental disorder | | | | | | | | | |
| No | 15599 (82%) | 15599 (82%) | 14608 (77%) | 14608 (77%) | | 33894 (89%) | 33894 (89%) | 31675 (83%) | 31675 (83%) |

**eTable 9. Descriptive characteristics in analyses including exposure to child's developmental disorder (F84)**

| | Finland | | | | Denmark | | | |
|---|---|---|---|---|---|---|---|---|
| Characteristic | Men Unexposed | Men Exposed | Women Unexposed | Women Exposed | Men Unexposed | Men Exposed | Women Unexposed | Women Exposed |
| N | 18978 | 18978 | 18978 | 18978 | 38065 | 38065 | 38065 | 38065 |
| Yes | 3379 (18%) | 3379 (18%) | 4370 (23%) | 4370 (23%) | 4171 (11%) | 4171 (11%) | 6390 (17%) | 6390 (17%) |
| Other parent's prior mental disorder | | | | | | | | |
| No | 14608 (77%) | 14608 (77%) | 15599 (82%) | 15599 (82%) | 31675 (83%) | 31675 (83%) | 33894 (89%) | 33894 (89%) |
| Yes | 4370 (23%) | 4370 (23%) | 3379 (18%) | 3379 (18%) | 6390 (17%) | 6390 (17%) | 4171 (11%) | 4171 (11%) |

* Thousand euros (€)

**eTable 10. Descriptive characteristics in analyses including exposure to child's childhood onset disorder (F90–F98)**

| Characteristic | Finland Men Unexposed | Finland Men Exposed | Finland Women Unexposed | Finland Women Exposed | Denmark Men Unexposed | Denmark Men Exposed | Denmark Women Unexposed | Denmark Women Exposed |
|---|---|---|---|---|---|---|---|---|
| N | 115128 | 115128 | 115128 | 115128 | 77967 | 77967 | 77967 | 77967 |
| | Mean (SD) | Mean (SD) | Mean (SD) | Mean (SD) | Mean (SD) | Mean (SD) | Mean (SD) | Mean (SD) |
| Sex of child, N (%) | | | | | | | | |
| Female | 57752 (50%) | 43656 (38%) | 57752 (50%) | 43656 (38%) | 38444 (49%) | 26822 (34%) | 38444 (49%) | 26822 (34%) |
| Male | 57376 (50%) | 71472 (62%) | 57376 (50%) | 71472 (62%) | 39523 (51%) | 51145 (66%) | 39523 (51%) | 51145 (66%) |
| Child's age at diagnosis | | 10.09 (4.68) | | 10.09 (4.68) | | 11.70 (5.15) | | 11.70 (5.15) |
| Parent's age | 42.44 (7.84) | 42.09 (8.02) | 39.83 (7.20) | 39.34 (7.33) | 44.06 (7.68) | 43.51 (7.75) | 41.22 (6.93) | 40.67 (7.02) |
| Earnings* | 35.62 (26.49) | 32.81 (25.83) | 22.35 (17.91) | 20.33 (17.71) | 55.6 (33.32) | 52.5 (29.74) | 43.73 (17.81) | 42.27 (16.51) |
| Disposable income* | 29.59 (17.23) | 27.87 (16.19) | 24.56 (10.28) | 24.09 (9.95) | 35.73 (19.46) | 33.63 (17.51) | 31.3 (11.56) | 30.16 (10.69) |
| Social welfare benefits* | 3.50 (6.94) | 4.08 (7.20) | 7.96 (7.54) | 9.08 (8.04) | 4.73 (8.87) | 5.46 (9.31) | 10.45 (10.81) | 12.19 (11.45) |
| Employed, N (%) | | | | | | | | |
| No | 21255 (19%) | 25568 (23%) | 28042 (25%) | 34358 (30%) | 20982 (28%) | 22622 (30%) | 24982 (33%) | 29084 (38%) |
| Yes | 89876 (81%) | 85291 (77%) | 85137 (75%) | 79496 (70%) | 53774 (72%) | 52351 (70%) | 51044 (67%) | 47860 (62%) |
| Matching criteria | | | | | | | | |
| | N (%) | N (%) | N (%) | N (%) | N (%) | N (%) | N (%) | N (%) |
| Child's year of birth, Mean (SD) | 1999.03 (7.88) | 1999.03 (7.88) | 1999.03 (7.88) | 1999.03 (7.88) | 1998.42 (7.42) | 1998.42 (7.42) | 1998.42 (7.42) | 1998.42 (7.42) |
| Child's birth region | | | | | | | | |

**eTable 10. Descriptive characteristics in analyses including exposure to child's childhood onset disorder (F90–F98)**

| | Finland | | | | | Denmark | | | |
|---|---|---|---|---|---|---|---|---|---|
| Characteristic | Men Unexposed | Men Exposed | Women Unexposed | Women Exposed | | Men Unexposed | Men Exposed | Women Unexposed | Women Exposed |
| N | 115128 | 115128 | 115128 | 115128 | | 77967 | 77967 | 77967 | 77967 |
| Helsinki-Uusimaa | 36472 (32%) | 36472 (32%) | 36472 (32%) | 36472 (32%) | Capital city | 23079 (30%) | 23079 (30%) | 23079 (30%) | 23079 (30%) |
| North & East Finland | 27799 (24%) | 27799 (24%) | 27799 (24%) | 27799 (24%) | Central Denmark | 19432 (25%) | 19432 (25%) | 19432 (25%) | 19432 (25%) |
| South Finland | 26563 (23%) | 26563 (23%) | 26563 (23%) | 26563 (23%) | North Jutland | 6598 (8.5%) | 6598 (8.5%) | 6598 (8.5%) | 6598 (8.5%) |
| West Finland | 24294 (21%) | 24294 (21%) | 24294 (21%) | 24294 (21%) | Southern Denmark | 17998 (23%) | 17998 (23%) | 17998 (23%) | 17998 (23%) |
| | | | | | Zealand | 10860 (14%) | 10860 (14%) | 10860 (14%) | 10860 (14%) |
| Education | | | | | | | | | |
| Lower secondary or less | 33470 (29%) | 33470 (29%) | 29711 (26%) | 29711 (26%) | | 28425 (36%) | 28425 (36%) | 31034 (40%) | 31034 (40%) |
| Upper secondary | 55035 (48%) | 55035 (48%) | 50114 (44%) | 50114 (44%) | | 39979 (51%) | 39979 (51%) | 33114 (42%) | 33114 (42%) |
| Post-secondary or tertiary | 26623 (23%) | 26623 (23%) | 35303 (31%) | 35303 (31%) | | 9563 (12%) | 9563 (12%) | 13819 (18%) | 13819 (18%) |
| Other parent's education | | | | | | | | | |
| Lower secondary or less | 29711 (26%) | 29711 (26%) | 33470 (29%) | 33470 (29%) | | 31034 (40%) | 31034 (40%) | 28425 (36%) | 28425 (36%) |
| Upper secondary | 50114 (44%) | 50114 (44%) | 55035 (48%) | 55035 (48%) | | 33114 (42%) | 33114 (42%) | 39979 (51%) | 39979 (51%) |
| Post-secondary or tertiary | 35303 (31%) | 35303 (31%) | 26623 (23%) | 26623 (23%) | | 13819 (18%) | 13819 (18%) | 9563 (12%) | 9563 (12%) |
| Prior mental disorder | | | | | | | | | |
| No | 91846 (80%) | 91846 (80%) | 87519 (76%) | 87519 (76%) | | 67010 (86%) | 67010 (86%) | 62345 (80%) | 62345 (80%) |

**eTable 10. Descriptive characteristics in analyses including exposure to child's childhood onset disorder (F90–F98)**

| | Finland | | | | Denmark | | | |
|---|---|---|---|---|---|---|---|---|
| Characteristic | Men Unexposed | Men Exposed | Women Unexposed | Women Exposed | Men Unexposed | Men Exposed | Women Unexposed | Women Exposed |
| N | 115128 | 115128 | 115128 | 115128 | 77967 | 77967 | 77967 | 77967 |
| Yes | 23282 (20%) | 23282 (20%) | 27609 (24%) | 27609 (24%) | 10957 (14%) | 10957 (14%) | 15622 (20%) | 15622 (20%) |
| Other parent's prior mental disorder | | | | | | | | |
| No | 87519 (76%) | 87519 (76%) | 91846 (80%) | 91846 (80%) | 62345 (80%) | 62345 (80%) | 67010 (86%) | 67010 (86%) |
| Yes | 27609 (24%) | 27609 (24%) | 23282 (20%) | 23282 (20%) | 15622 (20%) | 15622 (20%) | 10957 (14%) | 10957 (14%) |

* Thousand euros (€)

### Average and total amount of follow-up years during the observation period in each analysis

**eTable 11. Average and total amount of follow-up years during the observation period in each analysis**

| | | Finland | | | | Denmark | | | |
|---|---|---|---|---|---|---|---|---|---|
| | | Men | | Women | | Men | | Women | |
| | | Total | Mean | Total | Mean | Total | Mean | Total | Mean |
| Any mental disorder | Earnings | 6169556 | 9.3 | 6533178 | 9.9 | 3999028 | 9.3 | 4208014 | 9.8 |
| | Disposable income | 5908072 | 8.9 | 6257176 | 9.4 | 3999028 | 9.3 | 4208014 | 9.8 |
| | Social welfare benefits | 5908072 | 8.9 | 6257176 | 9.4 | 3999028 | 9.3 | 4208014 | 9.8 |
| | Employment | 6169622 | 9.3 | 6532938 | 9.9 | 3999176 | 9.3 | 4208316 | 9.8 |
| Substance use disorders | Earnings | 891712 | 8.5 | 995764 | 9.5 | 412900 | 8.6 | 452960 | 9.5 |
| | Disposable income | 862324 | 8.2 | 963032 | 9.2 | 412900 | 8.6 | 452960 | 9.5 |
| | Social welfare benefits | 862324 | 8.2 | 963032 | 9.2 | 412900 | 8.6 | 452960 | 9.5 |
| | Employment | 891700 | 8.5 | 995704 | 9.5 | 412900 | 8.6 | 452964 | 9.5 |
| Psychotic disorders | Earnings | 427558 | 8.8 | 471114 | 9.7 | 392934 | 8.6 | 426300 | 9.3 |
| | Disposable income | 408058 | 8.4 | 449938 | 9.3 | 392934 | 8.6 | 426300 | 9.3 |
| | Social welfare benefits | 408058 | 8.4 | 449938 | 9.3 | 392934 | 8.6 | 426300 | 9.3 |
| | Employment | 427560 | 8.8 | 471088 | 9.7 | 392944 | 8.6 | 426382 | 9.3 |
| Mood disorders | Earnings | 2077090 | 8.7 | 2260528 | 9.5 | 884338 | 8.8 | 948186 | 9.4 |
| | Disposable income | 2039332 | 8.6 | 2219494 | 9.3 | 884338 | 8.8 | 948186 | 9.4 |
| | Social welfare benefits | 2039332 | 8.6 | 2219494 | 9.3 | 884338 | 8.8 | 948186 | 9.4 |
| | Employment | 2077028 | 8.7 | 2260334 | 9.5 | 884366 | 8.8 | 948244 | 9.4 |
| Anxiety disorders | Earnings | 2305104 | 8.8 | 2497108 | 9.5 | 1851312 | 8.8 | 1975528 | 9.4 |
| | Disposable income | 2240288 | 8.5 | 2427296 | 9.2 | 1851312 | 8.8 | 1975528 | 9.4 |
| | Social welfare benefits | 2240288 | 8.5 | 2427296 | 9.2 | 1851312 | 8.8 | 1975528 | 9.4 |
| | Employment | 2305034 | 8.8 | 2496986 | 9.5 | 1851442 | 8.8 | 1975622 | 9.4 |
| Eating disorders | Earnings | 383462 | 9.2 | 407692 | 9.8 | 358652 | 9.2 | 376636 | 9.7 |
| | Disposable income | 375048 | 9 | 398800 | 9.6 | 358652 | 9.2 | 376636 | 9.7 |
| | Social welfare benefits | 375048 | 9 | 398800 | 9.6 | 358652 | 9.2 | 376636 | 9.7 |
| | Employment | 383468 | 9.2 | 407676 | 9.8 | 358674 | 9.2 | 376650 | 9.7 |
| Personality disorders | Earnings | 318544 | 8.4 | 359838 | 9.5 | 572632 | 8.9 | 619926 | 9.6 |
| | Disposable income | 303124 | 8 | 342312 | 9 | 572632 | 8.9 | 619926 | 9.6 |

**eTable 11. Average and total amount of follow-up years during the observation period in each analysis**

| | | Finland | | | | Denmark | | | |
|---|---|---|---|---|---|---|---|---|---|
| | | Men | | Women | | Men | | Women | |
| | | Total | Mean | Total | Mean | Total | Mean | Total | Mean |
| | Social welfare benefits | 303124 | 8 | 342312 | 9 | 572632 | 8.9 | 619926 | 9.6 |
| | Employment | 318554 | 8.4 | 359854 | 9.5 | 572638 | 8.9 | 619940 | 9.6 |
| Intellectual disability | Earnings | 281558 | 9.7 | 292506 | 10 | 271856 | 9.6 | 285058 | 10 |
| | Disposable income | 266878 | 9.2 | 277434 | 9.5 | 271856 | 9.6 | 285058 | 10 |
| | Social welfare benefits | 266878 | 9.2 | 277434 | 9.5 | 271856 | 9.6 | 285058 | 10 |
| | Employment | 281566 | 9.7 | 292500 | 10 | 271880 | 9.6 | 285102 | 10 |
| Developmental disorders | Earnings | 352006 | 9.3 | 365166 | 9.6 | 720490 | 9.5 | 742166 | 9.7 |
| | Disposable income | 341994 | 9 | 354916 | 9.4 | 720490 | 9.5 | 742166 | 9.7 |
| | Social welfare benefits | 341994 | 9 | 354916 | 9.4 | 720490 | 9.5 | 742166 | 9.7 |
| | Employment | 351972 | 9.3 | 365138 | 9.6 | 720550 | 9.5 | 742210 | 9.7 |
| Childhood onset disorder | Earnings | 2157862 | 9.4 | 2252936 | 9.8 | 1485932 | 9.5 | 1547454 | 9.9 |
| | Disposable income | 2110644 | 9.2 | 2204256 | 9.6 | 1485932 | 9.5 | 1547454 | 9.9 |
| | Social welfare benefits | 2110644 | 9.2 | 2204256 | 9.6 | 1485932 | 9.5 | 1547454 | 9.9 |
| | Employment | 2157790 | 9.4 | 2252888 | 9.8 | 1486052 | 9.5 | 1547554 | 9.9 |

### Marginal predictions of annual income and employment among parents exposed to child's mental disorder and unexposed parents in Finland and Denmark

#### eFigure 1. Marginal predictions of annual disposable income among parents exposed to child's mental disorder (F00–F99) and unexposed parents

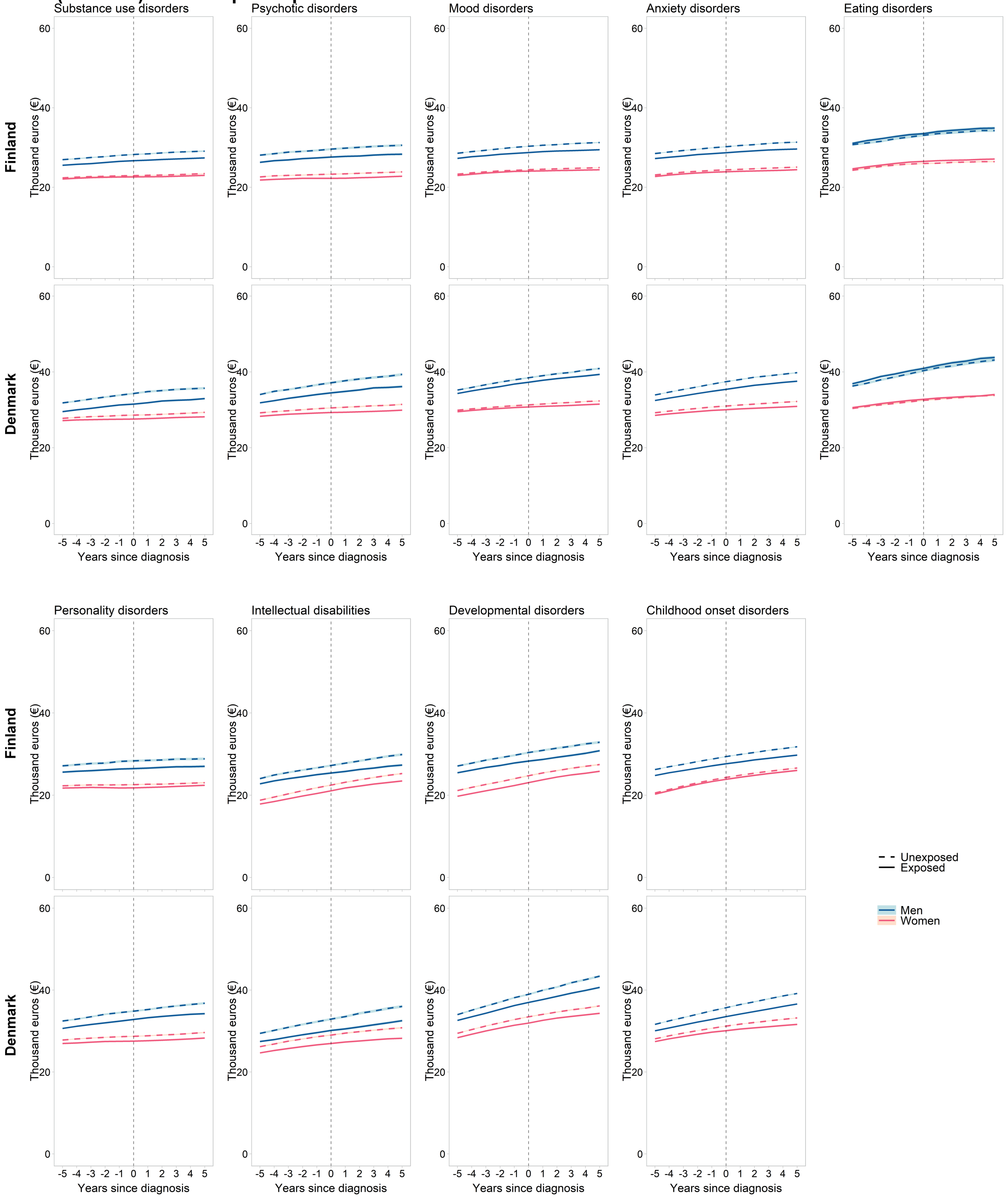

**eFigure 2. Marginal predictions of annual social welfare benefits among parents exposed to child's mental disorder (F00–F99) and unexposed parents**

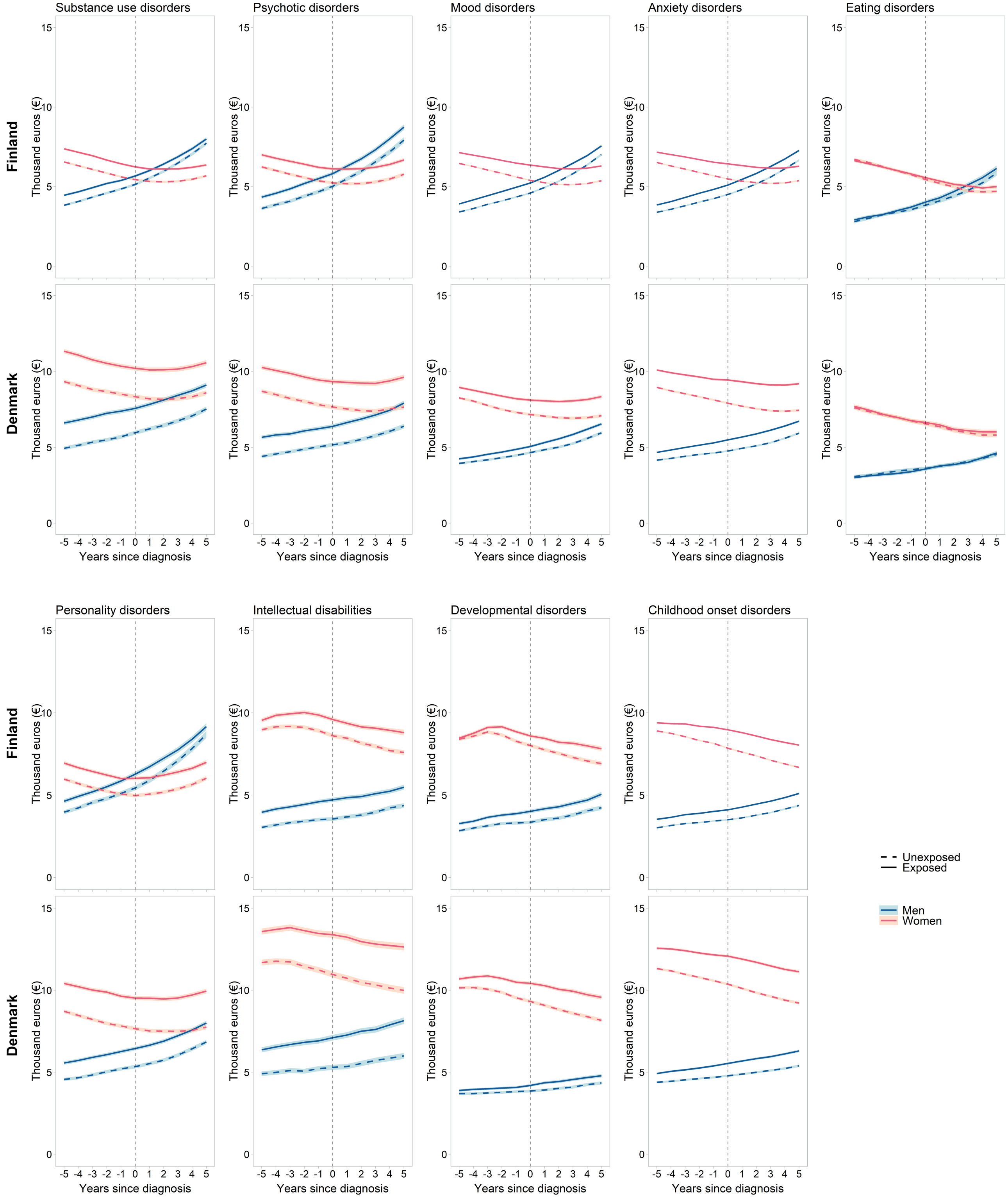

**eFigure 3. Marginal predictions of annual employment among parents exposed to child's mental disorder (F00–F99) and unexposed parents**

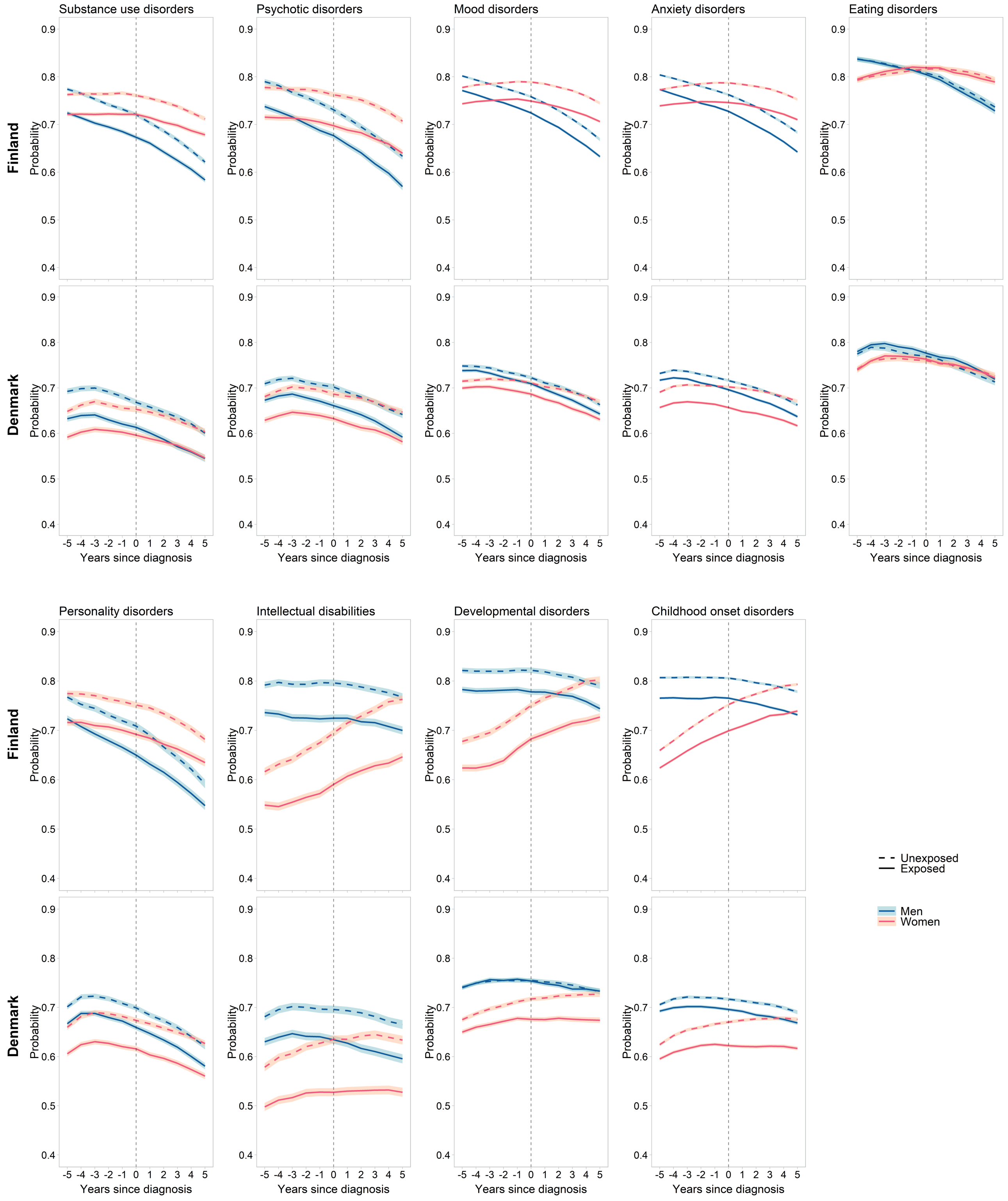

### Marginal predictions of annual income and employment among parents exposed to child's mental disorder and unexposed parents stratified by the child's age at diagnosis in Finland and Denmark

**eFigure 4. Marginal predictions of annual disposable income among parents exposed to child's any mental disorder (F00–F99) and unexposed parents stratified by the child's age at diagnosis**

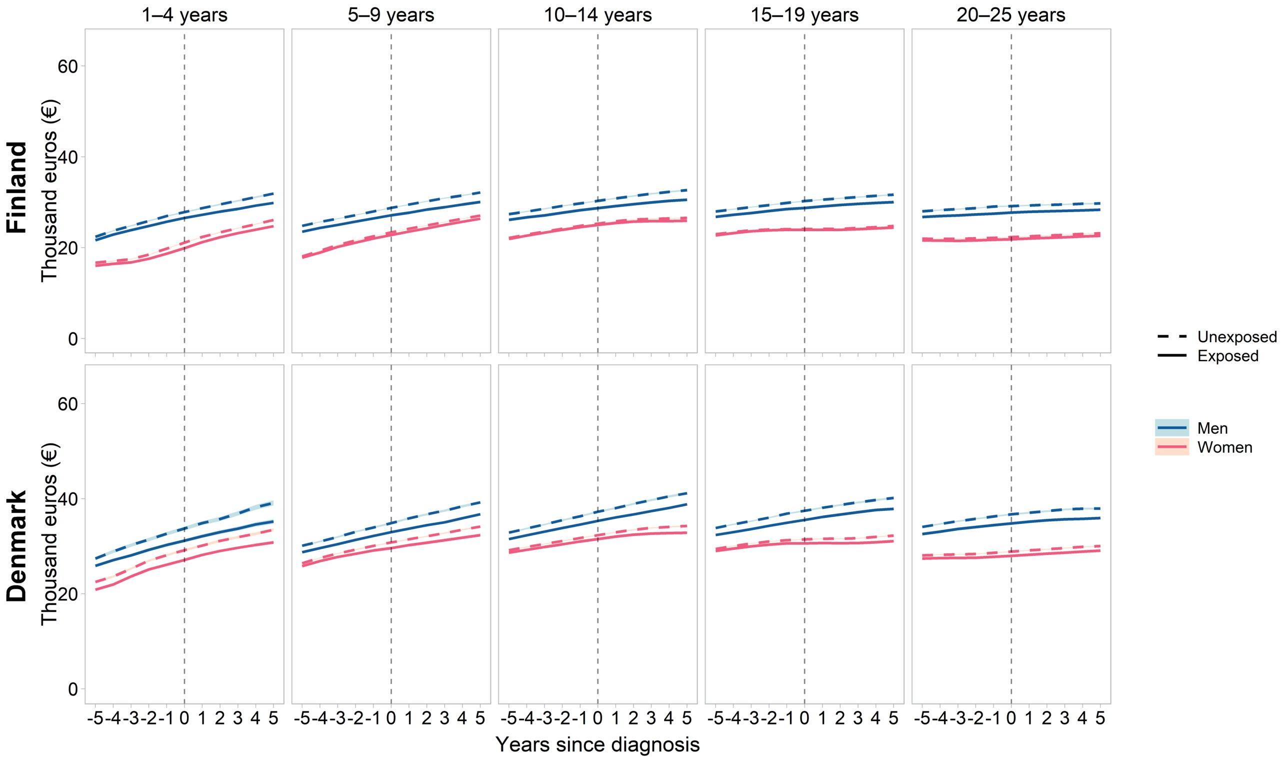

**eFigure 5. Marginal predictions of annual social welfare benefits among parents exposed to child's any mental disorder (F00–F99) and unexposed parents stratified by the child's age at diagnosis**

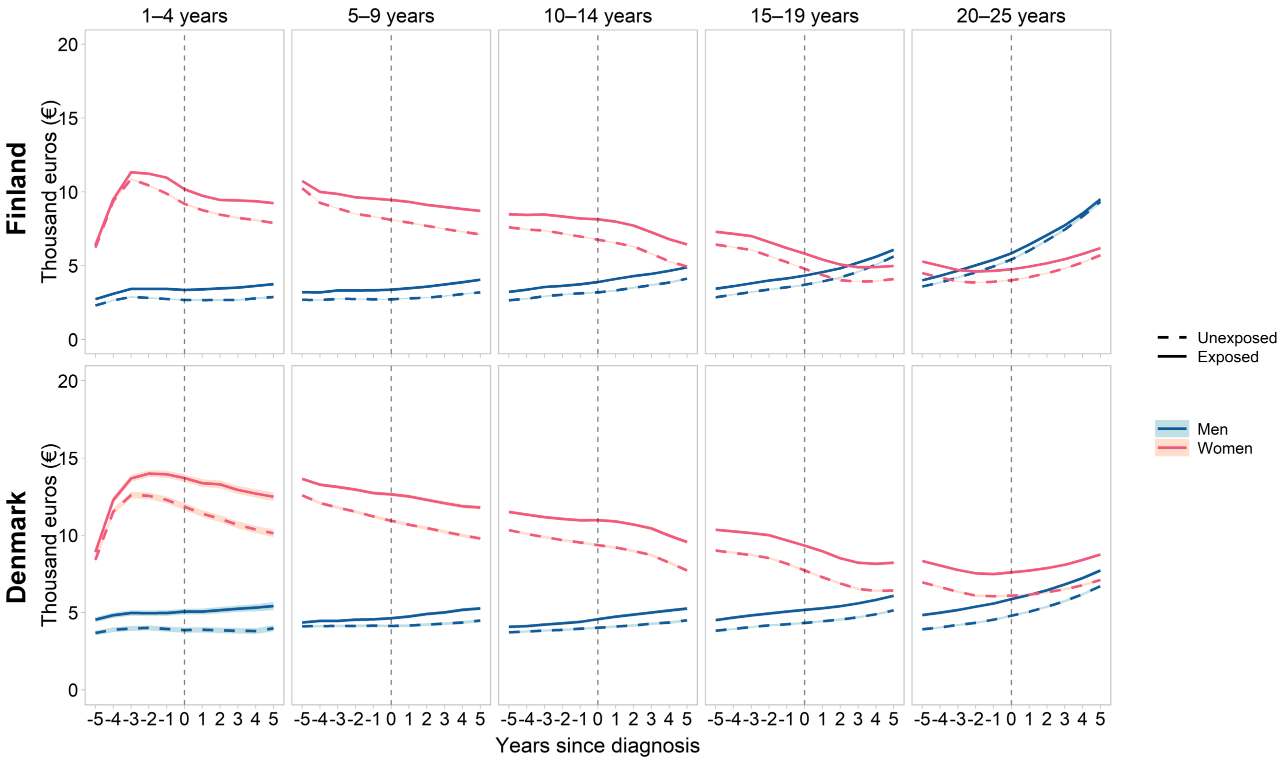

**eFigure 6. Marginal predictions of annual employment among parents exposed to child's any mental disorder (F00–F99) and unexposed parents stratified by the child's age at diagnosis**

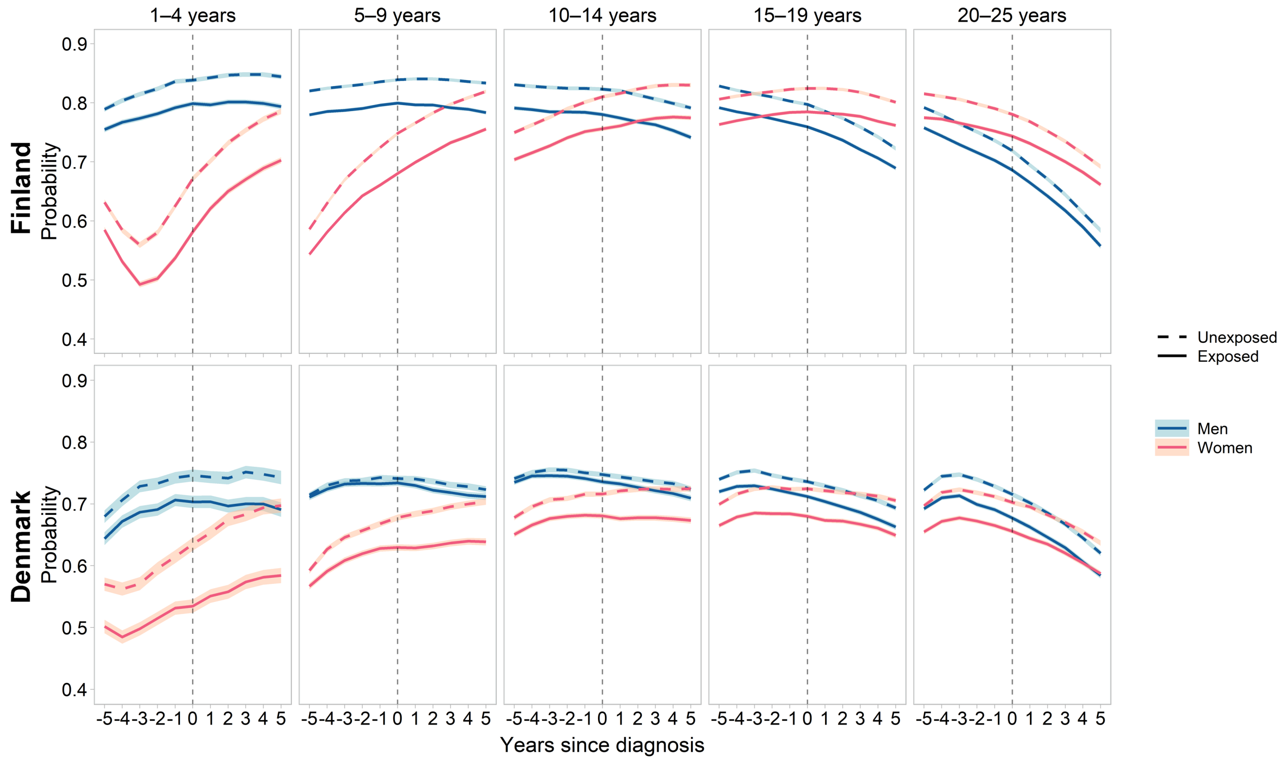

**eFigure 7. Marginal predictions of annual earnings among parents exposed to child's substance use disorder (F10–F19) and unexposed parents stratified by the child's age at diagnosis**

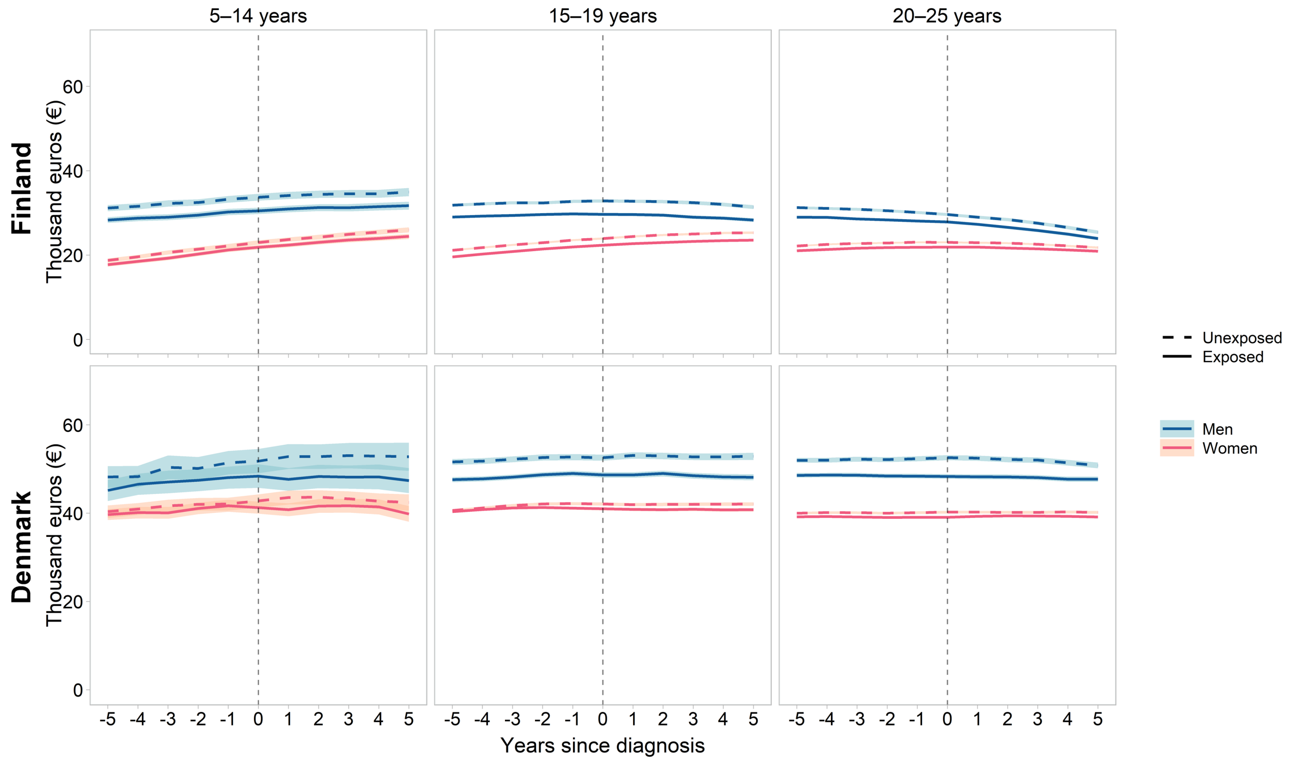

**eFigure 8. Marginal predictions of annual disposable income among parents exposed to child's substance use disorder (F10–F19) and unexposed parents stratified by the child's age at diagnosis**

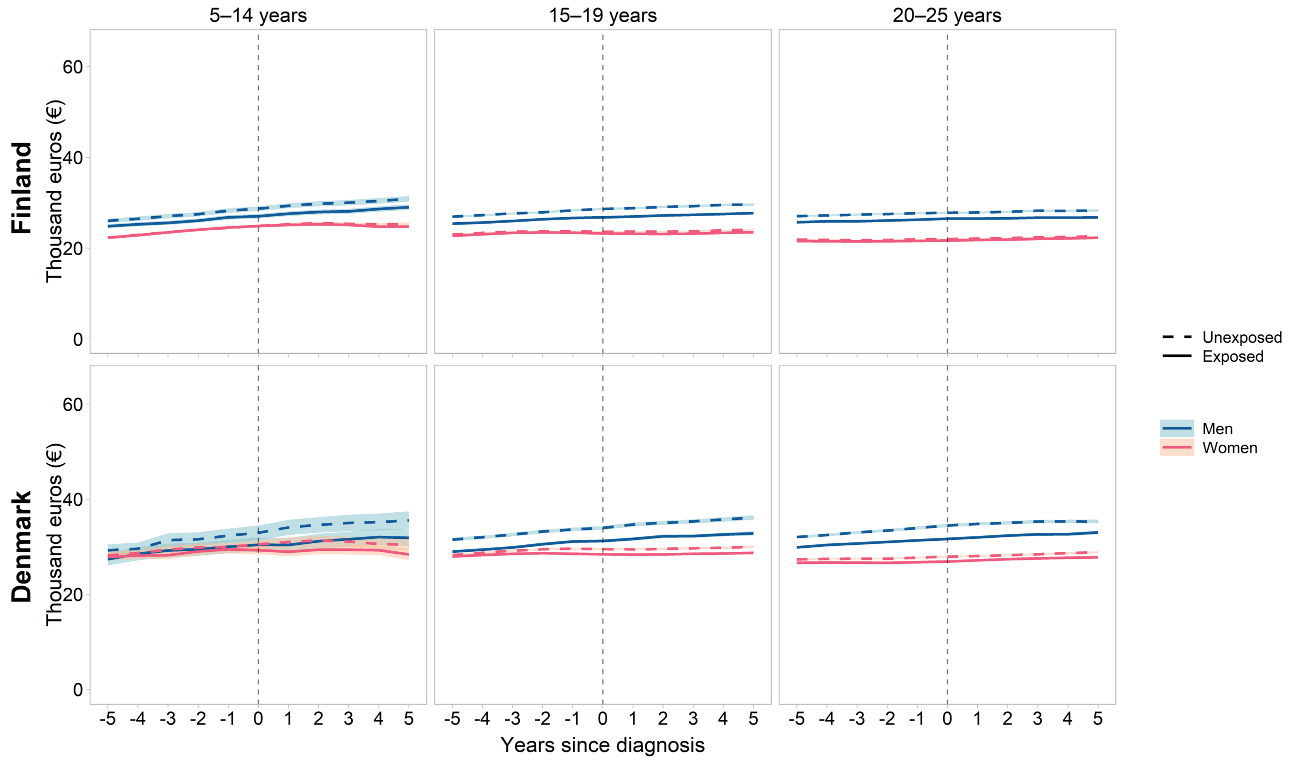

**eFigure 9. Marginal predictions of annual social welfare benefits among parents exposed to child's substance use disorder (F10–F19) and unexposed parents stratified by the child's age at diagnosis**

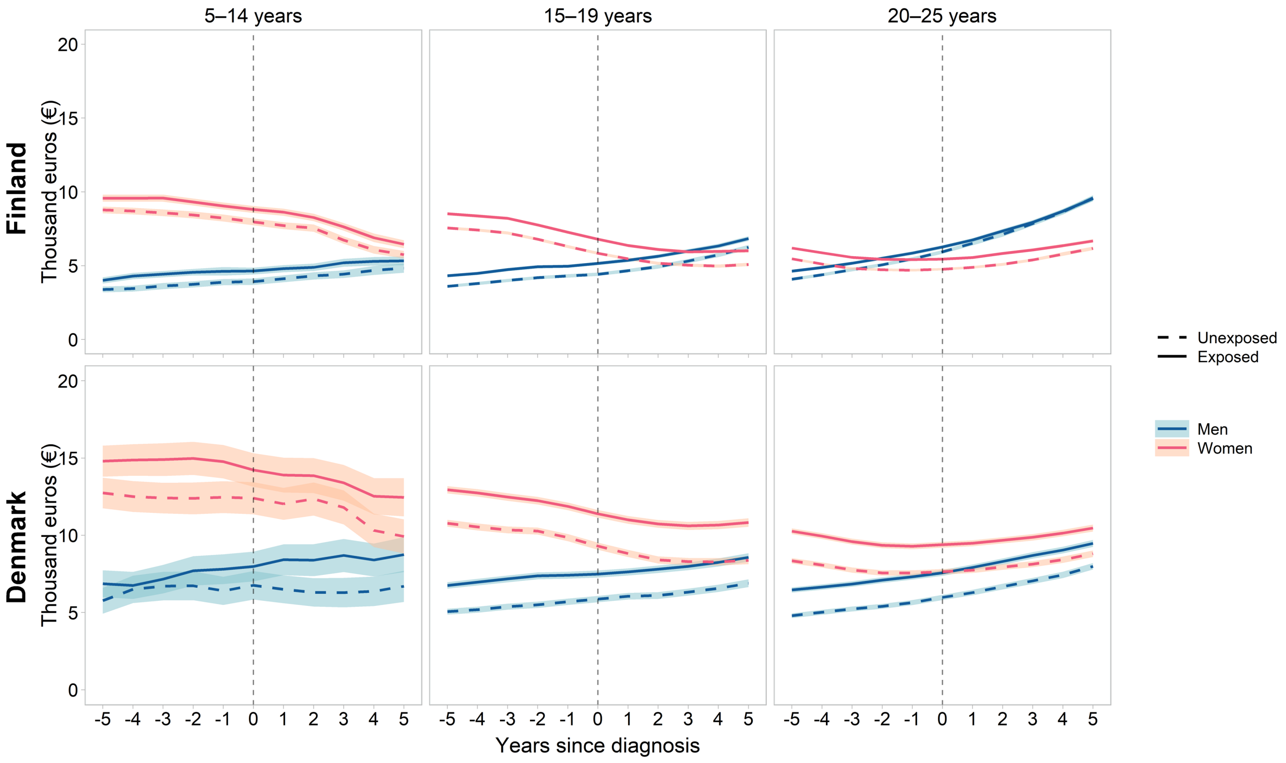

**eFigure 10. Marginal predictions of annual employment among parents exposed to child's substance use disorder (F10–F19) and unexposed parents stratified by the child's age at diagnosis**

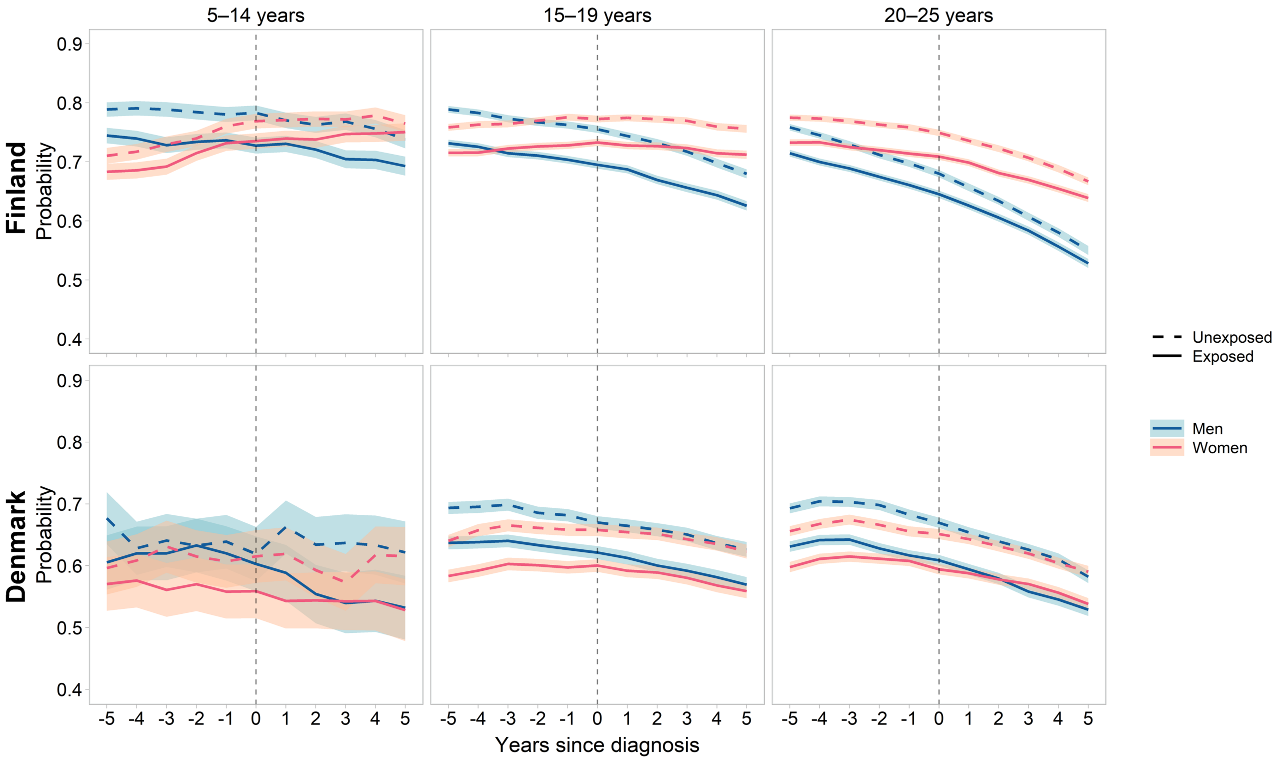

**eFigure 11. Marginal predictions of annual earnings among parents exposed to child's psychotic disorder (F20–F29) and unexposed parents stratified by the child's age at diagnosis**

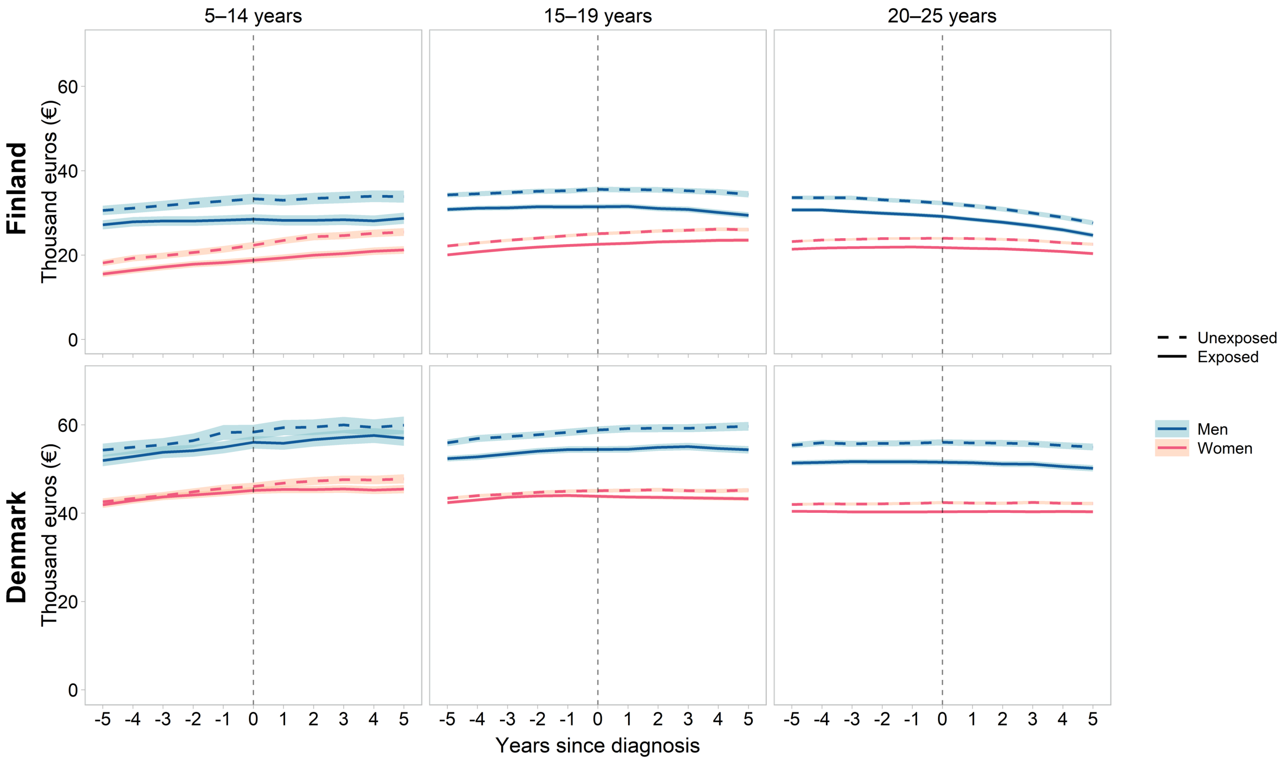

**eFigure 12. Marginal predictions of annual disposable income among parents exposed to child's psychotic disorder (F20–F29) and unexposed parents stratified by the child's age at diagnosis**

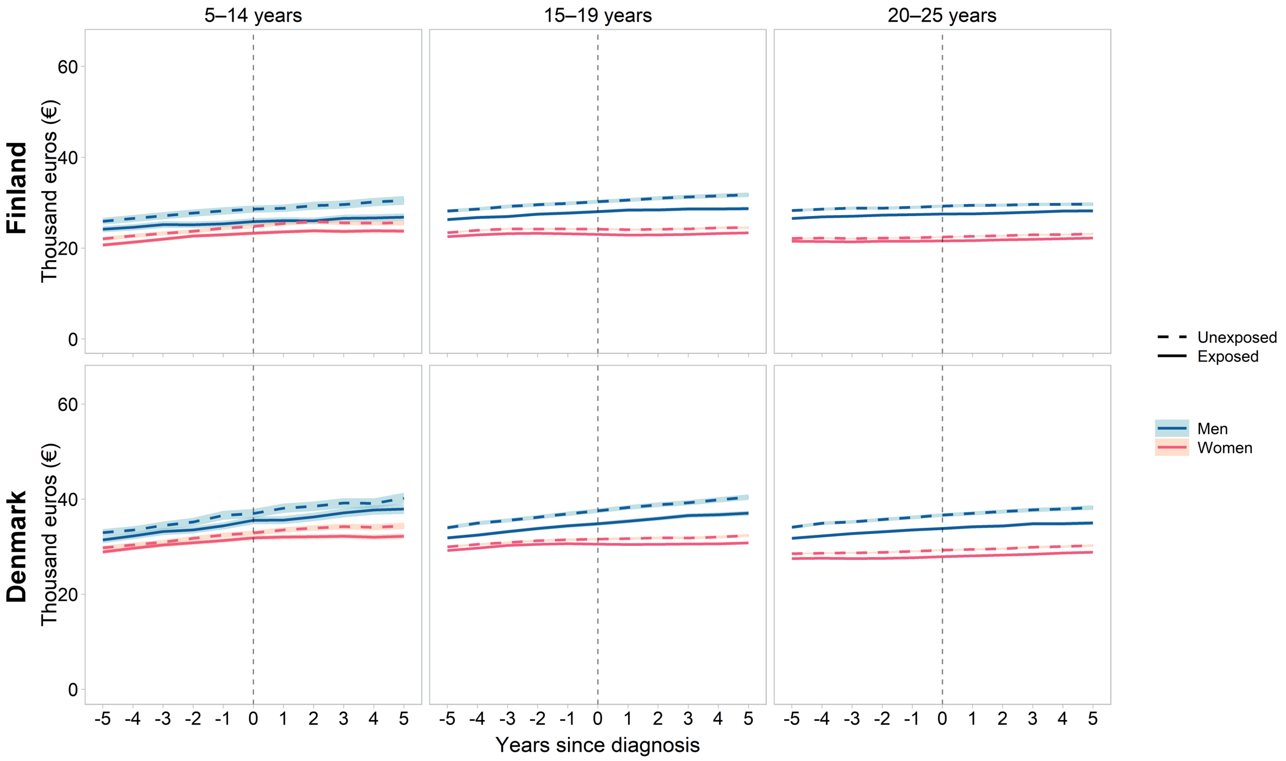

**eFigure 13. Marginal predictions of annual social welfare benefits among parents exposed to child's psychotic disorder (F20–F29) and unexposed parents stratified by the child's age at diagnosis**

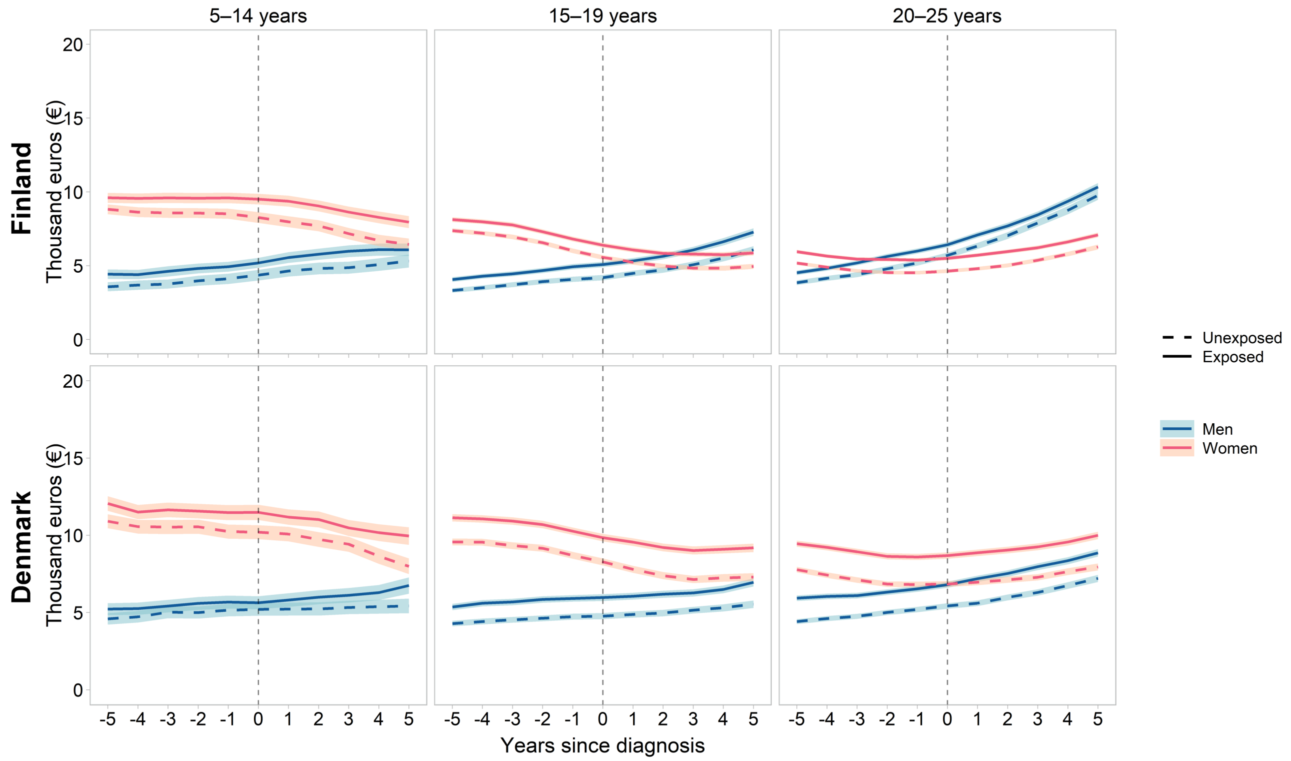

**eFigure 14. Marginal predictions of annual employment among parents exposed to child's psychotic disorder (F20–F29) and unexposed parents stratified by the child's age at diagnosis**

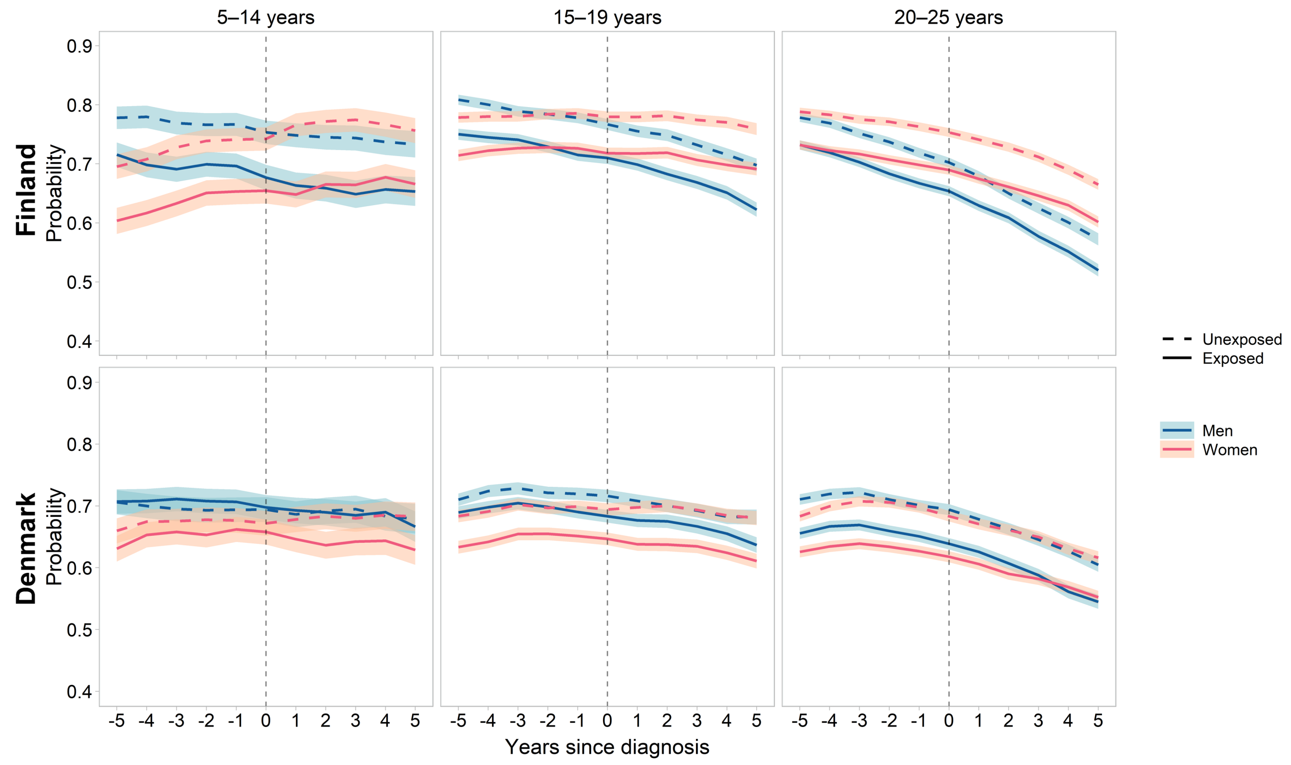

**eFigure 15. Marginal predictions of annual earnings among parents exposed to child's mood disorder (F30–F39) and unexposed parents stratified by the child's age at diagnosis**

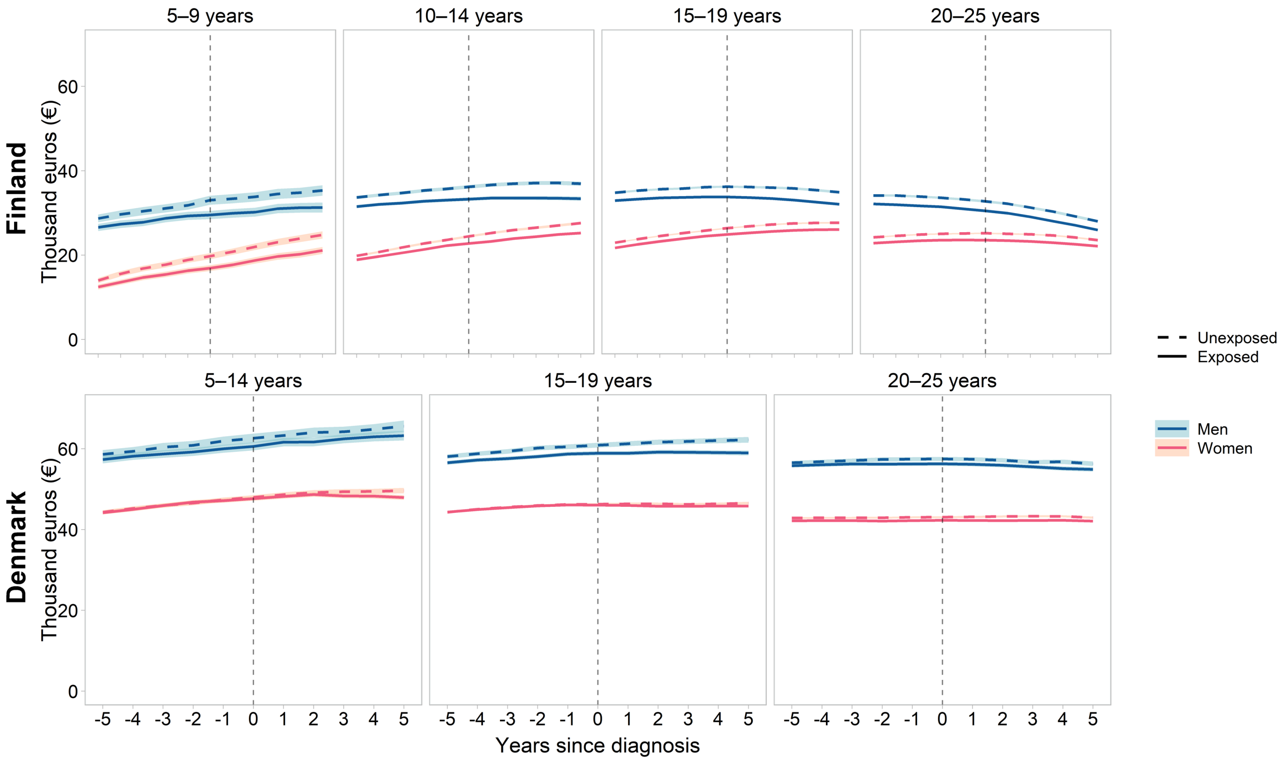

**eFigure 16. Marginal predictions of annual disposable income among parents exposed to child's mood disorder (F30–F39) and unexposed parents stratified by the child's age at diagnosis**

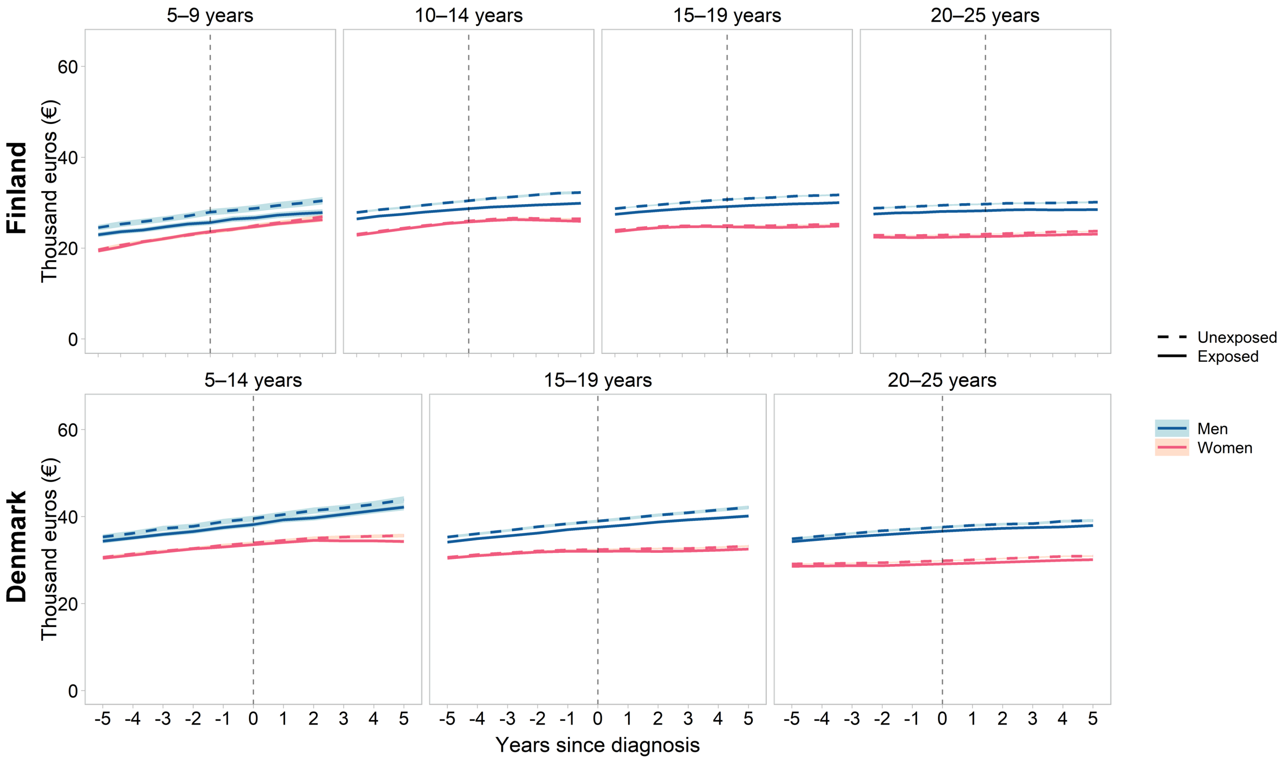

**eFigure 17. Marginal predictions of annual social welfare benefits among parents exposed to child's mood disorder (F30–F39) and unexposed parents stratified by the child's age at diagnosis**

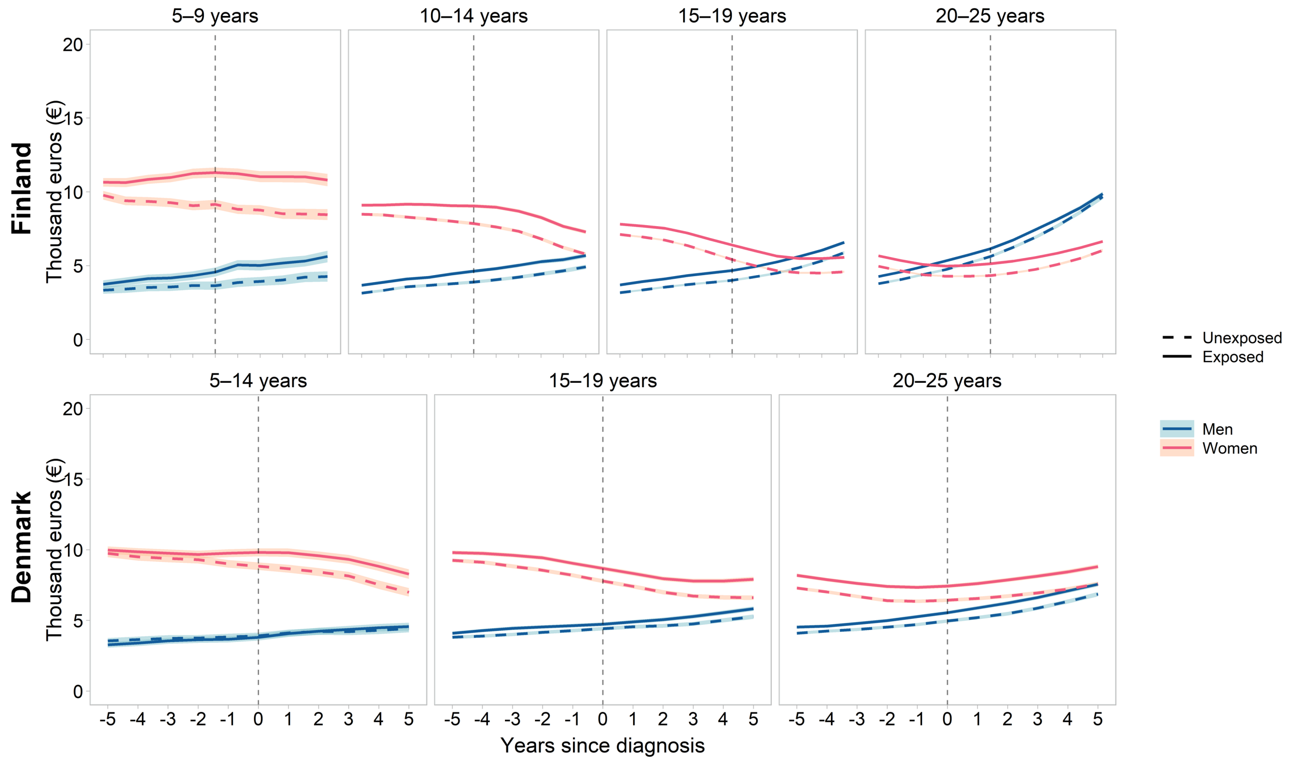

**eFigure 18. Marginal predictions of annual employment among parents exposed to child's mood disorder (F30–F39) and unexposed parents stratified by the child's age at diagnosis**

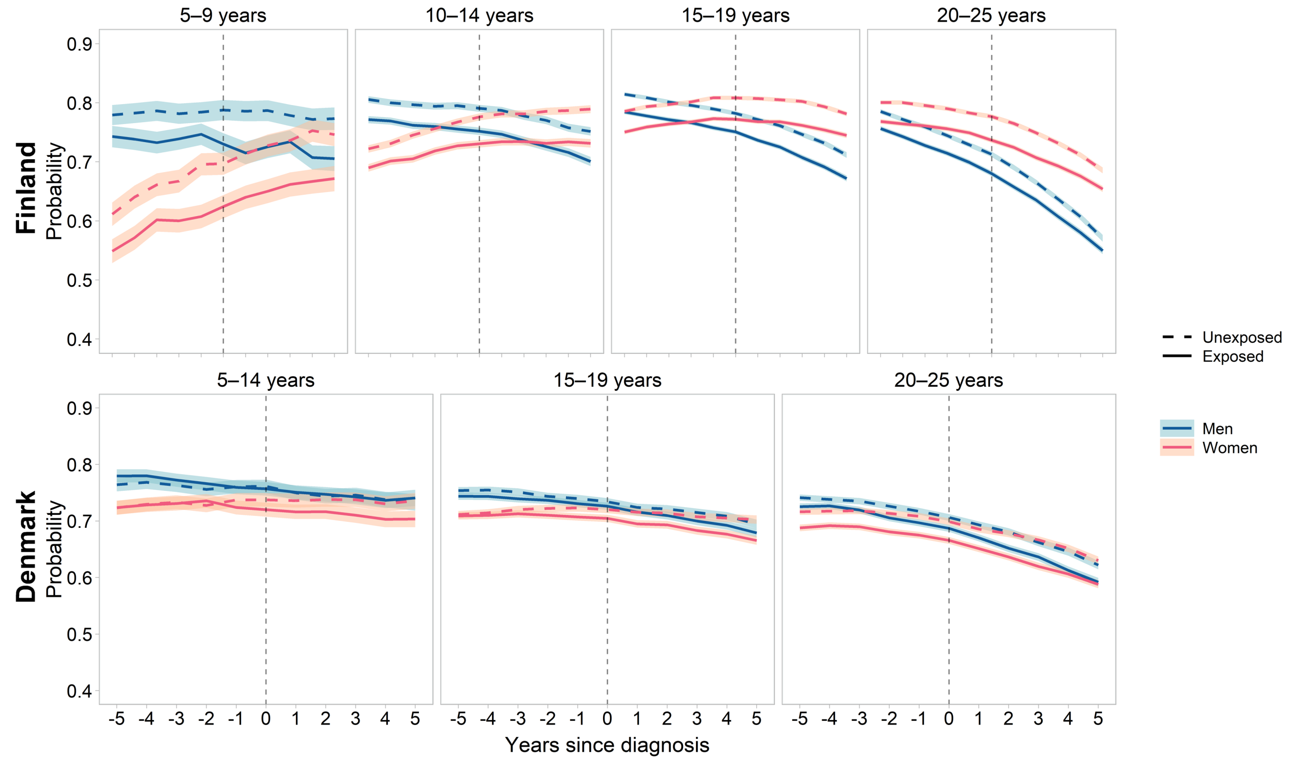

**eFigure 19. Marginal predictions of annual earnings among parents exposed to child's anxiety disorder (F40–F48) and unexposed parents stratified by the child's age at diagnosis**

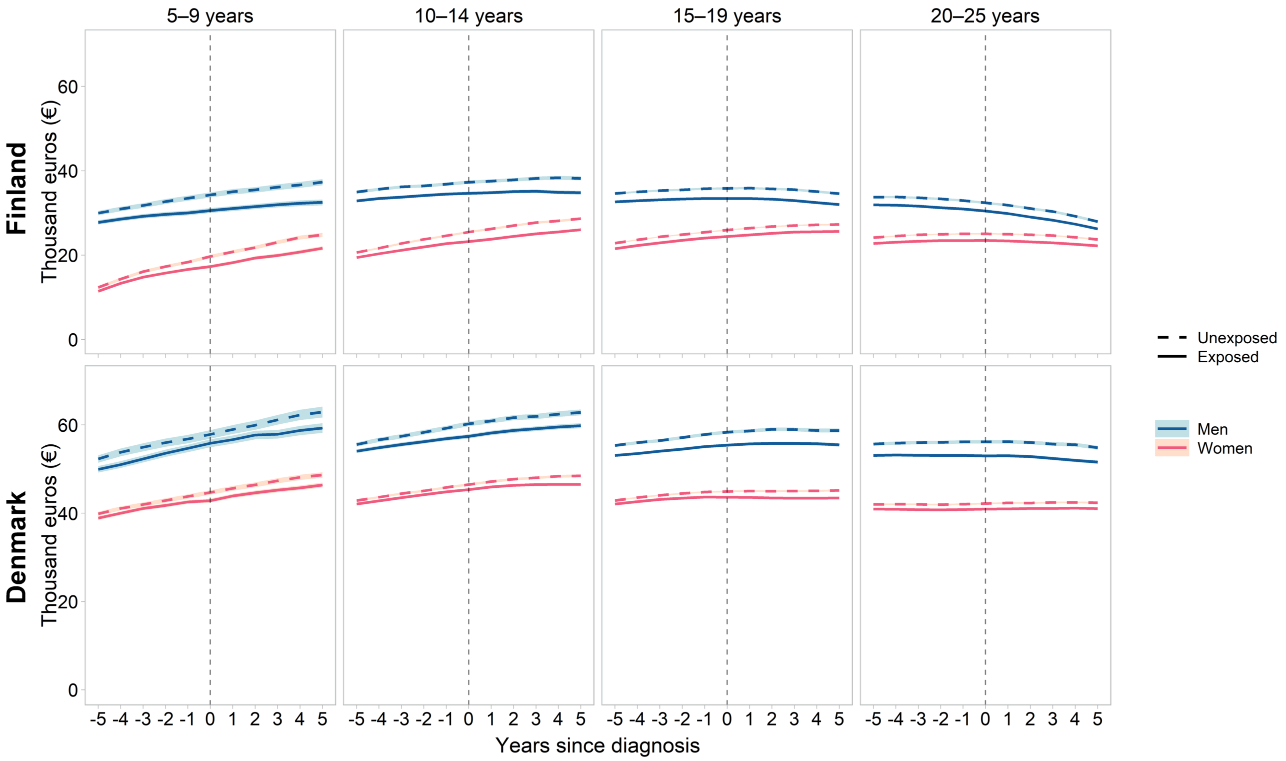

**eFigure 20. Marginal predictions of annual disposable income among parents exposed to child's anxiety disorder (F40–F48) and unexposed parents stratified by the child's age at diagnosis**

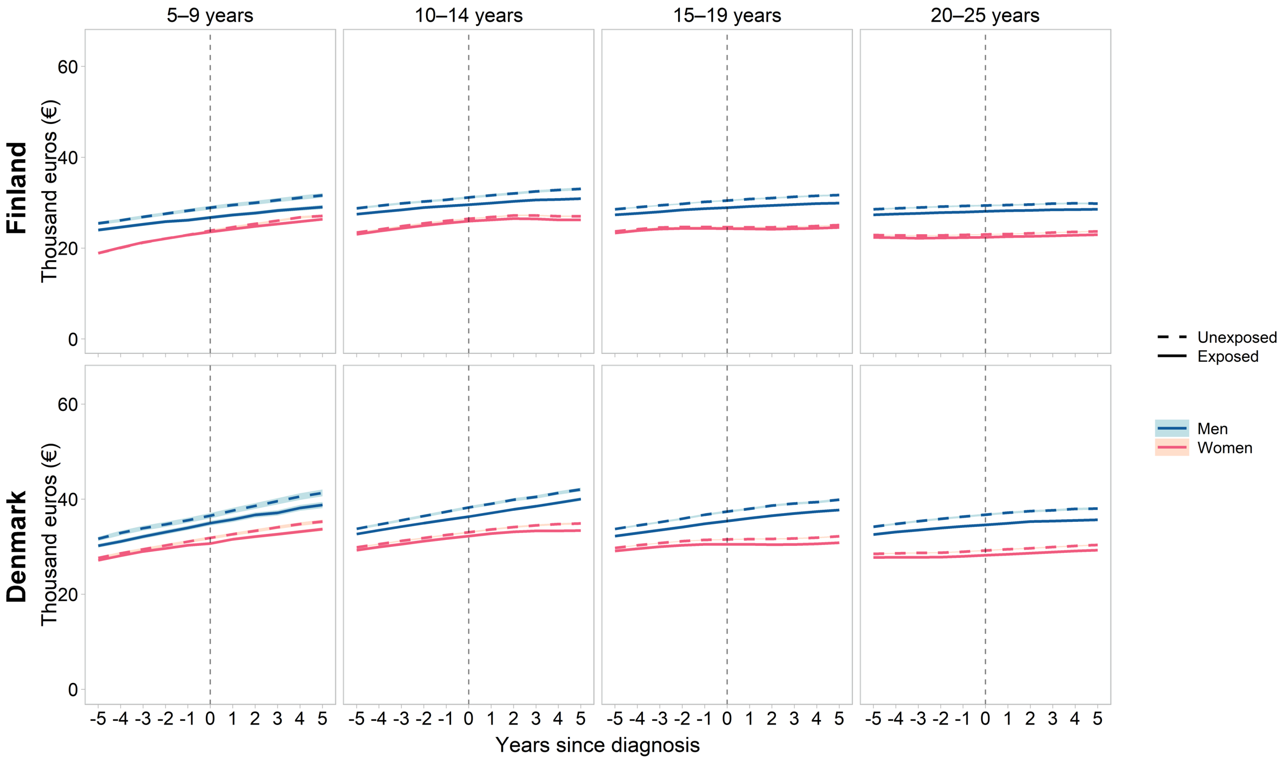

**eFigure 21. Marginal predictions of annual social welfare benefits among parents exposed to child's anxiety disorder (F40–F48) and unexposed parents stratified by the child's age at diagnosis**

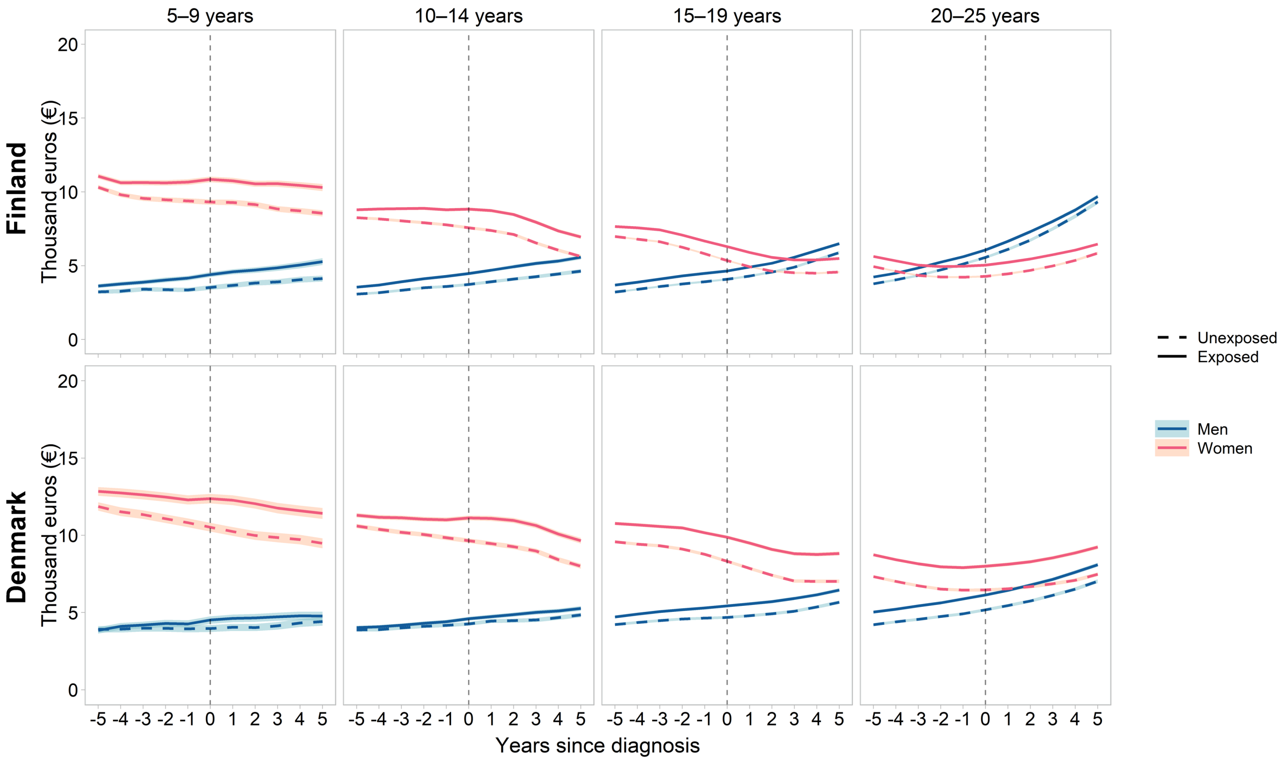

**eFigure 22. Marginal predictions of annual employment among parents exposed to child's anxiety disorder (F40–F48) and unexposed parents stratified by the child's age at diagnosis**

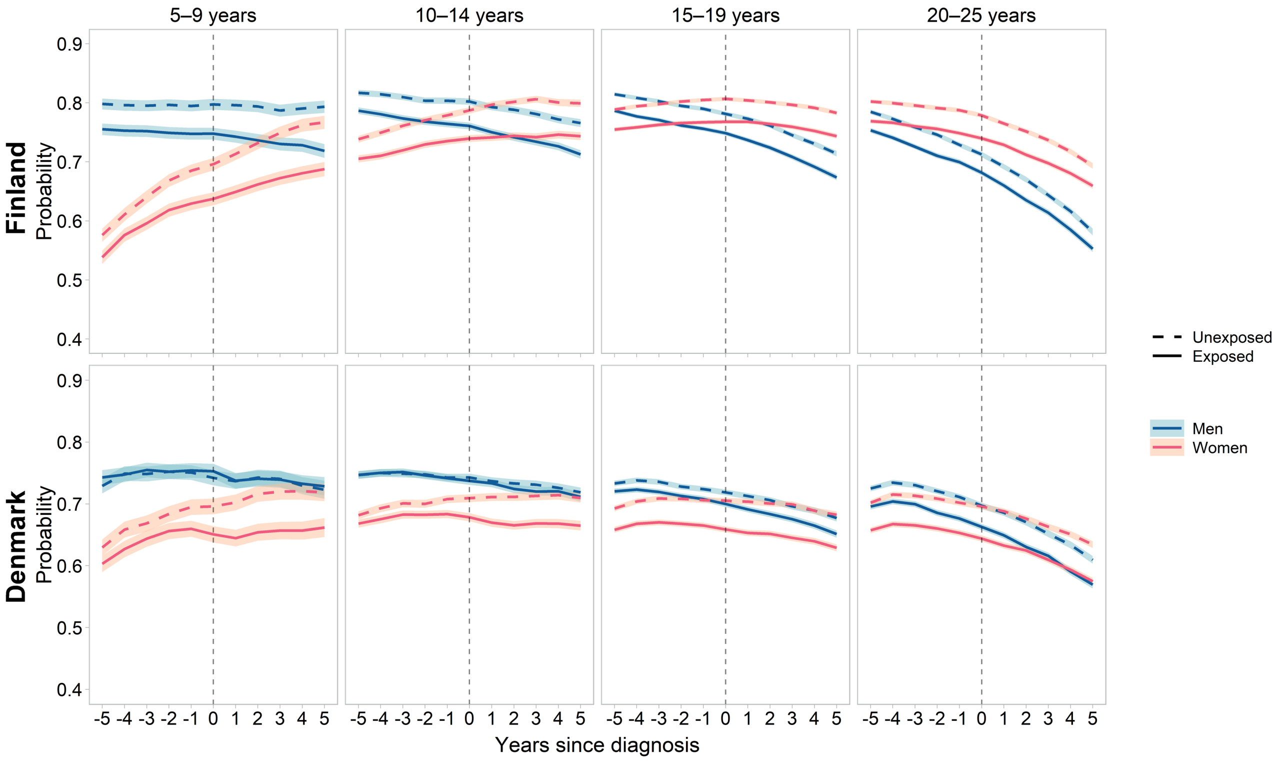

**eFigure 23. Marginal predictions of annual earnings among parents exposed to child's eating disorder (F50) and unexposed parents stratified by the child's age at diagnosis**

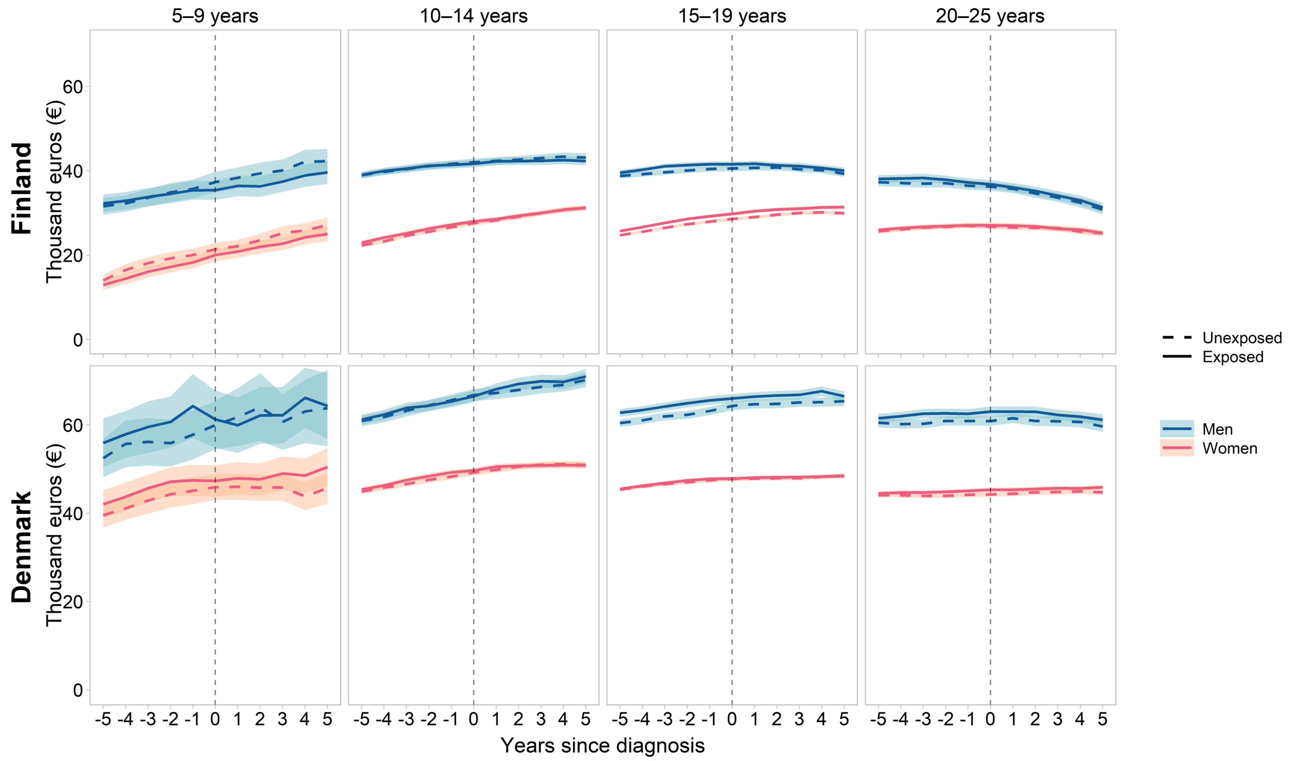

**eFigure 24. Marginal predictions of annual disposable income among parents exposed to child's eating disorder (F50) and unexposed parents stratified by the child's age at diagnosis**

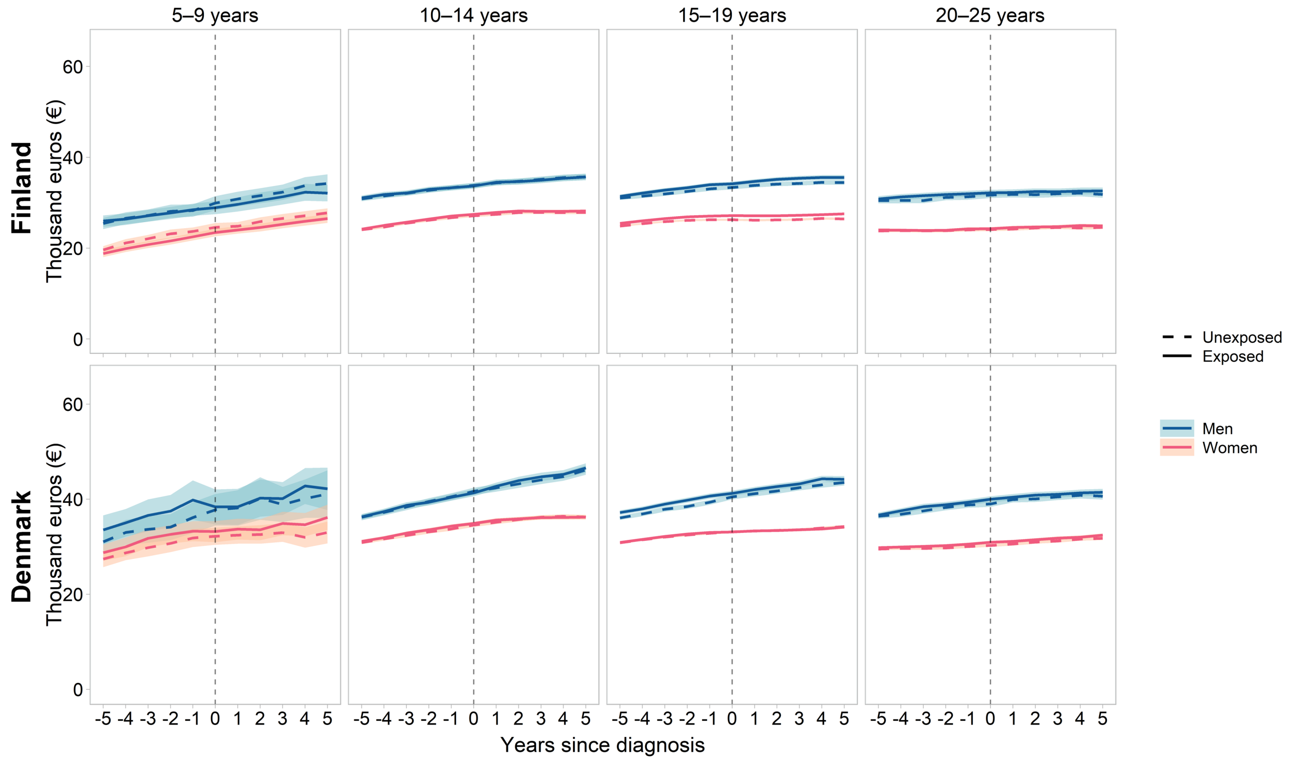

**eFigure 25. Marginal predictions of annual social welfare benefits among parents exposed to child's eating disorder (F50) and unexposed parents stratified by the child's age at diagnosis**

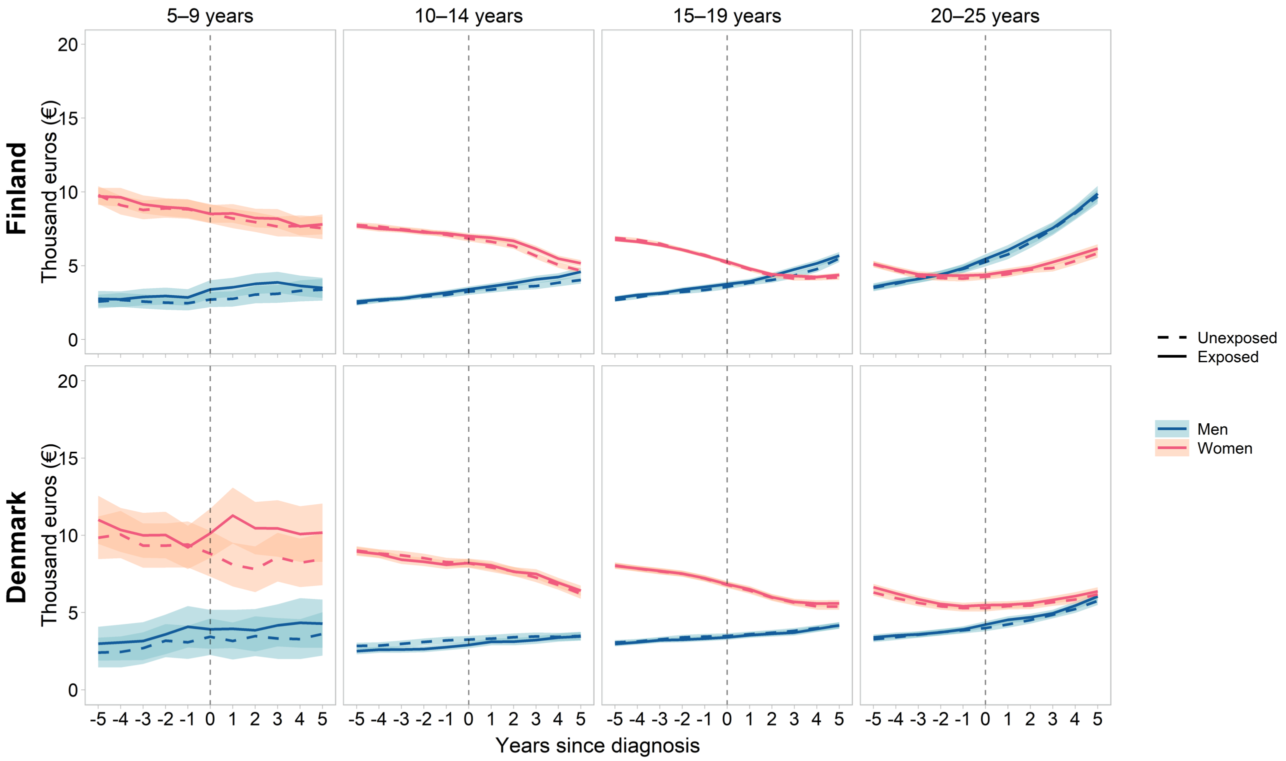

**eFigure 26. Marginal predictions of annual employment among parents exposed to child's eating disorder (F50) and unexposed parents stratified by the child's age at diagnosis**

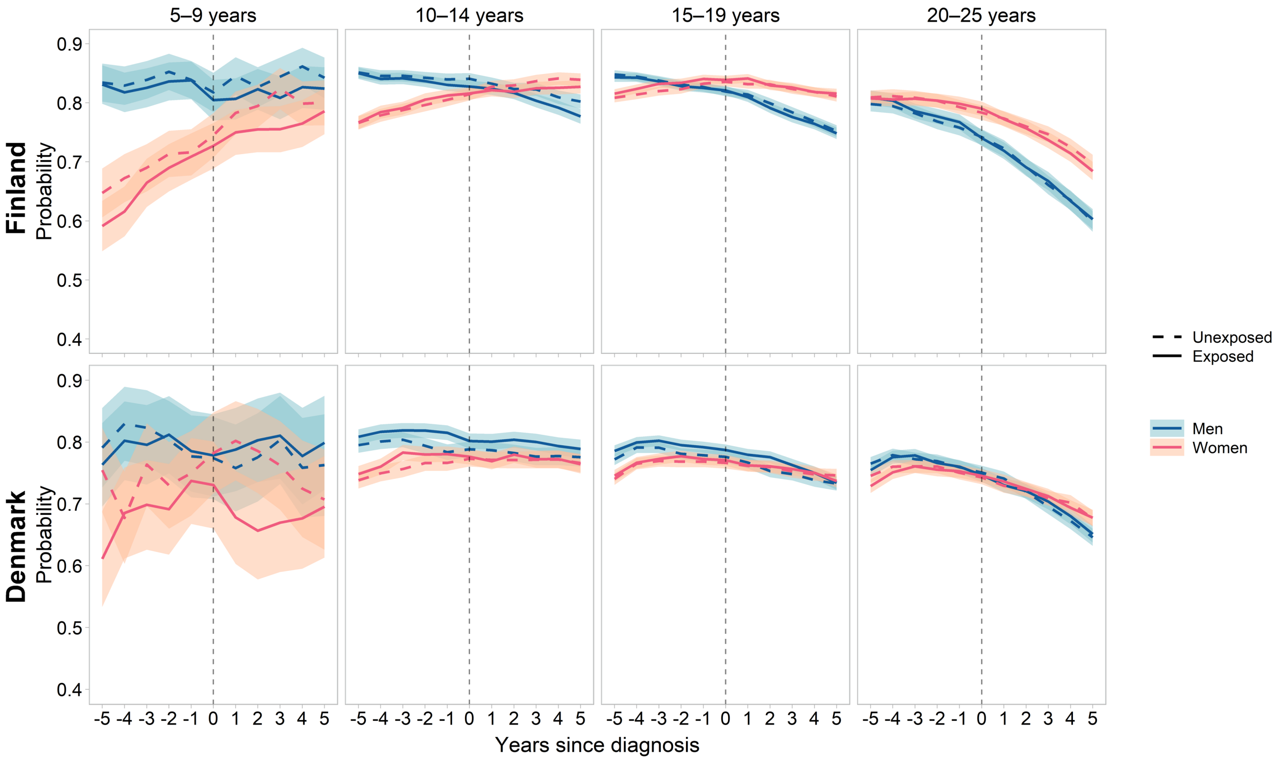

**eFigure 27. Marginal predictions of annual earnings among parents exposed to child's personality disorder (F60) and unexposed parents stratified by the child's age at diagnosis**

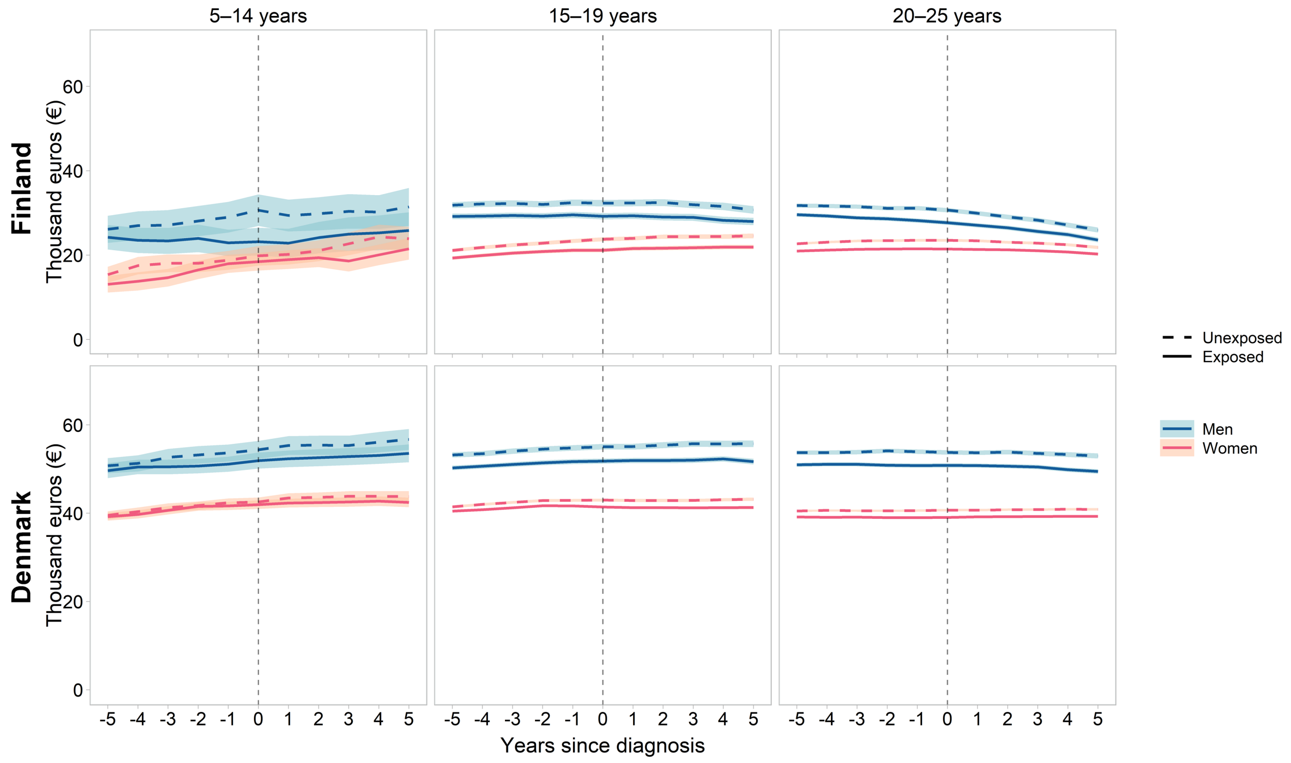

**eFigure 28. Marginal predictions of annual disposable income among parents exposed to child's personality disorder (F60) and unexposed parents stratified by the child's age at diagnosis**

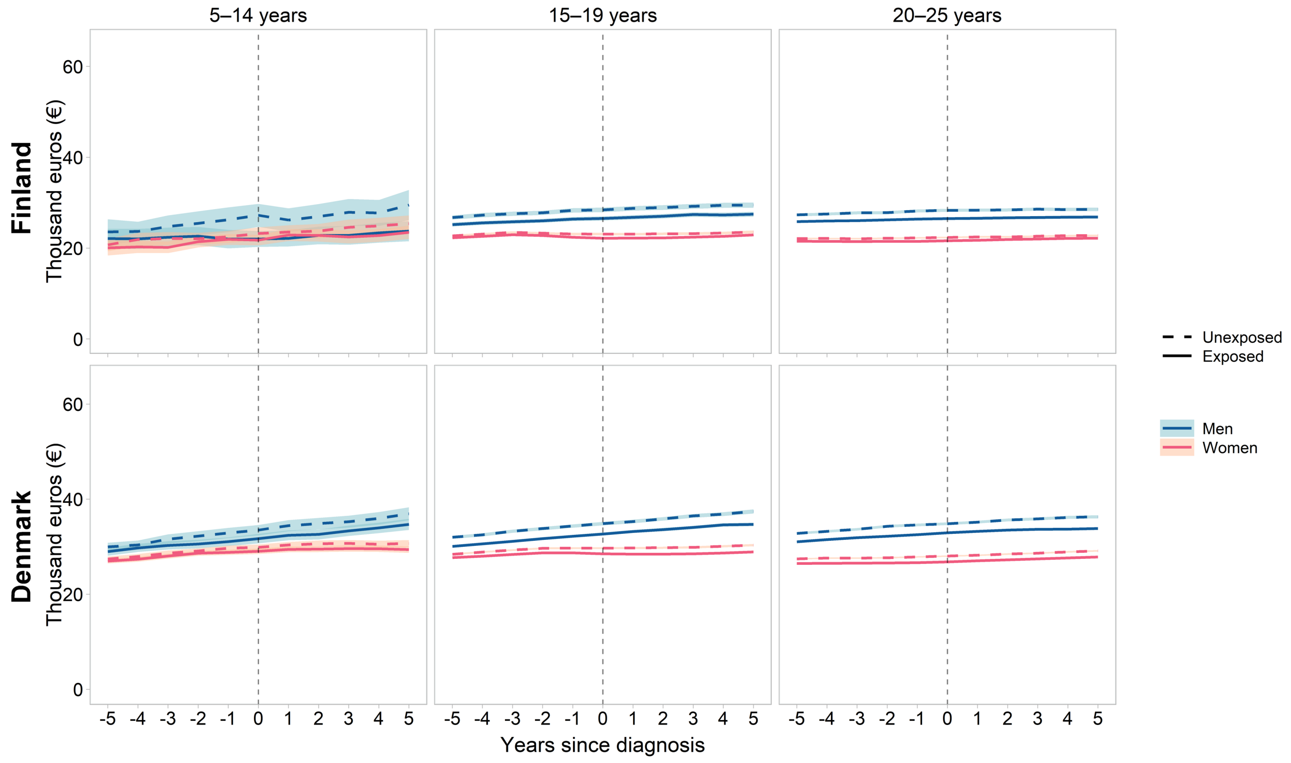

**eFigure 29. Marginal predictions of annual social welfare benefits among parents exposed to child's personality disorder (F60) and unexposed parents stratified by the child's age at diagnosis**

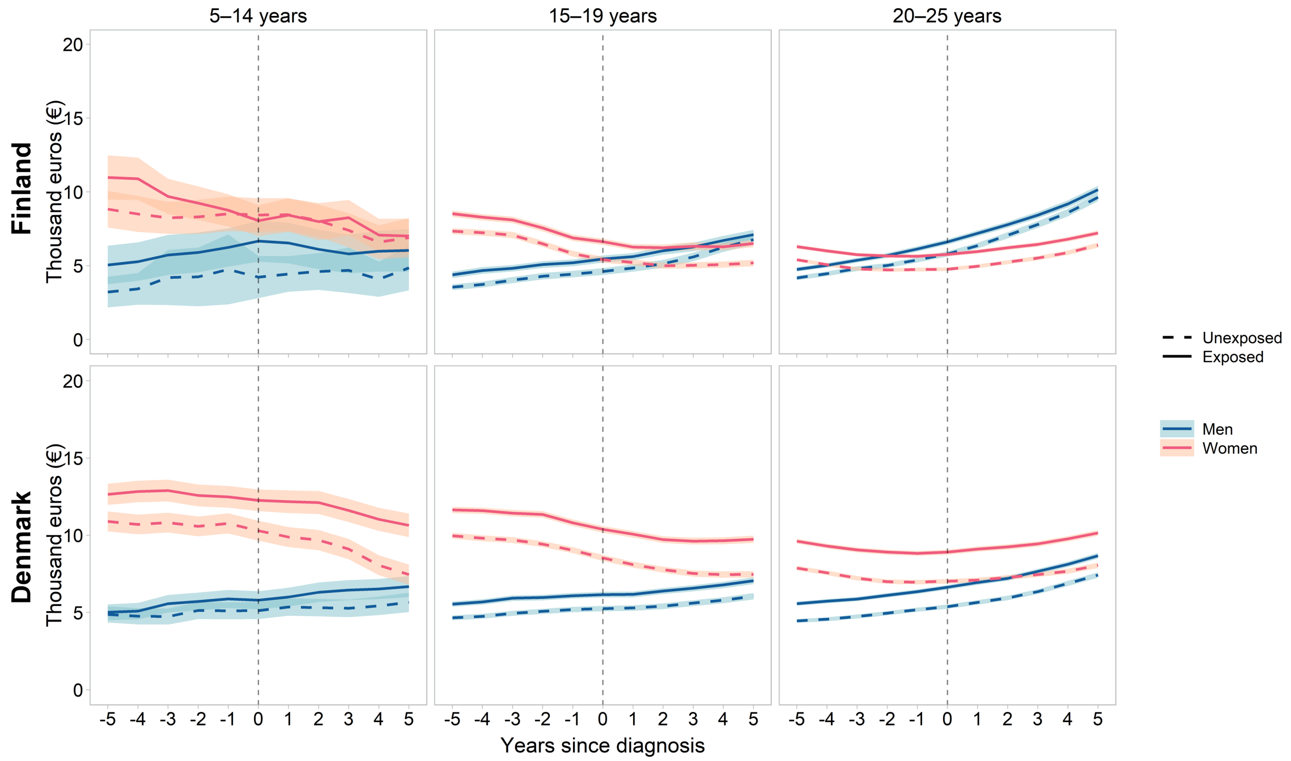

**eFigure 30. Marginal predictions of annual employment among parents exposed to child's personality disorder (F60) and unexposed parents stratified by the child's age at diagnosis**

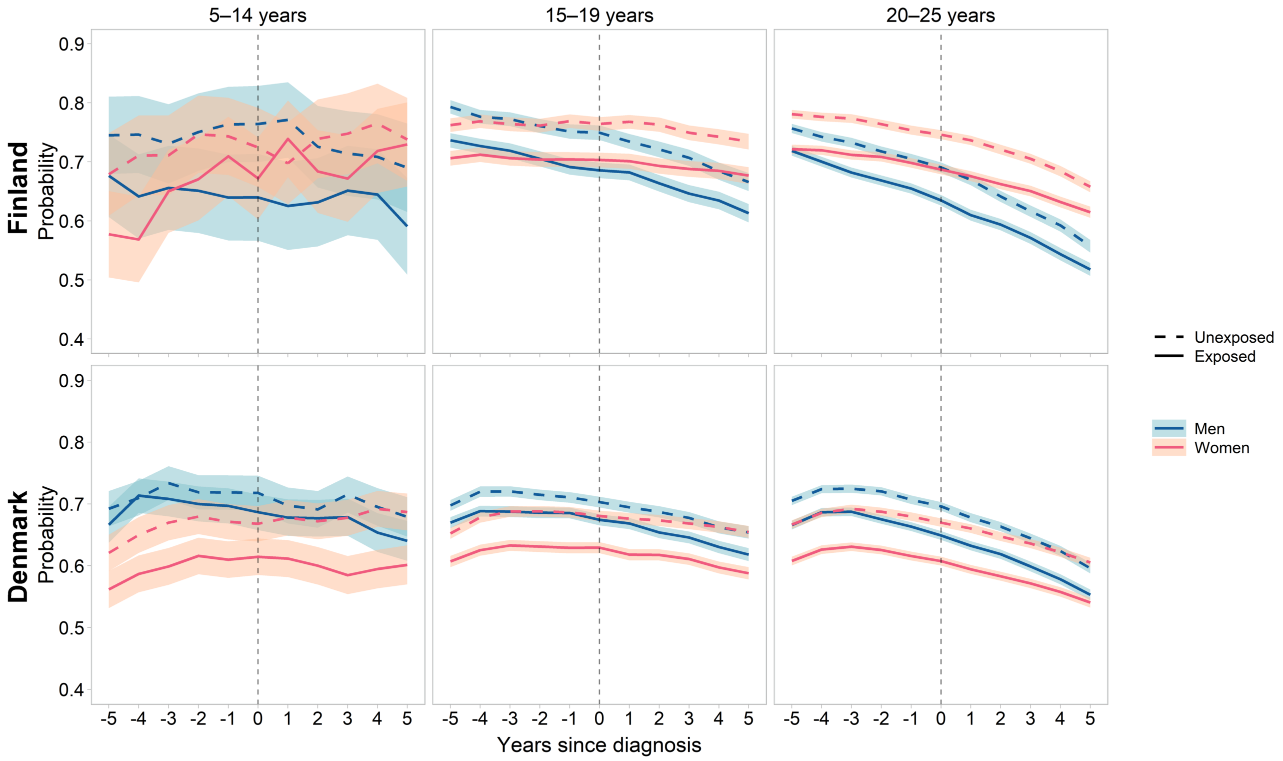

**eFigure 31. Marginal predictions of annual earnings among parents exposed to child's intellectual disability (F70–F79) and unexposed parents stratified by the child's age at diagnosis**

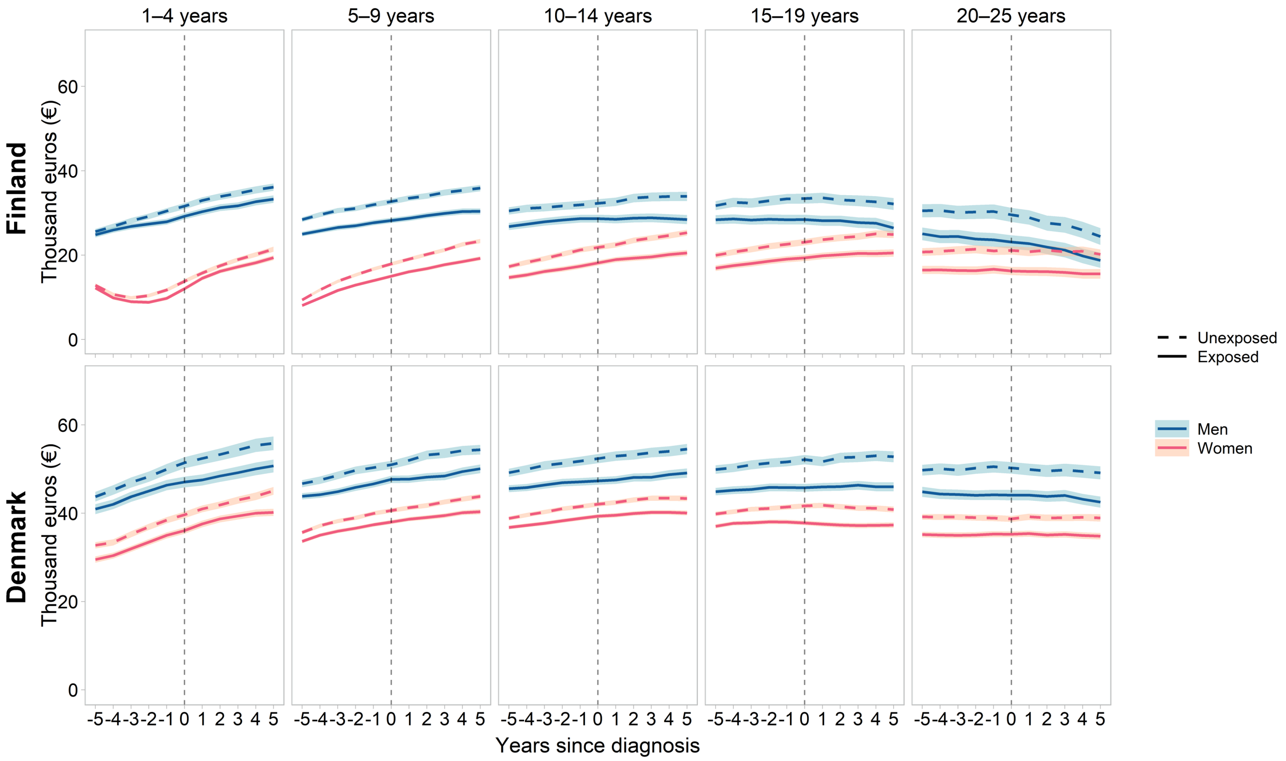

**eFigure 32. Marginal predictions of annual disposable income among parents exposed to child's intellectual disability (F70–F79) and unexposed parents stratified by the child's age at diagnosis**

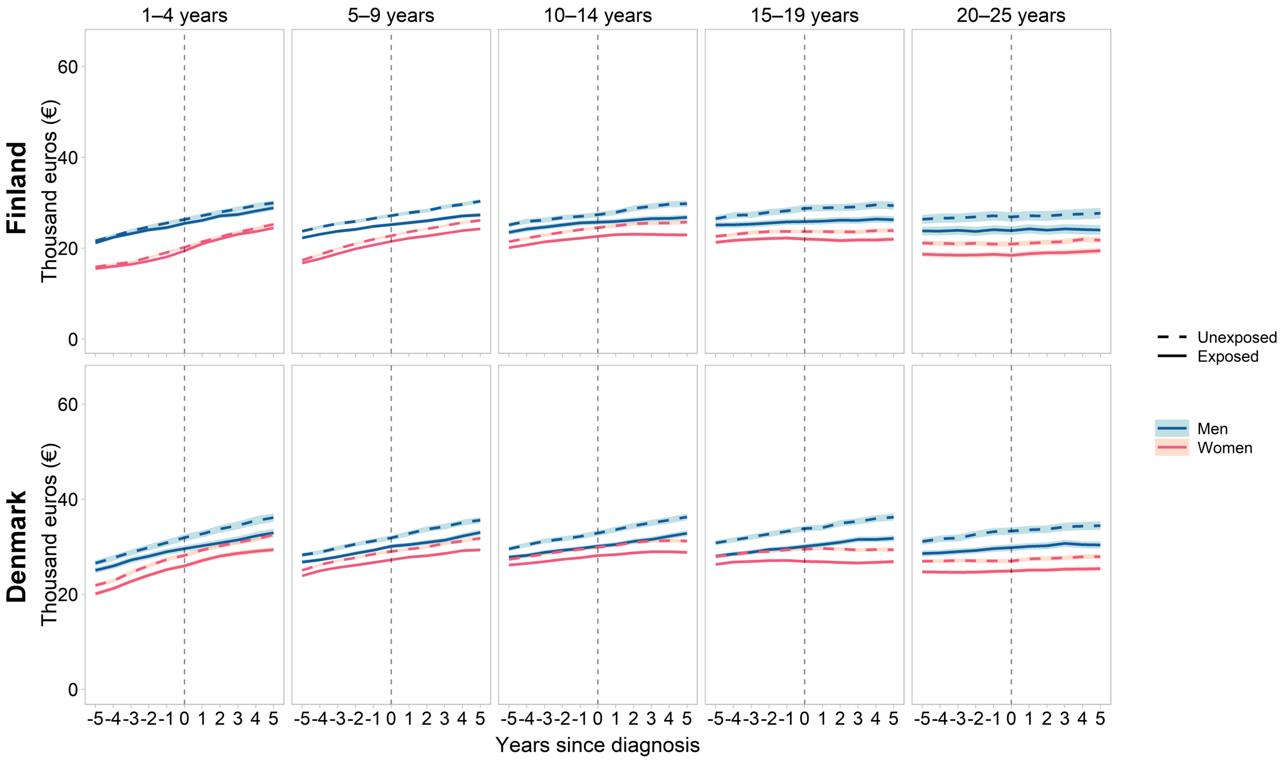

**eFigure 33. Marginal predictions of annual social welfare benefits among parents exposed to child's intellectual disability (F70–F79) and unexposed parents stratified by the child's age at diagnosis**

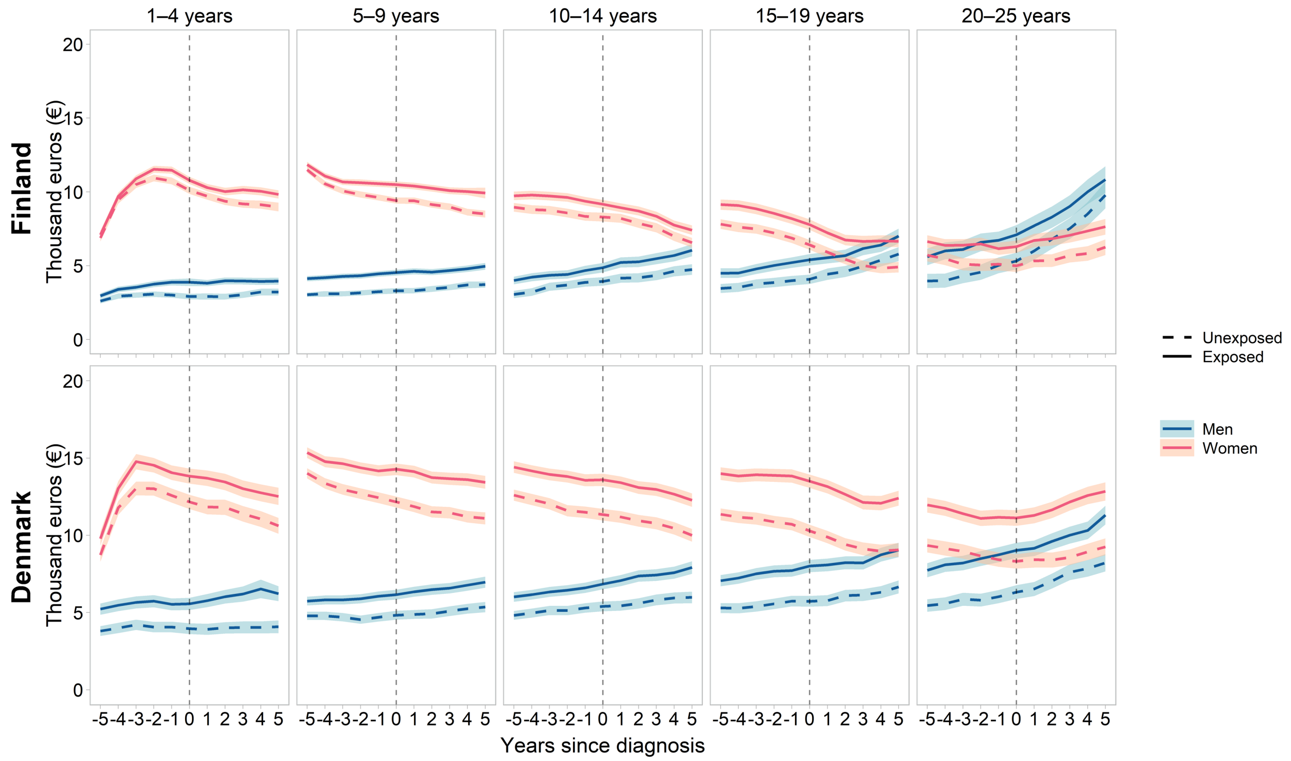

**eFigure 34. Marginal predictions of annual employment among parents exposed to child's intellectual disability (F70–F79) and unexposed parents stratified by the child's age at diagnosis**

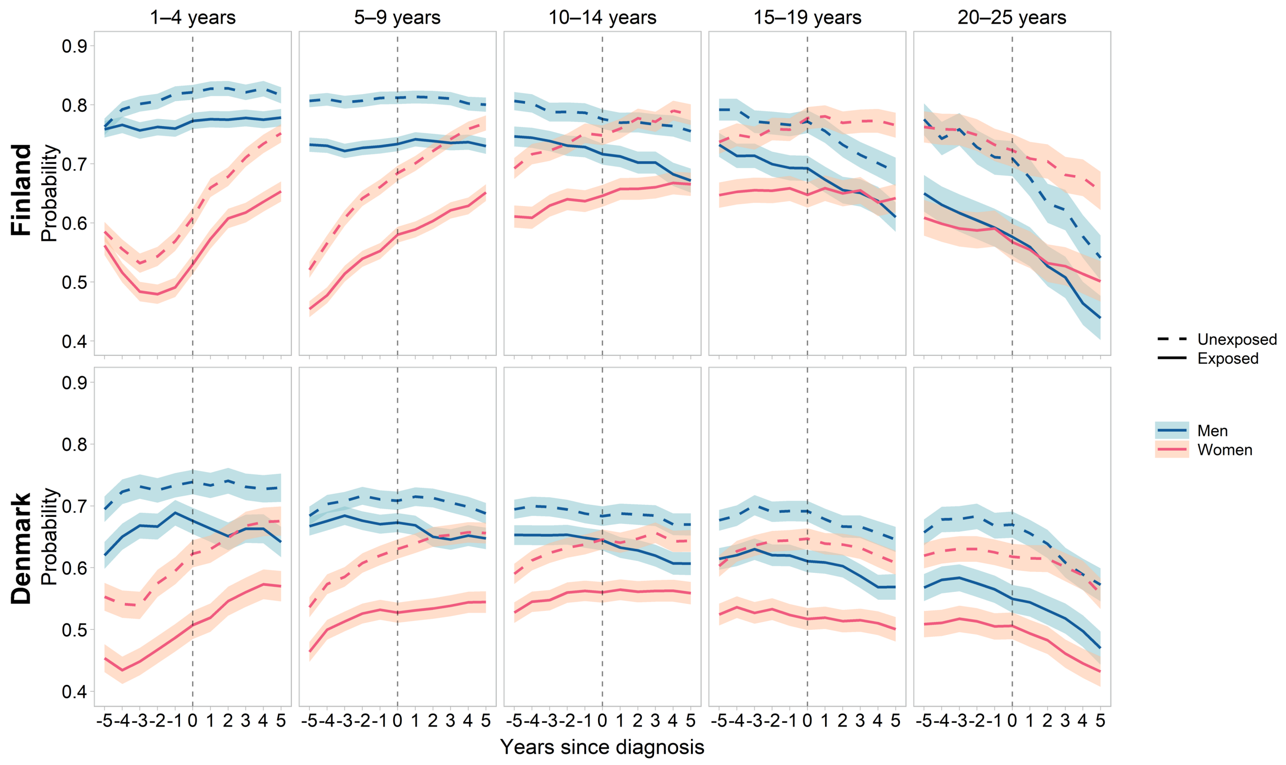

**eFigure 35. Marginal predictions of annual earnings among parents exposed to child's developmental disorder (F84) and unexposed parents stratified by the child's age at diagnosis**

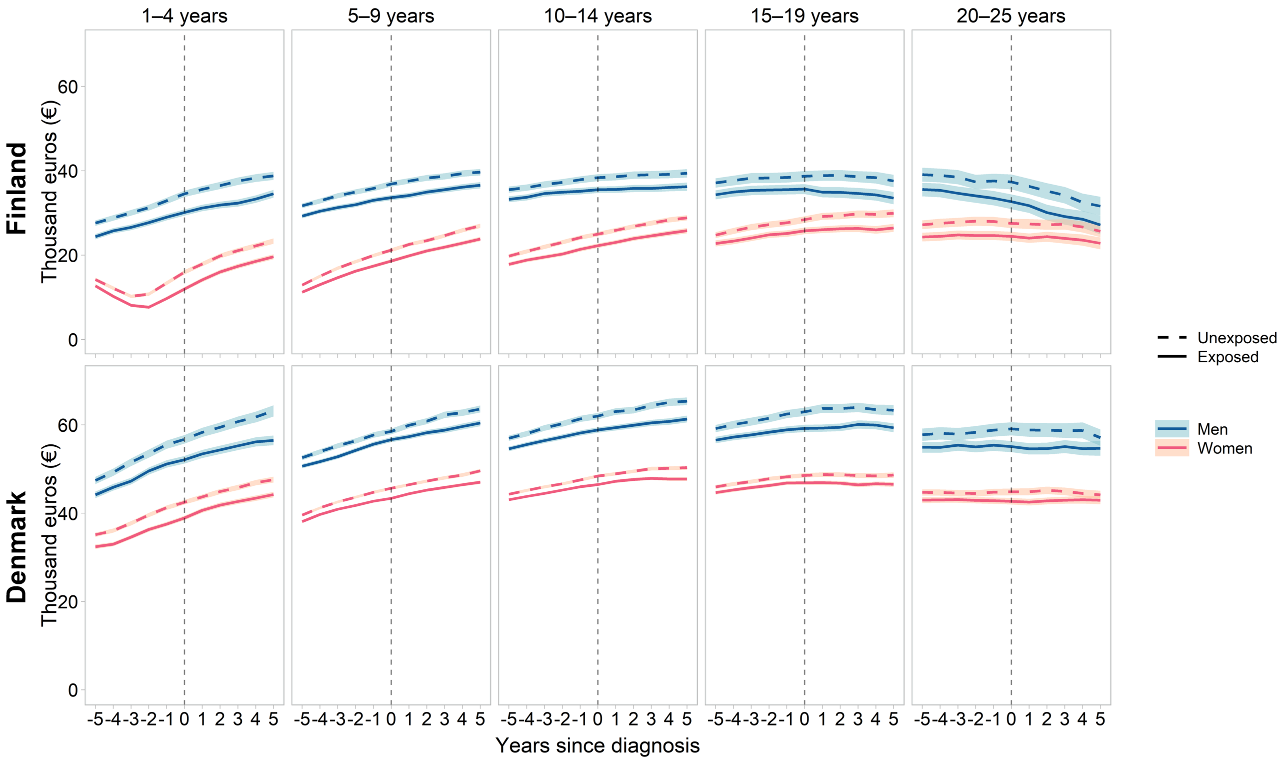

**eFigure 36. Marginal predictions of annual disposable income among parents exposed to child's developmental disorder (F84) and unexposed parents stratified by the child's age at diagnosis**

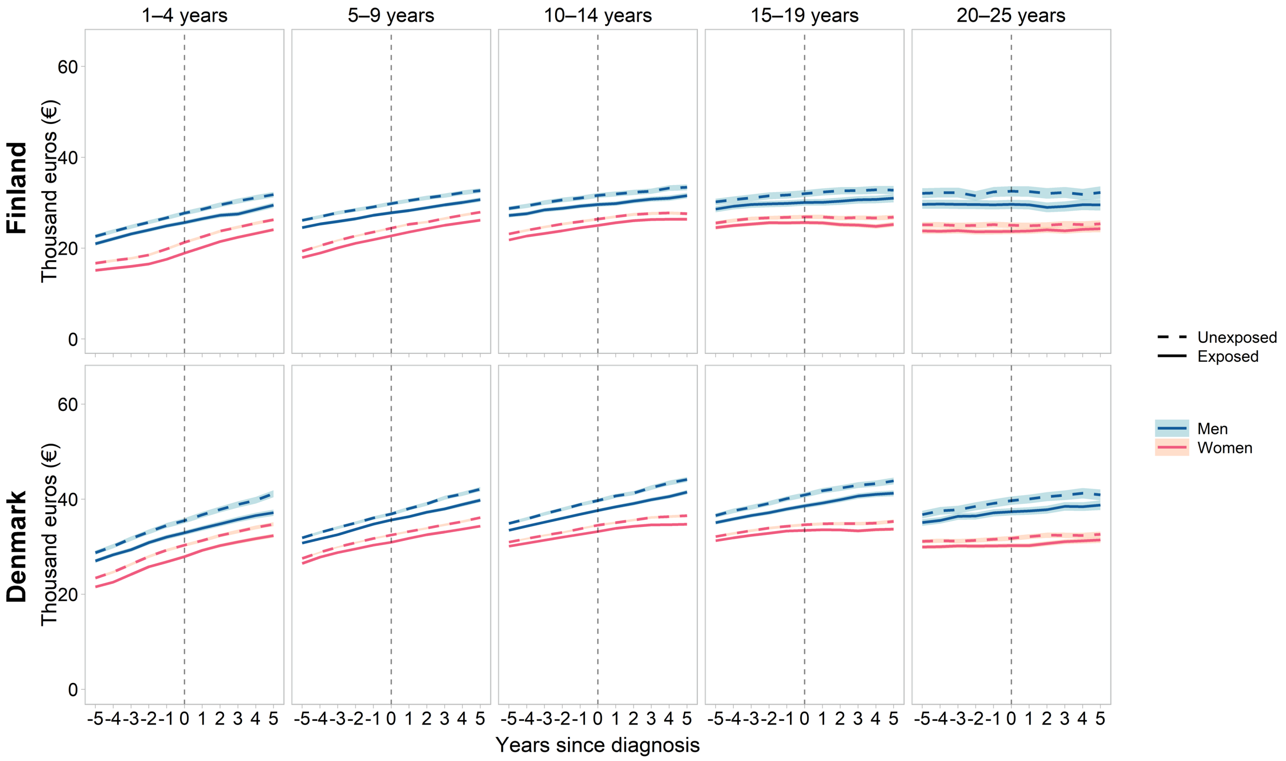

**eFigure 37. Marginal predictions of annual social welfare benefits among parents exposed to child's developmental disorder (F84) and unexposed parents stratified by the child's age at diagnosis**

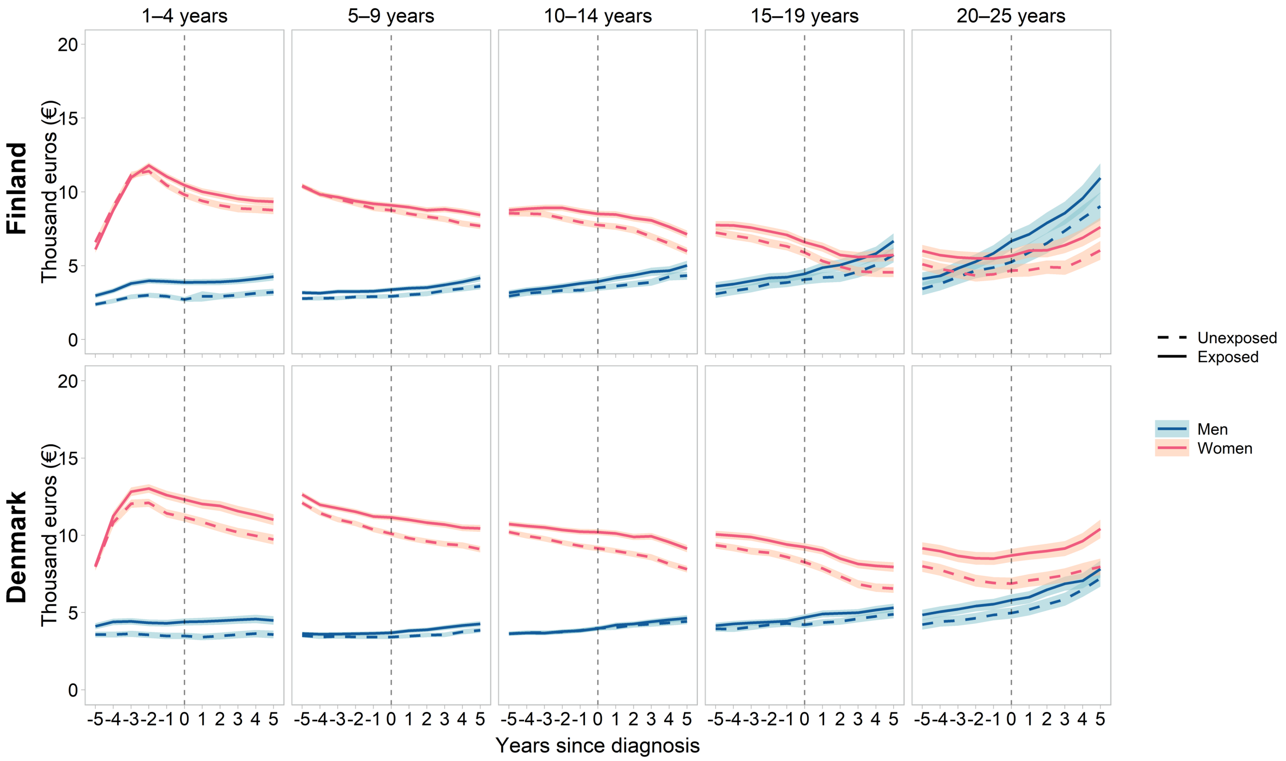

**eFigure 38. Marginal predictions of annual employment among parents exposed to child's developmental disorder (F84) and unexposed parents stratified by the child's age at diagnosis**

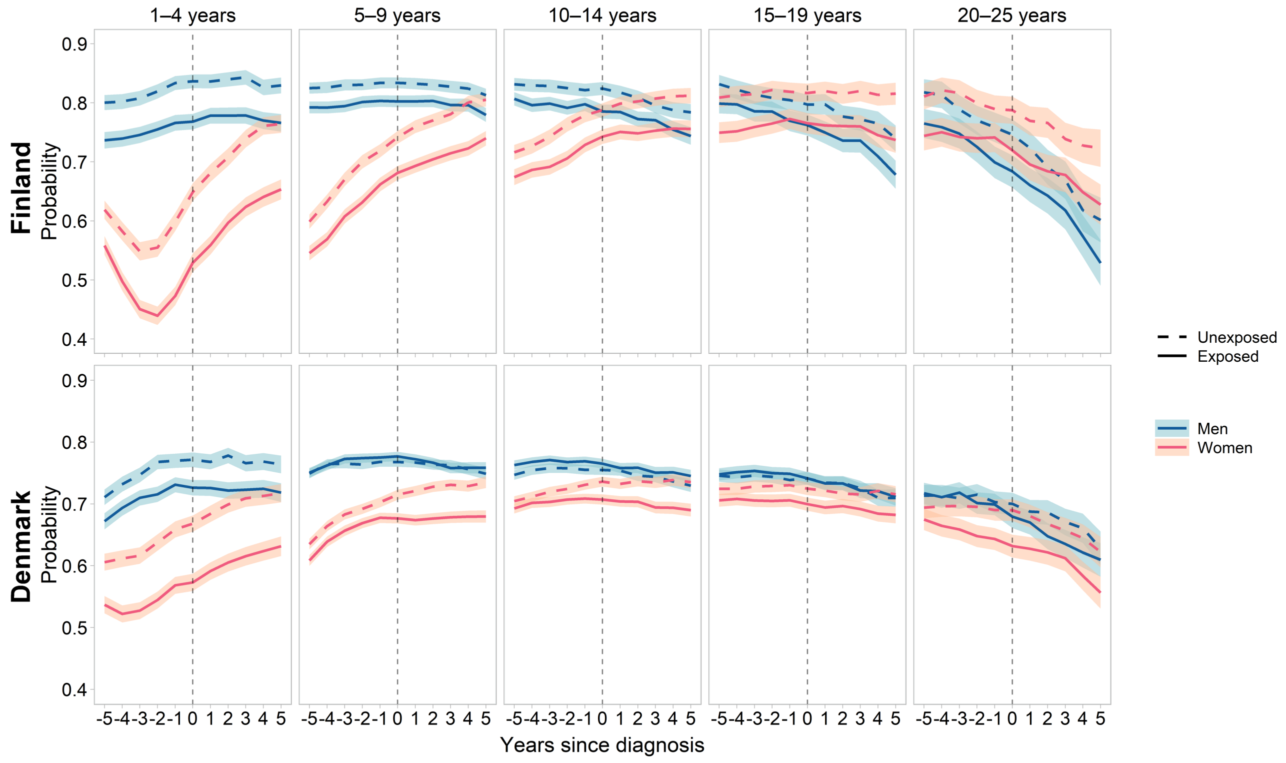

**eFigure 39. Marginal predictions of annual earnings among parents exposed to child's childhood onset disorder (F90–F98) and unexposed parents stratified by the child's age at diagnosis**

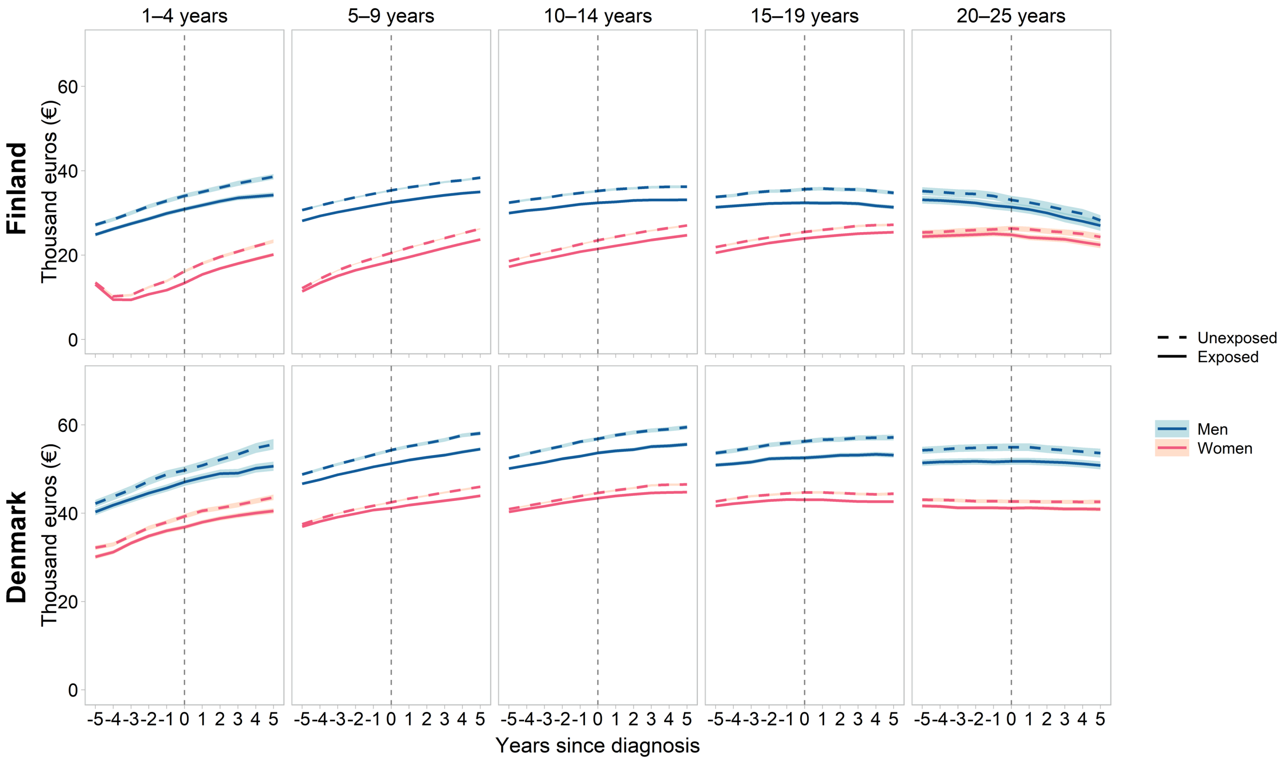

**eFigure 40. Marginal predictions of annual disposable income among parents exposed to child's childhood onset disorder (F90–F98) and unexposed parents stratified by the child's age at diagnosis**

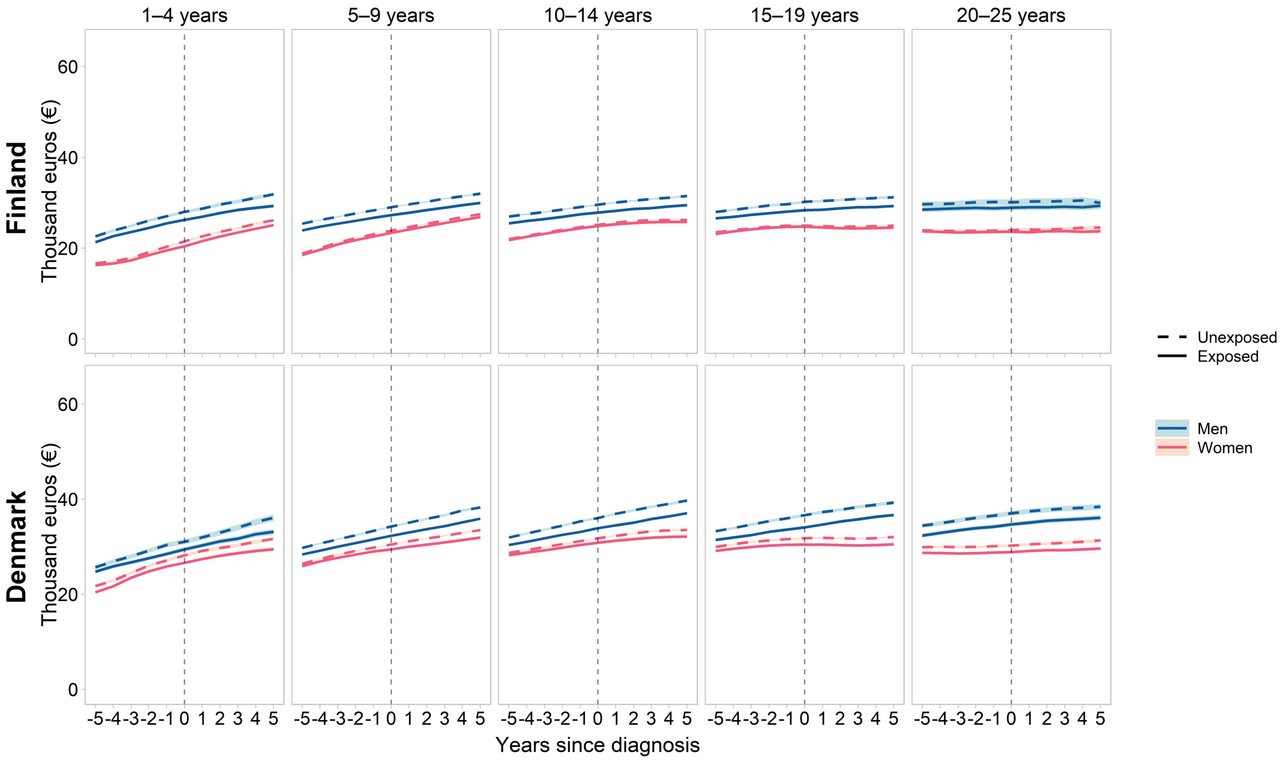

**eFigure 41. Marginal predictions of annual social welfare benefits among parents exposed to child's childhood onset disorder (F90–F98) and unexposed parents stratified by the child's age at diagnosis**

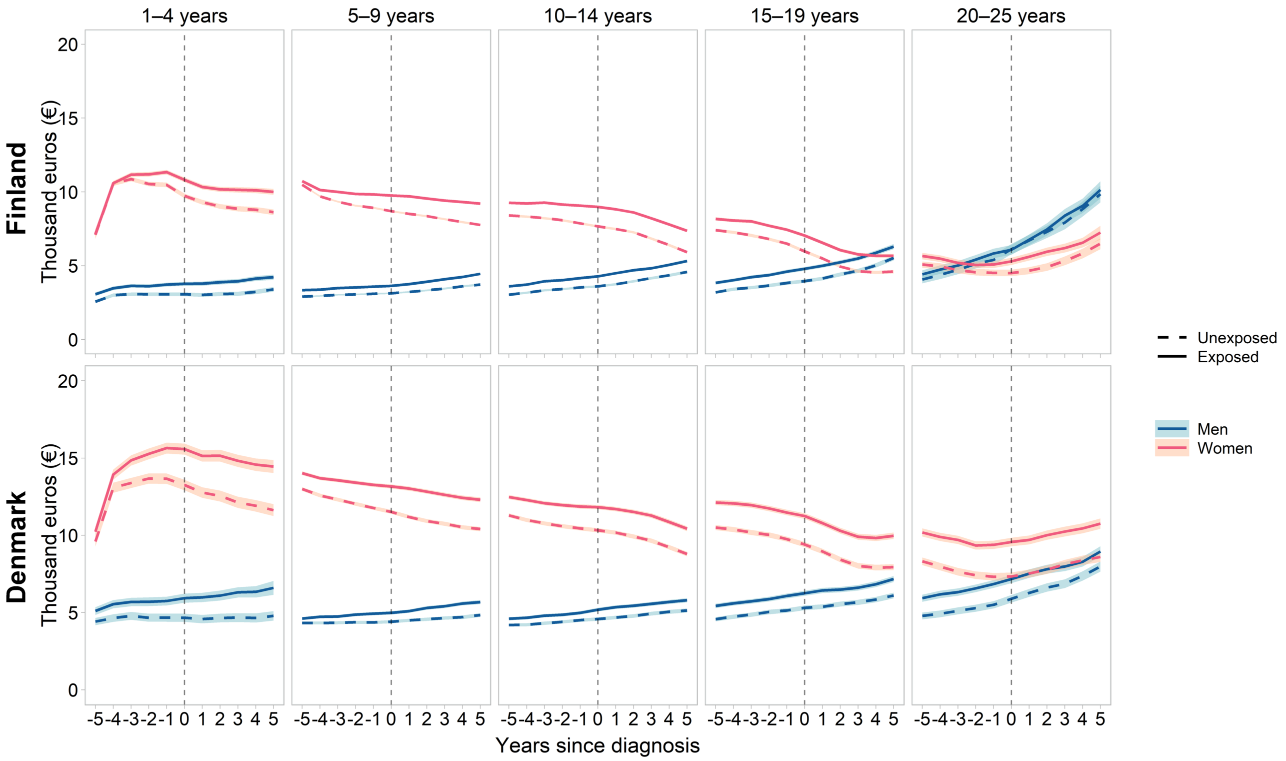

**eFigure 42. Marginal predictions of annual employment among parents exposed to child's childhood onset disorder (F90–F98) and unexposed parents stratified by the child's age at diagnosis**

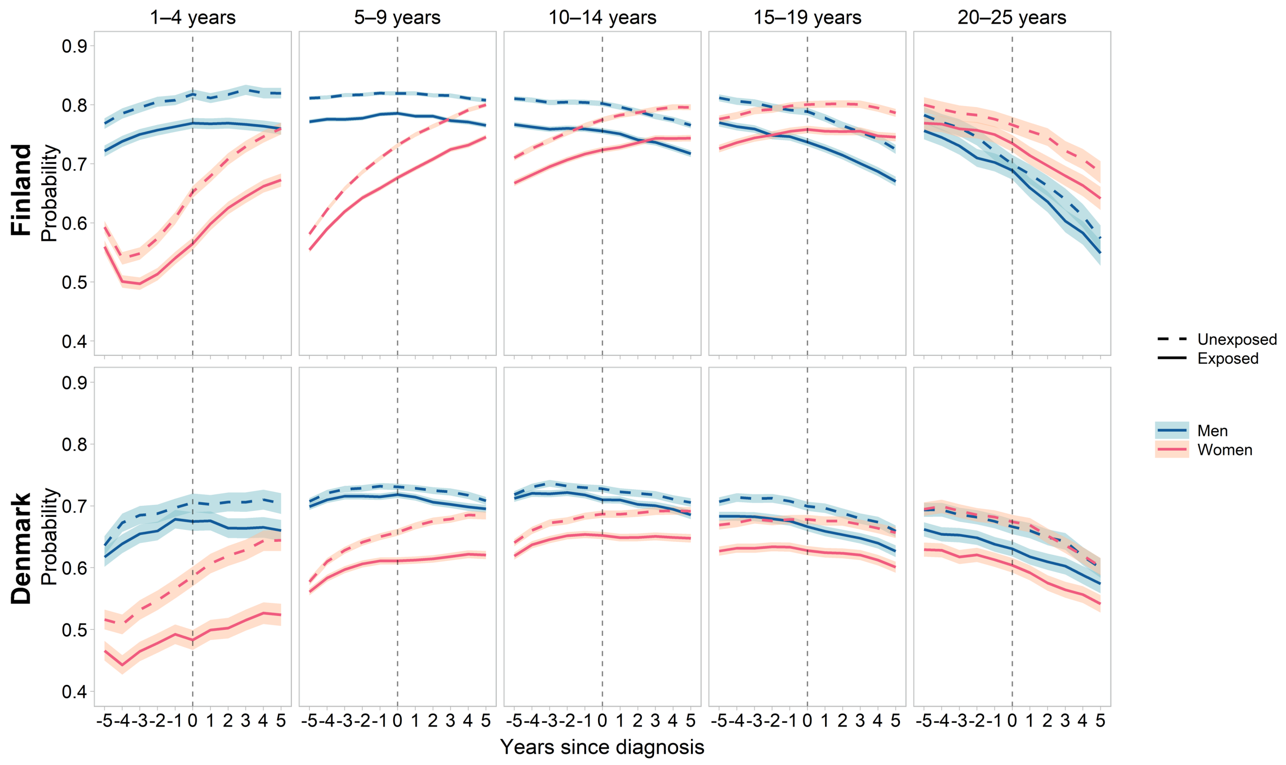

#### Estimates for marginal predictions of annual income and employment among parents exposed to child’s mental disorder and unexposed parents in Finland and Denmark

**eTable 12. Estimates for marginal predictions of annual income and employment among parents exposed to child's any mental disorder (F00–F99) and unexposed parents**

| | | Finland | | | | Denmark | | | |
|---|---|---|---|---|---|---|---|---|---|
| | | Men | | Women | | Men | | Women | |
| | | Unexposed | Exposed | Unexposed | Exposed | Unexposed | Exposed | Unexposed | Exposed |
| Years since diagnosis | Outcome | Marg. pred. (95% CI) | Marg. pred. (95% CI) | Marg. pred. (95% CI) | Marg. pred. (95% CI) | Marg. pred. (95% CI) | Marg. pred. (95% CI) | Marg. pred. (95% CI) | Marg. pred. (95% CI) |
| | | Unexposed | Exposed | Unexposed | Exposed | Unexposed | Exposed | Unexposed | Exposed |
| -5 | Earnings* | 32.3 (32.2–32.4) | 30.2 (30.1–30.3) | 19.1 (19–19.1) | 17.8 (17.7–17.8) | 54.3 (54.1–54.4) | 52 (51.9–52.1) | 41.2 (41.1–41.3) | 40.4 (40.3–40.4) |
| -4 | | 33 (32.9–33.1) | 30.8 (30.7–30.9) | 19.8 (19.7–19.8) | 18.4 (18.3–18.4) | 55 (54.9–55.2) | 52.6 (52.5–52.7) | 41.9 (41.8–42) | 41 (40.9–41) |
| -3 | | 33.7 (33.6–33.7) | 31.3 (31.2–31.4) | 20.6 (20.5–20.6) | 19 (19–19.1) | 55.7 (55.6–55.9) | 53.2 (53.1–53.3) | 42.5 (42.4–42.6) | 41.5 (41.4–41.6) |
| -2 | | 34.2 (34.1–34.3) | 31.8 (31.7–31.9) | 21.5 (21.5–21.6) | 19.8 (19.8–19.9) | 56.5 (56.3–56.6) | 53.8 (53.7–54) | 43.1 (43–43.2) | 42 (41.9–42.1) |
| -1 | | 34.8 (34.7–34.9) | 32.3 (32.2–32.4) | 22.5 (22.4–22.5) | 20.6 (20.6–20.7) | 57.2 (57–57.3) | 54.4 (54.2–54.5) | 43.6 (43.5–43.7) | 42.5 (42.4–42.5) |
| 0 | | 35.3 (35.2–35.4) | 32.7 (32.6–32.8) | 23.4 (23.3–23.5) | 21.4 (21.3–21.5) | 57.8 (57.6–57.9) | 54.8 (54.7–54.9) | 44 (43.9–44.1) | 42.7 (42.6–42.8) |
| 1 | | 35.6 (35.5–35.7) | 32.9 (32.8–33) | 24.2 (24.1–24.2) | 22.1 (22.1–22.2) | 58.2 (58.1–58.4) | 55.2 (55–55.3) | 44.4 (44.3–44.5) | 43.1 (43–43.1) |
| 2 | | 35.8 (35.7–35.9) | 33 (32.9–33.1) | 24.8 (24.8–24.9) | 22.8 (22.7–22.8) | 58.6 (58.4–58.7) | 55.4 (55.2–55.5) | 44.7 (44.6–44.8) | 43.3 (43.2–43.3) |

**eTable 12. Estimates for marginal predictions of annual income and employment among parents exposed to child's any mental disorder (F00–F99) and unexposed parents**

| | | Finland | | | | Denmark | | | |
|---|---|---|---|---|---|---|---|---|---|
| | | Men | | Women | | Men | | Women | |
| | | Unexposed | Exposed | Unexposed | Exposed | Unexposed | Exposed | Unexposed | Exposed |
| Years since diagnosis | Outcome | Marg. pred. (95% CI) | Marg. pred. (95% CI) | Marg. pred. (95% CI) | Marg. pred. (95% CI) | Marg. pred. (95% CI) | Marg. pred. (95% CI) | Marg. pred. (95% CI) | Marg. pred. (95% CI) |
| | | Unexposed | Exposed | Unexposed | Exposed | Unexposed | Exposed | Unexposed | Exposed |
| 3 | | 35.9 (35.8–36) | 33.1 (33–33.2) | 25.4 (25.4–25.5) | 23.3 (23.3–23.4) | 58.9 (58.7–59) | 55.5 (55.4–55.7) | 44.9 (44.8–45) | 43.4 (43.3–43.5) |
| 4 | | 35.9 (35.8–36) | 33 (32.9–33.1) | 26 (25.9–26) | 23.8 (23.7–23.9) | 59.1 (58.9–59.2) | 55.7 (55.5–55.8) | 45.2 (45.1–45.3) | 43.6 (43.5–43.7) |
| 5 | | 35.8 (35.7–35.9) | 32.9 (32.8–33) | 26.4 (26.3–26.4) | 24.2 (24.1–24.3) | 59.1 (58.9–59.2) | 55.7 (55.5–55.8) | 45.4 (45.3–45.5) | 43.7 (43.6–43.8) |
| -5 | Disposable income* | 26.5 (26.5–26.6) | 25.4 (25.3–25.4) | 20.8 (20.8–20.8) | 20.5 (20.4–20.5) | 33.1 (33–33.2) | 31.6 (31.6–31.7) | 28.4 (28.3–28.4) | 27.8 (27.7–27.8) |
| -4 | | 27.2 (27.1–27.3) | 26 (25.9–26) | 21.4 (21.3–21.4) | 21 (21–21.1) | 33.9 (33.8–34) | 32.3 (32.2–32.4) | 29 (28.9–29) | 28.3 (28.2–28.3) |
| -3 | | 27.8 (27.7–27.8) | 26.4 (26.4–26.5) | 21.9 (21.9–22) | 21.6 (21.5–21.6) | 34.6 (34.6–34.7) | 33 (32.9–33.1) | 29.5 (29.4–29.5) | 28.8 (28.7–28.8) |
| -2 | | 28.4 (28.3–28.4) | 27 (26.9–27.1) | 22.5 (22.4–22.5) | 22.1 (22–22.1) | 35.4 (35.4–35.5) | 33.7 (33.6–33.8) | 30 (29.9–30) | 29.2 (29.1–29.2) |
| -1 | | 29 (28.9–29.1) | 27.5 (27.5–27.6) | 23 (23–23.1) | 22.6 (22.6–22.6) | 36.2 (36.1–36.3) | 34.4 (34.3–34.4) | 30.4 (30.4–30.5) | 29.6 (29.6–29.7) |
| 0 | | 29.5 (29.4–29.6) | 28 (27.9–28.1) | 23.5 (23.5–23.6) | 23 (23–23.1) | 36.9 (36.9–37) | 35 (34.9–35.1) | 30.9 (30.8–30.9) | 29.9 (29.8–29.9) |

**eTable 12. Estimates for marginal predictions of annual income and employment among parents exposed to child's any mental disorder (F00–F99) and unexposed parents**

| | | Finland | | | | Denmark | | | |
|---|---|---|---|---|---|---|---|---|---|
| | | Men | | Women | | Men | | Women | |
| | | Unexposed | Exposed | Unexposed | Exposed | Unexposed | Exposed | Unexposed | Exposed |
| Years since diagnosis | Outcome | Marg. pred. (95% CI) | Marg. pred. (95% CI) | Marg. pred. (95% CI) | Marg. pred. (95% CI) | Marg. pred. (95% CI) | Marg. pred. (95% CI) | Marg. pred. (95% CI) | Marg. pred. (95% CI) |
| | | Unexposed | Exposed | Unexposed | Exposed | Unexposed | Exposed | Unexposed | Exposed |
| 1 | | 30 (29.9–30.1) | 28.4 (28.4–28.5) | 24 (23.9–24) | 23.5 (23.4–23.5) | 37.6 (37.5–37.7) | 35.6 (35.5–35.7) | 31.2 (31.2–31.3) | 30.2 (30.2–30.3) |
| 2 | | 30.5 (30.4–30.5) | 28.8 (28.8–28.9) | 24.4 (24.3–24.4) | 23.9 (23.8–23.9) | 38.2 (38.1–38.3) | 36.1 (36–36.2) | 31.6 (31.5–31.6) | 30.5 (30.4–30.5) |
| 3 | | 30.9 (30.9–31) | 29.2 (29.1–29.3) | 24.8 (24.7–24.8) | 24.2 (24.2–24.3) | 38.8 (38.7–38.9) | 36.6 (36.5–36.6) | 31.9 (31.8–31.9) | 30.7 (30.7–30.8) |
| 4 | | 31.3 (31.3–31.4) | 29.6 (29.5–29.6) | 25.2 (25.1–25.2) | 24.6 (24.5–24.6) | 39.4 (39.2–39.5) | 37 (36.9–37.1) | 32.2 (32.2–32.3) | 30.9 (30.9–31) |
| 5 | | 31.7 (31.7–31.8) | 29.9 (29.8–30) | 25.5 (25.5–25.6) | 24.9 (24.9–24.9) | 39.8 (39.7–39.9) | 37.5 (37.4–37.6) | 32.5 (32.5–32.6) | 31.2 (31.1–31.3) |
| -5 | Social welfare benefits* | 2.8 (2.8–2.9) | 3.3 (3.3–3.4) | 7 (6.9–7) | 7.6 (7.6–7.7) | 3.9 (3.9–3.9) | 4.5 (4.5–4.5) | 9.1 (9.1–9.2) | 10.3 (10.3–10.4) |
| -4 | | 3 (3–3) | 3.5 (3.5–3.5) | 6.9 (6.9–7) | 7.7 (7.7–7.7) | 4 (3.9–4) | 4.6 (4.6–4.7) | 9 (8.9–9) | 10.3 (10.2–10.3) |
| -3 | | 3.2 (3.2–3.2) | 3.7 (3.7–3.8) | 6.9 (6.9–6.9) | 7.8 (7.8–7.8) | 4.1 (4–4.1) | 4.7 (4.7–4.8) | 8.8 (8.8–8.9) | 10.1 (10.1–10.2) |
| -2 | | 3.3 (3.3–3.3) | 3.9 (3.9–3.9) | 6.6 (6.6–6.7) | 7.6 (7.6–7.6) | 4.2 (4.1–4.2) | 4.9 (4.8–4.9) | 8.6 (8.6–8.6) | 10 (10–10) |
| -1 | | 3.4 (3.4–3.4) | 4 (4–4) | 6.4 (6.4–6.4) | 7.4 (7.4–7.4) | 4.2 (4.2–4.3) | 5 (4.9–5) | 8.4 (8.3–8.4) | 9.8 (9.8–9.9) |

**eTable 12. Estimates for marginal predictions of annual income and employment among parents exposed to child's any mental disorder (F00–F99) and unexposed parents**

| | | Finland | | | | Denmark | | | |
|---|---|---|---|---|---|---|---|---|---|
| | | Men | | Women | | Men | | Women | |
| | | Unexposed | Exposed | Unexposed | Exposed | Unexposed | Exposed | Unexposed | Exposed |
| Years since diagnosis | Outcome | Marg. pred. (95% CI) | Marg. pred. (95% CI) | Marg. pred. (95% CI) | Marg. pred. (95% CI) | Marg. pred. (95% CI) | Marg. pred. (95% CI) | Marg. pred. (95% CI) | Marg. pred. (95% CI) |
| | | Unexposed | Exposed | Unexposed | Exposed | Unexposed | Exposed | Unexposed | Exposed |
| 0 | | 3.6 (3.5–3.6) | 4.2 (4.1–4.2) | 6.1 (6.1–6.2) | 7.2 (7.2–7.2) | 4.3 (4.3–4.4) | 5.1 (5.1–5.2) | 8.2 (8.1–8.2) | 9.7 (9.7–9.8) |
| 1 | | 3.8 (3.7–3.8) | 4.4 (4.4–4.4) | 6 (5.9–6) | 7.1 (7–7.1) | 4.5 (4.4–4.5) | 5.3 (5.3–5.3) | 8 (7.9–8) | 9.6 (9.6–9.7) |
| 2 | | 4 (4–4) | 4.6 (4.6–4.7) | 5.8 (5.8–5.8) | 6.9 (6.9–6.9) | 4.6 (4.6–4.7) | 5.5 (5.4–5.5) | 7.8 (7.8–7.8) | 9.4 (9.4–9.5) |
| 3 | | 4.3 (4.3–4.3) | 4.9 (4.9–5) | 5.7 (5.7–5.7) | 6.8 (6.8–6.8) | 4.8 (4.7–4.8) | 5.7 (5.6–5.7) | 7.6 (7.6–7.7) | 9.3 (9.3–9.4) |
| 4 | | 4.6 (4.6–4.7) | 5.2 (5.2–5.3) | 5.6 (5.6–5.7) | 6.8 (6.7–6.8) | 5 (4.9–5) | 5.9 (5.9–6) | 7.5 (7.5–7.6) | 9.3 (9.2–9.3) |
| 5 | | 5 (5–5.1) | 5.6 (5.6–5.7) | 5.7 (5.6–5.7) | 6.8 (6.7–6.8) | 5.3 (5.2–5.3) | 6.2 (6.1–6.2) | 7.5 (7.4–7.5) | 9.3 (9.2–9.3) |
| -5 | Employment** | 0.82 (0.82–0.82) | 0.78 (0.78–0.78) | 0.73 (0.73–0.73) | 0.68 (0.68–0.69) | 0.73 (0.73–0.73) | 0.71 (0.71–0.71) | 0.67 (0.67–0.67) | 0.64 (0.64–0.64) |
| -4 | | 0.81 (0.81–0.82) | 0.78 (0.78–0.78) | 0.74 (0.73–0.74) | 0.69 (0.69–0.69) | 0.74 (0.74–0.75) | 0.72 (0.72–0.73) | 0.69 (0.69–0.69) | 0.65 (0.65–0.65) |
| -3 | | 0.81 (0.81–0.81) | 0.77 (0.77–0.77) | 0.74 (0.74–0.75) | 0.7 (0.69–0.7) | 0.75 (0.75–0.75) | 0.73 (0.72–0.73) | 0.7 (0.7–0.7) | 0.66 (0.66–0.66) |
| -2 | | 0.81 (0.81–0.81) | 0.77 (0.77–0.77) | 0.76 (0.75–0.76) | 0.71 (0.7–0.71) | 0.74 (0.74–0.75) | 0.72 (0.72–0.72) | 0.7 (0.7–0.71) | 0.66 (0.66–0.67) |
| -1 | | 0.81 (0.81–0.81) | 0.77 (0.77–0.77) | 0.77 (0.77–0.77) | 0.72 (0.71–0.72) | 0.74 (0.74–0.74) | 0.72 (0.72–0.72) | 0.71 (0.7–0.71) | 0.66 (0.66–0.67) |

**eTable 12. Estimates for marginal predictions of annual income and employment among parents exposed to child's any mental disorder (F00–F99) and unexposed parents**

| | | Finland | | | | Denmark | | | |
|---|---|---|---|---|---|---|---|---|---|
| | | Men | | Women | | Men | | Women | |
| | | Unexposed | Exposed | Unexposed | Exposed | Unexposed | Exposed | Unexposed | Exposed |
| Years since diagnosis | Outcome | Marg. pred. (95% CI) | Marg. pred. (95% CI) | Marg. pred. (95% CI) | Marg. pred. (95% CI) | Marg. pred. (95% CI) | Marg. pred. (95% CI) | Marg. pred. (95% CI) | Marg. pred. (95% CI) |
| | | Unexposed | Exposed | Unexposed | Exposed | Unexposed | Exposed | Unexposed | Exposed |
| 0 | | 0.8 (0.8–0.8) | 0.76 (0.76–0.77) | 0.78 (0.78–0.78) | 0.73 (0.72–0.73) | 0.73 (0.73–0.74) | 0.71 (0.71–0.71) | 0.71 (0.7–0.71) | 0.66 (0.66–0.66) |
| 1 | | 0.8 (0.79–0.8) | 0.76 (0.75–0.76) | 0.79 (0.78–0.79) | 0.73 (0.73–0.73) | 0.73 (0.72–0.73) | 0.7 (0.7–0.7) | 0.71 (0.7–0.71) | 0.65 (0.65–0.66) |
| 2 | | 0.79 (0.79–0.79) | 0.75 (0.75–0.75) | 0.79 (0.79–0.79) | 0.74 (0.74–0.74) | 0.72 (0.72–0.72) | 0.69 (0.69–0.69) | 0.7 (0.7–0.71) | 0.65 (0.65–0.65) |
| 3 | | 0.78 (0.78–0.78) | 0.74 (0.74–0.74) | 0.79 (0.79–0.79) | 0.74 (0.74–0.74) | 0.71 (0.71–0.71) | 0.68 (0.68–0.69) | 0.7 (0.7–0.7) | 0.65 (0.65–0.65) |
| 4 | | 0.77 (0.76–0.77) | 0.73 (0.72–0.73) | 0.79 (0.79–0.79) | 0.74 (0.74–0.74) | 0.7 (0.7–0.7) | 0.67 (0.67–0.67) | 0.7 (0.69–0.7) | 0.64 (0.64–0.64) |
| 5 | | 0.75 (0.75–0.75) | 0.71 (0.71–0.71) | 0.79 (0.79–0.79) | 0.74 (0.73–0.74) | 0.69 (0.68–0.69) | 0.66 (0.66–0.66) | 0.69 (0.69–0.69) | 0.63 (0.63–0.63) |

Marg. pred. = Marginal prediction, * Thousand euros (€), ** Probability

**eTable 13. Estimates for marginal predictions of annual income and employment among parents exposed to child's substance use disorder (F10–F19) and unexposed parents**

| | | Finland | | | | Denmark | | | |
|---|---|---|---|---|---|---|---|---|---|
| | | Men | | Women | | Men | | Women | |
| | | Unexposed | Exposed | Unexposed | Exposed | Unexposed | Exposed | Unexposed | Exposed |
| Years since diagnosis | Outcome | Marg. pred. (95% CI) | Marg. pred. (95% CI) | Marg. pred. (95% CI) | Marg. pred. (95% CI) | Marg. pred. (95% CI) | Marg. pred. (95% CI) | Marg. pred. (95% CI) | Marg. pred. (95% CI) |
| | | Unexposed | Exposed | Unexposed | Exposed | Unexposed | Exposed | Unexposed | Exposed |
| -5 | Earnings* | 31.5 (31.3–31.8) | 29 (28.8–29.2) | 21.5 (21.4–21.7) | 20.2 (20.1–20.3) | 51.8 (51.4–52.2) | 48.2 (47.8–48.6) | 40.3 (40.1–40.5) | 39.7 (39.5–39.9) |
| -4 | | 31.6 (31.4–31.8) | 29.1 (28.9–29.3) | 22.1 (21.9–22.2) | 20.7 (20.6–20.9) | 51.9 (51.5–52.3) | 48.3 (47.9–48.7) | 40.6 (40.4–40.8) | 39.9 (39.7–40.1) |
| -3 | | 31.7 (31.4–31.9) | 29 (28.8–29.2) | 22.5 (22.4–22.6) | 21.2 (21–21.3) | 52.2 (51.8–52.7) | 48.5 (48.1–48.8) | 40.8 (40.6–41) | 40 (39.8–40.2) |
| -2 | | 31.5 (31.3–31.8) | 29 (28.8–29.3) | 22.9 (22.7–23) | 21.6 (21.4–21.7) | 52.4 (51.9–52.8) | 48.6 (48.2–49) | 40.9 (40.7–41.1) | 40 (39.8–40.2) |
| -1 | | 31.5 (31.3–31.8) | 29 (28.8–29.3) | 23.2 (23.1–23.4) | 21.9 (21.8–22.1) | 52.5 (52.1–53) | 48.7 (48.3–49.1) | 41 (40.8–41.2) | 40 (39.8–40.2) |
| 0 | | 31.4 (31.1–31.6) | 28.9 (28.7–29.1) | 23.5 (23.3–23.6) | 22.1 (22–22.3) | 52.6 (52.2–53.1) | 48.5 (48.1–48.9) | 41.1 (40.8–41.3) | 39.9 (39.7–40.1) |
| 1 | | 31.1 (30.8–31.3) | 28.6 (28.4–28.9) | 23.7 (23.5–23.8) | 22.3 (22.2–22.5) | 52.8 (52.3–53.2) | 48.5 (48.1–48.9) | 41 (40.8–41.3) | 40 (39.8–40.2) |
| 2 | | 30.8 (30.5–31) | 28.2 (28–28.5) | 23.8 (23.6–24) | 22.4 (22.2–22.5) | 52.6 (52.1–53) | 48.6 (48.2–49) | 41 (40.8–41.2) | 40 (39.8–40.3) |
| 3 | | 30.2 (30–30.5) | 27.7 (27.4–27.9) | 23.8 (23.7–24) | 22.4 (22.3–22.6) | 52.4 (51.9–52.9) | 48.3 (47.9–48.7) | 41 (40.8–41.2) | 40 (39.8–40.3) |

**eTable 13. Estimates for marginal predictions of annual income and employment among parents exposed to child's substance use disorder (F10–F19) and unexposed parents**

| | | Finland | | | | Denmark | | | |
|---|---|---|---|---|---|---|---|---|---|
| | | Men | | Women | | Men | | Women | |
| | | Unexposed | Exposed | Unexposed | Exposed | Unexposed | Exposed | Unexposed | Exposed |
| Years since diagnosis | Outcome | Marg. pred. (95% CI) | Marg. pred. (95% CI) | Marg. pred. (95% CI) | Marg. pred. (95% CI) | Marg. pred. (95% CI) | Marg. pred. (95% CI) | Marg. pred. (95% CI) | Marg. pred. (95% CI) |
| | | Unexposed | Exposed | Unexposed | Exposed | Unexposed | Exposed | Unexposed | Exposed |
| 4 | | 29.6 (29.3–29.8) | 27.1 (26.9–27.4) | 23.8 (23.6–23.9) | 22.4 (22.2–22.6) | 52.1 (51.6–52.5) | 48 (47.6–48.4) | 41.1 (40.8–41.3) | 39.9 (39.7–40.2) |
| 5 | | 28.8 (28.5–29.1) | 26.5 (26.2–26.8) | 23.6 (23.5–23.8) | 22.3 (22.2–22.5) | 51.7 (51.2–52.2) | 47.9 (47.5–48.4) | 41 (40.8–41.3) | 39.8 (39.6–40.1) |
| -5 | Disposable income* | 26.9 (26.8–27.1) | 25.5 (25.4–25.7) | 22.4 (22.3–22.5) | 22.1 (22–22.2) | 31.8 (31.6–32.1) | 29.5 (29.3–29.7) | 27.7 (27.6–27.9) | 27.2 (27–27.3) |
| -4 | | 27.2 (27–27.3) | 25.8 (25.6–25.9) | 22.6 (22.5–22.7) | 22.3 (22.2–22.3) | 32.3 (32.1–32.5) | 30 (29.8–30.2) | 28 (27.9–28.1) | 27.3 (27.2–27.5) |
| -3 | | 27.5 (27.3–27.7) | 26 (25.8–26.1) | 22.7 (22.6–22.8) | 22.4 (22.4–22.5) | 32.9 (32.6–33.1) | 30.4 (30.2–30.6) | 28.2 (28.1–28.3) | 27.4 (27.3–27.5) |
| -2 | | 27.7 (27.6–27.9) | 26.2 (26.1–26.4) | 22.8 (22.7–22.8) | 22.5 (22.4–22.6) | 33.4 (33.1–33.6) | 30.8 (30.6–31.1) | 28.3 (28.2–28.5) | 27.5 (27.3–27.6) |
| -1 | | 28 (27.9–28.2) | 26.5 (26.4–26.7) | 22.9 (22.8–23) | 22.6 (22.5–22.7) | 33.8 (33.6–34.1) | 31.3 (31–31.5) | 28.5 (28.4–28.6) | 27.5 (27.4–27.6) |
| 0 | | 28.3 (28.1–28.4) | 26.7 (26.6–26.9) | 22.9 (22.8–23) | 22.6 (22.5–22.7) | 34.3 (34.1–34.6) | 31.5 (31.3–31.7) | 28.6 (28.5–28.7) | 27.6 (27.4–27.7) |
| 1 | | 28.4 (28.3–28.6) | 26.8 (26.7–27) | 23 (22.9–23.1) | 22.7 (22.6–22.7) | 34.8 (34.5–35.1) | 31.9 (31.6–32.1) | 28.7 (28.5–28.8) | 27.7 (27.5–27.8) |

**eTable 13. Estimates for marginal predictions of annual income and employment among parents exposed to child's substance use disorder (F10–F19) and unexposed parents**

| | | Finland | | | | Denmark | | | |
|---|---|---|---|---|---|---|---|---|---|
| | | Men | | Women | | Men | | Women | |
| | | Unexposed | Exposed | Unexposed | Exposed | Unexposed | Exposed | Unexposed | Exposed |
| Years since diagnosis | Outcome | Marg. pred. (95% CI) | Marg. pred. (95% CI) | Marg. pred. (95% CI) | Marg. pred. (95% CI) | Marg. pred. (95% CI) | Marg. pred. (95% CI) | Marg. pred. (95% CI) | Marg. pred. (95% CI) |
| | | Unexposed | Exposed | Unexposed | Exposed | Unexposed | Exposed | Unexposed | Exposed |
| 2 | | 28.7 (28.5–28.8) | 27 (26.8–27.2) | 23.1 (23–23.2) | 22.7 (22.6–22.8) | 35.1 (34.8–35.4) | 32.3 (32.1–32.6) | 28.8 (28.7–29) | 27.8 (27.7–28) |
| 3 | | 28.8 (28.7–29) | 27.1 (27–27.3) | 23.2 (23.1–23.3) | 22.8 (22.7–22.9) | 35.4 (35.1–35.7) | 32.5 (32.2–32.8) | 29 (28.8–29.1) | 28 (27.8–28.1) |
| 4 | | 29 (28.8–29.2) | 27.3 (27.1–27.4) | 23.3 (23.2–23.4) | 22.9 (22.8–23) | 35.6 (35.3–35.8) | 32.7 (32.4–32.9) | 29.1 (29–29.3) | 28.1 (27.9–28.2) |
| 5 | | 29.1 (28.9–29.3) | 27.4 (27.2–27.6) | 23.4 (23.4–23.5) | 23 (22.9–23.1) | 35.7 (35.4–36) | 33 (32.7–33.2) | 29.3 (29.2–29.5) | 28.2 (28–28.3) |
| -5 | Social welfare benefits* | 3.8 (3.8–3.9) | 4.5 (4.4–4.5) | 6.6 (6.5–6.6) | 7.4 (7.3–7.4) | 4.9 (4.8–5.1) | 6.6 (6.5–6.7) | 9.3 (9.2–9.5) | 11.3 (11.2–11.5) |
| -4 | | 4.1 (4–4.1) | 4.7 (4.6–4.7) | 6.3 (6.3–6.4) | 7.2 (7.1–7.2) | 5.1 (5–5.3) | 6.8 (6.7–6.9) | 9.1 (8.9–9.2) | 11.1 (10.9–11.2) |
| -3 | | 4.3 (4.3–4.4) | 4.9 (4.9–5) | 6.1 (6–6.1) | 6.9 (6.9–7) | 5.3 (5.2–5.5) | 7 (6.9–7.1) | 8.8 (8.7–8.9) | 10.8 (10.6–10.9) |
| -2 | | 4.6 (4.5–4.7) | 5.2 (5.1–5.3) | 5.9 (5.8–5.9) | 6.7 (6.6–6.7) | 5.5 (5.4–5.6) | 7.2 (7.1–7.4) | 8.7 (8.5–8.8) | 10.5 (10.4–10.7) |
| -1 | | 4.9 (4.8–4.9) | 5.4 (5.3–5.5) | 5.6 (5.6–5.7) | 6.4 (6.4–6.5) | 5.7 (5.6–5.8) | 7.4 (7.2–7.5) | 8.5 (8.4–8.6) | 10.3 (10.2–10.5) |

**eTable 13. Estimates for marginal predictions of annual income and employment among parents exposed to child's substance use disorder (F10–F19) and unexposed parents**

| | | Finland | | | | Denmark | | | |
|---|---|---|---|---|---|---|---|---|---|
| | | Men | | Women | | Men | | Women | |
| | | Unexposed | Exposed | Unexposed | Exposed | Unexposed | Exposed | Unexposed | Exposed |
| Years since diagnosis | Outcome | Marg. pred. (95% CI) | Marg. pred. (95% CI) | Marg. pred. (95% CI) | Marg. pred. (95% CI) | Marg. pred. (95% CI) | Marg. pred. (95% CI) | Marg. pred. (95% CI) | Marg. pred. (95% CI) |
| | | Unexposed | Exposed | Unexposed | Exposed | Unexposed | Exposed | Unexposed | Exposed |
| 0 | | 5.1 (5.1–5.2) | 5.7 (5.6–5.8) | 5.4 (5.4–5.5) | 6.2 (6.2–6.3) | 6 (5.8–6.1) | 7.6 (7.4–7.7) | 8.3 (8.2–8.5) | 10.2 (10.1–10.4) |
| 1 | | 5.5 (5.5–5.6) | 6 (5.9–6.1) | 5.3 (5.3–5.4) | 6.1 (6–6.2) | 6.2 (6.1–6.4) | 7.8 (7.7–8) | 8.2 (8.1–8.3) | 10.1 (10–10.3) |
| 2 | | 6 (5.9–6.1) | 6.4 (6.3–6.5) | 5.3 (5.2–5.4) | 6.1 (6–6.2) | 6.4 (6.3–6.6) | 8.1 (8–8.3) | 8.2 (8–8.3) | 10.1 (10–10.3) |
| 3 | | 6.5 (6.4–6.6) | 6.9 (6.8–7) | 5.3 (5.3–5.4) | 6.1 (6–6.2) | 6.7 (6.6–6.9) | 8.4 (8.3–8.6) | 8.2 (8.1–8.4) | 10.2 (10–10.3) |
| 4 | | 7.1 (6.9–7.2) | 7.4 (7.3–7.5) | 5.5 (5.4–5.5) | 6.2 (6.1–6.3) | 7.1 (6.9–7.2) | 8.7 (8.6–8.9) | 8.3 (8.2–8.5) | 10.3 (10.2–10.5) |
| 5 | | 7.7 (7.6–7.8) | 8 (7.9–8.1) | 5.7 (5.6–5.8) | 6.4 (6.3–6.4) | 7.5 (7.4–7.7) | 9.1 (8.9–9.3) | 8.6 (8.4–8.8) | 10.6 (10.4–10.7) |
| -5 | Employment** | 0.77 (0.77–0.78) | 0.72 (0.72–0.73) | 0.76 (0.76–0.77) | 0.72 (0.72–0.73) | 0.69 (0.69–0.7) | 0.63 (0.63–0.64) | 0.65 (0.64–0.65) | 0.59 (0.59–0.6) |
| -4 | | 0.77 (0.76–0.77) | 0.71 (0.71–0.72) | 0.76 (0.76–0.77) | 0.72 (0.72–0.73) | 0.7 (0.69–0.7) | 0.64 (0.63–0.65) | 0.66 (0.66–0.67) | 0.6 (0.6–0.61) |
| -3 | | 0.75 (0.75–0.76) | 0.7 (0.7–0.71) | 0.76 (0.76–0.77) | 0.72 (0.72–0.73) | 0.7 (0.69–0.71) | 0.64 (0.63–0.65) | 0.67 (0.66–0.68) | 0.61 (0.6–0.62) |
| -2 | | 0.74 (0.74–0.75) | 0.69 (0.69–0.7) | 0.76 (0.76–0.77) | 0.72 (0.72–0.73) | 0.69 (0.69–0.7) | 0.63 (0.62–0.64) | 0.66 (0.66–0.67) | 0.61 (0.6–0.61) |

**eTable 13. Estimates for marginal predictions of annual income and employment among parents exposed to child's substance use disorder (F10–F19) and unexposed parents**

| | | Finland | | | | Denmark | | | |
|---|---|---|---|---|---|---|---|---|---|
| | | Men | | Women | | Men | | Women | |
| | | Unexposed | Exposed | Unexposed | Exposed | Unexposed | Exposed | Unexposed | Exposed |
| Years since diagnosis | Outcome | Marg. pred. (95% CI) | Marg. pred. (95% CI) | Marg. pred. (95% CI) | Marg. pred. (95% CI) | Marg. pred. (95% CI) | Marg. pred. (95% CI) | Marg. pred. (95% CI) | Marg. pred. (95% CI) |
| | | Unexposed | Exposed | Unexposed | Exposed | Unexposed | Exposed | Unexposed | Exposed |
| -1 | | 0.73 (0.73–0.74) | 0.69 (0.68–0.69) | 0.77 (0.76–0.77) | 0.72 (0.72–0.73) | 0.68 (0.67–0.69) | 0.62 (0.61–0.63) | 0.66 (0.65–0.66) | 0.6 (0.6–0.61) |
| 0 | | 0.72 (0.72–0.72) | 0.67 (0.67–0.68) | 0.76 (0.76–0.76) | 0.72 (0.72–0.73) | 0.67 (0.66–0.67) | 0.61 (0.61–0.62) | 0.65 (0.65–0.66) | 0.6 (0.59–0.6) |
| 1 | | 0.7 (0.7–0.71) | 0.66 (0.66–0.67) | 0.75 (0.75–0.76) | 0.71 (0.71–0.72) | 0.66 (0.65–0.66) | 0.6 (0.59–0.61) | 0.65 (0.64–0.65) | 0.59 (0.58–0.6) |
| 2 | | 0.69 (0.68–0.69) | 0.64 (0.64–0.65) | 0.75 (0.74–0.75) | 0.7 (0.7–0.71) | 0.65 (0.64–0.65) | 0.59 (0.58–0.59) | 0.64 (0.63–0.65) | 0.58 (0.57–0.59) |
| 3 | | 0.67 (0.66–0.67) | 0.62 (0.62–0.63) | 0.74 (0.73–0.74) | 0.7 (0.69–0.7) | 0.64 (0.63–0.64) | 0.57 (0.56–0.58) | 0.63 (0.62–0.63) | 0.57 (0.57–0.58) |
| 4 | | 0.65 (0.64–0.65) | 0.61 (0.6–0.61) | 0.73 (0.72–0.73) | 0.69 (0.68–0.69) | 0.62 (0.61–0.63) | 0.56 (0.55–0.57) | 0.62 (0.61–0.62) | 0.56 (0.55–0.57) |
| 5 | | 0.62 (0.62–0.63) | 0.58 (0.58–0.59) | 0.71 (0.71–0.72) | 0.68 (0.67–0.68) | 0.6 (0.59–0.61) | 0.55 (0.54–0.55) | 0.6 (0.6–0.61) | 0.55 (0.54–0.55) |

Marg. pred. = Marginal prediction, * Thousand euros (€), ** Probability

**eTable 14. Estimates for marginal predictions of annual income and employment among parents exposed to child's psychotic disorder (F20–F29) and unexposed parents**

| | | Finland | | | | Denmark | | | |
|---|---|---|---|---|---|---|---|---|---|
| | | Men | | Women | | Men | | Women | |
| | | Unexposed | Exposed | Unexposed | Exposed | Unexposed | Exposed | Unexposed | Exposed |
| Years since diagnosis | Outcome | Marg. pred. (95% CI) | Marg. pred. (95% CI) | Marg. pred. (95% CI) | Marg. pred. (95% CI) | Marg. pred. (95% CI) | Marg. pred. (95% CI) | Marg. pred. (95% CI) | Marg. pred. (95% CI) |
| | | Unexposed | Exposed | Unexposed | Exposed | Unexposed | Exposed | Unexposed | Exposed |
| -5 | Earnings* | 33.7 (33.4–34.1) | 30.6 (30.2–30.9) | 22.5 (22.3–22.7) | 20.5 (20.3–20.8) | 55.5 (55.1–56) | 51.8 (51.4–52.3) | 42.6 (42.3–42.8) | 41.3 (41.1–41.6) |
| -4 | | 33.8 (33.5–34.2) | 30.7 (30.4–31.1) | 23.1 (22.9–23.3) | 21 (20.8–21.2) | 56.3 (55.8–56.7) | 52.1 (51.7–52.6) | 42.9 (42.7–43.2) | 41.6 (41.4–41.9) |
| -3 | | 34 (33.6–34.4) | 30.5 (30.2–30.9) | 23.4 (23.2–23.7) | 21.4 (21.2–21.6) | 56.4 (55.9–56.9) | 52.6 (52.2–53) | 43.1 (42.9–43.4) | 41.9 (41.7–42.1) |
| -2 | | 33.9 (33.5–34.3) | 30.4 (30.1–30.8) | 23.8 (23.6–24) | 21.7 (21.4–21.9) | 56.7 (56.2–57.2) | 52.9 (52.4–53.3) | 43.4 (43.1–43.6) | 42 (41.8–42.3) |
| -1 | | 33.8 (33.4–34.2) | 30.3 (29.9–30.6) | 24.1 (23.9–24.3) | 21.9 (21.6–22.1) | 57.1 (56.6–57.6) | 53.1 (52.6–53.5) | 43.6 (43.4–43.9) | 42.1 (41.9–42.4) |
| 0 | | 33.7 (33.3–34.1) | 30.1 (29.7–30.4) | 24.4 (24.1–24.6) | 21.9 (21.7–22.2) | 57.4 (56.9–57.9) | 53.2 (52.7–53.6) | 43.8 (43.5–44) | 42.1 (41.9–42.4) |
| 1 | | 33.3 (32.9–33.7) | 29.7 (29.3–30.1) | 24.5 (24.3–24.8) | 21.9 (21.7–22.2) | 57.5 (57–58.1) | 53.1 (52.6–53.6) | 43.8 (43.6–44.1) | 42.1 (41.9–42.4) |
| 2 | | 32.9 (32.5–33.3) | 29.1 (28.7–29.5) | 24.6 (24.4–24.9) | 22.1 (21.8–22.3) | 57.6 (57–58.1) | 53.2 (52.7–53.7) | 43.9 (43.6–44.2) | 42.1 (41.8–42.4) |
| 3 | | 32.3 (31.9–32.7) | 28.6 (28.2–29) | 24.6 (24.3–24.8) | 22 (21.7–22.2) | 57.5 (57–58.1) | 53.3 (52.8–53.8) | 44 (43.7–44.2) | 42 (41.8–42.3) |

**eTable 14. Estimates for marginal predictions of annual income and employment among parents exposed to child's psychotic disorder (F20–F29) and unexposed parents**

| | | Finland | | | | Denmark | | | |
|---|---|---|---|---|---|---|---|---|---|
| | | Men | | Women | | Men | | Women | |
| | | Unexposed | Exposed | Unexposed | Exposed | Unexposed | Exposed | Unexposed | Exposed |
| Years since diagnosis | Outcome | Marg. pred. (95% CI) | Marg. pred. (95% CI) | Marg. pred. (95% CI) | Marg. pred. (95% CI) | Marg. pred. (95% CI) | Marg. pred. (95% CI) | Marg. pred. (95% CI) | Marg. pred. (95% CI) |
| | | Unexposed | Exposed | Unexposed | Exposed | Unexposed | Exposed | Unexposed | Exposed |
| 4 | | 31.7 (31.3–32.1) | 27.8 (27.4–28.2) | 24.4 (24.2–24.7) | 21.9 (21.7–22.2) | 57.4 (56.8–57.9) | 52.9 (52.3–53.4) | 43.8 (43.5–44.1) | 42 (41.7–42.3) |
| 5 | | 30.8 (30.4–31.3) | 26.9 (26.5–27.3) | 24.2 (23.9–24.5) | 21.7 (21.4–22) | 57.3 (56.7–57.9) | 52.5 (52–53) | 43.9 (43.6–44.2) | 41.9 (41.7–42.2) |
| -5 | Disposable income* | 28.1 (27.8–28.3) | 26.3 (26.1–26.5) | 22.6 (22.5–22.7) | 21.8 (21.7–22) | 34 (33.8–34.3) | 31.8 (31.6–32.1) | 29.2 (29.1–29.3) | 28.3 (28.2–28.4) |
| -4 | | 28.5 (28.2–28.7) | 26.7 (26.5–26.9) | 22.9 (22.8–23) | 22 (21.9–22.1) | 34.9 (34.6–35.2) | 32.4 (32.2–32.6) | 29.5 (29.4–29.7) | 28.6 (28.4–28.7) |
| -3 | | 28.8 (28.6–29.1) | 26.9 (26.7–27.1) | 23 (22.9–23.1) | 22.1 (22–22.3) | 35.4 (35.1–35.6) | 33 (32.8–33.3) | 29.7 (29.6–29.9) | 28.8 (28.7–29) |
| -2 | | 29 (28.8–29.3) | 27.2 (27–27.4) | 23.1 (23–23.2) | 22.3 (22.1–22.4) | 35.9 (35.7–36.2) | 33.5 (33.3–33.8) | 30 (29.9–30.2) | 29 (28.9–29.1) |
| -1 | | 29.3 (29–29.5) | 27.4 (27.1–27.6) | 23.2 (23.1–23.3) | 22.3 (22.1–22.4) | 36.6 (36.3–36.9) | 34 (33.8–34.3) | 30.3 (30.1–30.5) | 29.2 (29–29.3) |
| 0 | | 29.6 (29.4–29.9) | 27.6 (27.4–27.9) | 23.3 (23.2–23.4) | 22.3 (22.1–22.4) | 37.1 (36.8–37.4) | 34.5 (34.2–34.7) | 30.5 (30.4–30.7) | 29.3 (29.1–29.4) |
| 1 | | 29.8 (29.6–30.1) | 27.8 (27.5–28) | 23.4 (23.3–23.5) | 22.3 (22.2–22.4) | 37.7 (37.4–38) | 34.9 (34.6–35.1) | 30.7 (30.5–30.9) | 29.4 (29.2–29.5) |

**eTable 14. Estimates for marginal predictions of annual income and employment among parents exposed to child's psychotic disorder (F20–F29) and unexposed parents**

| | | Finland | | | | Denmark | | | |
|---|---|---|---|---|---|---|---|---|---|
| | | Men | | Women | | Men | | Women | |
| | | Unexposed | Exposed | Unexposed | Exposed | Unexposed | Exposed | Unexposed | Exposed |
| Years since diagnosis | Outcome | Marg. pred. (95% CI) | Marg. pred. (95% CI) | Marg. pred. (95% CI) | Marg. pred. (95% CI) | Marg. pred. (95% CI) | Marg. pred. (95% CI) | Marg. pred. (95% CI) | Marg. pred. (95% CI) |
| | | Unexposed | Exposed | Unexposed | Exposed | Unexposed | Exposed | Unexposed | Exposed |
| 2 | | 30.1 (29.8–30.3) | 27.9 (27.6–28.1) | 23.5 (23.4–23.7) | 22.4 (22.3–22.5) | 38.1 (37.8–38.5) | 35.2 (35–35.5) | 30.9 (30.7–31) | 29.5 (29.3–29.6) |
| 3 | | 30.3 (30–30.6) | 28.1 (27.9–28.4) | 23.6 (23.5–23.8) | 22.5 (22.4–22.6) | 38.5 (38.2–38.9) | 35.8 (35.5–36.1) | 31.1 (30.9–31.2) | 29.6 (29.4–29.8) |
| 4 | | 30.4 (30.2–30.7) | 28.3 (28–28.5) | 23.8 (23.6–23.9) | 22.6 (22.5–22.8) | 38.9 (38.5–39.2) | 35.9 (35.6–36.2) | 31.2 (31–31.4) | 29.7 (29.6–29.9) |
| 5 | | 30.6 (30.3–30.9) | 28.3 (28.1–28.6) | 23.9 (23.7–24) | 22.8 (22.6–22.9) | 39.3 (39–39.7) | 36.1 (35.8–36.4) | 31.4 (31.3–31.6) | 29.9 (29.7–30.1) |
| -5 | Social welfare benefits* | 3.6 (3.5–3.7) | 4.3 (4.2–4.4) | 6.2 (6.2–6.3) | 7 (6.9–7.1) | 4.4 (4.3–4.5) | 5.7 (5.5–5.8) | 8.7 (8.6–8.8) | 10.3 (10.1–10.4) |
| -4 | | 3.9 (3.8–4) | 4.6 (4.5–4.7) | 6 (5.9–6.1) | 6.8 (6.7–6.9) | 4.6 (4.4–4.7) | 5.8 (5.7–5.9) | 8.5 (8.3–8.6) | 10.1 (9.9–10.2) |
| -3 | | 4.1 (4–4.2) | 4.9 (4.8–5) | 5.8 (5.7–5.9) | 6.6 (6.5–6.7) | 4.7 (4.6–4.8) | 5.9 (5.8–6) | 8.2 (8.1–8.4) | 9.9 (9.7–10) |
| -2 | | 4.4 (4.3–4.5) | 5.2 (5.1–5.3) | 5.6 (5.5–5.7) | 6.4 (6.3–6.5) | 4.9 (4.8–5) | 6.1 (6–6.2) | 8 (7.9–8.2) | 9.6 (9.5–9.8) |
| -1 | | 4.7 (4.6–4.8) | 5.5 (5.4–5.6) | 5.4 (5.3–5.5) | 6.2 (6.1–6.3) | 5 (4.9–5.2) | 6.2 (6.1–6.4) | 7.8 (7.7–7.9) | 9.4 (9.3–9.6) |
| 0 | | 5 (4.9–5.2) | 5.8 (5.7–5.9) | 5.2 (5.1–5.3) | 6.1 (6–6.2) | 5.2 (5–5.3) | 6.4 (6.2–6.5) | 7.7 (7.5–7.8) | 9.3 (9.2–9.5) |
| 1 | | 5.5 (5.4–5.6) | 6.3 (6.2–6.4) | 5.2 (5.1–5.3) | 6.1 (6–6.2) | 5.3 (5.2–5.4) | 6.6 (6.5–6.8) | 7.5 (7.4–7.7) | 9.3 (9.1–9.4) |

**eTable 14. Estimates for marginal predictions of annual income and employment among parents exposed to child's psychotic disorder (F20–F29) and unexposed parents**

| | | Finland | | | | Denmark | | | |
|---|---|---|---|---|---|---|---|---|---|
| | | Men | | Women | | Men | | Women | |
| | | Unexposed | Exposed | Unexposed | Exposed | Unexposed | Exposed | Unexposed | Exposed |
| Years since diagnosis | Outcome | Marg. pred. (95% CI) | Marg. pred. (95% CI) | Marg. pred. (95% CI) | Marg. pred. (95% CI) | Marg. pred. (95% CI) | Marg. pred. (95% CI) | Marg. pred. (95% CI) | Marg. pred. (95% CI) |
| | | Unexposed | Exposed | Unexposed | Exposed | Unexposed | Exposed | Unexposed | Exposed |
| 2 | | 6 (5.8–6.1) | 6.7 (6.6–6.9) | 5.2 (5.1–5.3) | 6.1 (6–6.2) | 5.5 (5.4–5.7) | 6.9 (6.7–7) | 7.4 (7.3–7.6) | 9.2 (9.1–9.4) |
| 3 | | 6.6 (6.4–6.7) | 7.3 (7.2–7.5) | 5.3 (5.2–5.4) | 6.2 (6.1–6.3) | 5.8 (5.6–5.9) | 7.1 (7–7.3) | 7.4 (7.2–7.5) | 9.2 (9.1–9.4) |
| 4 | | 7.2 (7–7.4) | 8 (7.8–8.2) | 5.5 (5.4–5.6) | 6.4 (6.3–6.5) | 6.1 (5.9–6.2) | 7.4 (7.3–7.6) | 7.5 (7.4–7.7) | 9.4 (9.2–9.5) |
| 5 | | 7.9 (7.7–8.1) | 8.7 (8.6–8.9) | 5.8 (5.7–5.9) | 6.7 (6.6–6.8) | 6.4 (6.2–6.5) | 7.9 (7.7–8.1) | 7.7 (7.5–7.8) | 9.6 (9.4–9.8) |
| -5 | Employment** | 0.79 (0.78–0.8) | 0.74 (0.73–0.74) | 0.78 (0.77–0.78) | 0.72 (0.71–0.72) | 0.71 (0.7–0.72) | 0.67 (0.67–0.68) | 0.68 (0.67–0.69) | 0.63 (0.62–0.64) |
| -4 | | 0.78 (0.78–0.79) | 0.73 (0.72–0.73) | 0.78 (0.77–0.78) | 0.71 (0.71–0.72) | 0.72 (0.71–0.72) | 0.68 (0.68–0.69) | 0.69 (0.69–0.7) | 0.64 (0.63–0.65) |
| -3 | | 0.77 (0.76–0.77) | 0.72 (0.71–0.72) | 0.77 (0.77–0.78) | 0.71 (0.71–0.72) | 0.72 (0.72–0.73) | 0.69 (0.68–0.69) | 0.7 (0.7–0.71) | 0.65 (0.64–0.65) |
| -2 | | 0.76 (0.75–0.76) | 0.7 (0.7–0.71) | 0.77 (0.77–0.78) | 0.71 (0.7–0.72) | 0.71 (0.71–0.72) | 0.68 (0.67–0.68) | 0.7 (0.69–0.71) | 0.64 (0.64–0.65) |
| -1 | | 0.75 (0.74–0.75) | 0.69 (0.68–0.69) | 0.77 (0.76–0.78) | 0.71 (0.7–0.71) | 0.71 (0.7–0.71) | 0.67 (0.66–0.68) | 0.7 (0.69–0.7) | 0.64 (0.63–0.65) |
| 0 | | 0.73 (0.72–0.74) | 0.68 (0.67–0.68) | 0.76 (0.76–0.77) | 0.7 (0.69–0.7) | 0.7 (0.7–0.71) | 0.66 (0.65–0.67) | 0.69 (0.68–0.69) | 0.63 (0.63–0.64) |

**eTable 14. Estimates for marginal predictions of annual income and employment among parents exposed to child's psychotic disorder (F20–F29) and unexposed parents**

| | | Finland | | | | Denmark | | | |
|---|---|---|---|---|---|---|---|---|---|
| | | Men | | Women | | Men | | Women | |
| | | Unexposed | Exposed | Unexposed | Exposed | Unexposed | Exposed | Unexposed | Exposed |
| Years since diagnosis | Outcome | Marg. pred. (95% CI) | Marg. pred. (95% CI) | Marg. pred. (95% CI) | Marg. pred. (95% CI) | Marg. pred. (95% CI) | Marg. pred. (95% CI) | Marg. pred. (95% CI) | Marg. pred. (95% CI) |
| | | Unexposed | Exposed | Unexposed | Exposed | Unexposed | Exposed | Unexposed | Exposed |
| 1 | | 0.71 (0.71–0.72) | 0.66 (0.65–0.66) | 0.76 (0.75–0.76) | 0.69 (0.68–0.69) | 0.69 (0.68–0.7) | 0.65 (0.64–0.66) | 0.68 (0.68–0.69) | 0.62 (0.62–0.63) |
| 2 | | 0.69 (0.69–0.7) | 0.64 (0.63–0.65) | 0.75 (0.75–0.76) | 0.68 (0.68–0.69) | 0.68 (0.67–0.69) | 0.64 (0.63–0.65) | 0.68 (0.67–0.68) | 0.61 (0.61–0.62) |
| 3 | | 0.68 (0.67–0.68) | 0.62 (0.61–0.62) | 0.74 (0.73–0.75) | 0.67 (0.66–0.68) | 0.67 (0.66–0.68) | 0.63 (0.62–0.63) | 0.67 (0.66–0.68) | 0.61 (0.6–0.61) |
| 4 | | 0.66 (0.65–0.66) | 0.6 (0.59–0.61) | 0.73 (0.72–0.73) | 0.66 (0.65–0.67) | 0.65 (0.65–0.66) | 0.61 (0.6–0.62) | 0.66 (0.65–0.66) | 0.6 (0.59–0.6) |
| 5 | | 0.63 (0.63–0.64) | 0.57 (0.56–0.58) | 0.71 (0.7–0.71) | 0.64 (0.63–0.65) | 0.64 (0.63–0.65) | 0.59 (0.58–0.6) | 0.65 (0.64–0.65) | 0.58 (0.57–0.59) |

Marg. pred. = Marginal prediction, * Thousand euros (€), ** Probability

**eTable 15. Estimates for marginal predictions of annual income and employment among parents exposed to child's mood disorder (F30–F39) and unexposed parents**

| | | Finland | | | | Denmark | | | |
|---|---|---|---|---|---|---|---|---|---|
| | | Men | | Women | | Men | | Women | |
| | | Unexposed | Exposed | Unexposed | Exposed | Unexposed | Exposed | Unexposed | Exposed |
| Years since diagnosis | Outcome | Marg. pred. (95% CI) | Marg. pred. (95% CI) | Marg. pred. (95% CI) | Marg. pred. (95% CI) | Marg. pred. (95% CI) | Marg. pred. (95% CI) | Marg. pred. (95% CI) | Marg. pred. (95% CI) |
| | | Unexposed | Exposed | Unexposed | Exposed | Unexposed | Exposed | Unexposed | Exposed |
| -5 | Earnings* | 34.3 (34.2–34.5) | 32.3 (32.2–32.5) | 22.9 (22.8–23) | 21.6 (21.5–21.7) | 57.4 (57.1–57.7) | 56.3 (56–56.6) | 43.6 (43.4–43.7) | 43.2 (43.1–43.4) |
| -4 | | 34.6 (34.5–34.8) | 32.5 (32.4–32.7) | 23.5 (23.4–23.6) | 22.2 (22.1–22.3) | 57.9 (57.6–58.2) | 56.8 (56.4–57.1) | 44 (43.8–44.1) | 43.6 (43.4–43.7) |
| -3 | | 34.8 (34.6–35) | 32.6 (32.5–32.8) | 24.2 (24.1–24.3) | 22.8 (22.7–22.9) | 58.4 (58–58.7) | 57.1 (56.7–57.4) | 44.3 (44.1–44.4) | 43.9 (43.7–44) |
| -2 | | 34.9 (34.7–35) | 32.7 (32.5–32.8) | 24.7 (24.6–24.8) | 23.3 (23.2–23.4) | 58.9 (58.5–59.2) | 57.3 (57–57.6) | 44.5 (44.3–44.7) | 44.1 (43.9–44.2) |
| -1 | | 35 (34.8–35.1) | 32.6 (32.4–32.7) | 25.1 (25–25.3) | 23.7 (23.6–23.8) | 59.2 (58.8–59.5) | 57.6 (57.3–58) | 44.7 (44.6–44.9) | 44.2 (44.1–44.4) |
| 0 | | 34.9 (34.8–35.1) | 32.5 (32.3–32.6) | 25.6 (25.5–25.7) | 24 (23.9–24.1) | 59.4 (59.1–59.8) | 57.8 (57.5–58.1) | 44.8 (44.7–45) | 44.3 (44.2–44.5) |
| 1 | | 34.8 (34.6–34.9) | 32.2 (32.1–32.4) | 25.9 (25.8–26) | 24.2 (24.1–24.3) | 59.6 (59.2–60) | 57.9 (57.5–58.2) | 45 (44.8–45.2) | 44.3 (44.2–44.5) |
| 2 | | 34.4 (34.3–34.6) | 31.8 (31.6–32) | 26.1 (26–26.3) | 24.4 (24.3–24.5) | 59.7 (59.3–60.1) | 57.9 (57.5–58.2) | 45.1 (44.9–45.3) | 44.3 (44.1–44.5) |
| 3 | | 34 (33.8–34.2) | 31.3 (31.1–31.5) | 26.3 (26.2–26.4) | 24.5 (24.4–24.6) | 59.6 (59.2–60) | 57.7 (57.4–58.1) | 45.1 (44.9–45.3) | 44.3 (44.1–44.5) |

**eTable 15. Estimates for marginal predictions of annual income and employment among parents exposed to child's mood disorder (F30–F39) and unexposed parents**

| | | Finland | | | | Denmark | | | |
|---|---|---|---|---|---|---|---|---|---|
| | | Men | | Women | | Men | | Women | |
| | | Unexposed | Exposed | Unexposed | Exposed | Unexposed | Exposed | Unexposed | Exposed |
| Years since diagnosis | Outcome | Marg. pred. (95% CI) | Marg. pred. (95% CI) | Marg. pred. (95% CI) | Marg. pred. (95% CI) | Marg. pred. (95% CI) | Marg. pred. (95% CI) | Marg. pred. (95% CI) | Marg. pred. (95% CI) |
| | | Unexposed | Exposed | Unexposed | Exposed | Unexposed | Exposed | Unexposed | Exposed |
| 4 | | 33.4 (33.2–33.6) | 30.7 (30.5–30.9) | 26.2 (26.1–26.4) | 24.5 (24.4–24.6) | 59.8 (59.4–60.2) | 57.6 (57.2–57.9) | 45.1 (44.9–45.3) | 44.3 (44.1–44.5) |
| 5 | | 32.8 (32.6–33) | 30.1 (29.9–30.3) | 26.1 (26–26.3) | 24.4 (24.3–24.5) | 59.7 (59.2–60.1) | 57.4 (57–57.8) | 45.1 (44.9–45.3) | 44.2 (44–44.4) |
| -5 | Disposable income* | 28.6 (28.5–28.7) | 27.3 (27.2–27.4) | 23.3 (23.2–23.4) | 23 (22.9–23) | 35.2 (35–35.4) | 34.3 (34.1–34.5) | 29.9 (29.8–30) | 29.5 (29.4–29.6) |
| -4 | | 29 (28.9–29.1) | 27.7 (27.6–27.8) | 23.6 (23.6–23.7) | 23.3 (23.2–23.3) | 35.9 (35.7–36.1) | 35 (34.8–35.2) | 30.3 (30.2–30.4) | 29.8 (29.8–29.9) |
| -3 | | 29.3 (29.2–29.4) | 28 (27.9–28.1) | 23.9 (23.8–23.9) | 23.6 (23.5–23.6) | 36.6 (36.4–36.8) | 35.6 (35.4–35.8) | 30.5 (30.4–30.6) | 30.1 (30–30.2) |
| -2 | | 29.7 (29.6–29.8) | 28.3 (28.2–28.4) | 24.1 (24.1–24.2) | 23.8 (23.7–23.9) | 37.3 (37.1–37.5) | 36.1 (36–36.3) | 30.8 (30.7–30.9) | 30.4 (30.3–30.5) |
| -1 | | 30 (29.9–30.2) | 28.5 (28.4–28.6) | 24.3 (24.2–24.3) | 24 (23.9–24.1) | 37.9 (37.7–38.1) | 36.8 (36.6–37) | 31.1 (31–31.2) | 30.6 (30.5–30.7) |
| 0 | | 30.3 (30.2–30.4) | 28.7 (28.6–28.9) | 24.4 (24.4–24.5) | 24.1 (24–24.1) | 38.5 (38.3–38.7) | 37.3 (37.1–37.5) | 31.3 (31.2–31.4) | 30.8 (30.6–30.9) |
| 1 | | 30.6 (30.5–30.7) | 29 (28.9–29.1) | 24.5 (24.5–24.6) | 24.1 (24.1–24.2) | 39 (38.8–39.2) | 37.8 (37.6–38) | 31.5 (31.4–31.6) | 30.9 (30.8–31) |

**eTable 15. Estimates for marginal predictions of annual income and employment among parents exposed to child's mood disorder (F30–F39) and unexposed parents**

| | | Finland | | | | Denmark | | | |
|---|---|---|---|---|---|---|---|---|---|
| | | Men | | Women | | Men | | Women | |
| | | Unexposed | Exposed | Unexposed | Exposed | Unexposed | Exposed | Unexposed | Exposed |
| Years since diagnosis | Outcome | Marg. pred. (95% CI) | Marg. pred. (95% CI) | Marg. pred. (95% CI) | Marg. pred. (95% CI) | Marg. pred. (95% CI) | Marg. pred. (95% CI) | Marg. pred. (95% CI) | Marg. pred. (95% CI) |
| | | Unexposed | Exposed | Unexposed | Exposed | Unexposed | Exposed | Unexposed | Exposed |
| 2 | | 30.7 (30.6–30.9) | 29.1 (29–29.2) | 24.6 (24.6–24.7) | 24.2 (24.1–24.3) | 39.5 (39.3–39.8) | 38.2 (38–38.4) | 31.8 (31.7–31.9) | 31 (30.9–31.1) |
| 3 | | 31 (30.8–31.1) | 29.2 (29.1–29.3) | 24.8 (24.7–24.8) | 24.3 (24.2–24.3) | 39.9 (39.7–40.1) | 38.6 (38.4–38.8) | 31.9 (31.8–32.1) | 31.2 (31–31.3) |
| 4 | | 31.1 (31–31.3) | 29.3 (29.2–29.4) | 24.8 (24.8–24.9) | 24.3 (24.3–24.4) | 40.5 (40.2–40.7) | 38.9 (38.7–39.2) | 32.2 (32.1–32.3) | 31.3 (31.2–31.5) |
| 5 | | 31.3 (31.1–31.4) | 29.5 (29.3–29.6) | 24.9 (24.9–25) | 24.4 (24.3–24.5) | 40.9 (40.6–41.1) | 39.3 (39.1–39.6) | 32.3 (32.2–32.5) | 31.5 (31.4–31.6) |
| -5 | Social welfare benefits* | 3.4 (3.4–3.5) | 3.9 (3.9–4) | 6.5 (6.4–6.5) | 7.1 (7.1–7.2) | 3.9 (3.9–4) | 4.2 (4.2–4.3) | 8.3 (8.2–8.3) | 9 (8.9–9) |
| -4 | | 3.6 (3.6–3.7) | 4.2 (4.1–4.2) | 6.3 (6.2–6.3) | 7 (6.9–7) | 4.1 (4–4.1) | 4.4 (4.3–4.4) | 8 (8–8.1) | 8.8 (8.7–8.9) |
| -3 | | 3.9 (3.8–3.9) | 4.4 (4.4–4.5) | 6 (6–6.1) | 6.8 (6.8–6.9) | 4.2 (4.1–4.3) | 4.5 (4.5–4.6) | 7.8 (7.7–7.9) | 8.6 (8.5–8.7) |
| -2 | | 4.1 (4.1–4.2) | 4.7 (4.6–4.7) | 5.8 (5.8–5.9) | 6.6 (6.6–6.7) | 4.3 (4.2–4.4) | 4.7 (4.6–4.8) | 7.5 (7.4–7.6) | 8.4 (8.3–8.5) |
| -1 | | 4.3 (4.3–4.4) | 5 (4.9–5) | 5.6 (5.6–5.7) | 6.5 (6.4–6.5) | 4.5 (4.4–4.5) | 4.9 (4.8–5) | 7.3 (7.2–7.4) | 8.2 (8.1–8.3) |
| 0 | | 4.6 (4.5–4.7) | 5.2 (5.2–5.3) | 5.4 (5.4–5.4) | 6.4 (6.3–6.4) | 4.7 (4.6–4.7) | 5.1 (5–5.1) | 7.2 (7.1–7.3) | 8.1 (8–8.2) |
| 1 | | 5 (4.9–5) | 5.6 (5.5–5.7) | 5.3 (5.2–5.3) | 6.2 (6.2–6.3) | 4.8 (4.8–4.9) | 5.3 (5.2–5.4) | 7.1 (7–7.1) | 8.1 (8–8.2) |
| 2 | | 5.3 (5.3–5.4) | 6 (6–6.1) | 5.1 (5.1–5.2) | 6.1 (6.1–6.2) | 5 (4.9–5.1) | 5.6 (5.5–5.7) | 7 (6.9–7) | 8 (7.9–8.1) |

**eTable 15. Estimates for marginal predictions of annual income and employment among parents exposed to child's mood disorder (F30–F39) and unexposed parents**

| | | Finland | | | | Denmark | | | |
|---|---|---|---|---|---|---|---|---|---|
| | | Men | | Women | | Men | | Women | |
| | | Unexposed | Exposed | Unexposed | Exposed | Unexposed | Exposed | Unexposed | Exposed |
| Years since diagnosis | Outcome | Marg. pred. (95% CI) | Marg. pred. (95% CI) | Marg. pred. (95% CI) | Marg. pred. (95% CI) | Marg. pred. (95% CI) | Marg. pred. (95% CI) | Marg. pred. (95% CI) | Marg. pred. (95% CI) |
| | | Unexposed | Exposed | Unexposed | Exposed | Unexposed | Exposed | Unexposed | Exposed |
| 3 | | 5.8 (5.8–5.9) | 6.5 (6.4–6.5) | 5.1 (5.1–5.2) | 6.1 (6.1–6.2) | 5.3 (5.2–5.3) | 5.9 (5.8–5.9) | 6.9 (6.8–7) | 8.1 (8–8.2) |
| 4 | | 6.4 (6.3–6.4) | 7 (6.9–7) | 5.2 (5.1–5.2) | 6.2 (6.1–6.2) | 5.6 (5.5–5.7) | 6.2 (6.1–6.3) | 7 (6.9–7.1) | 8.2 (8.1–8.3) |
| 5 | | 7 (6.9–7.1) | 7.6 (7.5–7.6) | 5.4 (5.3–5.4) | 6.3 (6.2–6.4) | 6 (5.9–6.1) | 6.5 (6.4–6.6) | 7.1 (7–7.2) | 8.3 (8.2–8.5) |
| -5 | Employment** | 0.8 (0.8–0.8) | 0.77 (0.77–0.77) | 0.78 (0.78–0.78) | 0.74 (0.74–0.75) | 0.75 (0.74–0.75) | 0.74 (0.73–0.74) | 0.71 (0.71–0.72) | 0.7 (0.7–0.7) |
| -4 | | 0.79 (0.79–0.8) | 0.76 (0.76–0.77) | 0.78 (0.78–0.79) | 0.75 (0.75–0.75) | 0.75 (0.74–0.75) | 0.74 (0.73–0.74) | 0.72 (0.71–0.72) | 0.7 (0.7–0.71) |
| -3 | | 0.79 (0.78–0.79) | 0.75 (0.75–0.76) | 0.79 (0.78–0.79) | 0.75 (0.75–0.75) | 0.74 (0.74–0.75) | 0.73 (0.73–0.74) | 0.72 (0.72–0.72) | 0.7 (0.7–0.71) |
| -2 | | 0.78 (0.77–0.78) | 0.75 (0.74–0.75) | 0.79 (0.78–0.79) | 0.75 (0.75–0.75) | 0.74 (0.73–0.74) | 0.72 (0.72–0.73) | 0.72 (0.71–0.72) | 0.7 (0.69–0.7) |
| -1 | | 0.77 (0.77–0.77) | 0.74 (0.73–0.74) | 0.79 (0.79–0.79) | 0.75 (0.75–0.76) | 0.73 (0.73–0.73) | 0.72 (0.71–0.72) | 0.72 (0.71–0.72) | 0.69 (0.69–0.7) |
| 0 | | 0.76 (0.76–0.76) | 0.72 (0.72–0.73) | 0.79 (0.79–0.79) | 0.75 (0.75–0.75) | 0.72 (0.72–0.73) | 0.71 (0.71–0.71) | 0.71 (0.71–0.72) | 0.69 (0.68–0.69) |
| 1 | | 0.74 (0.74–0.75) | 0.71 (0.71–0.71) | 0.79 (0.78–0.79) | 0.74 (0.74–0.75) | 0.71 (0.71–0.72) | 0.7 (0.69–0.7) | 0.7 (0.7–0.71) | 0.68 (0.67–0.68) |

**eTable 15. Estimates for marginal predictions of annual income and employment among parents exposed to child's mood disorder (F30–F39) and unexposed parents**

| | | Finland | | | | Denmark | | | |
|---|---|---|---|---|---|---|---|---|---|
| | | Men | | Women | | Men | | Women | |
| | | Unexposed | Exposed | Unexposed | Exposed | Unexposed | Exposed | Unexposed | Exposed |
| Years since diagnosis | Outcome | Marg. pred. (95% CI) | Marg. pred. (95% CI) | Marg. pred. (95% CI) | Marg. pred. (95% CI) | Marg. pred. (95% CI) | Marg. pred. (95% CI) | Marg. pred. (95% CI) | Marg. pred. (95% CI) |
| | | Unexposed | Exposed | Unexposed | Exposed | Unexposed | Exposed | Unexposed | Exposed |
| 2 | | 0.73 (0.73–0.73) | 0.69 (0.69–0.7) | 0.78 (0.78–0.78) | 0.74 (0.73–0.74) | 0.7 (0.7–0.71) | 0.68 (0.68–0.69) | 0.7 (0.69–0.7) | 0.67 (0.66–0.67) |
| 3 | | 0.71 (0.71–0.71) | 0.67 (0.67–0.68) | 0.77 (0.77–0.77) | 0.73 (0.73–0.73) | 0.69 (0.69–0.7) | 0.67 (0.67–0.68) | 0.69 (0.69–0.69) | 0.65 (0.65–0.66) |
| 4 | | 0.69 (0.69–0.69) | 0.66 (0.65–0.66) | 0.76 (0.76–0.76) | 0.72 (0.72–0.72) | 0.68 (0.68–0.69) | 0.66 (0.65–0.66) | 0.68 (0.68–0.69) | 0.64 (0.64–0.65) |
| 5 | | 0.67 (0.67–0.67) | 0.63 (0.63–0.64) | 0.74 (0.74–0.75) | 0.71 (0.7–0.71) | 0.66 (0.66–0.67) | 0.64 (0.64–0.65) | 0.67 (0.66–0.67) | 0.63 (0.63–0.64) |

Marg. pred. = Marginal prediction, * Thousand euros (€), ** Probability

**eTable 16. Estimates for marginal predictions of annual income and employment among parents exposed to child's anxiety disorder (F40–F48) and unexposed parents**

| | | Finland | | | | Denmark | | | |
|---|---|---|---|---|---|---|---|---|---|
| | | Men | | Women | | Men | | Women | |
| | | Unexposed | Exposed | Unexposed | Exposed | Unexposed | Exposed | Unexposed | Exposed |
| Years since diagnosis | Outcome | Marg. pred. (95% CI) | Marg. pred. (95% CI) | Marg. pred. (95% CI) | Marg. pred. (95% CI) | Marg. pred. (95% CI) | Marg. pred. (95% CI) | Marg. pred. (95% CI) | Marg. pred. (95% CI) |
| | | Unexposed | Exposed | Unexposed | Exposed | Unexposed | Exposed | Unexposed | Exposed |
| -5 | Earnings* | 34.2 (34.1–34.4) | 32.2 (32.1–32.4) | 22.5 (22.4–22.6) | 21.2 (21.1–21.3) | 55.4 (55.2–55.6) | 53.2 (53–53.4) | 42.4 (42.3–42.5) | 41.5 (41.4–41.6) |
| -4 | | 34.5 (34.4–34.7) | 32.5 (32.3–32.6) | 23.2 (23.1–23.3) | 21.9 (21.8–22) | 56 (55.8–56.2) | 53.6 (53.4–53.8) | 42.9 (42.8–43) | 41.9 (41.8–42) |
| -3 | | 34.8 (34.6–34.9) | 32.6 (32.4–32.7) | 23.9 (23.8–24) | 22.4 (22.3–22.5) | 56.4 (56.2–56.7) | 54 (53.8–54.2) | 43.3 (43.2–43.4) | 42.3 (42.2–42.4) |
| -2 | | 34.8 (34.7–35) | 32.6 (32.5–32.8) | 24.4 (24.3–24.5) | 22.9 (22.8–23) | 57 (56.8–57.2) | 54.3 (54.1–54.6) | 43.6 (43.5–43.7) | 42.5 (42.4–42.6) |
| -1 | | 34.9 (34.8–35.1) | 32.6 (32.5–32.8) | 24.9 (24.8–25) | 23.3 (23.2–23.4) | 57.5 (57.3–57.8) | 54.8 (54.6–55) | 44 (43.9–44.1) | 42.8 (42.7–42.9) |
| 0 | | 34.9 (34.7–35.1) | 32.5 (32.4–32.7) | 25.4 (25.3–25.5) | 23.6 (23.5–23.7) | 58 (57.8–58.2) | 55 (54.8–55.2) | 44.3 (44.1–44.4) | 43 (42.9–43.1) |
| 1 | | 34.8 (34.7–35) | 32.4 (32.2–32.5) | 25.7 (25.6–25.8) | 23.9 (23.8–24) | 58.3 (58–58.5) | 55.3 (55.1–55.6) | 44.5 (44.4–44.6) | 43.1 (43–43.3) |
| 2 | | 34.6 (34.4–34.7) | 32.1 (31.9–32.3) | 26 (25.9–26.1) | 24.1 (24–24.3) | 58.6 (58.3–58.8) | 55.5 (55.2–55.7) | 44.6 (44.5–44.8) | 43.2 (43.1–43.3) |
| 3 | | 34.3 (34.2–34.5) | 31.7 (31.6–31.9) | 26.2 (26.1–26.3) | 24.3 (24.2–24.4) | 58.5 (58.3–58.8) | 55.4 (55.2–55.6) | 44.8 (44.7–44.9) | 43.3 (43.1–43.4) |

**eTable 16. Estimates for marginal predictions of annual income and employment among parents exposed to child's anxiety disorder (F40–F48) and unexposed parents**

| | | Finland | | | | Denmark | | | |
|---|---|---|---|---|---|---|---|---|---|
| | | Men | | Women | | Men | | Women | |
| | | Unexposed | Exposed | Unexposed | Exposed | Unexposed | Exposed | Unexposed | Exposed |
| Years since diagnosis | Outcome | Marg. pred. (95% CI) | Marg. pred. (95% CI) | Marg. pred. (95% CI) | Marg. pred. (95% CI) | Marg. pred. (95% CI) | Marg. pred. (95% CI) | Marg. pred. (95% CI) | Marg. pred. (95% CI) |
| | | Unexposed | Exposed | Unexposed | Exposed | Unexposed | Exposed | Unexposed | Exposed |
| 4 | | 33.8 (33.6–34) | 31.2 (31–31.4) | 26.3 (26.2–26.4) | 24.4 (24.2–24.5) | 58.5 (58.3–58.8) | 55.3 (55.1–55.5) | 44.9 (44.8–45) | 43.3 (43.2–43.4) |
| 5 | | 33.2 (33–33.4) | 30.6 (30.4–30.8) | 26.3 (26.1–26.4) | 24.4 (24.3–24.5) | 58.3 (58.1–58.6) | 55.1 (54.9–55.4) | 44.9 (44.8–45.1) | 43.3 (43.2–43.5) |
| -5 | Disposable income* | 28.5 (28.4–28.6) | 27.2 (27.1–27.3) | 23.1 (23–23.2) | 22.7 (22.7–22.8) | 33.9 (33.8–34) | 32.5 (32.4–32.6) | 29.2 (29.2–29.3) | 28.6 (28.5–28.6) |
| -4 | | 28.9 (28.8–29) | 27.6 (27.5–27.7) | 23.5 (23.4–23.5) | 23.1 (23–23.1) | 34.7 (34.6–34.8) | 33.1 (33–33.2) | 29.7 (29.6–29.7) | 28.9 (28.9–29) |
| -3 | | 29.3 (29.2–29.4) | 27.9 (27.8–28) | 23.8 (23.7–23.9) | 23.4 (23.3–23.4) | 35.4 (35.3–35.5) | 33.7 (33.6–33.8) | 30 (30–30.1) | 29.3 (29.2–29.3) |
| -2 | | 29.6 (29.5–29.7) | 28.2 (28.1–28.3) | 24 (24–24.1) | 23.6 (23.6–23.7) | 36.1 (35.9–36.2) | 34.3 (34.2–34.4) | 30.4 (30.3–30.5) | 29.5 (29.5–29.6) |
| -1 | | 29.9 (29.8–30) | 28.5 (28.4–28.6) | 24.2 (24.2–24.3) | 23.8 (23.7–23.8) | 36.8 (36.6–36.9) | 34.9 (34.8–35) | 30.7 (30.7–30.8) | 29.8 (29.8–29.9) |
| 0 | | 30.2 (30.1–30.3) | 28.7 (28.6–28.8) | 24.4 (24.3–24.4) | 23.9 (23.9–24) | 37.4 (37.3–37.6) | 35.4 (35.3–35.5) | 31 (30.9–31.1) | 30 (30–30.1) |
| 1 | | 30.5 (30.4–30.6) | 29 (28.9–29.1) | 24.5 (24.5–24.6) | 24 (24–24.1) | 38 (37.9–38.1) | 36 (35.8–36.1) | 31.3 (31.2–31.4) | 30.2 (30.2–30.3) |

**eTable 16. Estimates for marginal predictions of annual income and employment among parents exposed to child's anxiety disorder (F40–F48) and unexposed parents**

| | | Finland | | | | Denmark | | | |
|---|---|---|---|---|---|---|---|---|---|
| | | Men | | Women | | Men | | Women | |
| | | Unexposed | Exposed | Unexposed | Exposed | Unexposed | Exposed | Unexposed | Exposed |
| Years since diagnosis | Outcome | Marg. pred. (95% CI) | Marg. pred. (95% CI) | Marg. pred. (95% CI) | Marg. pred. (95% CI) | Marg. pred. (95% CI) | Marg. pred. (95% CI) | Marg. pred. (95% CI) | Marg. pred. (95% CI) |
| | | Unexposed | Exposed | Unexposed | Exposed | Unexposed | Exposed | Unexposed | Exposed |
| 2 | | 30.7 (30.6–30.9) | 29.2 (29.1–29.3) | 24.7 (24.6–24.7) | 24.1 (24–24.2) | 38.6 (38.5–38.7) | 36.5 (36.3–36.6) | 31.5 (31.4–31.6) | 30.4 (30.3–30.5) |
| 3 | | 31 (30.9–31.1) | 29.4 (29.3–29.5) | 24.8 (24.7–24.9) | 24.2 (24.1–24.3) | 39 (38.8–39.1) | 36.8 (36.7–37) | 31.7 (31.7–31.8) | 30.6 (30.5–30.6) |
| 4 | | 31.2 (31.1–31.4) | 29.5 (29.4–29.6) | 24.9 (24.9–25) | 24.3 (24.2–24.3) | 39.4 (39.2–39.6) | 37.2 (37.1–37.4) | 32 (31.9–32.1) | 30.7 (30.6–30.8) |
| 5 | | 31.4 (31.2–31.5) | 29.6 (29.5–29.8) | 25.1 (25–25.2) | 24.4 (24.4–24.5) | 39.8 (39.6–40) | 37.6 (37.4–37.7) | 32.2 (32.1–32.3) | 30.9 (30.8–31) |
| -5 | Social welfare benefits* | 3.4 (3.3–3.4) | 3.9 (3.8–3.9) | 6.5 (6.5–6.6) | 7.2 (7.1–7.2) | 4.2 (4.1–4.2) | 4.7 (4.6–4.7) | 9 (8.9–9) | 10.1 (10–10.2) |
| -4 | | 3.6 (3.5–3.6) | 4.1 (4–4.1) | 6.3 (6.3–6.3) | 7 (7–7.1) | 4.3 (4.2–4.3) | 4.8 (4.8–4.9) | 8.7 (8.7–8.8) | 9.9 (9.9–10) |
| -3 | | 3.8 (3.8–3.8) | 4.3 (4.3–4.4) | 6.1 (6.1–6.1) | 6.9 (6.8–6.9) | 4.4 (4.4–4.5) | 5 (4.9–5) | 8.5 (8.5–8.6) | 9.8 (9.7–9.8) |
| -2 | | 4 (4–4.1) | 4.6 (4.5–4.6) | 5.9 (5.9–5.9) | 6.7 (6.7–6.7) | 4.5 (4.5–4.6) | 5.1 (5.1–5.2) | 8.3 (8.3–8.4) | 9.6 (9.6–9.7) |
| -1 | | 4.3 (4.2–4.3) | 4.8 (4.8–4.9) | 5.7 (5.6–5.7) | 6.5 (6.5–6.6) | 4.6 (4.6–4.7) | 5.3 (5.2–5.4) | 8.1 (8.1–8.2) | 9.5 (9.4–9.6) |
| 0 | | 4.5 (4.5–4.6) | 5.1 (5.1–5.2) | 5.5 (5.4–5.5) | 6.4 (6.4–6.5) | 4.8 (4.7–4.8) | 5.5 (5.4–5.6) | 7.9 (7.8–8) | 9.4 (9.4–9.5) |
| 1 | | 4.8 (4.8–4.9) | 5.5 (5.4–5.5) | 5.3 (5.3–5.4) | 6.3 (6.3–6.4) | 4.9 (4.9–5) | 5.7 (5.6–5.7) | 7.7 (7.6–7.8) | 9.3 (9.3–9.4) |
| 2 | | 5.2 (5.1–5.2) | 5.8 (5.8–5.9) | 5.3 (5.2–5.3) | 6.2 (6.2–6.3) | 5.1 (5–5.2) | 5.9 (5.8–6) | 7.5 (7.5–7.6) | 9.2 (9.1–9.3) |

**eTable 16. Estimates for marginal predictions of annual income and employment among parents exposed to child's anxiety disorder (F40–F48) and unexposed parents**

| | | Finland | | | | Denmark | | | |
|---|---|---|---|---|---|---|---|---|---|
| | | Men | | Women | | Men | | Women | |
| | | Unexposed | Exposed | Unexposed | Exposed | Unexposed | Exposed | Unexposed | Exposed |
| Years since diagnosis | Outcome | Marg. pred. (95% CI) | Marg. pred. (95% CI) | Marg. pred. (95% CI) | Marg. pred. (95% CI) | Marg. pred. (95% CI) | Marg. pred. (95% CI) | Marg. pred. (95% CI) | Marg. pred. (95% CI) |
| | | Unexposed | Exposed | Unexposed | Exposed | Unexposed | Exposed | Unexposed | Exposed |
| 3 | | 5.6 (5.5–5.7) | 6.3 (6.2–6.3) | 5.2 (5.2–5.2) | 6.2 (6.1–6.2) | 5.3 (5.2–5.4) | 6.1 (6.1–6.2) | 7.4 (7.3–7.5) | 9.1 (9–9.2) |
| 4 | | 6.1 (6.1–6.2) | 6.7 (6.7–6.8) | 5.2 (5.2–5.3) | 6.2 (6.1–6.2) | 5.6 (5.5–5.7) | 6.4 (6.3–6.5) | 7.4 (7.3–7.5) | 9.1 (9–9.2) |
| 5 | | 6.7 (6.6–6.7) | 7.3 (7.2–7.3) | 5.4 (5.3–5.4) | 6.3 (6.2–6.3) | 5.9 (5.9–6) | 6.7 (6.7–6.8) | 7.4 (7.4–7.5) | 9.2 (9.1–9.3) |
| -5 | Employment** | 0.8 (0.8–0.81) | 0.77 (0.77–0.78) | 0.77 (0.77–0.77) | 0.74 (0.74–0.74) | 0.73 (0.73–0.73) | 0.72 (0.71–0.72) | 0.69 (0.69–0.69) | 0.66 (0.65–0.66) |
| -4 | | 0.8 (0.79–0.8) | 0.76 (0.76–0.77) | 0.78 (0.78–0.78) | 0.74 (0.74–0.75) | 0.74 (0.74–0.74) | 0.72 (0.72–0.73) | 0.7 (0.7–0.71) | 0.67 (0.66–0.67) |
| -3 | | 0.79 (0.79–0.79) | 0.75 (0.75–0.76) | 0.78 (0.78–0.78) | 0.75 (0.74–0.75) | 0.74 (0.73–0.74) | 0.72 (0.72–0.72) | 0.71 (0.7–0.71) | 0.67 (0.67–0.67) |
| -2 | | 0.78 (0.78–0.78) | 0.74 (0.74–0.75) | 0.79 (0.78–0.79) | 0.75 (0.75–0.75) | 0.73 (0.73–0.73) | 0.71 (0.71–0.71) | 0.71 (0.7–0.71) | 0.67 (0.66–0.67) |
| -1 | | 0.77 (0.77–0.77) | 0.74 (0.74–0.74) | 0.79 (0.79–0.79) | 0.75 (0.75–0.75) | 0.72 (0.72–0.73) | 0.7 (0.7–0.71) | 0.7 (0.7–0.71) | 0.66 (0.66–0.67) |
| 0 | | 0.76 (0.76–0.76) | 0.73 (0.72–0.73) | 0.79 (0.79–0.79) | 0.75 (0.74–0.75) | 0.72 (0.71–0.72) | 0.7 (0.69–0.7) | 0.7 (0.7–0.71) | 0.66 (0.65–0.66) |
| 1 | | 0.75 (0.75–0.75) | 0.71 (0.71–0.72) | 0.78 (0.78–0.79) | 0.74 (0.74–0.75) | 0.71 (0.71–0.71) | 0.69 (0.68–0.69) | 0.7 (0.7–0.7) | 0.65 (0.65–0.65) |

**eTable 16. Estimates for marginal predictions of annual income and employment among parents exposed to child's anxiety disorder (F40–F48) and unexposed parents**

| | | Finland | | | | Denmark | | | |
|---|---|---|---|---|---|---|---|---|---|
| | | Men | | Women | | Men | | Women | |
| | | Unexposed | Exposed | Unexposed | Exposed | Unexposed | Exposed | Unexposed | Exposed |
| Years since diagnosis | Outcome | Marg. pred. (95% CI) | Marg. pred. (95% CI) | Marg. pred. (95% CI) | Marg. pred. (95% CI) | Marg. pred. (95% CI) | Marg. pred. (95% CI) | Marg. pred. (95% CI) | Marg. pred. (95% CI) |
| | | Unexposed | Exposed | Unexposed | Exposed | Unexposed | Exposed | Unexposed | Exposed |
| 2 | | 0.74 (0.73–0.74) | 0.7 (0.69–0.7) | 0.78 (0.78–0.78) | 0.74 (0.73–0.74) | 0.7 (0.7–0.7) | 0.67 (0.67–0.68) | 0.69 (0.69–0.7) | 0.64 (0.64–0.65) |
| 3 | | 0.72 (0.72–0.72) | 0.68 (0.68–0.68) | 0.77 (0.77–0.78) | 0.73 (0.73–0.73) | 0.69 (0.69–0.69) | 0.67 (0.66–0.67) | 0.69 (0.69–0.69) | 0.64 (0.63–0.64) |
| 4 | | 0.7 (0.7–0.71) | 0.66 (0.66–0.67) | 0.76 (0.76–0.77) | 0.72 (0.72–0.72) | 0.68 (0.67–0.68) | 0.65 (0.65–0.66) | 0.68 (0.68–0.68) | 0.63 (0.63–0.63) |
| 5 | | 0.68 (0.68–0.69) | 0.64 (0.64–0.65) | 0.75 (0.75–0.76) | 0.71 (0.71–0.71) | 0.66 (0.66–0.67) | 0.64 (0.63–0.64) | 0.67 (0.67–0.67) | 0.62 (0.61–0.62) |

Marg. pred. = Marginal prediction, * Thousand euros (€), ** Probability

**eTable 17. Estimates for marginal predictions of annual income and employment among parents exposed to child's eating disorder (F50) and unexposed parents**

| | | Finland | | | | Denmark | | | |
|---|---|---|---|---|---|---|---|---|---|
| | | Men | | Women | | Men | | Women | |
| | | Unexposed | Exposed | Unexposed | Exposed | Unexposed | Exposed | Unexposed | Exposed |
| Years since diagnosis | Outcome | Marg. pred. (95% CI) | Marg. pred. (95% CI) | Marg. pred. (95% CI) | Marg. pred. (95% CI) | Marg. pred. (95% CI) | Marg. pred. (95% CI) | Marg. pred. (95% CI) | Marg. pred. (95% CI) |
| | | Unexposed | Exposed | Unexposed | Exposed | Unexposed | Exposed | Unexposed | Exposed |
| -5 | Earnings* | 38.3 (37.9–38.7) | 38.9 (38.5–39.3) | 24 (23.8–24.3) | 24.7 (24.4–25) | 60.5 (60–61.1) | 62 (61.4–62.5) | 44.8 (44.5–45.1) | 45.1 (44.8–45.4) |
| -4 | | 38.7 (38.3–39.1) | 39.5 (39.1–39.9) | 24.8 (24.6–25.1) | 25.6 (25.4–25.9) | 60.9 (60.3–61.5) | 62.7 (62.1–63.2) | 45.3 (45.1–45.6) | 45.7 (45.4–46) |
| -3 | | 39.1 (38.7–39.5) | 40.1 (39.7–40.6) | 25.8 (25.5–26) | 26.5 (26.2–26.8) | 61.7 (61.1–62.2) | 63.5 (62.9–64.2) | 45.7 (45.4–46) | 46.3 (46–46.5) |
| -2 | | 39.6 (39.1–40) | 40.4 (39.9–40.8) | 26.5 (26.3–26.8) | 27.3 (27–27.6) | 62.3 (61.7–62.9) | 64 (63.4–64.7) | 46.1 (45.8–46.4) | 46.8 (46.5–47.1) |
| -1 | | 39.8 (39.3–40.2) | 40.4 (40–40.9) | 27.2 (26.9–27.5) | 27.9 (27.7–28.2) | 63 (62.3–63.6) | 64.5 (63.9–65.2) | 46.5 (46.2–46.8) | 47.2 (46.9–47.5) |
| 0 | | 39.9 (39.5–40.4) | 40.4 (39.9–40.9) | 27.7 (27.5–28) | 28.5 (28.2–28.8) | 63.7 (63.1–64.4) | 65.1 (64.5–65.8) | 46.9 (46.6–47.2) | 47.4 (47.1–47.7) |
| 1 | | 40 (39.6–40.5) | 40.5 (40–41) | 28.1 (27.8–28.4) | 28.9 (28.6–29.2) | 64.2 (63.6–64.9) | 65.7 (65–66.3) | 47.1 (46.8–47.4) | 47.7 (47.4–48) |
| 2 | | 39.9 (39.5–40.4) | 40.1 (39.6–40.6) | 28.6 (28.3–28.9) | 29.3 (29–29.6) | 64.2 (63.6–64.9) | 66 (65.3–66.7) | 47.4 (47.1–47.7) | 47.8 (47.5–48.1) |
| 3 | | 39.6 (39.1–40.1) | 39.8 (39.3–40.3) | 29.1 (28.8–29.4) | 29.5 (29.2–29.8) | 64.4 (63.8–65.1) | 65.9 (65.3–66.6) | 47.5 (47.2–47.8) | 47.9 (47.6–48.2) |

**eTable 17. Estimates for marginal predictions of annual income and employment among parents exposed to child's eating disorder (F50) and unexposed parents**

| | | Finland | | | | Denmark | | | |
|---|---|---|---|---|---|---|---|---|---|
| | | Men | | Women | | Men | | Women | |
| | | Unexposed | Exposed | Unexposed | Exposed | Unexposed | Exposed | Unexposed | Exposed |
| Years since diagnosis | Outcome | Marg. pred. (95% CI) | Marg. pred. (95% CI) | Marg. pred. (95% CI) | Marg. pred. (95% CI) | Marg. pred. (95% CI) | Marg. pred. (95% CI) | Marg. pred. (95% CI) | Marg. pred. (95% CI) |
| | | Unexposed | Exposed | Unexposed | Exposed | Unexposed | Exposed | Unexposed | Exposed |
| 4 | | 39.4 (38.9–39.9) | 39.5 (39–40) | 29.2 (28.9–29.5) | 29.8 (29.5–30.2) | 64.5 (63.8–65.2) | 66.2 (65.4–66.9) | 47.7 (47.4–48.1) | 48 (47.6–48.3) |
| 5 | | 38.6 (38.1–39.1) | 38.7 (38.2–39.3) | 29.2 (28.9–29.5) | 29.8 (29.5–30.2) | 64.5 (63.8–65.2) | 65.6 (64.9–66.4) | 47.7 (47.3–48) | 48.1 (47.8–48.5) |
| -5 | Disposable income* | 30.7 (30.5–31) | 31.1 (30.8–31.3) | 24.3 (24.1–24.4) | 24.6 (24.5–24.8) | 36.3 (36–36.6) | 36.9 (36.6–37.2) | 30.4 (30.2–30.5) | 30.6 (30.4–30.7) |
| -4 | | 31.2 (30.9–31.4) | 31.7 (31.4–32) | 24.8 (24.6–24.9) | 25.2 (25–25.3) | 37 (36.7–37.3) | 37.8 (37.5–38.1) | 30.9 (30.7–31.1) | 31.1 (30.9–31.3) |
| -3 | | 31.6 (31.3–31.9) | 32.2 (31.9–32.5) | 25.2 (25.1–25.4) | 25.6 (25.4–25.7) | 37.9 (37.6–38.3) | 38.8 (38.5–39.1) | 31.3 (31.1–31.5) | 31.6 (31.5–31.8) |
| -2 | | 32.2 (31.9–32.5) | 32.8 (32.5–33.1) | 25.6 (25.4–25.7) | 26 (25.8–26.2) | 38.7 (38.4–39) | 39.5 (39.1–39.8) | 31.7 (31.5–31.9) | 32.1 (31.9–32.2) |
| -1 | | 32.7 (32.4–33) | 33.2 (32.9–33.6) | 25.8 (25.7–26) | 26.3 (26.2–26.5) | 39.5 (39.2–39.9) | 40.3 (39.9–40.6) | 32.1 (31.9–32.3) | 32.5 (32.3–32.7) |
| 0 | | 33.1 (32.8–33.4) | 33.5 (33.2–33.8) | 26 (25.8–26.2) | 26.5 (26.4–26.7) | 40.3 (40–40.7) | 40.9 (40.6–41.3) | 32.5 (32.3–32.7) | 32.8 (32.6–33) |
| 1 | | 33.5 (33.2–33.8) | 34 (33.7–34.4) | 26.1 (25.9–26.2) | 26.7 (26.5–26.9) | 41.1 (40.7–41.5) | 41.7 (41.4–42.1) | 32.8 (32.6–33) | 33.1 (32.9–33.3) |

**eTable 17. Estimates for marginal predictions of annual income and employment among parents exposed to child's eating disorder (F50) and unexposed parents**

| | | Finland | | | | Denmark | | | |
|---|---|---|---|---|---|---|---|---|---|
| | | Men | | Women | | Men | | Women | |
| | | Unexposed | Exposed | Unexposed | Exposed | Unexposed | Exposed | Unexposed | Exposed |
| Years since diagnosis | Outcome | Marg. pred. (95% CI) | Marg. pred. (95% CI) | Marg. pred. (95% CI) | Marg. pred. (95% CI) | Marg. pred. (95% CI) | Marg. pred. (95% CI) | Marg. pred. (95% CI) | Marg. pred. (95% CI) |
| | | Unexposed | Exposed | Unexposed | Exposed | Unexposed | Exposed | Unexposed | Exposed |
| 2 | | 33.7 (33.4–34.1) | 34.3 (34–34.7) | 26.3 (26.1–26.4) | 26.8 (26.6–27) | 41.6 (41.2–42) | 42.4 (42–42.8) | 33.1 (32.9–33.3) | 33.3 (33.1–33.5) |
| 3 | | 34 (33.7–34.3) | 34.6 (34.2–34.9) | 26.4 (26.2–26.5) | 26.9 (26.7–27) | 42.2 (41.8–42.6) | 42.9 (42.5–43.3) | 33.3 (33.1–33.5) | 33.5 (33.3–33.7) |
| 4 | | 34.3 (33.9–34.6) | 34.8 (34.5–35.2) | 26.4 (26.3–26.6) | 27 (26.8–27.2) | 42.7 (42.3–43.1) | 43.5 (43.1–44) | 33.7 (33.4–33.9) | 33.7 (33.5–33.9) |
| 5 | | 34.3 (33.9–34.6) | 34.9 (34.5–35.3) | 26.5 (26.3–26.6) | 27.1 (26.9–27.3) | 43.2 (42.7–43.6) | 43.8 (43.4–44.3) | 33.8 (33.6–34) | 34 (33.8–34.3) |
| -5 | Social welfare benefits* | 2.8 (2.7–2.9) | 2.9 (2.8–3) | 6.7 (6.6–6.8) | 6.6 (6.5–6.7) | 3.1 (3–3.2) | 3 (2.9–3.1) | 7.6 (7.5–7.7) | 7.7 (7.6–7.8) |
| -4 | | 3 (2.9–3.1) | 3.1 (3–3.2) | 6.5 (6.4–6.6) | 6.4 (6.3–6.5) | 3.2 (3.1–3.3) | 3.1 (3–3.2) | 7.4 (7.2–7.5) | 7.5 (7.3–7.6) |
| -3 | | 3.2 (3.1–3.3) | 3.2 (3.1–3.4) | 6.3 (6.2–6.4) | 6.2 (6.1–6.3) | 3.3 (3.2–3.4) | 3.2 (3.1–3.3) | 7.1 (7–7.3) | 7.2 (7.1–7.3) |
| -2 | | 3.4 (3.3–3.5) | 3.5 (3.4–3.6) | 6 (5.9–6.1) | 6 (5.9–6.1) | 3.4 (3.3–3.6) | 3.3 (3.2–3.4) | 7 (6.8–7.1) | 7 (6.8–7.1) |
| -1 | | 3.6 (3.5–3.7) | 3.7 (3.6–3.8) | 5.8 (5.7–5.9) | 5.8 (5.7–5.9) | 3.5 (3.4–3.6) | 3.4 (3.3–3.5) | 6.7 (6.6–6.9) | 6.8 (6.6–6.9) |
| 0 | | 3.8 (3.7–4) | 4 (3.9–4.2) | 5.5 (5.4–5.5) | 5.6 (5.5–5.7) | 3.6 (3.5–3.7) | 3.6 (3.5–3.7) | 6.5 (6.4–6.7) | 6.6 (6.5–6.8) |
| 1 | | 4.1 (4–4.2) | 4.3 (4.2–4.4) | 5.2 (5.1–5.3) | 5.4 (5.3–5.5) | 3.8 (3.6–3.9) | 3.8 (3.7–3.9) | 6.4 (6.2–6.5) | 6.5 (6.3–6.6) |
| 2 | | 4.4 (4.3–4.5) | 4.7 (4.6–4.8) | 5 (4.9–5.1) | 5.2 (5.1–5.3) | 3.9 (3.8–4) | 3.9 (3.7–4) | 6.1 (6–6.2) | 6.2 (6.1–6.3) |

**eTable 17. Estimates for marginal predictions of annual income and employment among parents exposed to child's eating disorder (F50) and unexposed parents**

| | | Finland | | | | Denmark | | | |
|---|---|---|---|---|---|---|---|---|---|
| | | Men | | Women | | Men | | Women | |
| | | Unexposed | Exposed | Unexposed | Exposed | Unexposed | Exposed | Unexposed | Exposed |
| Years since diagnosis | Outcome | Marg. pred. (95% CI) | Marg. pred. (95% CI) | Marg. pred. (95% CI) | Marg. pred. (95% CI) | Marg. pred. (95% CI) | Marg. pred. (95% CI) | Marg. pred. (95% CI) | Marg. pred. (95% CI) |
| | | Unexposed | Exposed | Unexposed | Exposed | Unexposed | Exposed | Unexposed | Exposed |
| 3 | | 4.8 (4.6–4.9) | 5.1 (5–5.3) | 4.7 (4.6–4.8) | 5 (4.9–5.2) | 4.1 (4–4.2) | 4 (3.9–4.1) | 6 (5.8–6.1) | 6.1 (6–6.2) |
| 4 | | 5.2 (5.1–5.4) | 5.6 (5.4–5.7) | 4.7 (4.6–4.8) | 4.9 (4.8–5) | 4.3 (4.1–4.4) | 4.3 (4.2–4.4) | 5.8 (5.7–6) | 6 (5.9–6.2) |
| 5 | | 5.9 (5.7–6) | 6.1 (6–6.3) | 4.7 (4.6–4.8) | 5 (4.9–5.1) | 4.5 (4.4–4.7) | 4.6 (4.5–4.8) | 5.8 (5.7–6) | 6 (5.9–6.2) |
| -5 | Employment** | 0.84 (0.83–0.84) | 0.84 (0.83–0.84) | 0.79 (0.79–0.8) | 0.79 (0.79–0.8) | 0.77 (0.77–0.78) | 0.78 (0.77–0.79) | 0.74 (0.74–0.75) | 0.74 (0.73–0.75) |
| -4 | | 0.83 (0.83–0.84) | 0.83 (0.83–0.84) | 0.8 (0.79–0.81) | 0.8 (0.8–0.81) | 0.79 (0.78–0.8) | 0.79 (0.79–0.8) | 0.76 (0.75–0.77) | 0.76 (0.75–0.77) |
| -3 | | 0.83 (0.82–0.83) | 0.83 (0.82–0.83) | 0.81 (0.8–0.81) | 0.81 (0.81–0.82) | 0.79 (0.78–0.79) | 0.8 (0.79–0.8) | 0.76 (0.76–0.77) | 0.77 (0.76–0.78) |
| -2 | | 0.82 (0.82–0.83) | 0.82 (0.81–0.82) | 0.81 (0.8–0.81) | 0.82 (0.81–0.82) | 0.78 (0.77–0.79) | 0.79 (0.78–0.8) | 0.76 (0.76–0.77) | 0.77 (0.76–0.78) |
| -1 | | 0.82 (0.81–0.82) | 0.81 (0.81–0.82) | 0.81 (0.81–0.82) | 0.82 (0.82–0.83) | 0.77 (0.77–0.78) | 0.79 (0.78–0.79) | 0.76 (0.76–0.77) | 0.77 (0.76–0.77) |
| 0 | | 0.81 (0.8–0.81) | 0.8 (0.8–0.81) | 0.82 (0.81–0.82) | 0.82 (0.81–0.82) | 0.77 (0.76–0.78) | 0.78 (0.77–0.78) | 0.76 (0.75–0.77) | 0.76 (0.76–0.77) |
| 1 | | 0.8 (0.79–0.81) | 0.79 (0.79–0.8) | 0.82 (0.81–0.82) | 0.82 (0.81–0.82) | 0.76 (0.76–0.77) | 0.77 (0.76–0.77) | 0.75 (0.75–0.76) | 0.75 (0.75–0.76) |

**eTable 17. Estimates for marginal predictions of annual income and employment among parents exposed to child's eating disorder (F50) and unexposed parents**

| | | Finland | | | | Denmark | | | |
|---|---|---|---|---|---|---|---|---|---|
| | | Men | | Women | | Men | | Women | |
| | | Unexposed | Exposed | Unexposed | Exposed | Unexposed | Exposed | Unexposed | Exposed |
| Years since diagnosis | Outcome | Marg. pred. (95% CI) | Marg. pred. (95% CI) | Marg. pred. (95% CI) | Marg. pred. (95% CI) | Marg. pred. (95% CI) | Marg. pred. (95% CI) | Marg. pred. (95% CI) | Marg. pred. (95% CI) |
| | | Unexposed | Exposed | Unexposed | Exposed | Unexposed | Exposed | Unexposed | Exposed |
| 2 | | 0.78 (0.78–0.79) | 0.78 (0.77–0.78) | 0.81 (0.81–0.82) | 0.81 (0.8–0.81) | 0.75 (0.74–0.75) | 0.76 (0.76–0.77) | 0.75 (0.74–0.76) | 0.75 (0.75–0.76) |
| 3 | | 0.77 (0.76–0.78) | 0.76 (0.76–0.77) | 0.81 (0.8–0.82) | 0.8 (0.8–0.81) | 0.74 (0.73–0.74) | 0.75 (0.74–0.76) | 0.74 (0.74–0.75) | 0.74 (0.74–0.75) |
| 4 | | 0.75 (0.75–0.76) | 0.75 (0.74–0.75) | 0.8 (0.8–0.81) | 0.8 (0.79–0.8) | 0.72 (0.72–0.73) | 0.74 (0.73–0.74) | 0.74 (0.73–0.74) | 0.74 (0.73–0.74) |
| 5 | | 0.74 (0.73–0.74) | 0.73 (0.72–0.74) | 0.79 (0.79–0.8) | 0.79 (0.78–0.8) | 0.71 (0.71–0.72) | 0.72 (0.71–0.73) | 0.73 (0.72–0.73) | 0.72 (0.72–0.73) |

Marg. pred. = Marginal prediction, * Thousand euros (€), ** Probability

**eTable 18. Estimates for marginal predictions of annual income and employment among parents exposed to child's personality disorder (F60) and unexposed parents**

| | | Finland | | | | Denmark | | | |
|---|---|---|---|---|---|---|---|---|---|
| | | Men | | Women | | Men | | Women | |
| | | Unexposed | Exposed | Unexposed | Exposed | Unexposed | Exposed | Unexposed | Exposed |
| Years since diagnosis | Outcome | Marg. pred. (95% CI) | Marg. pred. (95% CI) | Marg. pred. (95% CI) | Marg. pred. (95% CI) | Marg. pred. (95% CI) | Marg. pred. (95% CI) | Marg. pred. (95% CI) | Marg. pred. (95% CI) |
| | | Unexposed | Exposed | Unexposed | Exposed | Unexposed | Exposed | Unexposed | Exposed |
| -5 | Earnings* | 31.8 (31.4–32.2) | 29.4 (29.1–29.8) | 22.2 (22–22.4) | 20.5 (20.2–20.7) | 53.4 (53.1–53.8) | 50.7 (50.3–51) | 40.8 (40.6–41) | 39.6 (39.5–39.8) |
| -4 | | 31.8 (31.4–32.2) | 29.3 (28.9–29.6) | 22.7 (22.5–22.9) | 20.8 (20.6–21) | 53.6 (53.2–54) | 50.9 (50.6–51.3) | 41.1 (41–41.3) | 39.7 (39.6–39.9) |
| -3 | | 31.7 (31.3–32.1) | 29 (28.6–29.4) | 23.1 (22.8–23.3) | 21.1 (20.8–21.3) | 53.9 (53.5–54.3) | 51.1 (50.7–51.4) | 41.3 (41.1–41.5) | 40 (39.8–40.1) |
| -2 | | 31.4 (31–31.8) | 28.8 (28.4–29.2) | 23.3 (23–23.5) | 21.3 (21–21.5) | 54.3 (53.9–54.6) | 51.1 (50.7–51.4) | 41.5 (41.3–41.7) | 40.1 (39.9–40.3) |
| -1 | | 31.5 (31.1–32) | 28.6 (28.2–29) | 23.5 (23.2–23.7) | 21.4 (21.2–21.7) | 54.3 (53.9–54.7) | 51.2 (50.8–51.5) | 41.5 (41.3–41.7) | 40.1 (39.9–40.3) |
| 0 | | 31.2 (30.8–31.6) | 28.1 (27.7–28.5) | 23.6 (23.3–23.8) | 21.4 (21.1–21.6) | 54.3 (53.9–54.7) | 51.3 (50.9–51.6) | 41.6 (41.4–41.8) | 40 (39.8–40.2) |
| 1 | | 30.7 (30.3–31.1) | 27.7 (27.3–28.2) | 23.6 (23.3–23.9) | 21.4 (21.2–21.7) | 54.3 (53.9–54.7) | 51.3 (50.9–51.7) | 41.6 (41.4–41.8) | 40.1 (39.9–40.3) |
| 2 | | 30.1 (29.7–30.6) | 27.3 (26.8–27.7) | 23.5 (23.2–23.7) | 21.4 (21.1–21.7) | 54.5 (54.1–54.9) | 51.2 (50.9–51.6) | 41.7 (41.5–41.9) | 40.1 (39.9–40.3) |
| 3 | | 29.4 (29–29.9) | 26.6 (26.2–27.1) | 23.3 (23–23.6) | 21.3 (21–21.5) | 54.5 (54.1–54.9) | 51.2 (50.8–51.5) | 41.7 (41.5–41.9) | 40.1 (39.9–40.3) |

**eTable 18. Estimates for marginal predictions of annual income and employment among parents exposed to child's personality disorder (F60) and unexposed parents**

| | | Finland | | | | Denmark | | | |
|---|---|---|---|---|---|---|---|---|---|
| | | Men | | Women | | Men | | Women | |
| | | Unexposed | Exposed | Unexposed | Exposed | Unexposed | Exposed | Unexposed | Exposed |
| Years since diagnosis | Outcome | Marg. pred. (95% CI) Unexposed | Marg. pred. (95% CI) Exposed | Marg. pred. (95% CI) Unexposed | Marg. pred. (95% CI) Exposed | Marg. pred. (95% CI) Unexposed | Marg. pred. (95% CI) Exposed | Marg. pred. (95% CI) Unexposed | Marg. pred. (95% CI) Exposed |
| 4 | | 28.5 (28.1–29) | 25.9 (25.4–26.4) | 23.1 (22.8–23.4) | 21.1 (20.8–21.4) | 54.3 (53.9–54.8) | 50.9 (50.5–51.3) | 41.8 (41.6–42) | 40.2 (39.9–40.4) |
| 5 | | 27.5 (27–28) | 25 (24.5–25.5) | 22.7 (22.4–23) | 20.8 (20.5–21.1) | 54.2 (53.8–54.6) | 50.5 (50.1–50.9) | 41.8 (41.6–42) | 40.2 (39.9–40.4) |
| -5 | Disposable income* | 27.1 (26.9–27.4) | 25.6 (25.4–25.9) | 22.3 (22.1–22.4) | 21.8 (21.6–21.9) | 32.5 (32.3–32.7) | 30.6 (30.5–30.8) | 27.8 (27.7–27.9) | 27 (26.9–27.1) |
| -4 | | 27.4 (27.2–27.7) | 25.8 (25.6–26.1) | 22.4 (22.3–22.6) | 21.8 (21.7–21.9) | 32.9 (32.7–33.1) | 31.2 (31–31.3) | 28.1 (28–28.2) | 27.1 (27–27.2) |
| -3 | | 27.7 (27.4–28) | 26 (25.7–26.2) | 22.5 (22.4–22.6) | 21.9 (21.7–22) | 33.5 (33.3–33.7) | 31.6 (31.4–31.8) | 28.3 (28.2–28.4) | 27.3 (27.2–27.4) |
| -2 | | 27.8 (27.5–28.1) | 26.2 (25.9–26.4) | 22.5 (22.4–22.6) | 21.9 (21.7–22) | 34.1 (33.9–34.3) | 32 (31.8–32.2) | 28.5 (28.3–28.6) | 27.4 (27.3–27.6) |
| -1 | | 28.2 (27.9–28.5) | 26.4 (26.1–26.6) | 22.5 (22.4–22.7) | 21.8 (21.6–21.9) | 34.5 (34.3–34.7) | 32.4 (32.2–32.6) | 28.6 (28.5–28.7) | 27.5 (27.4–27.6) |
| 0 | | 28.4 (28.1–28.6) | 26.5 (26.2–26.7) | 22.6 (22.4–22.7) | 21.8 (21.7–21.9) | 34.9 (34.6–35.1) | 32.8 (32.6–33) | 28.7 (28.6–28.8) | 27.5 (27.4–27.6) |
| 1 | | 28.5 (28.2–28.7) | 26.6 (26.3–26.9) | 22.7 (22.5–22.8) | 21.9 (21.7–22) | 35.2 (35–35.5) | 33.2 (33–33.5) | 28.8 (28.7–29) | 27.6 (27.5–27.8) |

**eTable 18. Estimates for marginal predictions of annual income and employment among parents exposed to child's personality disorder (F60) and unexposed parents**

| | | Finland | | | | Denmark | | | |
|---|---|---|---|---|---|---|---|---|---|
| | | Men | | Women | | Men | | Women | |
| | | Unexposed | Exposed | Unexposed | Exposed | Unexposed | Exposed | Unexposed | Exposed |
| Years since diagnosis | Outcome | Marg. pred. (95% CI) | Marg. pred. (95% CI) | Marg. pred. (95% CI) | Marg. pred. (95% CI) | Marg. pred. (95% CI) | Marg. pred. (95% CI) | Marg. pred. (95% CI) | Marg. pred. (95% CI) |
| | | Unexposed | Exposed | Unexposed | Exposed | Unexposed | Exposed | Unexposed | Exposed |
| 2 | | 28.6 (28.3–28.9) | 26.8 (26.5–27) | 22.7 (22.5–22.8) | 22 (21.9–22.2) | 35.7 (35.5–36) | 33.6 (33.4–33.8) | 29 (28.9–29.2) | 27.8 (27.6–27.9) |
| 3 | | 28.8 (28.5–29.1) | 26.9 (26.6–27.2) | 22.8 (22.7–23) | 22.1 (22–22.3) | 36.1 (35.9–36.4) | 33.8 (33.6–34.1) | 29.2 (29–29.3) | 27.9 (27.8–28) |
| 4 | | 28.8 (28.5–29.1) | 26.9 (26.7–27.2) | 22.9 (22.8–23.1) | 22.3 (22.1–22.4) | 36.5 (36.2–36.7) | 34.1 (33.9–34.3) | 29.4 (29.3–29.6) | 28.1 (28–28.2) |
| 5 | | 28.9 (28.5–29.2) | 27 (26.7–27.3) | 23.1 (22.9–23.2) | 22.4 (22.3–22.6) | 36.8 (36.5–37.1) | 34.2 (34–34.5) | 29.6 (29.5–29.8) | 28.3 (28.2–28.4) |
| -5 | Social welfare benefits* | 4 (3.9–4.1) | 4.6 (4.5–4.8) | 6 (5.9–6.1) | 7 (6.8–7.1) | 4.6 (4.5–4.7) | 5.6 (5.5–5.7) | 8.7 (8.6–8.8) | 10.4 (10.3–10.5) |
| -4 | | 4.2 (4.1–4.3) | 4.9 (4.8–5.1) | 5.7 (5.6–5.8) | 6.7 (6.6–6.8) | 4.7 (4.6–4.8) | 5.7 (5.6–5.8) | 8.5 (8.4–8.6) | 10.2 (10.1–10.3) |
| -3 | | 4.6 (4.4–4.7) | 5.2 (5.1–5.3) | 5.5 (5.4–5.6) | 6.4 (6.3–6.6) | 4.8 (4.7–4.9) | 5.9 (5.8–6) | 8.2 (8.1–8.3) | 10 (9.9–10.1) |
| -2 | | 4.8 (4.7–4.9) | 5.5 (5.4–5.6) | 5.2 (5.1–5.3) | 6.2 (6.1–6.3) | 5 (4.9–5.1) | 6.1 (6–6.2) | 8 (7.9–8.1) | 9.9 (9.8–10) |
| -1 | | 5.1 (5–5.2) | 5.9 (5.7–6) | 5.1 (5–5.2) | 6 (5.9–6.1) | 5.2 (5.1–5.3) | 6.3 (6.1–6.4) | 7.8 (7.7–7.9) | 9.6 (9.5–9.8) |
| 0 | | 5.4 (5.3–5.6) | 6.3 (6.1–6.4) | 5 (4.9–5.1) | 6 (5.9–6.1) | 5.3 (5.2–5.4) | 6.4 (6.3–6.6) | 7.7 (7.5–7.8) | 9.5 (9.4–9.6) |
| 1 | | 5.9 (5.7–6) | 6.7 (6.6–6.9) | 5.1 (5–5.2) | 6.1 (5.9–6.2) | 5.5 (5.4–5.6) | 6.6 (6.5–6.8) | 7.5 (7.4–7.6) | 9.5 (9.4–9.6) |

**eTable 18. Estimates for marginal predictions of annual income and employment among parents exposed to child's personality disorder (F60) and unexposed parents**

| | | Finland | | | | Denmark | | | |
|---|---|---|---|---|---|---|---|---|---|
| | | Men | | Women | | Men | | Women | |
| | | Unexposed | Exposed | Unexposed | Exposed | Unexposed | Exposed | Unexposed | Exposed |
| Years since diagnosis | Outcome | Marg. pred. (95% CI) | Marg. pred. (95% CI) | Marg. pred. (95% CI) | Marg. pred. (95% CI) | Marg. pred. (95% CI) | Marg. pred. (95% CI) | Marg. pred. (95% CI) | Marg. pred. (95% CI) |
| | | Unexposed | Exposed | Unexposed | Exposed | Unexposed | Exposed | Unexposed | Exposed |
| 2 | | 6.5 (6.3–6.6) | 7.2 (7.1–7.4) | 5.2 (5.1–5.3) | 6.2 (6.1–6.3) | 5.7 (5.6–5.8) | 6.9 (6.8–7) | 7.5 (7.4–7.6) | 9.5 (9.3–9.6) |
| 3 | | 7.1 (6.9–7.3) | 7.8 (7.6–7.9) | 5.4 (5.3–5.5) | 6.4 (6.3–6.5) | 6 (5.9–6.2) | 7.2 (7.1–7.4) | 7.5 (7.4–7.6) | 9.5 (9.4–9.7) |
| 4 | | 7.8 (7.6–8) | 8.4 (8.2–8.6) | 5.6 (5.5–5.8) | 6.6 (6.5–6.8) | 6.4 (6.3–6.6) | 7.6 (7.4–7.7) | 7.5 (7.4–7.7) | 9.7 (9.6–9.8) |
| 5 | | 8.7 (8.5–8.9) | 9.2 (9–9.4) | 6 (5.9–6.2) | 7 (6.9–7.1) | 6.8 (6.7–7) | 8 (7.9–8.1) | 7.8 (7.6–7.9) | 9.9 (9.8–10.1) |
| -5 | Employment** | 0.77 (0.76–0.77) | 0.72 (0.72–0.73) | 0.77 (0.77–0.78) | 0.72 (0.71–0.72) | 0.7 (0.7–0.71) | 0.67 (0.66–0.67) | 0.66 (0.65–0.67) | 0.61 (0.6–0.61) |
| -4 | | 0.75 (0.75–0.76) | 0.71 (0.7–0.71) | 0.77 (0.77–0.78) | 0.72 (0.71–0.72) | 0.72 (0.72–0.73) | 0.69 (0.68–0.69) | 0.68 (0.68–0.69) | 0.62 (0.62–0.63) |
| -3 | | 0.74 (0.74–0.75) | 0.69 (0.69–0.7) | 0.77 (0.76–0.78) | 0.71 (0.7–0.72) | 0.72 (0.72–0.73) | 0.69 (0.68–0.69) | 0.69 (0.68–0.69) | 0.63 (0.62–0.64) |
| -2 | | 0.73 (0.72–0.74) | 0.68 (0.67–0.69) | 0.76 (0.76–0.77) | 0.71 (0.7–0.71) | 0.72 (0.71–0.72) | 0.68 (0.67–0.68) | 0.69 (0.68–0.69) | 0.63 (0.62–0.63) |
| -1 | | 0.72 (0.71–0.73) | 0.67 (0.66–0.67) | 0.76 (0.75–0.76) | 0.7 (0.69–0.71) | 0.71 (0.7–0.71) | 0.67 (0.67–0.68) | 0.68 (0.68–0.69) | 0.62 (0.61–0.63) |
| 0 | | 0.71 (0.7–0.72) | 0.65 (0.64–0.66) | 0.75 (0.74–0.76) | 0.69 (0.69–0.7) | 0.7 (0.69–0.7) | 0.66 (0.65–0.66) | 0.67 (0.67–0.68) | 0.62 (0.61–0.62) |

**eTable 18. Estimates for marginal predictions of annual income and employment among parents exposed to child's personality disorder (F60) and unexposed parents**

| | | Finland | | | | Denmark | | | |
|---|---|---|---|---|---|---|---|---|---|
| | | Men | | Women | | Men | | Women | |
| | | Unexposed | Exposed | Unexposed | Exposed | Unexposed | Exposed | Unexposed | Exposed |
| Years since diagnosis | Outcome | Marg. pred. (95% CI) | Marg. pred. (95% CI) | Marg. pred. (95% CI) | Marg. pred. (95% CI) | Marg. pred. (95% CI) | Marg. pred. (95% CI) | Marg. pred. (95% CI) | Marg. pred. (95% CI) |
| | | Unexposed | Exposed | Unexposed | Exposed | Unexposed | Exposed | Unexposed | Exposed |
| 1 | | 0.69 (0.68–0.7) | 0.63 (0.62–0.64) | 0.75 (0.74–0.75) | 0.68 (0.68–0.69) | 0.68 (0.68–0.69) | 0.65 (0.64–0.65) | 0.67 (0.66–0.67) | 0.6 (0.6–0.61) |
| 2 | | 0.67 (0.66–0.67) | 0.62 (0.61–0.62) | 0.73 (0.73–0.74) | 0.67 (0.66–0.68) | 0.67 (0.67–0.68) | 0.63 (0.63–0.64) | 0.66 (0.65–0.66) | 0.6 (0.59–0.6) |
| 3 | | 0.64 (0.64–0.65) | 0.59 (0.59–0.6) | 0.72 (0.71–0.73) | 0.66 (0.65–0.67) | 0.66 (0.65–0.67) | 0.62 (0.61–0.63) | 0.65 (0.64–0.66) | 0.59 (0.58–0.59) |
| 4 | | 0.62 (0.61–0.63) | 0.57 (0.56–0.58) | 0.7 (0.7–0.71) | 0.65 (0.64–0.66) | 0.64 (0.63–0.65) | 0.6 (0.59–0.61) | 0.64 (0.63–0.65) | 0.57 (0.57–0.58) |
| 5 | | 0.59 (0.58–0.6) | 0.55 (0.54–0.56) | 0.68 (0.67–0.69) | 0.63 (0.63–0.64) | 0.62 (0.61–0.63) | 0.58 (0.57–0.59) | 0.63 (0.62–0.63) | 0.56 (0.55–0.57) |

Marg. pred. = Marginal prediction, * Thousand euros (€), ** Probability

**eTable 19. Estimates for marginal predictions of annual income and employment among parents exposed to child's intellectual disability (F70–F79) and unexposed parents**

| | | Finland | | | | Denmark | | | |
|---|---|---|---|---|---|---|---|---|---|
| | | Men | | Women | | Men | | Women | |
| | | Unexposed | Exposed | Unexposed | Exposed | Unexposed | Exposed | Unexposed | Exposed |
| Years since diagnosis | Outcome | Marg. pred. (95% CI) | Marg. pred. (95% CI) | Marg. pred. (95% CI) | Marg. pred. (95% CI) | Marg. pred. (95% CI) | Marg. pred. (95% CI) | Marg. pred. (95% CI) | Marg. pred. (95% CI) |
| | | Unexposed | Exposed | Unexposed | Exposed | Unexposed | Exposed | Unexposed | Exposed |
| -5 | Earnings* | 28.7 (28.3–29) | 25.7 (25.4–26.1) | 14 (13.7–14.2) | 12.1 (11.9–12.4) | 48.1 (47.6–48.5) | 44.3 (43.9–44.7) | 37.6 (37.3–37.8) | 34.9 (34.7–35.1) |
| -4 | | 29.6 (29.3–30) | 26.4 (26–26.8) | 14.6 (14.4–14.9) | 12.4 (12.2–12.6) | 48.8 (48.4–49.3) | 44.6 (44.2–45) | 38.3 (38.1–38.6) | 35.6 (35.4–35.8) |
| -3 | | 30.3 (29.9–30.7) | 27 (26.6–27.3) | 15.4 (15.2–15.7) | 13 (12.8–13.3) | 49.6 (49.2–50.1) | 45.2 (44.8–45.6) | 39.1 (38.9–39.4) | 36.2 (35.9–36.4) |
| -2 | | 30.9 (30.5–31.3) | 27.3 (26.9–27.7) | 16.4 (16.2–16.7) | 13.7 (13.4–13.9) | 50.4 (49.9–50.8) | 45.8 (45.4–46.2) | 39.7 (39.5–40) | 36.7 (36.5–37) |
| -1 | | 31.7 (31.3–32) | 27.8 (27.4–28.1) | 17.5 (17.2–17.7) | 14.5 (14.2–14.7) | 51 (50.5–51.5) | 46.3 (45.8–46.7) | 40.4 (40.1–40.6) | 37.3 (37.1–37.6) |
| 0 | | 32.3 (31.9–32.7) | 28.3 (27.9–28.7) | 18.6 (18.4–18.9) | 15.6 (15.3–15.9) | 51.6 (51.1–52.1) | 46.7 (46.2–47.1) | 40.9 (40.6–41.1) | 37.7 (37.5–37.9) |
| 1 | | 33 (32.5–33.4) | 28.6 (28.2–29.1) | 19.7 (19.4–20) | 16.8 (16.5–17.1) | 52 (51.5–52.5) | 46.8 (46.4–47.3) | 41.4 (41.1–41.7) | 38.1 (37.8–38.4) |
| 2 | | 33.4 (33–33.8) | 29.1 (28.7–29.6) | 20.8 (20.5–21.1) | 17.6 (17.3–17.9) | 52.7 (52.1–53.2) | 47.2 (46.8–47.7) | 41.7 (41.4–42) | 38.4 (38.1–38.6) |
| 3 | | 33.9 (33.5–34.4) | 29.4 (29–29.8) | 21.7 (21.4–22) | 18.3 (18–18.6) | 53.1 (52.6–53.7) | 47.5 (47–48) | 42.1 (41.8–42.4) | 38.6 (38.4–38.9) |

**eTable 19. Estimates for marginal predictions of annual income and employment among parents exposed to child's intellectual disability (F70–F79) and unexposed parents**

| | | Finland | | | | Denmark | | | |
|---|---|---|---|---|---|---|---|---|---|
| | | Men | | Women | | Men | | Women | |
| | | Unexposed | Exposed | Unexposed | Exposed | Unexposed | Exposed | Unexposed | Exposed |
| Years since diagnosis | Outcome | Marg. pred. (95% CI) | Marg. pred. (95% CI) | Marg. pred. (95% CI) | Marg. pred. (95% CI) | Marg. pred. (95% CI) | Marg. pred. (95% CI) | Marg. pred. (95% CI) | Marg. pred. (95% CI) |
| | | Unexposed | Exposed | Unexposed | Exposed | Unexposed | Exposed | Unexposed | Exposed |
| 4 | | 34.3 (33.8–34.7) | 29.7 (29.2–30.1) | 22.7 (22.4–23) | 18.9 (18.6–19.2) | 53.5 (53–54.1) | 47.9 (47.4–48.4) | 42.4 (42.1–42.7) | 38.9 (38.6–39.2) |
| 5 | | 34.5 (34–35) | 29.6 (29.2–30.1) | 23.4 (23.1–23.7) | 19.6 (19.3–19.9) | 53.7 (53.2–54.3) | 48.2 (47.7–48.7) | 42.7 (42.4–43) | 38.9 (38.7–39.2) |
| -5 | Disposable income* | 24 (23.8–24.3) | 22.8 (22.5–23) | 18.8 (18.6–18.9) | 17.9 (17.7–18) | 29.4 (29.2–29.7) | 27.4 (27.2–27.7) | 26.2 (26–26.3) | 24.7 (24.5–24.8) |
| -4 | | 24.9 (24.7–25.1) | 23.5 (23.3–23.7) | 19.6 (19.4–19.7) | 18.5 (18.3–18.6) | 30.2 (29.9–30.4) | 27.9 (27.7–28.1) | 26.8 (26.7–27) | 25.3 (25.1–25.4) |
| -3 | | 25.5 (25.3–25.8) | 24 (23.8–24.3) | 20.3 (20.2–20.5) | 19.1 (19–19.3) | 30.9 (30.7–31.2) | 28.5 (28.3–28.8) | 27.5 (27.4–27.7) | 25.7 (25.6–25.9) |
| -2 | | 26.1 (25.9–26.4) | 24.5 (24.3–24.7) | 21.1 (21–21.3) | 19.8 (19.7–20) | 31.6 (31.4–31.9) | 29.1 (28.9–29.3) | 28.1 (27.9–28.2) | 26.2 (26.1–26.4) |
| -1 | | 26.7 (26.4–26.9) | 25 (24.8–25.2) | 21.8 (21.7–22) | 20.4 (20.3–20.6) | 32.3 (32–32.6) | 29.6 (29.4–29.8) | 28.6 (28.4–28.8) | 26.6 (26.5–26.8) |
| 0 | | 27.2 (27–27.5) | 25.4 (25.2–25.6) | 22.5 (22.3–22.6) | 21.1 (20.9–21.2) | 32.9 (32.6–33.2) | 30.1 (29.9–30.4) | 29 (28.9–29.2) | 27 (26.8–27.1) |
| 1 | | 27.8 (27.5–28.1) | 25.8 (25.5–26) | 23.1 (23–23.3) | 21.8 (21.6–21.9) | 33.5 (33.2–33.8) | 30.5 (30.3–30.8) | 29.5 (29.3–29.7) | 27.3 (27.2–27.5) |

**eTable 19. Estimates for marginal predictions of annual income and employment among parents exposed to child's intellectual disability (F70–F79) and unexposed parents**

| | | Finland | | | | Denmark | | | |
|---|---|---|---|---|---|---|---|---|---|
| | | Men | | Women | | Men | | Women | |
| | | Unexposed | Exposed | Unexposed | Exposed | Unexposed | Exposed | Unexposed | Exposed |
| Years since diagnosis | Outcome | Marg. pred. (95% CI) | Marg. pred. (95% CI) | Marg. pred. (95% CI) | Marg. pred. (95% CI) | Marg. pred. (95% CI) | Marg. pred. (95% CI) | Marg. pred. (95% CI) | Marg. pred. (95% CI) |
| | | Unexposed | Exposed | Unexposed | Exposed | Unexposed | Exposed | Unexposed | Exposed |
| 2 | | 28.3 (28.1–28.6) | 26.3 (26–26.5) | 23.7 (23.6–23.9) | 22.2 (22.1–22.4) | 34.3 (34–34.6) | 31 (30.8–31.3) | 29.9 (29.7–30) | 27.6 (27.4–27.8) |
| 3 | | 28.9 (28.7–29.2) | 26.6 (26.3–26.9) | 24.3 (24.1–24.5) | 22.7 (22.5–22.9) | 34.9 (34.6–35.2) | 31.5 (31.2–31.8) | 30.2 (30–30.4) | 27.8 (27.7–28) |
| 4 | | 29.5 (29.2–29.8) | 27 (26.8–27.3) | 24.8 (24.7–25) | 23.1 (22.9–23.3) | 35.5 (35.2–35.9) | 32 (31.7–32.3) | 30.5 (30.3–30.7) | 28.1 (27.9–28.3) |
| 5 | | 29.9 (29.6–30.2) | 27.3 (27–27.6) | 25.3 (25.1–25.5) | 23.5 (23.3–23.6) | 36 (35.7–36.4) | 32.5 (32.2–32.8) | 30.8 (30.6–31) | 28.2 (28–28.4) |
| -5 | Social welfare benefits* | 3 (2.9–3.1) | 4 (3.8–4.1) | 9 (8.9–9.1) | 9.5 (9.4–9.7) | 4.9 (4.8–5.1) | 6.4 (6.2–6.5) | 11.7 (11.5–11.9) | 13.6 (13.4–13.8) |
| -4 | | 3.2 (3.1–3.3) | 4.2 (4–4.3) | 9.2 (9–9.3) | 9.8 (9.7–10) | 5 (4.8–5.1) | 6.5 (6.4–6.7) | 11.8 (11.6–11.9) | 13.7 (13.5–13.9) |
| -3 | | 3.3 (3.2–3.4) | 4.3 (4.2–4.4) | 9.2 (9.1–9.3) | 9.9 (9.8–10.1) | 5.1 (5–5.2) | 6.7 (6.5–6.8) | 11.7 (11.5–11.9) | 13.8 (13.6–14) |
| -2 | | 3.4 (3.3–3.5) | 4.4 (4.3–4.6) | 9.1 (9–9.2) | 10 (9.9–10.2) | 5.1 (4.9–5.2) | 6.8 (6.7–7) | 11.5 (11.3–11.6) | 13.6 (13.4–13.8) |
| -1 | | 3.5 (3.4–3.6) | 4.6 (4.5–4.7) | 8.9 (8.8–9) | 9.9 (9.7–10) | 5.2 (5.1–5.4) | 6.9 (6.7–7.1) | 11.2 (11.1–11.4) | 13.5 (13.3–13.7) |
| 0 | | 3.6 (3.4–3.7) | 4.7 (4.6–4.9) | 8.6 (8.5–8.7) | 9.6 (9.5–9.7) | 5.3 (5.1–5.5) | 7.1 (6.9–7.3) | 11 (10.8–11.1) | 13.4 (13.2–13.6) |

**eTable 19. Estimates for marginal predictions of annual income and employment among parents exposed to child's intellectual disability (F70–F79) and unexposed parents**

| | | Finland | | | | Denmark | | | |
|---|---|---|---|---|---|---|---|---|---|
| | | Men | | Women | | Men | | Women | |
| | | Unexposed | Exposed | Unexposed | Exposed | Unexposed | Exposed | Unexposed | Exposed |
| Years since diagnosis | Outcome | Marg. pred. (95% CI) | Marg. pred. (95% CI) | Marg. pred. (95% CI) | Marg. pred. (95% CI) | Marg. pred. (95% CI) | Marg. pred. (95% CI) | Marg. pred. (95% CI) | Marg. pred. (95% CI) |
| | | Unexposed | Exposed | Unexposed | Exposed | Unexposed | Exposed | Unexposed | Exposed |
| 1 | | 3.7 (3.6–3.8) | 4.8 (4.7–5) | 8.5 (8.3–8.6) | 9.4 (9.2–9.5) | 5.3 (5.2–5.5) | 7.3 (7.1–7.4) | 10.7 (10.5–10.9) | 13.2 (13–13.4) |
| 2 | | 3.8 (3.7–3.9) | 4.9 (4.8–5.1) | 8.2 (8–8.3) | 9.2 (9–9.3) | 5.5 (5.4–5.7) | 7.5 (7.3–7.7) | 10.5 (10.3–10.7) | 13 (12.8–13.2) |
| 3 | | 4 (3.8–4.1) | 5.1 (4.9–5.2) | 8 (7.8–8.1) | 9.1 (8.9–9.2) | 5.7 (5.5–5.9) | 7.6 (7.4–7.8) | 10.3 (10.1–10.5) | 12.8 (12.6–13) |
| 4 | | 4.2 (4.1–4.4) | 5.2 (5.1–5.4) | 7.7 (7.6–7.8) | 8.9 (8.8–9.1) | 5.9 (5.7–6) | 7.9 (7.7–8.1) | 10.1 (9.9–10.3) | 12.7 (12.5–12.9) |
| 5 | | 4.4 (4.2–4.5) | 5.5 (5.3–5.6) | 7.6 (7.5–7.7) | 8.8 (8.6–9) | 6 (5.8–6.2) | 8.1 (7.9–8.3) | 10 (9.8–10.2) | 12.6 (12.4–12.9) |
| -5 | Employment** | 0.79 (0.79–0.8) | 0.74 (0.73–0.74) | 0.62 (0.61–0.62) | 0.55 (0.54–0.56) | 0.68 (0.67–0.69) | 0.63 (0.62–0.64) | 0.58 (0.57–0.59) | 0.5 (0.49–0.51) |
| -4 | | 0.8 (0.79–0.8) | 0.73 (0.73–0.74) | 0.63 (0.62–0.64) | 0.55 (0.54–0.55) | 0.7 (0.69–0.7) | 0.64 (0.63–0.65) | 0.6 (0.59–0.61) | 0.51 (0.5–0.52) |
| -3 | | 0.79 (0.79–0.8) | 0.73 (0.72–0.73) | 0.64 (0.63–0.65) | 0.55 (0.55–0.56) | 0.7 (0.69–0.71) | 0.65 (0.64–0.65) | 0.61 (0.6–0.61) | 0.52 (0.51–0.52) |
| -2 | | 0.79 (0.79–0.8) | 0.72 (0.72–0.73) | 0.66 (0.65–0.67) | 0.56 (0.56–0.57) | 0.7 (0.69–0.71) | 0.64 (0.63–0.65) | 0.62 (0.61–0.63) | 0.53 (0.52–0.53) |
| -1 | | 0.8 (0.79–0.8) | 0.72 (0.72–0.73) | 0.67 (0.67–0.68) | 0.57 (0.56–0.58) | 0.7 (0.69–0.7) | 0.64 (0.63–0.65) | 0.63 (0.62–0.64) | 0.53 (0.52–0.54) |

**eTable 19. Estimates for marginal predictions of annual income and employment among parents exposed to child's intellectual disability (F70–F79) and unexposed parents**

| | | Finland | | | | Denmark | | | |
|---|---|---|---|---|---|---|---|---|---|
| | | Men | | Women | | Men | | Women | |
| | | Unexposed | Exposed | Unexposed | Exposed | Unexposed | Exposed | Unexposed | Exposed |
| Years since diagnosis | Outcome | Marg. pred. (95% CI) | Marg. pred. (95% CI) | Marg. pred. (95% CI) | Marg. pred. (95% CI) | Marg. pred. (95% CI) | Marg. pred. (95% CI) | Marg. pred. (95% CI) | Marg. pred. (95% CI) |
| | | Unexposed | Exposed | Unexposed | Exposed | Unexposed | Exposed | Unexposed | Exposed |
| 0 | | 0.8 (0.79–0.8) | 0.72 (0.72–0.73) | 0.69 (0.69–0.7) | 0.59 (0.58–0.6) | 0.7 (0.69–0.7) | 0.63 (0.63–0.64) | 0.64 (0.63–0.64) | 0.53 (0.52–0.54) |
| 1 | | 0.79 (0.79–0.8) | 0.72 (0.72–0.73) | 0.71 (0.71–0.72) | 0.61 (0.6–0.62) | 0.69 (0.69–0.7) | 0.63 (0.62–0.64) | 0.64 (0.63–0.64) | 0.53 (0.52–0.54) |
| 2 | | 0.79 (0.78–0.79) | 0.72 (0.71–0.73) | 0.73 (0.72–0.74) | 0.62 (0.61–0.63) | 0.69 (0.68–0.7) | 0.62 (0.61–0.63) | 0.64 (0.63–0.65) | 0.53 (0.52–0.54) |
| 3 | | 0.78 (0.77–0.79) | 0.72 (0.71–0.72) | 0.74 (0.73–0.75) | 0.63 (0.62–0.64) | 0.68 (0.67–0.69) | 0.61 (0.6–0.62) | 0.64 (0.64–0.65) | 0.53 (0.52–0.54) |
| 4 | | 0.78 (0.77–0.78) | 0.71 (0.7–0.72) | 0.76 (0.75–0.77) | 0.63 (0.63–0.64) | 0.67 (0.66–0.68) | 0.6 (0.59–0.61) | 0.64 (0.63–0.65) | 0.53 (0.52–0.54) |
| 5 | | 0.77 (0.76–0.77) | 0.7 (0.69–0.71) | 0.76 (0.76–0.77) | 0.65 (0.64–0.66) | 0.67 (0.66–0.67) | 0.6 (0.59–0.6) | 0.63 (0.62–0.64) | 0.53 (0.52–0.54) |

Marg. pred. = Marginal prediction, * Thousand euros (€), ** Probability

**eTable 20. Estimates for marginal predictions of annual income and employment among parents exposed to child's developmental disorder (F84) and unexposed parents**

| | | Finland | | | | Denmark | | | |
|---|---|---|---|---|---|---|---|---|---|
| | | Men | | Women | | Men | | Women | |
| | | Unexposed | Exposed | Unexposed | Exposed | Unexposed | Exposed | Unexposed | Exposed |
| Years since diagnosis | Outcome | Marg. pred. (95% CI) | Marg. pred. (95% CI) | Marg. pred. (95% CI) | Marg. pred. (95% CI) | Marg. pred. (95% CI) | Marg. pred. (95% CI) | Marg. pred. (95% CI) | Marg. pred. (95% CI) |
| | | Unexposed | Exposed | Unexposed | Exposed | Unexposed | Exposed | Unexposed | Exposed |
| -5 | Earnings* | 33.1 (32.7–33.4) | 30.4 (30–30.7) | 17.5 (17.3–17.7) | 15.6 (15.4–15.9) | 55.2 (54.9–55.5) | 52.8 (52.5–53) | 42.3 (42.1–42.5) | 40.7 (40.6–40.9) |
| -4 | | 33.9 (33.5–34.2) | 31.2 (30.9–31.6) | 18.2 (18–18.5) | 16.1 (15.9–16.3) | 56.4 (56–56.7) | 53.7 (53.4–54) | 43.3 (43.1–43.5) | 41.7 (41.5–41.8) |
| -3 | | 34.8 (34.4–35.2) | 31.9 (31.6–32.3) | 18.9 (18.6–19.1) | 16.5 (16.3–16.8) | 57.6 (57.2–57.9) | 54.6 (54.3–55) | 44.3 (44.1–44.5) | 42.6 (42.4–42.7) |
| -2 | | 35.4 (35–35.8) | 32.5 (32.1–32.8) | 19.9 (19.6–20.2) | 17.2 (17–17.5) | 58.6 (58.3–59) | 55.7 (55.4–56) | 45.2 (45–45.4) | 43.3 (43.2–43.5) |
| -1 | | 36.3 (35.9–36.7) | 33.1 (32.8–33.5) | 21.3 (21–21.6) | 18.4 (18.2–18.7) | 59.9 (59.5–60.3) | 56.8 (56.5–57.1) | 46.1 (45.9–46.3) | 44.1 (43.9–44.3) |
| 0 | | 37.1 (36.7–37.5) | 33.7 (33.3–34) | 22.6 (22.3–22.9) | 19.6 (19.3–19.9) | 60.6 (60.2–61) | 57.5 (57.2–57.8) | 46.9 (46.7–47.1) | 44.7 (44.5–44.9) |
| 1 | | 37.7 (37.2–38.1) | 34 (33.6–34.4) | 23.8 (23.5–24.1) | 20.7 (20.4–21) | 61.7 (61.3–62.1) | 58 (57.7–58.4) | 47.5 (47.3–47.7) | 45.5 (45.3–45.7) |
| 2 | | 38.2 (37.8–38.6) | 34.4 (34–34.8) | 24.8 (24.5–25.1) | 21.8 (21.6–22.1) | 62.3 (61.9–62.7) | 58.7 (58.3–59) | 48.1 (47.9–48.4) | 46 (45.8–46.2) |
| 3 | | 38.5 (38.1–39) | 34.7 (34.2–35.1) | 25.8 (25.5–26) | 22.6 (22.3–22.9) | 63.3 (62.9–63.7) | 59.3 (59–59.7) | 48.7 (48.4–48.9) | 46.4 (46.2–46.6) |

**eTable 20. Estimates for marginal predictions of annual income and employment among parents exposed to child's developmental disorder (F84) and unexposed parents**

| | | Finland | | | | Denmark | | | |
|---|---|---|---|---|---|---|---|---|---|
| | | Men | | Women | | Men | | Women | |
| | | Unexposed | Exposed | Unexposed | Exposed | Unexposed | Exposed | Unexposed | Exposed |
| Years since diagnosis | Outcome | Marg. pred. (95% CI) | Marg. pred. (95% CI) | Marg. pred. (95% CI) | Marg. pred. (95% CI) | Marg. pred. (95% CI) | Marg. pred. (95% CI) | Marg. pred. (95% CI) | Marg. pred. (95% CI) |
| | | Unexposed | Exposed | Unexposed | Exposed | Unexposed | Exposed | Unexposed | Exposed |
| 4 | | 38.9 (38.4–39.3) | 35.1 (34.7–35.6) | 26.6 (26.3–26.9) | 23.3 (23–23.6) | 63.8 (63.4–64.3) | 59.8 (59.4–60.2) | 49.1 (48.8–49.3) | 46.7 (46.5–47) |
| 5 | | 39.1 (38.6–39.6) | 35.6 (35.1–36) | 27.4 (27.1–27.7) | 24.2 (23.8–24.5) | 64.3 (63.9–64.8) | 60.3 (59.8–60.7) | 49.6 (49.3–49.8) | 47.1 (46.8–47.3) |
| -5 | Disposable income* | 27.1 (26.9–27.3) | 25.5 (25.2–25.7) | 21.1 (21–21.3) | 19.7 (19.6–19.9) | 34 (33.8–34.2) | 32.6 (32.4–32.8) | 29.5 (29.3–29.6) | 28.4 (28.3–28.5) |
| -4 | | 27.8 (27.6–28) | 26.1 (25.9–26.3) | 21.9 (21.8–22.1) | 20.4 (20.3–20.6) | 35.1 (34.9–35.3) | 33.5 (33.3–33.7) | 30.4 (30.3–30.5) | 29.2 (29.1–29.3) |
| -3 | | 28.5 (28.3–28.8) | 26.8 (26.5–27) | 22.6 (22.5–22.8) | 21.1 (20.9–21.2) | 36.1 (35.9–36.3) | 34.4 (34.2–34.5) | 31.2 (31.1–31.3) | 30 (29.9–30.1) |
| -2 | | 29.1 (28.9–29.4) | 27.2 (27–27.5) | 23.3 (23.1–23.4) | 21.7 (21.6–21.9) | 37.1 (36.9–37.3) | 35.3 (35.1–35.5) | 32 (31.9–32.1) | 30.8 (30.6–30.9) |
| -1 | | 29.7 (29.5–30) | 27.8 (27.6–28.1) | 24 (23.9–24.2) | 22.4 (22.2–22.5) | 38.1 (37.9–38.4) | 36.2 (36–36.4) | 32.8 (32.7–32.9) | 31.4 (31.3–31.5) |
| 0 | | 30.4 (30.1–30.6) | 28.3 (28.1–28.5) | 24.7 (24.6–24.9) | 23.1 (22.9–23.2) | 39 (38.7–39.2) | 37 (36.8–37.2) | 33.5 (33.4–33.6) | 32 (31.8–32.1) |
| 1 | | 30.9 (30.7–31.2) | 28.7 (28.5–28.9) | 25.4 (25.3–25.6) | 23.8 (23.6–23.9) | 40 (39.7–40.2) | 37.7 (37.5–37.9) | 34.1 (33.9–34.2) | 32.6 (32.5–32.8) |

**eTable 20. Estimates for marginal predictions of annual income and employment among parents exposed to child's developmental disorder (F84) and unexposed parents**

| | | Finland | | | | Denmark | | | |
|---|---|---|---|---|---|---|---|---|---|
| | | Men | | Women | | Men | | Women | |
| | | Unexposed | Exposed | Unexposed | Exposed | Unexposed | Exposed | Unexposed | Exposed |
| Years since diagnosis | Outcome | Marg. pred. (95% CI) | Marg. pred. (95% CI) | Marg. pred. (95% CI) | Marg. pred. (95% CI) | Marg. pred. (95% CI) | Marg. pred. (95% CI) | Marg. pred. (95% CI) | Marg. pred. (95% CI) |
| | | Unexposed | Exposed | Unexposed | Exposed | Unexposed | Exposed | Unexposed | Exposed |
| 2 | | 31.5 (31.2–31.7) | 29.2 (29–29.5) | 26 (25.8–26.2) | 24.4 (24.3–24.6) | 40.7 (40.5–41) | 38.5 (38.3–38.7) | 34.6 (34.5–34.8) | 33.2 (33–33.3) |
| 3 | | 31.9 (31.6–32.2) | 29.7 (29.4–29.9) | 26.6 (26.4–26.7) | 24.9 (24.8–25.1) | 41.8 (41.5–42) | 39.2 (39–39.5) | 35.2 (35–35.3) | 33.6 (33.4–33.7) |
| 4 | | 32.5 (32.2–32.8) | 30.2 (29.9–30.5) | 27 (26.9–27.2) | 25.3 (25.2–25.5) | 42.6 (42.3–42.8) | 39.9 (39.7–40.2) | 35.6 (35.4–35.8) | 33.9 (33.8–34.1) |
| 5 | | 32.9 (32.6–33.2) | 30.8 (30.5–31.1) | 27.5 (27.3–27.6) | 25.8 (25.6–26) | 43.4 (43.1–43.7) | 40.7 (40.4–40.9) | 36.1 (36–36.3) | 34.3 (34.2–34.5) |
| -5 | Social welfare benefits* | 2.8 (2.8–2.9) | 3.3 (3.2–3.4) | 8.4 (8.3–8.5) | 8.5 (8.4–8.6) | 3.7 (3.6–3.8) | 3.9 (3.8–4) | 10.2 (10.1–10.2) | 10.7 (10.6–10.8) |
| -4 | | 3 (2.9–3.1) | 3.4 (3.3–3.5) | 8.6 (8.5–8.7) | 8.8 (8.7–8.9) | 3.7 (3.6–3.8) | 4 (3.9–4) | 10.2 (10.1–10.3) | 10.8 (10.7–10.9) |
| -3 | | 3.1 (3–3.2) | 3.7 (3.6–3.8) | 8.8 (8.7–8.9) | 9.1 (9–9.2) | 3.7 (3.7–3.8) | 4 (3.9–4.1) | 10.1 (10–10.2) | 10.9 (10.8–11) |
| -2 | | 3.3 (3.2–3.4) | 3.8 (3.7–3.9) | 8.7 (8.6–8.8) | 9.2 (9.1–9.3) | 3.8 (3.7–3.9) | 4 (4–4.1) | 9.9 (9.8–10) | 10.7 (10.6–10.8) |
| -1 | | 3.3 (3.2–3.4) | 3.9 (3.8–4) | 8.3 (8.2–8.4) | 8.9 (8.7–9) | 3.8 (3.7–3.9) | 4.1 (4–4.2) | 9.5 (9.4–9.6) | 10.5 (10.4–10.6) |
| 0 | | 3.4 (3.3–3.5) | 4 (3.9–4.1) | 8 (7.9–8.1) | 8.6 (8.5–8.7) | 3.9 (3.8–3.9) | 4.2 (4.1–4.3) | 9.3 (9.2–9.4) | 10.4 (10.3–10.5) |

**eTable 20. Estimates for marginal predictions of annual income and employment among parents exposed to child's developmental disorder (F84) and unexposed parents**

| | | Finland | | | | Denmark | | | |
|---|---|---|---|---|---|---|---|---|---|
| | | Men | | Women | | Men | | Women | |
| | | Unexposed | Exposed | Unexposed | Exposed | Unexposed | Exposed | Unexposed | Exposed |
| Years since diagnosis | Outcome | Marg. pred. (95% CI) | Marg. pred. (95% CI) | Marg. pred. (95% CI) | Marg. pred. (95% CI) | Marg. pred. (95% CI) | Marg. pred. (95% CI) | Marg. pred. (95% CI) | Marg. pred. (95% CI) |
| | | Unexposed | Exposed | Unexposed | Exposed | Unexposed | Exposed | Unexposed | Exposed |
| 1 | | 3.5 (3.4–3.6) | 4.2 (4.1–4.3) | 7.8 (7.6–7.9) | 8.5 (8.3–8.6) | 3.9 (3.8–4) | 4.4 (4.3–4.4) | 9.1 (9–9.2) | 10.3 (10.2–10.4) |
| 2 | | 3.6 (3.5–3.7) | 4.3 (4.2–4.4) | 7.5 (7.4–7.7) | 8.2 (8.1–8.3) | 4 (3.9–4.1) | 4.4 (4.3–4.5) | 8.8 (8.7–8.9) | 10.1 (9.9–10.2) |
| 3 | | 3.8 (3.7–3.9) | 4.5 (4.4–4.6) | 7.3 (7.2–7.4) | 8.2 (8–8.3) | 4.1 (4–4.2) | 4.6 (4.5–4.7) | 8.6 (8.5–8.7) | 9.9 (9.8–10.1) |
| 4 | | 4.1 (3.9–4.2) | 4.7 (4.6–4.8) | 7.1 (7–7.2) | 8 (7.9–8.1) | 4.2 (4.2–4.3) | 4.7 (4.6–4.8) | 8.4 (8.3–8.5) | 9.7 (9.6–9.9) |
| 5 | | 4.2 (4.1–4.4) | 5.1 (4.9–5.2) | 6.9 (6.8–7) | 7.8 (7.7–8) | 4.3 (4.2–4.4) | 4.8 (4.7–4.9) | 8.2 (8–8.3) | 9.6 (9.4–9.7) |
| -5 | Employment** | 0.82 (0.82–0.83) | 0.78 (0.78–0.79) | 0.68 (0.67–0.68) | 0.62 (0.62–0.63) | 0.74 (0.74–0.74) | 0.74 (0.74–0.75) | 0.68 (0.67–0.68) | 0.65 (0.65–0.65) |
| -4 | | 0.82 (0.81–0.83) | 0.78 (0.77–0.79) | 0.69 (0.68–0.69) | 0.62 (0.62–0.63) | 0.75 (0.75–0.75) | 0.75 (0.75–0.75) | 0.69 (0.68–0.69) | 0.66 (0.66–0.66) |
| -3 | | 0.82 (0.81–0.83) | 0.78 (0.77–0.79) | 0.7 (0.69–0.7) | 0.63 (0.62–0.64) | 0.75 (0.75–0.76) | 0.76 (0.75–0.76) | 0.7 (0.69–0.7) | 0.67 (0.66–0.67) |
| -2 | | 0.82 (0.81–0.83) | 0.78 (0.78–0.79) | 0.71 (0.71–0.72) | 0.64 (0.63–0.65) | 0.76 (0.75–0.76) | 0.75 (0.75–0.76) | 0.7 (0.7–0.71) | 0.67 (0.67–0.68) |
| -1 | | 0.82 (0.82–0.83) | 0.78 (0.78–0.79) | 0.73 (0.72–0.74) | 0.66 (0.66–0.67) | 0.75 (0.75–0.76) | 0.76 (0.75–0.76) | 0.71 (0.71–0.72) | 0.68 (0.67–0.68) |

**eTable 20. Estimates for marginal predictions of annual income and employment among parents exposed to child's developmental disorder (F84) and unexposed parents**

| | | Finland | | | | Denmark | | | |
|---|---|---|---|---|---|---|---|---|---|
| | | Men | | Women | | Men | | Women | |
| | | Unexposed | Exposed | Unexposed | Exposed | Unexposed | Exposed | Unexposed | Exposed |
| Years since diagnosis | Outcome | Marg. pred. (95% CI) | Marg. pred. (95% CI) | Marg. pred. (95% CI) | Marg. pred. (95% CI) | Marg. pred. (95% CI) | Marg. pred. (95% CI) | Marg. pred. (95% CI) | Marg. pred. (95% CI) |
| | | Unexposed | Exposed | Unexposed | Exposed | Unexposed | Exposed | Unexposed | Exposed |
| 0 | | 0.82 (0.82–0.83) | 0.78 (0.77–0.78) | 0.75 (0.74–0.76) | 0.68 (0.68–0.69) | 0.75 (0.75–0.76) | 0.75 (0.75–0.76) | 0.72 (0.71–0.72) | 0.68 (0.67–0.68) |
| 1 | | 0.82 (0.81–0.82) | 0.78 (0.77–0.78) | 0.77 (0.76–0.77) | 0.69 (0.69–0.7) | 0.75 (0.75–0.76) | 0.75 (0.74–0.75) | 0.72 (0.71–0.72) | 0.68 (0.67–0.68) |
| 2 | | 0.81 (0.81–0.82) | 0.77 (0.77–0.78) | 0.78 (0.77–0.78) | 0.7 (0.7–0.71) | 0.75 (0.75–0.75) | 0.74 (0.74–0.75) | 0.72 (0.72–0.73) | 0.68 (0.67–0.68) |
| 3 | | 0.81 (0.8–0.81) | 0.77 (0.76–0.78) | 0.79 (0.78–0.79) | 0.71 (0.71–0.72) | 0.75 (0.74–0.75) | 0.74 (0.73–0.74) | 0.72 (0.72–0.73) | 0.68 (0.67–0.68) |
| 4 | | 0.8 (0.79–0.8) | 0.76 (0.75–0.77) | 0.8 (0.79–0.81) | 0.72 (0.71–0.73) | 0.74 (0.73–0.74) | 0.74 (0.73–0.74) | 0.73 (0.72–0.73) | 0.67 (0.67–0.68) |
| 5 | | 0.79 (0.78–0.8) | 0.74 (0.74–0.75) | 0.8 (0.8–0.81) | 0.73 (0.72–0.73) | 0.73 (0.73–0.74) | 0.73 (0.73–0.74) | 0.73 (0.72–0.73) | 0.67 (0.67–0.68) |

Marg. pred. = Marginal prediction, * Thousand euros (€), ** Probability

**eTable 21. Estimates for marginal predictions of annual income and employment among parents exposed to child's childhood onset disorder (F90–F98) and unexposed parents**

| | | Finland | | | | Denmark | | | |
|---|---|---|---|---|---|---|---|---|---|
| | | Men | | Women | | Men | | Women | |
| | | Unexposed | Exposed | Unexposed | Exposed | Unexposed | Exposed | Unexposed | Exposed |
| Years since diagnosis | Outcome | Marg. pred. (95% CI) | Marg. pred. (95% CI) | Marg. pred. (95% CI) | Marg. pred. (95% CI) | Marg. pred. (95% CI) | Marg. pred. (95% CI) | Marg. pred. (95% CI) | Marg. pred. (95% CI) |
| | | Unexposed | Exposed | Unexposed | Exposed | Unexposed | Exposed | Unexposed | Exposed |
| -5 | Earnings* | 31.6 (31.5–31.8) | 29.1 (29–29.3) | 16.1 (16.1–16.2) | 15.2 (15.1–15.3) | 51.3 (51.1–51.5) | 48.9 (48.7–49.1) | 40 (39.9–40.2) | 39.2 (39.1–39.3) |
| -4 | | 32.4 (32.3–32.6) | 30 (29.8–30.1) | 17.4 (17.3–17.5) | 16.2 (16.1–16.3) | 52.2 (52–52.4) | 49.6 (49.4–49.8) | 40.9 (40.8–41) | 40 (39.9–40.1) |
| -3 | | 33.2 (33.1–33.4) | 30.6 (30.5–30.8) | 18.7 (18.6–18.8) | 17.3 (17.2–17.4) | 53.1 (52.9–53.3) | 50.4 (50.2–50.6) | 41.7 (41.6–41.8) | 40.7 (40.6–40.8) |
| -2 | | 34 (33.9–34.2) | 31.3 (31.1–31.4) | 19.9 (19.8–20) | 18.4 (18.3–18.5) | 54 (53.8–54.3) | 51.2 (51–51.4) | 42.4 (42.3–42.5) | 41.3 (41.2–41.4) |
| -1 | | 34.7 (34.5–34.8) | 31.9 (31.7–32) | 21 (20.9–21.1) | 19.3 (19.2–19.4) | 54.9 (54.7–55.1) | 51.8 (51.6–52) | 43.1 (42.9–43.2) | 41.9 (41.8–42) |
| 0 | | 35.3 (35.1–35.4) | 32.4 (32.3–32.6) | 22.2 (22.1–22.3) | 20.2 (20.1–20.3) | 55.6 (55.4–55.8) | 52.4 (52.2–52.6) | 43.7 (43.5–43.8) | 42.2 (42.1–42.4) |
| 1 | | 35.8 (35.7–36) | 32.8 (32.6–32.9) | 23.2 (23.1–23.3) | 21 (20.9–21.1) | 56.3 (56.1–56.6) | 52.9 (52.7–53.1) | 44.2 (44.1–44.3) | 42.7 (42.6–42.8) |
| 2 | | 36.2 (36–36.4) | 33.2 (33–33.4) | 24.1 (24–24.2) | 21.9 (21.8–22) | 56.8 (56.6–57.1) | 53.3 (53.1–53.5) | 44.6 (44.5–44.8) | 43 (42.9–43.1) |
| 3 | | 36.6 (36.5–36.8) | 33.5 (33.3–33.7) | 25 (24.9–25.1) | 22.7 (22.6–22.8) | 57.4 (57.1–57.6) | 53.7 (53.5–54) | 45.1 (44.9–45.2) | 43.3 (43.2–43.4) |

**eTable 21. Estimates for marginal predictions of annual income and employment among parents exposed to child's childhood onset disorder (F90–F98) and unexposed parents**

| | | Finland | | | | Denmark | | | |
|---|---|---|---|---|---|---|---|---|---|
| | | Men | | Women | | Men | | Women | |
| | | Unexposed | Exposed | Unexposed | Exposed | Unexposed | Exposed | Unexposed | Exposed |
| Years since diagnosis | Outcome | Marg. pred. (95% CI) | Marg. pred. (95% CI) | Marg. pred. (95% CI) | Marg. pred. (95% CI) | Marg. pred. (95% CI) | Marg. pred. (95% CI) | Marg. pred. (95% CI) | Marg. pred. (95% CI) |
| | | Unexposed | Exposed | Unexposed | Exposed | Unexposed | Exposed | Unexposed | Exposed |
| 4 | | 36.9 (36.7–37) | 33.6 (33.5–33.8) | 25.8 (25.7–25.9) | 23.4 (23.3–23.6) | 57.9 (57.6–58.2) | 54.1 (53.9–54.4) | 45.4 (45.3–45.5) | 43.5 (43.4–43.7) |
| 5 | | 37.1 (36.9–37.3) | 33.8 (33.6–34) | 26.5 (26.4–26.6) | 24.1 (24–24.3) | 58.3 (58–58.6) | 54.5 (54.2–54.7) | 45.8 (45.6–45.9) | 43.8 (43.7–44) |
| -5 | Disposable income* | 26.2 (26.2–26.3) | 24.8 (24.7–24.9) | 20.5 (20.5–20.6) | 20.3 (20.2–20.3) | 31.6 (31.5–31.7) | 30.1 (30–30.2) | 28.1 (28–28.2) | 27.4 (27.4–27.5) |
| -4 | | 26.9 (26.8–27) | 25.5 (25.4–25.6) | 21.4 (21.3–21.4) | 21.1 (21–21.1) | 32.5 (32.4–32.6) | 30.8 (30.6–30.9) | 28.8 (28.8–28.9) | 28.1 (28–28.2) |
| -3 | | 27.5 (27.4–27.6) | 26 (25.9–26.1) | 22.2 (22.2–22.3) | 21.9 (21.9–22) | 33.3 (33.2–33.4) | 31.5 (31.4–31.6) | 29.5 (29.4–29.6) | 28.7 (28.6–28.7) |
| -2 | | 28.2 (28.1–28.3) | 26.6 (26.5–26.7) | 23 (23–23.1) | 22.7 (22.6–22.7) | 34.1 (34–34.2) | 32.2 (32.1–32.3) | 30.1 (30–30.2) | 29.2 (29.2–29.3) |
| -1 | | 28.8 (28.7–28.9) | 27.2 (27.1–27.3) | 23.7 (23.6–23.8) | 23.3 (23.3–23.4) | 34.9 (34.8–35.1) | 32.8 (32.7–32.9) | 30.7 (30.6–30.8) | 29.7 (29.7–29.8) |
| 0 | | 29.4 (29.3–29.5) | 27.7 (27.6–27.8) | 24.3 (24.3–24.4) | 23.9 (23.8–23.9) | 35.7 (35.6–35.8) | 33.5 (33.4–33.6) | 31.2 (31.1–31.3) | 30.1 (30–30.2) |
| 1 | | 29.9 (29.8–30) | 28.1 (28–28.2) | 24.9 (24.8–24.9) | 24.4 (24.3–24.4) | 36.5 (36.3–36.6) | 34.2 (34–34.3) | 31.7 (31.6–31.8) | 30.5 (30.4–30.6) |

**eTable 21. Estimates for marginal predictions of annual income and employment among parents exposed to child's childhood onset disorder (F90–F98) and unexposed parents**

| | | Finland | | | | Denmark | | | |
|---|---|---|---|---|---|---|---|---|---|
| | | Men | | Women | | Men | | Women | |
| | | Unexposed | Exposed | Unexposed | Exposed | Unexposed | Exposed | Unexposed | Exposed |
| Years since diagnosis | Outcome | Marg. pred. (95% CI) | Marg. pred. (95% CI) | Marg. pred. (95% CI) | Marg. pred. (95% CI) | Marg. pred. (95% CI) | Marg. pred. (95% CI) | Marg. pred. (95% CI) | Marg. pred. (95% CI) |
| | | Unexposed | Exposed | Unexposed | Exposed | Unexposed | Exposed | Unexposed | Exposed |
| 2 | | 30.4 (30.3–30.5) | 28.6 (28.5–28.7) | 25.4 (25.3–25.4) | 24.8 (24.8–24.9) | 37.2 (37–37.3) | 34.8 (34.6–34.9) | 32.1 (32–32.2) | 30.8 (30.7–30.9) |
| 3 | | 30.9 (30.8–31) | 29 (28.9–29.1) | 25.8 (25.8–25.9) | 25.3 (25.2–25.3) | 37.9 (37.7–38) | 35.4 (35.2–35.5) | 32.5 (32.4–32.6) | 31.1 (31–31.1) |
| 4 | | 31.3 (31.2–31.5) | 29.4 (29.3–29.5) | 26.2 (26.2–26.3) | 25.7 (25.6–25.7) | 38.6 (38.4–38.7) | 36 (35.9–36.2) | 32.8 (32.7–32.9) | 31.4 (31.3–31.4) |
| 5 | | 31.8 (31.7–31.9) | 29.8 (29.6–29.9) | 26.6 (26.5–26.7) | 26 (26–26.1) | 39.2 (39–39.4) | 36.6 (36.5–36.8) | 33.2 (33.1–33.3) | 31.6 (31.5–31.7) |
| -5 | Social welfare benefits* | 3 (3–3.1) | 3.5 (3.5–3.6) | 8.9 (8.9–8.9) | 9.4 (9.4–9.4) | 4.4 (4.3–4.4) | 4.9 (4.9–5) | 11.3 (11.2–11.4) | 12.6 (12.5–12.6) |
| -4 | | 3.2 (3.1–3.2) | 3.7 (3.6–3.7) | 8.8 (8.7–8.8) | 9.3 (9.3–9.4) | 4.4 (4.4–4.5) | 5.1 (5–5.1) | 11.2 (11.1–11.3) | 12.5 (12.4–12.6) |
| -3 | | 3.3 (3.2–3.3) | 3.8 (3.8–3.9) | 8.6 (8.5–8.6) | 9.3 (9.3–9.4) | 4.5 (4.5–4.6) | 5.1 (5.1–5.2) | 11 (10.9–11) | 12.4 (12.3–12.5) |
| -2 | | 3.3 (3.3–3.4) | 3.9 (3.9–3.9) | 8.3 (8.3–8.4) | 9.2 (9.1–9.2) | 4.6 (4.6–4.7) | 5.3 (5.2–5.3) | 10.8 (10.7–10.9) | 12.3 (12.2–12.3) |
| -1 | | 3.4 (3.4–3.5) | 4 (4–4.1) | 8.1 (8.1–8.2) | 9.1 (9.1–9.2) | 4.7 (4.6–4.7) | 5.4 (5.3–5.4) | 10.6 (10.5–10.7) | 12.2 (12.1–12.2) |
| 0 | | 3.5 (3.5–3.5) | 4.1 (4.1–4.2) | 7.9 (7.8–7.9) | 9 (8.9–9) | 4.8 (4.7–4.8) | 5.5 (5.5–5.6) | 10.4 (10.3–10.4) | 12.1 (12–12.2) |

**eTable 21. Estimates for marginal predictions of annual income and employment among parents exposed to child's childhood onset disorder (F90–F98) and unexposed parents**

| | | Finland | | | | Denmark | | | |
|---|---|---|---|---|---|---|---|---|---|
| | | Men | | Women | | Men | | Women | |
| | | Unexposed | Exposed | Unexposed | Exposed | Unexposed | Exposed | Unexposed | Exposed |
| Years since diagnosis | Outcome | Marg. pred. (95% CI) | Marg. pred. (95% CI) | Marg. pred. (95% CI) | Marg. pred. (95% CI) | Marg. pred. (95% CI) | Marg. pred. (95% CI) | Marg. pred. (95% CI) | Marg. pred. (95% CI) |
| | | Unexposed | Exposed | Unexposed | Exposed | Unexposed | Exposed | Unexposed | Exposed |
| 1 | | 3.6 (3.6–3.7) | 4.3 (4.2–4.3) | 7.6 (7.6–7.7) | 8.8 (8.8–8.8) | 4.9 (4.8–4.9) | 5.7 (5.6–5.8) | 10.1 (10–10.2) | 11.9 (11.8–12) |
| 2 | | 3.8 (3.7–3.8) | 4.5 (4.4–4.5) | 7.4 (7.3–7.4) | 8.6 (8.5–8.6) | 5 (4.9–5.1) | 5.8 (5.8–5.9) | 9.8 (9.8–9.9) | 11.7 (11.6–11.8) |
| 3 | | 3.9 (3.9–4) | 4.6 (4.6–4.7) | 7.1 (7.1–7.2) | 8.4 (8.3–8.4) | 5.1 (5–5.2) | 5.9 (5.9–6) | 9.6 (9.5–9.7) | 11.5 (11.4–11.6) |
| 4 | | 4.2 (4.1–4.2) | 4.9 (4.8–4.9) | 6.9 (6.9–7) | 8.2 (8.2–8.3) | 5.2 (5.2–5.3) | 6.1 (6–6.2) | 9.4 (9.3–9.5) | 11.3 (11.2–11.4) |
| 5 | | 4.4 (4.3–4.4) | 5.1 (5.1–5.2) | 6.7 (6.6–6.7) | 8 (8–8.1) | 5.4 (5.3–5.5) | 6.3 (6.2–6.4) | 9.2 (9.1–9.3) | 11.1 (11–11.2) |
| -5 | Employment** | 0.81 (0.8–0.81) | 0.77 (0.76–0.77) | 0.66 (0.66–0.66) | 0.62 (0.62–0.63) | 0.71 (0.7–0.71) | 0.69 (0.69–0.7) | 0.62 (0.62–0.63) | 0.6 (0.59–0.6) |
| -4 | | 0.81 (0.8–0.81) | 0.77 (0.76–0.77) | 0.68 (0.68–0.68) | 0.64 (0.64–0.64) | 0.72 (0.71–0.72) | 0.7 (0.7–0.7) | 0.64 (0.64–0.65) | 0.61 (0.61–0.61) |
| -3 | | 0.81 (0.81–0.81) | 0.76 (0.76–0.77) | 0.7 (0.7–0.7) | 0.66 (0.66–0.66) | 0.72 (0.72–0.72) | 0.7 (0.7–0.7) | 0.65 (0.65–0.66) | 0.62 (0.61–0.62) |
| -2 | | 0.81 (0.8–0.81) | 0.76 (0.76–0.77) | 0.72 (0.72–0.72) | 0.67 (0.67–0.68) | 0.72 (0.72–0.72) | 0.7 (0.7–0.7) | 0.66 (0.66–0.66) | 0.62 (0.62–0.63) |
| -1 | | 0.81 (0.8–0.81) | 0.77 (0.76–0.77) | 0.74 (0.73–0.74) | 0.69 (0.68–0.69) | 0.72 (0.72–0.72) | 0.7 (0.7–0.7) | 0.67 (0.66–0.67) | 0.62 (0.62–0.63) |

**eTable 21. Estimates for marginal predictions of annual income and employment among parents exposed to child's childhood onset disorder (F90–F98) and unexposed parents**

| | | Finland | | | | Denmark | | | |
|---|---|---|---|---|---|---|---|---|---|
| | | Men | | Women | | Men | | Women | |
| | | Unexposed | Exposed | Unexposed | Exposed | Unexposed | Exposed | Unexposed | Exposed |
| Years since diagnosis | Outcome | Marg. pred. (95% CI) | Marg. pred. (95% CI) | Marg. pred. (95% CI) | Marg. pred. (95% CI) | Marg. pred. (95% CI) | Marg. pred. (95% CI) | Marg. pred. (95% CI) | Marg. pred. (95% CI) |
| | | Unexposed | Exposed | Unexposed | Exposed | Unexposed | Exposed | Unexposed | Exposed |
| 0 | | 0.81 (0.8–0.81) | 0.77 (0.76–0.77) | 0.75 (0.75–0.75) | 0.7 (0.7–0.7) | 0.72 (0.71–0.72) | 0.7 (0.69–0.7) | 0.67 (0.67–0.67) | 0.62 (0.62–0.63) |
| 1 | | 0.8 (0.8–0.8) | 0.76 (0.76–0.76) | 0.76 (0.76–0.77) | 0.71 (0.71–0.71) | 0.71 (0.71–0.72) | 0.69 (0.69–0.7) | 0.67 (0.67–0.68) | 0.62 (0.62–0.62) |
| 2 | | 0.8 (0.79–0.8) | 0.75 (0.75–0.76) | 0.77 (0.77–0.78) | 0.72 (0.72–0.72) | 0.71 (0.71–0.71) | 0.68 (0.68–0.69) | 0.68 (0.67–0.68) | 0.62 (0.62–0.62) |
| 3 | | 0.79 (0.79–0.79) | 0.75 (0.74–0.75) | 0.78 (0.78–0.79) | 0.73 (0.73–0.73) | 0.71 (0.7–0.71) | 0.68 (0.68–0.68) | 0.68 (0.67–0.68) | 0.62 (0.62–0.62) |
| 4 | | 0.79 (0.78–0.79) | 0.74 (0.74–0.74) | 0.79 (0.79–0.79) | 0.73 (0.73–0.74) | 0.7 (0.7–0.7) | 0.68 (0.67–0.68) | 0.68 (0.67–0.68) | 0.62 (0.62–0.62) |
| 5 | | 0.78 (0.78–0.78) | 0.73 (0.73–0.73) | 0.79 (0.79–0.8) | 0.74 (0.74–0.74) | 0.69 (0.69–0.69) | 0.67 (0.66–0.67) | 0.68 (0.67–0.68) | 0.62 (0.61–0.62) |

Marg. pred. = Marginal prediction, * Thousand euros (€), ** Probability

#### Estimates for marginal predictions of annual income and employment among parents exposed to child's mental disorder and unexposed parents stratified by the child's age at diagnosis in Finland and Denmark

##### eTable 22. Estimates for marginal predictions of annual earnings among parents exposed to child's any mental disorder (F00–F99) and unexposed parents stratified by the child's age at diagnosis

| | | Finland | | | | Denmark | | | |
|---|---|---|---|---|---|---|---|---|---|
| | | Men | | Women | | Men | | Women | |
| | | Unexposed | Exposed | Unexposed | Exposed | Unexposed | Exposed | Unexposed | Exposed |
| Years since diagnosis | Age group | Marg. pred. (95% CI) | Marg. pred. (95% CI) | Marg. pred. (95% CI) | Marg. pred. (95% CI) | Marg. pred. (95% CI) | Marg. pred. (95% CI) | Marg. pred. (95% CI) | Marg. pred. (95% CI) |
| | | Unexposed | Exposed | Unexposed | Exposed | Unexposed | Exposed | Unexposed | Exposed |
| -5 | 1–4 years | 27.2 (27–27.4) | 25.7 (25.5–25.9) | 14.6 (14.4–14.7) | 13.5 (13.3–13.6) | 45.3 (44.7–45.9) | 42.3 (41.8–42.9) | 33.6 (33.3–34) | 31.1 (30.8–31.5) |
| -4 | | 28.7 (28.4–28.9) | 27 (26.8–27.2) | 11.6 (11.5–11.8) | 10.5 (10.4–10.7) | 47.5 (46.9–48.1) | 43.9 (43.4–44.5) | 34.5 (34.2–34.9) | 32 (31.6–32.3) |
| -3 | | 30.1 (29.9–30.3) | 28.1 (27.9–28.4) | 10.3 (10.2–10.4) | 8.8 (8.7–8.9) | 49.3 (48.7–50) | 45.4 (44.8–45.9) | 36.3 (36–36.7) | 33.7 (33.4–34) |
| -2 | | 31.5 (31.3–31.8) | 29.3 (29.1–29.5) | 11.8 (11.6–11.9) | 9.6 (9.5–9.7) | 51 (50.3–51.6) | 47.1 (46.5–47.7) | 38.3 (37.9–38.7) | 35.4 (35–35.7) |
| -1 | | 33 (32.8–33.3) | 30.6 (30.4–30.8) | 14 (13.9–14.2) | 11.2 (11.1–11.4) | 52.7 (52–53.3) | 48.5 (47.9–49.1) | 39.6 (39.3–40) | 36.6 (36.2–36.9) |
| 0 | | 34.3 (34.1–34.6) | 31.8 (31.5–32) | 16.4 (16.3–16.6) | 13.6 (13.4–13.7) | 54.1 (53.4–54.8) | 49.7 (49–50.3) | 41 (40.6–41.4) | 37.8 (37.4–38.1) |
| 1 | | 35.6 (35.3–35.8) | 32.7 (32.4–32.9) | 18.5 (18.3–18.7) | 15.7 (15.5–15.9) | 55.4 (54.7–56.2) | 50.9 (50.2–51.5) | 42.1 (41.7–42.5) | 39.2 (38.8–39.5) |
| 2 | | 36.6 (36.4–36.9) | 33.5 (33.2–33.7) | 20.1 (19.9–20.3) | 17.4 (17.2–17.6) | 56.5 (55.8–57.3) | 51.8 (51.1–52.5) | 43.4 (43–43.8) | 40.2 (39.8–40.6) |

**eTable 22. Estimates for marginal predictions of annual earnings among parents exposed to child's any mental disorder (F00–F99) and unexposed parents stratified by the child's age at diagnosis**

| | | Finland | | | | Denmark | | | |
|---|---|---|---|---|---|---|---|---|---|
| | | Men | | Women | | Men | | Women | |
| | | Unexposed | Exposed | Unexposed | Exposed | Unexposed | Exposed | Unexposed | Exposed |
| Years since diagnosis | Age group | Marg. pred. (95% CI) | Marg. pred. (95% CI) | Marg. pred. (95% CI) | Marg. pred. (95% CI) | Marg. pred. (95% CI) | Marg. pred. (95% CI) | Marg. pred. (95% CI) | Marg. pred. (95% CI) |
| | | Unexposed | Exposed | Unexposed | Exposed | Unexposed | Exposed | Unexposed | Exposed |
| 3 | | 37.6 (37.3–37.9) | 34.2 (33.9–34.5) | 21.5 (21.3–21.7) | 18.5 (18.3–18.7) | 57.8 (57–58.6) | 52.4 (51.7–53.1) | 44.4 (43.9–44.8) | 40.9 (40.5–41.4) |
| 4 | | 38.5 (38.2–38.8) | 34.9 (34.6–35.2) | 22.8 (22.6–23) | 19.5 (19.4–19.7) | 59.2 (58.4–60) | 53.3 (52.6–54.1) | 45.2 (44.8–45.7) | 41.7 (41.2–42.1) |
| 5 | | 39.3 (39–39.6) | 35.5 (35.2–35.8) | 24.1 (23.9–24.3) | 20.6 (20.4–20.8) | 60.2 (59.3–61.1) | 53.8 (53.1–54.6) | 46.2 (45.8–46.7) | 42.3 (41.8–42.7) |
| -5 | 5–9 years | 30.1 (30–30.3) | 27.8 (27.6–27.9) | 11.7 (11.6–11.8) | 10.6 (10.5–10.7) | 49.7 (49.4–50) | 47.5 (47.2–47.8) | 37.8 (37.7–38) | 37 (36.8–37.1) |
| -4 | | 31.4 (31.2–31.6) | 29 (28.8–29.1) | 14.2 (14.1–14.3) | 12.7 (12.6–12.8) | 50.8 (50.5–51.1) | 48.5 (48.2–48.8) | 39.3 (39.1–39.4) | 38.3 (38.1–38.4) |
| -3 | | 32.5 (32.4–32.7) | 30 (29.8–30.1) | 16.3 (16.2–16.4) | 14.6 (14.5–14.7) | 52 (51.7–52.3) | 49.6 (49.3–49.9) | 40.4 (40.2–40.6) | 39.3 (39.2–39.5) |
| -2 | | 33.6 (33.4–33.8) | 30.9 (30.8–31.1) | 18 (17.9–18.1) | 16 (15.9–16.1) | 53.4 (53–53.7) | 50.7 (50.4–51) | 41.4 (41.2–41.5) | 40.1 (40–40.3) |
| -1 | | 34.6 (34.5–34.8) | 31.8 (31.6–31.9) | 19.3 (19.2–19.5) | 17.1 (17–17.3) | 54.4 (54.1–54.8) | 51.8 (51.5–52.1) | 42.3 (42.1–42.5) | 41 (40.8–41.1) |
| 0 | | 35.7 (35.5–35.9) | 32.7 (32.5–32.9) | 20.7 (20.6–20.8) | 18.3 (18.1–18.4) | 55.5 (55.2–55.9) | 52.6 (52.3–52.9) | 43.2 (43–43.4) | 41.5 (41.4–41.7) |

**eTable 22. Estimates for marginal predictions of annual earnings among parents exposed to child's any mental disorder (F00–F99) and unexposed parents stratified by the child's age at diagnosis**

| | | Finland | | | | Denmark | | | |
|---|---|---|---|---|---|---|---|---|---|
| | | Men | | Women | | Men | | Women | |
| | | Unexposed | Exposed | Unexposed | Exposed | Unexposed | Exposed | Unexposed | Exposed |
| Years since diagnosis | Age group | Marg. pred. (95% CI) | Marg. pred. (95% CI) | Marg. pred. (95% CI) | Marg. pred. (95% CI) | Marg. pred. (95% CI) | Marg. pred. (95% CI) | Marg. pred. (95% CI) | Marg. pred. (95% CI) |
| | | Unexposed | Exposed | Unexposed | Exposed | Unexposed | Exposed | Unexposed | Exposed |
| 1 | | 36.5 (36.3–36.7) | 33.3 (33.1–33.5) | 21.9 (21.8–22) | 19.3 (19.2–19.5) | 56.6 (56.3–57) | 53.4 (53.1–53.7) | 43.9 (43.7–44.1) | 42.3 (42.1–42.5) |
| 2 | | 37.3 (37.1–37.5) | 34 (33.8–34.1) | 23.1 (23–23.2) | 20.5 (20.3–20.6) | 57.5 (57.1–57.9) | 54 (53.7–54.4) | 44.7 (44.5–44.9) | 42.9 (42.7–43) |
| 3 | | 38.1 (37.9–38.3) | 34.5 (34.3–34.7) | 24.3 (24.2–24.4) | 21.6 (21.4–21.7) | 58.2 (57.8–58.6) | 54.5 (54.2–54.9) | 45.4 (45.2–45.6) | 43.5 (43.3–43.7) |
| 4 | | 38.6 (38.4–38.8) | 35 (34.8–35.2) | 25.4 (25.3–25.6) | 22.6 (22.5–22.8) | 59.1 (58.7–59.5) | 55.3 (55–55.7) | 46.3 (46–46.5) | 44 (43.8–44.2) |
| 5 | | 39.2 (39–39.4) | 35.5 (35.3–35.7) | 26.5 (26.4–26.7) | 23.6 (23.5–23.8) | 59.8 (59.3–60.2) | 56.1 (55.7–56.4) | 47 (46.8–47.2) | 44.6 (44.4–44.8) |
| -5 | 10–14 years | 33.7 (33.5–33.9) | 31.4 (31.2–31.6) | 20 (19.9–20.1) | 18.6 (18.5–18.8) | 54.4 (54.1–54.7) | 52.3 (52–52.5) | 41.9 (41.8–42.1) | 41.2 (41.1–41.3) |
| -4 | | 34.4 (34.2–34.6) | 32.1 (31.9–32.3) | 21.1 (20.9–21.2) | 19.5 (19.4–19.7) | 55.4 (55.1–55.7) | 53.2 (52.9–53.5) | 42.7 (42.6–42.9) | 41.9 (41.8–42.1) |
| -3 | | 35 (34.8–35.3) | 32.6 (32.4–32.8) | 22.1 (21.9–22.2) | 20.4 (20.3–20.5) | 56.3 (56–56.6) | 53.9 (53.7–54.2) | 43.5 (43.4–43.7) | 42.6 (42.5–42.8) |
| -2 | | 35.7 (35.5–35.9) | 33.2 (33–33.4) | 23.1 (23–23.3) | 21.3 (21.2–21.5) | 57.3 (57–57.7) | 54.8 (54.5–55) | 44.3 (44.1–44.4) | 43.4 (43.2–43.5) |

**eTable 22. Estimates for marginal predictions of annual earnings among parents exposed to child's any mental disorder (F00–F99) and unexposed parents stratified by the child's age at diagnosis**

| | | Finland | | | | Denmark | | | |
|---|---|---|---|---|---|---|---|---|---|
| | | Men | | Women | | Men | | Women | |
| | | Unexposed | Exposed | Unexposed | Exposed | Unexposed | Exposed | Unexposed | Exposed |
| Years since diagnosis | Age group | Marg. pred. (95% CI) | Marg. pred. (95% CI) | Marg. pred. (95% CI) | Marg. pred. (95% CI) | Marg. pred. (95% CI) | Marg. pred. (95% CI) | Marg. pred. (95% CI) | Marg. pred. (95% CI) |
| | | Unexposed | Exposed | Unexposed | Exposed | Unexposed | Exposed | Unexposed | Exposed |
| -1 | | 36.3 (36–36.5) | 33.7 (33.5–33.9) | 24.1 (24–24.3) | 22.3 (22.1–22.4) | 58.1 (57.8–58.4) | 55.5 (55.2–55.8) | 45 (44.9–45.2) | 44 (43.9–44.2) |
| 0 | | 36.8 (36.6–37) | 34.1 (33.8–34.3) | 25 (24.9–25.2) | 22.9 (22.7–23) | 59.2 (58.8–59.5) | 56.2 (55.9–56.5) | 45.7 (45.5–45.9) | 44.6 (44.4–44.7) |
| 1 | | 37.4 (37.1–37.6) | 34.4 (34.1–34.6) | 25.9 (25.8–26.1) | 23.6 (23.5–23.8) | 59.9 (59.5–60.2) | 56.9 (56.6–57.2) | 46.4 (46.2–46.5) | 45.2 (45–45.3) |
| 2 | | 37.7 (37.4–37.9) | 34.7 (34.5–34.9) | 26.7 (26.5–26.9) | 24.4 (24.2–24.5) | 60.6 (60.2–61) | 57.2 (56.9–57.6) | 46.9 (46.7–47.1) | 45.6 (45.4–45.8) |
| 3 | | 38 (37.7–38.2) | 35 (34.7–35.2) | 27.5 (27.3–27.6) | 25 (24.9–25.2) | 61.2 (60.8–61.6) | 57.8 (57.4–58.1) | 47.3 (47.2–47.5) | 45.8 (45.6–46) |
| 4 | | 38.2 (37.9–38.4) | 35 (34.7–35.2) | 28.2 (28–28.4) | 25.7 (25.5–25.8) | 61.7 (61.3–62.1) | 58.1 (57.7–58.4) | 47.5 (47.3–47.7) | 45.9 (45.7–46.1) |
| 5 | | 38.2 (38–38.5) | 35 (34.7–35.2) | 28.8 (28.6–28.9) | 26.2 (26–26.4) | 62 (61.6–62.4) | 58.5 (58.2–58.9) | 47.7 (47.5–48) | 45.9 (45.7–46.1) |
| -5 | 15–19 years | 34.4 (34.2–34.6) | 32.2 (32.1–32.4) | 22.8 (22.7–22.9) | 21.4 (21.3–21.5) | 55.9 (55.7–56.2) | 53.7 (53.5–54) | 42.8 (42.7–42.9) | 42.2 (42.1–42.3) |
| -4 | | 34.8 (34.6–35) | 32.6 (32.4–32.8) | 23.5 (23.4–23.7) | 22.2 (22–22.3) | 56.6 (56.3–56.9) | 54.2 (53.9–54.4) | 43.5 (43.3–43.6) | 42.8 (42.7–42.9) |

**eTable 22. Estimates for marginal predictions of annual earnings among parents exposed to child's any mental disorder (F00–F99) and unexposed parents stratified by the child's age at diagnosis**

| | | Finland | | | | Denmark | | | |
|---|---|---|---|---|---|---|---|---|---|
| | | Men | | Women | | Men | | Women | |
| | | Unexposed | Exposed | Unexposed | Exposed | Unexposed | Exposed | Unexposed | Exposed |
| Years since diagnosis | Age group | Marg. pred. (95% CI) | Marg. pred. (95% CI) | Marg. pred. (95% CI) | Marg. pred. (95% CI) | Marg. pred. (95% CI) | Marg. pred. (95% CI) | Marg. pred. (95% CI) | Marg. pred. (95% CI) |
| | | Unexposed | Exposed | Unexposed | Exposed | Unexposed | Exposed | Unexposed | Exposed |
| -3 | | 35.3 (35.1–35.5) | 33 (32.8–33.1) | 24.2 (24.1–24.4) | 22.8 (22.7–22.9) | 57.1 (56.9–57.4) | 54.6 (54.4–54.9) | 44.1 (44–44.2) | 43.3 (43.2–43.4) |
| -2 | | 35.5 (35.3–35.7) | 33.2 (33.1–33.4) | 24.9 (24.7–25) | 23.5 (23.4–23.6) | 57.8 (57.5–58) | 55.2 (54.9–55.5) | 44.6 (44.5–44.8) | 43.7 (43.6–43.8) |
| -1 | | 35.8 (35.7–36) | 33.5 (33.3–33.7) | 25.4 (25.3–25.5) | 24.1 (24–24.2) | 58.4 (58.1–58.7) | 55.7 (55.4–55.9) | 44.9 (44.8–45) | 44 (43.9–44.1) |
| 0 | | 36 (35.8–36.2) | 33.5 (33.3–33.7) | 26 (25.9–26.1) | 24.6 (24.4–24.7) | 58.8 (58.5–59.1) | 56 (55.7–56.3) | 45.1 (45–45.3) | 44 (43.9–44.1) |
| 1 | | 36.1 (35.8–36.3) | 33.5 (33.3–33.7) | 26.5 (26.4–26.6) | 25 (24.9–25.1) | 59.2 (58.9–59.5) | 56.3 (56–56.6) | 45.2 (45.1–45.4) | 44 (43.9–44.2) |
| 2 | | 35.9 (35.7–36.1) | 33.5 (33.3–33.7) | 26.9 (26.7–27) | 25.4 (25.2–25.5) | 59.4 (59.1–59.7) | 56.4 (56.1–56.7) | 45.2 (45–45.4) | 43.9 (43.8–44.1) |
| 3 | | 35.7 (35.5–35.9) | 33.2 (33–33.4) | 27.2 (27–27.3) | 25.7 (25.6–25.8) | 59.5 (59.2–59.9) | 56.5 (56.2–56.8) | 45.2 (45–45.3) | 43.9 (43.8–44.1) |
| 4 | | 35.3 (35.1–35.5) | 32.8 (32.6–33) | 27.3 (27.2–27.4) | 25.9 (25.7–26) | 59.6 (59.3–59.9) | 56.5 (56.2–56.8) | 45.4 (45.2–45.5) | 44 (43.8–44.1) |
| 5 | | 34.8 (34.6–35) | 32.3 (32.1–32.6) | 27.4 (27.2–27.5) | 26 (25.8–26.1) | 59.5 (59.2–59.9) | 56.1 (55.8–56.4) | 45.4 (45.3–45.6) | 44.1 (43.9–44.2) |

**eTable 22. Estimates for marginal predictions of annual earnings among parents exposed to child's any mental disorder (F00–F99) and unexposed parents stratified by the child's age at diagnosis**

| | | Finland | | | | Denmark | | | |
|---|---|---|---|---|---|---|---|---|---|
| | | Men | | Women | | Men | | Women | |
| | | Unexposed | Exposed | Unexposed | Exposed | Unexposed | Exposed | Unexposed | Exposed |
| Years since diagnosis | Age group | Marg. pred. (95% CI) | Marg. pred. (95% CI) | Marg. pred. (95% CI) | Marg. pred. (95% CI) | Marg. pred. (95% CI) | Marg. pred. (95% CI) | Marg. pred. (95% CI) | Marg. pred. (95% CI) |
| | | Unexposed | Exposed | Unexposed | Exposed | Unexposed | Exposed | Unexposed | Exposed |
| -5 | 20–25 years | 33.2 (33–33.4) | 31.4 (31.2–31.6) | 23.6 (23.5–23.7) | 22.3 (22.2–22.4) | 55.8 (55.5–56.1) | 53.5 (53.2–53.7) | 41.6 (41.4–41.7) | 40.6 (40.5–40.8) |
| -4 | | 33.2 (33–33.4) | 31.3 (31.1–31.5) | 23.9 (23.8–24.1) | 22.6 (22.4–22.7) | 56.1 (55.8–56.4) | 53.5 (53.3–53.8) | 41.7 (41.5–41.8) | 40.7 (40.5–40.8) |
| -3 | | 33.1 (32.9–33.4) | 31.1 (30.9–31.3) | 24.3 (24.1–24.4) | 22.8 (22.7–22.9) | 56.3 (56–56.5) | 53.7 (53.5–54) | 41.7 (41.5–41.8) | 40.6 (40.5–40.8) |
| -2 | | 32.9 (32.7–33.1) | 30.8 (30.6–31.1) | 24.4 (24.3–24.5) | 23 (22.9–23.1) | 56.4 (56.1–56.7) | 53.7 (53.4–54) | 41.7 (41.5–41.8) | 40.6 (40.5–40.8) |
| -1 | | 32.7 (32.5–32.9) | 30.6 (30.3–30.8) | 24.5 (24.3–24.6) | 23 (22.9–23.1) | 56.5 (56.2–56.8) | 53.7 (53.4–54) | 41.8 (41.7–41.9) | 40.8 (40.6–40.9) |
| 0 | | 32.2 (32–32.5) | 30.2 (30–30.4) | 24.5 (24.4–24.6) | 23 (22.9–23.2) | 56.6 (56.3–56.9) | 53.7 (53.4–54) | 41.9 (41.8–42.1) | 40.8 (40.7–40.9) |
| 1 | | 31.6 (31.4–31.8) | 29.6 (29.4–29.8) | 24.4 (24.2–24.5) | 23 (22.8–23.1) | 56.4 (56.1–56.8) | 53.7 (53.4–54) | 42.1 (41.9–42.2) | 40.9 (40.8–41.1) |
| 2 | | 30.8 (30.5–31) | 28.9 (28.7–29.1) | 24.2 (24–24.3) | 22.8 (22.7–22.9) | 56.3 (56–56.6) | 53.5 (53.2–53.8) | 42.1 (42–42.3) | 40.9 (40.8–41.1) |
| 3 | | 29.9 (29.7–30.1) | 28 (27.8–28.3) | 23.9 (23.7–24) | 22.6 (22.4–22.7) | 56.2 (55.8–56.5) | 53.2 (52.9–53.5) | 42.2 (42–42.3) | 41 (40.8–41.1) |

**eTable 22. Estimates for marginal predictions of annual earnings among parents exposed to child's any mental disorder (F00–F99) and unexposed parents stratified by the child's age at diagnosis**

| | | Finland | | | | Denmark | | | |
|---|---|---|---|---|---|---|---|---|---|
| | | Men | | Women | | Men | | Women | |
| | | Unexposed | Exposed | Unexposed | Exposed | Unexposed | Exposed | Unexposed | Exposed |
| Years since diagnosis | Age group | Marg. pred. (95% CI) | Marg. pred. (95% CI) | Marg. pred. (95% CI) | Marg. pred. (95% CI) | Marg. pred. (95% CI) | Marg. pred. (95% CI) | Marg. pred. (95% CI) | Marg. pred. (95% CI) |
| | | Unexposed | Exposed | Unexposed | Exposed | Unexposed | Exposed | Unexposed | Exposed |
| 4 | | 28.9 (28.7–29.2) | 27.1 (26.8–27.3) | 23.5 (23.4–23.7) | 22.2 (22.1–22.4) | 55.8 (55.4–56.1) | 52.8 (52.5–53.1) | 42.2 (42.1–42.4) | 41 (40.9–41.2) |
| 5 | | 27.7 (27.4–28) | 26 (25.7–26.2) | 23 (22.8–23.1) | 21.8 (21.7–22) | 55 (54.7–55.4) | 52.4 (52.1–52.7) | 42.1 (42–42.3) | 41 (40.8–41.1) |

Marg. pred. = Marginal predictions in thousand euros (€)

**eTable 23. Estimates for marginal predictions of annual disposable income among parents exposed to child's any mental disorder (F00–F99) and unexposed parents stratified by the child's age at diagnosis**

| | | Finland | | | | Denmark | | | |
|---|---|---|---|---|---|---|---|---|---|
| | | Men | | Women | | Men | | Women | |
| | | Unexposed | Exposed | Unexposed | Exposed | Unexposed | Exposed | Unexposed | Exposed |
| Years since diagnosis | Age group | Marg. pred. (95% CI) | Marg. pred. (95% CI) | Marg. pred. (95% CI) | Marg. pred. (95% CI) | Marg. pred. (95% CI) | Marg. pred. (95% CI) | Marg. pred. (95% CI) | Marg. pred. (95% CI) |
| | | Unexposed | Exposed | Unexposed | Exposed | Unexposed | Exposed | Unexposed | Exposed |
| -5 | 1–4 years | 22.4 (22.3–22.5) | 21.6 (21.5–21.8) | 16.7 (16.6–16.8) | 16 (15.9–16.1) | 27.4 (27.1–27.7) | 25.9 (25.6–26.2) | 22.5 (22.3–22.7) | 20.9 (20.6–21.1) |
| -4 | | 23.7 (23.6–23.9) | 22.9 (22.7–23) | 17.1 (17–17.2) | 16.4 (16.3–16.5) | 28.9 (28.6–29.2) | 27.1 (26.8–27.4) | 23.6 (23.4–23.8) | 22 (21.8–22.2) |
| -3 | | 24.8 (24.7–25) | 23.9 (23.7–24) | 17.5 (17.4–17.6) | 16.8 (16.7–16.9) | 30.2 (29.9–30.6) | 28.1 (27.8–28.4) | 25.3 (25.1–25.5) | 23.6 (23.4–23.8) |
| -2 | | 25.9 (25.7–26) | 24.8 (24.6–24.9) | 18.5 (18.4–18.5) | 17.5 (17.5–17.6) | 31.4 (31–31.8) | 29.2 (28.9–29.6) | 27 (26.7–27.2) | 25.1 (24.9–25.3) |
| -1 | | 26.9 (26.8–27.1) | 25.7 (25.6–25.9) | 19.8 (19.7–19.9) | 18.7 (18.6–18.8) | 32.6 (32.3–33) | 30.3 (29.9–30.6) | 28.1 (27.9–28.3) | 26.1 (25.9–26.3) |
| 0 | | 27.8 (27.7–28) | 26.5 (26.4–26.7) | 21.1 (21–21.2) | 19.9 (19.8–20) | 33.7 (33.3–34.1) | 31.2 (30.8–31.5) | 29.2 (28.9–29.4) | 27.1 (26.9–27.4) |
| 1 | | 28.7 (28.5–28.9) | 27.2 (27.1–27.4) | 22.4 (22.3–22.5) | 21.2 (21.1–21.3) | 34.9 (34.5–35.3) | 32.1 (31.7–32.5) | 30.1 (29.9–30.4) | 28.2 (27.9–28.4) |
| 2 | | 29.5 (29.4–29.7) | 27.9 (27.8–28.1) | 23.4 (23.3–23.5) | 22.3 (22.2–22.4) | 35.7 (35.3–36.2) | 33 (32.6–33.4) | 31.1 (30.9–31.4) | 29 (28.8–29.3) |
| 3 | | 30.3 (30.1–30.5) | 28.5 (28.4–28.7) | 24.3 (24.2–24.4) | 23.2 (23.1–23.3) | 36.9 (36.5–37.4) | 33.7 (33.3–34.1) | 31.9 (31.6–32.2) | 29.7 (29.4–29.9) |

**eTable 23. Estimates for marginal predictions of annual disposable income among parents exposed to child's any mental disorder (F00–F99) and unexposed parents stratified by the child's age at diagnosis**

| | | Finland | | | | Denmark | | | |
|---|---|---|---|---|---|---|---|---|---|
| | | Men | | Women | | Men | | Women | |
| | | Unexposed | Exposed | Unexposed | Exposed | Unexposed | Exposed | Unexposed | Exposed |
| Years since diagnosis | Age group | Marg. pred. (95% CI) | Marg. pred. (95% CI) | Marg. pred. (95% CI) | Marg. pred. (95% CI) | Marg. pred. (95% CI) | Marg. pred. (95% CI) | Marg. pred. (95% CI) | Marg. pred. (95% CI) |
| | | Unexposed | Exposed | Unexposed | Exposed | Unexposed | Exposed | Unexposed | Exposed |
| 4 | | 31.1 (30.9–31.3) | 29.3 (29.1–29.5) | 25.2 (25.1–25.3) | 24 (23.9–24.1) | 38.2 (37.7–38.7) | 34.6 (34.1–35) | 32.6 (32.3–32.9) | 30.3 (30–30.6) |
| 5 | | 31.9 (31.7–32.1) | 29.9 (29.7–30.1) | 26.1 (26–26.2) | 24.7 (24.6–24.9) | 39.1 (38.6–39.7) | 35.2 (34.7–35.7) | 33.5 (33.1–33.8) | 30.8 (30.5–31.1) |
| -5 | 5–9 years | 24.8 (24.7–24.9) | 23.5 (23.4–23.6) | 18.1 (18–18.2) | 17.8 (17.7–17.8) | 30.1 (29.9–30.3) | 28.8 (28.6–28.9) | 26.4 (26.3–26.5) | 25.8 (25.7–25.9) |
| -4 | | 25.7 (25.6–25.8) | 24.4 (24.3–24.5) | 19.3 (19.2–19.4) | 18.9 (18.8–19) | 31 (30.8–31.2) | 29.6 (29.5–29.7) | 27.5 (27.4–27.6) | 26.9 (26.8–27) |
| -3 | | 26.4 (26.3–26.5) | 25 (24.9–25.1) | 20.6 (20.5–20.7) | 20.2 (20.1–20.2) | 31.9 (31.8–32.1) | 30.4 (30.3–30.6) | 28.5 (28.4–28.6) | 27.7 (27.6–27.8) |
| -2 | | 27.2 (27.1–27.3) | 25.7 (25.6–25.8) | 21.6 (21.5–21.7) | 21.1 (21.1–21.2) | 33 (32.9–33.2) | 31.3 (31.2–31.5) | 29.3 (29.2–29.4) | 28.4 (28.3–28.5) |
| -1 | | 28 (27.9–28.1) | 26.4 (26.3–26.5) | 22.5 (22.5–22.6) | 22 (21.9–22.1) | 33.9 (33.7–34.1) | 32.2 (32–32.3) | 30.1 (30–30.2) | 29.1 (29–29.2) |
| 0 | | 28.8 (28.7–28.9) | 27.1 (27–27.3) | 23.4 (23.3–23.5) | 22.8 (22.7–22.9) | 34.9 (34.7–35.1) | 33 (32.8–33.1) | 30.8 (30.7–30.9) | 29.6 (29.5–29.7) |
| 1 | | 29.5 (29.4–29.6) | 27.7 (27.6–27.8) | 24.2 (24.1–24.2) | 23.6 (23.5–23.6) | 35.9 (35.7–36.1) | 33.7 (33.5–33.9) | 31.5 (31.3–31.6) | 30.2 (30.1–30.4) |

**eTable 23. Estimates for marginal predictions of annual disposable income among parents exposed to child's any mental disorder (F00–F99) and unexposed parents stratified by the child's age at diagnosis**

| | | Finland | | | | Denmark | | | |
|---|---|---|---|---|---|---|---|---|---|
| | | Men | | Women | | Men | | Women | |
| | | Unexposed | Exposed | Unexposed | Exposed | Unexposed | Exposed | Unexposed | Exposed |
| Years since diagnosis | Age group | Marg. pred. (95% CI) | Marg. pred. (95% CI) | Marg. pred. (95% CI) | Marg. pred. (95% CI) | Marg. pred. (95% CI) | Marg. pred. (95% CI) | Marg. pred. (95% CI) | Marg. pred. (95% CI) |
| | | Unexposed | Exposed | Unexposed | Exposed | Unexposed | Exposed | Unexposed | Exposed |
| 2 | | 30.2 (30.1–30.3) | 28.4 (28.3–28.5) | 24.9 (24.8–25) | 24.2 (24.2–24.3) | 36.7 (36.5–36.9) | 34.4 (34.2–34.6) | 32.1 (32–32.2) | 30.7 (30.6–30.8) |
| 3 | | 30.9 (30.8–31) | 28.9 (28.8–29) | 25.7 (25.6–25.8) | 25 (24.9–25.1) | 37.5 (37.3–37.7) | 35 (34.8–35.2) | 32.8 (32.6–32.9) | 31.3 (31.1–31.4) |
| 4 | | 31.5 (31.3–31.6) | 29.5 (29.4–29.6) | 26.4 (26.3–26.5) | 25.7 (25.6–25.8) | 38.4 (38.2–38.6) | 35.9 (35.7–36.1) | 33.5 (33.3–33.6) | 31.8 (31.7–31.9) |
| 5 | | 32.1 (32–32.3) | 30.1 (29.9–30.2) | 27.1 (27–27.2) | 26.4 (26.3–26.5) | 39.2 (39–39.5) | 36.7 (36.5–36.9) | 34.1 (34–34.3) | 32.3 (32.2–32.5) |
| -5 | 10–14 years | 27.4 (27.3–27.5) | 26.1 (26–26.3) | 22.2 (22.1–22.3) | 21.9 (21.9–22) | 32.9 (32.7–33) | 31.5 (31.3–31.6) | 29.2 (29.1–29.3) | 28.6 (28.5–28.7) |
| -4 | | 28 (27.8–28.1) | 26.7 (26.5–26.8) | 22.9 (22.8–23) | 22.6 (22.6–22.7) | 33.7 (33.6–33.9) | 32.3 (32.2–32.5) | 29.8 (29.7–29.9) | 29.2 (29.2–29.3) |
| -3 | | 28.6 (28.4–28.7) | 27.1 (27–27.2) | 23.5 (23.5–23.6) | 23.3 (23.2–23.4) | 34.6 (34.4–34.7) | 33.1 (32.9–33.2) | 30.5 (30.4–30.6) | 29.8 (29.7–29.9) |
| -2 | | 29.2 (29–29.3) | 27.7 (27.6–27.8) | 24.2 (24.1–24.3) | 23.9 (23.8–24) | 35.5 (35.3–35.7) | 33.8 (33.7–34) | 31.1 (31–31.2) | 30.4 (30.3–30.5) |
| -1 | | 29.8 (29.6–29.9) | 28.2 (28.1–28.4) | 24.8 (24.7–24.9) | 24.5 (24.5–24.6) | 36.3 (36.1–36.5) | 34.5 (34.4–34.7) | 31.7 (31.6–31.8) | 31 (30.9–31.1) |

**eTable 23. Estimates for marginal predictions of annual disposable income among parents exposed to child's any mental disorder (F00–F99) and unexposed parents stratified by the child's age at diagnosis**

| | | Finland | | | | Denmark | | | |
|---|---|---|---|---|---|---|---|---|---|
| | | Men | | Women | | Men | | Women | |
| | | Unexposed | Exposed | Unexposed | Exposed | Unexposed | Exposed | Unexposed | Exposed |
| Years since diagnosis | Age group | Marg. pred. (95% CI) | Marg. pred. (95% CI) | Marg. pred. (95% CI) | Marg. pred. (95% CI) | Marg. pred. (95% CI) | Marg. pred. (95% CI) | Marg. pred. (95% CI) | Marg. pred. (95% CI) |
| | | Unexposed | Exposed | Unexposed | Exposed | Unexposed | Exposed | Unexposed | Exposed |
| 0 | | 30.3 (30.2–30.5) | 28.7 (28.5–28.8) | 25.3 (25.2–25.4) | 25 (24.9–25.1) | 37.3 (37.1–37.5) | 35.3 (35.2–35.5) | 32.3 (32.2–32.4) | 31.5 (31.4–31.6) |
| 1 | | 30.9 (30.7–31) | 29.2 (29–29.3) | 25.8 (25.7–25.9) | 25.4 (25.3–25.5) | 38 (37.8–38.2) | 36.1 (35.9–36.3) | 32.9 (32.8–33) | 32 (31.9–32.1) |
| 2 | | 31.4 (31.2–31.5) | 29.6 (29.4–29.7) | 26.2 (26.1–26.3) | 25.7 (25.7–25.8) | 38.9 (38.7–39.1) | 36.7 (36.5–36.9) | 33.5 (33.4–33.6) | 32.4 (32.3–32.5) |
| 3 | | 31.9 (31.7–32) | 30 (29.8–30.1) | 26.3 (26.2–26.4) | 25.8 (25.8–25.9) | 39.7 (39.5–39.9) | 37.4 (37.2–37.6) | 33.9 (33.7–34) | 32.7 (32.6–32.8) |
| 4 | | 32.3 (32.1–32.5) | 30.3 (30.1–30.5) | 26.4 (26.3–26.5) | 25.9 (25.8–26) | 40.5 (40.2–40.7) | 38 (37.8–38.3) | 34.1 (33.9–34.2) | 32.8 (32.6–32.9) |
| 5 | | 32.7 (32.5–32.9) | 30.6 (30.4–30.7) | 26.5 (26.4–26.6) | 25.9 (25.8–26) | 41.2 (40.9–41.4) | 38.8 (38.6–39) | 34.3 (34.1–34.4) | 32.8 (32.7–33) |
| -5 | 15–19 years | 28 (27.9–28.1) | 26.8 (26.7–26.9) | 23 (22.9–23) | 22.7 (22.7–22.8) | 33.8 (33.7–34) | 32.4 (32.2–32.5) | 29.5 (29.4–29.5) | 29 (28.9–29.1) |
| -4 | | 28.5 (28.4–28.6) | 27.2 (27.1–27.3) | 23.5 (23.4–23.5) | 23.2 (23.2–23.3) | 34.6 (34.5–34.8) | 33 (32.8–33.1) | 30 (30–30.1) | 29.5 (29.4–29.6) |
| -3 | | 28.9 (28.8–29.1) | 27.6 (27.5–27.8) | 23.8 (23.8–23.9) | 23.6 (23.5–23.7) | 35.3 (35.1–35.4) | 33.6 (33.5–33.8) | 30.6 (30.5–30.7) | 30 (29.9–30) |

**eTable 23. Estimates for marginal predictions of annual disposable income among parents exposed to child's any mental disorder (F00–F99) and unexposed parents stratified by the child's age at diagnosis**

| | | Finland | | | | Denmark | | | |
|---|---|---|---|---|---|---|---|---|---|
| | | Men | | Women | | Men | | Women | |
| | | Unexposed | Exposed | Unexposed | Exposed | Unexposed | Exposed | Unexposed | Exposed |
| Years since diagnosis | Age group | Marg. pred. (95% CI) | Marg. pred. (95% CI) | Marg. pred. (95% CI) | Marg. pred. (95% CI) | Marg. pred. (95% CI) | Marg. pred. (95% CI) | Marg. pred. (95% CI) | Marg. pred. (95% CI) |
| | | Unexposed | Exposed | Unexposed | Exposed | Unexposed | Exposed | Unexposed | Exposed |
| -2 | | 29.4 (29.2–29.5) | 28.1 (28–28.2) | 24 (23.9–24.1) | 23.8 (23.8–23.9) | 36 (35.9–36.2) | 34.3 (34.2–34.4) | 31 (30.9–31.1) | 30.3 (30.3–30.4) |
| -1 | | 29.8 (29.7–30) | 28.5 (28.4–28.6) | 24.1 (24–24.2) | 23.9 (23.9–24) | 36.8 (36.7–37) | 34.9 (34.8–35.1) | 31.3 (31.2–31.3) | 30.6 (30.5–30.7) |
| 0 | | 30.3 (30.1–30.4) | 28.7 (28.6–28.9) | 24.1 (24.1–24.2) | 23.9 (23.9–24) | 37.5 (37.3–37.6) | 35.5 (35.4–35.7) | 31.4 (31.3–31.5) | 30.6 (30.5–30.7) |
| 1 | | 30.6 (30.4–30.7) | 29.1 (29–29.2) | 24.1 (24.1–24.2) | 23.9 (23.9–24) | 38.1 (37.9–38.3) | 36.2 (36–36.3) | 31.6 (31.5–31.7) | 30.7 (30.6–30.7) |
| 2 | | 30.9 (30.7–31) | 29.4 (29.3–29.5) | 24.1 (24.1–24.2) | 23.9 (23.9–24) | 38.7 (38.5–38.9) | 36.7 (36.5–36.8) | 31.6 (31.5–31.7) | 30.6 (30.5–30.7) |
| 3 | | 31.1 (31–31.3) | 29.6 (29.5–29.8) | 24.3 (24.3–24.4) | 24.1 (24–24.1) | 39.2 (39–39.4) | 37.2 (37–37.3) | 31.7 (31.6–31.8) | 30.7 (30.6–30.8) |
| 4 | | 31.4 (31.3–31.6) | 29.8 (29.7–30) | 24.5 (24.4–24.6) | 24.2 (24.2–24.3) | 39.7 (39.5–39.9) | 37.6 (37.4–37.8) | 32 (31.9–32.1) | 30.8 (30.7–30.9) |
| 5 | | 31.7 (31.5–31.8) | 30 (29.9–30.2) | 24.8 (24.7–24.8) | 24.4 (24.3–24.5) | 40.2 (39.9–40.4) | 37.8 (37.7–38) | 32.2 (32.1–32.3) | 31.1 (31–31.2) |
| -5 | 20–25 years | 28 (27.9–28.1) | 26.8 (26.6–26.9) | 22 (21.9–22) | 21.6 (21.5–21.7) | 34.1 (33.9–34.2) | 32.6 (32.4–32.7) | 28.1 (28–28.2) | 27.4 (27.3–27.5) |

**eTable 23. Estimates for marginal predictions of annual disposable income among parents exposed to child's any mental disorder (F00–F99) and unexposed parents stratified by the child’s age at diagnosis**

| | | Finland | | | | Denmark | | | |
|---|---|---|---|---|---|---|---|---|---|
| | | Men | | Women | | Men | | Women | |
| | | Unexposed | Exposed | Unexposed | Exposed | Unexposed | Exposed | Unexposed | Exposed |
| Years since diagnosis | Age group | Marg. pred. (95% CI) | Marg. pred. (95% CI) | Marg. pred. (95% CI) | Marg. pred. (95% CI) | Marg. pred. (95% CI) | Marg. pred. (95% CI) | Marg. pred. (95% CI) | Marg. pred. (95% CI) |
| | | Unexposed | Exposed | Unexposed | Exposed | Unexposed | Exposed | Unexposed | Exposed |
| -4 | | 28.3 (28.1–28.4) | 27 (26.8–27.1) | 21.9 (21.9–22) | 21.6 (21.5–21.6) | 34.7 (34.5–34.9) | 33.1 (32.9–33.2) | 28.2 (28.1–28.3) | 27.5 (27.4–27.6) |
| -3 | | 28.5 (28.4–28.6) | 27.1 (27–27.2) | 21.9 (21.9–22) | 21.5 (21.4–21.6) | 35.3 (35.1–35.4) | 33.6 (33.5–33.8) | 28.3 (28.2–28.4) | 27.5 (27.4–27.6) |
| -2 | | 28.7 (28.6–28.9) | 27.3 (27.2–27.5) | 22 (22–22.1) | 21.6 (21.5–21.7) | 35.8 (35.6–35.9) | 34 (33.9–34.2) | 28.4 (28.3–28.5) | 27.6 (27.5–27.7) |
| -1 | | 29.1 (28.9–29.2) | 27.5 (27.4–27.6) | 22.2 (22.1–22.2) | 21.7 (21.7–21.8) | 36.3 (36.1–36.4) | 34.4 (34.3–34.6) | 28.6 (28.5–28.7) | 27.8 (27.7–27.9) |
| 0 | | 29.1 (29–29.3) | 27.7 (27.6–27.9) | 22.3 (22.3–22.4) | 21.9 (21.8–21.9) | 36.7 (36.6–36.9) | 34.8 (34.6–35) | 28.9 (28.8–29) | 28 (27.9–28.1) |
| 1 | | 29.3 (29.2–29.5) | 27.9 (27.8–28) | 22.5 (22.4–22.6) | 22 (21.9–22.1) | 37 (36.9–37.2) | 35.2 (35–35.3) | 29.1 (29–29.2) | 28.2 (28.1–28.3) |
| 2 | | 29.4 (29.3–29.6) | 28 (27.9–28.2) | 22.7 (22.6–22.7) | 22.2 (22.1–22.2) | 37.4 (37.2–37.6) | 35.5 (35.3–35.6) | 29.4 (29.3–29.5) | 28.4 (28.3–28.5) |
| 3 | | 29.5 (29.4–29.7) | 28.1 (28–28.3) | 22.8 (22.7–22.9) | 22.3 (22.2–22.4) | 37.7 (37.6–37.9) | 35.7 (35.5–35.8) | 29.6 (29.5–29.7) | 28.6 (28.6–28.7) |
| 4 | | 29.7 (29.5–29.8) | 28.2 (28.1–28.4) | 23 (22.9–23.1) | 22.5 (22.4–22.6) | 37.9 (37.7–38.1) | 35.8 (35.6–35.9) | 29.9 (29.8–30) | 28.9 (28.8–29) |

**eTable 23. Estimates for marginal predictions of annual disposable income among parents exposed to child's any mental disorder (F00–F99) and unexposed parents stratified by the child's age at diagnosis**

| | | Finland | | | | Denmark | | | |
|---|---|---|---|---|---|---|---|---|---|
| | | Men | | Women | | Men | | Women | |
| | | Unexposed | Exposed | Unexposed | Exposed | Unexposed | Exposed | Unexposed | Exposed |
| Years since diagnosis | Age group | Marg. pred. (95% CI) | Marg. pred. (95% CI) | Marg. pred. (95% CI) | Marg. pred. (95% CI) | Marg. pred. (95% CI) | Marg. pred. (95% CI) | Marg. pred. (95% CI) | Marg. pred. (95% CI) |
| | | Unexposed | Exposed | Unexposed | Exposed | Unexposed | Exposed | Unexposed | Exposed |
| 5 | | 29.7 (29.6–29.9) | 28.3 (28.2–28.5) | 23.2 (23.1–23.3) | 22.6 (22.5–22.7) | 37.9 (37.7–38.1) | 35.9 (35.7–36.1) | 30 (29.9–30.2) | 29.1 (29–29.2) |

Marg. pred. = Marginal predictions in thousand euros (€)

**eTable 24. Estimates for marginal predictions of annual social welfare benefits among parents exposed to child's any mental disorder (F00–F99) and unexposed parents stratified by the child's age at diagnosis**

| | | Finland | | | | Denmark | | | |
|---|---|---|---|---|---|---|---|---|---|
| | | Men | | Women | | Men | | Women | |
| | | Unexposed | Exposed | Unexposed | Exposed | Unexposed | Exposed | Unexposed | Exposed |
| Years since diagnosis | Age group | Marg. pred. (95% CI) | Marg. pred. (95% CI) | Marg. pred. (95% CI) | Marg. pred. (95% CI) | Marg. pred. (95% CI) | Marg. pred. (95% CI) | Marg. pred. (95% CI) | Marg. pred. (95% CI) |
| | | Unexposed | Exposed | Unexposed | Exposed | Unexposed | Exposed | Unexposed | Exposed |
| -5 | 1–4 years | 2.3 (2.3–2.4) | 2.7 (2.7–2.8) | 6.2 (6.2–6.3) | 6.4 (6.3–6.5) | 3.7 (3.5–3.8) | 4.5 (4.4–4.7) | 8.4 (8.2–8.6) | 8.9 (8.7–9.1) |
| -4 | | 2.7 (2.6–2.7) | 3.1 (3–3.2) | 9.4 (9.3–9.5) | 9.5 (9.4–9.6) | 3.9 (3.7–4) | 4.8 (4.7–5) | 11.5 (11.3–11.7) | 12.3 (12.1–12.5) |
| -3 | | 2.9 (2.8–2.9) | 3.4 (3.4–3.5) | 10.9 (10.8–10.9) | 11.3 (11.3–11.4) | 4 (3.8–4.1) | 5 (4.8–5.1) | 12.6 (12.4–12.8) | 13.7 (13.5–13.9) |
| -2 | | 2.8 (2.8–2.9) | 3.4 (3.4–3.5) | 10.4 (10.4–10.5) | 11.2 (11.2–11.3) | 4 (3.8–4.2) | 5 (4.8–5.1) | 12.6 (12.3–12.8) | 14 (13.8–14.2) |
| -1 | | 2.7 (2.7–2.8) | 3.4 (3.4–3.5) | 9.9 (9.8–10) | 11 (10.9–11) | 3.9 (3.8–4.1) | 5 (4.8–5.2) | 12.3 (12.1–12.5) | 14 (13.7–14.2) |
| 0 | | 2.7 (2.6–2.7) | 3.4 (3.3–3.4) | 9.2 (9.1–9.3) | 10.2 (10.1–10.3) | 3.9 (3.7–4) | 5.1 (4.9–5.2) | 11.9 (11.6–12.1) | 13.7 (13.5–13.9) |
| 1 | | 2.7 (2.6–2.7) | 3.4 (3.3–3.5) | 8.8 (8.7–8.8) | 9.7 (9.7–9.8) | 3.9 (3.7–4.1) | 5.1 (4.9–5.3) | 11.4 (11.2–11.6) | 13.4 (13.1–13.6) |
| 2 | | 2.7 (2.6–2.7) | 3.5 (3.4–3.5) | 8.5 (8.4–8.5) | 9.5 (9.4–9.5) | 3.8 (3.7–4) | 5.2 (5–5.4) | 11.1 (10.9–11.3) | 13.3 (13–13.5) |
| 3 | | 2.7 (2.6–2.7) | 3.5 (3.4–3.6) | 8.2 (8.2–8.3) | 9.4 (9.3–9.5) | 3.8 (3.6–4) | 5.3 (5–5.5) | 10.7 (10.4–10.9) | 12.9 (12.7–13.2) |
| 4 | | 2.8 (2.7–2.9) | 3.6 (3.6–3.7) | 8.1 (8–8.2) | 9.4 (9.3–9.4) | 3.8 (3.6–4) | 5.3 (5.1–5.5) | 10.4 (10.1–10.6) | 12.7 (12.4–13) |

**eTable 24. Estimates for marginal predictions of annual social welfare benefits among parents exposed to child's any mental disorder (F00–F99) and unexposed parents stratified by the child's age at diagnosis**

| | | Finland | | | | Denmark | | | |
|---|---|---|---|---|---|---|---|---|---|
| | | Men | | Women | | Men | | Women | |
| | | Unexposed | Exposed | Unexposed | Exposed | Unexposed | Exposed | Unexposed | Exposed |
| Years since diagnosis | Age group | Marg. pred. (95% CI) | Marg. pred. (95% CI) | Marg. pred. (95% CI) | Marg. pred. (95% CI) | Marg. pred. (95% CI) | Marg. pred. (95% CI) | Marg. pred. (95% CI) | Marg. pred. (95% CI) |
| | | Unexposed | Exposed | Unexposed | Exposed | Unexposed | Exposed | Unexposed | Exposed |
| 5 | | 2.9 (2.8–3) | 3.8 (3.7–3.8) | 7.9 (7.8–8) | 9.2 (9.2–9.3) | 4 (3.8–4.2) | 5.4 (5.2–5.7) | 10.1 (9.9–10.4) | 12.5 (12.2–12.8) |
| -5 | 5–9 years | 2.7 (2.6–2.7) | 3.2 (3.2–3.2) | 10.3 (10.2–10.3) | 10.7 (10.7–10.8) | 4.1 (4–4.2) | 4.4 (4.3–4.4) | 12.6 (12.5–12.7) | 13.7 (13.5–13.8) |
| -4 | | 2.7 (2.6–2.7) | 3.2 (3.2–3.2) | 9.3 (9.2–9.3) | 10 (10–10.1) | 4.1 (4–4.2) | 4.5 (4.4–4.5) | 12.1 (12–12.2) | 13.3 (13.2–13.4) |
| -3 | | 2.8 (2.7–2.8) | 3.3 (3.3–3.4) | 8.9 (8.8–8.9) | 9.9 (9.8–9.9) | 4.1 (4–4.2) | 4.5 (4.4–4.5) | 11.8 (11.7–11.9) | 13.1 (13–13.2) |
| -2 | | 2.7 (2.7–2.8) | 3.3 (3.3–3.4) | 8.5 (8.5–8.6) | 9.6 (9.6–9.7) | 4.1 (4.1–4.2) | 4.5 (4.5–4.6) | 11.5 (11.4–11.6) | 13 (12.8–13.1) |
| -1 | | 2.7 (2.7–2.8) | 3.3 (3.3–3.4) | 8.3 (8.3–8.4) | 9.6 (9.5–9.6) | 4.2 (4.1–4.2) | 4.6 (4.5–4.7) | 11.2 (11.1–11.3) | 12.7 (12.6–12.8) |
| 0 | | 2.7 (2.7–2.8) | 3.4 (3.3–3.4) | 8.1 (8.1–8.2) | 9.4 (9.4–9.5) | 4.1 (4–4.2) | 4.6 (4.5–4.7) | 10.9 (10.8–11) | 12.7 (12.5–12.8) |
| 1 | | 2.8 (2.7–2.8) | 3.5 (3.4–3.5) | 7.9 (7.9–8) | 9.3 (9.3–9.4) | 4.2 (4.1–4.2) | 4.8 (4.7–4.8) | 10.7 (10.6–10.8) | 12.5 (12.4–12.6) |
| 2 | | 2.8 (2.8–2.9) | 3.6 (3.5–3.6) | 7.7 (7.6–7.7) | 9.1 (9.1–9.2) | 4.2 (4.1–4.3) | 4.9 (4.8–5) | 10.5 (10.4–10.6) | 12.3 (12.2–12.4) |
| 3 | | 2.9 (2.9–3) | 3.7 (3.7–3.8) | 7.5 (7.4–7.5) | 9 (8.9–9) | 4.3 (4.2–4.4) | 5 (4.9–5.1) | 10.2 (10.1–10.3) | 12.1 (12–12.2) |

**eTable 24. Estimates for marginal predictions of annual social welfare benefits among parents exposed to child's any mental disorder (F00–F99) and unexposed parents stratified by the child's age at diagnosis**

| | | Finland | | | | Denmark | | | |
|---|---|---|---|---|---|---|---|---|---|
| | | Men | | Women | | Men | | Women | |
| | | Unexposed | Exposed | Unexposed | Exposed | Unexposed | Exposed | Unexposed | Exposed |
| Years since diagnosis | Age group | Marg. pred. (95% CI) | Marg. pred. (95% CI) | Marg. pred. (95% CI) | Marg. pred. (95% CI) | Marg. pred. (95% CI) | Marg. pred. (95% CI) | Marg. pred. (95% CI) | Marg. pred. (95% CI) |
| | | Unexposed | Exposed | Unexposed | Exposed | Unexposed | Exposed | Unexposed | Exposed |
| 4 | | 3.1 (3–3.1) | 3.9 (3.8–3.9) | 7.3 (7.2–7.4) | 8.8 (8.8–8.9) | 4.4 (4.3–4.5) | 5.2 (5.1–5.3) | 10 (9.9–10.1) | 11.9 (11.8–12) |
| 5 | | 3.2 (3.1–3.3) | 4 (4–4.1) | 7.1 (7.1–7.2) | 8.7 (8.6–8.8) | 4.5 (4.4–4.6) | 5.3 (5.2–5.4) | 9.8 (9.7–9.9) | 11.8 (11.7–11.9) |
| -5 | 10–14 years | 2.7 (2.6–2.7) | 3.2 (3.2–3.3) | 7.6 (7.5–7.7) | 8.5 (8.4–8.5) | 3.7 (3.7–3.8) | 4.1 (4–4.2) | 10.3 (10.2–10.4) | 11.5 (11.4–11.6) |
| -4 | | 2.8 (2.7–2.8) | 3.4 (3.3–3.4) | 7.5 (7.4–7.5) | 8.5 (8.4–8.5) | 3.8 (3.7–3.8) | 4.1 (4.1–4.2) | 10.1 (10–10.2) | 11.3 (11.2–11.4) |
| -3 | | 2.9 (2.9–3) | 3.6 (3.5–3.6) | 7.4 (7.3–7.4) | 8.5 (8.4–8.5) | 3.8 (3.8–3.9) | 4.2 (4.1–4.3) | 9.9 (9.8–10) | 11.2 (11.1–11.3) |
| -2 | | 3 (3–3.1) | 3.6 (3.6–3.7) | 7.2 (7.1–7.2) | 8.3 (8.3–8.4) | 3.9 (3.8–4) | 4.3 (4.2–4.4) | 9.7 (9.6–9.8) | 11.1 (11–11.2) |
| -1 | | 3.1 (3.1–3.2) | 3.7 (3.7–3.8) | 7 (6.9–7) | 8.2 (8.1–8.3) | 4 (3.9–4) | 4.4 (4.3–4.5) | 9.5 (9.4–9.6) | 11 (10.9–11.1) |
| 0 | | 3.2 (3.1–3.3) | 3.9 (3.8–4) | 6.8 (6.7–6.8) | 8.1 (8.1–8.2) | 4 (3.9–4.1) | 4.6 (4.5–4.7) | 9.4 (9.3–9.5) | 11 (10.9–11.1) |
| 1 | | 3.3 (3.3–3.4) | 4.1 (4–4.2) | 6.5 (6.5–6.6) | 8 (7.9–8.1) | 4.1 (4–4.2) | 4.7 (4.6–4.8) | 9.2 (9.1–9.3) | 10.9 (10.8–11) |
| 2 | | 3.5 (3.4–3.6) | 4.3 (4.2–4.4) | 6.3 (6.2–6.4) | 7.7 (7.7–7.8) | 4.2 (4.1–4.2) | 4.9 (4.8–5) | 9 (8.9–9.1) | 10.7 (10.6–10.8) |
| 3 | | 3.7 (3.6–3.8) | 4.5 (4.4–4.5) | 5.8 (5.7–5.9) | 7.3 (7.2–7.3) | 4.3 (4.2–4.4) | 5 (4.9–5.1) | 8.7 (8.6–8.8) | 10.5 (10.3–10.6) |

**eTable 24. Estimates for marginal predictions of annual social welfare benefits among parents exposed to child's any mental disorder (F00–F99) and unexposed parents stratified by the child's age at diagnosis**

| | | Finland | | | | Denmark | | | |
|---|---|---|---|---|---|---|---|---|---|
| | | Men | | Women | | Men | | Women | |
| | | Unexposed | Exposed | Unexposed | Exposed | Unexposed | Exposed | Unexposed | Exposed |
| Years since diagnosis | Age group | Marg. pred. (95% CI) | Marg. pred. (95% CI) | Marg. pred. (95% CI) | Marg. pred. (95% CI) | Marg. pred. (95% CI) | Marg. pred. (95% CI) | Marg. pred. (95% CI) | Marg. pred. (95% CI) |
| | | Unexposed | Exposed | Unexposed | Exposed | Unexposed | Exposed | Unexposed | Exposed |
| 4 | | 3.9 (3.8–3.9) | 4.7 (4.6–4.7) | 5.3 (5.2–5.4) | 6.8 (6.7–6.9) | 4.4 (4.3–4.4) | 5.1 (5–5.2) | 8.2 (8.1–8.3) | 10 (9.9–10.1) |
| 5 | | 4.1 (4–4.2) | 4.9 (4.8–5) | 4.9 (4.9–5) | 6.4 (6.4–6.5) | 4.5 (4.4–4.6) | 5.3 (5.2–5.4) | 7.7 (7.6–7.8) | 9.6 (9.5–9.7) |
| -5 | 15–19 years | 2.9 (2.8–2.9) | 3.4 (3.4–3.5) | 6.4 (6.4–6.5) | 7.3 (7.3–7.4) | 3.8 (3.8–3.9) | 4.5 (4.4–4.6) | 9 (8.9–9.1) | 10.4 (10.3–10.4) |
| -4 | | 3 (3–3.1) | 3.6 (3.6–3.7) | 6.3 (6.2–6.3) | 7.2 (7.1–7.2) | 3.9 (3.9–4) | 4.7 (4.6–4.7) | 8.9 (8.8–8.9) | 10.3 (10.2–10.3) |
| -3 | | 3.2 (3.2–3.3) | 3.8 (3.8–3.9) | 6.1 (6–6.1) | 7 (7–7.1) | 4.1 (4–4.1) | 4.8 (4.8–4.9) | 8.7 (8.7–8.8) | 10.1 (10.1–10.2) |
| -2 | | 3.4 (3.3–3.4) | 4 (3.9–4) | 5.7 (5.6–5.7) | 6.6 (6.6–6.7) | 4.2 (4.1–4.2) | 4.9 (4.9–5) | 8.5 (8.5–8.6) | 10 (9.9–10.1) |
| -1 | | 3.5 (3.5–3.6) | 4.1 (4.1–4.2) | 5.2 (5.2–5.3) | 6.2 (6.1–6.2) | 4.2 (4.2–4.3) | 5.1 (5–5.1) | 8.2 (8.1–8.2) | 9.7 (9.6–9.8) |
| 0 | | 3.7 (3.6–3.8) | 4.3 (4.3–4.4) | 4.8 (4.7–4.8) | 5.8 (5.8–5.9) | 4.3 (4.3–4.4) | 5.2 (5.1–5.2) | 7.7 (7.6–7.8) | 9.3 (9.2–9.4) |
| 1 | | 3.9 (3.9–4) | 4.6 (4.5–4.6) | 4.4 (4.3–4.4) | 5.4 (5.4–5.5) | 4.4 (4.4–4.5) | 5.3 (5.2–5.4) | 7.3 (7.2–7.4) | 9 (8.9–9.1) |
| 2 | | 4.2 (4.1–4.3) | 4.8 (4.8–4.9) | 4 (4–4.1) | 5.1 (5–5.1) | 4.6 (4.5–4.6) | 5.4 (5.3–5.5) | 6.9 (6.8–7) | 8.5 (8.4–8.6) |
| 3 | | 4.6 (4.5–4.7) | 5.2 (5.1–5.3) | 3.9 (3.9–4) | 4.9 (4.8–5) | 4.7 (4.6–4.8) | 5.6 (5.5–5.7) | 6.5 (6.4–6.6) | 8.2 (8.1–8.3) |
| 4 | | 5.1 (5–5.1) | 5.6 (5.5–5.7) | 4 (3.9–4) | 4.9 (4.9–5) | 4.9 (4.8–5) | 5.8 (5.7–5.9) | 6.4 (6.3–6.5) | 8.2 (8.1–8.2) |

**eTable 24. Estimates for marginal predictions of annual social welfare benefits among parents exposed to child's any mental disorder (F00–F99) and unexposed parents stratified by the child's age at diagnosis**

| | | Finland | | | | Denmark | | | |
|---|---|---|---|---|---|---|---|---|---|
| | | Men | | Women | | Men | | Women | |
| | | Unexposed | Exposed | Unexposed | Exposed | Unexposed | Exposed | Unexposed | Exposed |
| Years since diagnosis | Age group | Marg. pred. (95% CI) | Marg. pred. (95% CI) | Marg. pred. (95% CI) | Marg. pred. (95% CI) | Marg. pred. (95% CI) | Marg. pred. (95% CI) | Marg. pred. (95% CI) | Marg. pred. (95% CI) |
| | | Unexposed | Exposed | Unexposed | Exposed | Unexposed | Exposed | Unexposed | Exposed |
| 5 | | 5.6 (5.5–5.7) | 6.1 (6–6.2) | 4.1 (4–4.1) | 5 (4.9–5) | 5.2 (5.1–5.2) | 6.1 (6–6.2) | 6.4 (6.3–6.5) | 8.2 (8.1–8.3) |
| -5 | 20–25 years | 3.6 (3.5–3.7) | 4 (3.9–4.1) | 4.5 (4.5–4.6) | 5.3 (5.2–5.3) | 3.9 (3.9–4) | 4.8 (4.8–4.9) | 7 (6.9–7) | 8.3 (8.3–8.4) |
| -4 | | 3.9 (3.8–3.9) | 4.3 (4.3–4.4) | 4.2 (4.2–4.2) | 5 (4.9–5) | 4 (4–4.1) | 5 (4.9–5.1) | 6.7 (6.6–6.7) | 8 (8–8.1) |
| -3 | | 4.2 (4.1–4.3) | 4.7 (4.6–4.7) | 3.9 (3.9–4) | 4.7 (4.7–4.8) | 4.2 (4.1–4.3) | 5.2 (5.1–5.2) | 6.3 (6.3–6.4) | 7.8 (7.7–7.8) |
| -2 | | 4.5 (4.5–4.6) | 5 (5–5.1) | 3.9 (3.8–3.9) | 4.6 (4.6–4.7) | 4.3 (4.3–4.4) | 5.4 (5.3–5.5) | 6.1 (6–6.2) | 7.5 (7.5–7.6) |
| -1 | | 4.9 (4.9–5) | 5.4 (5.3–5.5) | 3.9 (3.9–4) | 4.7 (4.6–4.7) | 4.5 (4.5–4.6) | 5.6 (5.5–5.7) | 6.1 (6–6.1) | 7.5 (7.4–7.6) |
| 0 | | 5.4 (5.3–5.5) | 5.8 (5.8–5.9) | 4 (4–4.1) | 4.8 (4.7–4.8) | 4.8 (4.7–4.9) | 5.9 (5.8–5.9) | 6.1 (6–6.2) | 7.6 (7.5–7.7) |
| 1 | | 6 (5.9–6.1) | 6.4 (6.3–6.5) | 4.2 (4.2–4.3) | 4.9 (4.9–5) | 5.1 (5–5.1) | 6.1 (6.1–6.2) | 6.2 (6.1–6.2) | 7.7 (7.6–7.8) |
| 2 | | 6.7 (6.6–6.8) | 7.1 (7–7.2) | 4.5 (4.4–4.5) | 5.2 (5.1–5.2) | 5.4 (5.3–5.5) | 6.4 (6.4–6.5) | 6.3 (6.2–6.4) | 7.9 (7.8–8) |
| 3 | | 7.5 (7.4–7.5) | 7.7 (7.6–7.8) | 4.8 (4.8–4.9) | 5.5 (5.4–5.5) | 5.7 (5.7–5.8) | 6.8 (6.7–6.9) | 6.5 (6.4–6.6) | 8.1 (8–8.2) |
| 4 | | 8.4 (8.2–8.5) | 8.6 (8.5–8.7) | 5.2 (5.2–5.3) | 5.8 (5.7–5.9) | 6.2 (6.1–6.3) | 7.2 (7.1–7.3) | 6.8 (6.7–6.9) | 8.4 (8.3–8.5) |
| 5 | | 9.3 (9.2–9.5) | 9.5 (9.4–9.6) | 5.7 (5.6–5.8) | 6.2 (6.1–6.3) | 6.7 (6.6–6.8) | 7.7 (7.6–7.8) | 7.1 (7–7.2) | 8.8 (8.7–8.9) |

Marg. pred. = Marginal predictions in thousand euros (€)

**eTable 25. Estimates for marginal predictions of annual employment among parents exposed to child's any mental disorder (F00–F99) and unexposed parents stratified by the child's age at diagnosis**

| | | Finland | | | | Denmark | | | |
|---|---|---|---|---|---|---|---|---|---|
| | | Men | | Women | | Men | | Women | |
| | | Unexposed | Exposed | Unexposed | Exposed | Unexposed | Exposed | Unexposed | Exposed |
| Years since diagnosis | Age group | Marg. pred. (95% CI) | Marg. pred. (95% CI) | Marg. pred. (95% CI) | Marg. pred. (95% CI) | Marg. pred. (95% CI) | Marg. pred. (95% CI) | Marg. pred. (95% CI) | Marg. pred. (95% CI) |
| | | Unexposed | Exposed | Unexposed | Exposed | Unexposed | Exposed | Unexposed | Exposed |
| -5 | 1–4 years | 0.79 (0.78–0.79) | 0.75 (0.75–0.76) | 0.63 (0.63–0.64) | 0.58 (0.58–0.59) | 0.68 (0.67–0.69) | 0.64 (0.63–0.65) | 0.57 (0.56–0.58) | 0.5 (0.49–0.51) |
| -4 | | 0.8 (0.8–0.81) | 0.77 (0.76–0.77) | 0.58 (0.58–0.59) | 0.53 (0.53–0.54) | 0.71 (0.7–0.72) | 0.67 (0.66–0.68) | 0.56 (0.55–0.57) | 0.48 (0.47–0.5) |
| -3 | | 0.81 (0.81–0.82) | 0.77 (0.77–0.78) | 0.56 (0.55–0.56) | 0.49 (0.49–0.5) | 0.73 (0.72–0.74) | 0.69 (0.68–0.7) | 0.57 (0.56–0.58) | 0.5 (0.49–0.51) |
| -2 | | 0.82 (0.82–0.83) | 0.78 (0.78–0.79) | 0.58 (0.58–0.59) | 0.5 (0.5–0.51) | 0.73 (0.72–0.74) | 0.69 (0.68–0.7) | 0.6 (0.58–0.61) | 0.51 (0.5–0.53) |
| -1 | | 0.84 (0.83–0.84) | 0.79 (0.79–0.8) | 0.63 (0.62–0.63) | 0.54 (0.53–0.54) | 0.74 (0.73–0.75) | 0.71 (0.7–0.72) | 0.61 (0.6–0.63) | 0.53 (0.52–0.54) |
| 0 | | 0.84 (0.83–0.84) | 0.8 (0.79–0.8) | 0.67 (0.67–0.68) | 0.58 (0.58–0.59) | 0.75 (0.74–0.76) | 0.7 (0.69–0.71) | 0.63 (0.62–0.65) | 0.53 (0.52–0.55) |
| 1 | | 0.84 (0.84–0.85) | 0.8 (0.79–0.8) | 0.7 (0.7–0.71) | 0.62 (0.62–0.63) | 0.74 (0.73–0.75) | 0.7 (0.69–0.71) | 0.65 (0.64–0.66) | 0.55 (0.54–0.56) |
| 2 | | 0.85 (0.84–0.85) | 0.8 (0.8–0.81) | 0.73 (0.73–0.74) | 0.65 (0.65–0.66) | 0.74 (0.73–0.75) | 0.7 (0.69–0.71) | 0.67 (0.66–0.69) | 0.56 (0.55–0.57) |
| 3 | | 0.85 (0.84–0.85) | 0.8 (0.8–0.81) | 0.75 (0.75–0.76) | 0.67 (0.67–0.68) | 0.75 (0.74–0.76) | 0.7 (0.69–0.71) | 0.68 (0.67–0.69) | 0.57 (0.56–0.59) |

**eTable 25. Estimates for marginal predictions of annual employment among parents exposed to child's any mental disorder (F00–F99) and unexposed parents stratified by the child’s age at diagnosis**

| | | Finland | | | | Denmark | | | |
|---|---|---|---|---|---|---|---|---|---|
| | | Men | | Women | | Men | | Women | |
| | | Unexposed | Exposed | Unexposed | Exposed | Unexposed | Exposed | Unexposed | Exposed |
| Years since diagnosis | Age group | Marg. pred. (95% CI) | Marg. pred. (95% CI) | Marg. pred. (95% CI) | Marg. pred. (95% CI) | Marg. pred. (95% CI) | Marg. pred. (95% CI) | Marg. pred. (95% CI) | Marg. pred. (95% CI) |
| | | Unexposed | Exposed | Unexposed | Exposed | Unexposed | Exposed | Unexposed | Exposed |
| 4 | | 0.85 (0.84–0.85) | 0.8 (0.79–0.8) | 0.77 (0.77–0.78) | 0.69 (0.68–0.69) | 0.75 (0.74–0.76) | 0.7 (0.69–0.71) | 0.69 (0.68–0.71) | 0.58 (0.57–0.59) |
| 5 | | 0.84 (0.84–0.85) | 0.79 (0.79–0.8) | 0.79 (0.78–0.79) | 0.7 (0.7–0.71) | 0.74 (0.73–0.75) | 0.69 (0.68–0.7) | 0.7 (0.69–0.71) | 0.58 (0.57–0.6) |
| -5 | 5–9 years | 0.82 (0.82–0.82) | 0.78 (0.78–0.78) | 0.59 (0.58–0.59) | 0.54 (0.54–0.55) | 0.71 (0.71–0.72) | 0.71 (0.71–0.72) | 0.59 (0.59–0.6) | 0.57 (0.56–0.57) |
| -4 | | 0.82 (0.82–0.83) | 0.79 (0.78–0.79) | 0.63 (0.63–0.63) | 0.58 (0.58–0.58) | 0.73 (0.73–0.73) | 0.72 (0.72–0.73) | 0.63 (0.62–0.63) | 0.59 (0.59–0.6) |
| -3 | | 0.83 (0.83–0.83) | 0.79 (0.78–0.79) | 0.67 (0.67–0.67) | 0.61 (0.61–0.62) | 0.74 (0.73–0.74) | 0.73 (0.73–0.74) | 0.65 (0.64–0.65) | 0.61 (0.6–0.61) |
| -2 | | 0.83 (0.83–0.83) | 0.79 (0.79–0.79) | 0.7 (0.7–0.7) | 0.64 (0.64–0.65) | 0.74 (0.73–0.74) | 0.73 (0.73–0.74) | 0.66 (0.65–0.66) | 0.62 (0.61–0.62) |
| -1 | | 0.84 (0.83–0.84) | 0.8 (0.79–0.8) | 0.72 (0.72–0.73) | 0.66 (0.66–0.66) | 0.74 (0.74–0.75) | 0.73 (0.73–0.74) | 0.67 (0.66–0.67) | 0.63 (0.62–0.63) |
| 0 | | 0.84 (0.84–0.84) | 0.8 (0.8–0.8) | 0.75 (0.75–0.75) | 0.68 (0.68–0.68) | 0.74 (0.74–0.75) | 0.73 (0.73–0.74) | 0.68 (0.67–0.68) | 0.63 (0.62–0.63) |
| 1 | | 0.84 (0.84–0.84) | 0.8 (0.79–0.8) | 0.77 (0.76–0.77) | 0.7 (0.7–0.7) | 0.74 (0.74–0.75) | 0.73 (0.73–0.73) | 0.68 (0.68–0.69) | 0.63 (0.62–0.63) |
| 2 | | 0.84 (0.84–0.84) | 0.8 (0.79–0.8) | 0.78 (0.78–0.79) | 0.72 (0.71–0.72) | 0.74 (0.73–0.74) | 0.72 (0.72–0.73) | 0.69 (0.68–0.69) | 0.63 (0.63–0.64) |

**eTable 25. Estimates for marginal predictions of annual employment among parents exposed to child's any mental disorder (F00–F99) and unexposed parents stratified by the child’s age at diagnosis**

| | | Finland | | | | Denmark | | | |
|---|---|---|---|---|---|---|---|---|---|
| | | Men | | Women | | Men | | Women | |
| | | Unexposed | Exposed | Unexposed | Exposed | Unexposed | Exposed | Unexposed | Exposed |
| Years since diagnosis | Age group | Marg. pred. (95% CI) | Marg. pred. (95% CI) | Marg. pred. (95% CI) | Marg. pred. (95% CI) | Marg. pred. (95% CI) | Marg. pred. (95% CI) | Marg. pred. (95% CI) | Marg. pred. (95% CI) |
| | | Unexposed | Exposed | Unexposed | Exposed | Unexposed | Exposed | Unexposed | Exposed |
| 3 | | 0.84 (0.84–0.84) | 0.79 (0.79–0.79) | 0.8 (0.79–0.8) | 0.73 (0.73–0.74) | 0.73 (0.73–0.74) | 0.72 (0.71–0.72) | 0.7 (0.69–0.7) | 0.64 (0.63–0.64) |
| 4 | | 0.84 (0.83–0.84) | 0.79 (0.79–0.79) | 0.81 (0.81–0.81) | 0.74 (0.74–0.75) | 0.73 (0.72–0.73) | 0.71 (0.71–0.72) | 0.7 (0.69–0.71) | 0.64 (0.63–0.65) |
| 5 | | 0.83 (0.83–0.84) | 0.78 (0.78–0.79) | 0.82 (0.82–0.82) | 0.76 (0.75–0.76) | 0.72 (0.72–0.73) | 0.71 (0.71–0.72) | 0.7 (0.7–0.71) | 0.64 (0.63–0.64) |
| -5 | 10–14 years | 0.83 (0.83–0.83) | 0.79 (0.79–0.79) | 0.75 (0.75–0.75) | 0.7 (0.7–0.71) | 0.74 (0.74–0.75) | 0.73 (0.73–0.74) | 0.68 (0.67–0.68) | 0.65 (0.65–0.65) |
| -4 | | 0.83 (0.82–0.83) | 0.79 (0.79–0.79) | 0.76 (0.76–0.77) | 0.72 (0.71–0.72) | 0.75 (0.75–0.75) | 0.75 (0.74–0.75) | 0.7 (0.69–0.7) | 0.67 (0.66–0.67) |
| -3 | | 0.83 (0.82–0.83) | 0.78 (0.78–0.79) | 0.78 (0.77–0.78) | 0.73 (0.72–0.73) | 0.76 (0.75–0.76) | 0.75 (0.74–0.75) | 0.71 (0.7–0.71) | 0.68 (0.67–0.68) |
| -2 | | 0.82 (0.82–0.83) | 0.78 (0.78–0.79) | 0.79 (0.79–0.79) | 0.74 (0.74–0.74) | 0.75 (0.75–0.76) | 0.74 (0.74–0.75) | 0.71 (0.7–0.71) | 0.68 (0.68–0.68) |
| -1 | | 0.82 (0.82–0.83) | 0.78 (0.78–0.79) | 0.8 (0.8–0.8) | 0.75 (0.75–0.75) | 0.75 (0.75–0.75) | 0.74 (0.74–0.75) | 0.72 (0.71–0.72) | 0.68 (0.68–0.69) |
| 0 | | 0.82 (0.82–0.83) | 0.78 (0.78–0.78) | 0.81 (0.81–0.81) | 0.76 (0.75–0.76) | 0.75 (0.74–0.75) | 0.74 (0.73–0.74) | 0.72 (0.71–0.72) | 0.68 (0.68–0.69) |

**eTable 25. Estimates for marginal predictions of annual employment among parents exposed to child's any mental disorder (F00–F99) and unexposed parents stratified by the child's age at diagnosis**

| | | Finland | | | | Denmark | | | |
|---|---|---|---|---|---|---|---|---|---|
| | | Men | | Women | | Men | | Women | |
| | | Unexposed | Exposed | Unexposed | Exposed | Unexposed | Exposed | Unexposed | Exposed |
| Years since diagnosis | Age group | Marg. pred. (95% CI) | Marg. pred. (95% CI) | Marg. pred. (95% CI) | Marg. pred. (95% CI) | Marg. pred. (95% CI) | Marg. pred. (95% CI) | Marg. pred. (95% CI) | Marg. pred. (95% CI) |
| | | Unexposed | Exposed | Unexposed | Exposed | Unexposed | Exposed | Unexposed | Exposed |
| 1 | | 0.82 (0.82–0.82) | 0.77 (0.77–0.78) | 0.82 (0.81–0.82) | 0.76 (0.76–0.76) | 0.74 (0.74–0.75) | 0.73 (0.73–0.74) | 0.72 (0.72–0.73) | 0.68 (0.67–0.68) |
| 2 | | 0.81 (0.81–0.82) | 0.77 (0.76–0.77) | 0.82 (0.82–0.83) | 0.77 (0.77–0.77) | 0.74 (0.74–0.74) | 0.73 (0.72–0.73) | 0.72 (0.72–0.73) | 0.68 (0.67–0.68) |
| 3 | | 0.81 (0.8–0.81) | 0.76 (0.76–0.77) | 0.83 (0.83–0.83) | 0.77 (0.77–0.78) | 0.74 (0.73–0.74) | 0.72 (0.72–0.73) | 0.72 (0.72–0.73) | 0.68 (0.67–0.68) |
| 4 | | 0.8 (0.8–0.8) | 0.75 (0.75–0.76) | 0.83 (0.83–0.83) | 0.78 (0.77–0.78) | 0.73 (0.73–0.74) | 0.72 (0.71–0.72) | 0.72 (0.72–0.73) | 0.68 (0.67–0.68) |
| 5 | | 0.79 (0.79–0.8) | 0.74 (0.74–0.75) | 0.83 (0.83–0.83) | 0.77 (0.77–0.78) | 0.72 (0.72–0.73) | 0.71 (0.7–0.71) | 0.72 (0.72–0.73) | 0.67 (0.67–0.68) |
| -5 | 15–19 years | 0.83 (0.83–0.83) | 0.79 (0.79–0.79) | 0.81 (0.8–0.81) | 0.76 (0.76–0.77) | 0.74 (0.74–0.74) | 0.72 (0.72–0.72) | 0.7 (0.7–0.7) | 0.67 (0.66–0.67) |
| -4 | | 0.82 (0.82–0.82) | 0.78 (0.78–0.79) | 0.81 (0.81–0.81) | 0.77 (0.77–0.77) | 0.75 (0.75–0.75) | 0.73 (0.72–0.73) | 0.72 (0.71–0.72) | 0.68 (0.68–0.68) |
| -3 | | 0.82 (0.81–0.82) | 0.78 (0.78–0.78) | 0.82 (0.81–0.82) | 0.78 (0.77–0.78) | 0.75 (0.75–0.76) | 0.73 (0.73–0.73) | 0.72 (0.72–0.73) | 0.69 (0.68–0.69) |
| -2 | | 0.81 (0.81–0.81) | 0.77 (0.77–0.78) | 0.82 (0.82–0.82) | 0.78 (0.78–0.78) | 0.75 (0.74–0.75) | 0.72 (0.72–0.73) | 0.73 (0.72–0.73) | 0.68 (0.68–0.69) |

**eTable 25. Estimates for marginal predictions of annual employment among parents exposed to child's any mental disorder (F00–F99) and unexposed parents stratified by the child's age at diagnosis**

| | | Finland | | | | Denmark | | | |
|---|---|---|---|---|---|---|---|---|---|
| | | Men | | Women | | Men | | Women | |
| | | Unexposed | Exposed | Unexposed | Exposed | Unexposed | Exposed | Unexposed | Exposed |
| Years since diagnosis | Age group | Marg. pred. (95% CI) | Marg. pred. (95% CI) | Marg. pred. (95% CI) | Marg. pred. (95% CI) | Marg. pred. (95% CI) | Marg. pred. (95% CI) | Marg. pred. (95% CI) | Marg. pred. (95% CI) |
| | | Unexposed | Exposed | Unexposed | Exposed | Unexposed | Exposed | Unexposed | Exposed |
| -1 | | 0.8 (0.8–0.81) | 0.77 (0.76–0.77) | 0.82 (0.82–0.83) | 0.78 (0.78–0.79) | 0.74 (0.74–0.74) | 0.72 (0.71–0.72) | 0.72 (0.72–0.73) | 0.68 (0.68–0.69) |
| 0 | | 0.8 (0.79–0.8) | 0.76 (0.76–0.76) | 0.82 (0.82–0.83) | 0.78 (0.78–0.79) | 0.74 (0.73–0.74) | 0.71 (0.71–0.72) | 0.72 (0.72–0.73) | 0.68 (0.68–0.68) |
| 1 | | 0.79 (0.78–0.79) | 0.75 (0.75–0.75) | 0.82 (0.82–0.83) | 0.78 (0.78–0.79) | 0.73 (0.73–0.73) | 0.7 (0.7–0.71) | 0.72 (0.72–0.72) | 0.67 (0.67–0.68) |
| 2 | | 0.77 (0.77–0.78) | 0.74 (0.73–0.74) | 0.82 (0.82–0.83) | 0.78 (0.78–0.78) | 0.72 (0.72–0.73) | 0.7 (0.69–0.7) | 0.72 (0.71–0.72) | 0.67 (0.67–0.68) |
| 3 | | 0.76 (0.76–0.76) | 0.72 (0.72–0.72) | 0.82 (0.82–0.82) | 0.78 (0.77–0.78) | 0.71 (0.71–0.72) | 0.69 (0.68–0.69) | 0.72 (0.71–0.72) | 0.67 (0.66–0.67) |
| 4 | | 0.74 (0.74–0.75) | 0.71 (0.7–0.71) | 0.81 (0.81–0.81) | 0.77 (0.77–0.77) | 0.71 (0.7–0.71) | 0.68 (0.67–0.68) | 0.71 (0.71–0.72) | 0.66 (0.66–0.66) |
| 5 | | 0.72 (0.72–0.73) | 0.69 (0.69–0.69) | 0.8 (0.8–0.8) | 0.76 (0.76–0.76) | 0.69 (0.69–0.7) | 0.66 (0.66–0.67) | 0.71 (0.7–0.71) | 0.65 (0.64–0.65) |
| -5 | 20–25 years | 0.79 (0.79–0.8) | 0.76 (0.75–0.76) | 0.82 (0.81–0.82) | 0.77 (0.77–0.78) | 0.72 (0.72–0.73) | 0.69 (0.69–0.7) | 0.7 (0.69–0.7) | 0.65 (0.65–0.66) |
| -4 | | 0.78 (0.78–0.78) | 0.74 (0.74–0.75) | 0.81 (0.81–0.81) | 0.77 (0.77–0.78) | 0.74 (0.74–0.75) | 0.71 (0.71–0.71) | 0.72 (0.71–0.72) | 0.67 (0.67–0.68) |

**eTable 25. Estimates for marginal predictions of annual employment among parents exposed to child's any mental disorder (F00–F99) and unexposed parents stratified by the child's age at diagnosis**

| | | Finland | | | | Denmark | | | |
|---|---|---|---|---|---|---|---|---|---|
| | | Men | | Women | | Men | | Women | |
| | | Unexposed | Exposed | Unexposed | Exposed | Unexposed | Exposed | Unexposed | Exposed |
| Years since diagnosis | Age group | Marg. pred. (95% CI) | Marg. pred. (95% CI) | Marg. pred. (95% CI) | Marg. pred. (95% CI) | Marg. pred. (95% CI) | Marg. pred. (95% CI) | Marg. pred. (95% CI) | Marg. pred. (95% CI) |
| | | Unexposed | Exposed | Unexposed | Exposed | Unexposed | Exposed | Unexposed | Exposed |
| -3 | | 0.76 (0.76–0.77) | 0.73 (0.73–0.73) | 0.81 (0.8–0.81) | 0.77 (0.76–0.77) | 0.75 (0.74–0.75) | 0.71 (0.71–0.72) | 0.72 (0.72–0.73) | 0.68 (0.67–0.68) |
| -2 | | 0.75 (0.75–0.75) | 0.72 (0.71–0.72) | 0.8 (0.8–0.8) | 0.76 (0.76–0.76) | 0.74 (0.74–0.74) | 0.7 (0.7–0.7) | 0.72 (0.71–0.72) | 0.67 (0.67–0.68) |
| -1 | | 0.74 (0.73–0.74) | 0.7 (0.7–0.71) | 0.79 (0.79–0.79) | 0.75 (0.75–0.76) | 0.73 (0.72–0.73) | 0.69 (0.69–0.69) | 0.71 (0.71–0.72) | 0.67 (0.66–0.67) |
| 0 | | 0.72 (0.72–0.72) | 0.69 (0.68–0.69) | 0.78 (0.78–0.78) | 0.74 (0.74–0.75) | 0.72 (0.71–0.72) | 0.68 (0.67–0.68) | 0.7 (0.7–0.71) | 0.66 (0.65–0.66) |
| 1 | | 0.69 (0.69–0.7) | 0.66 (0.66–0.67) | 0.77 (0.76–0.77) | 0.73 (0.73–0.73) | 0.7 (0.7–0.71) | 0.66 (0.66–0.67) | 0.7 (0.69–0.7) | 0.64 (0.64–0.65) |
| 2 | | 0.67 (0.67–0.67) | 0.64 (0.64–0.65) | 0.75 (0.75–0.76) | 0.72 (0.71–0.72) | 0.69 (0.68–0.69) | 0.65 (0.64–0.65) | 0.68 (0.68–0.69) | 0.64 (0.63–0.64) |
| 3 | | 0.64 (0.64–0.65) | 0.62 (0.61–0.62) | 0.73 (0.73–0.74) | 0.7 (0.7–0.7) | 0.67 (0.66–0.67) | 0.63 (0.62–0.63) | 0.67 (0.67–0.67) | 0.62 (0.62–0.62) |
| 4 | | 0.61 (0.61–0.62) | 0.59 (0.59–0.59) | 0.71 (0.71–0.72) | 0.68 (0.68–0.69) | 0.64 (0.64–0.65) | 0.61 (0.6–0.61) | 0.65 (0.65–0.66) | 0.6 (0.6–0.61) |
| 5 | | 0.58 (0.58–0.59) | 0.56 (0.55–0.56) | 0.69 (0.69–0.7) | 0.66 (0.66–0.67) | 0.62 (0.62–0.63) | 0.58 (0.58–0.59) | 0.64 (0.63–0.64) | 0.59 (0.58–0.59) |

Marg. pred. = Marginal predictions in probabilities

**eTable 26. Estimates for marginal predictions of annual earnings among parents exposed to child's substance use disorder (F10–F19) and unexposed parents stratified by the child’s age at diagnosis**

| | | Finland | | | | Denmark | | | |
|---|---|---|---|---|---|---|---|---|---|
| | | Men | | Women | | Men | | Women | |
| | | Unexposed | Exposed | Unexposed | Exposed | Unexposed | Exposed | Unexposed | Exposed |
| Years since diagnosis | Age group | Marg. pred. (95% CI) | Marg. pred. (95% CI) | Marg. pred. (95% CI) | Marg. pred. (95% CI) | Marg. pred. (95% CI) | Marg. pred. (95% CI) | Marg. pred. (95% CI) | Marg. pred. (95% CI) |
| | | Unexposed | Exposed | Unexposed | Exposed | Unexposed | Exposed | Unexposed | Exposed |
| -5 | 5–14 years | 31.2 (30.4–31.9) | 28.3 (27.7–29) | 18.8 (18.3–19.2) | 17.8 (17.3–18.2) | 48.2 (45.8–50.7) | 45.2 (42.8–47.5) | 40.4 (39–41.8) | 39.7 (38.5–40.9) |
| -4 | | 31.6 (30.9–32.3) | 28.8 (28.1–29.5) | 19.6 (19.1–20.1) | 18.5 (18.1–19) | 48.3 (45.9–50.7) | 46.6 (44.2–49) | 41 (39.6–42.3) | 40.1 (38.8–41.5) |
| -3 | | 32.3 (31.5–33) | 29 (28.3–29.7) | 20.6 (20.1–21.1) | 19.3 (18.8–19.8) | 50.4 (47.7–53.1) | 47.1 (44.5–49.6) | 41.7 (40.4–43.1) | 40.1 (38.8–41.4) |
| -2 | | 32.5 (31.7–33.3) | 29.5 (28.7–30.2) | 21.4 (20.9–21.9) | 20.3 (19.8–20.7) | 50.2 (47.6–52.7) | 47.5 (45–50) | 42 (40.6–43.5) | 41.1 (39.8–42.4) |
| -1 | | 33.2 (32.5–34) | 30.2 (29.5–31) | 22.2 (21.7–22.7) | 21.2 (20.7–21.7) | 51.5 (48.8–54.1) | 48.1 (45.6–50.6) | 42.1 (40.6–43.5) | 41.7 (40.3–43.1) |
| 0 | | 33.7 (32.9–34.5) | 30.5 (29.8–31.3) | 23 (22.5–23.5) | 21.9 (21.4–22.4) | 51.8 (49.1–54.5) | 48.4 (45.8–51.1) | 42.8 (41.3–44.3) | 41.3 (39.9–42.6) |
| 1 | | 34.2 (33.3–35) | 30.9 (30.2–31.7) | 23.7 (23.2–24.2) | 22.4 (21.9–22.9) | 52.9 (50–55.7) | 47.7 (45.1–50.2) | 43.6 (42–45.2) | 40.8 (39.3–42.3) |
| 2 | | 34.5 (33.6–35.3) | 31.3 (30.4–32.1) | 24.3 (23.8–24.8) | 23 (22.5–23.6) | 52.8 (50.1–55.6) | 48.3 (45.7–51) | 43.6 (42–45.2) | 41.6 (40.1–43.2) |
| 3 | | 34.6 (33.6–35.5) | 31.2 (30.4–32.1) | 25 (24.4–25.5) | 23.6 (23–24.2) | 53.1 (50.2–55.9) | 48.2 (45.6–50.8) | 43.3 (41.6–44.9) | 41.7 (40.1–43.3) |

**eTable 26. Estimates for marginal predictions of annual earnings among parents exposed to child's substance use disorder (F10–F19) and unexposed parents stratified by the child's age at diagnosis**

| | | Finland | | | | Denmark | | | |
|---|---|---|---|---|---|---|---|---|---|
| | | Men | | Women | | Men | | Women | |
| | | Unexposed | Exposed | Unexposed | Exposed | Unexposed | Exposed | Unexposed | Exposed |
| Years since diagnosis | Age group | Marg. pred. (95% CI) | Marg. pred. (95% CI) | Marg. pred. (95% CI) | Marg. pred. (95% CI) | Marg. pred. (95% CI) | Marg. pred. (95% CI) | Marg. pred. (95% CI) | Marg. pred. (95% CI) |
| | | Unexposed | Exposed | Unexposed | Exposed | Unexposed | Exposed | Unexposed | Exposed |
| 4 | | 34.5 (33.6–35.5) | 31.5 (30.6–32.4) | 25.5 (24.9–26) | 24 (23.4–24.6) | 52.9 (49.9–55.9) | 48.2 (45.4–51) | 42.7 (41–44.5) | 41.4 (39.7–43.1) |
| 5 | | 35 (34–35.9) | 31.7 (30.8–32.7) | 26 (25.4–26.6) | 24.4 (23.8–25) | 52.8 (49.6–56) | 47.4 (44.6–50.2) | 42.4 (40.6–44.3) | 39.8 (38.1–41.6) |
| -5 | 15–19 years | 31.9 (31.5–32.2) | 29 (28.7–29.4) | 21.2 (21–21.4) | 19.6 (19.4–19.8) | 51.6 (51–52.3) | 47.6 (47–48.2) | 40.6 (40.3–40.9) | 40.4 (40.1–40.7) |
| -4 | | 32.2 (31.8–32.5) | 29.3 (28.9–29.6) | 21.8 (21.6–22) | 20.3 (20–20.5) | 51.8 (51.1–52.5) | 47.8 (47.2–48.4) | 41.2 (40.9–41.5) | 40.8 (40.5–41.2) |
| -3 | | 32.4 (32.1–32.8) | 29.4 (29.1–29.8) | 22.4 (22.2–22.6) | 20.9 (20.6–21.1) | 52.2 (51.6–52.9) | 48.2 (47.6–48.8) | 41.8 (41.4–42.1) | 41.2 (40.9–41.5) |
| -2 | | 32.4 (32–32.7) | 29.7 (29.3–30) | 23 (22.7–23.2) | 21.4 (21.2–21.7) | 52.6 (52–53.3) | 48.7 (48.1–49.3) | 42.1 (41.8–42.5) | 41.3 (41–41.6) |
| -1 | | 32.7 (32.4–33.1) | 29.8 (29.4–30.2) | 23.6 (23.3–23.8) | 22 (21.7–22.2) | 52.7 (52–53.4) | 49 (48.4–49.6) | 42.2 (41.8–42.5) | 41.2 (40.8–41.5) |
| 0 | | 32.9 (32.5–33.2) | 29.7 (29.3–30.1) | 24 (23.7–24.2) | 22.3 (22.1–22.6) | 52.6 (51.9–53.3) | 48.7 (48.1–49.3) | 42.1 (41.7–42.5) | 41 (40.7–41.3) |
| 1 | | 32.8 (32.4–33.2) | 29.6 (29.3–30) | 24.4 (24.2–24.7) | 22.7 (22.5–23) | 53.1 (52.4–53.8) | 48.7 (48.1–49.3) | 42 (41.6–42.3) | 40.9 (40.5–41.2) |

**eTable 26. Estimates for marginal predictions of annual earnings among parents exposed to child's substance use disorder (F10–F19) and unexposed parents stratified by the child's age at diagnosis**

| | | Finland | | | | Denmark | | | |
|---|---|---|---|---|---|---|---|---|---|
| | | Men | | Women | | Men | | Women | |
| | | Unexposed | Exposed | Unexposed | Exposed | Unexposed | Exposed | Unexposed | Exposed |
| Years since diagnosis | Age group | Marg. pred. (95% CI) | Marg. pred. (95% CI) | Marg. pred. (95% CI) | Marg. pred. (95% CI) | Marg. pred. (95% CI) | Marg. pred. (95% CI) | Marg. pred. (95% CI) | Marg. pred. (95% CI) |
| | | Unexposed | Exposed | Unexposed | Exposed | Unexposed | Exposed | Unexposed | Exposed |
| 2 | | 32.7 (32.3–33.1) | 29.5 (29.1–29.9) | 24.8 (24.5–25) | 23 (22.7–23.2) | 53 (52.2–53.7) | 49 (48.3–49.6) | 42 (41.6–42.4) | 40.8 (40.5–41.2) |
| 3 | | 32.4 (32–32.8) | 29 (28.6–29.4) | 25 (24.7–25.3) | 23.3 (23–23.5) | 52.8 (52–53.5) | 48.5 (47.8–49.2) | 42 (41.7–42.4) | 40.9 (40.5–41.3) |
| 4 | | 32 (31.6–32.5) | 28.8 (28.3–29.2) | 25.2 (25–25.5) | 23.4 (23.2–23.7) | 52.8 (52–53.6) | 48.2 (47.6–48.9) | 42 (41.6–42.4) | 40.8 (40.4–41.2) |
| 5 | | 31.4 (31–31.9) | 28.3 (27.9–28.8) | 25.3 (25.1–25.6) | 23.5 (23.3–23.8) | 52.9 (52.1–53.7) | 48.1 (47.5–48.8) | 42.1 (41.7–42.5) | 40.8 (40.4–41.2) |
| -5 | 20–25 years | 31.3 (31–31.6) | 29 (28.7–29.3) | 22.2 (22–22.4) | 21 (20.8–21.2) | 52 (51.4–52.5) | 48.6 (48.1–49.1) | 40 (39.8–40.3) | 39.2 (38.9–39.5) |
| -4 | | 31.1 (30.8–31.4) | 29 (28.6–29.3) | 22.6 (22.4–22.8) | 21.3 (21.1–21.5) | 52 (51.5–52.6) | 48.7 (48.2–49.2) | 40.2 (39.9–40.4) | 39.2 (39–39.5) |
| -3 | | 30.9 (30.5–31.2) | 28.6 (28.3–28.9) | 22.8 (22.6–23) | 21.7 (21.5–21.9) | 52.2 (51.7–52.8) | 48.6 (48.1–49.1) | 40.1 (39.8–40.4) | 39.2 (38.9–39.4) |
| -2 | | 30.5 (30.2–30.9) | 28.4 (28–28.7) | 22.9 (22.7–23.1) | 21.8 (21.6–22) | 52.1 (51.6–52.7) | 48.4 (47.9–48.9) | 40 (39.7–40.3) | 39.1 (38.8–39.3) |
| -1 | | 30.1 (29.8–30.5) | 28.1 (27.8–28.4) | 23.1 (22.8–23.3) | 21.9 (21.7–22.1) | 52.3 (51.8–52.9) | 48.4 (47.9–48.9) | 40.1 (39.8–40.4) | 39.1 (38.8–39.4) |

**eTable 26. Estimates for marginal predictions of annual earnings among parents exposed to child's substance use disorder (F10–F19) and unexposed parents stratified by the child’s age at diagnosis**

| | | Finland | | | | Denmark | | | |
|---|---|---|---|---|---|---|---|---|---|
| | | Men | | Women | | Men | | Women | |
| | | Unexposed | Exposed | Unexposed | Exposed | Unexposed | Exposed | Unexposed | Exposed |
| Years since diagnosis | Age group | Marg. pred. (95% CI) | Marg. pred. (95% CI) | Marg. pred. (95% CI) | Marg. pred. (95% CI) | Marg. pred. (95% CI) | Marg. pred. (95% CI) | Marg. pred. (95% CI) | Marg. pred. (95% CI) |
| | | Unexposed | Exposed | Unexposed | Exposed | Unexposed | Exposed | Unexposed | Exposed |
| 0 | | 29.6 (29.3–30) | 27.9 (27.5–28.2) | 23 (22.8–23.2) | 21.9 (21.7–22.1) | 52.6 (52–53.2) | 48.3 (47.8–48.9) | 40.3 (40–40.6) | 39.1 (38.8–39.4) |
| 1 | | 29 (28.7–29.4) | 27.3 (27–27.7) | 23 (22.8–23.2) | 21.9 (21.7–22.1) | 52.5 (51.9–53.1) | 48.3 (47.7–48.8) | 40.3 (40–40.6) | 39.3 (39–39.6) |
| 2 | | 28.4 (28.1–28.8) | 26.6 (26.2–27) | 22.8 (22.6–23.1) | 21.7 (21.4–21.9) | 52.2 (51.6–52.8) | 48.2 (47.7–48.8) | 40.2 (39.9–40.5) | 39.4 (39.1–39.7) |
| 3 | | 27.6 (27.2–28) | 25.9 (25.5–26.2) | 22.6 (22.3–22.8) | 21.5 (21.3–21.7) | 52 (51.4–52.7) | 48.1 (47.5–48.6) | 40.2 (39.9–40.5) | 39.4 (39.1–39.7) |
| 4 | | 26.6 (26.2–27) | 25 (24.6–25.4) | 22.2 (22–22.4) | 21.2 (21–21.5) | 51.4 (50.8–52.1) | 47.7 (47.1–48.3) | 40.3 (40–40.6) | 39.3 (39–39.6) |
| 5 | | 25.4 (25–25.9) | 24 (23.6–24.4) | 21.8 (21.5–22) | 20.9 (20.7–21.2) | 50.8 (50.2–51.5) | 47.7 (47.1–48.3) | 40.2 (39.9–40.5) | 39.2 (38.8–39.5) |

Marg. pred. = Marginal predictions in thousand euros (€)

**eTable 27. Estimates for marginal predictions of annual disposable income among parents exposed to child's substance use disorder (F10–F19) and unexposed parents stratified by the child's age at diagnosis**

| | | Finland | | | | Denmark | | | |
|---|---|---|---|---|---|---|---|---|---|
| | | Men | | Women | | Men | | Women | |
| | | Unexposed | Exposed | Unexposed | Exposed | Unexposed | Exposed | Unexposed | Exposed |
| Years since diagnosis | Age group | Marg. pred. (95% CI) | Marg. pred. (95% CI) | Marg. pred. (95% CI) | Marg. pred. (95% CI) | Marg. pred. (95% CI) | Marg. pred. (95% CI) | Marg. pred. (95% CI) | Marg. pred. (95% CI) |
| | | Unexposed | Exposed | Unexposed | Exposed | Unexposed | Exposed | Unexposed | Exposed |
| -5 | 5–14 years | 26 (25.6–26.5) | 24.9 (24.4–25.3) | 22.3 (22.1–22.6) | 22.3 (22–22.5) | 29.2 (28–30.5) | 27.4 (26.1–28.7) | 28.2 (27.4–29) | 27.9 (27.1–28.6) |
| -4 | | 26.5 (26–27) | 25.3 (24.8–25.7) | 22.9 (22.6–23.2) | 22.9 (22.6–23.1) | 29.6 (28.2–30.9) | 28.5 (27.1–29.9) | 28.7 (27.9–29.5) | 28.1 (27.3–28.9) |
| -3 | | 27.1 (26.6–27.5) | 25.6 (25.1–26) | 23.5 (23.3–23.8) | 23.5 (23.2–23.7) | 31.4 (29.9–32.8) | 29.2 (27.8–30.6) | 29.4 (28.6–30.3) | 28.2 (27.4–29) |
| -2 | | 27.5 (27–28) | 26.1 (25.6–26.5) | 24 (23.7–24.3) | 24.1 (23.8–24.3) | 31.6 (30.1–33) | 29.5 (28.1–30.9) | 29.8 (29–30.7) | 29 (28.2–29.8) |
| -1 | | 28.3 (27.8–28.8) | 26.8 (26.3–27.3) | 24.5 (24.2–24.8) | 24.5 (24.3–24.8) | 32.3 (30.9–33.8) | 30 (28.6–31.4) | 30 (29.1–30.9) | 29.4 (28.6–30.3) |
| 0 | | 28.7 (28.2–29.2) | 27 (26.6–27.5) | 24.9 (24.6–25.2) | 24.9 (24.6–25.2) | 32.9 (31.4–34.5) | 30.4 (28.9–31.9) | 30.5 (29.6–31.4) | 29.3 (28.4–30.1) |
| 1 | | 29.3 (28.8–29.9) | 27.6 (27.1–28.1) | 25.3 (25–25.6) | 25.1 (24.8–25.4) | 34 (32.5–35.6) | 30.4 (28.9–31.8) | 31 (30–32) | 28.9 (28–29.9) |
| 2 | | 29.8 (29.2–30.4) | 28 (27.4–28.5) | 25.5 (25.2–25.8) | 25.3 (25–25.6) | 34.6 (33–36.2) | 31.2 (29.6–32.7) | 31.3 (30.3–32.3) | 29.4 (28.4–30.3) |
| 3 | | 30 (29.4–30.6) | 28.1 (27.6–28.7) | 25.4 (25–25.7) | 25.1 (24.8–25.4) | 35 (33.2–36.7) | 31.6 (30–33.1) | 31.1 (30–32.1) | 29.4 (28.4–30.3) |

**eTable 27. Estimates for marginal predictions of annual disposable income among parents exposed to child's substance use disorder (F10–F19) and unexposed parents stratified by the child's age at diagnosis**

| | | Finland | | | | Denmark | | | |
|---|---|---|---|---|---|---|---|---|---|
| | | Men | | Women | | Men | | Women | |
| | | Unexposed | Exposed | Unexposed | Exposed | Unexposed | Exposed | Unexposed | Exposed |
| Years since diagnosis | Age group | Marg. pred. (95% CI) | Marg. pred. (95% CI) | Marg. pred. (95% CI) | Marg. pred. (95% CI) | Marg. pred. (95% CI) | Marg. pred. (95% CI) | Marg. pred. (95% CI) | Marg. pred. (95% CI) |
| | | Unexposed | Exposed | Unexposed | Exposed | Unexposed | Exposed | Unexposed | Exposed |
| 4 | | 30.4 (29.8–31) | 28.6 (28–29.2) | 25.2 (24.9–25.6) | 24.7 (24.4–25) | 35.2 (33.4–37) | 32 (30.3–33.7) | 30.6 (29.5–31.7) | 29.3 (28.2–30.4) |
| 5 | | 30.9 (30.2–31.5) | 29 (28.4–29.6) | 25.3 (25–25.7) | 24.7 (24.4–25.1) | 35.5 (33.6–37.4) | 31.9 (30.2–33.6) | 30.4 (29.2–31.6) | 28.4 (27.2–29.5) |
| -5 | 15–19 years | 26.9 (26.7–27.2) | 25.4 (25.2–25.6) | 23 (22.9–23.1) | 22.7 (22.6–22.8) | 31.5 (31.1–31.9) | 29 (28.7–29.3) | 28.3 (28.1–28.4) | 27.9 (27.7–28.1) |
| -4 | | 27.3 (27–27.5) | 25.6 (25.4–25.9) | 23.4 (23.3–23.5) | 23.1 (22.9–23.2) | 32 (31.6–32.4) | 29.4 (29.1–29.7) | 28.7 (28.5–28.9) | 28.2 (28–28.4) |
| -3 | | 27.7 (27.4–27.9) | 26 (25.8–26.2) | 23.7 (23.5–23.8) | 23.4 (23.2–23.5) | 32.5 (32.2–32.9) | 29.9 (29.5–30.2) | 29.2 (29–29.4) | 28.5 (28.3–28.7) |
| -2 | | 27.9 (27.7–28.1) | 26.4 (26.1–26.6) | 23.7 (23.6–23.8) | 23.4 (23.3–23.6) | 33.2 (32.8–33.6) | 30.5 (30.2–30.9) | 29.5 (29.2–29.7) | 28.6 (28.4–28.8) |
| -1 | | 28.3 (28.1–28.6) | 26.6 (26.4–26.9) | 23.7 (23.6–23.8) | 23.4 (23.3–23.5) | 33.6 (33.2–34) | 31.1 (30.7–31.4) | 29.6 (29.4–29.8) | 28.5 (28.3–28.7) |
| 0 | | 28.6 (28.4–28.9) | 26.8 (26.5–27) | 23.6 (23.5–23.7) | 23.2 (23.1–23.4) | 34 (33.6–34.4) | 31.2 (30.9–31.6) | 29.5 (29.3–29.7) | 28.4 (28.2–28.6) |
| 1 | | 28.8 (28.5–29) | 27 (26.7–27.2) | 23.6 (23.5–23.8) | 23.2 (23–23.3) | 34.7 (34.3–35.1) | 31.6 (31.3–32) | 29.5 (29.2–29.7) | 28.3 (28.1–28.5) |

**eTable 27. Estimates for marginal predictions of annual disposable income among parents exposed to child's substance use disorder (F10–F19) and unexposed parents stratified by the child's age at diagnosis**

| | | Finland | | | | Denmark | | | |
|---|---|---|---|---|---|---|---|---|---|
| | | Men | | Women | | Men | | Women | |
| | | Unexposed | Exposed | Unexposed | Exposed | Unexposed | Exposed | Unexposed | Exposed |
| Years since diagnosis | Age group | Marg. pred. (95% CI) | Marg. pred. (95% CI) | Marg. pred. (95% CI) | Marg. pred. (95% CI) | Marg. pred. (95% CI) | Marg. pred. (95% CI) | Marg. pred. (95% CI) | Marg. pred. (95% CI) |
| | | Unexposed | Exposed | Unexposed | Exposed | Unexposed | Exposed | Unexposed | Exposed |
| 2 | | 29.1 (28.8–29.4) | 27.2 (27–27.5) | 23.6 (23.5–23.8) | 23.1 (23–23.3) | 35 (34.6–35.5) | 32.2 (31.8–32.6) | 29.6 (29.3–29.8) | 28.4 (28.2–28.6) |
| 3 | | 29.3 (29–29.5) | 27.3 (27.1–27.6) | 23.7 (23.6–23.9) | 23.2 (23.1–23.4) | 35.3 (34.9–35.8) | 32.2 (31.9–32.6) | 29.7 (29.4–29.9) | 28.5 (28.3–28.7) |
| 4 | | 29.5 (29.3–29.8) | 27.5 (27.3–27.8) | 23.9 (23.8–24.1) | 23.4 (23.2–23.5) | 35.7 (35.3–36.2) | 32.6 (32.2–33) | 29.8 (29.5–30) | 28.6 (28.3–28.8) |
| 5 | | 29.6 (29.3–29.9) | 27.7 (27.5–28) | 24.1 (24–24.3) | 23.5 (23.4–23.7) | 36.1 (35.7–36.6) | 32.8 (32.4–33.2) | 30 (29.8–30.3) | 28.7 (28.4–28.9) |
| -5 | 20–25 years | 27.1 (26.8–27.3) | 25.7 (25.5–25.9) | 21.9 (21.8–22) | 21.6 (21.5–21.7) | 32 (31.7–32.3) | 29.9 (29.6–30.1) | 27.4 (27.2–27.5) | 26.6 (26.4–26.7) |
| -4 | | 27.2 (27–27.4) | 25.9 (25.7–26.1) | 21.9 (21.7–22) | 21.5 (21.4–21.6) | 32.5 (32.2–32.8) | 30.4 (30.1–30.6) | 27.5 (27.3–27.6) | 26.7 (26.5–26.8) |
| -3 | | 27.4 (27.2–27.6) | 25.9 (25.7–26.1) | 21.7 (21.6–21.9) | 21.5 (21.4–21.6) | 33 (32.7–33.3) | 30.7 (30.4–31) | 27.5 (27.3–27.6) | 26.6 (26.5–26.8) |
| -2 | | 27.5 (27.3–27.7) | 26.1 (25.9–26.3) | 21.8 (21.7–21.9) | 21.5 (21.4–21.6) | 33.4 (33.1–33.7) | 31 (30.7–31.3) | 27.5 (27.3–27.6) | 26.6 (26.4–26.8) |
| -1 | | 27.7 (27.5–27.9) | 26.3 (26.1–26.5) | 22 (21.8–22.1) | 21.6 (21.5–21.7) | 33.9 (33.6–34.2) | 31.3 (31–31.6) | 27.7 (27.5–27.9) | 26.7 (26.6–26.9) |

**eTable 27. Estimates for marginal predictions of annual disposable income among parents exposed to child's substance use disorder (F10–F19) and unexposed parents stratified by the child’s age at diagnosis**

| | | Finland | | | | Denmark | | | |
|---|---|---|---|---|---|---|---|---|---|
| | | Men | | Women | | Men | | Women | |
| | | Unexposed | Exposed | Unexposed | Exposed | Unexposed | Exposed | Unexposed | Exposed |
| Years since diagnosis | Age group | Marg. pred. (95% CI) | Marg. pred. (95% CI) | Marg. pred. (95% CI) | Marg. pred. (95% CI) | Marg. pred. (95% CI) | Marg. pred. (95% CI) | Marg. pred. (95% CI) | Marg. pred. (95% CI) |
| | | Unexposed | Exposed | Unexposed | Exposed | Unexposed | Exposed | Unexposed | Exposed |
| 0 | | 27.8 (27.6–28) | 26.5 (26.3–26.7) | 22 (21.9–22.1) | 21.7 (21.6–21.8) | 34.5 (34.1–34.8) | 31.6 (31.3–31.9) | 27.9 (27.7–28.1) | 26.9 (26.7–27.1) |
| 1 | | 27.9 (27.6–28.1) | 26.5 (26.3–26.7) | 22.1 (22–22.2) | 21.8 (21.7–21.9) | 34.8 (34.5–35.1) | 32 (31.7–32.3) | 28.1 (27.9–28.2) | 27.2 (27–27.3) |
| 2 | | 28 (27.8–28.3) | 26.6 (26.4–26.8) | 22.2 (22.1–22.4) | 21.9 (21.8–22) | 35 (34.7–35.4) | 32.3 (32–32.7) | 28.2 (28–28.4) | 27.4 (27.2–27.6) |
| 3 | | 28.2 (28–28.5) | 26.7 (26.5–26.9) | 22.4 (22.3–22.5) | 22.1 (21.9–22.2) | 35.3 (34.9–35.7) | 32.6 (32.3–32.9) | 28.4 (28.2–28.6) | 27.5 (27.3–27.7) |
| 4 | | 28.2 (28–28.5) | 26.7 (26.5–27) | 22.5 (22.3–22.6) | 22.2 (22–22.3) | 35.3 (34.9–35.7) | 32.6 (32.3–33) | 28.7 (28.5–28.9) | 27.7 (27.5–27.9) |
| 5 | | 28.3 (28–28.6) | 26.8 (26.5–27) | 22.6 (22.5–22.7) | 22.3 (22.2–22.4) | 35.3 (34.9–35.7) | 33 (32.6–33.3) | 28.8 (28.6–29) | 27.8 (27.6–28) |

Marg. pred. = Marginal predictions in thousand euros (€)

**eTable 28. Estimates for marginal predictions of annual social welfare benefits among parents exposed to child's substance use disorder (F10–F19) and unexposed parents stratified by the child's age at diagnosis**

| | | Finland | | | | Denmark | | | |
|---|---|---|---|---|---|---|---|---|---|
| | | Men | | Women | | Men | | Women | |
| | | Unexposed | Exposed | Unexposed | Exposed | Unexposed | Exposed | Unexposed | Exposed |
| Years since diagnosis | Age group | Marg. pred. (95% CI) | Marg. pred. (95% CI) | Marg. pred. (95% CI) | Marg. pred. (95% CI) | Marg. pred. (95% CI) | Marg. pred. (95% CI) | Marg. pred. (95% CI) | Marg. pred. (95% CI) |
| | | Unexposed | Exposed | Unexposed | Exposed | Unexposed | Exposed | Unexposed | Exposed |
| -5 | 5–14 years | 3.4 (3.2–3.6) | 4 (3.8–4.2) | 8.8 (8.6–9) | 9.6 (9.3–9.8) | 5.8 (4.9–6.6) | 6.9 (6–7.7) | 12.7 (11.8–13.7) | 14.8 (13.8–15.8) |
| -4 | | 3.5 (3.3–3.6) | 4.3 (4.1–4.5) | 8.7 (8.5–8.9) | 9.6 (9.3–9.8) | 6.5 (5.6–7.4) | 6.8 (5.9–7.6) | 12.5 (11.5–13.5) | 14.9 (13.9–15.9) |
| -3 | | 3.6 (3.4–3.8) | 4.4 (4.2–4.6) | 8.6 (8.4–8.8) | 9.6 (9.3–9.8) | 6.7 (5.8–7.6) | 7.2 (6.2–8.1) | 12.4 (11.4–13.4) | 14.9 (13.8–16) |
| -2 | | 3.7 (3.5–3.9) | 4.6 (4.3–4.8) | 8.4 (8.2–8.7) | 9.3 (9.1–9.6) | 6.7 (5.8–7.7) | 7.7 (6.8–8.6) | 12.4 (11.4–13.4) | 15 (13.9–16) |
| -1 | | 3.9 (3.7–4.1) | 4.6 (4.3–4.9) | 8.2 (8–8.5) | 9 (8.8–9.3) | 6.4 (5.5–7.3) | 7.8 (6.8–8.8) | 12.5 (11.4–13.5) | 14.8 (13.7–15.8) |
| 0 | | 3.9 (3.7–4.1) | 4.6 (4.4–4.9) | 8 (7.7–8.2) | 8.8 (8.6–9) | 6.8 (5.8–7.7) | 8 (7–8.9) | 12.4 (11.4–13.4) | 14.2 (13.1–15.3) |
| 1 | | 4.1 (3.9–4.3) | 4.8 (4.6–5) | 7.7 (7.5–7.9) | 8.6 (8.4–8.9) | 6.5 (5.6–7.4) | 8.4 (7.4–9.4) | 12 (11–13.1) | 13.9 (12.8–15) |
| 2 | | 4.3 (4.1–4.6) | 4.9 (4.6–5.1) | 7.5 (7.3–7.8) | 8.3 (8–8.5) | 6.3 (5.4–7.2) | 8.4 (7.4–9.4) | 12.4 (11.3–13.4) | 13.9 (12.7–15) |
| 3 | | 4.4 (4.2–4.7) | 5.2 (4.9–5.5) | 6.7 (6.5–7) | 7.6 (7.4–7.9) | 6.3 (5.3–7.2) | 8.7 (7.6–9.8) | 11.8 (10.7–12.9) | 13.4 (12.2–14.6) |
| 4 | | 4.7 (4.4–5) | 5.3 (5–5.6) | 6.1 (5.8–6.3) | 6.9 (6.6–7.2) | 6.4 (5.4–7.3) | 8.4 (7.3–9.5) | 10.3 (9.2–11.4) | 12.5 (11.4–13.7) |

**eTable 28. Estimates for marginal predictions of annual social welfare benefits among parents exposed to child's substance use disorder (F10–F19) and unexposed parents stratified by the child's age at diagnosis**

| | | Finland | | | | Denmark | | | |
|---|---|---|---|---|---|---|---|---|---|
| | | Men | | Women | | Men | | Women | |
| | | Unexposed | Exposed | Unexposed | Exposed | Unexposed | Exposed | Unexposed | Exposed |
| Years since diagnosis | Age group | Marg. pred. (95% CI) | Marg. pred. (95% CI) | Marg. pred. (95% CI) | Marg. pred. (95% CI) | Marg. pred. (95% CI) | Marg. pred. (95% CI) | Marg. pred. (95% CI) | Marg. pred. (95% CI) |
| | | Unexposed | Exposed | Unexposed | Exposed | Unexposed | Exposed | Unexposed | Exposed |
| 5 | | 4.8 (4.5–5.1) | 5.3 (5–5.6) | 5.8 (5.5–6) | 6.5 (6.2–6.7) | 6.7 (5.7–7.7) | 8.8 (7.6–9.9) | 9.9 (8.8–11) | 12.5 (11.2–13.7) |
| -5 | 15–19 years | 3.6 (3.5–3.7) | 4.3 (4.2–4.4) | 7.6 (7.5–7.7) | 8.5 (8.4–8.6) | 5.1 (4.9–5.3) | 6.8 (6.5–7) | 10.8 (10.6–11) | 13 (12.7–13.2) |
| -4 | | 3.8 (3.7–3.9) | 4.5 (4.4–4.6) | 7.4 (7.3–7.5) | 8.4 (8.3–8.5) | 5.2 (5–5.4) | 7 (6.8–7.2) | 10.5 (10.3–10.8) | 12.7 (12.5–13) |
| -3 | | 4 (3.9–4.1) | 4.7 (4.6–4.8) | 7.2 (7.1–7.3) | 8.2 (8.1–8.3) | 5.4 (5.2–5.6) | 7.2 (7–7.4) | 10.3 (10.1–10.6) | 12.5 (12.2–12.7) |
| -2 | | 4.2 (4.1–4.3) | 4.9 (4.8–5) | 6.8 (6.7–6.9) | 7.8 (7.7–7.9) | 5.5 (5.3–5.7) | 7.4 (7.2–7.6) | 10.3 (10.1–10.5) | 12.2 (12–12.5) |
| -1 | | 4.3 (4.2–4.4) | 5 (4.9–5.1) | 6.3 (6.2–6.4) | 7.3 (7.2–7.4) | 5.7 (5.5–5.9) | 7.4 (7.2–7.6) | 9.9 (9.6–10.1) | 11.9 (11.6–12.1) |
| 0 | | 4.4 (4.3–4.5) | 5.2 (5–5.3) | 5.8 (5.7–5.9) | 6.8 (6.7–6.9) | 5.9 (5.7–6.1) | 7.5 (7.3–7.7) | 9.3 (9.1–9.5) | 11.4 (11.1–11.6) |
| 1 | | 4.7 (4.5–4.8) | 5.4 (5.2–5.5) | 5.5 (5.4–5.6) | 6.4 (6.3–6.5) | 6.1 (5.8–6.3) | 7.6 (7.4–7.9) | 8.8 (8.6–9.1) | 11 (10.8–11.3) |
| 2 | | 4.9 (4.8–5.1) | 5.6 (5.5–5.8) | 5.2 (5.1–5.3) | 6.1 (6–6.2) | 6.1 (5.9–6.3) | 7.8 (7.6–8) | 8.4 (8.2–8.6) | 10.7 (10.5–11) |
| 3 | | 5.3 (5.2–5.5) | 6 (5.9–6.1) | 5 (4.9–5.1) | 5.9 (5.8–6.1) | 6.3 (6.1–6.5) | 8 (7.7–8.2) | 8.3 (8.1–8.5) | 10.6 (10.4–10.9) |
| 4 | | 5.7 (5.6–5.9) | 6.3 (6.2–6.5) | 5 (4.9–5.1) | 6 (5.9–6.1) | 6.6 (6.3–6.8) | 8.2 (8–8.5) | 8.3 (8–8.5) | 10.7 (10.4–10.9) |

**eTable 28. Estimates for marginal predictions of annual social welfare benefits among parents exposed to child's substance use disorder (F10–F19) and unexposed parents stratified by the child's age at diagnosis**

| | | Finland | | | | Denmark | | | |
|---|---|---|---|---|---|---|---|---|---|
| | | Men | | Women | | Men | | Women | |
| | | Unexposed | Exposed | Unexposed | Exposed | Unexposed | Exposed | Unexposed | Exposed |
| Years since diagnosis | Age group | Marg. pred. (95% CI) | Marg. pred. (95% CI) | Marg. pred. (95% CI) | Marg. pred. (95% CI) | Marg. pred. (95% CI) | Marg. pred. (95% CI) | Marg. pred. (95% CI) | Marg. pred. (95% CI) |
| | | Unexposed | Exposed | Unexposed | Exposed | Unexposed | Exposed | Unexposed | Exposed |
| 5 | | 6.2 (6.1–6.4) | 6.8 (6.7–7) | 5.1 (5–5.2) | 6 (5.9–6.1) | 6.9 (6.7–7.2) | 8.6 (8.3–8.8) | 8.4 (8.1–8.6) | 10.8 (10.6–11.1) |
| -5 | 20–25 years | 4.1 (4–4.2) | 4.6 (4.5–4.7) | 5.5 (5.4–5.5) | 6.2 (6.1–6.3) | 4.8 (4.6–5) | 6.5 (6.3–6.6) | 8.3 (8.2–8.5) | 10.3 (10.1–10.5) |
| -4 | | 4.4 (4.3–4.5) | 4.9 (4.8–5) | 5.1 (5–5.2) | 5.9 (5.8–6) | 5 (4.9–5.2) | 6.7 (6.5–6.8) | 8.1 (7.9–8.2) | 10 (9.8–10.1) |
| -3 | | 4.7 (4.6–4.8) | 5.2 (5.1–5.3) | 4.8 (4.7–4.9) | 5.6 (5.5–5.6) | 5.2 (5.1–5.4) | 6.9 (6.7–7) | 7.8 (7.6–7.9) | 9.6 (9.4–9.8) |
| -2 | | 5.1 (4.9–5.2) | 5.5 (5.4–5.6) | 4.7 (4.6–4.8) | 5.4 (5.3–5.5) | 5.4 (5.2–5.6) | 7.1 (6.9–7.3) | 7.6 (7.4–7.7) | 9.4 (9.2–9.5) |
| -1 | | 5.5 (5.3–5.6) | 5.9 (5.7–6) | 4.7 (4.6–4.8) | 5.4 (5.3–5.5) | 5.6 (5.5–5.8) | 7.3 (7.1–7.5) | 7.6 (7.4–7.7) | 9.3 (9.1–9.5) |
| 0 | | 5.9 (5.8–6.1) | 6.3 (6.1–6.4) | 4.8 (4.7–4.8) | 5.4 (5.4–5.5) | 6 (5.8–6.2) | 7.6 (7.4–7.8) | 7.6 (7.5–7.8) | 9.4 (9.2–9.6) |
| 1 | | 6.5 (6.4–6.7) | 6.7 (6.6–6.9) | 4.9 (4.8–5) | 5.6 (5.5–5.6) | 6.3 (6.1–6.5) | 7.9 (7.7–8.1) | 7.7 (7.6–7.9) | 9.5 (9.3–9.7) |
| 2 | | 7.1 (7–7.3) | 7.4 (7.2–7.5) | 5.1 (5–5.2) | 5.8 (5.7–5.9) | 6.7 (6.5–6.9) | 8.3 (8.1–8.5) | 8 (7.8–8.1) | 9.7 (9.5–9.9) |
| 3 | | 7.9 (7.7–8) | 7.9 (7.8–8.1) | 5.4 (5.3–5.5) | 6.1 (6–6.2) | 7 (6.9–7.2) | 8.7 (8.5–8.9) | 8.1 (7.9–8.3) | 9.9 (9.7–10.1) |
| 4 | | 8.6 (8.5–8.8) | 8.7 (8.5–8.9) | 5.8 (5.7–5.9) | 6.4 (6.3–6.5) | 7.4 (7.2–7.6) | 9 (8.8–9.3) | 8.4 (8.2–8.6) | 10.1 (9.9–10.4) |
| 5 | | 9.6 (9.4–9.8) | 9.5 (9.4–9.7) | 6.2 (6.1–6.3) | 6.7 (6.6–6.8) | 8 (7.8–8.2) | 9.5 (9.3–9.7) | 8.8 (8.6–9) | 10.5 (10.2–10.7) |

**eTable 28. Estimates for marginal predictions of annual social welfare benefits among parents exposed to child's substance use disorder (F10–F19) and unexposed parents stratified by the child’s age at diagnosis**

| | | Finland | | | | Denmark | | | |
|---|---|---|---|---|---|---|---|---|---|
| | | Men | | Women | | Men | | Women | |
| | | Unexposed | Exposed | Unexposed | Exposed | Unexposed | Exposed | Unexposed | Exposed |
| Years since diagnosis | Age group | Marg. pred. (95% CI) | Marg. pred. (95% CI) | Marg. pred. (95% CI) | Marg. pred. (95% CI) | Marg. pred. (95% CI) | Marg. pred. (95% CI) | Marg. pred. (95% CI) | Marg. pred. (95% CI) |
| | | Unexposed | Exposed | Unexposed | Exposed | Unexposed | Exposed | Unexposed | Exposed |

Marg. pred. = Marginal predictions in thousand euros (€)

**eTable 29. Estimates for marginal predictions of annual employment among parents exposed to child's substance use disorder (F10–F19) and unexposed parents stratified by the child’s age at diagnosis**

| | | Finland | | | | Denmark | | | |
|---|---|---|---|---|---|---|---|---|---|
| | | Men | | Women | | Men | | Women | |
| | | Unexposed | Exposed | Unexposed | Exposed | Unexposed | Exposed | Unexposed | Exposed |
| Years since diagnosis | Age group | Marg. pred. (95% CI) | Marg. pred. (95% CI) | Marg. pred. (95% CI) | Marg. pred. (95% CI) | Marg. pred. (95% CI) | Marg. pred. (95% CI) | Marg. pred. (95% CI) | Marg. pred. (95% CI) |
| | | Unexposed | Exposed | Unexposed | Exposed | Unexposed | Exposed | Unexposed | Exposed |
| -5 | 5–14 years | 0.79 (0.78–0.8) | 0.74 (0.73–0.76) | 0.71 (0.7–0.72) | 0.68 (0.67–0.7) | 0.68 (0.64–0.72) | 0.61 (0.56–0.65) | 0.6 (0.55–0.64) | 0.57 (0.53–0.61) |
| -4 | | 0.79 (0.78–0.8) | 0.74 (0.73–0.75) | 0.72 (0.7–0.73) | 0.69 (0.67–0.7) | 0.63 (0.59–0.67) | 0.62 (0.58–0.66) | 0.61 (0.57–0.65) | 0.58 (0.53–0.62) |
| -3 | | 0.79 (0.78–0.8) | 0.73 (0.71–0.74) | 0.73 (0.72–0.74) | 0.69 (0.68–0.71) | 0.64 (0.6–0.68) | 0.62 (0.58–0.66) | 0.63 (0.59–0.67) | 0.56 (0.52–0.6) |
| -2 | | 0.78 (0.77–0.8) | 0.73 (0.72–0.75) | 0.74 (0.73–0.75) | 0.71 (0.7–0.73) | 0.63 (0.59–0.68) | 0.63 (0.59–0.68) | 0.61 (0.57–0.66) | 0.57 (0.53–0.61) |
| -1 | | 0.78 (0.77–0.79) | 0.74 (0.72–0.75) | 0.76 (0.75–0.77) | 0.73 (0.72–0.74) | 0.64 (0.6–0.68) | 0.62 (0.58–0.66) | 0.61 (0.56–0.65) | 0.56 (0.51–0.6) |
| 0 | | 0.78 (0.77–0.8) | 0.73 (0.71–0.74) | 0.77 (0.76–0.78) | 0.74 (0.72–0.75) | 0.62 (0.58–0.66) | 0.6 (0.56–0.65) | 0.62 (0.57–0.66) | 0.56 (0.52–0.6) |
| 1 | | 0.77 (0.76–0.78) | 0.73 (0.72–0.74) | 0.77 (0.76–0.78) | 0.74 (0.73–0.75) | 0.66 (0.62–0.71) | 0.59 (0.54–0.63) | 0.62 (0.58–0.66) | 0.54 (0.5–0.59) |
| 2 | | 0.76 (0.75–0.78) | 0.72 (0.71–0.74) | 0.77 (0.76–0.79) | 0.74 (0.72–0.75) | 0.63 (0.59–0.68) | 0.55 (0.51–0.6) | 0.59 (0.55–0.64) | 0.54 (0.5–0.59) |
| 3 | | 0.77 (0.75–0.78) | 0.7 (0.69–0.72) | 0.77 (0.76–0.79) | 0.75 (0.73–0.76) | 0.64 (0.59–0.68) | 0.54 (0.49–0.59) | 0.57 (0.53–0.62) | 0.54 (0.5–0.59) |

**eTable 29. Estimates for marginal predictions of annual employment among parents exposed to child's substance use disorder (F10–F19) and unexposed parents stratified by the child's age at diagnosis**

| | | Finland | | | | Denmark | | | |
|---|---|---|---|---|---|---|---|---|---|
| | | Men | | Women | | Men | | Women | |
| | | Unexposed | Exposed | Unexposed | Exposed | Unexposed | Exposed | Unexposed | Exposed |
| Years since diagnosis | Age group | Marg. pred. (95% CI) | Marg. pred. (95% CI) | Marg. pred. (95% CI) | Marg. pred. (95% CI) | Marg. pred. (95% CI) | Marg. pred. (95% CI) | Marg. pred. (95% CI) | Marg. pred. (95% CI) |
| | | Unexposed | Exposed | Unexposed | Exposed | Unexposed | Exposed | Unexposed | Exposed |
| 4 | | 0.76 (0.74–0.77) | 0.7 (0.69–0.72) | 0.78 (0.77–0.79) | 0.75 (0.73–0.76) | 0.63 (0.59–0.68) | 0.54 (0.49–0.59) | 0.62 (0.57–0.66) | 0.54 (0.5–0.59) |
| 5 | | 0.74 (0.72–0.75) | 0.69 (0.68–0.71) | 0.76 (0.75–0.78) | 0.75 (0.74–0.77) | 0.62 (0.57–0.67) | 0.53 (0.48–0.58) | 0.62 (0.57–0.66) | 0.53 (0.48–0.58) |
| -5 | 15–19 years | 0.79 (0.78–0.79) | 0.73 (0.73–0.74) | 0.76 (0.75–0.76) | 0.72 (0.71–0.72) | 0.69 (0.68–0.7) | 0.64 (0.63–0.65) | 0.64 (0.63–0.65) | 0.58 (0.57–0.59) |
| -4 | | 0.78 (0.78–0.79) | 0.73 (0.72–0.73) | 0.76 (0.76–0.77) | 0.72 (0.71–0.72) | 0.7 (0.69–0.7) | 0.64 (0.63–0.65) | 0.66 (0.65–0.67) | 0.59 (0.58–0.6) |
| -3 | | 0.77 (0.77–0.78) | 0.71 (0.71–0.72) | 0.76 (0.76–0.77) | 0.72 (0.72–0.73) | 0.7 (0.69–0.71) | 0.64 (0.63–0.65) | 0.67 (0.66–0.68) | 0.6 (0.59–0.61) |
| -2 | | 0.77 (0.76–0.77) | 0.71 (0.7–0.72) | 0.77 (0.76–0.78) | 0.73 (0.72–0.73) | 0.69 (0.68–0.7) | 0.63 (0.62–0.64) | 0.66 (0.65–0.67) | 0.6 (0.59–0.61) |
| -1 | | 0.76 (0.76–0.77) | 0.7 (0.7–0.71) | 0.78 (0.77–0.78) | 0.73 (0.72–0.73) | 0.68 (0.67–0.69) | 0.63 (0.62–0.64) | 0.66 (0.65–0.67) | 0.6 (0.59–0.61) |
| 0 | | 0.76 (0.75–0.76) | 0.7 (0.69–0.7) | 0.77 (0.77–0.78) | 0.73 (0.73–0.74) | 0.67 (0.66–0.68) | 0.62 (0.61–0.63) | 0.66 (0.65–0.67) | 0.6 (0.59–0.61) |
| 1 | | 0.74 (0.74–0.75) | 0.69 (0.68–0.69) | 0.77 (0.77–0.78) | 0.73 (0.72–0.73) | 0.66 (0.65–0.68) | 0.61 (0.6–0.62) | 0.65 (0.64–0.66) | 0.59 (0.58–0.6) |

**eTable 29. Estimates for marginal predictions of annual employment among parents exposed to child's substance use disorder (F10–F19) and unexposed parents stratified by the child's age at diagnosis**

| | | Finland | | | | Denmark | | | |
|---|---|---|---|---|---|---|---|---|---|
| | | Men | | Women | | Men | | Women | |
| | | Unexposed | Exposed | Unexposed | Exposed | Unexposed | Exposed | Unexposed | Exposed |
| Years since diagnosis | Age group | Marg. pred. (95% CI) | Marg. pred. (95% CI) | Marg. pred. (95% CI) | Marg. pred. (95% CI) | Marg. pred. (95% CI) | Marg. pred. (95% CI) | Marg. pred. (95% CI) | Marg. pred. (95% CI) |
| | | Unexposed | Exposed | Unexposed | Exposed | Unexposed | Exposed | Unexposed | Exposed |
| 2 | | 0.73 (0.73–0.74) | 0.67 (0.66–0.68) | 0.77 (0.77–0.78) | 0.73 (0.72–0.73) | 0.66 (0.65–0.67) | 0.6 (0.59–0.61) | 0.65 (0.64–0.66) | 0.59 (0.58–0.6) |
| 3 | | 0.72 (0.71–0.72) | 0.66 (0.65–0.66) | 0.77 (0.76–0.78) | 0.72 (0.72–0.73) | 0.65 (0.64–0.66) | 0.59 (0.58–0.6) | 0.64 (0.63–0.65) | 0.58 (0.57–0.59) |
| 4 | | 0.7 (0.69–0.71) | 0.64 (0.64–0.65) | 0.76 (0.75–0.77) | 0.71 (0.71–0.72) | 0.64 (0.63–0.65) | 0.58 (0.57–0.59) | 0.64 (0.62–0.65) | 0.57 (0.56–0.58) |
| 5 | | 0.68 (0.67–0.69) | 0.63 (0.62–0.63) | 0.76 (0.75–0.76) | 0.71 (0.71–0.72) | 0.63 (0.61–0.64) | 0.57 (0.56–0.58) | 0.62 (0.61–0.63) | 0.56 (0.55–0.57) |
| -5 | 20–25 years | 0.76 (0.75–0.76) | 0.71 (0.71–0.72) | 0.77 (0.77–0.78) | 0.73 (0.73–0.74) | 0.69 (0.68–0.7) | 0.63 (0.62–0.64) | 0.66 (0.65–0.66) | 0.6 (0.59–0.61) |
| -4 | | 0.74 (0.74–0.75) | 0.7 (0.69–0.71) | 0.77 (0.77–0.78) | 0.73 (0.73–0.74) | 0.7 (0.7–0.71) | 0.64 (0.63–0.65) | 0.67 (0.66–0.68) | 0.61 (0.6–0.62) |
| -3 | | 0.73 (0.72–0.74) | 0.69 (0.68–0.69) | 0.77 (0.76–0.77) | 0.73 (0.72–0.73) | 0.7 (0.7–0.71) | 0.64 (0.63–0.65) | 0.67 (0.67–0.68) | 0.61 (0.61–0.62) |
| -2 | | 0.71 (0.71–0.72) | 0.67 (0.67–0.68) | 0.76 (0.76–0.77) | 0.72 (0.71–0.73) | 0.7 (0.69–0.71) | 0.63 (0.62–0.64) | 0.67 (0.66–0.67) | 0.61 (0.6–0.62) |
| -1 | | 0.7 (0.69–0.7) | 0.66 (0.65–0.67) | 0.76 (0.75–0.76) | 0.71 (0.71–0.72) | 0.68 (0.67–0.69) | 0.62 (0.61–0.63) | 0.66 (0.65–0.66) | 0.61 (0.6–0.62) |

**eTable 29. Estimates for marginal predictions of annual employment among parents exposed to child's substance use disorder (F10–F19) and unexposed parents stratified by the child's age at diagnosis**

| | | Finland | | | | Denmark | | | |
|---|---|---|---|---|---|---|---|---|---|
| | | Men | | Women | | Men | | Women | |
| | | Unexposed | Exposed | Unexposed | Exposed | Unexposed | Exposed | Unexposed | Exposed |
| Years since diagnosis | Age group | Marg. pred. (95% CI) | Marg. pred. (95% CI) | Marg. pred. (95% CI) | Marg. pred. (95% CI) | Marg. pred. (95% CI) | Marg. pred. (95% CI) | Marg. pred. (95% CI) | Marg. pred. (95% CI) |
| | | Unexposed | Exposed | Unexposed | Exposed | Unexposed | Exposed | Unexposed | Exposed |
| 0 | | 0.68 (0.67–0.69) | 0.65 (0.64–0.65) | 0.75 (0.74–0.75) | 0.71 (0.7–0.71) | 0.67 (0.66–0.68) | 0.61 (0.6–0.62) | 0.65 (0.64–0.66) | 0.59 (0.59–0.6) |
| 1 | | 0.66 (0.65–0.66) | 0.63 (0.62–0.63) | 0.74 (0.73–0.74) | 0.7 (0.69–0.7) | 0.65 (0.65–0.66) | 0.59 (0.59–0.6) | 0.64 (0.63–0.65) | 0.59 (0.58–0.6) |
| 2 | | 0.63 (0.63–0.64) | 0.61 (0.6–0.61) | 0.72 (0.72–0.73) | 0.68 (0.68–0.69) | 0.64 (0.63–0.65) | 0.58 (0.57–0.59) | 0.63 (0.62–0.64) | 0.58 (0.57–0.59) |
| 3 | | 0.61 (0.6–0.61) | 0.58 (0.58–0.59) | 0.71 (0.7–0.71) | 0.67 (0.66–0.68) | 0.63 (0.62–0.64) | 0.56 (0.55–0.57) | 0.62 (0.61–0.63) | 0.57 (0.56–0.58) |
| 4 | | 0.58 (0.57–0.59) | 0.56 (0.55–0.56) | 0.69 (0.68–0.69) | 0.65 (0.65–0.66) | 0.61 (0.6–0.62) | 0.55 (0.54–0.56) | 0.6 (0.6–0.61) | 0.56 (0.55–0.57) |
| 5 | | 0.55 (0.54–0.56) | 0.53 (0.52–0.54) | 0.67 (0.66–0.67) | 0.64 (0.63–0.65) | 0.58 (0.57–0.59) | 0.53 (0.52–0.54) | 0.59 (0.58–0.6) | 0.54 (0.53–0.55) |

Marg. pred. = Marginal predictions in probabilities

**eTable 30. Estimates for marginal predictions of annual earnings among parents exposed to child's psychotic disorder (F20–F29) and unexposed parents stratified by the child's age at diagnosis**

| | | Finland | | | | Denmark | | | |
|---|---|---|---|---|---|---|---|---|---|
| | | Men | | Women | | Men | | Women | |
| | | Unexposed | Exposed | Unexposed | Exposed | Unexposed | Exposed | Unexposed | Exposed |
| Years since diagnosis | Age group | Marg. pred. (95% CI) | Marg. pred. (95% CI) | Marg. pred. (95% CI) | Marg. pred. (95% CI) | Marg. pred. (95% CI) | Marg. pred. (95% CI) | Marg. pred. (95% CI) | Marg. pred. (95% CI) |
| | | Unexposed | Exposed | Unexposed | Exposed | Unexposed | Exposed | Unexposed | Exposed |
| -5 | 5–14 years | 30.6 (29.5–31.7) | 27.2 (26.1–28.3) | 18.1 (17.4–18.9) | 15.5 (14.8–16.2) | 54.3 (52.8–55.8) | 51.9 (50.7–53.2) | 42.6 (41.9–43.4) | 41.9 (41.2–42.6) |
| -4 | | 31.1 (30–32.3) | 27.9 (26.8–29) | 19.3 (18.5–20.1) | 16.4 (15.7–17.1) | 55 (53.5–56.5) | 52.8 (51.5–54.1) | 43.4 (42.6–44.1) | 42.9 (42.2–43.6) |
| -3 | | 31.7 (30.5–32.9) | 28.1 (27–29.2) | 19.9 (19.1–20.6) | 17.2 (16.4–17.9) | 55.5 (54–57) | 53.8 (52.5–55.2) | 44 (43.2–44.8) | 43.7 (43–44.4) |
| -2 | | 32.3 (31.1–33.5) | 28.1 (26.9–29.2) | 20.6 (19.9–21.4) | 17.9 (17.1–18.6) | 56.5 (54.9–58.1) | 54.2 (52.8–55.5) | 44.9 (44.1–45.7) | 44.1 (43.4–44.9) |
| -1 | | 32.8 (31.6–34.1) | 28.3 (27.1–29.5) | 21.4 (20.6–22.2) | 18.2 (17.5–19) | 58.3 (56.6–60) | 54.9 (53.6–56.3) | 45.6 (44.8–46.5) | 44.6 (43.8–45.3) |
| 0 | | 33.3 (32.1–34.6) | 28.5 (27.3–29.7) | 22.3 (21.5–23.2) | 18.8 (18–19.6) | 58.4 (56.7–60.1) | 56.1 (54.6–57.5) | 46.1 (45.2–46.9) | 45.2 (44.4–45.9) |
| 1 | | 33 (31.7–34.3) | 28.2 (27–29.5) | 23.5 (22.7–24.3) | 19.4 (18.6–20.2) | 59.4 (57.6–61.1) | 55.9 (54.4–57.4) | 46.8 (45.9–47.7) | 45.4 (44.6–46.2) |
| 2 | | 33.5 (32.1–34.8) | 28.2 (27–29.5) | 24.4 (23.5–25.2) | 20 (19.2–20.8) | 59.5 (57.7–61.3) | 56.7 (55.1–58.2) | 47.3 (46.4–48.2) | 45.4 (44.5–46.2) |
| 3 | | 33.7 (32.3–35) | 28.4 (27.1–29.7) | 24.6 (23.8–25.5) | 20.4 (19.5–21.2) | 60 (58.2–61.8) | 57.2 (55.6–58.8) | 47.6 (46.7–48.6) | 45.5 (44.6–46.3) |

**eTable 30. Estimates for marginal predictions of annual earnings among parents exposed to child's psychotic disorder (F20–F29) and unexposed parents stratified by the child's age at diagnosis**

| | | Finland | | | | Denmark | | | |
|---|---|---|---|---|---|---|---|---|---|
| | | Men | | Women | | Men | | Women | |
| | | Unexposed | Exposed | Unexposed | Exposed | Unexposed | Exposed | Unexposed | Exposed |
| Years since diagnosis | Age group | Marg. pred. (95% CI) | Marg. pred. (95% CI) | Marg. pred. (95% CI) | Marg. pred. (95% CI) | Marg. pred. (95% CI) | Marg. pred. (95% CI) | Marg. pred. (95% CI) | Marg. pred. (95% CI) |
| | | Unexposed | Exposed | Unexposed | Exposed | Unexposed | Exposed | Unexposed | Exposed |
| 4 | | 34 (32.6–35.4) | 28.1 (26.9–29.4) | 25.2 (24.3–26.1) | 20.9 (20.1–21.8) | 59.4 (57.6–61.3) | 57.6 (55.9–59.3) | 47.5 (46.6–48.5) | 45.2 (44.3–46.1) |
| 5 | | 33.9 (32.4–35.3) | 28.8 (27.4–30.1) | 25.5 (24.6–26.4) | 21.2 (20.3–22.1) | 59.9 (58–61.9) | 57 (55.3–58.7) | 47.8 (46.8–48.8) | 45.4 (44.5–46.4) |
| -5 | 15–19 years | 34.3 (33.7–34.9) | 30.8 (30.3–31.4) | 22.2 (21.8–22.5) | 20.1 (19.7–20.4) | 55.9 (55.2–56.7) | 52.4 (51.7–53) | 43.4 (43–43.8) | 42.4 (42–42.8) |
| -4 | | 34.5 (34–35.1) | 31.1 (30.6–31.7) | 22.9 (22.6–23.3) | 20.8 (20.4–21.1) | 56.9 (56.1–57.7) | 52.8 (52.1–53.4) | 44 (43.6–44.4) | 43 (42.6–43.4) |
| -3 | | 34.9 (34.3–35.5) | 31.2 (30.7–31.8) | 23.5 (23.2–23.9) | 21.4 (21.1–21.8) | 57.4 (56.6–58.2) | 53.4 (52.7–54.1) | 44.3 (43.9–44.7) | 43.7 (43.3–44) |
| -2 | | 35.1 (34.5–35.8) | 31.5 (30.9–32.1) | 24 (23.7–24.4) | 21.9 (21.5–22.3) | 57.8 (57–58.6) | 54.1 (53.3–54.8) | 44.7 (44.3–45.1) | 43.9 (43.5–44.3) |
| -1 | | 35.3 (34.6–35.9) | 31.5 (30.8–32.1) | 24.6 (24.2–25) | 22.3 (21.9–22.7) | 58.3 (57.5–59.2) | 54.4 (53.7–55.2) | 45 (44.6–45.4) | 44 (43.6–44.4) |
| 0 | | 35.6 (35–36.2) | 31.5 (30.9–32.1) | 25.1 (24.7–25.5) | 22.6 (22.2–23) | 58.9 (58–59.7) | 54.4 (53.7–55.2) | 45.1 (44.7–45.5) | 43.8 (43.4–44.3) |
| 1 | | 35.5 (34.9–36.2) | 31.6 (30.9–32.2) | 25.4 (25–25.8) | 22.8 (22.4–23.2) | 59.2 (58.3–60) | 54.5 (53.7–55.3) | 45.2 (44.8–45.6) | 43.7 (43.3–44.1) |

**eTable 30. Estimates for marginal predictions of annual earnings among parents exposed to child's psychotic disorder (F20–F29) and unexposed parents stratified by the child's age at diagnosis**

| | | Finland | | | | Denmark | | | |
|---|---|---|---|---|---|---|---|---|---|
| | | Men | | Women | | Men | | Women | |
| | | Unexposed | Exposed | Unexposed | Exposed | Unexposed | Exposed | Unexposed | Exposed |
| Years since diagnosis | Age group | Marg. pred. (95% CI) | Marg. pred. (95% CI) | Marg. pred. (95% CI) | Marg. pred. (95% CI) | Marg. pred. (95% CI) | Marg. pred. (95% CI) | Marg. pred. (95% CI) | Marg. pred. (95% CI) |
| | | Unexposed | Exposed | Unexposed | Exposed | Unexposed | Exposed | Unexposed | Exposed |
| 2 | | 35.5 (34.8–36.2) | 31.1 (30.4–31.7) | 25.7 (25.3–26.1) | 23.1 (22.7–23.5) | 59.3 (58.4–60.1) | 54.9 (54.1–55.7) | 45.3 (44.9–45.8) | 43.6 (43.2–44) |
| 3 | | 35.2 (34.6–35.9) | 30.8 (30.2–31.5) | 25.9 (25.5–26.3) | 23.3 (22.9–23.7) | 59.3 (58.4–60.2) | 55.1 (54.3–55.9) | 45.1 (44.7–45.6) | 43.5 (43–43.9) |
| 4 | | 35 (34.2–35.7) | 30.1 (29.5–30.8) | 26.2 (25.7–26.6) | 23.5 (23.1–23.9) | 59.5 (58.5–60.4) | 54.7 (53.8–55.5) | 45.1 (44.6–45.5) | 43.4 (42.9–43.8) |
| 5 | | 34.4 (33.7–35.2) | 29.4 (28.7–30.1) | 26.1 (25.6–26.5) | 23.6 (23.1–24) | 59.7 (58.7–60.7) | 54.4 (53.5–55.2) | 45.2 (44.7–45.7) | 43.3 (42.8–43.8) |
| -5 | 20–25 years | 33.6 (33.1–34.1) | 30.7 (30.3–31.2) | 23.2 (22.9–23.5) | 21.4 (21.1–21.7) | 55.4 (54.8–56.1) | 51.4 (50.8–52) | 42 (41.7–42.3) | 40.5 (40.1–40.8) |
| -4 | | 33.6 (33.1–34.1) | 30.7 (30.2–31.2) | 23.6 (23.3–23.9) | 21.7 (21.4–22) | 56 (55.3–56.6) | 51.5 (50.9–52.1) | 42.1 (41.8–42.5) | 40.4 (40.1–40.7) |
| -3 | | 33.6 (33.1–34.1) | 30.3 (29.8–30.8) | 23.8 (23.4–24.1) | 21.8 (21.5–22.1) | 55.7 (55.1–56.4) | 51.7 (51.1–52.3) | 42.1 (41.7–42.4) | 40.3 (40–40.6) |
| -2 | | 33.1 (32.6–33.6) | 30 (29.5–30.5) | 23.9 (23.6–24.2) | 21.9 (21.6–22.2) | 55.8 (55.1–56.5) | 51.6 (51–52.3) | 42.1 (41.8–42.5) | 40.3 (40–40.6) |
| -1 | | 32.8 (32.3–33.3) | 29.6 (29.1–30.1) | 24 (23.7–24.3) | 21.9 (21.6–22.2) | 55.9 (55.2–56.6) | 51.6 (51–52.3) | 42.2 (41.9–42.6) | 40.3 (39.9–40.6) |

**eTable 30. Estimates for marginal predictions of annual earnings among parents exposed to child's psychotic disorder (F20–F29) and unexposed parents stratified by the child's age at diagnosis**

| | | Finland | | | | Denmark | | | |
|---|---|---|---|---|---|---|---|---|---|
| | | Men | | Women | | Men | | Women | |
| | | Unexposed | Exposed | Unexposed | Exposed | Unexposed | Exposed | Unexposed | Exposed |
| Years since diagnosis | Age group | Marg. pred. (95% CI) | Marg. pred. (95% CI) | Marg. pred. (95% CI) | Marg. pred. (95% CI) | Marg. pred. (95% CI) | Marg. pred. (95% CI) | Marg. pred. (95% CI) | Marg. pred. (95% CI) |
| | | Unexposed | Exposed | Unexposed | Exposed | Unexposed | Exposed | Unexposed | Exposed |
| 0 | | 32.3 (31.8–32.9) | 29.2 (28.7–29.7) | 24 (23.7–24.3) | 21.8 (21.5–22.1) | 56.1 (55.4–56.8) | 51.6 (50.9–52.2) | 42.4 (42.1–42.8) | 40.3 (40–40.7) |
| 1 | | 31.7 (31.1–32.2) | 28.5 (27.9–29) | 23.9 (23.6–24.3) | 21.6 (21.3–21.9) | 55.9 (55.2–56.6) | 51.5 (50.8–52.1) | 42.3 (42–42.7) | 40.4 (40–40.7) |
| 2 | | 30.9 (30.4–31.5) | 27.8 (27.3–28.3) | 23.7 (23.4–24.1) | 21.5 (21.2–21.8) | 55.9 (55.1–56.6) | 51.1 (50.5–51.8) | 42.3 (41.9–42.6) | 40.4 (40.1–40.8) |
| 3 | | 30 (29.4–30.5) | 27 (26.4–27.5) | 23.5 (23.1–23.8) | 21.2 (20.9–21.6) | 55.7 (55–56.5) | 51.1 (50.5–51.8) | 42.5 (42.1–42.8) | 40.3 (40–40.7) |
| 4 | | 28.9 (28.4–29.5) | 26 (25.4–26.6) | 23 (22.6–23.3) | 20.8 (20.5–21.2) | 55.4 (54.6–56.1) | 50.6 (49.9–51.3) | 42.3 (41.9–42.7) | 40.4 (40–40.8) |
| 5 | | 27.6 (27–28.3) | 24.7 (24.1–25.3) | 22.6 (22.2–23) | 20.4 (20–20.7) | 55 (54.2–55.8) | 50.2 (49.5–50.9) | 42.2 (41.8–42.6) | 40.4 (40–40.7) |

Marg. pred. = Marginal predictions in thousand euros (€)

**eTable 31. Estimates for marginal predictions of annual disposable income among parents exposed to child's psychotic disorder (F20–F29) and unexposed parents stratified by the child's age at diagnosis**

| | | Finland | | | | Denmark | | | |
|---|---|---|---|---|---|---|---|---|---|
| | | Men | | Women | | Men | | Women | |
| | | Unexposed | Exposed | Unexposed | Exposed | Unexposed | Exposed | Unexposed | Exposed |
| Years since diagnosis | Age group | Marg. pred. (95% CI) | Marg. pred. (95% CI) | Marg. pred. (95% CI) | Marg. pred. (95% CI) | Marg. pred. (95% CI) | Marg. pred. (95% CI) | Marg. pred. (95% CI) | Marg. pred. (95% CI) |
| | | Unexposed | Exposed | Unexposed | Exposed | Unexposed | Exposed | Unexposed | Exposed |
| -5 | 5–14 years | 25.9 (25.2–26.7) | 24.2 (23.5–24.9) | 22 (21.6–22.5) | 20.7 (20.3–21.1) | 33 (32.2–33.7) | 31.4 (30.7–32.1) | 29.8 (29.3–30.2) | 28.9 (28.5–29.3) |
| -4 | | 26.5 (25.7–27.3) | 24.6 (23.9–25.3) | 22.7 (22.3–23.2) | 21.3 (20.9–21.7) | 33.5 (32.7–34.3) | 32.3 (31.6–33) | 30.4 (29.9–30.9) | 29.7 (29.2–30.1) |
| -3 | | 27.1 (26.3–27.9) | 25.2 (24.4–25.9) | 23.2 (22.8–23.7) | 22 (21.6–22.4) | 34.4 (33.6–35.3) | 33.2 (32.4–33.9) | 31 (30.5–31.5) | 30.4 (29.9–30.8) |
| -2 | | 27.7 (26.9–28.5) | 25.2 (24.4–25.9) | 23.7 (23.3–24.2) | 22.7 (22.2–23.1) | 35.2 (34.3–36.1) | 33.5 (32.8–34.3) | 31.8 (31.3–32.3) | 30.8 (30.4–31.3) |
| -1 | | 28.2 (27.4–29) | 25.4 (24.6–26.1) | 24.4 (23.9–24.9) | 22.9 (22.5–23.4) | 36.6 (35.7–37.5) | 34.4 (33.6–35.2) | 32.5 (31.9–33) | 31.3 (30.9–31.8) |
| 0 | | 28.6 (27.8–29.4) | 25.9 (25.1–26.6) | 24.8 (24.3–25.3) | 23.3 (22.9–23.7) | 37 (36.1–37.9) | 35.5 (34.7–36.4) | 32.9 (32.4–33.4) | 31.9 (31.4–32.3) |
| 1 | | 28.8 (28–29.6) | 26 (25.3–26.8) | 25.4 (25–25.9) | 23.6 (23.2–24) | 38.1 (37.1–39.1) | 35.6 (34.8–36.4) | 33.5 (33–34.1) | 32 (31.5–32.5) |
| 2 | | 29.4 (28.5–30.2) | 26 (25.3–26.8) | 25.8 (25.4–26.3) | 23.8 (23.4–24.3) | 38.5 (37.5–39.5) | 36.3 (35.4–37.2) | 33.9 (33.4–34.5) | 32.1 (31.6–32.6) |
| 3 | | 29.6 (28.7–30.5) | 26.6 (25.8–27.4) | 25.6 (25.1–26.1) | 23.7 (23.2–24.2) | 39.2 (38.1–40.2) | 37.1 (36.2–38.1) | 34.2 (33.6–34.8) | 32.2 (31.7–32.7) |

**eTable 31. Estimates for marginal predictions of annual disposable income among parents exposed to child's psychotic disorder (F20–F29) and unexposed parents stratified by the child's age at diagnosis**

| | | Finland | | | | Denmark | | | |
|---|---|---|---|---|---|---|---|---|---|
| | | Men | | Women | | Men | | Women | |
| | | Unexposed | Exposed | Unexposed | Exposed | Unexposed | Exposed | Unexposed | Exposed |
| Years since diagnosis | Age group | Marg. pred. (95% CI) | Marg. pred. (95% CI) | Marg. pred. (95% CI) | Marg. pred. (95% CI) | Marg. pred. (95% CI) | Marg. pred. (95% CI) | Marg. pred. (95% CI) | Marg. pred. (95% CI) |
| | | Unexposed | Exposed | Unexposed | Exposed | Unexposed | Exposed | Unexposed | Exposed |
| 4 | | 30.2 (29.2–31.1) | 26.6 (25.8–27.4) | 25.5 (25–26) | 23.8 (23.3–24.3) | 39.1 (38.1–40.2) | 37.7 (36.7–38.7) | 34.1 (33.5–34.7) | 32 (31.4–32.6) |
| 5 | | 30.5 (29.5–31.5) | 26.8 (26–27.7) | 25.6 (25.1–26.1) | 23.8 (23.3–24.3) | 40.2 (39–41.3) | 37.9 (36.9–39) | 34.4 (33.7–35.1) | 32.2 (31.6–32.8) |
| -5 | 15–19 years | 28.2 (27.8–28.6) | 26.3 (25.9–26.7) | 23.4 (23.2–23.6) | 22.5 (22.3–22.8) | 34 (33.6–34.4) | 31.9 (31.5–32.2) | 30 (29.7–30.2) | 29.2 (29–29.4) |
| -4 | | 28.6 (28.2–29) | 26.7 (26.4–27.1) | 23.9 (23.7–24.2) | 22.9 (22.7–23.1) | 35 (34.5–35.4) | 32.4 (32.1–32.8) | 30.5 (30.3–30.7) | 29.7 (29.5–29.9) |
| -3 | | 29.2 (28.8–29.6) | 27 (26.6–27.3) | 24.2 (24–24.5) | 23.2 (23–23.4) | 35.5 (35.1–36) | 33.2 (32.8–33.6) | 30.9 (30.6–31.1) | 30.3 (30–30.5) |
| -2 | | 29.6 (29.2–30) | 27.5 (27.1–27.9) | 24.2 (24–24.4) | 23.3 (23–23.5) | 36.2 (35.7–36.7) | 33.8 (33.4–34.2) | 31.3 (31–31.5) | 30.5 (30.3–30.7) |
| -1 | | 29.8 (29.4–30.3) | 27.7 (27.3–28.1) | 24.2 (24–24.5) | 23.2 (22.9–23.4) | 36.9 (36.4–37.4) | 34.4 (34–34.8) | 31.5 (31.2–31.7) | 30.6 (30.4–30.9) |
| 0 | | 30.2 (29.8–30.7) | 28 (27.6–28.4) | 24.2 (24–24.4) | 23 (22.8–23.2) | 37.6 (37.1–38) | 34.8 (34.4–35.2) | 31.6 (31.3–31.9) | 30.5 (30.3–30.8) |
| 1 | | 30.6 (30.2–31) | 28.4 (28–28.8) | 24.1 (23.9–24.3) | 22.9 (22.7–23.1) | 38.3 (37.8–38.7) | 35.4 (34.9–35.8) | 31.7 (31.4–32) | 30.5 (30.2–30.7) |

**eTable 31. Estimates for marginal predictions of annual disposable income among parents exposed to child's psychotic disorder (F20–F29) and unexposed parents stratified by the child's age at diagnosis**

| | | Finland | | | | Denmark | | | |
|---|---|---|---|---|---|---|---|---|---|
| | | Men | | Women | | Men | | Women | |
| | | Unexposed | Exposed | Unexposed | Exposed | Unexposed | Exposed | Unexposed | Exposed |
| Years since diagnosis | Age group | Marg. pred. (95% CI) | Marg. pred. (95% CI) | Marg. pred. (95% CI) | Marg. pred. (95% CI) | Marg. pred. (95% CI) | Marg. pred. (95% CI) | Marg. pred. (95% CI) | Marg. pred. (95% CI) |
| | | Unexposed | Exposed | Unexposed | Exposed | Unexposed | Exposed | Unexposed | Exposed |
| 2 | | 31 (30.6–31.5) | 28.4 (28–28.8) | 24.2 (23.9–24.4) | 22.9 (22.7–23.1) | 38.8 (38.3–39.3) | 35.9 (35.5–36.4) | 31.9 (31.6–32.1) | 30.5 (30.2–30.8) |
| 3 | | 31.3 (30.8–31.8) | 28.6 (28.2–29.1) | 24.2 (24–24.5) | 23 (22.8–23.3) | 39.2 (38.7–39.8) | 36.5 (36.1–37) | 31.8 (31.6–32.1) | 30.6 (30.3–30.8) |
| 4 | | 31.5 (31–32) | 28.6 (28.2–29.1) | 24.5 (24.2–24.7) | 23.2 (23–23.5) | 39.8 (39.3–40.4) | 36.7 (36.2–37.2) | 32 (31.7–32.3) | 30.6 (30.3–30.9) |
| 5 | | 31.8 (31.3–32.3) | 28.7 (28.3–29.2) | 24.5 (24.3–24.8) | 23.4 (23.1–23.6) | 40.4 (39.8–41) | 37 (36.5–37.6) | 32.4 (32.1–32.7) | 30.8 (30.5–31.1) |
| -5 | 20–25 years | 28.3 (28–28.6) | 26.5 (26.2–26.9) | 22.1 (22–22.3) | 21.5 (21.3–21.7) | 34.1 (33.7–34.5) | 31.8 (31.4–32.1) | 28.5 (28.3–28.7) | 27.5 (27.3–27.7) |
| -4 | | 28.6 (28.2–28.9) | 26.9 (26.6–27.2) | 22.2 (22–22.4) | 21.5 (21.3–21.6) | 34.9 (34.6–35.3) | 32.3 (31.9–32.6) | 28.7 (28.5–28.9) | 27.6 (27.4–27.8) |
| -3 | | 28.8 (28.5–29.2) | 27 (26.7–27.3) | 22.1 (22–22.3) | 21.4 (21.2–21.6) | 35.2 (34.9–35.6) | 32.8 (32.4–33.1) | 28.7 (28.5–28.9) | 27.5 (27.3–27.7) |
| -2 | | 28.8 (28.5–29.2) | 27.3 (26.9–27.6) | 22.2 (22–22.4) | 21.5 (21.3–21.7) | 35.7 (35.3–36.1) | 33.1 (32.8–33.5) | 28.8 (28.6–29) | 27.5 (27.3–27.7) |
| -1 | | 29 (28.6–29.3) | 27.4 (27.1–27.7) | 22.3 (22.1–22.5) | 21.5 (21.4–21.7) | 36.2 (35.8–36.5) | 33.5 (33.2–33.9) | 29 (28.8–29.2) | 27.7 (27.5–27.9) |

**eTable 31. Estimates for marginal predictions of annual disposable income among parents exposed to child's psychotic disorder (F20–F29) and unexposed parents stratified by the child’s age at diagnosis**

| | | Finland | | | | Denmark | | | |
|---|---|---|---|---|---|---|---|---|---|
| | | Men | | Women | | Men | | Women | |
| | | Unexposed | Exposed | Unexposed | Exposed | Unexposed | Exposed | Unexposed | Exposed |
| Years since diagnosis | Age group | Marg. pred. (95% CI) | Marg. pred. (95% CI) | Marg. pred. (95% CI) | Marg. pred. (95% CI) | Marg. pred. (95% CI) | Marg. pred. (95% CI) | Marg. pred. (95% CI) | Marg. pred. (95% CI) |
| | | Unexposed | Exposed | Unexposed | Exposed | Unexposed | Exposed | Unexposed | Exposed |
| 0 | | 29.3 (28.9–29.6) | 27.5 (27.2–27.9) | 22.5 (22.3–22.7) | 21.6 (21.4–21.8) | 36.7 (36.3–37.1) | 33.8 (33.5–34.2) | 29.3 (29.1–29.5) | 27.9 (27.7–28.1) |
| 1 | | 29.4 (29.1–29.8) | 27.6 (27.2–27.9) | 22.6 (22.4–22.8) | 21.7 (21.5–21.9) | 37 (36.6–37.4) | 34.2 (33.8–34.6) | 29.4 (29.2–29.6) | 28.1 (27.8–28.3) |
| 2 | | 29.5 (29.1–29.8) | 27.7 (27.4–28.1) | 22.7 (22.5–22.9) | 21.8 (21.7–22) | 37.4 (37–37.8) | 34.4 (34–34.8) | 29.6 (29.4–29.8) | 28.2 (28–28.5) |
| 3 | | 29.6 (29.3–30) | 27.9 (27.6–28.3) | 22.9 (22.8–23.1) | 21.9 (21.8–22.1) | 37.7 (37.3–38.2) | 34.8 (34.4–35.2) | 29.9 (29.7–30.2) | 28.4 (28.2–28.6) |
| 4 | | 29.7 (29.3–30) | 28.2 (27.8–28.5) | 23 (22.8–23.2) | 22.1 (21.9–22.3) | 37.9 (37.5–38.4) | 34.8 (34.4–35.3) | 30 (29.8–30.3) | 28.7 (28.4–28.9) |
| 5 | | 29.7 (29.3–30.1) | 28.2 (27.8–28.6) | 23.2 (22.9–23.4) | 22.2 (22–22.4) | 38.2 (37.8–38.7) | 35 (34.6–35.4) | 30.3 (30–30.5) | 28.8 (28.6–29.1) |

Marg. pred. = Marginal predictions in thousand euros (€)

**eTable 32. Estimates for marginal predictions of annual social welfare benefits among parents exposed to child's psychotic disorder (F20–F29) and unexposed parents stratified by the child's age at diagnosis**

| | | Finland | | | | Denmark | | | |
|---|---|---|---|---|---|---|---|---|---|
| | | Men | | Women | | Men | | Women | |
| | | Unexposed | Exposed | Unexposed | Exposed | Unexposed | Exposed | Unexposed | Exposed |
| Years since diagnosis | Age group | Marg. pred. (95% CI) | Marg. pred. (95% CI) | Marg. pred. (95% CI) | Marg. pred. (95% CI) | Marg. pred. (95% CI) | Marg. pred. (95% CI) | Marg. pred. (95% CI) | Marg. pred. (95% CI) |
| | | Unexposed | Exposed | Unexposed | Exposed | Unexposed | Exposed | Unexposed | Exposed |
| -5 | 5–14 years | 3.6 (3.3–3.9) | 4.4 (4.1–4.7) | 8.8 (8.5–9.2) | 9.6 (9.3–9.9) | 4.6 (4.2–5) | 5.2 (4.8–5.6) | 10.9 (10.5–11.4) | 12.1 (11.6–12.5) |
| -4 | | 3.7 (3.4–4) | 4.4 (4.1–4.7) | 8.6 (8.3–9) | 9.6 (9.2–9.9) | 4.7 (4.4–5.1) | 5.3 (4.9–5.7) | 10.6 (10.1–11) | 11.5 (11–12) |
| -3 | | 3.8 (3.5–4.1) | 4.6 (4.3–5) | 8.6 (8.2–8.9) | 9.6 (9.2–9.9) | 5 (4.6–5.4) | 5.4 (5–5.8) | 10.5 (10.1–11) | 11.6 (11.2–12.1) |
| -2 | | 4 (3.6–4.3) | 4.8 (4.5–5.1) | 8.6 (8.2–8.9) | 9.6 (9.2–9.9) | 5 (4.6–5.4) | 5.6 (5.2–6) | 10.6 (10.1–11) | 11.6 (11.1–12) |
| -1 | | 4.1 (3.8–4.5) | 4.9 (4.6–5.3) | 8.5 (8.2–8.9) | 9.6 (9.2–10) | 5.2 (4.8–5.6) | 5.7 (5.3–6.1) | 10.2 (9.8–10.7) | 11.5 (11–12) |
| 0 | | 4.4 (4–4.7) | 5.2 (4.9–5.5) | 8.3 (7.9–8.6) | 9.5 (9.1–9.9) | 5.2 (4.8–5.6) | 5.6 (5.2–6.1) | 10.2 (9.7–10.7) | 11.5 (11–12) |
| 1 | | 4.6 (4.3–5) | 5.5 (5.2–5.9) | 8 (7.6–8.3) | 9.4 (9–9.7) | 5.2 (4.8–5.6) | 5.8 (5.4–6.3) | 10.1 (9.6–10.6) | 11.2 (10.7–11.7) |
| 2 | | 4.8 (4.4–5.2) | 5.8 (5.4–6.2) | 7.7 (7.3–8.1) | 9 (8.7–9.4) | 5.2 (4.8–5.7) | 6 (5.5–6.4) | 9.7 (9.3–10.2) | 11 (10.5–11.5) |
| 3 | | 4.9 (4.5–5.3) | 6 (5.6–6.4) | 7.2 (6.8–7.6) | 8.6 (8.3–9) | 5.3 (4.9–5.8) | 6.1 (5.6–6.6) | 9.4 (8.9–9.9) | 10.5 (10–11) |
| 4 | | 5.1 (4.7–5.5) | 6.1 (5.7–6.5) | 6.7 (6.3–7.1) | 8.3 (7.9–8.7) | 5.4 (4.9–5.8) | 6.3 (5.8–6.8) | 8.7 (8.2–9.1) | 10.2 (9.6–10.7) |
| 5 | | 5.3 (4.9–5.8) | 6.1 (5.7–6.5) | 6.4 (6–6.8) | 8 (7.5–8.4) | 5.4 (5–5.9) | 6.8 (6.2–7.3) | 8 (7.5–8.5) | 10 (9.4–10.5) |
| -5 | 15–19 years | 3.3 (3.2–3.5) | 4.1 (3.9–4.2) | 7.4 (7.2–7.5) | 8.1 (8–8.3) | 4.3 (4.1–4.5) | 5.4 (5.2–5.6) | 9.6 (9.4–9.8) | 11.1 (10.9–11.4) |

**eTable 32. Estimates for marginal predictions of annual social welfare benefits among parents exposed to child's psychotic disorder (F20–F29) and unexposed parents stratified by the child's age at diagnosis**

| | | Finland | | | | Denmark | | | |
|---|---|---|---|---|---|---|---|---|---|
| | | Men | | Women | | Men | | Women | |
| | | Unexposed | Exposed | Unexposed | Exposed | Unexposed | Exposed | Unexposed | Exposed |
| Years since diagnosis | Age group | Marg. pred. (95% CI) | Marg. pred. (95% CI) | Marg. pred. (95% CI) | Marg. pred. (95% CI) | Marg. pred. (95% CI) | Marg. pred. (95% CI) | Marg. pred. (95% CI) | Marg. pred. (95% CI) |
| | | Unexposed | Exposed | Unexposed | Exposed | Unexposed | Exposed | Unexposed | Exposed |
| -4 | | 3.5 (3.3–3.7) | 4.3 (4.1–4.5) | 7.2 (7–7.4) | 8 (7.8–8.1) | 4.4 (4.2–4.6) | 5.6 (5.4–5.8) | 9.6 (9.3–9.8) | 11.1 (10.8–11.3) |
| -3 | | 3.7 (3.5–3.9) | 4.5 (4.3–4.6) | 6.9 (6.8–7.1) | 7.8 (7.6–7.9) | 4.5 (4.3–4.7) | 5.7 (5.5–5.9) | 9.3 (9.1–9.6) | 10.9 (10.7–11.2) |
| -2 | | 3.9 (3.8–4.1) | 4.7 (4.5–4.8) | 6.6 (6.4–6.7) | 7.3 (7.1–7.5) | 4.6 (4.4–4.8) | 5.8 (5.6–6.1) | 9.2 (8.9–9.4) | 10.7 (10.5–10.9) |
| -1 | | 4.1 (3.9–4.3) | 4.9 (4.7–5.1) | 6 (5.9–6.2) | 6.8 (6.6–7) | 4.7 (4.5–4.9) | 5.9 (5.7–6.1) | 8.7 (8.5–8.9) | 10.3 (10–10.5) |
| 0 | | 4.2 (4–4.4) | 5.1 (4.9–5.3) | 5.6 (5.4–5.7) | 6.4 (6.2–6.6) | 4.8 (4.6–5) | 6 (5.8–6.2) | 8.3 (8.1–8.5) | 9.8 (9.6–10.1) |
| 1 | | 4.5 (4.3–4.7) | 5.3 (5.1–5.5) | 5.2 (5.1–5.4) | 6.1 (5.9–6.2) | 4.9 (4.7–5.1) | 6.1 (5.8–6.3) | 7.8 (7.6–8) | 9.6 (9.3–9.8) |
| 2 | | 4.7 (4.5–4.9) | 5.6 (5.4–5.8) | 5 (4.8–5.1) | 5.8 (5.7–6) | 5 (4.8–5.2) | 6.2 (6–6.4) | 7.4 (7.2–7.6) | 9.2 (9–9.5) |
| 3 | | 5.1 (4.9–5.3) | 6.1 (5.9–6.3) | 4.8 (4.7–5) | 5.8 (5.6–6) | 5.2 (4.9–5.4) | 6.3 (6–6.5) | 7.1 (6.9–7.4) | 9 (8.8–9.3) |
| 4 | | 5.5 (5.3–5.7) | 6.6 (6.4–6.9) | 4.8 (4.7–5) | 5.7 (5.6–5.9) | 5.3 (5.1–5.5) | 6.5 (6.3–6.8) | 7.2 (7–7.5) | 9.1 (8.8–9.4) |
| 5 | | 6.1 (5.8–6.3) | 7.3 (7–7.5) | 5 (4.8–5.1) | 5.9 (5.7–6) | 5.5 (5.3–5.8) | 7 (6.7–7.2) | 7.3 (7–7.6) | 9.2 (8.9–9.5) |
| -5 | 20–25 years | 3.9 (3.7–4) | 4.5 (4.4–4.7) | 5.2 (5.1–5.3) | 6 (5.8–6.1) | 4.4 (4.3–4.6) | 5.9 (5.8–6.1) | 7.8 (7.6–8) | 9.5 (9.3–9.7) |
| -4 | | 4.1 (4–4.3) | 4.8 (4.7–5) | 4.9 (4.8–5) | 5.7 (5.5–5.8) | 4.6 (4.4–4.8) | 6 (5.9–6.2) | 7.4 (7.2–7.6) | 9.2 (9–9.4) |

**eTable 32. Estimates for marginal predictions of annual social welfare benefits among parents exposed to child's psychotic disorder (F20–F29) and unexposed parents stratified by the child's age at diagnosis**

| | | Finland | | | | Denmark | | | |
|---|---|---|---|---|---|---|---|---|---|
| | | Men | | Women | | Men | | Women | |
| | | Unexposed | Exposed | Unexposed | Exposed | Unexposed | Exposed | Unexposed | Exposed |
| Years since diagnosis | Age group | Marg. pred. (95% CI) | Marg. pred. (95% CI) | Marg. pred. (95% CI) | Marg. pred. (95% CI) | Marg. pred. (95% CI) | Marg. pred. (95% CI) | Marg. pred. (95% CI) | Marg. pred. (95% CI) |
| | | Unexposed | Exposed | Unexposed | Exposed | Unexposed | Exposed | Unexposed | Exposed |
| -3 | | 4.4 (4.2–4.6) | 5.2 (5–5.4) | 4.7 (4.5–4.8) | 5.4 (5.3–5.6) | 4.8 (4.6–4.9) | 6.1 (5.9–6.3) | 7.1 (6.9–7.3) | 8.9 (8.7–9.1) |
| -2 | | 4.8 (4.6–5) | 5.6 (5.4–5.8) | 4.5 (4.4–4.6) | 5.4 (5.3–5.5) | 5 (4.8–5.2) | 6.3 (6.1–6.5) | 6.8 (6.7–7) | 8.6 (8.4–8.8) |
| -1 | | 5.2 (5–5.4) | 6 (5.8–6.2) | 4.5 (4.4–4.6) | 5.4 (5.3–5.5) | 5.2 (5–5.4) | 6.5 (6.3–6.7) | 6.8 (6.6–7) | 8.6 (8.4–8.8) |
| 0 | | 5.7 (5.5–5.9) | 6.4 (6.2–6.6) | 4.6 (4.5–4.8) | 5.5 (5.4–5.6) | 5.4 (5.3–5.6) | 6.8 (6.6–7) | 6.9 (6.7–7) | 8.7 (8.5–8.9) |
| 1 | | 6.3 (6.1–6.5) | 7.1 (6.9–7.3) | 4.8 (4.7–4.9) | 5.7 (5.6–5.8) | 5.6 (5.4–5.8) | 7.2 (7–7.4) | 7 (6.8–7.2) | 8.9 (8.7–9.1) |
| 2 | | 7 (6.8–7.3) | 7.7 (7.5–7.9) | 5 (4.9–5.2) | 6 (5.8–6.1) | 6 (5.8–6.2) | 7.5 (7.3–7.7) | 7.1 (6.9–7.3) | 9.1 (8.8–9.3) |
| 3 | | 7.9 (7.7–8.2) | 8.4 (8.2–8.7) | 5.4 (5.2–5.5) | 6.2 (6.1–6.4) | 6.3 (6.1–6.5) | 8 (7.7–8.2) | 7.3 (7.1–7.5) | 9.3 (9–9.5) |
| 4 | | 8.8 (8.5–9) | 9.4 (9.1–9.6) | 5.8 (5.6–5.9) | 6.6 (6.5–6.8) | 6.8 (6.5–7) | 8.4 (8.1–8.6) | 7.7 (7.4–7.9) | 9.6 (9.3–9.8) |
| 5 | | 9.8 (9.5–10.1) | 10.3 (10.1–10.6) | 6.3 (6.1–6.4) | 7.1 (6.9–7.2) | 7.2 (7–7.4) | 8.9 (8.6–9.1) | 8 (7.7–8.2) | 10 (9.8–10.2) |

Marg. pred. = Marginal predictions in thousand euros (€)

**eTable 33. Estimates for marginal predictions of annual employment among parents exposed to child's psychotic disorder (F20–F29) and unexposed parents stratified by the child's age at diagnosis**

| | | Finland | | | | Denmark | | | |
|---|---|---|---|---|---|---|---|---|---|
| | | Men | | Women | | Men | | Women | |
| | | Unexposed | Exposed | Unexposed | Exposed | Unexposed | Exposed | Unexposed | Exposed |
| Years since diagnosis | Age group | Marg. pred. (95% CI) | Marg. pred. (95% CI) | Marg. pred. (95% CI) | Marg. pred. (95% CI) | Marg. pred. (95% CI) | Marg. pred. (95% CI) | Marg. pred. (95% CI) | Marg. pred. (95% CI) |
| | | Unexposed | Exposed | Unexposed | Exposed | Unexposed | Exposed | Unexposed | Exposed |
| -5 | 5–14 years | 0.78 (0.76–0.8) | 0.72 (0.69–0.74) | 0.7 (0.67–0.72) | 0.6 (0.58–0.63) | 0.71 (0.69–0.73) | 0.71 (0.69–0.73) | 0.66 (0.64–0.68) | 0.63 (0.61–0.65) |
| -4 | | 0.78 (0.76–0.8) | 0.7 (0.68–0.72) | 0.71 (0.69–0.73) | 0.62 (0.59–0.64) | 0.7 (0.68–0.72) | 0.71 (0.69–0.73) | 0.67 (0.65–0.69) | 0.65 (0.63–0.67) |
| -3 | | 0.77 (0.75–0.79) | 0.69 (0.67–0.71) | 0.73 (0.71–0.75) | 0.63 (0.61–0.65) | 0.7 (0.68–0.72) | 0.71 (0.69–0.73) | 0.68 (0.66–0.7) | 0.66 (0.64–0.68) |
| -2 | | 0.77 (0.75–0.79) | 0.7 (0.68–0.72) | 0.74 (0.72–0.76) | 0.65 (0.63–0.67) | 0.69 (0.67–0.71) | 0.71 (0.69–0.73) | 0.68 (0.66–0.7) | 0.65 (0.63–0.67) |
| -1 | | 0.77 (0.75–0.79) | 0.7 (0.68–0.72) | 0.74 (0.72–0.76) | 0.65 (0.63–0.67) | 0.69 (0.67–0.71) | 0.71 (0.69–0.73) | 0.68 (0.66–0.7) | 0.66 (0.64–0.68) |
| 0 | | 0.75 (0.73–0.77) | 0.68 (0.66–0.7) | 0.74 (0.72–0.76) | 0.65 (0.63–0.68) | 0.69 (0.67–0.71) | 0.7 (0.68–0.72) | 0.67 (0.65–0.69) | 0.66 (0.64–0.68) |
| 1 | | 0.75 (0.73–0.77) | 0.66 (0.64–0.69) | 0.77 (0.75–0.79) | 0.65 (0.63–0.67) | 0.69 (0.67–0.71) | 0.69 (0.67–0.71) | 0.68 (0.66–0.7) | 0.65 (0.63–0.67) |
| 2 | | 0.74 (0.72–0.77) | 0.66 (0.64–0.68) | 0.77 (0.75–0.79) | 0.67 (0.64–0.69) | 0.69 (0.67–0.71) | 0.69 (0.67–0.71) | 0.68 (0.66–0.7) | 0.64 (0.61–0.66) |
| 3 | | 0.74 (0.72–0.76) | 0.65 (0.63–0.67) | 0.77 (0.75–0.79) | 0.66 (0.64–0.69) | 0.69 (0.67–0.72) | 0.68 (0.66–0.71) | 0.68 (0.66–0.7) | 0.64 (0.62–0.66) |

**eTable 33. Estimates for marginal predictions of annual employment among parents exposed to child's psychotic disorder (F20–F29) and unexposed parents stratified by the child's age at diagnosis**

| | | Finland | | | | Denmark | | | |
|---|---|---|---|---|---|---|---|---|---|
| | | Men | | Women | | Men | | Women | |
| | | Unexposed | Exposed | Unexposed | Exposed | Unexposed | Exposed | Unexposed | Exposed |
| Years since diagnosis | Age group | Marg. pred. (95% CI) | Marg. pred. (95% CI) | Marg. pred. (95% CI) | Marg. pred. (95% CI) | Marg. pred. (95% CI) | Marg. pred. (95% CI) | Marg. pred. (95% CI) | Marg. pred. (95% CI) |
| | | Unexposed | Exposed | Unexposed | Exposed | Unexposed | Exposed | Unexposed | Exposed |
| 4 | | 0.74 (0.72–0.76) | 0.66 (0.63–0.68) | 0.77 (0.75–0.79) | 0.68 (0.65–0.7) | 0.68 (0.66–0.71) | 0.69 (0.67–0.71) | 0.69 (0.66–0.71) | 0.64 (0.62–0.67) |
| 5 | | 0.73 (0.71–0.76) | 0.65 (0.63–0.68) | 0.76 (0.74–0.78) | 0.67 (0.64–0.69) | 0.68 (0.66–0.7) | 0.67 (0.64–0.69) | 0.68 (0.66–0.71) | 0.63 (0.6–0.65) |
| -5 | 15–19 years | 0.81 (0.8–0.82) | 0.75 (0.74–0.76) | 0.78 (0.77–0.79) | 0.71 (0.7–0.72) | 0.71 (0.7–0.72) | 0.69 (0.68–0.7) | 0.68 (0.67–0.69) | 0.63 (0.62–0.64) |
| -4 | | 0.8 (0.79–0.81) | 0.74 (0.74–0.75) | 0.78 (0.77–0.79) | 0.72 (0.71–0.73) | 0.72 (0.71–0.73) | 0.7 (0.69–0.71) | 0.69 (0.68–0.7) | 0.64 (0.63–0.65) |
| -3 | | 0.79 (0.78–0.8) | 0.74 (0.73–0.75) | 0.78 (0.77–0.79) | 0.73 (0.72–0.74) | 0.73 (0.72–0.74) | 0.7 (0.69–0.71) | 0.7 (0.69–0.71) | 0.65 (0.64–0.66) |
| -2 | | 0.78 (0.78–0.79) | 0.73 (0.72–0.74) | 0.78 (0.78–0.79) | 0.73 (0.72–0.74) | 0.72 (0.71–0.73) | 0.7 (0.69–0.71) | 0.7 (0.69–0.71) | 0.65 (0.64–0.67) |
| -1 | | 0.78 (0.77–0.79) | 0.72 (0.71–0.73) | 0.79 (0.78–0.79) | 0.73 (0.72–0.74) | 0.72 (0.71–0.73) | 0.69 (0.68–0.7) | 0.7 (0.69–0.71) | 0.65 (0.64–0.66) |
| 0 | | 0.77 (0.76–0.78) | 0.71 (0.7–0.72) | 0.78 (0.77–0.79) | 0.72 (0.71–0.73) | 0.72 (0.71–0.73) | 0.68 (0.67–0.69) | 0.69 (0.68–0.7) | 0.65 (0.64–0.66) |
| 1 | | 0.76 (0.75–0.77) | 0.7 (0.69–0.71) | 0.78 (0.77–0.79) | 0.72 (0.71–0.73) | 0.71 (0.7–0.72) | 0.68 (0.67–0.69) | 0.7 (0.69–0.71) | 0.64 (0.63–0.65) |

**eTable 33. Estimates for marginal predictions of annual employment among parents exposed to child's psychotic disorder (F20–F29) and unexposed parents stratified by the child's age at diagnosis**

| | | Finland | | | | Denmark | | | |
|---|---|---|---|---|---|---|---|---|---|
| | | Men | | Women | | Men | | Women | |
| | | Unexposed | Exposed | Unexposed | Exposed | Unexposed | Exposed | Unexposed | Exposed |
| Years since diagnosis | Age group | Marg. pred. (95% CI) | Marg. pred. (95% CI) | Marg. pred. (95% CI) | Marg. pred. (95% CI) | Marg. pred. (95% CI) | Marg. pred. (95% CI) | Marg. pred. (95% CI) | Marg. pred. (95% CI) |
| | | Unexposed | Exposed | Unexposed | Exposed | Unexposed | Exposed | Unexposed | Exposed |
| 2 | | 0.75 (0.74–0.76) | 0.68 (0.67–0.69) | 0.78 (0.77–0.79) | 0.72 (0.71–0.73) | 0.7 (0.69–0.71) | 0.68 (0.66–0.69) | 0.7 (0.69–0.71) | 0.64 (0.63–0.65) |
| 3 | | 0.73 (0.72–0.74) | 0.67 (0.66–0.68) | 0.77 (0.77–0.78) | 0.71 (0.7–0.72) | 0.69 (0.68–0.7) | 0.67 (0.66–0.68) | 0.69 (0.68–0.7) | 0.63 (0.62–0.65) |
| 4 | | 0.72 (0.71–0.73) | 0.65 (0.64–0.66) | 0.77 (0.76–0.78) | 0.7 (0.69–0.71) | 0.68 (0.67–0.69) | 0.66 (0.64–0.67) | 0.68 (0.67–0.7) | 0.62 (0.61–0.64) |
| 5 | | 0.7 (0.69–0.71) | 0.62 (0.61–0.63) | 0.76 (0.75–0.77) | 0.69 (0.68–0.7) | 0.68 (0.67–0.69) | 0.64 (0.62–0.65) | 0.68 (0.67–0.69) | 0.61 (0.6–0.62) |
| -5 | 20–25 years | 0.78 (0.77–0.79) | 0.73 (0.72–0.74) | 0.79 (0.78–0.8) | 0.73 (0.72–0.74) | 0.71 (0.7–0.72) | 0.66 (0.65–0.66) | 0.68 (0.67–0.69) | 0.63 (0.62–0.63) |
| -4 | | 0.77 (0.76–0.78) | 0.72 (0.71–0.73) | 0.78 (0.78–0.79) | 0.72 (0.71–0.73) | 0.72 (0.71–0.73) | 0.67 (0.66–0.68) | 0.7 (0.69–0.71) | 0.63 (0.63–0.64) |
| -3 | | 0.75 (0.74–0.76) | 0.7 (0.69–0.71) | 0.77 (0.77–0.78) | 0.72 (0.71–0.72) | 0.72 (0.71–0.73) | 0.67 (0.66–0.68) | 0.71 (0.7–0.72) | 0.64 (0.63–0.65) |
| -2 | | 0.74 (0.73–0.75) | 0.68 (0.67–0.69) | 0.77 (0.76–0.78) | 0.71 (0.7–0.71) | 0.71 (0.7–0.72) | 0.66 (0.65–0.67) | 0.71 (0.7–0.71) | 0.63 (0.62–0.64) |
| -1 | | 0.72 (0.71–0.73) | 0.67 (0.66–0.68) | 0.76 (0.76–0.77) | 0.7 (0.69–0.71) | 0.7 (0.69–0.71) | 0.65 (0.64–0.66) | 0.7 (0.69–0.71) | 0.63 (0.62–0.64) |

**eTable 33. Estimates for marginal predictions of annual employment among parents exposed to child's psychotic disorder (F20–F29) and unexposed parents stratified by the child's age at diagnosis**

| | | Finland | | | | Denmark | | | |
|---|---|---|---|---|---|---|---|---|---|
| | | Men | | Women | | Men | | Women | |
| | | Unexposed | Exposed | Unexposed | Exposed | Unexposed | Exposed | Unexposed | Exposed |
| Years since diagnosis | Age group | Marg. pred. (95% CI) | Marg. pred. (95% CI) | Marg. pred. (95% CI) | Marg. pred. (95% CI) | Marg. pred. (95% CI) | Marg. pred. (95% CI) | Marg. pred. (95% CI) | Marg. pred. (95% CI) |
| | | Unexposed | Exposed | Unexposed | Exposed | Unexposed | Exposed | Unexposed | Exposed |
| 0 | | 0.7 (0.69–0.71) | 0.65 (0.65–0.66) | 0.75 (0.75–0.76) | 0.69 (0.68–0.7) | 0.69 (0.69–0.7) | 0.64 (0.63–0.65) | 0.68 (0.67–0.69) | 0.62 (0.61–0.63) |
| 1 | | 0.68 (0.67–0.69) | 0.63 (0.62–0.64) | 0.74 (0.73–0.75) | 0.67 (0.67–0.68) | 0.68 (0.67–0.69) | 0.63 (0.62–0.64) | 0.67 (0.66–0.68) | 0.61 (0.6–0.62) |
| 2 | | 0.65 (0.64–0.66) | 0.61 (0.6–0.62) | 0.73 (0.72–0.74) | 0.66 (0.65–0.67) | 0.66 (0.65–0.67) | 0.61 (0.6–0.62) | 0.66 (0.65–0.67) | 0.59 (0.58–0.6) |
| 3 | | 0.63 (0.62–0.63) | 0.58 (0.57–0.59) | 0.71 (0.7–0.72) | 0.65 (0.64–0.65) | 0.65 (0.64–0.66) | 0.59 (0.58–0.6) | 0.65 (0.64–0.66) | 0.58 (0.57–0.59) |
| 4 | | 0.6 (0.59–0.61) | 0.55 (0.54–0.56) | 0.69 (0.68–0.7) | 0.63 (0.62–0.64) | 0.63 (0.62–0.64) | 0.56 (0.55–0.57) | 0.63 (0.62–0.64) | 0.57 (0.56–0.58) |
| 5 | | 0.57 (0.56–0.58) | 0.52 (0.51–0.53) | 0.67 (0.66–0.67) | 0.6 (0.59–0.61) | 0.6 (0.59–0.62) | 0.54 (0.53–0.56) | 0.62 (0.61–0.63) | 0.55 (0.54–0.56) |

Marg. pred. = Marginal predictions in probabilities

**eTable 34. Estimates for marginal predictions of annual earnings among parents exposed to child's mood disorder (F30–F39) and unexposed parents stratified by the child's age at diagnosis**

| | | Finland | | | | Denmark | | | |
|---|---|---|---|---|---|---|---|---|---|
| | | Men | | Women | | Men | | Women | |
| | | Unexposed | Exposed | Unexposed | Exposed | Unexposed | Exposed | Unexposed | Exposed |
| Years since diagnosis | Age group | Marg. pred. (95% CI) | Marg. pred. (95% CI) | Marg. pred. (95% CI) | Marg. pred. (95% CI) | Marg. pred. (95% CI) | Marg. pred. (95% CI) | Marg. pred. (95% CI) | Marg. pred. (95% CI) |
| | | Unexposed | Exposed | Unexposed | Exposed | Unexposed | Exposed | Unexposed | Exposed |
| -5 | 5–9 years | 28.7 (27.8–29.6) | 26.6 (25.7–27.4) | 14 (13.4–14.6) | 12.5 (11.9–13.1) | | | | |
| -4 | | 29.7 (28.7–30.6) | 27.3 (26.5–28.2) | 15.6 (15–16.2) | 13.6 (13–14.2) | | | | |
| -3 | | 30.4 (29.5–31.4) | 27.8 (26.9–28.7) | 16.8 (16.2–17.5) | 14.7 (14.1–15.4) | | | | |
| -2 | | 31.1 (30.1–32.1) | 28.7 (27.8–29.7) | 17.7 (17.1–18.4) | 15.4 (14.8–16.1) | | | | |
| -1 | | 31.8 (30.8–32.8) | 29.3 (28.4–30.3) | 18.9 (18.2–19.6) | 16.3 (15.7–17) | | | | |
| 0 | | 33 (32–34.1) | 29.5 (28.6–30.5) | 19.8 (19–20.5) | 16.9 (16.2–17.6) | | | | |
| 1 | | 33.4 (32.3–34.5) | 29.9 (28.9–31) | 20.8 (20.1–21.6) | 17.7 (17–18.4) | | | | |
| 2 | | 33.8 (32.7–34.9) | 30.2 (29.1–31.3) | 22 (21.2–22.8) | 18.8 (18–19.5) | | | | |
| 3 | | 34.5 (33.3–35.7) | 31 (29.9–32.1) | 23.1 (22.3–23.9) | 19.7 (18.9–20.5) | | | | |
| 4 | | 34.8 (33.6–36) | 31.2 (30.1–32.4) | 23.9 (23.1–24.8) | 20.2 (19.4–21) | | | | |

**eTable 34. Estimates for marginal predictions of annual earnings among parents exposed to child's mood disorder (F30–F39) and unexposed parents stratified by the child’s age at diagnosis**

| | | Finland | | | | Denmark | | | |
|---|---|---|---|---|---|---|---|---|---|
| | | Men | | Women | | Men | | Women | |
| | | Unexposed | Exposed | Unexposed | Exposed | Unexposed | Exposed | Unexposed | Exposed |
| Years since diagnosis | Age group | Marg. pred. (95% CI) | Marg. pred. (95% CI) | Marg. pred. (95% CI) | Marg. pred. (95% CI) | Marg. pred. (95% CI) | Marg. pred. (95% CI) | Marg. pred. (95% CI) | Marg. pred. (95% CI) |
| | | Unexposed | Exposed | Unexposed | Exposed | Unexposed | Exposed | Unexposed | Exposed |
| 5 | | 35.3 (34.1–36.6) | 31.3 (30.1–32.5) | 24.8 (23.9–25.6) | 21.1 (20.3–21.9) | | | | |
| -5 | 10–14 years | 33.7 (33.3–34.1) | 31.5 (31.2–31.9) | 19.8 (19.6–20.1) | 18.9 (18.7–19.1) | | | | |
| -4 | | 34.3 (33.9–34.6) | 32 (31.7–32.4) | 20.8 (20.5–21) | 19.7 (19.5–19.9) | | | | |
| -3 | | 34.7 (34.3–35.1) | 32.4 (32–32.7) | 21.8 (21.6–22) | 20.5 (20.3–20.8) | | | | |
| -2 | | 35.3 (34.9–35.7) | 32.8 (32.4–33.2) | 22.8 (22.5–23) | 21.3 (21.1–21.6) | | | | |
| -1 | | 35.7 (35.3–36.1) | 33 (32.7–33.4) | 23.6 (23.3–23.9) | 22.2 (22–22.5) | | | | |
| 0 | | 36.2 (35.8–36.6) | 33.3 (32.9–33.7) | 24.5 (24.2–24.7) | 22.8 (22.5–23) | | | | |
| 1 | | 36.7 (36.2–37.1) | 33.5 (33.1–33.9) | 25.2 (25–25.5) | 23.2 (23–23.5) | | | | |
| 2 | | 36.9 (36.5–37.4) | 33.5 (33.1–34) | 26 (25.7–26.2) | 23.9 (23.6–24.2) | | | | |
| 3 | | 37.1 (36.6–37.5) | 33.5 (33.1–34) | 26.5 (26.2–26.8) | 24.4 (24.1–24.7) | | | | |

**eTable 34. Estimates for marginal predictions of annual earnings among parents exposed to child's mood disorder (F30–F39) and unexposed parents stratified by the child’s age at diagnosis**

| | | Finland | | | | Denmark | | | |
|---|---|---|---|---|---|---|---|---|---|
| | | Men | | Women | | Men | | Women | |
| | | Unexposed | Exposed | Unexposed | Exposed | Unexposed | Exposed | Unexposed | Exposed |
| Years since diagnosis | Age group | Marg. pred. (95% CI) | Marg. pred. (95% CI) | Marg. pred. (95% CI) | Marg. pred. (95% CI) | Marg. pred. (95% CI) | Marg. pred. (95% CI) | Marg. pred. (95% CI) | Marg. pred. (95% CI) |
| | | Unexposed | Exposed | Unexposed | Exposed | Unexposed | Exposed | Unexposed | Exposed |
| 4 | | 37.1 (36.6–37.6) | 33.5 (33–34) | 27.1 (26.8–27.4) | 24.9 (24.6–25.2) | | | | |
| 5 | | 36.9 (36.4–37.4) | 33.4 (32.9–33.9) | 27.6 (27.3–27.9) | 25.2 (24.9–25.6) | | | | |
| -5 | 5–14 years | | | | | 58.6 (57.6–59.7) | 57.3 (56.4–58.2) | 44.3 (43.8–44.8) | 44.2 (43.7–44.7) |
| -4 | | | | | | 59.4 (58.3–60.4) | 58.2 (57.3–59.1) | 45.2 (44.7–45.8) | 45 (44.5–45.5) |
| -3 | | | | | | 60.4 (59.3–61.5) | 58.7 (57.8–59.7) | 46 (45.4–46.5) | 46 (45.5–46.4) |
| -2 | | | | | | 60.9 (59.8–62) | 59.2 (58.3–60.2) | 46.6 (46–47.1) | 46.8 (46.3–47.3) |
| -1 | | | | | | 61.9 (60.8–63) | 60 (59–61) | 47.4 (46.8–47.9) | 47.2 (46.6–47.7) |
| 0 | | | | | | 62.6 (61.5–63.8) | 60.6 (59.6–61.6) | 48 (47.4–48.5) | 47.7 (47.2–48.2) |
| 1 | | | | | | 63.3 (62.1–64.5) | 61.7 (60.6–62.7) | 48.7 (48.1–49.2) | 48.2 (47.7–48.7) |

**eTable 34. Estimates for marginal predictions of annual earnings among parents exposed to child's mood disorder (F30–F39) and unexposed parents stratified by the child's age at diagnosis**

| | | Finland | | | | Denmark | | | |
|---|---|---|---|---|---|---|---|---|---|
| | | Men | | Women | | Men | | Women | |
| | | Unexposed | Exposed | Unexposed | Exposed | Unexposed | Exposed | Unexposed | Exposed |
| Years since diagnosis | Age group | Marg. pred. (95% CI) | Marg. pred. (95% CI) | Marg. pred. (95% CI) | Marg. pred. (95% CI) | Marg. pred. (95% CI) | Marg. pred. (95% CI) | Marg. pred. (95% CI) | Marg. pred. (95% CI) |
| | | Unexposed | Exposed | Unexposed | Exposed | Unexposed | Exposed | Unexposed | Exposed |
| 2 | | | | | | 64 (62.8–65.2) | 61.7 (60.6–62.8) | 49.2 (48.6–49.8) | 48.7 (48.2–49.3) |
| 3 | | | | | | 64.2 (63–65.5) | 62.5 (61.4–63.6) | 49.4 (48.8–50) | 48.4 (47.8–48.9) |
| 4 | | | | | | 64.8 (63.5–66.1) | 63 (61.9–64.1) | 49.5 (48.8–50.1) | 48.3 (47.7–48.9) |
| 5 | | | | | | 65.6 (64.2–67) | 63.3 (62.1–64.5) | 49.7 (49–50.3) | 47.9 (47.3–48.6) |
| -5 | 15–19 years | 34.8 (34.6–35.1) | 32.9 (32.7–33.2) | 23 (22.8–23.1) | 21.7 (21.6–21.9) | 58.1 (57.6–58.6) | 56.6 (56.1–57) | 44.3 (44.1–44.6) | 44.3 (44.1–44.6) |
| -4 | | 35.3 (35–35.5) | 33.3 (33–33.5) | 23.8 (23.6–23.9) | 22.5 (22.4–22.7) | 58.8 (58.2–59.3) | 57.2 (56.7–57.7) | 45.1 (44.8–45.3) | 45 (44.7–45.2) |
| -3 | | 35.6 (35.4–35.9) | 33.6 (33.3–33.8) | 24.6 (24.4–24.7) | 23.3 (23.1–23.4) | 59.4 (58.8–59.9) | 57.6 (57.1–58.1) | 45.5 (45.3–45.8) | 45.5 (45.2–45.7) |
| -2 | | 35.8 (35.6–36.1) | 33.7 (33.4–33.9) | 25.2 (25–25.4) | 23.9 (23.7–24) | 60.2 (59.6–60.8) | 58.1 (57.6–58.6) | 46 (45.7–46.3) | 45.9 (45.6–46.1) |
| -1 | | 36.1 (35.9–36.4) | 33.8 (33.5–34) | 25.8 (25.6–26) | 24.5 (24.3–24.6) | 60.5 (59.9–61.1) | 58.7 (58.2–59.2) | 46.2 (45.9–46.5) | 46.1 (45.8–46.4) |

**eTable 34. Estimates for marginal predictions of annual earnings among parents exposed to child's mood disorder (F30–F39) and unexposed parents stratified by the child's age at diagnosis**

| | | Finland | | | | Denmark | | | |
|---|---|---|---|---|---|---|---|---|---|
| | | Men | | Women | | Men | | Women | |
| | | Unexposed | Exposed | Unexposed | Exposed | Unexposed | Exposed | Unexposed | Exposed |
| Years since diagnosis | Age group | Marg. pred. (95% CI) | Marg. pred. (95% CI) | Marg. pred. (95% CI) | Marg. pred. (95% CI) | Marg. pred. (95% CI) | Marg. pred. (95% CI) | Marg. pred. (95% CI) | Marg. pred. (95% CI) |
| | | Unexposed | Exposed | Unexposed | Exposed | Unexposed | Exposed | Unexposed | Exposed |
| 0 | | 36.2 (36–36.5) | 33.8 (33.5–34.1) | 26.4 (26.3–26.6) | 24.9 (24.7–25.1) | 60.9 (60.3–61.5) | 58.9 (58.4–59.4) | 46.3 (46–46.6) | 46.1 (45.8–46.4) |
| 1 | | 36.2 (35.9–36.4) | 33.7 (33.4–33.9) | 26.8 (26.7–27) | 25.3 (25.1–25.4) | 61.3 (60.7–61.9) | 58.9 (58.4–59.5) | 46.4 (46.1–46.7) | 46 (45.7–46.3) |
| 2 | | 36 (35.7–36.3) | 33.4 (33.1–33.7) | 27.3 (27.1–27.4) | 25.6 (25.5–25.8) | 61.6 (61–62.3) | 59.2 (58.6–59.7) | 46.4 (46.1–46.7) | 45.8 (45.5–46.1) |
| 3 | | 35.8 (35.5–36.1) | 33 (32.8–33.3) | 27.5 (27.4–27.7) | 25.9 (25.7–26.1) | 61.8 (61.2–62.5) | 59.2 (58.6–59.7) | 46.2 (45.9–46.6) | 45.8 (45.5–46.1) |
| 4 | | 35.4 (35.1–35.7) | 32.6 (32.3–32.9) | 27.7 (27.5–27.8) | 26 (25.9–26.2) | 62 (61.4–62.7) | 59.1 (58.5–59.6) | 46.4 (46.1–46.7) | 45.8 (45.5–46.1) |
| 5 | | 34.9 (34.6–35.2) | 32.1 (31.8–32.4) | 27.7 (27.5–27.9) | 26.1 (25.9–26.3) | 62.2 (61.5–62.9) | 59 (58.4–59.6) | 46.6 (46.2–46.9) | 45.8 (45.5–46.1) |
| -5 | 20–25 years | 34.2 (33.9–34.4) | 32.1 (31.9–32.4) | 24.2 (24.1–24.4) | 22.9 (22.7–23) | 56.6 (56.1–57) | 55.8 (55.4–56.3) | 42.9 (42.7–43.1) | 42.2 (42–42.4) |
| -4 | | 34.1 (33.9–34.4) | 32 (31.7–32.2) | 24.6 (24.4–24.7) | 23.1 (23–23.3) | 56.8 (56.4–57.3) | 56 (55.6–56.5) | 42.9 (42.7–43.1) | 42.2 (42–42.5) |
| -3 | | 33.9 (33.6–34.2) | 31.7 (31.4–32) | 24.9 (24.7–25) | 23.4 (23.3–23.6) | 57.1 (56.6–57.6) | 56.2 (55.8–56.7) | 42.9 (42.7–43.2) | 42.2 (42–42.4) |
| -2 | | 33.6 (33.3–33.9) | 31.5 (31.2–31.7) | 25 (24.9–25.2) | 23.6 (23.4–23.7) | 57.4 (56.9–57.9) | 56.2 (55.8–56.7) | 42.9 (42.7–43.2) | 42.1 (41.9–42.4) |

**eTable 34. Estimates for marginal predictions of annual earnings among parents exposed to child's mood disorder (F30–F39) and unexposed parents stratified by the child's age at diagnosis**

| | | Finland | | | | Denmark | | | |
|---|---|---|---|---|---|---|---|---|---|
| | | Men | | Women | | Men | | Women | |
| | | Unexposed | Exposed | Unexposed | Exposed | Unexposed | Exposed | Unexposed | Exposed |
| Years since diagnosis | Age group | Marg. pred. (95% CI) | Marg. pred. (95% CI) | Marg. pred. (95% CI) | Marg. pred. (95% CI) | Marg. pred. (95% CI) | Marg. pred. (95% CI) | Marg. pred. (95% CI) | Marg. pred. (95% CI) |
| | | Unexposed | Exposed | Unexposed | Exposed | Unexposed | Exposed | Unexposed | Exposed |
| -1 | | 33.2 (32.9–33.5) | 31 (30.7–31.3) | 25.1 (25–25.3) | 23.6 (23.4–23.8) | 57.5 (57–57.9) | 56.2 (55.8–56.7) | 43 (42.8–43.3) | 42.2 (42–42.5) |
| 0 | | 32.8 (32.5–33) | 30.5 (30.2–30.8) | 25.2 (25–25.4) | 23.5 (23.4–23.7) | 57.5 (57–58) | 56.3 (55.8–56.8) | 43.1 (42.8–43.3) | 42.3 (42.1–42.5) |
| 1 | | 32.2 (31.9–32.5) | 29.9 (29.7–30.2) | 25.1 (24.9–25.2) | 23.5 (23.3–23.6) | 57.4 (56.9–57.9) | 56.2 (55.7–56.7) | 43.2 (42.9–43.4) | 42.3 (42–42.5) |
| 2 | | 31.3 (31–31.6) | 29.1 (28.8–29.4) | 24.9 (24.8–25.1) | 23.3 (23.1–23.4) | 57.2 (56.7–57.7) | 55.9 (55.4–56.4) | 43.3 (43–43.5) | 42.2 (42–42.5) |
| 3 | | 30.3 (30–30.7) | 28.1 (27.8–28.4) | 24.7 (24.5–24.9) | 23 (22.8–23.2) | 56.7 (56.2–57.2) | 55.5 (55–56.1) | 43.3 (43.1–43.6) | 42.3 (42–42.5) |
| 4 | | 29.2 (28.9–29.5) | 27.1 (26.8–27.4) | 24.2 (24–24.4) | 22.6 (22.4–22.8) | 56.8 (56.3–57.4) | 55.1 (54.6–55.7) | 43.3 (43–43.6) | 42.3 (42.1–42.6) |
| 5 | | 28 (27.7–28.4) | 26 (25.7–26.3) | 23.6 (23.4–23.8) | 22.1 (21.9–22.3) | 56.3 (55.7–56.9) | 54.9 (54.4–55.4) | 43 (42.7–43.2) | 42.1 (41.9–42.4) |

Marg. pred. = Marginal predictions in thousand euros (€)

**eTable 35. Estimates for marginal predictions of annual disposable income among parents exposed to child's mood disorder (F30–F39) and unexposed parents stratified by the child's age at diagnosis**

| | | Finland | | | | Denmark | | | |
|---|---|---|---|---|---|---|---|---|---|
| | | Men | | Women | | Men | | Women | |
| | | Unexposed | Exposed | Unexposed | Exposed | Unexposed | Exposed | Unexposed | Exposed |
| Years since diagnosis | Age group | Marg. pred. (95% CI) | Marg. pred. (95% CI) | Marg. pred. (95% CI) | Marg. pred. (95% CI) | Marg. pred. (95% CI) | Marg. pred. (95% CI) | Marg. pred. (95% CI) | Marg. pred. (95% CI) |
| | | Unexposed | Exposed | Unexposed | Exposed | Unexposed | Exposed | Unexposed | Exposed |
| -5 | 5–9 years | 24.5 (23.9–25.1) | 23 (22.5–23.5) | 19.7 (19.3–20) | 19.4 (19–19.7) | | | | |
| -4 | | 25.3 (24.7–25.9) | 23.6 (23.1–24.2) | 20.7 (20.3–21) | 20.3 (19.9–20.6) | | | | |
| -3 | | 25.8 (25.2–26.5) | 24 (23.5–24.5) | 21.5 (21.1–21.8) | 21.4 (21–21.7) | | | | |
| -2 | | 26.4 (25.8–27.1) | 24.7 (24.1–25.3) | 22.2 (21.8–22.5) | 22.1 (21.7–22.5) | | | | |
| -1 | | 27.1 (26.4–27.7) | 25.3 (24.7–25.9) | 22.9 (22.5–23.3) | 23 (22.6–23.4) | | | | |
| 0 | | 27.9 (27.2–28.6) | 25.7 (25.1–26.2) | 23.6 (23.2–24) | 23.6 (23.2–24) | | | | |
| 1 | | 28.3 (27.6–29) | 26.4 (25.8–27.1) | 24.1 (23.7–24.5) | 24.2 (23.8–24.6) | | | | |
| 2 | | 28.8 (28.1–29.5) | 26.7 (26–27.3) | 25 (24.5–25.4) | 24.7 (24.3–25.2) | | | | |
| 3 | | 29.4 (28.6–30.2) | 27.3 (26.6–28) | 25.6 (25.1–26) | 25.4 (24.9–25.8) | | | | |
| 4 | | 29.9 (29.1–30.7) | 27.6 (26.9–28.3) | 26.2 (25.8–26.7) | 25.8 (25.4–26.3) | | | | |

**eTable 35. Estimates for marginal predictions of annual disposable income among parents exposed to child's mood disorder (F30–F39) and unexposed parents stratified by the child's age at diagnosis**

| | | Finland | | | | Denmark | | | |
|---|---|---|---|---|---|---|---|---|---|
| | | Men | | Women | | Men | | Women | |
| | | Unexposed | Exposed | Unexposed | Exposed | Unexposed | Exposed | Unexposed | Exposed |
| Years since diagnosis | Age group | Marg. pred. (95% CI) | Marg. pred. (95% CI) | Marg. pred. (95% CI) | Marg. pred. (95% CI) | Marg. pred. (95% CI) | Marg. pred. (95% CI) | Marg. pred. (95% CI) | Marg. pred. (95% CI) |
| | | Unexposed | Exposed | Unexposed | Exposed | Unexposed | Exposed | Unexposed | Exposed |
| 5 | | 30.5 (29.6–31.3) | 27.8 (27.1–28.6) | 26.9 (26.4–27.4) | 26.3 (25.8–26.7) | | | | |
| -5 | 10–14 years | 27.9 (27.6–28.1) | 26.4 (26.2–26.7) | 23.1 (22.9–23.2) | 22.8 (22.7–23) | | | | |
| -4 | | 28.5 (28.2–28.7) | 27.1 (26.8–27.3) | 23.7 (23.6–23.9) | 23.5 (23.4–23.6) | | | | |
| -3 | | 28.9 (28.7–29.2) | 27.5 (27.2–27.7) | 24.3 (24.2–24.5) | 24.2 (24–24.3) | | | | |
| -2 | | 29.5 (29.2–29.7) | 27.9 (27.7–28.2) | 25 (24.8–25.1) | 24.8 (24.7–24.9) | | | | |
| -1 | | 30 (29.7–30.2) | 28.3 (28.1–28.6) | 25.5 (25.4–25.6) | 25.4 (25.3–25.5) | | | | |
| 0 | | 30.4 (30.2–30.7) | 28.7 (28.5–29) | 26 (25.8–26.1) | 25.8 (25.6–25.9) | | | | |
| 1 | | 31 (30.7–31.2) | 29 (28.8–29.3) | 26.4 (26.2–26.5) | 26.1 (25.9–26.2) | | | | |
| 2 | | 31.3 (31–31.6) | 29.3 (29–29.5) | 26.6 (26.4–26.8) | 26.3 (26.2–26.5) | | | | |

**eTable 35. Estimates for marginal predictions of annual disposable income among parents exposed to child's mood disorder (F30–F39) and unexposed parents stratified by the child's age at diagnosis**

| | | Finland | | | | Denmark | | | |
|---|---|---|---|---|---|---|---|---|---|
| | | Men | | Women | | Men | | Women | |
| | | Unexposed | Exposed | Unexposed | Exposed | Unexposed | Exposed | Unexposed | Exposed |
| Years since diagnosis | Age group | Marg. pred. (95% CI) | Marg. pred. (95% CI) | Marg. pred. (95% CI) | Marg. pred. (95% CI) | Marg. pred. (95% CI) | Marg. pred. (95% CI) | Marg. pred. (95% CI) | Marg. pred. (95% CI) |
| | | Unexposed | Exposed | Unexposed | Exposed | Unexposed | Exposed | Unexposed | Exposed |
| 3 | | 31.7 (31.4–32.1) | 29.5 (29.2–29.8) | 26.6 (26.4–26.7) | 26.2 (26–26.4) | | | | |
| 4 | | 32.1 (31.8–32.4) | 29.7 (29.4–30) | 26.4 (26.3–26.6) | 26.1 (25.9–26.2) | | | | |
| 5 | | 32.3 (31.9–32.6) | 29.9 (29.6–30.2) | 26.5 (26.3–26.7) | 25.9 (25.8–26.1) | | | | |
| -5 | 5–14 years | | | | | 35.4 (34.8–35.9) | 34.4 (33.9–34.9) | 30.7 (30.4–31) | 30.5 (30.2–30.7) |
| -4 | | | | | | 36.2 (35.6–36.8) | 35.2 (34.6–35.7) | 31.5 (31.2–31.8) | 31.2 (30.9–31.4) |
| -3 | | | | | | 37.3 (36.7–37.9) | 35.9 (35.4–36.5) | 32.1 (31.8–32.4) | 31.9 (31.6–32.2) |
| -2 | | | | | | 37.8 (37.2–38.4) | 36.6 (36–37.1) | 32.7 (32.4–33) | 32.6 (32.3–32.9) |
| -1 | | | | | | 38.9 (38.2–39.5) | 37.5 (36.9–38.1) | 33.4 (33.1–33.8) | 33 (32.7–33.3) |
| 0 | | | | | | 39.6 (38.9–40.2) | 38.2 (37.6–38.8) | 34 (33.6–34.3) | 33.6 (33.2–33.9) |

**eTable 35. Estimates for marginal predictions of annual disposable income among parents exposed to child's mood disorder (F30–F39) and unexposed parents stratified by the child's age at diagnosis**

| | | Finland | | | | Denmark | | | |
|---|---|---|---|---|---|---|---|---|---|
| | | Men | | Women | | Men | | Women | |
| | | Unexposed | Exposed | Unexposed | Exposed | Unexposed | Exposed | Unexposed | Exposed |
| Years since diagnosis | Age group | Marg. pred. (95% CI) | Marg. pred. (95% CI) | Marg. pred. (95% CI) | Marg. pred. (95% CI) | Marg. pred. (95% CI) | Marg. pred. (95% CI) | Marg. pred. (95% CI) | Marg. pred. (95% CI) |
| | | Unexposed | Exposed | Unexposed | Exposed | Unexposed | Exposed | Unexposed | Exposed |
| 1 | | | | | | 40.5 (39.8–41.2) | 39.3 (38.7–39.9) | 34.5 (34.2–34.9) | 34.1 (33.8–34.4) |
| 2 | | | | | | 41.4 (40.7–42.1) | 39.7 (39.1–40.4) | 35.1 (34.7–35.4) | 34.6 (34.2–34.9) |
| 3 | | | | | | 42 (41.3–42.7) | 40.6 (39.9–41.2) | 35.3 (35–35.7) | 34.4 (34.1–34.8) |
| 4 | | | | | | 42.8 (42.1–43.6) | 41.4 (40.7–42.1) | 35.5 (35.1–35.9) | 34.4 (34.1–34.8) |
| 5 | | | | | | 43.9 (43–44.7) | 42.2 (41.5–42.9) | 35.7 (35.2–36.1) | 34.3 (33.9–34.7) |
| -5 | 15–19 years | 28.7 (28.5–28.9) | 27.5 (27.3–27.6) | 23.9 (23.8–24) | 23.6 (23.5–23.7) | 35.3 (35–35.6) | 34.2 (33.9–34.4) | 30.7 (30.5–30.8) | 30.4 (30.3–30.6) |
| -4 | | 29.2 (29–29.3) | 27.9 (27.8–28.1) | 24.4 (24.3–24.5) | 24.1 (24–24.2) | 36.1 (35.8–36.4) | 35 (34.7–35.2) | 31.3 (31.1–31.4) | 31 (30.9–31.2) |
| -3 | | 29.6 (29.4–29.8) | 28.3 (28.2–28.5) | 24.8 (24.7–24.9) | 24.5 (24.4–24.6) | 36.8 (36.5–37.2) | 35.6 (35.3–35.8) | 31.7 (31.5–31.9) | 31.5 (31.3–31.6) |
| -2 | | 30 (29.8–30.2) | 28.7 (28.5–28.8) | 24.9 (24.8–25) | 24.7 (24.6–24.8) | 37.7 (37.4–38) | 36.2 (35.9–36.5) | 32.1 (31.9–32.3) | 31.9 (31.7–32) |

**eTable 35. Estimates for marginal predictions of annual disposable income among parents exposed to child's mood disorder (F30–F39) and unexposed parents stratified by the child's age at diagnosis**

| | | Finland | | | | Denmark | | | |
|---|---|---|---|---|---|---|---|---|---|
| | | Men | | Women | | Men | | Women | |
| | | Unexposed | Exposed | Unexposed | Exposed | Unexposed | Exposed | Unexposed | Exposed |
| Years since diagnosis | Age group | Marg. pred. (95% CI) | Marg. pred. (95% CI) | Marg. pred. (95% CI) | Marg. pred. (95% CI) | Marg. pred. (95% CI) | Marg. pred. (95% CI) | Marg. pred. (95% CI) | Marg. pred. (95% CI) |
| | | Unexposed | Exposed | Unexposed | Exposed | Unexposed | Exposed | Unexposed | Exposed |
| -1 | | 30.4 (30.2–30.6) | 28.9 (28.8–29.1) | 25 (24.9–25) | 24.8 (24.7–24.9) | 38.4 (38–38.7) | 37 (36.7–37.3) | 32.3 (32.2–32.5) | 32 (31.9–32.2) |
| 0 | | 30.7 (30.6–30.9) | 29.2 (29–29.3) | 25 (24.9–25.1) | 24.7 (24.6–24.8) | 39 (38.6–39.3) | 37.6 (37.3–37.9) | 32.5 (32.3–32.6) | 32.1 (31.9–32.2) |
| 1 | | 31 (30.8–31.2) | 29.4 (29.2–29.6) | 25 (24.9–25.1) | 24.6 (24.5–24.7) | 39.7 (39.3–40) | 38.1 (37.8–38.4) | 32.6 (32.4–32.8) | 32.1 (31.9–32.3) |
| 2 | | 31.2 (31–31.3) | 29.6 (29.4–29.7) | 25 (24.9–25.1) | 24.6 (24.5–24.7) | 40.4 (40–40.7) | 38.8 (38.4–39.1) | 32.7 (32.5–32.9) | 32 (31.8–32.2) |
| 3 | | 31.5 (31.3–31.7) | 29.7 (29.6–29.9) | 25.1 (24.9–25.2) | 24.6 (24.5–24.7) | 40.9 (40.6–41.3) | 39.3 (38.9–39.6) | 32.7 (32.5–32.9) | 32.1 (31.9–32.3) |
| 4 | | 31.6 (31.4–31.8) | 29.9 (29.7–30.1) | 25.2 (25.1–25.3) | 24.7 (24.6–24.9) | 41.6 (41.2–41.9) | 39.7 (39.4–40) | 33 (32.8–33.2) | 32.3 (32.1–32.5) |
| 5 | | 31.8 (31.5–32) | 30 (29.8–30.2) | 25.3 (25.2–25.4) | 24.9 (24.8–25) | 42.1 (41.7–42.6) | 40.1 (39.8–40.5) | 33.2 (33–33.5) | 32.5 (32.3–32.8) |
| -5 | 20–25 years | 28.8 (28.6–29) | 27.5 (27.3–27.7) | 22.8 (22.7–22.9) | 22.5 (22.4–22.5) | 34.9 (34.7–35.2) | 34.2 (34–34.5) | 29.1 (29–29.3) | 28.6 (28.4–28.7) |
| -4 | | 29 (28.8–29.2) | 27.8 (27.6–27.9) | 22.8 (22.7–22.9) | 22.4 (22.3–22.5) | 35.6 (35.3–35.8) | 34.8 (34.6–35.1) | 29.2 (29.1–29.4) | 28.7 (28.5–28.8) |

**eTable 35. Estimates for marginal predictions of annual disposable income among parents exposed to child's mood disorder (F30–F39) and unexposed parents stratified by the child's age at diagnosis**

| | | Finland | | | | Denmark | | | |
|---|---|---|---|---|---|---|---|---|---|
| | | Men | | Women | | Men | | Women | |
| | | Unexposed | Exposed | Unexposed | Exposed | Unexposed | Exposed | Unexposed | Exposed |
| Years since diagnosis | Age group | Marg. pred. (95% CI) | Marg. pred. (95% CI) | Marg. pred. (95% CI) | Marg. pred. (95% CI) | Marg. pred. (95% CI) | Marg. pred. (95% CI) | Marg. pred. (95% CI) | Marg. pred. (95% CI) |
| | | Unexposed | Exposed | Unexposed | Exposed | Unexposed | Exposed | Unexposed | Exposed |
| -3 | | 29.2 (29–29.4) | 27.8 (27.7–28) | 22.8 (22.7–22.9) | 22.3 (22.2–22.4) | 36.2 (35.9–36.4) | 35.4 (35.1–35.7) | 29.3 (29.2–29.5) | 28.7 (28.6–28.9) |
| -2 | | 29.4 (29.3–29.6) | 28.1 (27.9–28.2) | 22.9 (22.8–23) | 22.4 (22.3–22.5) | 36.8 (36.5–37) | 35.8 (35.5–36.1) | 29.4 (29.3–29.6) | 28.7 (28.6–28.9) |
| -1 | | 29.6 (29.4–29.8) | 28.2 (28–28.3) | 23 (22.9–23.1) | 22.5 (22.4–22.6) | 37.2 (36.9–37.4) | 36.3 (36–36.5) | 29.6 (29.5–29.8) | 29 (28.8–29.1) |
| 0 | | 29.7 (29.5–29.9) | 28.3 (28.1–28.4) | 23.1 (23–23.2) | 22.6 (22.5–22.7) | 37.6 (37.3–37.9) | 36.7 (36.4–36.9) | 29.9 (29.7–30) | 29.1 (29–29.3) |
| 1 | | 29.9 (29.7–30.1) | 28.4 (28.2–28.6) | 23.2 (23.1–23.3) | 22.7 (22.6–22.8) | 38 (37.7–38.3) | 37 (36.7–37.3) | 30.1 (29.9–30.2) | 29.3 (29.2–29.5) |
| 2 | | 29.9 (29.7–30.1) | 28.5 (28.3–28.7) | 23.4 (23.3–23.5) | 22.8 (22.7–22.9) | 38.3 (38–38.6) | 37.3 (37–37.6) | 30.4 (30.2–30.5) | 29.5 (29.4–29.7) |
| 3 | | 30 (29.8–30.2) | 28.4 (28.2–28.6) | 23.6 (23.5–23.7) | 22.9 (22.8–23) | 38.4 (38.1–38.7) | 37.5 (37.2–37.8) | 30.6 (30.5–30.8) | 29.8 (29.6–29.9) |
| 4 | | 30.1 (29.9–30.3) | 28.4 (28.3–28.6) | 23.6 (23.5–23.7) | 23 (22.9–23.1) | 39 (38.6–39.3) | 37.6 (37.3–38) | 30.9 (30.7–31.1) | 30 (29.8–30.1) |
| 5 | | 30.1 (29.9–30.4) | 28.5 (28.3–28.7) | 23.7 (23.6–23.9) | 23.1 (23–23.2) | 39.1 (38.8–39.5) | 38 (37.6–38.3) | 31 (30.8–31.1) | 30.1 (29.9–30.3) |

Marg. pred. = Marginal predictions in thousand euros (€)

**eTable 36. Estimates for marginal predictions of annual social welfare benefits among parents exposed to child's mood disorder (F30–F39) and unexposed parents stratified by the child's age at diagnosis**

| | | Finland | | | | Denmark | | | |
|---|---|---|---|---|---|---|---|---|---|
| | | Men | | Women | | Men | | Women | |
| | | Unexposed | Exposed | Unexposed | Exposed | Unexposed | Exposed | Unexposed | Exposed |
| Years since diagnosis | Age group | Marg. pred. (95% CI) | Marg. pred. (95% CI) | Marg. pred. (95% CI) | Marg. pred. (95% CI) | Marg. pred. (95% CI) | Marg. pred. (95% CI) | Marg. pred. (95% CI) | Marg. pred. (95% CI) |
| | | Unexposed | Exposed | Unexposed | Exposed | Unexposed | Exposed | Unexposed | Exposed |
| -5 | 5–9 years | 3.3 (3.1–3.6) | 3.7 (3.5–4) | 9.8 (9.5–10.1) | 10.7 (10.4–10.9) | | | | |
| -4 | | 3.4 (3.2–3.7) | 3.9 (3.7–4.2) | 9.4 (9.1–9.7) | 10.6 (10.3–10.9) | | | | |
| -3 | | 3.5 (3.3–3.8) | 4.1 (3.9–4.4) | 9.4 (9.1–9.6) | 10.8 (10.5–11.2) | | | | |
| -2 | | 3.6 (3.3–3.8) | 4.2 (3.9–4.5) | 9.3 (9–9.6) | 11 (10.7–11.3) | | | | |
| -1 | | 3.7 (3.4–3.9) | 4.3 (4–4.6) | 9.1 (8.8–9.4) | 11.2 (10.9–11.6) | | | | |
| 0 | | 3.6 (3.4–3.9) | 4.6 (4.3–4.9) | 9.2 (8.8–9.5) | 11.3 (11–11.7) | | | | |
| 1 | | 3.9 (3.6–4.2) | 5 (4.7–5.4) | 8.8 (8.5–9.1) | 11.2 (10.9–11.6) | | | | |
| 2 | | 3.9 (3.6–4.2) | 5 (4.7–5.4) | 8.8 (8.4–9.1) | 11 (10.7–11.4) | | | | |
| 3 | | 4 (3.7–4.3) | 5.2 (4.8–5.6) | 8.5 (8.2–8.9) | 11 (10.7–11.4) | | | | |
| 4 | | 4.2 (3.9–4.6) | 5.3 (5–5.7) | 8.5 (8.2–8.8) | 11 (10.6–11.4) | | | | |
| 5 | | 4.3 (3.9–4.6) | 5.6 (5.2–6) | 8.5 (8.1–8.8) | 10.8 (10.4–11.2) | | | | |

**eTable 36. Estimates for marginal predictions of annual social welfare benefits among parents exposed to child's mood disorder (F30–F39) and unexposed parents stratified by the child's age at diagnosis**

| | | Finland | | | | Denmark | | | |
|---|---|---|---|---|---|---|---|---|---|
| | | Men | | Women | | Men | | Women | |
| | | Unexposed | Exposed | Unexposed | Exposed | Unexposed | Exposed | Unexposed | Exposed |
| Years since diagnosis | Age group | Marg. pred. (95% CI) | Marg. pred. (95% CI) | Marg. pred. (95% CI) | Marg. pred. (95% CI) | Marg. pred. (95% CI) | Marg. pred. (95% CI) | Marg. pred. (95% CI) | Marg. pred. (95% CI) |
| | | Unexposed | Exposed | Unexposed | Exposed | Unexposed | Exposed | Unexposed | Exposed |
| -5 | 10–14 years | 3.1 (3.1–3.2) | 3.7 (3.6–3.8) | 8.5 (8.4–8.6) | 9.1 (9–9.2) | | | | |
| -4 | | 3.3 (3.2–3.4) | 3.9 (3.8–4) | 8.4 (8.3–8.5) | 9.1 (9–9.2) | | | | |
| -3 | | 3.6 (3.5–3.7) | 4.1 (4–4.2) | 8.3 (8.2–8.4) | 9.2 (9.1–9.3) | | | | |
| -2 | | 3.7 (3.6–3.8) | 4.2 (4.1–4.3) | 8.2 (8.1–8.3) | 9.1 (9–9.3) | | | | |
| -1 | | 3.8 (3.7–3.9) | 4.4 (4.3–4.6) | 8 (7.9–8.1) | 9.1 (8.9–9.2) | | | | |
| 0 | | 3.9 (3.8–4) | 4.6 (4.5–4.7) | 7.9 (7.7–8) | 9 (8.9–9.2) | | | | |
| 1 | | 4 (3.9–4.2) | 4.8 (4.7–4.9) | 7.6 (7.5–7.7) | 9 (8.8–9.1) | | | | |
| 2 | | 4.2 (4.1–4.3) | 5 (4.9–5.2) | 7.3 (7.2–7.4) | 8.7 (8.6–8.8) | | | | |
| 3 | | 4.4 (4.3–4.6) | 5.3 (5.1–5.4) | 6.8 (6.7–6.9) | 8.3 (8.1–8.4) | | | | |
| 4 | | 4.7 (4.5–4.8) | 5.4 (5.3–5.6) | 6.2 (6.1–6.4) | 7.7 (7.5–7.8) | | | | |
| 5 | | 4.9 (4.8–5.1) | 5.7 (5.5–5.8) | 5.8 (5.7–5.9) | 7.3 (7.1–7.4) | | | | |
| -5 | 5–14 years | | | | | 3.6 (3.3–3.8) | 3.3 (3.1–3.5) | 9.7 (9.5–10) | 10 (9.7–10.2) |
| -4 | | | | | | 3.6 (3.4–3.9) | 3.4 (3.2–3.6) | 9.5 (9.2–9.8) | 9.9 (9.6–10.1) |
| -3 | | | | | | 3.7 (3.5–4) | 3.6 (3.3–3.8) | 9.4 (9.1–9.7) | 9.8 (9.5–10) |

**eTable 36. Estimates for marginal predictions of annual social welfare benefits among parents exposed to child's mood disorder (F30–F39) and unexposed parents stratified by the child's age at diagnosis**

| | | Finland | | | | Denmark | | | |
|---|---|---|---|---|---|---|---|---|---|
| | | Men | | Women | | Men | | Women | |
| | | Unexposed | Exposed | Unexposed | Exposed | Unexposed | Exposed | Unexposed | Exposed |
| Years since diagnosis | Age group | Marg. pred. (95% CI) | Marg. pred. (95% CI) | Marg. pred. (95% CI) | Marg. pred. (95% CI) | Marg. pred. (95% CI) | Marg. pred. (95% CI) | Marg. pred. (95% CI) | Marg. pred. (95% CI) |
| | | Unexposed | Exposed | Unexposed | Exposed | Unexposed | Exposed | Unexposed | Exposed |
| -2 | | | | | | 3.8 (3.5–4) | 3.6 (3.4–3.9) | 9.3 (9–9.6) | 9.7 (9.4–9.9) |
| -1 | | | | | | 3.8 (3.6–4.1) | 3.7 (3.5–3.9) | 9 (8.7–9.3) | 9.8 (9.5–10) |
| 0 | | | | | | 3.9 (3.7–4.1) | 3.8 (3.6–4) | 8.8 (8.6–9.1) | 9.8 (9.5–10.1) |
| 1 | | | | | | 4.1 (3.9–4.4) | 4.1 (3.8–4.3) | 8.7 (8.4–8.9) | 9.8 (9.5–10.1) |
| 2 | | | | | | 4.2 (4–4.5) | 4.2 (4–4.5) | 8.4 (8.1–8.7) | 9.6 (9.3–9.9) |
| 3 | | | | | | 4.2 (4–4.5) | 4.4 (4.1–4.6) | 8.1 (7.9–8.4) | 9.3 (9–9.6) |
| 4 | | | | | | 4.3 (4–4.6) | 4.5 (4.2–4.8) | 7.5 (7.2–7.8) | 8.8 (8.5–9.2) |
| 5 | | | | | | 4.4 (4.2–4.7) | 4.6 (4.3–4.8) | 7 (6.7–7.3) | 8.3 (8–8.6) |
| -5 | 15–19 years | 3.2 (3.1–3.2) | 3.7 (3.6–3.8) | 7.1 (7.1–7.2) | 7.8 (7.7–7.9) | 3.8 (3.7–3.9) | 4.1 (4–4.2) | 9.3 (9.1–9.4) | 9.8 (9.7–10) |
| -4 | | 3.4 (3.3–3.4) | 3.9 (3.9–4) | 6.9 (6.9–7) | 7.7 (7.6–7.8) | 3.9 (3.8–4) | 4.3 (4.2–4.4) | 9.1 (9–9.3) | 9.7 (9.6–9.9) |
| -3 | | 3.6 (3.5–3.6) | 4.1 (4–4.2) | 6.7 (6.7–6.8) | 7.5 (7.5–7.6) | 4 (3.9–4.1) | 4.4 (4.3–4.6) | 8.8 (8.7–9) | 9.6 (9.5–9.8) |
| -2 | | 3.7 (3.7–3.8) | 4.3 (4.3–4.4) | 6.4 (6.3–6.4) | 7.2 (7.1–7.3) | 4.2 (4–4.3) | 4.5 (4.4–4.7) | 8.6 (8.4–8.7) | 9.4 (9.3–9.6) |
| -1 | | 3.9 (3.8–3.9) | 4.5 (4.4–4.6) | 5.9 (5.8–6) | 6.8 (6.7–6.9) | 4.3 (4.2–4.4) | 4.6 (4.5–4.8) | 8.2 (8.1–8.3) | 9 (8.9–9.2) |

**eTable 36. Estimates for marginal predictions of annual social welfare benefits among parents exposed to child's mood disorder (F30–F39) and unexposed parents stratified by the child's age at diagnosis**

| | | Finland | | | | Denmark | | | |
|---|---|---|---|---|---|---|---|---|---|
| | | Men | | Women | | Men | | Women | |
| | | Unexposed | Exposed | Unexposed | Exposed | Unexposed | Exposed | Unexposed | Exposed |
| Years since diagnosis | Age group | Marg. pred. (95% CI) | Marg. pred. (95% CI) | Marg. pred. (95% CI) | Marg. pred. (95% CI) | Marg. pred. (95% CI) | Marg. pred. (95% CI) | Marg. pred. (95% CI) | Marg. pred. (95% CI) |
| | | Unexposed | Exposed | Unexposed | Exposed | Unexposed | Exposed | Unexposed | Exposed |
| 0 | | 4 (3.9–4.1) | 4.7 (4.6–4.8) | 5.4 (5.4–5.5) | 6.4 (6.3–6.5) | 4.4 (4.3–4.5) | 4.7 (4.6–4.9) | 7.8 (7.6–7.9) | 8.7 (8.5–8.8) |
| 1 | | 4.2 (4.2–4.3) | 4.9 (4.8–5) | 5 (4.9–5.1) | 6 (5.9–6.1) | 4.6 (4.4–4.7) | 4.9 (4.8–5) | 7.4 (7.3–7.5) | 8.3 (8.2–8.5) |
| 2 | | 4.5 (4.4–4.6) | 5.3 (5.2–5.3) | 4.7 (4.6–4.7) | 5.6 (5.6–5.7) | 4.6 (4.5–4.8) | 5.1 (4.9–5.2) | 7 (6.9–7.1) | 8 (7.8–8.1) |
| 3 | | 4.9 (4.8–5) | 5.6 (5.5–5.7) | 4.5 (4.5–4.6) | 5.5 (5.4–5.6) | 4.8 (4.6–4.9) | 5.3 (5.1–5.4) | 6.7 (6.6–6.9) | 7.8 (7.6–8) |
| 4 | | 5.3 (5.2–5.4) | 6 (5.9–6.1) | 4.5 (4.4–4.6) | 5.5 (5.4–5.6) | 5 (4.9–5.2) | 5.6 (5.4–5.7) | 6.6 (6.5–6.8) | 7.8 (7.6–8) |
| 5 | | 5.9 (5.8–6) | 6.6 (6.5–6.7) | 4.6 (4.5–4.7) | 5.6 (5.5–5.6) | 5.3 (5.1–5.4) | 5.8 (5.7–6) | 6.6 (6.4–6.8) | 7.9 (7.7–8.1) |
| -5 | 20–25 years | 3.8 (3.7–3.9) | 4.3 (4.2–4.3) | 5 (4.9–5) | 5.7 (5.6–5.7) | 4.1 (4–4.2) | 4.5 (4.4–4.6) | 7.3 (7.2–7.4) | 8.2 (8.1–8.3) |
| -4 | | 4.1 (4–4.1) | 4.6 (4.5–4.7) | 4.6 (4.6–4.7) | 5.4 (5.3–5.4) | 4.2 (4.1–4.4) | 4.6 (4.5–4.7) | 7 (6.9–7.1) | 7.9 (7.8–8) |
| -3 | | 4.4 (4.3–4.5) | 4.9 (4.9–5) | 4.4 (4.3–4.4) | 5.1 (5–5.1) | 4.4 (4.3–4.5) | 4.8 (4.7–4.9) | 6.7 (6.6–6.8) | 7.6 (7.5–7.8) |
| -2 | | 4.7 (4.7–4.8) | 5.3 (5.2–5.4) | 4.3 (4.2–4.4) | 5 (4.9–5) | 4.5 (4.4–4.6) | 5 (4.9–5.1) | 6.4 (6.3–6.5) | 7.4 (7.3–7.5) |
| -1 | | 5.2 (5.1–5.3) | 5.7 (5.6–5.8) | 4.3 (4.2–4.3) | 5 (5–5.1) | 4.7 (4.6–4.8) | 5.3 (5.2–5.4) | 6.4 (6.2–6.5) | 7.3 (7.2–7.5) |
| 0 | | 5.6 (5.5–5.7) | 6.1 (6–6.2) | 4.3 (4.3–4.4) | 5.1 (5.1–5.2) | 5 (4.8–5.1) | 5.5 (5.4–5.7) | 6.4 (6.3–6.6) | 7.4 (7.3–7.6) |
| 1 | | 6.2 (6.1–6.3) | 6.7 (6.6–6.8) | 4.5 (4.4–4.6) | 5.3 (5.2–5.4) | 5.2 (5.1–5.3) | 5.9 (5.8–6) | 6.6 (6.4–6.7) | 7.6 (7.5–7.8) |

**eTable 36. Estimates for marginal predictions of annual social welfare benefits among parents exposed to child's mood disorder (F30–F39) and unexposed parents stratified by the child's age at diagnosis**

| | | Finland | | | | Denmark | | | |
|---|---|---|---|---|---|---|---|---|---|
| | | Men | | Women | | Men | | Women | |
| | | Unexposed | Exposed | Unexposed | Exposed | Unexposed | Exposed | Unexposed | Exposed |
| Years since diagnosis | Age group | Marg. pred. (95% CI) | Marg. pred. (95% CI) | Marg. pred. (95% CI) | Marg. pred. (95% CI) | Marg. pred. (95% CI) | Marg. pred. (95% CI) | Marg. pred. (95% CI) | Marg. pred. (95% CI) |
| | | Unexposed | Exposed | Unexposed | Exposed | Unexposed | Exposed | Unexposed | Exposed |
| 2 | | 6.9 (6.8–7) | 7.4 (7.3–7.6) | 4.7 (4.7–4.8) | 5.6 (5.5–5.6) | 5.5 (5.4–5.6) | 6.2 (6.1–6.4) | 6.7 (6.6–6.9) | 7.9 (7.7–8) |
| 3 | | 7.7 (7.6–7.8) | 8.2 (8–8.3) | 5.1 (5–5.2) | 5.9 (5.8–5.9) | 5.9 (5.7–6) | 6.6 (6.5–6.8) | 6.9 (6.8–7.1) | 8.1 (8–8.3) |
| 4 | | 8.6 (8.5–8.8) | 8.9 (8.8–9.1) | 5.5 (5.4–5.6) | 6.2 (6.1–6.3) | 6.3 (6.2–6.5) | 7.1 (6.9–7.2) | 7.2 (7.1–7.4) | 8.4 (8.3–8.6) |
| 5 | | 9.7 (9.5–9.8) | 9.9 (9.7–10) | 6 (5.9–6.1) | 6.6 (6.6–6.7) | 6.9 (6.7–7) | 7.6 (7.4–7.7) | 7.6 (7.4–7.8) | 8.8 (8.6–9) |

Marg. pred. = Marginal predictions in thousand euros (€)

**eTable 37. Estimates for marginal predictions of annual employment among parents exposed to child's mood disorder (F30–F39) and unexposed parents stratified by the child's age at diagnosis**

| | | Finland | | | | Denmark | | | |
|---|---|---|---|---|---|---|---|---|---|
| | | Men | | Women | | Men | | Women | |
| | | Unexposed | Exposed | Unexposed | Exposed | Unexposed | Exposed | Unexposed | Exposed |
| Years since diagnosis | Age group | Marg. pred. (95% CI) | Marg. pred. (95% CI) | Marg. pred. (95% CI) | Marg. pred. (95% CI) | Marg. pred. (95% CI) | Marg. pred. (95% CI) | Marg. pred. (95% CI) | Marg. pred. (95% CI) |
| | | Unexposed | Exposed | Unexposed | Exposed | Unexposed | Exposed | Unexposed | Exposed |
| -5 | 5–9 years | 0.78 (0.76–0.8) | 0.74 (0.72–0.76) | 0.61 (0.59–0.63) | 0.55 (0.53–0.57) | | | | |
| -4 | | 0.78 (0.77–0.8) | 0.74 (0.72–0.76) | 0.64 (0.62–0.66) | 0.57 (0.55–0.59) | | | | |
| -3 | | 0.79 (0.77–0.8) | 0.73 (0.71–0.75) | 0.66 (0.64–0.68) | 0.6 (0.58–0.62) | | | | |
| -2 | | 0.78 (0.76–0.8) | 0.74 (0.72–0.76) | 0.67 (0.65–0.69) | 0.6 (0.58–0.62) | | | | |
| -1 | | 0.78 (0.77–0.8) | 0.75 (0.73–0.76) | 0.7 (0.68–0.71) | 0.61 (0.59–0.63) | | | | |
| 0 | | 0.79 (0.77–0.8) | 0.73 (0.71–0.75) | 0.7 (0.68–0.72) | 0.62 (0.6–0.64) | | | | |
| 1 | | 0.79 (0.77–0.8) | 0.72 (0.7–0.73) | 0.71 (0.7–0.73) | 0.64 (0.62–0.66) | | | | |
| 2 | | 0.79 (0.77–0.8) | 0.73 (0.71–0.74) | 0.73 (0.71–0.75) | 0.65 (0.63–0.67) | | | | |
| 3 | | 0.78 (0.76–0.8) | 0.73 (0.71–0.75) | 0.74 (0.72–0.76) | 0.66 (0.64–0.68) | | | | |

**eTable 37. Estimates for marginal predictions of annual employment among parents exposed to child's mood disorder (F30–F39) and unexposed parents stratified by the child's age at diagnosis**

| | | Finland | | | | Denmark | | | |
|---|---|---|---|---|---|---|---|---|---|
| | | Men | | Women | | Men | | Women | |
| | | Unexposed | Exposed | Unexposed | Exposed | Unexposed | Exposed | Unexposed | Exposed |
| Years since diagnosis | Age group | Marg. pred. (95% CI) | Marg. pred. (95% CI) | Marg. pred. (95% CI) | Marg. pred. (95% CI) | Marg. pred. (95% CI) | Marg. pred. (95% CI) | Marg. pred. (95% CI) | Marg. pred. (95% CI) |
| | | Unexposed | Exposed | Unexposed | Exposed | Unexposed | Exposed | Unexposed | Exposed |
| 4 | | 0.77 (0.75–0.79) | 0.71 (0.69–0.73) | 0.75 (0.73–0.77) | 0.67 (0.65–0.69) | | | | |
| 5 | | 0.77 (0.75–0.79) | 0.71 (0.68–0.73) | 0.75 (0.73–0.77) | 0.67 (0.65–0.69) | | | | |
| -5 | 10–14 years | 0.81 (0.8–0.81) | 0.77 (0.77–0.78) | 0.72 (0.72–0.73) | 0.69 (0.68–0.7) | | | | |
| -4 | | 0.8 (0.79–0.81) | 0.77 (0.76–0.78) | 0.73 (0.72–0.74) | 0.7 (0.7–0.71) | | | | |
| -3 | | 0.8 (0.79–0.8) | 0.76 (0.76–0.77) | 0.75 (0.74–0.75) | 0.71 (0.7–0.71) | | | | |
| -2 | | 0.79 (0.79–0.8) | 0.76 (0.75–0.77) | 0.76 (0.75–0.76) | 0.72 (0.71–0.72) | | | | |
| -1 | | 0.8 (0.79–0.8) | 0.76 (0.75–0.76) | 0.77 (0.76–0.77) | 0.73 (0.72–0.73) | | | | |
| 0 | | 0.79 (0.78–0.8) | 0.75 (0.75–0.76) | 0.78 (0.77–0.78) | 0.73 (0.72–0.74) | | | | |
| 1 | | 0.79 (0.78–0.79) | 0.75 (0.74–0.75) | 0.78 (0.78–0.79) | 0.73 (0.73–0.74) | | | | |

**eTable 37. Estimates for marginal predictions of annual employment among parents exposed to child's mood disorder (F30–F39) and unexposed parents stratified by the child's age at diagnosis**

| | | Finland | | | | Denmark | | | |
|---|---|---|---|---|---|---|---|---|---|
| | | Men | | Women | | Men | | Women | |
| | | Unexposed | Exposed | Unexposed | Exposed | Unexposed | Exposed | Unexposed | Exposed |
| Years since diagnosis | Age group | Marg. pred. (95% CI) | Marg. pred. (95% CI) | Marg. pred. (95% CI) | Marg. pred. (95% CI) | Marg. pred. (95% CI) | Marg. pred. (95% CI) | Marg. pred. (95% CI) | Marg. pred. (95% CI) |
| | | Unexposed | Exposed | Unexposed | Exposed | Unexposed | Exposed | Unexposed | Exposed |
| 2 | | 0.78 (0.77–0.78) | 0.74 (0.73–0.74) | 0.78 (0.78–0.79) | 0.73 (0.73–0.74) | | | | |
| 3 | | 0.77 (0.76–0.78) | 0.73 (0.72–0.73) | 0.79 (0.78–0.79) | 0.73 (0.72–0.74) | | | | |
| 4 | | 0.76 (0.75–0.76) | 0.72 (0.71–0.72) | 0.79 (0.78–0.79) | 0.73 (0.73–0.74) | | | | |
| 5 | | 0.75 (0.74–0.76) | 0.7 (0.69–0.71) | 0.79 (0.78–0.8) | 0.73 (0.72–0.74) | | | | |
| -5 | 5–14 years | | | | | 0.76 (0.75–0.78) | 0.78 (0.77–0.79) | 0.72 (0.71–0.74) | 0.72 (0.71–0.74) |
| -4 | | | | | | 0.77 (0.76–0.78) | 0.78 (0.77–0.79) | 0.73 (0.72–0.74) | 0.73 (0.72–0.74) |
| -3 | | | | | | 0.76 (0.75–0.77) | 0.77 (0.76–0.78) | 0.73 (0.72–0.75) | 0.73 (0.72–0.74) |
| -2 | | | | | | 0.76 (0.74–0.77) | 0.77 (0.75–0.78) | 0.73 (0.72–0.74) | 0.74 (0.72–0.75) |
| -1 | | | | | | 0.76 (0.75–0.77) | 0.76 (0.75–0.77) | 0.74 (0.73–0.75) | 0.72 (0.71–0.74) |

**eTable 37. Estimates for marginal predictions of annual employment among parents exposed to child's mood disorder (F30–F39) and unexposed parents stratified by the child's age at diagnosis**

| | | Finland | | | | Denmark | | | |
|---|---|---|---|---|---|---|---|---|---|
| | | Men | | Women | | Men | | Women | |
| | | Unexposed | Exposed | Unexposed | Exposed | Unexposed | Exposed | Unexposed | Exposed |
| Years since diagnosis | Age group | Marg. pred. (95% CI) | Marg. pred. (95% CI) | Marg. pred. (95% CI) | Marg. pred. (95% CI) | Marg. pred. (95% CI) | Marg. pred. (95% CI) | Marg. pred. (95% CI) | Marg. pred. (95% CI) |
| | | Unexposed | Exposed | Unexposed | Exposed | Unexposed | Exposed | Unexposed | Exposed |
| 0 | | | | | | 0.76 (0.75–0.77) | 0.76 (0.74–0.77) | 0.74 (0.73–0.75) | 0.72 (0.71–0.73) |
| 1 | | | | | | 0.75 (0.74–0.76) | 0.75 (0.74–0.76) | 0.74 (0.72–0.75) | 0.72 (0.7–0.73) |
| 2 | | | | | | 0.74 (0.73–0.76) | 0.75 (0.73–0.76) | 0.74 (0.73–0.75) | 0.72 (0.7–0.73) |
| 3 | | | | | | 0.75 (0.73–0.76) | 0.74 (0.73–0.76) | 0.74 (0.72–0.75) | 0.71 (0.7–0.72) |
| 4 | | | | | | 0.74 (0.72–0.75) | 0.74 (0.72–0.75) | 0.73 (0.72–0.74) | 0.7 (0.69–0.72) |
| 5 | | | | | | 0.73 (0.72–0.75) | 0.74 (0.73–0.76) | 0.74 (0.72–0.75) | 0.7 (0.69–0.72) |
| -5 | 15–19 years | 0.81 (0.81–0.82) | 0.78 (0.78–0.79) | 0.79 (0.78–0.79) | 0.75 (0.75–0.75) | 0.75 (0.75–0.76) | 0.74 (0.74–0.75) | 0.71 (0.7–0.72) | 0.71 (0.7–0.72) |
| -4 | | 0.81 (0.81–0.81) | 0.78 (0.77–0.78) | 0.79 (0.79–0.8) | 0.76 (0.76–0.76) | 0.75 (0.75–0.76) | 0.74 (0.74–0.75) | 0.71 (0.71–0.72) | 0.71 (0.7–0.72) |
| -3 | | 0.8 (0.8–0.81) | 0.77 (0.77–0.78) | 0.8 (0.79–0.8) | 0.76 (0.76–0.77) | 0.75 (0.75–0.76) | 0.74 (0.73–0.75) | 0.72 (0.71–0.73) | 0.71 (0.71–0.72) |

**eTable 37. Estimates for marginal predictions of annual employment among parents exposed to child's mood disorder (F30–F39) and unexposed parents stratified by the child’s age at diagnosis**

| | | Finland | | | | Denmark | | | |
|---|---|---|---|---|---|---|---|---|---|
| | | Men | | Women | | Men | | Women | |
| | | Unexposed | Exposed | Unexposed | Exposed | Unexposed | Exposed | Unexposed | Exposed |
| Years since diagnosis | Age group | Marg. pred. (95% CI) | Marg. pred. (95% CI) | Marg. pred. (95% CI) | Marg. pred. (95% CI) | Marg. pred. (95% CI) | Marg. pred. (95% CI) | Marg. pred. (95% CI) | Marg. pred. (95% CI) |
| | | Unexposed | Exposed | Unexposed | Exposed | Unexposed | Exposed | Unexposed | Exposed |
| -2 | | 0.8 (0.79–0.8) | 0.77 (0.76–0.77) | 0.8 (0.8–0.81) | 0.77 (0.76–0.77) | 0.74 (0.74–0.75) | 0.74 (0.73–0.74) | 0.72 (0.72–0.73) | 0.71 (0.7–0.72) |
| -1 | | 0.79 (0.79–0.79) | 0.76 (0.75–0.76) | 0.81 (0.81–0.81) | 0.77 (0.77–0.78) | 0.74 (0.73–0.75) | 0.73 (0.72–0.74) | 0.72 (0.72–0.73) | 0.71 (0.7–0.71) |
| 0 | | 0.78 (0.78–0.79) | 0.75 (0.75–0.75) | 0.81 (0.8–0.81) | 0.77 (0.77–0.78) | 0.73 (0.73–0.74) | 0.73 (0.72–0.73) | 0.72 (0.71–0.73) | 0.7 (0.7–0.71) |
| 1 | | 0.77 (0.77–0.78) | 0.74 (0.73–0.74) | 0.81 (0.8–0.81) | 0.77 (0.76–0.77) | 0.72 (0.72–0.73) | 0.72 (0.71–0.72) | 0.72 (0.71–0.72) | 0.7 (0.69–0.7) |
| 2 | | 0.76 (0.76–0.77) | 0.72 (0.72–0.73) | 0.81 (0.8–0.81) | 0.77 (0.76–0.77) | 0.72 (0.71–0.73) | 0.71 (0.7–0.72) | 0.71 (0.71–0.72) | 0.69 (0.69–0.7) |
| 3 | | 0.75 (0.74–0.75) | 0.71 (0.7–0.71) | 0.8 (0.8–0.81) | 0.76 (0.76–0.77) | 0.71 (0.71–0.72) | 0.7 (0.69–0.71) | 0.71 (0.7–0.71) | 0.68 (0.68–0.69) |
| 4 | | 0.73 (0.73–0.74) | 0.69 (0.69–0.7) | 0.79 (0.79–0.8) | 0.75 (0.75–0.76) | 0.71 (0.7–0.72) | 0.69 (0.68–0.7) | 0.71 (0.7–0.71) | 0.68 (0.67–0.68) |
| 5 | | 0.71 (0.71–0.72) | 0.67 (0.67–0.68) | 0.78 (0.78–0.79) | 0.74 (0.74–0.75) | 0.69 (0.69–0.7) | 0.68 (0.67–0.69) | 0.7 (0.7–0.71) | 0.67 (0.66–0.67) |
| -5 | 20–25 years | 0.79 (0.78–0.79) | 0.76 (0.75–0.76) | 0.8 (0.8–0.8) | 0.77 (0.76–0.77) | 0.74 (0.74–0.75) | 0.73 (0.72–0.73) | 0.72 (0.71–0.72) | 0.69 (0.68–0.69) |

**eTable 37. Estimates for marginal predictions of annual employment among parents exposed to child's mood disorder (F30–F39) and unexposed parents stratified by the child’s age at diagnosis**

| | | Finland | | | | Denmark | | | |
|---|---|---|---|---|---|---|---|---|---|
| | | Men | | Women | | Men | | Women | |
| | | Unexposed | Exposed | Unexposed | Exposed | Unexposed | Exposed | Unexposed | Exposed |
| Years since diagnosis | Age group | Marg. pred. (95% CI) | Marg. pred. (95% CI) | Marg. pred. (95% CI) | Marg. pred. (95% CI) | Marg. pred. (95% CI) | Marg. pred. (95% CI) | Marg. pred. (95% CI) | Marg. pred. (95% CI) |
| | | Unexposed | Exposed | Unexposed | Exposed | Unexposed | Exposed | Unexposed | Exposed |
| -4 | | 0.77 (0.77–0.78) | 0.74 (0.74–0.75) | 0.8 (0.8–0.8) | 0.76 (0.76–0.77) | 0.74 (0.73–0.74) | 0.73 (0.72–0.73) | 0.72 (0.71–0.72) | 0.69 (0.69–0.7) |
| -3 | | 0.76 (0.76–0.76) | 0.73 (0.72–0.73) | 0.8 (0.79–0.8) | 0.76 (0.76–0.76) | 0.74 (0.73–0.74) | 0.72 (0.71–0.73) | 0.72 (0.71–0.72) | 0.69 (0.68–0.7) |
| -2 | | 0.74 (0.74–0.75) | 0.71 (0.71–0.72) | 0.79 (0.79–0.79) | 0.76 (0.75–0.76) | 0.73 (0.72–0.73) | 0.71 (0.7–0.71) | 0.71 (0.71–0.72) | 0.68 (0.67–0.69) |
| -1 | | 0.73 (0.72–0.73) | 0.7 (0.69–0.7) | 0.78 (0.78–0.79) | 0.75 (0.75–0.75) | 0.72 (0.71–0.72) | 0.7 (0.69–0.7) | 0.71 (0.7–0.71) | 0.68 (0.67–0.68) |
| 0 | | 0.71 (0.71–0.72) | 0.68 (0.68–0.68) | 0.78 (0.77–0.78) | 0.74 (0.73–0.74) | 0.71 (0.7–0.71) | 0.69 (0.68–0.69) | 0.7 (0.69–0.7) | 0.67 (0.66–0.67) |
| 1 | | 0.69 (0.69–0.69) | 0.66 (0.65–0.66) | 0.76 (0.76–0.77) | 0.72 (0.72–0.73) | 0.69 (0.69–0.7) | 0.67 (0.66–0.68) | 0.69 (0.68–0.69) | 0.65 (0.65–0.66) |
| 2 | | 0.66 (0.66–0.67) | 0.64 (0.63–0.64) | 0.75 (0.74–0.75) | 0.71 (0.7–0.71) | 0.68 (0.67–0.69) | 0.65 (0.65–0.66) | 0.68 (0.67–0.68) | 0.64 (0.63–0.64) |
| 3 | | 0.64 (0.63–0.64) | 0.61 (0.6–0.61) | 0.73 (0.73–0.74) | 0.69 (0.69–0.7) | 0.66 (0.66–0.67) | 0.64 (0.63–0.64) | 0.67 (0.66–0.67) | 0.62 (0.61–0.63) |
| 4 | | 0.61 (0.6–0.61) | 0.58 (0.57–0.59) | 0.71 (0.71–0.72) | 0.68 (0.67–0.68) | 0.65 (0.64–0.65) | 0.61 (0.61–0.62) | 0.65 (0.65–0.66) | 0.61 (0.6–0.61) |

**eTable 37. Estimates for marginal predictions of annual employment among parents exposed to child's mood disorder (F30–F39) and unexposed parents stratified by the child's age at diagnosis**

| | | Finland | | | | Denmark | | | |
|---|---|---|---|---|---|---|---|---|---|
| | | Men | | Women | | Men | | Women | |
| | | Unexposed | Exposed | Unexposed | Exposed | Unexposed | Exposed | Unexposed | Exposed |
| Years since diagnosis | Age group | Marg. pred. (95% CI) | Marg. pred. (95% CI) | Marg. pred. (95% CI) | Marg. pred. (95% CI) | Marg. pred. (95% CI) | Marg. pred. (95% CI) | Marg. pred. (95% CI) | Marg. pred. (95% CI) |
| | | Unexposed | Exposed | Unexposed | Exposed | Unexposed | Exposed | Unexposed | Exposed |
| 5 | | 0.57 (0.56–0.58) | 0.55 (0.54–0.56) | 0.69 (0.68–0.69) | 0.65 (0.65–0.66) | 0.62 (0.61–0.63) | 0.59 (0.58–0.6) | 0.63 (0.62–0.64) | 0.59 (0.58–0.6) |

Marg. pred. = Marginal predictions in probabilities

**eTable 38. Estimates for marginal predictions of annual earnings among parents exposed to child's anxiety disorder (F40–F48) and unexposed parents stratified by the child's age at diagnosis**

| | | Finland | | | | Denmark | | | |
|---|---|---|---|---|---|---|---|---|---|
| | | Men | | Women | | Men | | Women | |
| | | Unexposed | Exposed | Unexposed | Exposed | Unexposed | Exposed | Unexposed | Exposed |
| Years since diagnosis | Age group | Marg. pred. (95% CI) | Marg. pred. (95% CI) | Marg. pred. (95% CI) | Marg. pred. (95% CI) | Marg. pred. (95% CI) | Marg. pred. (95% CI) | Marg. pred. (95% CI) | Marg. pred. (95% CI) |
| | | Unexposed | Exposed | Unexposed | Exposed | Unexposed | Exposed | Unexposed | Exposed |
| -5 | 5–9 years | 30 (29.4–30.5) | 27.8 (27.3–28.3) | 12.3 (12–12.7) | 11.4 (11.1–11.8) | 52.3 (51.5–53.1) | 49.9 (49.2–50.7) | 39.9 (39.4–40.3) | 38.9 (38.5–39.3) |
| -4 | | 30.9 (30.4–31.5) | 28.6 (28–29.1) | 14.3 (14–14.7) | 13.3 (13–13.7) | 53.8 (52.9–54.7) | 51 (50.3–51.8) | 41.1 (40.6–41.6) | 40 (39.6–40.4) |
| -3 | | 31.8 (31.2–32.3) | 29.2 (28.7–29.8) | 16.1 (15.7–16.5) | 14.8 (14.4–15.1) | 54.9 (54–55.8) | 52.3 (51.5–53.1) | 42 (41.5–42.4) | 41.1 (40.7–41.5) |
| -2 | | 32.7 (32.1–33.3) | 29.7 (29.1–30.2) | 17.3 (16.9–17.7) | 15.8 (15.4–16.1) | 56 (55.1–56.9) | 53.6 (52.8–54.4) | 42.9 (42.4–43.4) | 41.8 (41.3–42.2) |
| -1 | | 33.5 (32.9–34.1) | 30 (29.4–30.6) | 18.3 (18–18.7) | 16.6 (16.2–17) | 56.8 (55.8–57.8) | 54.7 (53.9–55.5) | 43.8 (43.3–44.3) | 42.6 (42.1–43) |
| 0 | | 34.3 (33.7–34.9) | 30.6 (30–31.2) | 19.7 (19.3–20.1) | 17.3 (16.9–17.7) | 57.9 (56.9–58.8) | 55.9 (55–56.8) | 44.7 (44.2–45.2) | 42.9 (42.4–43.3) |
| 1 | | 35 (34.4–35.7) | 31 (30.4–31.6) | 20.8 (20.4–21.2) | 18.2 (17.8–18.6) | 59 (58–60) | 56.7 (55.8–57.6) | 45.6 (45.1–46.2) | 43.9 (43.4–44.4) |
| 2 | | 35.5 (34.8–36.1) | 31.5 (30.9–32.1) | 21.8 (21.4–22.2) | 19.3 (18.9–19.7) | 60 (58.9–61) | 57.7 (56.8–58.7) | 46.4 (45.9–47) | 44.6 (44.2–45.1) |
| 3 | | 36.1 (35.5–36.8) | 32 (31.3–32.6) | 23.1 (22.6–23.5) | 19.9 (19.5–20.4) | 61.2 (60.1–62.3) | 57.9 (56.9–58.9) | 47.4 (46.8–47.9) | 45.2 (44.7–45.8) |
| 4 | | 36.7 (36–37.4) | 32.3 (31.6–33) | 24.2 (23.7–24.6) | 20.8 (20.3–21.2) | 62.3 (61.1–63.5) | 58.7 (57.7–59.8) | 48.1 (47.5–48.7) | 45.8 (45.2–46.3) |

**eTable 38. Estimates for marginal predictions of annual earnings among parents exposed to child's anxiety disorder (F40–F48) and unexposed parents stratified by the child's age at diagnosis**

| | | Finland | | | | Denmark | | | |
|---|---|---|---|---|---|---|---|---|---|
| | | Men | | Women | | Men | | Women | |
| | | Unexposed | Exposed | Unexposed | Exposed | Unexposed | Exposed | Unexposed | Exposed |
| Years since diagnosis | Age group | Marg. pred. (95% CI) | Marg. pred. (95% CI) | Marg. pred. (95% CI) | Marg. pred. (95% CI) | Marg. pred. (95% CI) | Marg. pred. (95% CI) | Marg. pred. (95% CI) | Marg. pred. (95% CI) |
| | | Unexposed | Exposed | Unexposed | Exposed | Unexposed | Exposed | Unexposed | Exposed |
| 5 | | 37.3 (36.6–38.1) | 32.5 (31.8–33.2) | 24.8 (24.3–25.3) | 21.7 (21.2–22.1) | 62.9 (61.7–64.2) | 59.3 (58.2–60.4) | 48.7 (48.1–49.3) | 46.4 (45.8–47) |
| -5 | 10–14 years | 35 (34.6–35.3) | 32.9 (32.5–33.2) | 20.6 (20.4–20.9) | 19.4 (19.2–19.7) | 55.6 (55.1–56) | 54.1 (53.6–54.5) | 42.9 (42.6–43.1) | 42.1 (41.9–42.3) |
| -4 | | 35.6 (35.3–36) | 33.4 (33.1–33.8) | 21.6 (21.4–21.9) | 20.4 (20.1–20.6) | 56.6 (56.1–57.1) | 54.9 (54.4–55.3) | 43.6 (43.4–43.9) | 42.8 (42.6–43) |
| -3 | | 36.2 (35.8–36.6) | 33.8 (33.4–34.1) | 22.8 (22.5–23) | 21.2 (20.9–21.4) | 57.4 (56.9–57.9) | 55.6 (55.1–56) | 44.5 (44.2–44.7) | 43.5 (43.3–43.7) |
| -2 | | 36.4 (36.1–36.8) | 34.2 (33.8–34.5) | 23.7 (23.5–24) | 21.9 (21.7–22.2) | 58.3 (57.8–58.9) | 56.2 (55.8–56.7) | 45.1 (44.8–45.3) | 44.2 (44–44.4) |
| -1 | | 36.8 (36.5–37.2) | 34.5 (34.1–34.8) | 24.6 (24.4–24.9) | 22.7 (22.5–23) | 59.3 (58.8–59.8) | 56.9 (56.4–57.3) | 45.8 (45.6–46.1) | 44.9 (44.6–45.1) |
| 0 | | 37.3 (36.9–37.7) | 34.7 (34.3–35) | 25.5 (25.2–25.7) | 23.2 (23–23.5) | 60.3 (59.7–60.8) | 57.4 (56.9–57.9) | 46.5 (46.2–46.8) | 45.4 (45.1–45.6) |
| 1 | | 37.6 (37.2–38) | 34.8 (34.4–35.2) | 26.2 (26–26.5) | 23.8 (23.5–24) | 60.9 (60.4–61.5) | 58.2 (57.7–58.7) | 47.2 (46.9–47.5) | 46 (45.7–46.2) |
| 2 | | 37.8 (37.4–38.3) | 35.1 (34.7–35.5) | 27 (26.8–27.3) | 24.4 (24.2–24.7) | 61.7 (61.1–62.2) | 58.8 (58.2–59.3) | 47.7 (47.4–48) | 46.3 (46.1–46.6) |

**eTable 38. Estimates for marginal predictions of annual earnings among parents exposed to child's anxiety disorder (F40–F48) and unexposed parents stratified by the child's age at diagnosis**

| | | Finland | | | | Denmark | | | |
|---|---|---|---|---|---|---|---|---|---|
| | | Men | | Women | | Men | | Women | |
| | | Unexposed | Exposed | Unexposed | Exposed | Unexposed | Exposed | Unexposed | Exposed |
| Years since diagnosis | Age group | Marg. pred. (95% CI) | Marg. pred. (95% CI) | Marg. pred. (95% CI) | Marg. pred. (95% CI) | Marg. pred. (95% CI) | Marg. pred. (95% CI) | Marg. pred. (95% CI) | Marg. pred. (95% CI) |
| | | Unexposed | Exposed | Unexposed | Exposed | Unexposed | Exposed | Unexposed | Exposed |
| 3 | | 38.2 (37.8–38.6) | 35.2 (34.7–35.6) | 27.7 (27.5–28) | 25 (24.8–25.3) | 61.9 (61.4–62.5) | 59.2 (58.6–59.7) | 48 (47.7–48.3) | 46.5 (46.2–46.8) |
| 4 | | 38.3 (37.9–38.8) | 34.9 (34.5–35.3) | 28.1 (27.8–28.4) | 25.5 (25.2–25.8) | 62.5 (61.8–63.1) | 59.5 (59–60.1) | 48.4 (48.1–48.7) | 46.5 (46.2–46.8) |
| 5 | | 38.2 (37.7–38.7) | 34.8 (34.3–35.3) | 28.6 (28.4–28.9) | 26 (25.7–26.3) | 62.9 (62.2–63.5) | 59.8 (59.2–60.4) | 48.5 (48.1–48.8) | 46.5 (46.2–46.8) |
| -5 | 15–19 years | 34.6 (34.4–34.9) | 32.7 (32.4–32.9) | 22.8 (22.7–23) | 21.5 (21.4–21.7) | 55.4 (55–55.7) | 53.1 (52.8–53.4) | 42.9 (42.7–43.1) | 42.1 (41.9–42.3) |
| -4 | | 35 (34.8–35.2) | 32.9 (32.7–33.1) | 23.6 (23.5–23.8) | 22.3 (22.1–22.4) | 56 (55.6–56.3) | 53.5 (53.2–53.9) | 43.6 (43.4–43.7) | 42.6 (42.5–42.8) |
| -3 | | 35.3 (35.1–35.5) | 33.1 (32.9–33.4) | 24.3 (24.2–24.5) | 22.9 (22.8–23.1) | 56.4 (56.1–56.8) | 54.1 (53.7–54.4) | 44.1 (43.9–44.2) | 43.1 (43–43.3) |
| -2 | | 35.5 (35.2–35.7) | 33.3 (33.1–33.6) | 24.9 (24.7–25) | 23.5 (23.4–23.7) | 57.1 (56.8–57.5) | 54.5 (54.2–54.8) | 44.5 (44.3–44.7) | 43.4 (43.2–43.6) |
| -1 | | 35.8 (35.5–36) | 33.4 (33.2–33.7) | 25.4 (25.3–25.6) | 24.1 (23.9–24.2) | 57.8 (57.5–58.2) | 55.1 (54.8–55.4) | 44.8 (44.6–45) | 43.7 (43.5–43.8) |
| 0 | | 35.8 (35.6–36.1) | 33.4 (33.2–33.7) | 26 (25.8–26.1) | 24.4 (24.3–24.6) | 58.4 (58–58.8) | 55.4 (55.1–55.8) | 44.9 (44.8–45.1) | 43.6 (43.4–43.8) |

**eTable 38. Estimates for marginal predictions of annual earnings among parents exposed to child's anxiety disorder (F40–F48) and unexposed parents stratified by the child’s age at diagnosis**

| | | Finland | | | | Denmark | | | |
|---|---|---|---|---|---|---|---|---|---|
| | | Men | | Women | | Men | | Women | |
| | | Unexposed | Exposed | Unexposed | Exposed | Unexposed | Exposed | Unexposed | Exposed |
| Years since diagnosis | Age group | Marg. pred. (95% CI) | Marg. pred. (95% CI) | Marg. pred. (95% CI) | Marg. pred. (95% CI) | Marg. pred. (95% CI) | Marg. pred. (95% CI) | Marg. pred. (95% CI) | Marg. pred. (95% CI) |
| | | Unexposed | Exposed | Unexposed | Exposed | Unexposed | Exposed | Unexposed | Exposed |
| 1 | | 35.9 (35.7–36.2) | 33.4 (33.2–33.7) | 26.4 (26.2–26.6) | 24.8 (24.7–25) | 58.6 (58.2–59) | 55.7 (55.4–56.1) | 45 (44.8–45.2) | 43.6 (43.4–43.8) |
| 2 | | 35.7 (35.5–36) | 33.3 (33–33.6) | 26.8 (26.6–27) | 25.2 (25–25.4) | 59 (58.6–59.4) | 55.8 (55.4–56.2) | 45 (44.8–45.2) | 43.5 (43.3–43.7) |
| 3 | | 35.5 (35.3–35.8) | 32.9 (32.7–33.2) | 27 (26.9–27.2) | 25.5 (25.3–25.6) | 59 (58.6–59.4) | 55.8 (55.5–56.2) | 45 (44.8–45.2) | 43.4 (43.2–43.6) |
| 4 | | 35.1 (34.8–35.4) | 32.4 (32.2–32.7) | 27.2 (27–27.4) | 25.5 (25.4–25.7) | 58.7 (58.3–59.2) | 55.7 (55.4–56.1) | 45.1 (44.9–45.3) | 43.4 (43.2–43.6) |
| 5 | | 34.6 (34.3–34.9) | 32 (31.7–32.3) | 27.3 (27.1–27.5) | 25.6 (25.4–25.8) | 58.7 (58.3–59.2) | 55.5 (55.1–55.9) | 45.2 (45–45.4) | 43.5 (43.3–43.7) |
| -5 | 20–25 years | 33.8 (33.5–34) | 31.9 (31.7–32.2) | 24.2 (24–24.3) | 22.8 (22.6–22.9) | 55.7 (55.3–56) | 53.1 (52.8–53.4) | 42 (41.8–42.2) | 40.9 (40.8–41.1) |
| -4 | | 33.8 (33.5–34) | 31.8 (31.6–32.1) | 24.5 (24.4–24.7) | 23.1 (22.9–23.2) | 55.9 (55.5–56.3) | 53.2 (52.8–53.5) | 42 (41.9–42.2) | 40.9 (40.7–41.1) |
| -3 | | 33.6 (33.4–33.9) | 31.6 (31.4–31.9) | 24.8 (24.7–25) | 23.3 (23.1–23.5) | 56 (55.6–56.4) | 53.1 (52.8–53.5) | 42 (41.8–42.2) | 40.8 (40.6–41) |
| -2 | | 33.3 (33.1–33.6) | 31.3 (31–31.5) | 24.9 (24.8–25.1) | 23.4 (23.3–23.6) | 56.1 (55.7–56.4) | 53.1 (52.7–53.4) | 42 (41.8–42.1) | 40.8 (40.6–40.9) |

**eTable 38. Estimates for marginal predictions of annual earnings among parents exposed to child's anxiety disorder (F40–F48) and unexposed parents stratified by the child's age at diagnosis**

| | | Finland | | | | Denmark | | | |
|---|---|---|---|---|---|---|---|---|---|
| | | Men | | Women | | Men | | Women | |
| | | Unexposed | Exposed | Unexposed | Exposed | Unexposed | Exposed | Unexposed | Exposed |
| Years since diagnosis | Age group | Marg. pred. (95% CI) | Marg. pred. (95% CI) | Marg. pred. (95% CI) | Marg. pred. (95% CI) | Marg. pred. (95% CI) | Marg. pred. (95% CI) | Marg. pred. (95% CI) | Marg. pred. (95% CI) |
| | | Unexposed | Exposed | Unexposed | Exposed | Unexposed | Exposed | Unexposed | Exposed |
| -1 | | 32.9 (32.7–33.2) | 30.9 (30.7–31.2) | 25.1 (24.9–25.2) | 23.5 (23.3–23.6) | 56.2 (55.8–56.5) | 53.1 (52.7–53.4) | 42 (41.9–42.2) | 40.9 (40.7–41) |
| 0 | | 32.4 (32.1–32.7) | 30.4 (30.2–30.7) | 25.1 (24.9–25.2) | 23.5 (23.3–23.6) | 56.2 (55.8–56.6) | 53 (52.6–53.4) | 42.2 (42–42.4) | 40.9 (40.8–41.1) |
| 1 | | 31.8 (31.6–32.1) | 29.9 (29.6–30.1) | 25 (24.8–25.2) | 23.4 (23.2–23.5) | 56.2 (55.8–56.6) | 53 (52.7–53.4) | 42.3 (42.1–42.5) | 41 (40.8–41.2) |
| 2 | | 31.1 (30.8–31.4) | 29 (28.8–29.3) | 24.8 (24.7–25) | 23.2 (23–23.3) | 56 (55.6–56.4) | 52.8 (52.5–53.2) | 42.3 (42.1–42.5) | 41.1 (40.9–41.3) |
| 3 | | 30.3 (30–30.6) | 28.3 (28–28.6) | 24.6 (24.4–24.8) | 22.9 (22.7–23.1) | 55.7 (55.3–56.1) | 52.4 (52.1–52.8) | 42.4 (42.2–42.6) | 41.1 (40.9–41.3) |
| 4 | | 29.2 (28.9–29.6) | 27.4 (27–27.7) | 24.2 (24–24.4) | 22.6 (22.4–22.8) | 55.5 (55.1–56) | 52 (51.6–52.4) | 42.5 (42.3–42.7) | 41.1 (40.9–41.3) |
| 5 | | 27.9 (27.6–28.3) | 26.2 (25.9–26.5) | 23.7 (23.5–23.9) | 22.2 (22–22.4) | 54.8 (54.4–55.3) | 51.6 (51.2–52) | 42.4 (42.2–42.6) | 41.1 (40.9–41.3) |

Marg. pred. = Marginal predictions in thousand euros (€)

**eTable 39. Estimates for marginal predictions of annual disposable income among parents exposed to child's anxiety disorder (F40–F48) and unexposed parents stratified by the child's age at diagnosis**

| | | Finland | | | | Denmark | | | |
|---|---|---|---|---|---|---|---|---|---|
| | | Men | | Women | | Men | | Women | |
| | | Unexposed | Exposed | Unexposed | Exposed | Unexposed | Exposed | Unexposed | Exposed |
| Years since diagnosis | Age group | Marg. pred. (95% CI) | Marg. pred. (95% CI) | Marg. pred. (95% CI) | Marg. pred. (95% CI) | Marg. pred. (95% CI) | Marg. pred. (95% CI) | Marg. pred. (95% CI) | Marg. pred. (95% CI) |
| | | Unexposed | Exposed | Unexposed | Exposed | Unexposed | Exposed | Unexposed | Exposed |
| -5 | 5–9 years | 25.5 (25.1–25.8) | 24 (23.7–24.3) | 18.9 (18.7–19.1) | 18.9 (18.7–19.1) | 31.7 (31.2–32.1) | 30.2 (29.8–30.6) | 27.6 (27.4–27.9) | 27.2 (26.9–27.4) |
| -4 | | 26.1 (25.8–26.5) | 24.6 (24.3–25) | 20.1 (19.8–20.3) | 20.1 (19.9–20.3) | 32.9 (32.4–33.4) | 31.1 (30.7–31.5) | 28.6 (28.3–28.9) | 28.1 (27.9–28.4) |
| -3 | | 26.9 (26.5–27.3) | 25.3 (24.9–25.6) | 21.2 (21–21.4) | 21.3 (21.1–21.5) | 33.9 (33.4–34.4) | 32.1 (31.7–32.6) | 29.5 (29.2–29.7) | 29 (28.8–29.3) |
| -2 | | 27.6 (27.2–28) | 25.8 (25.5–26.2) | 22.1 (21.9–22.3) | 22.1 (21.9–22.3) | 34.7 (34.2–35.2) | 33.1 (32.6–33.5) | 30.3 (30–30.6) | 29.6 (29.4–29.9) |
| -1 | | 28.3 (27.9–28.7) | 26.2 (25.8–26.5) | 22.9 (22.7–23.1) | 22.9 (22.7–23.1) | 35.6 (35–36.1) | 34 (33.5–34.4) | 31.1 (30.8–31.4) | 30.3 (30–30.6) |
| 0 | | 28.9 (28.5–29.3) | 26.8 (26.4–27.1) | 23.9 (23.6–24.1) | 23.6 (23.4–23.8) | 36.5 (36–37.1) | 35 (34.4–35.5) | 31.9 (31.5–32.2) | 30.7 (30.4–31) |
| 1 | | 29.5 (29.1–29.9) | 27.3 (26.9–27.7) | 24.6 (24.4–24.9) | 24.2 (24–24.5) | 37.6 (37–38.2) | 35.7 (35.2–36.2) | 32.6 (32.3–32.9) | 31.6 (31.3–31.9) |
| 2 | | 30 (29.6–30.4) | 27.7 (27.3–28.1) | 25.3 (25.1–25.6) | 24.8 (24.6–25) | 38.6 (38–39.2) | 36.7 (36.1–37.2) | 33.3 (33–33.7) | 32.1 (31.8–32.4) |
| 3 | | 30.6 (30.1–31) | 28.3 (27.9–28.7) | 26.1 (25.8–26.3) | 25.3 (25.1–25.6) | 39.6 (38.9–40.2) | 37.1 (36.6–37.7) | 34.1 (33.7–34.4) | 32.6 (32.3–32.9) |

**eTable 39. Estimates for marginal predictions of annual disposable income among parents exposed to child's anxiety disorder (F40–F48) and unexposed parents stratified by the child's age at diagnosis**

| | | Finland | | | | Denmark | | | |
|---|---|---|---|---|---|---|---|---|---|
| | | Men | | Women | | Men | | Women | |
| | | Unexposed | Exposed | Unexposed | Exposed | Unexposed | Exposed | Unexposed | Exposed |
| Years since diagnosis | Age group | Marg. pred. (95% CI) | Marg. pred. (95% CI) | Marg. pred. (95% CI) | Marg. pred. (95% CI) | Marg. pred. (95% CI) | Marg. pred. (95% CI) | Marg. pred. (95% CI) | Marg. pred. (95% CI) |
| | | Unexposed | Exposed | Unexposed | Exposed | Unexposed | Exposed | Unexposed | Exposed |
| 4 | | 31.1 (30.6–31.6) | 28.7 (28.2–29.1) | 26.8 (26.5–27.1) | 25.8 (25.6–26.1) | 40.6 (39.9–41.3) | 38.2 (37.5–38.8) | 34.8 (34.4–35.1) | 33.2 (32.8–33.5) |
| 5 | | 31.6 (31.1–32.1) | 29.1 (28.6–29.5) | 27.1 (26.8–27.4) | 26.4 (26.1–26.7) | 41.3 (40.6–42.1) | 38.8 (38.1–39.4) | 35.3 (34.9–35.7) | 33.7 (33.3–34.1) |
| -5 | 10–14 years | 28.8 (28.5–29) | 27.5 (27.3–27.7) | 23.5 (23.3–23.6) | 23 (22.9–23.2) | 33.8 (33.5–34) | 32.7 (32.4–32.9) | 29.9 (29.7–30) | 29.3 (29.1–29.4) |
| -4 | | 29.3 (29.1–29.6) | 28 (27.8–28.2) | 24.1 (24–24.2) | 23.7 (23.6–23.9) | 34.7 (34.4–34.9) | 33.5 (33.2–33.7) | 30.5 (30.4–30.7) | 29.9 (29.8–30.1) |
| -3 | | 29.8 (29.6–30.1) | 28.4 (28.2–28.7) | 24.8 (24.7–25) | 24.4 (24.2–24.5) | 35.5 (35.3–35.8) | 34.2 (34–34.5) | 31.2 (31.1–31.4) | 30.6 (30.4–30.7) |
| -2 | | 30.3 (30–30.5) | 28.9 (28.7–29.2) | 25.5 (25.3–25.6) | 25 (24.8–25.1) | 36.5 (36.2–36.7) | 35 (34.7–35.2) | 31.8 (31.7–32) | 31.2 (31–31.3) |
| -1 | | 30.7 (30.4–30.9) | 29.3 (29–29.5) | 26 (25.9–26.2) | 25.5 (25.4–25.6) | 37.3 (37–37.6) | 35.7 (35.4–35.9) | 32.5 (32.3–32.6) | 31.7 (31.6–31.9) |
| 0 | | 31.2 (30.9–31.4) | 29.6 (29.3–29.8) | 26.5 (26.4–26.6) | 25.9 (25.8–26.1) | 38.2 (37.9–38.5) | 36.3 (36–36.6) | 33 (32.9–33.2) | 32.2 (32.1–32.4) |
| 1 | | 31.6 (31.4–31.9) | 29.9 (29.7–30.2) | 26.9 (26.7–27) | 26.2 (26.1–26.4) | 39 (38.7–39.3) | 37.1 (36.8–37.4) | 33.6 (33.5–33.8) | 32.8 (32.6–32.9) |

**eTable 39. Estimates for marginal predictions of annual disposable income among parents exposed to child's anxiety disorder (F40–F48) and unexposed parents stratified by the child's age at diagnosis**

| | | Finland | | | | Denmark | | | |
|---|---|---|---|---|---|---|---|---|---|
| | | Men | | Women | | Men | | Women | |
| | | Unexposed | Exposed | Unexposed | Exposed | Unexposed | Exposed | Unexposed | Exposed |
| Years since diagnosis | Age group | Marg. pred. (95% CI) | Marg. pred. (95% CI) | Marg. pred. (95% CI) | Marg. pred. (95% CI) | Marg. pred. (95% CI) | Marg. pred. (95% CI) | Marg. pred. (95% CI) | Marg. pred. (95% CI) |
| | | Unexposed | Exposed | Unexposed | Exposed | Unexposed | Exposed | Unexposed | Exposed |
| 2 | | 32.1 (31.8–32.3) | 30.3 (30–30.6) | 27.2 (27.1–27.4) | 26.5 (26.4–26.6) | 39.9 (39.6–40.2) | 37.9 (37.6–38.2) | 34.1 (34–34.3) | 33.1 (33–33.3) |
| 3 | | 32.5 (32.2–32.8) | 30.6 (30.3–30.9) | 27.2 (27.1–27.4) | 26.4 (26.3–26.6) | 40.5 (40.1–40.8) | 38.5 (38.2–38.8) | 34.5 (34.3–34.7) | 33.3 (33.2–33.5) |
| 4 | | 32.8 (32.5–33.1) | 30.7 (30.4–31) | 27 (26.9–27.2) | 26.2 (26.1–26.4) | 41.3 (41–41.7) | 39.3 (38.9–39.6) | 34.8 (34.6–35) | 33.3 (33.2–33.5) |
| 5 | | 33.1 (32.8–33.4) | 30.9 (30.6–31.2) | 27 (26.9–27.2) | 26.2 (26.1–26.4) | 42 (41.7–42.4) | 40 (39.7–40.4) | 34.9 (34.7–35.1) | 33.4 (33.2–33.6) |
| -5 | 15–19 years | 28.6 (28.4–28.7) | 27.3 (27.2–27.5) | 23.7 (23.6–23.8) | 23.4 (23.3–23.4) | 33.7 (33.5–33.9) | 32.2 (32.1–32.4) | 29.7 (29.6–29.8) | 29.1 (29–29.2) |
| -4 | | 29 (28.9–29.2) | 27.7 (27.5–27.8) | 24.2 (24.1–24.3) | 23.8 (23.8–23.9) | 34.5 (34.3–34.7) | 32.9 (32.7–33) | 30.3 (30.2–30.4) | 29.6 (29.5–29.7) |
| -3 | | 29.4 (29.3–29.6) | 28 (27.9–28.2) | 24.5 (24.5–24.6) | 24.2 (24.1–24.3) | 35.1 (34.9–35.3) | 33.5 (33.3–33.7) | 30.8 (30.7–30.9) | 30 (29.9–30.1) |
| -2 | | 29.8 (29.6–29.9) | 28.4 (28.3–28.6) | 24.6 (24.6–24.7) | 24.3 (24.3–24.4) | 35.9 (35.7–36.1) | 34.1 (34–34.3) | 31.2 (31.1–31.3) | 30.3 (30.2–30.4) |
| -1 | | 30.2 (30–30.4) | 28.7 (28.5–28.9) | 24.7 (24.6–24.8) | 24.4 (24.3–24.4) | 36.7 (36.5–36.9) | 34.8 (34.6–35) | 31.4 (31.3–31.5) | 30.5 (30.4–30.6) |

**eTable 39. Estimates for marginal predictions of annual disposable income among parents exposed to child's anxiety disorder (F40–F48) and unexposed parents stratified by the child's age at diagnosis**

| | | Finland | | | | Denmark | | | |
|---|---|---|---|---|---|---|---|---|---|
| | | Men | | Women | | Men | | Women | |
| | | Unexposed | Exposed | Unexposed | Exposed | Unexposed | Exposed | Unexposed | Exposed |
| Years since diagnosis | Age group | Marg. pred. (95% CI) | Marg. pred. (95% CI) | Marg. pred. (95% CI) | Marg. pred. (95% CI) | Marg. pred. (95% CI) | Marg. pred. (95% CI) | Marg. pred. (95% CI) | Marg. pred. (95% CI) |
| | | Unexposed | Exposed | Unexposed | Exposed | Unexposed | Exposed | Unexposed | Exposed |
| 0 | | 30.5 (30.3–30.6) | 28.9 (28.7–29.1) | 24.6 (24.6–24.7) | 24.3 (24.2–24.4) | 37.4 (37.2–37.6) | 35.4 (35.2–35.6) | 31.5 (31.4–31.6) | 30.5 (30.4–30.6) |
| 1 | | 30.8 (30.7–31) | 29.2 (29–29.4) | 24.6 (24.5–24.7) | 24.2 (24.1–24.3) | 38 (37.7–38.2) | 36 (35.8–36.2) | 31.6 (31.5–31.7) | 30.5 (30.4–30.6) |
| 2 | | 31 (30.8–31.2) | 29.4 (29.2–29.6) | 24.6 (24.5–24.7) | 24.2 (24.1–24.3) | 38.7 (38.4–38.9) | 36.5 (36.3–36.7) | 31.6 (31.5–31.8) | 30.4 (30.3–30.6) |
| 3 | | 31.3 (31.1–31.5) | 29.6 (29.4–29.8) | 24.7 (24.6–24.8) | 24.3 (24.2–24.4) | 39.1 (38.8–39.3) | 37 (36.8–37.2) | 31.7 (31.6–31.9) | 30.5 (30.3–30.6) |
| 4 | | 31.5 (31.3–31.7) | 29.8 (29.6–30) | 24.9 (24.8–25) | 24.4 (24.3–24.5) | 39.4 (39.1–39.6) | 37.4 (37.2–37.6) | 31.9 (31.8–32) | 30.6 (30.5–30.7) |
| 5 | | 31.7 (31.5–31.9) | 29.9 (29.7–30.1) | 25.1 (25–25.2) | 24.6 (24.5–24.7) | 39.9 (39.6–40.2) | 37.7 (37.5–38) | 32.2 (32.1–32.3) | 30.8 (30.7–31) |
| -5 | 20–25 years | 28.6 (28.4–28.7) | 27.3 (27.2–27.5) | 22.9 (22.8–23) | 22.4 (22.3–22.5) | 34.2 (34–34.4) | 32.6 (32.4–32.8) | 28.5 (28.4–28.6) | 27.7 (27.6–27.8) |
| -4 | | 28.8 (28.6–29) | 27.5 (27.3–27.7) | 22.8 (22.7–22.9) | 22.3 (22.2–22.4) | 34.8 (34.6–35) | 33.1 (32.9–33.3) | 28.6 (28.5–28.7) | 27.8 (27.7–27.9) |
| -3 | | 29 (28.8–29.1) | 27.7 (27.5–27.8) | 22.8 (22.7–22.9) | 22.2 (22.1–22.3) | 35.4 (35.2–35.6) | 33.5 (33.3–33.7) | 28.7 (28.6–28.8) | 27.8 (27.7–27.9) |

**eTable 39. Estimates for marginal predictions of annual disposable income among parents exposed to child's anxiety disorder (F40–F48) and unexposed parents stratified by the child's age at diagnosis**

| | | Finland | | | | Denmark | | | |
|---|---|---|---|---|---|---|---|---|---|
| | | Men | | Women | | Men | | Women | |
| | | Unexposed | Exposed | Unexposed | Exposed | Unexposed | Exposed | Unexposed | Exposed |
| Years since diagnosis | Age group | Marg. pred. (95% CI) | Marg. pred. (95% CI) | Marg. pred. (95% CI) | Marg. pred. (95% CI) | Marg. pred. (95% CI) | Marg. pred. (95% CI) | Marg. pred. (95% CI) | Marg. pred. (95% CI) |
| | | Unexposed | Exposed | Unexposed | Exposed | Unexposed | Exposed | Unexposed | Exposed |
| -2 | | 29.1 (29–29.3) | 27.8 (27.7–28) | 22.8 (22.7–22.9) | 22.3 (22.2–22.3) | 35.9 (35.7–36.1) | 33.9 (33.7–34.1) | 28.8 (28.6–28.9) | 27.8 (27.7–27.9) |
| -1 | | 29.3 (29.1–29.5) | 27.9 (27.8–28.1) | 22.9 (22.8–23) | 22.3 (22.2–22.4) | 36.3 (36.1–36.5) | 34.3 (34.1–34.5) | 29 (28.8–29.1) | 28 (27.9–28.1) |
| 0 | | 29.4 (29.2–29.6) | 28.1 (28–28.3) | 23 (22.9–23.1) | 22.4 (22.3–22.5) | 36.7 (36.5–37) | 34.6 (34.4–34.8) | 29.2 (29.1–29.3) | 28.2 (28.1–28.3) |
| 1 | | 29.5 (29.3–29.7) | 28.2 (28.1–28.4) | 23.1 (23–23.2) | 22.5 (22.4–22.6) | 37.1 (36.9–37.4) | 34.9 (34.7–35.2) | 29.5 (29.3–29.6) | 28.4 (28.3–28.5) |
| 2 | | 29.6 (29.4–29.8) | 28.3 (28.1–28.5) | 23.3 (23.2–23.4) | 22.6 (22.5–22.7) | 37.5 (37.2–37.7) | 35.3 (35.1–35.5) | 29.7 (29.5–29.8) | 28.7 (28.5–28.8) |
| 3 | | 29.8 (29.6–30) | 28.5 (28.3–28.6) | 23.4 (23.3–23.5) | 22.7 (22.6–22.8) | 37.7 (37.4–37.9) | 35.4 (35.2–35.6) | 29.9 (29.8–30.1) | 28.9 (28.8–29) |
| 4 | | 29.9 (29.7–30.1) | 28.5 (28.3–28.7) | 23.6 (23.5–23.7) | 22.9 (22.8–23) | 37.9 (37.7–38.2) | 35.5 (35.3–35.8) | 30.2 (30–30.3) | 29.1 (29–29.2) |
| 5 | | 29.8 (29.6–30) | 28.6 (28.4–28.8) | 23.7 (23.6–23.8) | 23 (22.9–23.1) | 38 (37.8–38.3) | 35.7 (35.4–35.9) | 30.4 (30.2–30.5) | 29.3 (29.1–29.4) |

Marg. pred. = Marginal predictions in thousand euros (€)

**eTable 40. Estimates for marginal predictions of annual social welfare benefits among parents exposed to child's anxiety disorder (F40–F48) and unexposed parents stratified by the child's age at diagnosis**

| | | Finland | | | | Denmark | | | |
|---|---|---|---|---|---|---|---|---|---|
| | | Men | | Women | | Men | | Women | |
| | | Unexposed | Exposed | Unexposed | Exposed | Unexposed | Exposed | Unexposed | Exposed |
| Years since diagnosis | Age group | Marg. pred. (95% CI) | Marg. pred. (95% CI) | Marg. pred. (95% CI) | Marg. pred. (95% CI) | Marg. pred. (95% CI) | Marg. pred. (95% CI) | Marg. pred. (95% CI) | Marg. pred. (95% CI) |
| | | Unexposed | Exposed | Unexposed | Exposed | Unexposed | Exposed | Unexposed | Exposed |
| -5 | 5–9 years | 3.2 (3.1–3.4) | 3.6 (3.5–3.8) | 10.3 (10.2–10.5) | 11.1 (10.9–11.2) | 3.9 (3.7–4.1) | 3.9 (3.6–4.1) | 11.9 (11.6–12.1) | 12.8 (12.6–13.1) |
| -4 | | 3.3 (3.1–3.4) | 3.8 (3.6–3.9) | 9.8 (9.6–10) | 10.6 (10.5–10.8) | 3.9 (3.7–4.1) | 4.1 (3.9–4.3) | 11.5 (11.3–11.8) | 12.7 (12.5–13) |
| -3 | | 3.4 (3.3–3.6) | 3.9 (3.7–4) | 9.6 (9.4–9.7) | 10.6 (10.5–10.8) | 4 (3.8–4.2) | 4.2 (4–4.4) | 11.3 (11.1–11.6) | 12.6 (12.3–12.9) |
| -2 | | 3.4 (3.2–3.5) | 4 (3.9–4.2) | 9.5 (9.3–9.6) | 10.6 (10.4–10.8) | 4 (3.8–4.2) | 4.3 (4.1–4.5) | 11.1 (10.8–11.4) | 12.5 (12.2–12.8) |
| -1 | | 3.4 (3.2–3.5) | 4.2 (4–4.3) | 9.4 (9.2–9.6) | 10.7 (10.5–10.9) | 3.9 (3.7–4.2) | 4.3 (4–4.5) | 10.8 (10.5–11.1) | 12.3 (12–12.6) |
| 0 | | 3.5 (3.4–3.7) | 4.4 (4.2–4.6) | 9.3 (9.1–9.5) | 10.8 (10.7–11) | 4 (3.7–4.2) | 4.5 (4.3–4.8) | 10.5 (10.2–10.8) | 12.4 (12.1–12.7) |
| 1 | | 3.7 (3.5–3.8) | 4.6 (4.4–4.8) | 9.3 (9.1–9.5) | 10.8 (10.6–10.9) | 4 (3.8–4.3) | 4.6 (4.4–4.9) | 10.3 (10–10.5) | 12.3 (12–12.6) |
| 2 | | 3.8 (3.7–4) | 4.7 (4.5–4.9) | 9.1 (9–9.3) | 10.5 (10.3–10.8) | 4 (3.8–4.3) | 4.7 (4.4–4.9) | 10 (9.7–10.3) | 12 (11.7–12.4) |
| 3 | | 3.9 (3.7–4.1) | 4.9 (4.7–5.1) | 8.8 (8.6–9) | 10.6 (10.3–10.8) | 4.1 (3.9–4.4) | 4.7 (4.5–5) | 9.9 (9.6–10.2) | 11.8 (11.4–12.1) |

**eTable 40. Estimates for marginal predictions of annual social welfare benefits among parents exposed to child's anxiety disorder (F40–F48) and unexposed parents stratified by the child’s age at diagnosis**

| | | Finland | | | | Denmark | | | |
|---|---|---|---|---|---|---|---|---|---|
| | | Men | | Women | | Men | | Women | |
| | | Unexposed | Exposed | Unexposed | Exposed | Unexposed | Exposed | Unexposed | Exposed |
| Years since diagnosis | Age group | Marg. pred. (95% CI) | Marg. pred. (95% CI) | Marg. pred. (95% CI) | Marg. pred. (95% CI) | Marg. pred. (95% CI) | Marg. pred. (95% CI) | Marg. pred. (95% CI) | Marg. pred. (95% CI) |
| | | Unexposed | Exposed | Unexposed | Exposed | Unexposed | Exposed | Unexposed | Exposed |
| 4 | | 4.1 (3.9–4.3) | 5 (4.8–5.3) | 8.7 (8.5–8.9) | 10.4 (10.2–10.7) | 4.3 (4.1–4.6) | 4.8 (4.5–5.1) | 9.7 (9.4–10) | 11.6 (11.2–11.9) |
| 5 | | 4.1 (3.9–4.3) | 5.3 (5–5.5) | 8.6 (8.3–8.8) | 10.3 (10.1–10.5) | 4.4 (4.2–4.7) | 4.8 (4.5–5.1) | 9.5 (9.2–9.8) | 11.4 (11.1–11.8) |
| -5 | 10–14 years | 3.1 (3–3.2) | 3.5 (3.5–3.6) | 8.3 (8.2–8.3) | 8.8 (8.7–8.9) | 3.9 (3.8–4) | 4 (3.9–4.1) | 10.6 (10.5–10.7) | 11.3 (11.2–11.4) |
| -4 | | 3.2 (3.1–3.3) | 3.7 (3.6–3.8) | 8.2 (8.1–8.3) | 8.8 (8.7–8.9) | 3.9 (3.8–4) | 4.1 (4–4.2) | 10.4 (10.2–10.5) | 11.2 (11–11.3) |
| -3 | | 3.3 (3.2–3.4) | 3.9 (3.8–4) | 8 (7.9–8.1) | 8.9 (8.8–9) | 4 (3.9–4.1) | 4.2 (4.1–4.3) | 10.2 (10.1–10.3) | 11.1 (11–11.3) |
| -2 | | 3.5 (3.4–3.6) | 4.1 (4–4.2) | 7.9 (7.8–8) | 8.9 (8.8–9) | 4.1 (4–4.2) | 4.3 (4.2–4.4) | 10.1 (9.9–10.2) | 11 (10.9–11.2) |
| -1 | | 3.6 (3.5–3.7) | 4.3 (4.2–4.4) | 7.8 (7.7–7.9) | 8.8 (8.7–8.9) | 4.2 (4.1–4.3) | 4.4 (4.3–4.5) | 9.8 (9.7–10) | 11 (10.9–11.2) |
| 0 | | 3.7 (3.6–3.8) | 4.5 (4.4–4.6) | 7.6 (7.5–7.7) | 8.8 (8.7–8.9) | 4.3 (4.1–4.4) | 4.6 (4.5–4.7) | 9.7 (9.5–9.8) | 11.1 (11–11.3) |
| 1 | | 3.9 (3.8–4) | 4.7 (4.6–4.8) | 7.4 (7.3–7.5) | 8.7 (8.6–8.8) | 4.5 (4.3–4.6) | 4.7 (4.6–4.9) | 9.5 (9.3–9.6) | 11.1 (10.9–11.3) |
| 2 | | 4.1 (4–4.2) | 4.9 (4.8–5.1) | 7.1 (7–7.2) | 8.5 (8.4–8.6) | 4.5 (4.3–4.6) | 4.9 (4.7–5) | 9.3 (9.1–9.4) | 11 (10.8–11.1) |
| 3 | | 4.3 (4.1–4.4) | 5.2 (5–5.3) | 6.5 (6.4–6.7) | 7.9 (7.8–8.1) | 4.5 (4.4–4.7) | 5 (4.9–5.2) | 9 (8.8–9.1) | 10.6 (10.5–10.8) |

**eTable 40. Estimates for marginal predictions of annual social welfare benefits among parents exposed to child's anxiety disorder (F40–F48) and unexposed parents stratified by the child's age at diagnosis**

| | | Finland | | | | Denmark | | | |
|---|---|---|---|---|---|---|---|---|---|
| | | Men | | Women | | Men | | Women | |
| | | Unexposed | Exposed | Unexposed | Exposed | Unexposed | Exposed | Unexposed | Exposed |
| Years since diagnosis | Age group | Marg. pred. (95% CI) | Marg. pred. (95% CI) | Marg. pred. (95% CI) | Marg. pred. (95% CI) | Marg. pred. (95% CI) | Marg. pred. (95% CI) | Marg. pred. (95% CI) | Marg. pred. (95% CI) |
| | | Unexposed | Exposed | Unexposed | Exposed | Unexposed | Exposed | Unexposed | Exposed |
| 4 | | 4.4 (4.3–4.6) | 5.3 (5.2–5.4) | 6.1 (5.9–6.2) | 7.4 (7.2–7.5) | 4.7 (4.5–4.8) | 5.1 (5–5.2) | 8.4 (8.3–8.6) | 10.1 (9.9–10.3) |
| 5 | | 4.6 (4.5–4.8) | 5.6 (5.4–5.7) | 5.6 (5.5–5.7) | 6.9 (6.8–7.1) | 4.8 (4.7–5) | 5.3 (5.1–5.4) | 8 (7.8–8.2) | 9.7 (9.5–9.8) |
| -5 | 15–19 years | 3.2 (3.1–3.3) | 3.7 (3.6–3.8) | 7 (6.9–7) | 7.7 (7.6–7.7) | 4.2 (4.1–4.3) | 4.7 (4.6–4.8) | 9.6 (9.5–9.7) | 10.8 (10.7–10.9) |
| -4 | | 3.4 (3.3–3.5) | 3.9 (3.8–4) | 6.8 (6.7–6.9) | 7.6 (7.5–7.6) | 4.4 (4.3–4.4) | 4.9 (4.8–5) | 9.4 (9.3–9.5) | 10.7 (10.6–10.8) |
| -3 | | 3.6 (3.5–3.7) | 4.1 (4–4.2) | 6.6 (6.6–6.7) | 7.4 (7.4–7.5) | 4.5 (4.4–4.6) | 5.1 (5–5.2) | 9.3 (9.2–9.4) | 10.6 (10.5–10.7) |
| -2 | | 3.8 (3.7–3.8) | 4.3 (4.2–4.4) | 6.2 (6.2–6.3) | 7.1 (7–7.1) | 4.6 (4.5–4.7) | 5.2 (5.1–5.3) | 9.1 (9–9.2) | 10.5 (10.4–10.6) |
| -1 | | 3.9 (3.8–4) | 4.5 (4.4–4.6) | 5.8 (5.8–5.9) | 6.7 (6.6–6.7) | 4.6 (4.6–4.7) | 5.3 (5.2–5.4) | 8.8 (8.7–8.9) | 10.2 (10.1–10.3) |
| 0 | | 4.1 (4–4.2) | 4.6 (4.6–4.7) | 5.4 (5.3–5.4) | 6.3 (6.2–6.4) | 4.7 (4.6–4.8) | 5.4 (5.3–5.5) | 8.3 (8.2–8.4) | 9.9 (9.8–10) |
| 1 | | 4.3 (4.2–4.4) | 4.9 (4.8–5) | 4.9 (4.9–5) | 5.9 (5.8–6) | 4.8 (4.7–4.9) | 5.6 (5.5–5.7) | 7.9 (7.8–8) | 9.5 (9.4–9.6) |
| 2 | | 4.6 (4.5–4.6) | 5.2 (5.1–5.3) | 4.6 (4.6–4.7) | 5.6 (5.5–5.6) | 4.9 (4.8–5) | 5.7 (5.6–5.8) | 7.4 (7.3–7.5) | 9.1 (9–9.2) |
| 3 | | 4.9 (4.8–5) | 5.6 (5.5–5.7) | 4.5 (4.4–4.6) | 5.4 (5.3–5.5) | 5.1 (5–5.2) | 5.9 (5.8–6) | 7 (6.9–7.2) | 8.8 (8.7–8.9) |

**eTable 40. Estimates for marginal predictions of annual social welfare benefits among parents exposed to child's anxiety disorder (F40–F48) and unexposed parents stratified by the child's age at diagnosis**

| | | Finland | | | | Denmark | | | |
|---|---|---|---|---|---|---|---|---|---|
| | | Men | | Women | | Men | | Women | |
| | | Unexposed | Exposed | Unexposed | Exposed | Unexposed | Exposed | Unexposed | Exposed |
| Years since diagnosis | Age group | Marg. pred. (95% CI) | Marg. pred. (95% CI) | Marg. pred. (95% CI) | Marg. pred. (95% CI) | Marg. pred. (95% CI) | Marg. pred. (95% CI) | Marg. pred. (95% CI) | Marg. pred. (95% CI) |
| | | Unexposed | Exposed | Unexposed | Exposed | Unexposed | Exposed | Unexposed | Exposed |
| 4 | | 5.4 (5.3–5.5) | 6 (5.9–6.1) | 4.5 (4.4–4.6) | 5.4 (5.3–5.5) | 5.4 (5.3–5.5) | 6.2 (6–6.3) | 7 (6.9–7.1) | 8.8 (8.6–8.9) |
| 5 | | 5.9 (5.8–6) | 6.5 (6.4–6.6) | 4.6 (4.5–4.6) | 5.5 (5.4–5.6) | 5.7 (5.6–5.8) | 6.5 (6.3–6.6) | 7 (6.9–7.1) | 8.8 (8.7–9) |
| -5 | 20–25 years | 3.8 (3.7–3.8) | 4.2 (4.2–4.3) | 4.9 (4.9–5) | 5.6 (5.6–5.7) | 4.2 (4.1–4.3) | 5 (4.9–5.1) | 7.3 (7.2–7.4) | 8.7 (8.6–8.8) |
| -4 | | 4 (4–4.1) | 4.5 (4.4–4.6) | 4.6 (4.6–4.7) | 5.3 (5.3–5.4) | 4.4 (4.3–4.5) | 5.2 (5.1–5.3) | 7 (6.9–7.1) | 8.4 (8.3–8.5) |
| -3 | | 4.3 (4.3–4.4) | 4.9 (4.8–4.9) | 4.3 (4.3–4.4) | 5 (5–5.1) | 4.6 (4.5–4.7) | 5.4 (5.3–5.5) | 6.7 (6.6–6.8) | 8.2 (8.1–8.3) |
| -2 | | 4.7 (4.6–4.8) | 5.2 (5.2–5.3) | 4.2 (4.2–4.3) | 4.9 (4.9–5) | 4.7 (4.7–4.8) | 5.6 (5.5–5.7) | 6.5 (6.4–6.6) | 8 (7.9–8.1) |
| -1 | | 5.1 (5–5.2) | 5.6 (5.5–5.7) | 4.2 (4.2–4.3) | 5 (4.9–5) | 4.9 (4.8–5) | 5.9 (5.8–6) | 6.5 (6.4–6.6) | 7.9 (7.8–8) |
| 0 | | 5.6 (5.5–5.7) | 6.1 (6–6.2) | 4.3 (4.2–4.3) | 5 (5–5.1) | 5.2 (5.1–5.3) | 6.1 (6–6.2) | 6.5 (6.4–6.6) | 8 (7.9–8.1) |
| 1 | | 6.1 (6–6.2) | 6.7 (6.6–6.8) | 4.5 (4.4–4.5) | 5.2 (5.2–5.3) | 5.5 (5.4–5.6) | 6.4 (6.3–6.5) | 6.5 (6.4–6.6) | 8.1 (8–8.2) |
| 2 | | 6.7 (6.6–6.8) | 7.3 (7.2–7.4) | 4.7 (4.6–4.8) | 5.5 (5.4–5.5) | 5.8 (5.6–5.9) | 6.8 (6.7–6.9) | 6.7 (6.6–6.8) | 8.3 (8.2–8.4) |
| 3 | | 7.5 (7.4–7.6) | 8 (7.9–8.1) | 5 (4.9–5.1) | 5.7 (5.7–5.8) | 6.1 (6–6.2) | 7.2 (7–7.3) | 6.9 (6.8–7) | 8.5 (8.4–8.7) |
| 4 | | 8.4 (8.2–8.5) | 8.8 (8.7–8.9) | 5.4 (5.3–5.4) | 6.1 (6–6.1) | 6.5 (6.4–6.6) | 7.6 (7.5–7.7) | 7.1 (7–7.2) | 8.9 (8.7–9) |
| 5 | | 9.3 (9.2–9.5) | 9.7 (9.5–9.8) | 5.9 (5.8–5.9) | 6.5 (6.4–6.6) | 7 (6.9–7.2) | 8.1 (8–8.2) | 7.5 (7.4–7.6) | 9.2 (9.1–9.4) |

Marg. pred. = Marginal predictions in thousand euros (€)

**eTable 41. Estimates for marginal predictions of annual employment among parents exposed to child's anxiety disorder (F40–F48) and unexposed parents stratified by the child's age at diagnosis**

| | | Finland | | | | Denmark | | | |
|---|---|---|---|---|---|---|---|---|---|
| | | Men | | Women | | Men | | Women | |
| | | Unexposed | Exposed | Unexposed | Exposed | Unexposed | Exposed | Unexposed | Exposed |
| Years since diagnosis | Age group | Marg. pred. (95% CI) | Marg. pred. (95% CI) | Marg. pred. (95% CI) | Marg. pred. (95% CI) | Marg. pred. (95% CI) | Marg. pred. (95% CI) | Marg. pred. (95% CI) | Marg. pred. (95% CI) |
| | | Unexposed | Exposed | Unexposed | Exposed | Unexposed | Exposed | Unexposed | Exposed |
| -5 | 5–9 years | 0.8 (0.79–0.81) | 0.76 (0.75–0.77) | 0.58 (0.56–0.59) | 0.54 (0.53–0.55) | 0.73 (0.72–0.74) | 0.74 (0.73–0.75) | 0.63 (0.62–0.64) | 0.6 (0.59–0.62) |
| -4 | | 0.8 (0.79–0.81) | 0.75 (0.74–0.76) | 0.61 (0.6–0.62) | 0.58 (0.56–0.59) | 0.75 (0.74–0.76) | 0.75 (0.74–0.76) | 0.66 (0.65–0.67) | 0.63 (0.61–0.64) |
| -3 | | 0.8 (0.79–0.8) | 0.75 (0.74–0.76) | 0.64 (0.63–0.65) | 0.6 (0.59–0.61) | 0.75 (0.74–0.76) | 0.75 (0.74–0.77) | 0.67 (0.66–0.68) | 0.64 (0.63–0.66) |
| -2 | | 0.8 (0.79–0.81) | 0.75 (0.74–0.76) | 0.67 (0.66–0.68) | 0.62 (0.61–0.63) | 0.75 (0.74–0.76) | 0.75 (0.74–0.76) | 0.68 (0.67–0.7) | 0.66 (0.64–0.67) |
| -1 | | 0.79 (0.79–0.8) | 0.75 (0.74–0.76) | 0.69 (0.67–0.7) | 0.63 (0.62–0.64) | 0.75 (0.74–0.76) | 0.75 (0.74–0.77) | 0.69 (0.68–0.71) | 0.66 (0.65–0.67) |
| 0 | | 0.8 (0.79–0.81) | 0.75 (0.74–0.76) | 0.7 (0.69–0.71) | 0.64 (0.63–0.65) | 0.74 (0.73–0.75) | 0.75 (0.74–0.76) | 0.7 (0.68–0.71) | 0.65 (0.64–0.66) |
| 1 | | 0.8 (0.79–0.81) | 0.74 (0.73–0.75) | 0.71 (0.7–0.72) | 0.65 (0.64–0.66) | 0.74 (0.72–0.75) | 0.74 (0.72–0.75) | 0.7 (0.69–0.72) | 0.64 (0.63–0.66) |
| 2 | | 0.79 (0.78–0.8) | 0.74 (0.73–0.75) | 0.73 (0.72–0.74) | 0.66 (0.65–0.67) | 0.74 (0.73–0.76) | 0.74 (0.73–0.75) | 0.72 (0.7–0.73) | 0.65 (0.64–0.67) |
| 3 | | 0.79 (0.78–0.8) | 0.73 (0.72–0.74) | 0.75 (0.74–0.76) | 0.67 (0.66–0.68) | 0.74 (0.73–0.75) | 0.74 (0.73–0.75) | 0.72 (0.71–0.73) | 0.66 (0.64–0.67) |

**eTable 41. Estimates for marginal predictions of annual employment among parents exposed to child's anxiety disorder (F40–F48) and unexposed parents stratified by the child's age at diagnosis**

| | | Finland | | | | Denmark | | | |
|---|---|---|---|---|---|---|---|---|---|
| | | Men | | Women | | Men | | Women | |
| | | Unexposed | Exposed | Unexposed | Exposed | Unexposed | Exposed | Unexposed | Exposed |
| Years since diagnosis | Age group | Marg. pred. (95% CI) | Marg. pred. (95% CI) | Marg. pred. (95% CI) | Marg. pred. (95% CI) | Marg. pred. (95% CI) | Marg. pred. (95% CI) | Marg. pred. (95% CI) | Marg. pred. (95% CI) |
| | | Unexposed | Exposed | Unexposed | Exposed | Unexposed | Exposed | Unexposed | Exposed |
| 4 | | 0.79 (0.78–0.8) | 0.73 (0.72–0.74) | 0.76 (0.75–0.77) | 0.68 (0.67–0.69) | 0.73 (0.72–0.74) | 0.73 (0.72–0.75) | 0.72 (0.71–0.73) | 0.66 (0.64–0.67) |
| 5 | | 0.79 (0.78–0.8) | 0.72 (0.71–0.73) | 0.77 (0.76–0.78) | 0.69 (0.68–0.7) | 0.72 (0.71–0.74) | 0.73 (0.71–0.74) | 0.72 (0.7–0.73) | 0.66 (0.65–0.68) |
| -5 | 10–14 years | 0.82 (0.81–0.82) | 0.79 (0.78–0.79) | 0.74 (0.73–0.74) | 0.71 (0.7–0.71) | 0.75 (0.74–0.75) | 0.75 (0.74–0.75) | 0.68 (0.68–0.69) | 0.67 (0.66–0.67) |
| -4 | | 0.81 (0.81–0.82) | 0.78 (0.78–0.79) | 0.75 (0.74–0.76) | 0.71 (0.7–0.72) | 0.75 (0.74–0.76) | 0.75 (0.74–0.76) | 0.69 (0.69–0.7) | 0.68 (0.67–0.68) |
| -3 | | 0.81 (0.8–0.81) | 0.77 (0.77–0.78) | 0.76 (0.76–0.77) | 0.72 (0.71–0.73) | 0.75 (0.74–0.76) | 0.75 (0.75–0.76) | 0.7 (0.69–0.71) | 0.68 (0.68–0.69) |
| -2 | | 0.8 (0.8–0.81) | 0.77 (0.76–0.77) | 0.77 (0.77–0.78) | 0.73 (0.72–0.74) | 0.75 (0.74–0.75) | 0.75 (0.74–0.75) | 0.7 (0.69–0.71) | 0.68 (0.68–0.69) |
| -1 | | 0.8 (0.8–0.81) | 0.76 (0.76–0.77) | 0.78 (0.77–0.78) | 0.74 (0.73–0.74) | 0.74 (0.74–0.75) | 0.74 (0.74–0.75) | 0.71 (0.7–0.71) | 0.68 (0.68–0.69) |
| 0 | | 0.8 (0.8–0.81) | 0.76 (0.76–0.77) | 0.79 (0.78–0.79) | 0.74 (0.73–0.75) | 0.74 (0.74–0.75) | 0.74 (0.73–0.74) | 0.71 (0.7–0.72) | 0.68 (0.67–0.68) |
| 1 | | 0.79 (0.79–0.8) | 0.75 (0.74–0.76) | 0.8 (0.79–0.8) | 0.74 (0.74–0.75) | 0.74 (0.73–0.74) | 0.73 (0.73–0.74) | 0.71 (0.7–0.72) | 0.67 (0.66–0.68) |

**eTable 41. Estimates for marginal predictions of annual employment among parents exposed to child's anxiety disorder (F40–F48) and unexposed parents stratified by the child's age at diagnosis**

| | | Finland | | | | Denmark | | | |
|---|---|---|---|---|---|---|---|---|---|
| | | Men | | Women | | Men | | Women | |
| | | Unexposed | Exposed | Unexposed | Exposed | Unexposed | Exposed | Unexposed | Exposed |
| Years since diagnosis | Age group | Marg. pred. (95% CI) | Marg. pred. (95% CI) | Marg. pred. (95% CI) | Marg. pred. (95% CI) | Marg. pred. (95% CI) | Marg. pred. (95% CI) | Marg. pred. (95% CI) | Marg. pred. (95% CI) |
| | | Unexposed | Exposed | Unexposed | Exposed | Unexposed | Exposed | Unexposed | Exposed |
| 2 | | 0.79 (0.78–0.79) | 0.74 (0.74–0.75) | 0.8 (0.8–0.81) | 0.74 (0.74–0.75) | 0.73 (0.73–0.74) | 0.72 (0.72–0.73) | 0.71 (0.7–0.72) | 0.67 (0.66–0.67) |
| 3 | | 0.78 (0.78–0.79) | 0.73 (0.73–0.74) | 0.81 (0.8–0.81) | 0.74 (0.74–0.75) | 0.73 (0.72–0.74) | 0.72 (0.71–0.73) | 0.71 (0.71–0.72) | 0.67 (0.66–0.68) |
| 4 | | 0.77 (0.77–0.78) | 0.73 (0.72–0.73) | 0.8 (0.79–0.81) | 0.75 (0.74–0.75) | 0.73 (0.72–0.73) | 0.72 (0.71–0.73) | 0.71 (0.71–0.72) | 0.67 (0.66–0.68) |
| 5 | | 0.77 (0.76–0.77) | 0.71 (0.71–0.72) | 0.8 (0.79–0.81) | 0.74 (0.74–0.75) | 0.72 (0.71–0.73) | 0.71 (0.7–0.72) | 0.71 (0.7–0.72) | 0.66 (0.66–0.67) |
| -5 | 15–19 years | 0.81 (0.81–0.82) | 0.79 (0.78–0.79) | 0.79 (0.78–0.79) | 0.75 (0.75–0.76) | 0.73 (0.73–0.74) | 0.72 (0.72–0.72) | 0.69 (0.69–0.7) | 0.66 (0.65–0.66) |
| -4 | | 0.81 (0.8–0.81) | 0.78 (0.77–0.78) | 0.79 (0.79–0.8) | 0.76 (0.75–0.76) | 0.74 (0.73–0.74) | 0.72 (0.72–0.73) | 0.7 (0.7–0.71) | 0.67 (0.66–0.67) |
| -3 | | 0.8 (0.8–0.81) | 0.77 (0.77–0.77) | 0.8 (0.79–0.8) | 0.76 (0.76–0.77) | 0.74 (0.73–0.74) | 0.72 (0.71–0.72) | 0.71 (0.7–0.71) | 0.67 (0.67–0.67) |
| -2 | | 0.79 (0.79–0.8) | 0.76 (0.76–0.77) | 0.8 (0.8–0.81) | 0.77 (0.76–0.77) | 0.73 (0.72–0.73) | 0.71 (0.71–0.72) | 0.71 (0.7–0.71) | 0.67 (0.66–0.67) |
| -1 | | 0.79 (0.79–0.79) | 0.76 (0.75–0.76) | 0.8 (0.8–0.81) | 0.77 (0.76–0.77) | 0.72 (0.72–0.73) | 0.71 (0.7–0.71) | 0.71 (0.7–0.71) | 0.67 (0.66–0.67) |

**eTable 41. Estimates for marginal predictions of annual employment among parents exposed to child's anxiety disorder (F40–F48) and unexposed parents stratified by the child's age at diagnosis**

| | | Finland | | | | Denmark | | | |
|---|---|---|---|---|---|---|---|---|---|
| | | Men | | Women | | Men | | Women | |
| | | Unexposed | Exposed | Unexposed | Exposed | Unexposed | Exposed | Unexposed | Exposed |
| Years since diagnosis | Age group | Marg. pred. (95% CI) | Marg. pred. (95% CI) | Marg. pred. (95% CI) | Marg. pred. (95% CI) | Marg. pred. (95% CI) | Marg. pred. (95% CI) | Marg. pred. (95% CI) | Marg. pred. (95% CI) |
| | | Unexposed | Exposed | Unexposed | Exposed | Unexposed | Exposed | Unexposed | Exposed |
| 0 | | 0.78 (0.78–0.78) | 0.75 (0.74–0.75) | 0.81 (0.8–0.81) | 0.77 (0.76–0.77) | 0.72 (0.71–0.72) | 0.7 (0.7–0.7) | 0.71 (0.7–0.71) | 0.66 (0.65–0.66) |
| 1 | | 0.77 (0.77–0.78) | 0.74 (0.73–0.74) | 0.8 (0.8–0.81) | 0.77 (0.76–0.77) | 0.71 (0.71–0.72) | 0.69 (0.69–0.7) | 0.7 (0.7–0.71) | 0.65 (0.65–0.66) |
| 2 | | 0.76 (0.76–0.77) | 0.72 (0.72–0.73) | 0.8 (0.8–0.8) | 0.76 (0.76–0.77) | 0.71 (0.7–0.71) | 0.68 (0.68–0.69) | 0.7 (0.7–0.71) | 0.65 (0.65–0.66) |
| 3 | | 0.75 (0.74–0.75) | 0.71 (0.7–0.71) | 0.8 (0.79–0.8) | 0.76 (0.76–0.76) | 0.7 (0.69–0.7) | 0.68 (0.67–0.68) | 0.7 (0.69–0.7) | 0.65 (0.64–0.65) |
| 4 | | 0.73 (0.73–0.73) | 0.69 (0.69–0.7) | 0.79 (0.79–0.8) | 0.75 (0.75–0.76) | 0.69 (0.68–0.69) | 0.66 (0.66–0.67) | 0.69 (0.68–0.69) | 0.64 (0.63–0.64) |
| 5 | | 0.71 (0.71–0.72) | 0.67 (0.67–0.68) | 0.78 (0.78–0.79) | 0.74 (0.74–0.75) | 0.68 (0.67–0.68) | 0.65 (0.65–0.66) | 0.68 (0.68–0.69) | 0.63 (0.62–0.63) |
| -5 | 20–25 years | 0.78 (0.78–0.79) | 0.75 (0.75–0.76) | 0.8 (0.8–0.81) | 0.77 (0.77–0.77) | 0.72 (0.72–0.73) | 0.7 (0.69–0.7) | 0.7 (0.7–0.71) | 0.66 (0.65–0.66) |
| -4 | | 0.77 (0.77–0.78) | 0.74 (0.74–0.75) | 0.8 (0.8–0.8) | 0.77 (0.76–0.77) | 0.73 (0.73–0.74) | 0.7 (0.7–0.71) | 0.72 (0.71–0.72) | 0.67 (0.66–0.67) |
| -3 | | 0.76 (0.75–0.76) | 0.73 (0.72–0.73) | 0.8 (0.79–0.8) | 0.76 (0.76–0.76) | 0.73 (0.73–0.74) | 0.7 (0.69–0.7) | 0.71 (0.71–0.72) | 0.67 (0.66–0.67) |

**eTable 41. Estimates for marginal predictions of annual employment among parents exposed to child's anxiety disorder (F40–F48) and unexposed parents stratified by the child's age at diagnosis**

| | | Finland | | | | Denmark | | | |
|---|---|---|---|---|---|---|---|---|---|
| | | Men | | Women | | Men | | Women | |
| | | Unexposed | Exposed | Unexposed | Exposed | Unexposed | Exposed | Unexposed | Exposed |
| Years since diagnosis | Age group | Marg. pred. (95% CI) | Marg. pred. (95% CI) | Marg. pred. (95% CI) | Marg. pred. (95% CI) | Marg. pred. (95% CI) | Marg. pred. (95% CI) | Marg. pred. (95% CI) | Marg. pred. (95% CI) |
| | | Unexposed | Exposed | Unexposed | Exposed | Unexposed | Exposed | Unexposed | Exposed |
| -2 | | 0.75 (0.74–0.75) | 0.71 (0.71–0.71) | 0.79 (0.79–0.79) | 0.76 (0.75–0.76) | 0.72 (0.72–0.73) | 0.69 (0.68–0.69) | 0.71 (0.7–0.71) | 0.66 (0.66–0.67) |
| -1 | | 0.73 (0.72–0.73) | 0.7 (0.7–0.7) | 0.79 (0.78–0.79) | 0.75 (0.74–0.75) | 0.71 (0.71–0.72) | 0.68 (0.67–0.68) | 0.7 (0.7–0.71) | 0.65 (0.65–0.66) |
| 0 | | 0.71 (0.71–0.72) | 0.68 (0.68–0.69) | 0.78 (0.77–0.78) | 0.74 (0.74–0.74) | 0.7 (0.69–0.7) | 0.66 (0.66–0.67) | 0.7 (0.69–0.7) | 0.64 (0.64–0.65) |
| 1 | | 0.69 (0.69–0.7) | 0.66 (0.66–0.66) | 0.77 (0.76–0.77) | 0.73 (0.72–0.73) | 0.69 (0.68–0.69) | 0.65 (0.64–0.65) | 0.69 (0.68–0.69) | 0.63 (0.63–0.64) |
| 2 | | 0.67 (0.67–0.67) | 0.64 (0.63–0.64) | 0.75 (0.75–0.76) | 0.71 (0.71–0.72) | 0.67 (0.67–0.68) | 0.63 (0.62–0.64) | 0.68 (0.67–0.68) | 0.62 (0.62–0.63) |
| 3 | | 0.64 (0.64–0.65) | 0.61 (0.61–0.62) | 0.74 (0.73–0.74) | 0.7 (0.69–0.7) | 0.65 (0.65–0.66) | 0.62 (0.61–0.62) | 0.66 (0.66–0.67) | 0.61 (0.6–0.61) |
| 4 | | 0.62 (0.61–0.62) | 0.58 (0.58–0.59) | 0.72 (0.71–0.72) | 0.68 (0.68–0.69) | 0.63 (0.63–0.64) | 0.59 (0.58–0.6) | 0.65 (0.65–0.66) | 0.59 (0.59–0.6) |
| 5 | | 0.58 (0.58–0.59) | 0.55 (0.55–0.56) | 0.69 (0.69–0.7) | 0.66 (0.65–0.66) | 0.61 (0.6–0.62) | 0.57 (0.56–0.58) | 0.63 (0.63–0.64) | 0.57 (0.57–0.58) |

Marg. pred. = Marginal predictions in probabilities

**eTable 42. Estimates for marginal predictions of annual earnings among parents exposed to child's eating disorder (F50) and unexposed parents stratified by the child's age at diagnosis**

| | | Finland | | | | Denmark | | | |
|---|---|---|---|---|---|---|---|---|---|
| | | Men | | Women | | Men | | Women | |
| | | Unexposed | Exposed | Unexposed | Exposed | Unexposed | Exposed | Unexposed | Exposed |
| Years since diagnosis | Age group | Marg. pred. (95% CI) | Marg. pred. (95% CI) | Marg. pred. (95% CI) | Marg. pred. (95% CI) | Marg. pred. (95% CI) | Marg. pred. (95% CI) | Marg. pred. (95% CI) | Marg. pred. (95% CI) |
| | | Unexposed | Exposed | Unexposed | Exposed | Unexposed | Exposed | Unexposed | Exposed |
| -5 | 5–9 years | 31.6 (29.6–33.6) | 32.3 (30.2–34.4) | 14 (12.7–15.4) | 12.9 (11.7–14.1) | 52.5 (48.2–56.8) | 55.9 (50.4–61.5) | 39.5 (36.8–42.3) | 42.1 (38.9–45.2) |
| -4 | | 32.3 (30.3–34.2) | 32.9 (30.9–35) | 16.5 (15.1–17.9) | 14.4 (13.1–15.7) | 55.7 (50.4–61) | 57.9 (52.6–63.2) | 41.1 (38.5–43.7) | 43.8 (40.5–47) |
| -3 | | 33.6 (31.5–35.7) | 33.8 (31.6–35.9) | 18.1 (16.7–19.5) | 16.1 (14.7–17.4) | 56.2 (50.8–61.5) | 59.5 (53.7–65.4) | 42.9 (39.9–45.8) | 45.7 (42.6–48.7) |
| -2 | | 34.9 (32.7–37.1) | 34.6 (32.3–36.8) | 19.2 (17.7–20.7) | 17.2 (15.9–18.5) | 55.9 (50.6–61.2) | 60.7 (54.9–66.5) | 44.3 (41.3–47.3) | 47.2 (43.9–50.4) |
| -1 | | 35.7 (33.5–37.9) | 35.4 (33.2–37.6) | 20 (18.6–21.5) | 18.3 (16.9–19.6) | 57.8 (52.2–63.3) | 64.3 (57.1–71.5) | 45.1 (42.1–48.2) | 47.5 (44–51) |
| 0 | | 37.3 (34.9–39.7) | 35.4 (33.1–37.7) | 21.4 (19.9–22.9) | 20 (18.6–21.5) | 60 (54.5–65.4) | 61.3 (54.8–67.8) | 45.9 (43.1–48.8) | 47.3 (43.8–50.9) |
| 1 | | 38.3 (35.9–40.8) | 36.4 (34–38.9) | 22.1 (20.6–23.6) | 20.9 (19.4–22.3) | 61.8 (55.2–68.3) | 60 (53.7–66.2) | 46 (43–49) | 47.9 (44.2–51.6) |
| 2 | | 39.4 (36.8–42) | 36.3 (33.8–38.8) | 23.6 (22–25.1) | 22 (20.4–23.5) | 64.1 (56.4–71.7) | 62.2 (55.7–68.6) | 45.8 (42.9–48.8) | 47.7 (44.1–51.3) |
| 3 | | 40.1 (37.5–42.7) | 37.4 (35–39.9) | 25.2 (23.6–26.8) | 22.7 (21.2–24.3) | 60.7 (54.4–67) | 62.2 (55.8–68.6) | 45.9 (42.9–48.8) | 49 (45.2–52.9) |

**eTable 42. Estimates for marginal predictions of annual earnings among parents exposed to child's eating disorder (F50) and unexposed parents stratified by the child's age at diagnosis**

| | | Finland | | | | Denmark | | | |
|---|---|---|---|---|---|---|---|---|---|
| | | Men | | Women | | Men | | Women | |
| | | Unexposed | Exposed | Unexposed | Exposed | Unexposed | Exposed | Unexposed | Exposed |
| Years since diagnosis | Age group | Marg. pred. (95% CI) | Marg. pred. (95% CI) | Marg. pred. (95% CI) | Marg. pred. (95% CI) | Marg. pred. (95% CI) | Marg. pred. (95% CI) | Marg. pred. (95% CI) | Marg. pred. (95% CI) |
| | | Unexposed | Exposed | Unexposed | Exposed | Unexposed | Exposed | Unexposed | Exposed |
| 4 | | 42.2 (39.4–45) | 38.9 (36.2–41.6) | 25.9 (24.2–27.6) | 24.2 (22.6–25.9) | 63 (55.9–70.2) | 66.2 (59.3–73) | 43.9 (40.7–47) | 48.6 (44.7–52.4) |
| 5 | | 42.3 (39.4–45.2) | 39.6 (36.9–42.4) | 27.1 (25.3–28.9) | 25 (23.3–26.8) | 63.8 (55.2–72.5) | 64.3 (56.9–71.7) | 45.7 (42.2–49.3) | 50.5 (46.2–54.8) |
| -5 | 10–14 years | 38.9 (38.2–39.7) | 39 (38.3–39.8) | 22.3 (21.8–22.8) | 22.9 (22.5–23.4) | 60.9 (59.7–62.1) | 61.2 (60.1–62.3) | 44.9 (44.4–45.5) | 45.4 (44.8–45.9) |
| -4 | | 39.8 (39–40.5) | 39.9 (39.1–40.7) | 23.2 (22.8–23.7) | 24.1 (23.6–24.6) | 61.8 (60.6–63) | 62.3 (61.2–63.5) | 45.8 (45.3–46.4) | 46.3 (45.7–46.8) |
| -3 | | 40.4 (39.6–41.2) | 40.5 (39.7–41.3) | 24.6 (24.1–25.1) | 25.2 (24.7–25.7) | 63.2 (61.9–64.4) | 63.8 (62.6–65) | 46.6 (46–47.2) | 47.5 (46.9–48.1) |
| -2 | | 41.1 (40.3–41.9) | 41.2 (40.3–42) | 25.6 (25.1–26.1) | 26.3 (25.8–26.8) | 64.4 (63.1–65.7) | 64.4 (63.1–65.6) | 47.6 (47–48.2) | 48.4 (47.8–49) |
| -1 | | 41.6 (40.8–42.5) | 41.4 (40.6–42.3) | 26.6 (26.1–27.2) | 27.2 (26.7–27.7) | 65.6 (64.3–66.9) | 65.3 (64.1–66.5) | 48.3 (47.7–48.9) | 49.3 (48.6–49.9) |
| 0 | | 42 (41.2–42.9) | 41.7 (40.8–42.5) | 27.6 (27–28.1) | 28 (27.4–28.5) | 66.8 (65.4–68.1) | 66.5 (65.2–67.8) | 49.2 (48.6–49.8) | 49.7 (49.1–50.3) |
| 1 | | 42.3 (41.5–43.2) | 42.2 (41.3–43.1) | 28.3 (27.7–28.8) | 28.5 (28–29.1) | 67.2 (65.9–68.6) | 68.1 (66.8–69.4) | 49.9 (49.2–50.5) | 50.5 (49.9–51.2) |

**eTable 42. Estimates for marginal predictions of annual earnings among parents exposed to child's eating disorder (F50) and unexposed parents stratified by the child's age at diagnosis**

| | | Finland | | | | Denmark | | | |
|---|---|---|---|---|---|---|---|---|---|
| | | Men | | Women | | Men | | Women | |
| | | Unexposed | Exposed | Unexposed | Exposed | Unexposed | Exposed | Unexposed | Exposed |
| Years since diagnosis | Age group | Marg. pred. (95% CI) | Marg. pred. (95% CI) | Marg. pred. (95% CI) | Marg. pred. (95% CI) | Marg. pred. (95% CI) | Marg. pred. (95% CI) | Marg. pred. (95% CI) | Marg. pred. (95% CI) |
| | | Unexposed | Exposed | Unexposed | Exposed | Unexposed | Exposed | Unexposed | Exposed |
| 2 | | 42.7 (41.8–43.6) | 42.3 (41.4–43.2) | 29.1 (28.5–29.6) | 29.3 (28.8–29.9) | 67.9 (66.5–69.4) | 69.3 (67.9–70.7) | 50.6 (49.9–51.2) | 50.7 (50.1–51.4) |
| 3 | | 43 (42.1–43.9) | 42.3 (41.4–43.3) | 30 (29.5–30.6) | 30 (29.4–30.6) | 68.6 (67.1–70.1) | 69.9 (68.4–71.4) | 51 (50.3–51.7) | 50.8 (50.1–51.5) |
| 4 | | 43.4 (42.4–44.3) | 42.5 (41.6–43.5) | 30.7 (30.1–31.3) | 30.8 (30.2–31.4) | 69.1 (67.6–70.6) | 69.8 (68.2–71.3) | 51.2 (50.5–51.9) | 50.9 (50.2–51.6) |
| 5 | | 43.1 (42.1–44.1) | 42.3 (41.3–43.3) | 31.2 (30.7–31.8) | 31.2 (30.6–31.8) | 70.2 (68.5–71.9) | 71 (69.3–72.6) | 50.9 (50.2–51.7) | 50.9 (50.1–51.6) |
| -5 | 15–19 years | 38.8 (38.2–39.4) | 39.6 (38.9–40.2) | 24.7 (24.4–25.1) | 25.7 (25.3–26.1) | 60.4 (59.6–61.3) | 62.8 (61.9–63.6) | 45.4 (45–45.8) | 45.5 (45.1–45.9) |
| -4 | | 39.2 (38.6–39.8) | 40.3 (39.6–40.9) | 25.6 (25.2–25.9) | 26.7 (26.3–27) | 61 (60.2–61.9) | 63.4 (62.5–64.3) | 46.1 (45.7–46.5) | 46.2 (45.8–46.6) |
| -3 | | 39.7 (39.1–40.3) | 41.1 (40.4–41.7) | 26.5 (26.1–26.9) | 27.6 (27.3–28) | 62 (61.1–62.8) | 64.2 (63.2–65.1) | 46.6 (46.2–47) | 46.8 (46.4–47.3) |
| -2 | | 40.1 (39.5–40.7) | 41.4 (40.7–42) | 27.4 (27–27.7) | 28.6 (28.2–29) | 62.3 (61.4–63.2) | 64.9 (64–65.9) | 47 (46.6–47.5) | 47.4 (47–47.8) |
| -1 | | 40.4 (39.8–41) | 41.6 (40.9–42.2) | 28 (27.6–28.3) | 29.2 (28.8–29.6) | 63.2 (62.2–64.1) | 65.6 (64.6–66.5) | 47.5 (47–47.9) | 47.8 (47.4–48.2) |
| 0 | | 40.5 (39.9–41.2) | 41.6 (40.9–42.2) | 28.6 (28.2–29) | 29.8 (29.4–30.2) | 64.3 (63.3–65.3) | 66 (65–67) | 47.8 (47.3–48.2) | 47.9 (47.4–48.3) |

**eTable 42. Estimates for marginal predictions of annual earnings among parents exposed to child's eating disorder (F50) and unexposed parents stratified by the child's age at diagnosis**

| | | Finland | | | | Denmark | | | |
|---|---|---|---|---|---|---|---|---|---|
| | | Men | | Women | | Men | | Women | |
| | | Unexposed | Exposed | Unexposed | Exposed | Unexposed | Exposed | Unexposed | Exposed |
| Years since diagnosis | Age group | Marg. pred. (95% CI) | Marg. pred. (95% CI) | Marg. pred. (95% CI) | Marg. pred. (95% CI) | Marg. pred. (95% CI) | Marg. pred. (95% CI) | Marg. pred. (95% CI) | Marg. pred. (95% CI) |
| | | Unexposed | Exposed | Unexposed | Exposed | Unexposed | Exposed | Unexposed | Exposed |
| 1 | | 40.7 (40–41.4) | 41.7 (41–42.4) | 29 (28.6–29.4) | 30.4 (30–30.8) | 64.7 (63.7–65.7) | 66.4 (65.4–67.4) | 47.9 (47.4–48.3) | 48.1 (47.6–48.5) |
| 2 | | 40.8 (40.1–41.4) | 41.3 (40.6–42) | 29.6 (29.2–30) | 30.9 (30.4–31.3) | 64.8 (63.8–65.8) | 66.7 (65.6–67.7) | 47.9 (47.5–48.4) | 48.2 (47.7–48.6) |
| 3 | | 40.4 (39.7–41.1) | 41.1 (40.4–41.8) | 30 (29.6–30.4) | 31 (30.6–31.5) | 65.1 (64.1–66.1) | 66.8 (65.8–67.9) | 47.9 (47.5–48.4) | 48.2 (47.7–48.6) |
| 4 | | 40.1 (39.4–40.8) | 40.7 (39.9–41.4) | 30.2 (29.7–30.6) | 31.3 (30.9–31.8) | 65.2 (64.1–66.2) | 67.6 (66.5–68.7) | 48.3 (47.8–48.8) | 48.3 (47.8–48.8) |
| 5 | | 39.3 (38.5–40) | 40 (39.3–40.8) | 29.9 (29.5–30.4) | 31.4 (30.9–31.9) | 65.4 (64.3–66.5) | 66.5 (65.4–67.6) | 48.4 (47.9–49) | 48.5 (48–49) |
| -5 | 20–25 years | 37.3 (36.4–38.2) | 38.1 (37.1–39) | 25.7 (25.1–26.2) | 26 (25.4–26.5) | 60.5 (59.5–61.5) | 61.5 (60.5–62.5) | 44.1 (43.6–44.6) | 44.5 (44–45) |
| -4 | | 37.1 (36.2–38) | 38.2 (37.2–39.1) | 26.1 (25.6–26.6) | 26.4 (25.9–27) | 60.2 (59.2–61.2) | 62 (60.9–63) | 44.1 (43.6–44.5) | 44.7 (44.2–45.2) |
| -3 | | 36.9 (36–37.9) | 38.3 (37.3–39.3) | 26.5 (26–27.1) | 26.7 (26.1–27.3) | 60.3 (59.3–61.3) | 62.6 (61.5–63.6) | 44 (43.5–44.4) | 44.7 (44.2–45.2) |
| -2 | | 37 (36.1–38) | 37.9 (36.9–38.9) | 26.8 (26.2–27.3) | 26.9 (26.3–27.5) | 60.9 (59.9–62) | 62.6 (61.5–63.7) | 44 (43.5–44.4) | 44.8 (44.4–45.3) |

**eTable 42. Estimates for marginal predictions of annual earnings among parents exposed to child's eating disorder (F50) and unexposed parents stratified by the child’s age at diagnosis**

| | | Finland | | | | Denmark | | | |
|---|---|---|---|---|---|---|---|---|---|
| | | Men | | Women | | Men | | Women | |
| | | Unexposed | Exposed | Unexposed | Exposed | Unexposed | Exposed | Unexposed | Exposed |
| Years since diagnosis | Age group | Marg. pred. (95% CI) | Marg. pred. (95% CI) | Marg. pred. (95% CI) | Marg. pred. (95% CI) | Marg. pred. (95% CI) | Marg. pred. (95% CI) | Marg. pred. (95% CI) | Marg. pred. (95% CI) |
| | | Unexposed | Exposed | Unexposed | Exposed | Unexposed | Exposed | Unexposed | Exposed |
| -1 | | 36.5 (35.5–37.4) | 37.2 (36.3–38.2) | 27 (26.4–27.6) | 27.1 (26.5–27.7) | 60.9 (59.9–61.9) | 62.5 (61.5–63.6) | 44.2 (43.7–44.7) | 45.1 (44.6–45.6) |
| 0 | | 36.3 (35.3–37.3) | 36.8 (35.8–37.8) | 26.8 (26.2–27.4) | 27.1 (26.5–27.7) | 60.9 (59.8–61.9) | 63 (61.9–64.2) | 44.3 (43.8–44.8) | 45.3 (44.8–45.8) |
| 1 | | 35.8 (34.8–36.7) | 36.1 (35–37.1) | 26.6 (26–27.2) | 27 (26.4–27.6) | 61.5 (60.4–62.6) | 63 (61.9–64.2) | 44.4 (43.9–44.9) | 45.3 (44.8–45.8) |
| 2 | | 34.6 (33.6–35.6) | 35.2 (34.2–36.3) | 26.5 (25.9–27.1) | 26.8 (26.2–27.4) | 60.9 (59.8–62) | 63 (61.8–64.2) | 44.7 (44.2–45.3) | 45.5 (45–46) |
| 3 | | 33.6 (32.6–34.7) | 34.1 (33–35.2) | 26.4 (25.8–27) | 26.3 (25.6–26.9) | 60.8 (59.7–62) | 62.3 (61.1–63.5) | 44.8 (44.3–45.3) | 45.7 (45.1–46.2) |
| 4 | | 32.4 (31.4–33.5) | 33 (31.9–34.1) | 25.6 (25–26.2) | 26 (25.4–26.7) | 60.7 (59.5–61.9) | 61.9 (60.7–63.1) | 45 (44.4–45.5) | 45.6 (45.1–46.2) |
| 5 | | 30.8 (29.7–31.9) | 31.3 (30.1–32.4) | 25.2 (24.5–25.9) | 25.2 (24.6–25.9) | 59.6 (58.4–60.8) | 61.2 (59.9–62.4) | 44.7 (44.2–45.3) | 45.9 (45.3–46.5) |

Marg. pred. = Marginal predictions in thousand euros (€)

**eTable 43. Estimates for marginal predictions of annual disposable income among parents exposed to child's eating disorder (F50) and unexposed parents stratified by the child's age at diagnosis**

| | | Finland | | | | Denmark | | | |
|---|---|---|---|---|---|---|---|---|---|
| | | Men | | Women | | Men | | Women | |
| | | Unexposed | Exposed | Unexposed | Exposed | Unexposed | Exposed | Unexposed | Exposed |
| Years since diagnosis | Age group | Marg. pred. (95% CI) | Marg. pred. (95% CI) | Marg. pred. (95% CI) | Marg. pred. (95% CI) | Marg. pred. (95% CI) | Marg. pred. (95% CI) | Marg. pred. (95% CI) | Marg. pred. (95% CI) |
| | | Unexposed | Exposed | Unexposed | Exposed | Unexposed | Exposed | Unexposed | Exposed |
| -5 | 5–9 years | 25.4 (24.2–26.7) | 26 (24.7–27.2) | 19.6 (18.8–20.4) | 18.8 (18–19.6) | 31 (28.7–33.3) | 33.5 (30.5–36.6) | 27.4 (25.7–29.1) | 28.8 (27–30.5) |
| -4 | | 26.6 (25.3–27.9) | 26.4 (25.1–27.7) | 21.2 (20.3–22) | 19.9 (19.1–20.7) | 33 (30.4–35.6) | 35 (32.1–37.9) | 28.7 (27.2–30.2) | 30 (28–31.9) |
| -3 | | 27.2 (25.9–28.5) | 27.1 (25.9–28.4) | 22.1 (21.2–22.9) | 20.8 (20–21.6) | 33.7 (30.8–36.5) | 36.6 (33.2–40) | 29.8 (28–31.7) | 31.8 (29.9–33.7) |
| -2 | | 28.1 (26.7–29.6) | 27.8 (26.5–29.1) | 23.2 (22.2–24.1) | 21.6 (20.8–22.4) | 34.1 (31.4–36.9) | 37.5 (34.1–40.9) | 30.7 (28.9–32.6) | 32.6 (30.7–34.5) |
| -1 | | 28.3 (26.9–29.6) | 28.5 (27.1–29.8) | 23.7 (22.8–24.6) | 22.5 (21.7–23.3) | 36.1 (32.9–39.4) | 39.8 (35.7–43.9) | 31.8 (29.9–33.7) | 33.3 (31.2–35.3) |
| 0 | | 29.9 (28.3–31.5) | 28.9 (27.5–30.3) | 24.6 (23.7–25.5) | 23.5 (22.6–24.3) | 37.8 (34.5–41.1) | 38.4 (34.7–42.1) | 32.2 (30.4–34) | 33.2 (31–35.4) |
| 1 | | 30.8 (29.2–32.4) | 29.6 (28.1–31.2) | 24.8 (24–25.7) | 24 (23.2–24.8) | 38.2 (34.4–41.9) | 38.4 (34.6–42.2) | 32.4 (30.6–34.3) | 33.7 (31.5–35.9) |
| 2 | | 31.5 (29.9–33.2) | 30.5 (28.8–32.2) | 25.9 (24.9–26.8) | 24.5 (23.7–25.4) | 40.1 (35.7–44.6) | 40.2 (36.3–44.1) | 32.6 (30.6–34.5) | 33.6 (31.4–35.8) |
| 3 | | 32.3 (30.6–34) | 31.3 (29.6–33) | 26.5 (25.6–27.5) | 25.3 (24.4–26.2) | 38.9 (35.2–42.6) | 40.1 (36.7–43.6) | 32.9 (31–34.9) | 34.9 (32.4–37.4) |

**eTable 43. Estimates for marginal predictions of annual disposable income among parents exposed to child's eating disorder (F50) and unexposed parents stratified by the child's age at diagnosis**

| | | Finland | | | | Denmark | | | |
|---|---|---|---|---|---|---|---|---|---|
| | | Men | | Women | | Men | | Women | |
| | | Unexposed | Exposed | Unexposed | Exposed | Unexposed | Exposed | Unexposed | Exposed |
| Years since diagnosis | Age group | Marg. pred. (95% CI) | Marg. pred. (95% CI) | Marg. pred. (95% CI) | Marg. pred. (95% CI) | Marg. pred. (95% CI) | Marg. pred. (95% CI) | Marg. pred. (95% CI) | Marg. pred. (95% CI) |
| | | Unexposed | Exposed | Unexposed | Exposed | Unexposed | Exposed | Unexposed | Exposed |
| 4 | | 33.8 (31.9–35.6) | 32.3 (30.5–34.2) | 27.2 (26.2–28.2) | 25.9 (25–26.8) | 40.1 (36.1–44.1) | 42.8 (39–46.6) | 32 (29.8–34.1) | 34.6 (32.2–37.1) |
| 5 | | 34.3 (32.3–36.3) | 32.1 (30.3–33.9) | 27.8 (26.8–28.8) | 26.5 (25.6–27.5) | 41.1 (36.1–46.1) | 42.2 (37.7–46.6) | 33 (30.7–35.3) | 36.2 (33.5–38.8) |
| -5 | 10–14 years | 30.9 (30.3–31.4) | 31.1 (30.5–31.6) | 24.1 (23.8–24.3) | 24.2 (23.9–24.5) | 36.2 (35.6–36.8) | 36.3 (35.7–36.9) | 30.9 (30.5–31.2) | 31.1 (30.8–31.5) |
| -4 | | 31.5 (31–32) | 31.8 (31.3–32.4) | 24.6 (24.3–24.9) | 25 (24.7–25.3) | 37.2 (36.6–37.9) | 37.4 (36.8–38) | 31.6 (31.3–32) | 31.9 (31.6–32.2) |
| -3 | | 32.1 (31.5–32.6) | 32.2 (31.6–32.7) | 25.5 (25.2–25.8) | 25.7 (25.4–26) | 38.4 (37.7–39) | 38.7 (38–39.3) | 32.4 (32–32.7) | 32.9 (32.5–33.2) |
| -2 | | 32.7 (32.2–33.3) | 32.9 (32.4–33.5) | 26.1 (25.9–26.4) | 26.4 (26.1–26.7) | 39.5 (38.8–40.2) | 39.3 (38.7–40) | 33.1 (32.8–33.5) | 33.6 (33.2–33.9) |
| -1 | | 33.3 (32.7–33.9) | 33.3 (32.7–33.9) | 26.7 (26.4–27) | 27.1 (26.8–27.4) | 40.5 (39.8–41.3) | 40.4 (39.7–41.1) | 33.7 (33.4–34.1) | 34.3 (33.9–34.7) |
| 0 | | 33.8 (33.2–34.4) | 33.7 (33.1–34.3) | 27.2 (26.9–27.5) | 27.5 (27.2–27.8) | 41.7 (40.9–42.4) | 41.3 (40.6–42) | 34.5 (34.2–34.9) | 34.9 (34.5–35.3) |
| 1 | | 34.4 (33.8–35) | 34.5 (33.9–35.2) | 27.5 (27.2–27.8) | 27.9 (27.5–28.2) | 42.4 (41.6–43.2) | 42.7 (42–43.5) | 35.2 (34.8–35.6) | 35.6 (35.2–36) |

**eTable 43. Estimates for marginal predictions of annual disposable income among parents exposed to child's eating disorder (F50) and unexposed parents stratified by the child’s age at diagnosis**

| | | Finland | | | | Denmark | | | |
|---|---|---|---|---|---|---|---|---|---|
| | | Men | | Women | | Men | | Women | |
| | | Unexposed | Exposed | Unexposed | Exposed | Unexposed | Exposed | Unexposed | Exposed |
| Years since diagnosis | Age group | Marg. pred. (95% CI) | Marg. pred. (95% CI) | Marg. pred. (95% CI) | Marg. pred. (95% CI) | Marg. pred. (95% CI) | Marg. pred. (95% CI) | Marg. pred. (95% CI) | Marg. pred. (95% CI) |
| | | Unexposed | Exposed | Unexposed | Exposed | Unexposed | Exposed | Unexposed | Exposed |
| 2 | | 34.7 (34.1–35.4) | 34.7 (34–35.3) | 27.8 (27.5–28.2) | 28.2 (27.8–28.5) | 43.2 (42.4–44) | 43.9 (43.1–44.7) | 35.7 (35.3–36.1) | 35.9 (35.5–36.3) |
| 3 | | 35.1 (34.5–35.8) | 35 (34.3–35.6) | 27.9 (27.5–28.2) | 28.1 (27.8–28.4) | 44.1 (43.2–44.9) | 44.7 (43.8–45.6) | 36.2 (35.8–36.6) | 36.1 (35.7–36.5) |
| 4 | | 35.5 (34.9–36.2) | 35.4 (34.7–36.1) | 27.8 (27.5–28.2) | 28.1 (27.8–28.5) | 44.7 (43.9–45.6) | 45.2 (44.3–46.1) | 36.4 (35.9–36.8) | 36.1 (35.7–36.6) |
| 5 | | 35.7 (35–36.4) | 35.7 (35–36.4) | 27.9 (27.5–28.2) | 28.2 (27.8–28.5) | 46.1 (45.2–47.1) | 46.6 (45.6–47.5) | 36.2 (35.8–36.7) | 36.2 (35.8–36.7) |
| -5 | 15–19 years | 31 (30.6–31.4) | 31.4 (31–31.8) | 24.9 (24.7–25.1) | 25.5 (25.3–25.7) | 36.1 (35.6–36.5) | 37.2 (36.7–37.7) | 30.8 (30.6–31.1) | 30.9 (30.7–31.1) |
| -4 | | 31.4 (31–31.8) | 32.1 (31.7–32.6) | 25.4 (25.2–25.6) | 26 (25.8–26.2) | 36.9 (36.4–37.3) | 38 (37.5–38.5) | 31.5 (31.2–31.7) | 31.5 (31.3–31.8) |
| -3 | | 31.9 (31.5–32.3) | 32.8 (32.3–33.2) | 25.9 (25.6–26.1) | 26.5 (26.3–26.7) | 37.9 (37.4–38.4) | 38.9 (38.4–39.5) | 32 (31.8–32.3) | 32.2 (31.9–32.4) |
| -2 | | 32.5 (32.1–32.9) | 33.3 (32.9–33.7) | 26.1 (25.9–26.4) | 26.9 (26.7–27.1) | 38.4 (38–38.9) | 39.8 (39.3–40.3) | 32.5 (32.2–32.7) | 32.7 (32.4–32.9) |
| -1 | | 33.1 (32.6–33.5) | 34 (33.5–34.4) | 26.3 (26–26.5) | 27.1 (26.8–27.3) | 39.3 (38.8–39.9) | 40.6 (40.1–41.2) | 32.8 (32.6–33.1) | 33 (32.7–33.3) |

**eTable 43. Estimates for marginal predictions of annual disposable income among parents exposed to child's eating disorder (F50) and unexposed parents stratified by the child's age at diagnosis**

| | | Finland | | | | Denmark | | | |
|---|---|---|---|---|---|---|---|---|---|
| | | Men | | Women | | Men | | Women | |
| | | Unexposed | Exposed | Unexposed | Exposed | Unexposed | Exposed | Unexposed | Exposed |
| Years since diagnosis | Age group | Marg. pred. (95% CI) | Marg. pred. (95% CI) | Marg. pred. (95% CI) | Marg. pred. (95% CI) | Marg. pred. (95% CI) | Marg. pred. (95% CI) | Marg. pred. (95% CI) | Marg. pred. (95% CI) |
| | | Unexposed | Exposed | Unexposed | Exposed | Unexposed | Exposed | Unexposed | Exposed |
| 0 | | 33.4 (32.9–33.8) | 34.2 (33.7–34.6) | 26.3 (26.1–26.5) | 27.2 (26.9–27.4) | 40.5 (39.9–41) | 41.2 (40.7–41.8) | 33.1 (32.9–33.4) | 33.1 (32.8–33.4) |
| 1 | | 33.8 (33.4–34.3) | 34.7 (34.2–35.2) | 26.2 (25.9–26.4) | 27.1 (26.9–27.4) | 41.1 (40.6–41.7) | 42 (41.5–42.6) | 33.3 (33–33.6) | 33.3 (33–33.6) |
| 2 | | 34.1 (33.7–34.6) | 35.1 (34.6–35.6) | 26.2 (26–26.5) | 27.1 (26.9–27.4) | 41.7 (41.1–42.3) | 42.6 (42.1–43.2) | 33.4 (33.1–33.7) | 33.4 (33.1–33.7) |
| 3 | | 34.3 (33.8–34.7) | 35.4 (34.9–35.9) | 26.3 (26.1–26.6) | 27.2 (27–27.5) | 42.4 (41.8–43) | 43.2 (42.6–43.8) | 33.6 (33.3–33.9) | 33.5 (33.3–33.8) |
| 4 | | 34.5 (34–35) | 35.6 (35–36.1) | 26.5 (26.3–26.8) | 27.4 (27.1–27.7) | 43 (42.4–43.6) | 44.3 (43.6–44.9) | 33.9 (33.6–34.2) | 33.7 (33.4–34) |
| 5 | | 34.5 (34–35) | 35.6 (35–36.1) | 26.5 (26.2–26.7) | 27.5 (27.3–27.8) | 43.5 (42.9–44.2) | 44.2 (43.5–44.8) | 34.2 (33.9–34.6) | 34.1 (33.8–34.5) |
| -5 | 20–25 years | 30.5 (29.9–31.1) | 30.8 (30.2–31.5) | 23.8 (23.5–24.1) | 24 (23.7–24.3) | 36.4 (35.9–37) | 36.7 (36.1–37.2) | 29.5 (29.2–29.8) | 29.8 (29.5–30.1) |
| -4 | | 30.5 (29.9–31.1) | 31.3 (30.6–31.9) | 23.9 (23.5–24.2) | 24 (23.6–24.3) | 36.8 (36.3–37.4) | 37.5 (37–38.1) | 29.6 (29.4–29.9) | 30 (29.7–30.2) |
| -3 | | 30.5 (29.9–31.1) | 31.6 (30.9–32.2) | 23.8 (23.5–24.1) | 23.9 (23.6–24.2) | 37.5 (37–38.1) | 38.4 (37.9–39) | 29.6 (29.3–29.9) | 30.1 (29.8–30.4) |

**eTable 43. Estimates for marginal predictions of annual disposable income among parents exposed to child's eating disorder (F50) and unexposed parents stratified by the child's age at diagnosis**

| | | Finland | | | | Denmark | | | |
|---|---|---|---|---|---|---|---|---|---|
| | | Men | | Women | | Men | | Women | |
| | | Unexposed | Exposed | Unexposed | Exposed | Unexposed | Exposed | Unexposed | Exposed |
| Years since diagnosis | Age group | Marg. pred. (95% CI) | Marg. pred. (95% CI) | Marg. pred. (95% CI) | Marg. pred. (95% CI) | Marg. pred. (95% CI) | Marg. pred. (95% CI) | Marg. pred. (95% CI) | Marg. pred. (95% CI) |
| | | Unexposed | Exposed | Unexposed | Exposed | Unexposed | Exposed | Unexposed | Exposed |
| -2 | | 31.2 (30.5–31.8) | 31.8 (31.2–32.5) | 23.9 (23.5–24.2) | 23.9 (23.6–24.3) | 38.2 (37.7–38.8) | 38.8 (38.2–39.4) | 29.7 (29.5–30) | 30.2 (29.9–30.5) |
| -1 | | 31.3 (30.7–32) | 32 (31.3–32.7) | 24 (23.7–24.4) | 24.3 (23.9–24.6) | 38.8 (38.2–39.4) | 39.3 (38.7–39.9) | 30 (29.7–30.3) | 30.5 (30.2–30.8) |
| 0 | | 31.7 (31–32.4) | 32.2 (31.5–32.9) | 24.1 (23.8–24.4) | 24.3 (23.9–24.6) | 39 (38.4–39.6) | 40 (39.3–40.6) | 30.3 (30–30.6) | 30.9 (30.6–31.2) |
| 1 | | 31.8 (31.2–32.5) | 32.3 (31.6–33) | 24.2 (23.9–24.6) | 24.6 (24.2–24.9) | 39.9 (39.3–40.5) | 40.4 (39.8–41.1) | 30.6 (30.2–30.9) | 31.1 (30.8–31.4) |
| 2 | | 31.8 (31.1–32.4) | 32.5 (31.7–33.2) | 24.4 (24.1–24.8) | 24.7 (24.3–25) | 40.1 (39.4–40.7) | 40.9 (40.2–41.5) | 31 (30.6–31.3) | 31.5 (31.1–31.8) |
| 3 | | 32 (31.3–32.7) | 32.4 (31.7–33.1) | 24.6 (24.2–24.9) | 24.7 (24.4–25.1) | 40.5 (39.8–41.1) | 41 (40.3–41.7) | 31.2 (30.9–31.6) | 31.8 (31.5–32.2) |
| 4 | | 32.1 (31.4–32.9) | 32.6 (31.8–33.3) | 24.5 (24.1–24.8) | 25 (24.6–25.4) | 40.8 (40.1–41.5) | 41.3 (40.6–42) | 31.6 (31.3–32) | 32 (31.7–32.4) |
| 5 | | 31.9 (31.1–32.6) | 32.6 (31.9–33.4) | 24.6 (24.2–25) | 24.9 (24.5–25.3) | 40.6 (39.9–41.3) | 41.5 (40.7–42.2) | 31.8 (31.4–32.1) | 32.4 (32.1–32.8) |

Marg. pred. = Marginal predictions in thousand euros (€)

**eTable 44. Estimates for marginal predictions of annual social welfare benefits among parents exposed to child's eating disorder (F50) and unexposed parents stratified by the child's age at diagnosis**

| | | Finland | | | | Denmark | | | |
|---|---|---|---|---|---|---|---|---|---|
| | | Men | | Women | | Men | | Women | |
| | | Unexposed | Exposed | Unexposed | Exposed | Unexposed | Exposed | Unexposed | Exposed |
| Years since diagnosis | Age group | Marg. pred. (95% CI) | Marg. pred. (95% CI) | Marg. pred. (95% CI) | Marg. pred. (95% CI) | Marg. pred. (95% CI) | Marg. pred. (95% CI) | Marg. pred. (95% CI) | Marg. pred. (95% CI) |
| | | Unexposed | Exposed | Unexposed | Exposed | Unexposed | Exposed | Unexposed | Exposed |
| -5 | 5–9 years | 2.6 (2.1–3) | 2.8 (2.2–3.3) | 9.8 (9.2–10.4) | 9.7 (9.1–10.3) | 2.4 (1.4–3.4) | 3 (1.9–4.1) | 9.8 (8.5–11.2) | 11 (9.5–12.6) |
| -4 | | 2.7 (2.2–3.2) | 2.7 (2.2–3.3) | 9.1 (8.5–9.7) | 9.6 (9–10.3) | 2.5 (1.5–3.5) | 3.1 (1.9–4.2) | 10.1 (8.5–11.6) | 10.3 (8.9–11.8) |
| -3 | | 2.6 (2.1–3.1) | 2.9 (2.3–3.4) | 8.8 (8.1–9.4) | 9.2 (8.5–9.8) | 2.7 (1.7–3.7) | 3.2 (1.9–4.4) | 9.3 (7.9–10.8) | 10 (8.6–11.5) |
| -2 | | 2.5 (2–3) | 2.9 (2.4–3.5) | 8.9 (8.3–9.5) | 9 (8.4–9.6) | 3.2 (2.1–4.3) | 3.6 (2.3–4.9) | 9.3 (7.9–10.8) | 10 (8.5–11.5) |
| -1 | | 2.5 (2–3) | 2.9 (2.3–3.4) | 8.8 (8.2–9.5) | 8.9 (8.2–9.5) | 3.1 (2–4.2) | 4.1 (2.7–5.4) | 9.4 (7.9–10.9) | 9.2 (7.8–10.6) |
| 0 | | 2.7 (2.2–3.2) | 3.4 (2.8–4) | 8.5 (7.9–9.2) | 8.5 (7.9–9.1) | 3.4 (2.3–4.6) | 3.9 (2.7–5.2) | 8.8 (7.3–10.3) | 10.1 (8.5–11.7) |
| 1 | | 2.8 (2.2–3.3) | 3.5 (2.9–4.2) | 8.2 (7.5–8.9) | 8.5 (7.9–9.2) | 3.2 (2–4.4) | 3.9 (2.7–5.2) | 8.1 (6.7–9.5) | 11.3 (9.5–13.1) |
| 2 | | 3 (2.5–3.6) | 3.8 (3.1–4.5) | 8 (7.3–8.6) | 8.2 (7.6–8.9) | 3.5 (2.2–4.8) | 3.9 (2.5–5.2) | 7.8 (6.3–9.3) | 10.5 (8.8–12.2) |
| 3 | | 3.1 (2.5–3.7) | 3.9 (3.2–4.6) | 7.7 (7–8.3) | 8.2 (7.5–8.8) | 3.3 (2–4.6) | 4.2 (2.8–5.5) | 8.6 (7–10.2) | 10.4 (8.6–12.3) |
| 4 | | 3.3 (2.6–4) | 3.6 (2.9–4.3) | 7.7 (7–8.4) | 7.7 (7.1–8.3) | 3.3 (2–4.6) | 4.3 (2.7–5.9) | 8.2 (6.6–9.8) | 10.1 (8.3–11.9) |
| 5 | | 3.4 (2.6–4.1) | 3.5 (2.8–4.2) | 7.6 (6.8–8.3) | 7.8 (7.1–8.5) | 3.6 (2.2–5) | 4.3 (2.7–5.8) | 8.5 (6.8–10.1) | 10.2 (8.3–12.1) |
| -5 | 10–14 years | 2.5 (2.3–2.6) | 2.6 (2.4–2.7) | 7.8 (7.6–7.9) | 7.7 (7.5–7.9) | 2.8 (2.6–3) | 2.5 (2.3–2.7) | 9 (8.7–9.3) | 9 (8.7–9.2) |
| -4 | | 2.6 (2.5–2.8) | 2.7 (2.5–2.9) | 7.7 (7.5–7.8) | 7.5 (7.3–7.7) | 2.9 (2.7–3.1) | 2.6 (2.4–2.8) | 8.8 (8.5–9.1) | 8.8 (8.5–9.1) |

**eTable 44. Estimates for marginal predictions of annual social welfare benefits among parents exposed to child's eating disorder (F50) and unexposed parents stratified by the child's age at diagnosis**

| | | Finland | | | | Denmark | | | |
|---|---|---|---|---|---|---|---|---|---|
| | | Men | | Women | | Men | | Women | |
| | | Unexposed | Exposed | Unexposed | Exposed | Unexposed | Exposed | Unexposed | Exposed |
| Years since diagnosis | Age group | Marg. pred. (95% CI) | Marg. pred. (95% CI) | Marg. pred. (95% CI) | Marg. pred. (95% CI) | Marg. pred. (95% CI) | Marg. pred. (95% CI) | Marg. pred. (95% CI) | Marg. pred. (95% CI) |
| | | Unexposed | Exposed | Unexposed | Exposed | Unexposed | Exposed | Unexposed | Exposed |
| -3 | | 2.8 (2.6–3) | 2.8 (2.6–2.9) | 7.5 (7.3–7.7) | 7.4 (7.2–7.6) | 3 (2.8–3.2) | 2.6 (2.4–2.8) | 8.7 (8.4–9) | 8.4 (8.2–8.7) |
| -2 | | 2.9 (2.7–3.1) | 3 (2.8–3.2) | 7.3 (7.1–7.5) | 7.3 (7.1–7.4) | 3.1 (2.9–3.3) | 2.6 (2.4–2.8) | 8.5 (8.2–8.8) | 8.3 (8–8.6) |
| -1 | | 3 (2.8–3.2) | 3.2 (3–3.4) | 7.1 (6.9–7.3) | 7.2 (7–7.4) | 3.2 (3–3.4) | 2.8 (2.6–3) | 8.3 (8–8.5) | 8.1 (7.8–8.4) |
| 0 | | 3.2 (3–3.5) | 3.4 (3.2–3.6) | 6.8 (6.6–7) | 7 (6.8–7.2) | 3.3 (3–3.5) | 2.9 (2.7–3.1) | 8.2 (7.9–8.5) | 8.2 (7.9–8.5) |
| 1 | | 3.4 (3.2–3.6) | 3.6 (3.4–3.8) | 6.6 (6.4–6.8) | 6.9 (6.7–7.1) | 3.3 (3.1–3.6) | 3.1 (2.9–3.3) | 7.9 (7.6–8.2) | 8.1 (7.8–8.4) |
| 2 | | 3.5 (3.3–3.8) | 3.8 (3.6–4) | 6.3 (6.1–6.5) | 6.7 (6.5–6.9) | 3.4 (3.2–3.7) | 3.1 (2.9–3.4) | 7.7 (7.4–8) | 7.6 (7.4–7.9) |
| 3 | | 3.6 (3.4–3.8) | 4.1 (3.8–4.3) | 5.7 (5.5–5.9) | 6.1 (5.9–6.3) | 3.5 (3.2–3.7) | 3.2 (3–3.5) | 7.3 (7–7.6) | 7.5 (7.2–7.8) |
| 4 | | 3.8 (3.6–4.1) | 4.2 (4–4.5) | 5.1 (4.9–5.3) | 5.5 (5.3–5.7) | 3.4 (3.2–3.7) | 3.4 (3.1–3.6) | 6.8 (6.5–7.1) | 6.9 (6.6–7.2) |
| 5 | | 4 (3.8–4.3) | 4.6 (4.3–4.9) | 4.7 (4.5–4.9) | 5.2 (4.9–5.4) | 3.5 (3.2–3.8) | 3.5 (3.2–3.7) | 6.2 (5.9–6.5) | 6.4 (6.1–6.7) |
| -5 | 15–19 years | 2.7 (2.5–2.8) | 2.8 (2.7–2.9) | 6.9 (6.8–7) | 6.8 (6.6–6.9) | 3.1 (2.9–3.2) | 3 (2.8–3.1) | 8.1 (7.9–8.2) | 8 (7.8–8.2) |
| -4 | | 2.9 (2.7–3) | 3 (2.9–3.2) | 6.8 (6.6–6.9) | 6.6 (6.5–6.8) | 3.1 (3–3.3) | 3.1 (2.9–3.2) | 7.9 (7.7–8) | 7.9 (7.7–8) |
| -3 | | 3.1 (3–3.2) | 3.1 (3–3.3) | 6.5 (6.4–6.6) | 6.4 (6.3–6.5) | 3.3 (3.1–3.4) | 3.2 (3–3.4) | 7.7 (7.5–7.9) | 7.7 (7.5–7.9) |
| -2 | | 3.2 (3.1–3.4) | 3.4 (3.2–3.5) | 6.1 (5.9–6.2) | 6.1 (5.9–6.2) | 3.4 (3.2–3.6) | 3.2 (3.1–3.4) | 7.5 (7.3–7.7) | 7.5 (7.3–7.7) |

**eTable 44. Estimates for marginal predictions of annual social welfare benefits among parents exposed to child's eating disorder (F50) and unexposed parents stratified by the child's age at diagnosis**

| | | Finland | | | | Denmark | | | |
|---|---|---|---|---|---|---|---|---|---|
| | | Men | | Women | | Men | | Women | |
| | | Unexposed | Exposed | Unexposed | Exposed | Unexposed | Exposed | Unexposed | Exposed |
| Years since diagnosis | Age group | Marg. pred. (95% CI) | Marg. pred. (95% CI) | Marg. pred. (95% CI) | Marg. pred. (95% CI) | Marg. pred. (95% CI) | Marg. pred. (95% CI) | Marg. pred. (95% CI) | Marg. pred. (95% CI) |
| | | Unexposed | Exposed | Unexposed | Exposed | Unexposed | Exposed | Unexposed | Exposed |
| -1 | | 3.4 (3.2–3.5) | 3.6 (3.4–3.7) | 5.7 (5.6–5.9) | 5.7 (5.5–5.8) | 3.5 (3.3–3.6) | 3.3 (3.1–3.5) | 7.2 (7–7.4) | 7.2 (7–7.4) |
| 0 | | 3.6 (3.4–3.7) | 3.7 (3.6–3.9) | 5.2 (5.1–5.3) | 5.3 (5.1–5.4) | 3.5 (3.3–3.7) | 3.4 (3.2–3.6) | 6.8 (6.6–7) | 6.8 (6.6–7) |
| 1 | | 3.9 (3.7–4) | 3.9 (3.8–4.1) | 4.7 (4.6–4.9) | 4.8 (4.7–4.9) | 3.6 (3.4–3.8) | 3.5 (3.4–3.7) | 6.4 (6.2–6.6) | 6.5 (6.3–6.7) |
| 2 | | 4 (3.8–4.2) | 4.3 (4.1–4.5) | 4.3 (4.2–4.5) | 4.4 (4.3–4.5) | 3.7 (3.5–3.9) | 3.6 (3.4–3.8) | 6 (5.8–6.2) | 6 (5.8–6.2) |
| 3 | | 4.3 (4.1–4.5) | 4.7 (4.5–5) | 4.1 (4–4.3) | 4.3 (4.2–4.5) | 3.8 (3.6–4) | 3.7 (3.5–3.9) | 5.6 (5.4–5.8) | 5.7 (5.5–5.9) |
| 4 | | 4.8 (4.6–5) | 5.2 (4.9–5.4) | 4.1 (4–4.3) | 4.2 (4.1–4.4) | 4 (3.8–4.2) | 3.9 (3.7–4.1) | 5.4 (5.2–5.6) | 5.6 (5.4–5.8) |
| 5 | | 5.5 (5.2–5.7) | 5.7 (5.4–5.9) | 4.2 (4–4.4) | 4.4 (4.2–4.5) | 4.2 (4–4.4) | 4.2 (4–4.4) | 5.4 (5.2–5.6) | 5.6 (5.4–5.8) |
| -5 | 20–25 years | 3.5 (3.3–3.7) | 3.6 (3.3–3.8) | 5.1 (4.9–5.3) | 5.1 (4.9–5.3) | 3.3 (3.1–3.5) | 3.4 (3.2–3.6) | 6.3 (6.1–6.5) | 6.6 (6.4–6.9) |
| -4 | | 3.8 (3.5–4) | 3.9 (3.6–4.1) | 4.7 (4.5–4.9) | 4.7 (4.5–5) | 3.4 (3.2–3.6) | 3.5 (3.3–3.7) | 5.9 (5.7–6.2) | 6.2 (6–6.5) |
| -3 | | 4.1 (3.9–4.4) | 4.1 (3.8–4.4) | 4.3 (4.1–4.5) | 4.4 (4.2–4.6) | 3.6 (3.4–3.8) | 3.6 (3.4–3.8) | 5.6 (5.4–5.8) | 5.9 (5.6–6.1) |
| -2 | | 4.4 (4.1–4.7) | 4.4 (4.1–4.7) | 4.2 (4–4.3) | 4.3 (4.1–4.5) | 3.7 (3.5–3.9) | 3.7 (3.5–3.9) | 5.4 (5.2–5.6) | 5.6 (5.3–5.8) |
| -1 | | 4.8 (4.5–5.1) | 4.8 (4.5–5.1) | 4.1 (3.9–4.3) | 4.3 (4.1–4.5) | 3.9 (3.6–4.1) | 3.9 (3.7–4.1) | 5.3 (5.1–5.5) | 5.4 (5.2–5.6) |
| 0 | | 5.3 (5–5.6) | 5.5 (5.1–5.8) | 4.3 (4.1–4.5) | 4.4 (4.2–4.6) | 4 (3.8–4.2) | 4.2 (4–4.4) | 5.3 (5.1–5.5) | 5.5 (5.3–5.7) |

**eTable 44. Estimates for marginal predictions of annual social welfare benefits among parents exposed to child's eating disorder (F50) and unexposed parents stratified by the child's age at diagnosis**

| | | Finland | | | | Denmark | | | |
|---|---|---|---|---|---|---|---|---|---|
| | | Men | | Women | | Men | | Women | |
| | | Unexposed | Exposed | Unexposed | Exposed | Unexposed | Exposed | Unexposed | Exposed |
| Years since diagnosis | Age group | Marg. pred. (95% CI) | Marg. pred. (95% CI) | Marg. pred. (95% CI) | Marg. pred. (95% CI) | Marg. pred. (95% CI) | Marg. pred. (95% CI) | Marg. pred. (95% CI) | Marg. pred. (95% CI) |
| | | Unexposed | Exposed | Unexposed | Exposed | Unexposed | Exposed | Unexposed | Exposed |
| 1 | | 5.8 (5.4–6.1) | 6.1 (5.7–6.4) | 4.4 (4.2–4.6) | 4.6 (4.4–4.8) | 4.2 (4–4.5) | 4.5 (4.3–4.8) | 5.4 (5.2–5.7) | 5.5 (5.3–5.7) |
| 2 | | 6.6 (6.2–6.9) | 6.8 (6.4–7.2) | 4.7 (4.5–5) | 4.8 (4.6–5.1) | 4.5 (4.3–4.7) | 4.7 (4.5–4.9) | 5.5 (5.2–5.7) | 5.6 (5.4–5.8) |
| 3 | | 7.5 (7.1–7.9) | 7.6 (7.2–7.9) | 4.8 (4.6–5.1) | 5.2 (5–5.5) | 4.9 (4.6–5.1) | 4.9 (4.7–5.2) | 5.7 (5.4–5.9) | 5.8 (5.6–6.1) |
| 4 | | 8.5 (8.1–9) | 8.6 (8.2–9.1) | 5.3 (5.1–5.6) | 5.7 (5.4–6) | 5.2 (5–5.5) | 5.5 (5.2–5.7) | 5.8 (5.6–6.1) | 6.1 (5.8–6.3) |
| 5 | | 9.7 (9.2–10.2) | 9.9 (9.4–10.4) | 5.8 (5.5–6.1) | 6.2 (5.9–6.5) | 5.8 (5.5–6) | 6 (5.8–6.3) | 6.2 (5.9–6.4) | 6.4 (6.1–6.6) |

Marg. pred. = Marginal predictions in thousand euros (€)

**eTable 45. Estimates for marginal predictions of annual employment among parents exposed to child's eating disorder (F50) and unexposed parents stratified by the child's age at diagnosis**

| | | Finland | | | | Denmark | | | |
|---|---|---|---|---|---|---|---|---|---|
| | | Men | | Women | | Men | | Women | |
| | | Unexposed | Exposed | Unexposed | Exposed | Unexposed | Exposed | Unexposed | Exposed |
| Years since diagnosis | Age group | Marg. pred. (95% CI) | Marg. pred. (95% CI) | Marg. pred. (95% CI) | Marg. pred. (95% CI) | Marg. pred. (95% CI) | Marg. pred. (95% CI) | Marg. pred. (95% CI) | Marg. pred. (95% CI) |
| | | Unexposed | Exposed | Unexposed | Exposed | Unexposed | Exposed | Unexposed | Exposed |
| -5 | 5–9 years | 0.83 (0.8–0.87) | 0.83 (0.8–0.86) | 0.65 (0.61–0.69) | 0.59 (0.55–0.63) | 0.79 (0.73–0.86) | 0.76 (0.7–0.83) | 0.75 (0.69–0.82) | 0.61 (0.53–0.69) |
| -4 | | 0.83 (0.8–0.86) | 0.82 (0.78–0.85) | 0.67 (0.63–0.71) | 0.62 (0.57–0.66) | 0.83 (0.77–0.89) | 0.8 (0.74–0.87) | 0.68 (0.6–0.75) | 0.69 (0.61–0.76) |
| -3 | | 0.84 (0.81–0.87) | 0.83 (0.79–0.86) | 0.69 (0.65–0.73) | 0.66 (0.62–0.71) | 0.82 (0.76–0.88) | 0.8 (0.73–0.86) | 0.76 (0.7–0.83) | 0.7 (0.63–0.77) |
| -2 | | 0.85 (0.82–0.88) | 0.84 (0.8–0.87) | 0.71 (0.67–0.75) | 0.69 (0.65–0.73) | 0.8 (0.74–0.87) | 0.81 (0.75–0.87) | 0.73 (0.66–0.8) | 0.69 (0.62–0.76) |
| -1 | | 0.84 (0.81–0.87) | 0.84 (0.81–0.87) | 0.72 (0.68–0.76) | 0.71 (0.67–0.75) | 0.78 (0.71–0.84) | 0.79 (0.72–0.85) | 0.75 (0.68–0.82) | 0.74 (0.67–0.81) |
| 0 | | 0.82 (0.78–0.85) | 0.8 (0.77–0.84) | 0.74 (0.71–0.78) | 0.73 (0.69–0.77) | 0.77 (0.71–0.84) | 0.78 (0.71–0.84) | 0.78 (0.72–0.85) | 0.73 (0.66–0.8) |
| 1 | | 0.85 (0.81–0.88) | 0.81 (0.77–0.84) | 0.78 (0.75–0.82) | 0.75 (0.71–0.79) | 0.76 (0.69–0.83) | 0.79 (0.72–0.86) | 0.8 (0.74–0.87) | 0.68 (0.6–0.75) |
| 2 | | 0.83 (0.79–0.86) | 0.82 (0.79–0.86) | 0.79 (0.76–0.83) | 0.75 (0.72–0.79) | 0.78 (0.7–0.85) | 0.8 (0.74–0.87) | 0.79 (0.72–0.85) | 0.66 (0.58–0.74) |
| 3 | | 0.84 (0.81–0.88) | 0.81 (0.77–0.84) | 0.82 (0.79–0.86) | 0.76 (0.72–0.79) | 0.8 (0.73–0.87) | 0.81 (0.74–0.88) | 0.76 (0.69–0.83) | 0.67 (0.59–0.75) |

**eTable 45. Estimates for marginal predictions of annual employment among parents exposed to child's eating disorder (F50) and unexposed parents stratified by the child's age at diagnosis**

| | | Finland | | | | Denmark | | | |
|---|---|---|---|---|---|---|---|---|---|
| | | Men | | Women | | Men | | Women | |
| | | Unexposed | Exposed | Unexposed | Exposed | Unexposed | Exposed | Unexposed | Exposed |
| Years since diagnosis | Age group | Marg. pred. (95% CI) | Marg. pred. (95% CI) | Marg. pred. (95% CI) | Marg. pred. (95% CI) | Marg. pred. (95% CI) | Marg. pred. (95% CI) | Marg. pred. (95% CI) | Marg. pred. (95% CI) |
| | | Unexposed | Exposed | Unexposed | Exposed | Unexposed | Exposed | Unexposed | Exposed |
| 4 | | 0.86 (0.83–0.89) | 0.83 (0.79–0.86) | 0.8 (0.76–0.84) | 0.76 (0.73–0.8) | 0.76 (0.68–0.84) | 0.78 (0.7–0.86) | 0.72 (0.65–0.8) | 0.68 (0.6–0.76) |
| 5 | | 0.84 (0.81–0.88) | 0.82 (0.79–0.86) | 0.8 (0.76–0.84) | 0.79 (0.75–0.82) | 0.76 (0.68–0.85) | 0.8 (0.72–0.87) | 0.71 (0.63–0.79) | 0.7 (0.61–0.78) |
| -5 | 10–14 years | 0.85 (0.84–0.86) | 0.85 (0.84–0.86) | 0.77 (0.75–0.78) | 0.77 (0.76–0.78) | 0.8 (0.78–0.81) | 0.81 (0.8–0.82) | 0.74 (0.72–0.75) | 0.75 (0.73–0.76) |
| -4 | | 0.85 (0.84–0.86) | 0.84 (0.83–0.85) | 0.78 (0.77–0.79) | 0.78 (0.77–0.8) | 0.8 (0.79–0.81) | 0.82 (0.8–0.83) | 0.75 (0.74–0.76) | 0.76 (0.75–0.77) |
| -3 | | 0.85 (0.84–0.86) | 0.84 (0.83–0.85) | 0.79 (0.78–0.8) | 0.79 (0.78–0.8) | 0.8 (0.79–0.82) | 0.82 (0.81–0.83) | 0.76 (0.74–0.77) | 0.78 (0.77–0.8) |
| -2 | | 0.84 (0.83–0.85) | 0.84 (0.83–0.85) | 0.8 (0.79–0.81) | 0.81 (0.79–0.82) | 0.79 (0.78–0.81) | 0.82 (0.81–0.83) | 0.77 (0.75–0.78) | 0.78 (0.77–0.79) |
| -1 | | 0.84 (0.83–0.85) | 0.83 (0.82–0.84) | 0.8 (0.79–0.82) | 0.81 (0.8–0.82) | 0.78 (0.77–0.8) | 0.82 (0.8–0.83) | 0.77 (0.75–0.78) | 0.78 (0.77–0.79) |
| 0 | | 0.84 (0.83–0.85) | 0.83 (0.82–0.84) | 0.81 (0.8–0.82) | 0.82 (0.81–0.83) | 0.79 (0.78–0.8) | 0.8 (0.79–0.81) | 0.77 (0.76–0.79) | 0.78 (0.76–0.79) |
| 1 | | 0.83 (0.82–0.84) | 0.82 (0.81–0.83) | 0.83 (0.82–0.84) | 0.82 (0.81–0.83) | 0.79 (0.77–0.8) | 0.8 (0.79–0.81) | 0.77 (0.76–0.78) | 0.77 (0.76–0.78) |

**eTable 45. Estimates for marginal predictions of annual employment among parents exposed to child's eating disorder (F50) and unexposed parents stratified by the child's age at diagnosis**

| | | Finland | | | | Denmark | | | |
|---|---|---|---|---|---|---|---|---|---|
| | | Men | | Women | | Men | | Women | |
| | | Unexposed | Exposed | Unexposed | Exposed | Unexposed | Exposed | Unexposed | Exposed |
| Years since diagnosis | Age group | Marg. pred. (95% CI) | Marg. pred. (95% CI) | Marg. pred. (95% CI) | Marg. pred. (95% CI) | Marg. pred. (95% CI) | Marg. pred. (95% CI) | Marg. pred. (95% CI) | Marg. pred. (95% CI) |
| | | Unexposed | Exposed | Unexposed | Exposed | Unexposed | Exposed | Unexposed | Exposed |
| 2 | | 0.82 (0.81–0.83) | 0.82 (0.81–0.83) | 0.83 (0.82–0.84) | 0.82 (0.81–0.83) | 0.78 (0.77–0.8) | 0.8 (0.79–0.82) | 0.77 (0.76–0.78) | 0.78 (0.77–0.79) |
| 3 | | 0.82 (0.81–0.83) | 0.8 (0.79–0.81) | 0.84 (0.83–0.85) | 0.82 (0.81–0.84) | 0.78 (0.76–0.79) | 0.8 (0.79–0.81) | 0.77 (0.76–0.79) | 0.77 (0.76–0.79) |
| 4 | | 0.81 (0.8–0.82) | 0.79 (0.78–0.8) | 0.84 (0.83–0.85) | 0.83 (0.81–0.84) | 0.78 (0.76–0.79) | 0.79 (0.78–0.81) | 0.77 (0.76–0.79) | 0.77 (0.76–0.79) |
| 5 | | 0.8 (0.79–0.81) | 0.78 (0.76–0.79) | 0.84 (0.83–0.85) | 0.83 (0.82–0.84) | 0.78 (0.76–0.79) | 0.79 (0.77–0.8) | 0.77 (0.75–0.78) | 0.76 (0.75–0.78) |
| -5 | 15–19 years | 0.85 (0.84–0.86) | 0.84 (0.84–0.85) | 0.81 (0.8–0.82) | 0.82 (0.81–0.82) | 0.77 (0.76–0.78) | 0.79 (0.78–0.79) | 0.74 (0.73–0.75) | 0.75 (0.74–0.76) |
| -4 | | 0.84 (0.84–0.85) | 0.84 (0.84–0.85) | 0.81 (0.81–0.82) | 0.82 (0.82–0.83) | 0.79 (0.78–0.8) | 0.8 (0.79–0.81) | 0.77 (0.76–0.77) | 0.77 (0.76–0.78) |
| -3 | | 0.84 (0.83–0.85) | 0.84 (0.83–0.84) | 0.82 (0.81–0.83) | 0.83 (0.82–0.84) | 0.79 (0.78–0.8) | 0.8 (0.79–0.81) | 0.77 (0.76–0.78) | 0.77 (0.76–0.78) |
| -2 | | 0.83 (0.82–0.84) | 0.83 (0.82–0.84) | 0.82 (0.82–0.83) | 0.83 (0.83–0.84) | 0.78 (0.77–0.79) | 0.8 (0.79–0.8) | 0.77 (0.76–0.78) | 0.78 (0.77–0.79) |
| -1 | | 0.83 (0.82–0.83) | 0.83 (0.82–0.83) | 0.83 (0.83–0.84) | 0.84 (0.83–0.85) | 0.78 (0.77–0.79) | 0.79 (0.78–0.8) | 0.77 (0.76–0.78) | 0.77 (0.76–0.78) |

**eTable 45. Estimates for marginal predictions of annual employment among parents exposed to child's eating disorder (F50) and unexposed parents stratified by the child's age at diagnosis**

| | | Finland | | | | Denmark | | | |
|---|---|---|---|---|---|---|---|---|---|
| | | Men | | Women | | Men | | Women | |
| | | Unexposed | Exposed | Unexposed | Exposed | Unexposed | Exposed | Unexposed | Exposed |
| Years since diagnosis | Age group | Marg. pred. (95% CI) | Marg. pred. (95% CI) | Marg. pred. (95% CI) | Marg. pred. (95% CI) | Marg. pred. (95% CI) | Marg. pred. (95% CI) | Marg. pred. (95% CI) | Marg. pred. (95% CI) |
| | | Unexposed | Exposed | Unexposed | Exposed | Unexposed | Exposed | Unexposed | Exposed |
| 0 | | 0.82 (0.81–0.83) | 0.82 (0.81–0.83) | 0.84 (0.83–0.84) | 0.84 (0.83–0.85) | 0.78 (0.77–0.79) | 0.79 (0.78–0.8) | 0.77 (0.76–0.78) | 0.77 (0.76–0.78) |
| 1 | | 0.81 (0.81–0.82) | 0.81 (0.8–0.82) | 0.83 (0.82–0.84) | 0.84 (0.83–0.85) | 0.77 (0.76–0.78) | 0.78 (0.77–0.79) | 0.76 (0.75–0.77) | 0.76 (0.75–0.77) |
| 2 | | 0.8 (0.79–0.81) | 0.79 (0.78–0.8) | 0.83 (0.82–0.84) | 0.83 (0.82–0.84) | 0.75 (0.74–0.76) | 0.78 (0.77–0.79) | 0.76 (0.75–0.77) | 0.76 (0.75–0.77) |
| 3 | | 0.78 (0.78–0.79) | 0.78 (0.77–0.78) | 0.82 (0.82–0.83) | 0.83 (0.82–0.83) | 0.75 (0.74–0.76) | 0.76 (0.75–0.77) | 0.75 (0.74–0.76) | 0.76 (0.75–0.77) |
| 4 | | 0.77 (0.76–0.78) | 0.76 (0.75–0.77) | 0.82 (0.81–0.83) | 0.82 (0.81–0.83) | 0.74 (0.73–0.75) | 0.75 (0.74–0.76) | 0.75 (0.74–0.76) | 0.75 (0.74–0.76) |
| 5 | | 0.75 (0.74–0.76) | 0.75 (0.74–0.76) | 0.81 (0.8–0.82) | 0.82 (0.81–0.82) | 0.73 (0.72–0.74) | 0.73 (0.72–0.75) | 0.75 (0.74–0.76) | 0.74 (0.73–0.75) |
| -5 | 20–25 years | 0.8 (0.79–0.81) | 0.81 (0.8–0.82) | 0.81 (0.8–0.82) | 0.81 (0.8–0.82) | 0.77 (0.75–0.78) | 0.75 (0.74–0.77) | 0.75 (0.73–0.76) | 0.73 (0.72–0.74) |
| -4 | | 0.79 (0.78–0.81) | 0.8 (0.79–0.82) | 0.81 (0.8–0.82) | 0.81 (0.79–0.82) | 0.78 (0.77–0.79) | 0.78 (0.77–0.79) | 0.76 (0.75–0.77) | 0.75 (0.74–0.76) |
| -3 | | 0.78 (0.77–0.79) | 0.79 (0.77–0.8) | 0.81 (0.8–0.82) | 0.81 (0.8–0.82) | 0.77 (0.76–0.78) | 0.78 (0.77–0.79) | 0.76 (0.75–0.77) | 0.76 (0.75–0.77) |

**eTable 45. Estimates for marginal predictions of annual employment among parents exposed to child's eating disorder (F50) and unexposed parents stratified by the child's age at diagnosis**

| | | Finland | | | | Denmark | | | |
|---|---|---|---|---|---|---|---|---|---|
| | | Men | | Women | | Men | | Women | |
| | | Unexposed | Exposed | Unexposed | Exposed | Unexposed | Exposed | Unexposed | Exposed |
| Years since diagnosis | Age group | Marg. pred. (95% CI) | Marg. pred. (95% CI) | Marg. pred. (95% CI) | Marg. pred. (95% CI) | Marg. pred. (95% CI) | Marg. pred. (95% CI) | Marg. pred. (95% CI) | Marg. pred. (95% CI) |
| | | Unexposed | Exposed | Unexposed | Exposed | Unexposed | Exposed | Unexposed | Exposed |
| -2 | | 0.77 (0.76–0.78) | 0.78 (0.76–0.79) | 0.8 (0.79–0.82) | 0.8 (0.79–0.82) | 0.77 (0.76–0.78) | 0.77 (0.76–0.78) | 0.76 (0.75–0.77) | 0.76 (0.74–0.77) |
| -1 | | 0.76 (0.74–0.77) | 0.77 (0.75–0.78) | 0.79 (0.78–0.8) | 0.8 (0.79–0.81) | 0.76 (0.75–0.77) | 0.76 (0.75–0.77) | 0.75 (0.74–0.76) | 0.75 (0.74–0.76) |
| 0 | | 0.74 (0.73–0.76) | 0.74 (0.73–0.75) | 0.78 (0.77–0.8) | 0.79 (0.78–0.8) | 0.75 (0.74–0.76) | 0.75 (0.74–0.76) | 0.74 (0.73–0.75) | 0.75 (0.73–0.76) |
| 1 | | 0.72 (0.71–0.74) | 0.72 (0.7–0.73) | 0.77 (0.76–0.79) | 0.77 (0.76–0.79) | 0.74 (0.73–0.75) | 0.73 (0.72–0.74) | 0.73 (0.72–0.74) | 0.74 (0.73–0.75) |
| 2 | | 0.69 (0.68–0.71) | 0.69 (0.68–0.71) | 0.76 (0.75–0.77) | 0.76 (0.74–0.77) | 0.72 (0.71–0.73) | 0.72 (0.71–0.73) | 0.72 (0.71–0.73) | 0.72 (0.71–0.74) |
| 3 | | 0.66 (0.64–0.68) | 0.67 (0.65–0.68) | 0.75 (0.73–0.76) | 0.74 (0.72–0.75) | 0.7 (0.68–0.71) | 0.7 (0.69–0.72) | 0.71 (0.7–0.72) | 0.71 (0.7–0.72) |
| 4 | | 0.63 (0.62–0.65) | 0.63 (0.62–0.65) | 0.73 (0.71–0.74) | 0.71 (0.7–0.73) | 0.67 (0.66–0.69) | 0.68 (0.67–0.69) | 0.7 (0.69–0.71) | 0.69 (0.68–0.71) |
| 5 | | 0.6 (0.58–0.62) | 0.6 (0.59–0.62) | 0.7 (0.68–0.71) | 0.68 (0.67–0.7) | 0.65 (0.63–0.66) | 0.65 (0.64–0.67) | 0.68 (0.66–0.69) | 0.68 (0.66–0.69) |

Marg. pred. = Marginal predictions in probabilities

**eTable 46. Estimates for marginal predictions of annual earnings among parents exposed to child's personality disorder (F60) and unexposed parents stratified by the child's age at diagnosis**

| | | Finland | | | | Denmark | | | |
|---|---|---|---|---|---|---|---|---|---|
| | | Men | | Women | | Men | | Women | |
| | | Unexposed | Exposed | Unexposed | Exposed | Unexposed | Exposed | Unexposed | Exposed |
| Years since diagnosis | Age group | Marg. pred. (95% CI) | Marg. pred. (95% CI) | Marg. pred. (95% CI) | Marg. pred. (95% CI) | Marg. pred. (95% CI) | Marg. pred. (95% CI) | Marg. pred. (95% CI) | Marg. pred. (95% CI) |
| | | Unexposed | Exposed | Unexposed | Exposed | Unexposed | Exposed | Unexposed | Exposed |
| -5 | 5–14 years | 26.1 (22.9–29.3) | 24.2 (21.3–27.1) | 15.4 (13.5–17.3) | 13.1 (11.1–15.1) | 50.8 (49.1–52.5) | 49.7 (48–51.3) | 39.5 (38.6–40.4) | 39.2 (38.3–40.1) |
| -4 | | 27 (23.6–30.4) | 23.5 (20.5–26.5) | 17.5 (15.4–19.6) | 13.8 (11.6–16) | 51.3 (49.5–53.1) | 50.5 (48.9–52.1) | 40.4 (39.4–41.3) | 39.7 (38.8–40.7) |
| -3 | | 27.1 (23.6–30.7) | 23.4 (20.3–26.5) | 18.1 (16–20.2) | 14.7 (12.6–16.7) | 52.7 (50.8–54.6) | 50.5 (48.8–52.2) | 41.3 (40.3–42.2) | 40.7 (39.7–41.6) |
| -2 | | 28.1 (24.5–31.7) | 24 (20.7–27.2) | 18.1 (16–20.1) | 16.5 (14.3–18.6) | 53.2 (51.2–55.2) | 50.7 (49.1–52.3) | 41.7 (40.8–42.7) | 41.6 (40.6–42.5) |
| -1 | | 29 (25.4–32.7) | 22.9 (19.8–26.1) | 18.8 (16.6–21) | 18 (15.8–20.1) | 53.7 (51.7–55.6) | 51.1 (49.4–52.8) | 42.4 (41.4–43.4) | 41.7 (40.7–42.6) |
| 0 | | 30.7 (26.9–34.4) | 23.2 (19.9–26.5) | 19.8 (17.5–22.2) | 18.4 (16.4–20.5) | 54.4 (52.4–56.4) | 51.9 (50.1–53.7) | 42.6 (41.6–43.6) | 42 (41–42.9) |
| 1 | | 29.4 (25.6–33.2) | 22.8 (19.4–26.3) | 20.2 (17.8–22.6) | 18.9 (16.7–21.2) | 55.3 (53.2–57.5) | 52.3 (50.5–54.2) | 43.5 (42.4–44.5) | 42.3 (41.3–43.3) |
| 2 | | 29.8 (25.9–33.7) | 24.1 (20.4–27.9) | 21 (18.6–23.5) | 19.4 (17.2–21.6) | 55.4 (53.3–57.6) | 52.6 (50.7–54.5) | 43.6 (42.5–44.7) | 42.4 (41.4–43.5) |
| 3 | | 30.4 (26.3–34.5) | 25 (21–29) | 22.7 (20–25.4) | 18.6 (16.2–21.1) | 55.4 (53.1–57.6) | 52.8 (50.9–54.8) | 43.8 (42.7–45) | 42.6 (41.5–43.6) |

**eTable 46. Estimates for marginal predictions of annual earnings among parents exposed to child's personality disorder (F60) and unexposed parents stratified by the child’s age at diagnosis**

| | | Finland | | | | Denmark | | | |
|---|---|---|---|---|---|---|---|---|---|
| | | Men | | Women | | Men | | Women | |
| | | Unexposed | Exposed | Unexposed | Exposed | Unexposed | Exposed | Unexposed | Exposed |
| Years since diagnosis | Age group | Marg. pred. (95% CI) | Marg. pred. (95% CI) | Marg. pred. (95% CI) | Marg. pred. (95% CI) | Marg. pred. (95% CI) | Marg. pred. (95% CI) | Marg. pred. (95% CI) | Marg. pred. (95% CI) |
| | | Unexposed | Exposed | Unexposed | Exposed | Unexposed | Exposed | Unexposed | Exposed |
| 4 | | 30.2 (26.2–34.2) | 25.3 (21.2–29.4) | 24.3 (21.4–27.2) | 20.1 (17.6–22.5) | 56.1 (53.8–58.4) | 53.1 (51.2–55) | 43.9 (42.7–45) | 42.7 (41.6–43.8) |
| 5 | | 31.4 (27–35.9) | 25.8 (21.5–30.2) | 23.9 (21.1–26.8) | 21.5 (18.9–24.1) | 56.7 (54.4–59.1) | 53.6 (51.5–55.6) | 43.8 (42.6–45) | 42.5 (41.4–43.5) |
| -5 | 15–19 years | 31.9 (31.2–32.5) | 29.2 (28.5–29.9) | 21.1 (20.7–21.6) | 19.3 (18.9–19.7) | 53.2 (52.6–53.8) | 50.3 (49.8–50.8) | 41.5 (41.2–41.8) | 40.5 (40.2–40.8) |
| -4 | | 32.2 (31.4–32.9) | 29.3 (28.6–30) | 21.8 (21.4–22.3) | 19.9 (19.5–20.4) | 53.5 (52.9–54.1) | 50.6 (50.1–51.2) | 42 (41.7–42.3) | 40.8 (40.5–41.1) |
| -3 | | 32.3 (31.6–33) | 29.4 (28.7–30.1) | 22.4 (22–22.8) | 20.5 (20–20.9) | 54.1 (53.4–54.7) | 51 (50.5–51.6) | 42.5 (42.1–42.8) | 41.2 (40.9–41.5) |
| -2 | | 32.1 (31.3–32.8) | 29.3 (28.5–30) | 22.9 (22.4–23.3) | 20.8 (20.4–21.3) | 54.5 (53.9–55.2) | 51.4 (50.9–52) | 42.9 (42.6–43.2) | 41.7 (41.4–42) |
| -1 | | 32.4 (31.7–33.2) | 29.5 (28.8–30.3) | 23.3 (22.9–23.8) | 21.1 (20.7–21.6) | 54.8 (54.2–55.4) | 51.7 (51.1–52.3) | 42.9 (42.6–43.2) | 41.7 (41.4–42) |
| 0 | | 32.3 (31.6–33.1) | 29.2 (28.5–30) | 23.8 (23.3–24.2) | 21.2 (20.7–21.6) | 55.1 (54.4–55.7) | 51.8 (51.2–52.4) | 43 (42.7–43.3) | 41.4 (41.1–41.7) |
| 1 | | 32.4 (31.6–33.2) | 29.3 (28.6–30.1) | 24 (23.5–24.5) | 21.6 (21.1–22) | 55.1 (54.4–55.8) | 51.9 (51.4–52.5) | 42.9 (42.6–43.2) | 41.3 (41–41.6) |

**eTable 46. Estimates for marginal predictions of annual earnings among parents exposed to child's personality disorder (F60) and unexposed parents stratified by the child's age at diagnosis**

| | | Finland | | | | Denmark | | | |
|---|---|---|---|---|---|---|---|---|---|
| | | Men | | Women | | Men | | Women | |
| | | Unexposed | Exposed | Unexposed | Exposed | Unexposed | Exposed | Unexposed | Exposed |
| Years since diagnosis | Age group | Marg. pred. (95% CI) | Marg. pred. (95% CI) | Marg. pred. (95% CI) | Marg. pred. (95% CI) | Marg. pred. (95% CI) | Marg. pred. (95% CI) | Marg. pred. (95% CI) | Marg. pred. (95% CI) |
| | | Unexposed | Exposed | Unexposed | Exposed | Unexposed | Exposed | Unexposed | Exposed |
| 2 | | 32.5 (31.7–33.3) | 29.1 (28.3–29.9) | 24.4 (23.9–24.9) | 21.7 (21.2–22.2) | 55.4 (54.8–56.1) | 51.9 (51.3–52.5) | 42.9 (42.5–43.2) | 41.3 (40.9–41.6) |
| 3 | | 32 (31.1–32.8) | 29 (28.1–29.8) | 24.4 (23.9–24.9) | 21.8 (21.3–22.3) | 55.8 (55.1–56.5) | 52 (51.4–52.6) | 42.9 (42.5–43.3) | 41.2 (40.9–41.6) |
| 4 | | 31.6 (30.7–32.4) | 28.3 (27.4–29.1) | 24.4 (23.9–24.9) | 21.9 (21.4–22.4) | 55.7 (55–56.4) | 52.3 (51.7–52.9) | 43.1 (42.7–43.4) | 41.3 (40.9–41.6) |
| 5 | | 30.7 (29.8–31.6) | 28 (27.1–28.9) | 24.6 (24–25.1) | 21.9 (21.4–22.5) | 55.7 (55–56.5) | 51.7 (51.1–52.3) | 43.2 (42.8–43.6) | 41.3 (41–41.7) |
| -5 | 20–25 years | 31.8 (31.3–32.3) | 29.6 (29.1–30.1) | 22.7 (22.4–23) | 21 (20.7–21.3) | 53.7 (53.2–54.2) | 51 (50.5–51.4) | 40.5 (40.3–40.8) | 39.1 (38.9–39.4) |
| -4 | | 31.7 (31.2–32.1) | 29.3 (28.8–29.8) | 23.1 (22.8–23.4) | 21.2 (20.9–21.5) | 53.8 (53.3–54.2) | 51.1 (50.7–51.6) | 40.6 (40.4–40.9) | 39.1 (38.8–39.3) |
| -3 | | 31.5 (31–32) | 28.9 (28.4–29.4) | 23.4 (23.1–23.7) | 21.4 (21.1–21.7) | 53.8 (53.3–54.3) | 51.1 (50.6–51.6) | 40.6 (40.3–40.8) | 39.1 (38.9–39.4) |
| -2 | | 31.1 (30.6–31.6) | 28.6 (28.1–29.1) | 23.5 (23.2–23.8) | 21.5 (21.2–21.8) | 54.1 (53.6–54.6) | 50.9 (50.4–51.3) | 40.6 (40.3–40.8) | 39 (38.8–39.3) |
| -1 | | 31.2 (30.6–31.7) | 28.2 (27.7–28.7) | 23.5 (23.2–23.8) | 21.5 (21.2–21.8) | 54 (53.4–54.5) | 50.8 (50.3–51.3) | 40.6 (40.3–40.9) | 39 (38.8–39.3) |

**eTable 46. Estimates for marginal predictions of annual earnings among parents exposed to child's personality disorder (F60) and unexposed parents stratified by the child’s age at diagnosis**

| | | Finland | | | | Denmark | | | |
|---|---|---|---|---|---|---|---|---|---|
| | | Men | | Women | | Men | | Women | |
| | | Unexposed | Exposed | Unexposed | Exposed | Unexposed | Exposed | Unexposed | Exposed |
| Years since diagnosis | Age group | Marg. pred. (95% CI) | Marg. pred. (95% CI) | Marg. pred. (95% CI) | Marg. pred. (95% CI) | Marg. pred. (95% CI) | Marg. pred. (95% CI) | Marg. pred. (95% CI) | Marg. pred. (95% CI) |
| | | Unexposed | Exposed | Unexposed | Exposed | Unexposed | Exposed | Unexposed | Exposed |
| 0 | | 30.7 (30.2–31.2) | 27.7 (27.2–28.2) | 23.5 (23.2–23.8) | 21.5 (21.2–21.8) | 53.8 (53.3–54.3) | 50.9 (50.4–51.3) | 40.7 (40.4–41) | 39.1 (38.8–39.3) |
| 1 | | 29.9 (29.4–30.5) | 27.1 (26.6–27.6) | 23.4 (23.1–23.7) | 21.4 (21–21.7) | 53.7 (53.2–54.2) | 50.8 (50.3–51.3) | 40.7 (40.4–41) | 39.2 (38.9–39.4) |
| 2 | | 29.1 (28.5–29.6) | 26.5 (26–27) | 23.1 (22.7–23.4) | 21.3 (20.9–21.6) | 53.9 (53.3–54.4) | 50.7 (50.2–51.2) | 40.8 (40.5–41.1) | 39.2 (39–39.5) |
| 3 | | 28.3 (27.7–28.9) | 25.6 (25.1–26.2) | 22.8 (22.5–23.2) | 21 (20.7–21.4) | 53.6 (53–54.1) | 50.5 (50–51) | 40.8 (40.6–41.1) | 39.3 (39–39.5) |
| 4 | | 27.2 (26.6–27.7) | 24.9 (24.3–25.4) | 22.5 (22.1–22.8) | 20.7 (20.4–21.1) | 53.3 (52.7–53.8) | 49.9 (49.4–50.4) | 40.9 (40.6–41.2) | 39.3 (39.1–39.6) |
| 5 | | 25.9 (25.4–26.5) | 23.6 (23–24.2) | 21.8 (21.5–22.2) | 20.3 (19.9–20.6) | 53 (52.4–53.6) | 49.5 (48.9–50) | 40.9 (40.6–41.1) | 39.3 (39–39.6) |

Marg. pred. = Marginal predictions in thousand euros (€)

**eTable 47. Estimates for marginal predictions of annual disposable income among parents exposed to child's personality disorder (F60) and unexposed parents stratified by the child's age at diagnosis**

| | | Finland | | | | Denmark | | | |
|---|---|---|---|---|---|---|---|---|---|
| | | Men | | Women | | Men | | Women | |
| | | Unexposed | Exposed | Unexposed | Exposed | Unexposed | Exposed | Unexposed | Exposed |
| Years since diagnosis | Age group | Marg. pred. (95% CI) | Marg. pred. (95% CI) | Marg. pred. (95% CI) | Marg. pred. (95% CI) | Marg. pred. (95% CI) | Marg. pred. (95% CI) | Marg. pred. (95% CI) | Marg. pred. (95% CI) |
| | | Unexposed | Exposed | Unexposed | Exposed | Unexposed | Exposed | Unexposed | Exposed |
| -5 | 5–14 years | 23.6 (20.8–26.4) | 22.1 (19.9–24.3) | 20.7 (19.3–22.1) | 20.1 (18.4–21.7) | 30 (29.1–30.8) | 29 (28.1–29.8) | 27.4 (26.9–27.9) | 27 (26.5–27.5) |
| -4 | | 23.7 (21.5–25.8) | 22 (19.8–24.2) | 21.9 (20.5–23.4) | 20.3 (18.9–21.6) | 30.4 (29.4–31.3) | 29.7 (28.9–30.6) | 27.9 (27.4–28.5) | 27.4 (26.8–27.9) |
| -3 | | 24.8 (22.3–27.2) | 22.4 (20.3–24.5) | 22.1 (20.7–23.6) | 20.2 (18.9–21.4) | 31.6 (30.6–32.6) | 30.3 (29.4–31.2) | 28.6 (28.1–29.2) | 28 (27.5–28.6) |
| -2 | | 25.5 (22.8–28.1) | 22.7 (20.6–24.7) | 22.1 (20.8–23.4) | 21.4 (20.2–22.7) | 32.2 (31.2–33.3) | 30.6 (29.7–31.5) | 29.1 (28.5–29.7) | 28.6 (28.1–29.2) |
| -1 | | 26.3 (23.6–29) | 22 (20–24) | 22.5 (21.1–23.8) | 21.9 (20.7–23.1) | 32.9 (31.8–34) | 31.1 (30.1–32) | 29.7 (29.1–30.3) | 28.8 (28.3–29.4) |
| 0 | | 27.2 (24.7–29.8) | 22 (20.3–23.8) | 23.2 (21.8–24.6) | 21.8 (20.6–22.9) | 33.5 (32.4–34.6) | 31.7 (30.7–32.7) | 29.9 (29.2–30.5) | 29.1 (28.5–29.6) |
| 1 | | 26.2 (23.6–28.8) | 22.2 (20.4–23.9) | 23.5 (22–25) | 22.9 (21.6–24.2) | 34.4 (33.3–35.6) | 32.4 (31.4–33.4) | 30.4 (29.7–31) | 29.4 (28.9–30) |
| 2 | | 26.9 (24.2–29.7) | 22.8 (20.9–24.7) | 23.8 (22.2–25.3) | 22.8 (21.5–24.2) | 34.9 (33.7–36.1) | 32.6 (31.5–33.6) | 30.6 (30–31.3) | 29.5 (28.9–30.1) |
| 3 | | 27.9 (25–30.8) | 22.8 (20.8–24.9) | 24.6 (22.9–26.3) | 22.5 (21–23.9) | 35.3 (34–36.5) | 33.3 (32.3–34.4) | 30.7 (30–31.4) | 29.6 (29–30.3) |

**eTable 47. Estimates for marginal predictions of annual disposable income among parents exposed to child's personality disorder (F60) and unexposed parents stratified by the child's age at diagnosis**

| | | Finland | | | | Denmark | | | |
|---|---|---|---|---|---|---|---|---|---|
| | | Men | | Women | | Men | | Women | |
| | | Unexposed | Exposed | Unexposed | Exposed | Unexposed | Exposed | Unexposed | Exposed |
| Years since diagnosis | Age group | Marg. pred. (95% CI) | Marg. pred. (95% CI) | Marg. pred. (95% CI) | Marg. pred. (95% CI) | Marg. pred. (95% CI) | Marg. pred. (95% CI) | Marg. pred. (95% CI) | Marg. pred. (95% CI) |
| | | Unexposed | Exposed | Unexposed | Exposed | Unexposed | Exposed | Unexposed | Exposed |
| 4 | | 27.8 (24.9–30.6) | 23.4 (21.2–25.6) | 24.9 (23.1–26.7) | 22.8 (21.4–24.2) | 36 (34.7–37.3) | 34 (32.9–35.1) | 30.5 (29.8–31.3) | 29.6 (28.9–30.2) |
| 5 | | 29.5 (26.2–32.8) | 23.7 (21.5–26) | 25.4 (23.5–27.2) | 23.5 (22–25) | 36.9 (35.6–38.3) | 34.7 (33.5–35.8) | 30.7 (29.9–31.4) | 29.4 (28.7–30.1) |
| -5 | 15–19 years | 26.8 (26.3–27.2) | 25.2 (24.8–25.6) | 22.7 (22.4–23) | 22.3 (22.1–22.5) | 32 (31.7–32.3) | 30.1 (29.8–30.3) | 28.4 (28.2–28.6) | 27.7 (27.5–27.9) |
| -4 | | 27.3 (26.8–27.7) | 25.6 (25.1–26) | 23.1 (22.8–23.3) | 22.6 (22.4–22.9) | 32.5 (32.2–32.8) | 30.6 (30.3–30.9) | 28.9 (28.7–29.1) | 28 (27.9–28.2) |
| -3 | | 27.6 (27.1–28) | 25.8 (25.4–26.2) | 23.5 (23.2–23.7) | 23 (22.7–23.2) | 33.3 (32.9–33.6) | 31.1 (30.9–31.4) | 29.3 (29.1–29.5) | 28.4 (28.2–28.6) |
| -2 | | 27.8 (27.3–28.3) | 26 (25.6–26.5) | 23.3 (23–23.6) | 22.8 (22.5–23) | 33.8 (33.5–34.2) | 31.7 (31.4–32) | 29.7 (29.5–29.9) | 28.7 (28.5–28.9) |
| -1 | | 28.3 (27.8–28.8) | 26.4 (26–26.9) | 23.1 (22.9–23.4) | 22.4 (22.2–22.7) | 34.4 (34–34.7) | 32.2 (31.9–32.5) | 29.7 (29.5–29.9) | 28.7 (28.5–28.9) |
| 0 | | 28.4 (27.9–28.9) | 26.5 (26.1–27) | 23.1 (22.8–23.4) | 22.2 (22–22.5) | 34.9 (34.5–35.2) | 32.7 (32.4–33) | 29.7 (29.5–29.9) | 28.5 (28.3–28.7) |
| 1 | | 28.8 (28.2–29.3) | 26.8 (26.3–27.3) | 23.1 (22.8–23.4) | 22.2 (22–22.5) | 35.3 (34.9–35.7) | 33.2 (32.9–33.5) | 29.7 (29.5–29.9) | 28.4 (28.2–28.6) |

**eTable 47. Estimates for marginal predictions of annual disposable income among parents exposed to child's personality disorder (F60) and unexposed parents stratified by the child's age at diagnosis**

| | | Finland | | | | Denmark | | | |
|---|---|---|---|---|---|---|---|---|---|
| | | Men | | Women | | Men | | Women | |
| | | Unexposed | Exposed | Unexposed | Exposed | Unexposed | Exposed | Unexposed | Exposed |
| Years since diagnosis | Age group | Marg. pred. (95% CI) | Marg. pred. (95% CI) | Marg. pred. (95% CI) | Marg. pred. (95% CI) | Marg. pred. (95% CI) | Marg. pred. (95% CI) | Marg. pred. (95% CI) | Marg. pred. (95% CI) |
| | | Unexposed | Exposed | Unexposed | Exposed | Unexposed | Exposed | Unexposed | Exposed |
| 2 | | 29 (28.4–29.5) | 27 (26.6–27.5) | 23.2 (22.9–23.5) | 22.3 (22–22.5) | 35.9 (35.5–36.2) | 33.6 (33.3–34) | 29.8 (29.6–30) | 28.5 (28.3–28.6) |
| 3 | | 29.2 (28.7–29.8) | 27.4 (26.9–27.9) | 23.2 (22.9–23.5) | 22.4 (22.2–22.7) | 36.5 (36.1–36.9) | 34 (33.7–34.4) | 29.9 (29.7–30.1) | 28.5 (28.3–28.7) |
| 4 | | 29.5 (28.9–30) | 27.3 (26.8–27.8) | 23.3 (23–23.6) | 22.6 (22.3–22.9) | 36.9 (36.4–37.3) | 34.6 (34.2–34.9) | 30.1 (29.9–30.3) | 28.7 (28.5–28.9) |
| 5 | | 29.5 (28.9–30.1) | 27.5 (26.9–28) | 23.6 (23.3–23.9) | 22.9 (22.6–23.2) | 37.4 (37–37.9) | 34.7 (34.3–35.1) | 30.3 (30.1–30.6) | 28.9 (28.7–29.1) |
| -5 | 20–25 years | 27.3 (27–27.6) | 25.8 (25.5–26.1) | 22.1 (22–22.3) | 21.5 (21.4–21.7) | 32.8 (32.6–33.1) | 31 (30.8–31.3) | 27.4 (27.3–27.6) | 26.5 (26.4–26.6) |
| -4 | | 27.5 (27.2–27.8) | 26 (25.7–26.3) | 22.1 (22–22.3) | 21.5 (21.3–21.7) | 33.2 (33–33.5) | 31.5 (31.3–31.8) | 27.6 (27.4–27.7) | 26.5 (26.4–26.6) |
| -3 | | 27.8 (27.5–28.1) | 26.1 (25.8–26.4) | 22.1 (21.9–22.3) | 21.5 (21.3–21.6) | 33.7 (33.4–33.9) | 31.9 (31.6–32.1) | 27.6 (27.5–27.8) | 26.5 (26.4–26.7) |
| -2 | | 27.8 (27.5–28.1) | 26.2 (25.9–26.6) | 22.2 (22–22.4) | 21.5 (21.3–21.7) | 34.3 (34–34.6) | 32.2 (31.9–32.4) | 27.7 (27.5–27.8) | 26.6 (26.4–26.7) |
| -1 | | 28.2 (27.9–28.5) | 26.4 (26.1–26.7) | 22.3 (22.1–22.4) | 21.5 (21.3–21.7) | 34.6 (34.3–34.9) | 32.5 (32.3–32.8) | 27.8 (27.7–28) | 26.7 (26.5–26.8) |

**eTable 47. Estimates for marginal predictions of annual disposable income among parents exposed to child's personality disorder (F60) and unexposed parents stratified by the child's age at diagnosis**

| | | Finland | | | | Denmark | | | |
|---|---|---|---|---|---|---|---|---|---|
| | | Men | | Women | | Men | | Women | |
| | | Unexposed | Exposed | Unexposed | Exposed | Unexposed | Exposed | Unexposed | Exposed |
| Years since diagnosis | Age group | Marg. pred. (95% CI) | Marg. pred. (95% CI) | Marg. pred. (95% CI) | Marg. pred. (95% CI) | Marg. pred. (95% CI) | Marg. pred. (95% CI) | Marg. pred. (95% CI) | Marg. pred. (95% CI) |
| | | Unexposed | Exposed | Unexposed | Exposed | Unexposed | Exposed | Unexposed | Exposed |
| 0 | | 28.3 (28–28.7) | 26.5 (26.2–26.8) | 22.4 (22.2–22.5) | 21.6 (21.5–21.8) | 34.8 (34.6–35.1) | 32.9 (32.7–33.2) | 28 (27.9–28.2) | 26.8 (26.7–27) |
| 1 | | 28.4 (28–28.7) | 26.6 (26.2–26.9) | 22.5 (22.3–22.7) | 21.7 (21.6–21.9) | 35.2 (34.9–35.5) | 33.2 (33–33.5) | 28.2 (28–28.4) | 27 (26.9–27.2) |
| 2 | | 28.4 (28.1–28.8) | 26.7 (26.4–27) | 22.5 (22.3–22.7) | 21.9 (21.7–22.1) | 35.6 (35.3–35.9) | 33.5 (33.2–33.8) | 28.5 (28.3–28.6) | 27.2 (27.1–27.4) |
| 3 | | 28.6 (28.2–29) | 26.8 (26.4–27.1) | 22.6 (22.4–22.8) | 22 (21.8–22.2) | 35.9 (35.6–36.2) | 33.7 (33.4–33.9) | 28.6 (28.5–28.8) | 27.4 (27.3–27.6) |
| 4 | | 28.5 (28.1–28.9) | 26.8 (26.5–27.2) | 22.7 (22.5–22.9) | 22.1 (22–22.3) | 36.2 (35.9–36.5) | 33.7 (33.4–34) | 28.9 (28.7–29.1) | 27.6 (27.5–27.8) |
| 5 | | 28.6 (28.2–28.9) | 26.9 (26.5–27.2) | 22.8 (22.6–23) | 22.2 (22–22.4) | 36.3 (36–36.6) | 33.8 (33.5–34.1) | 29.1 (29–29.3) | 27.8 (27.7–28) |

Marg. pred. = Marginal predictions in thousand euros (€)

**eTable 48. Estimates for marginal predictions of annual social welfare benefits among parents exposed to child's personality disorder (F60) and unexposed parents stratified by the child's age at diagnosis**

| | | Finland | | | | Denmark | | | |
|---|---|---|---|---|---|---|---|---|---|
| | | Men | | Women | | Men | | Women | |
| | | Unexposed | Exposed | Unexposed | Exposed | Unexposed | Exposed | Unexposed | Exposed |
| Years since diagnosis | Age group | Marg. pred. (95% CI) | Marg. pred. (95% CI) | Marg. pred. (95% CI) | Marg. pred. (95% CI) | Marg. pred. (95% CI) | Marg. pred. (95% CI) | Marg. pred. (95% CI) | Marg. pred. (95% CI) |
| | | Unexposed | Exposed | Unexposed | Exposed | Unexposed | Exposed | Unexposed | Exposed |
| -5 | 5–14 years | 3.2 (2.2–4.3) | 5.1 (3.8–6.4) | 8.8 (7.6–10.1) | 11 (9.5–12.5) | 4.9 (4.4–5.4) | 5 (4.5–5.5) | 10.9 (10.3–11.5) | 12.7 (12–13.3) |
| -4 | | 3.4 (2.4–4.5) | 5.3 (4–6.6) | 8.5 (7.3–9.7) | 10.9 (9.4–12.3) | 4.8 (4.2–5.3) | 5.1 (4.6–5.6) | 10.7 (10.1–11.3) | 12.8 (12.1–13.5) |
| -3 | | 4.2 (2.3–6.1) | 5.7 (4.4–7.1) | 8.2 (7.2–9.3) | 9.7 (8.5–10.9) | 4.7 (4.2–5.3) | 5.6 (5–6.1) | 10.8 (10.2–11.5) | 12.9 (12.2–13.6) |
| -2 | | 4.2 (2.3–6.2) | 5.9 (4.6–7.2) | 8.3 (7.1–9.5) | 9.2 (8.1–10.4) | 5.1 (4.6–5.7) | 5.7 (5.1–6.3) | 10.6 (9.9–11.2) | 12.6 (11.9–13.3) |
| -1 | | 4.7 (2.4–7.1) | 6.2 (4.9–7.5) | 8.5 (7.3–9.7) | 8.8 (7.7–9.8) | 5.1 (4.6–5.7) | 5.9 (5.3–6.5) | 10.8 (10.1–11.4) | 12.5 (11.8–13.2) |
| 0 | | 4.2 (2.8–5.6) | 6.7 (5.3–8) | 8.4 (7.3–9.6) | 8.1 (7–9.1) | 5.1 (4.6–5.7) | 5.8 (5.2–6.4) | 10.3 (9.7–10.9) | 12.3 (11.6–13) |
| 1 | | 4.4 (3.2–5.6) | 6.5 (5.2–7.9) | 8.5 (7.3–9.6) | 8.4 (7.3–9.6) | 5.4 (4.8–5.9) | 6 (5.4–6.6) | 9.9 (9.2–10.5) | 12.2 (11.4–12.9) |
| 2 | | 4.6 (3.4–5.8) | 6.1 (4.8–7.4) | 8.1 (7–9.1) | 8 (6.7–9.2) | 5.3 (4.8–5.9) | 6.3 (5.7–6.9) | 9.7 (9–10.3) | 12.1 (11.4–12.9) |
| 3 | | 4.7 (3.2–6.2) | 5.8 (4.5–7.1) | 7.4 (6.2–8.6) | 8.3 (7.1–9.5) | 5.3 (4.7–5.8) | 6.5 (5.8–7.1) | 9.1 (8.5–9.8) | 11.6 (10.9–12.4) |
| 4 | | 4.1 (2.9–5.3) | 6 (4.6–7.4) | 6.6 (5.4–7.7) | 7.1 (6–8.2) | 5.4 (4.8–6) | 6.5 (5.9–7.2) | 8.1 (7.4–8.7) | 11 (10.3–11.8) |

**eTable 48. Estimates for marginal predictions of annual social welfare benefits among parents exposed to child's personality disorder (F60) and unexposed parents stratified by the child's age at diagnosis**

| | | Finland | | | | Denmark | | | |
|---|---|---|---|---|---|---|---|---|---|
| | | Men | | Women | | Men | | Women | |
| | | Unexposed | Exposed | Unexposed | Exposed | Unexposed | Exposed | Unexposed | Exposed |
| Years since diagnosis | Age group | Marg. pred. (95% CI) | Marg. pred. (95% CI) | Marg. pred. (95% CI) | Marg. pred. (95% CI) | Marg. pred. (95% CI) | Marg. pred. (95% CI) | Marg. pred. (95% CI) | Marg. pred. (95% CI) |
| | | Unexposed | Exposed | Unexposed | Exposed | Unexposed | Exposed | Unexposed | Exposed |
| 5 | | 4.9 (3.3–6.4) | 6 (4.6–7.5) | 6.9 (5.6–8.2) | 7 (5.8–8.2) | 5.7 (5–6.3) | 6.7 (6–7.4) | 7.5 (6.8–8.1) | 10.6 (9.9–11.4) |
| -5 | 15–19 years | 3.5 (3.3–3.7) | 4.4 (4.2–4.6) | 7.3 (7.1–7.5) | 8.5 (8.3–8.7) | 4.7 (4.5–4.8) | 5.5 (5.4–5.7) | 10 (9.8–10.2) | 11.6 (11.4–11.8) |
| -4 | | 3.7 (3.5–3.9) | 4.7 (4.5–4.9) | 7.2 (7–7.4) | 8.3 (8.1–8.5) | 4.8 (4.6–4.9) | 5.7 (5.5–5.9) | 9.8 (9.6–10) | 11.6 (11.4–11.8) |
| -3 | | 4 (3.8–4.2) | 4.8 (4.6–5.1) | 7.1 (6.9–7.3) | 8.1 (7.9–8.3) | 5 (4.8–5.1) | 5.9 (5.8–6.1) | 9.7 (9.5–9.9) | 11.4 (11.2–11.6) |
| -2 | | 4.3 (4.1–4.5) | 5.1 (4.8–5.3) | 6.5 (6.3–6.7) | 7.6 (7.3–7.8) | 5.1 (4.9–5.3) | 6 (5.8–6.2) | 9.4 (9.2–9.6) | 11.4 (11.1–11.6) |
| -1 | | 4.4 (4.2–4.7) | 5.2 (5–5.4) | 5.8 (5.6–6) | 6.9 (6.7–7.1) | 5.2 (5–5.4) | 6.1 (5.9–6.3) | 9 (8.8–9.2) | 10.8 (10.6–11) |
| 0 | | 4.6 (4.4–4.8) | 5.5 (5.2–5.7) | 5.4 (5.2–5.6) | 6.6 (6.4–6.8) | 5.3 (5.1–5.4) | 6.2 (6–6.3) | 8.5 (8.3–8.7) | 10.4 (10.2–10.6) |
| 1 | | 4.8 (4.6–5.1) | 5.6 (5.4–5.9) | 5.2 (5–5.4) | 6.3 (6.1–6.5) | 5.3 (5.1–5.5) | 6.2 (6–6.4) | 8.1 (7.9–8.3) | 10.1 (9.9–10.3) |
| 2 | | 5.1 (4.9–5.4) | 6 (5.7–6.3) | 5 (4.8–5.2) | 6.2 (6–6.4) | 5.4 (5.2–5.6) | 6.4 (6.2–6.6) | 7.8 (7.6–8) | 9.7 (9.5–9.9) |
| 3 | | 5.6 (5.3–5.9) | 6.3 (6–6.6) | 5 (4.8–5.2) | 6.3 (6–6.6) | 5.6 (5.4–5.8) | 6.6 (6.4–6.8) | 7.5 (7.3–7.7) | 9.6 (9.4–9.8) |
| 4 | | 6.3 (5.9–6.6) | 6.7 (6.4–7) | 5.1 (4.9–5.3) | 6.3 (6.1–6.5) | 5.8 (5.6–6) | 6.8 (6.6–7) | 7.5 (7.3–7.7) | 9.7 (9.4–9.9) |

**eTable 48. Estimates for marginal predictions of annual social welfare benefits among parents exposed to child's personality disorder (F60) and unexposed parents stratified by the child's age at diagnosis**

| | | Finland | | | | Denmark | | | |
|---|---|---|---|---|---|---|---|---|---|
| | | Men | | Women | | Men | | Women | |
| | | Unexposed | Exposed | Unexposed | Exposed | Unexposed | Exposed | Unexposed | Exposed |
| Years since diagnosis | Age group | Marg. pred. (95% CI) | Marg. pred. (95% CI) | Marg. pred. (95% CI) | Marg. pred. (95% CI) | Marg. pred. (95% CI) | Marg. pred. (95% CI) | Marg. pred. (95% CI) | Marg. pred. (95% CI) |
| | | Unexposed | Exposed | Unexposed | Exposed | Unexposed | Exposed | Unexposed | Exposed |
| 5 | | 6.8 (6.4–7.1) | 7.1 (6.8–7.4) | 5.2 (5–5.4) | 6.5 (6.3–6.8) | 6.1 (5.9–6.3) | 7.1 (6.8–7.3) | 7.5 (7.3–7.7) | 9.7 (9.5–10) |
| -5 | 20–25 years | 4.2 (4–4.3) | 4.7 (4.6–4.9) | 5.4 (5.3–5.5) | 6.3 (6.2–6.4) | 4.5 (4.3–4.6) | 5.6 (5.4–5.7) | 7.9 (7.7–8) | 9.6 (9.5–9.8) |
| -4 | | 4.5 (4.3–4.6) | 5 (4.9–5.2) | 5.1 (5–5.2) | 6 (5.9–6.1) | 4.6 (4.4–4.7) | 5.7 (5.6–5.9) | 7.6 (7.4–7.7) | 9.3 (9.2–9.5) |
| -3 | | 4.8 (4.6–5) | 5.4 (5.2–5.5) | 4.8 (4.7–4.9) | 5.7 (5.6–5.9) | 4.7 (4.6–4.9) | 5.9 (5.7–6) | 7.2 (7.1–7.4) | 9.1 (8.9–9.2) |
| -2 | | 5 (4.9–5.2) | 5.7 (5.5–5.9) | 4.7 (4.6–4.8) | 5.7 (5.5–5.8) | 5 (4.8–5.1) | 6.1 (6–6.3) | 7 (6.9–7.1) | 8.9 (8.8–9.1) |
| -1 | | 5.4 (5.2–5.6) | 6.1 (6–6.3) | 4.7 (4.6–4.9) | 5.6 (5.5–5.8) | 5.2 (5–5.3) | 6.4 (6.2–6.5) | 7 (6.8–7.1) | 8.8 (8.7–9) |
| 0 | | 5.8 (5.6–6) | 6.6 (6.4–6.8) | 4.8 (4.6–4.9) | 5.8 (5.6–5.9) | 5.4 (5.2–5.5) | 6.6 (6.5–6.8) | 7 (6.9–7.2) | 8.9 (8.7–9.1) |
| 1 | | 6.4 (6.2–6.6) | 7.2 (7–7.4) | 5 (4.8–5.1) | 6 (5.8–6.1) | 5.7 (5.5–5.8) | 7 (6.8–7.1) | 7.1 (7–7.3) | 9.1 (8.9–9.3) |
| 2 | | 7.1 (6.9–7.3) | 7.8 (7.6–8) | 5.2 (5.1–5.4) | 6.2 (6.1–6.4) | 5.9 (5.8–6.1) | 7.2 (7.1–7.4) | 7.3 (7.1–7.4) | 9.2 (9.1–9.4) |
| 3 | | 7.8 (7.6–8) | 8.4 (8.2–8.7) | 5.5 (5.4–5.7) | 6.4 (6.3–6.6) | 6.4 (6.2–6.5) | 7.7 (7.5–7.8) | 7.5 (7.3–7.6) | 9.4 (9.3–9.6) |
| 4 | | 8.6 (8.3–8.8) | 9.2 (9–9.4) | 5.9 (5.7–6) | 6.8 (6.6–7) | 6.9 (6.7–7.1) | 8.1 (8–8.3) | 7.7 (7.5–7.8) | 9.8 (9.6–10) |
| 5 | | 9.6 (9.4–9.9) | 10.2 (9.9–10.4) | 6.4 (6.2–6.6) | 7.2 (7.1–7.4) | 7.4 (7.3–7.6) | 8.7 (8.5–8.9) | 8.1 (7.9–8.2) | 10.1 (10–10.3) |

Marg. pred. = Marginal predictions in thousand euros (€)

**eTable 49. Estimates for marginal predictions of annual employment among parents exposed to child's personality disorder (F60) and unexposed parents stratified by the child's age at diagnosis**

| | | Finland | | | | Denmark | | | |
|---|---|---|---|---|---|---|---|---|---|
| | | Men | | Women | | Men | | Women | |
| | | Unexposed | Exposed | Unexposed | Exposed | Unexposed | Exposed | Unexposed | Exposed |
| Years since diagnosis | Age group | Marg. pred. (95% CI) | Marg. pred. (95% CI) | Marg. pred. (95% CI) | Marg. pred. (95% CI) | Marg. pred. (95% CI) | Marg. pred. (95% CI) | Marg. pred. (95% CI) | Marg. pred. (95% CI) |
| | | Unexposed | Exposed | Unexposed | Exposed | Unexposed | Exposed | Unexposed | Exposed |
| -5 | 5–14 years | 0.75 (0.68–0.81) | 0.68 (0.61–0.75) | 0.68 (0.61–0.75) | 0.58 (0.5–0.65) | 0.69 (0.66–0.72) | 0.67 (0.64–0.7) | 0.62 (0.59–0.65) | 0.56 (0.53–0.59) |
| -4 | | 0.75 (0.68–0.81) | 0.64 (0.57–0.71) | 0.71 (0.64–0.78) | 0.57 (0.5–0.64) | 0.71 (0.68–0.74) | 0.71 (0.69–0.74) | 0.65 (0.62–0.68) | 0.59 (0.56–0.62) |
| -3 | | 0.73 (0.66–0.8) | 0.66 (0.58–0.73) | 0.71 (0.64–0.78) | 0.65 (0.58–0.72) | 0.73 (0.71–0.76) | 0.71 (0.68–0.74) | 0.67 (0.64–0.7) | 0.6 (0.57–0.63) |
| -2 | | 0.75 (0.68–0.82) | 0.65 (0.58–0.72) | 0.75 (0.68–0.81) | 0.67 (0.6–0.74) | 0.72 (0.69–0.75) | 0.7 (0.67–0.73) | 0.68 (0.65–0.71) | 0.62 (0.59–0.65) |
| -1 | | 0.76 (0.7–0.83) | 0.64 (0.57–0.71) | 0.74 (0.68–0.81) | 0.71 (0.64–0.78) | 0.72 (0.69–0.75) | 0.7 (0.67–0.73) | 0.67 (0.64–0.7) | 0.61 (0.58–0.64) |
| 0 | | 0.76 (0.7–0.83) | 0.64 (0.57–0.71) | 0.72 (0.66–0.79) | 0.67 (0.6–0.74) | 0.72 (0.69–0.75) | 0.69 (0.66–0.72) | 0.67 (0.64–0.7) | 0.61 (0.58–0.64) |
| 1 | | 0.77 (0.71–0.83) | 0.63 (0.55–0.7) | 0.7 (0.63–0.77) | 0.74 (0.67–0.8) | 0.7 (0.67–0.73) | 0.68 (0.65–0.71) | 0.68 (0.65–0.71) | 0.61 (0.58–0.64) |
| 2 | | 0.73 (0.66–0.79) | 0.63 (0.56–0.71) | 0.74 (0.67–0.81) | 0.68 (0.61–0.75) | 0.69 (0.66–0.72) | 0.68 (0.65–0.71) | 0.67 (0.64–0.7) | 0.6 (0.57–0.63) |
| 3 | | 0.71 (0.64–0.79) | 0.65 (0.58–0.73) | 0.75 (0.68–0.82) | 0.67 (0.6–0.74) | 0.72 (0.69–0.74) | 0.68 (0.65–0.71) | 0.68 (0.65–0.71) | 0.58 (0.55–0.62) |

**eTable 49. Estimates for marginal predictions of annual employment among parents exposed to child's personality disorder (F60) and unexposed parents stratified by the child's age at diagnosis**

| | | Finland | | | | Denmark | | | |
|---|---|---|---|---|---|---|---|---|---|
| | | Men | | Women | | Men | | Women | |
| | | Unexposed | Exposed | Unexposed | Exposed | Unexposed | Exposed | Unexposed | Exposed |
| Years since diagnosis | Age group | Marg. pred. (95% CI) | Marg. pred. (95% CI) | Marg. pred. (95% CI) | Marg. pred. (95% CI) | Marg. pred. (95% CI) | Marg. pred. (95% CI) | Marg. pred. (95% CI) | Marg. pred. (95% CI) |
| | | Unexposed | Exposed | Unexposed | Exposed | Unexposed | Exposed | Unexposed | Exposed |
| 4 | | 0.71 (0.64–0.78) | 0.64 (0.57–0.72) | 0.77 (0.7–0.83) | 0.72 (0.65–0.79) | 0.69 (0.66–0.73) | 0.65 (0.62–0.68) | 0.69 (0.66–0.72) | 0.59 (0.56–0.63) |
| 5 | | 0.69 (0.62–0.76) | 0.59 (0.51–0.67) | 0.74 (0.67–0.81) | 0.73 (0.66–0.8) | 0.68 (0.65–0.71) | 0.64 (0.61–0.67) | 0.69 (0.66–0.72) | 0.6 (0.57–0.63) |
| -5 | 15–19 years | 0.79 (0.78–0.8) | 0.74 (0.72–0.75) | 0.76 (0.75–0.77) | 0.71 (0.69–0.72) | 0.7 (0.69–0.71) | 0.67 (0.66–0.68) | 0.65 (0.64–0.66) | 0.61 (0.6–0.62) |
| -4 | | 0.78 (0.76–0.79) | 0.73 (0.71–0.74) | 0.77 (0.76–0.78) | 0.71 (0.7–0.72) | 0.72 (0.71–0.73) | 0.69 (0.68–0.7) | 0.68 (0.67–0.69) | 0.63 (0.62–0.63) |
| -3 | | 0.77 (0.76–0.78) | 0.72 (0.71–0.73) | 0.76 (0.75–0.78) | 0.71 (0.69–0.72) | 0.72 (0.71–0.73) | 0.69 (0.68–0.7) | 0.69 (0.68–0.7) | 0.63 (0.62–0.64) |
| -2 | | 0.76 (0.75–0.77) | 0.71 (0.69–0.72) | 0.76 (0.75–0.77) | 0.7 (0.69–0.72) | 0.71 (0.71–0.72) | 0.69 (0.68–0.69) | 0.69 (0.68–0.7) | 0.63 (0.62–0.64) |
| -1 | | 0.75 (0.74–0.76) | 0.69 (0.68–0.7) | 0.77 (0.76–0.78) | 0.7 (0.69–0.72) | 0.71 (0.7–0.72) | 0.69 (0.68–0.69) | 0.69 (0.68–0.69) | 0.63 (0.62–0.64) |
| 0 | | 0.75 (0.74–0.76) | 0.69 (0.67–0.7) | 0.76 (0.75–0.78) | 0.7 (0.69–0.72) | 0.7 (0.69–0.71) | 0.67 (0.67–0.68) | 0.68 (0.67–0.69) | 0.63 (0.62–0.64) |
| 1 | | 0.73 (0.72–0.75) | 0.68 (0.67–0.7) | 0.77 (0.76–0.78) | 0.7 (0.69–0.71) | 0.69 (0.69–0.7) | 0.67 (0.66–0.68) | 0.68 (0.67–0.69) | 0.62 (0.61–0.63) |

**eTable 49. Estimates for marginal predictions of annual employment among parents exposed to child's personality disorder (F60) and unexposed parents stratified by the child's age at diagnosis**

| | | Finland | | | | Denmark | | | |
|---|---|---|---|---|---|---|---|---|---|
| | | Men | | Women | | Men | | Women | |
| | | Unexposed | Exposed | Unexposed | Exposed | Unexposed | Exposed | Unexposed | Exposed |
| Years since diagnosis | Age group | Marg. pred. (95% CI) | Marg. pred. (95% CI) | Marg. pred. (95% CI) | Marg. pred. (95% CI) | Marg. pred. (95% CI) | Marg. pred. (95% CI) | Marg. pred. (95% CI) | Marg. pred. (95% CI) |
| | | Unexposed | Exposed | Unexposed | Exposed | Unexposed | Exposed | Unexposed | Exposed |
| 2 | | 0.72 (0.71–0.73) | 0.66 (0.65–0.68) | 0.76 (0.75–0.78) | 0.69 (0.68–0.71) | 0.69 (0.68–0.7) | 0.65 (0.64–0.66) | 0.67 (0.66–0.68) | 0.62 (0.61–0.63) |
| 3 | | 0.71 (0.69–0.72) | 0.65 (0.63–0.66) | 0.75 (0.74–0.76) | 0.69 (0.67–0.7) | 0.68 (0.67–0.69) | 0.65 (0.64–0.66) | 0.67 (0.66–0.68) | 0.61 (0.6–0.62) |
| 4 | | 0.68 (0.67–0.7) | 0.63 (0.62–0.65) | 0.74 (0.73–0.76) | 0.68 (0.67–0.7) | 0.66 (0.65–0.67) | 0.63 (0.62–0.64) | 0.66 (0.65–0.67) | 0.6 (0.59–0.61) |
| 5 | | 0.67 (0.65–0.68) | 0.61 (0.6–0.63) | 0.73 (0.72–0.75) | 0.68 (0.66–0.69) | 0.65 (0.64–0.66) | 0.62 (0.61–0.63) | 0.65 (0.65–0.66) | 0.59 (0.58–0.6) |
| -5 | 20–25 years | 0.76 (0.75–0.76) | 0.72 (0.71–0.73) | 0.78 (0.77–0.79) | 0.72 (0.71–0.73) | 0.71 (0.7–0.71) | 0.67 (0.66–0.67) | 0.67 (0.66–0.67) | 0.61 (0.6–0.61) |
| -4 | | 0.74 (0.73–0.75) | 0.7 (0.69–0.71) | 0.78 (0.77–0.78) | 0.72 (0.71–0.73) | 0.72 (0.72–0.73) | 0.69 (0.68–0.69) | 0.69 (0.68–0.69) | 0.63 (0.62–0.63) |
| -3 | | 0.73 (0.72–0.74) | 0.68 (0.67–0.69) | 0.77 (0.77–0.78) | 0.71 (0.7–0.72) | 0.72 (0.72–0.73) | 0.69 (0.68–0.69) | 0.69 (0.69–0.7) | 0.63 (0.62–0.64) |
| -2 | | 0.72 (0.71–0.73) | 0.67 (0.66–0.68) | 0.76 (0.76–0.77) | 0.71 (0.7–0.72) | 0.72 (0.71–0.73) | 0.67 (0.67–0.68) | 0.69 (0.68–0.69) | 0.63 (0.62–0.63) |
| -1 | | 0.71 (0.7–0.71) | 0.65 (0.65–0.66) | 0.75 (0.75–0.76) | 0.7 (0.69–0.71) | 0.71 (0.7–0.71) | 0.66 (0.66–0.67) | 0.68 (0.67–0.69) | 0.62 (0.61–0.62) |

**eTable 49. Estimates for marginal predictions of annual employment among parents exposed to child's personality disorder (F60) and unexposed parents stratified by the child's age at diagnosis**

| | | Finland | | | | Denmark | | | |
|---|---|---|---|---|---|---|---|---|---|
| | | Men | | Women | | Men | | Women | |
| | | Unexposed | Exposed | Unexposed | Exposed | Unexposed | Exposed | Unexposed | Exposed |
| Years since diagnosis | Age group | Marg. pred. (95% CI) | Marg. pred. (95% CI) | Marg. pred. (95% CI) | Marg. pred. (95% CI) | Marg. pred. (95% CI) | Marg. pred. (95% CI) | Marg. pred. (95% CI) | Marg. pred. (95% CI) |
| | | Unexposed | Exposed | Unexposed | Exposed | Unexposed | Exposed | Unexposed | Exposed |
| 0 | | 0.69 (0.68–0.7) | 0.63 (0.63–0.64) | 0.75 (0.74–0.75) | 0.69 (0.68–0.7) | 0.7 (0.69–0.7) | 0.65 (0.64–0.66) | 0.67 (0.66–0.68) | 0.61 (0.6–0.61) |
| 1 | | 0.67 (0.66–0.68) | 0.61 (0.6–0.62) | 0.74 (0.73–0.74) | 0.68 (0.67–0.68) | 0.68 (0.67–0.68) | 0.63 (0.62–0.64) | 0.66 (0.65–0.67) | 0.59 (0.59–0.6) |
| 2 | | 0.64 (0.63–0.65) | 0.59 (0.58–0.6) | 0.72 (0.71–0.73) | 0.66 (0.65–0.67) | 0.66 (0.66–0.67) | 0.62 (0.61–0.63) | 0.65 (0.64–0.65) | 0.58 (0.58–0.59) |
| 3 | | 0.62 (0.61–0.63) | 0.57 (0.56–0.58) | 0.71 (0.7–0.71) | 0.65 (0.64–0.66) | 0.64 (0.64–0.65) | 0.6 (0.59–0.61) | 0.64 (0.63–0.64) | 0.57 (0.56–0.58) |
| 4 | | 0.59 (0.58–0.6) | 0.54 (0.53–0.55) | 0.68 (0.68–0.69) | 0.63 (0.62–0.64) | 0.62 (0.62–0.63) | 0.58 (0.57–0.59) | 0.62 (0.61–0.63) | 0.56 (0.55–0.57) |
| 5 | | 0.56 (0.55–0.57) | 0.52 (0.51–0.53) | 0.66 (0.65–0.67) | 0.61 (0.61–0.62) | 0.6 (0.59–0.6) | 0.55 (0.54–0.56) | 0.61 (0.6–0.61) | 0.54 (0.53–0.55) |

Marg. pred. = Marginal predictions in probabilities

**eTable 50. Estimates for marginal predictions of annual earnings among parents exposed to child's intellectual disability (F70–F79) and unexposed parents stratified by the child's age at diagnosis**

| | | Finland | | | | Denmark | | | |
|---|---|---|---|---|---|---|---|---|---|
| | | Men | | Women | | Men | | Women | |
| | | Unexposed | Exposed | Unexposed | Exposed | Unexposed | Exposed | Unexposed | Exposed |
| Years since diagnosis | Age group | Marg. pred. (95% CI) | Marg. pred. (95% CI) | Marg. pred. (95% CI) | Marg. pred. (95% CI) | Marg. pred. (95% CI) | Marg. pred. (95% CI) | Marg. pred. (95% CI) | Marg. pred. (95% CI) |
| | | Unexposed | Exposed | Unexposed | Exposed | Unexposed | Exposed | Unexposed | Exposed |
| -5 | 1–4 years | 25.6 (25–26.2) | 24.8 (24.2–25.5) | 12.9 (12.4–13.3) | 12.2 (11.8–12.7) | 43.8 (42.7–44.9) | 40.9 (39.8–42.1) | 32.7 (32–33.5) | 29.5 (28.8–30.2) |
| -4 | | 26.8 (26.2–27.4) | 26 (25.3–26.6) | 10.7 (10.3–11.1) | 9.9 (9.5–10.3) | 45.3 (44.2–46.4) | 42 (40.9–43.1) | 33.4 (32.7–34.1) | 30.4 (29.7–31.1) |
| -3 | | 28.1 (27.5–28.8) | 26.8 (26.2–27.5) | 9.9 (9.5–10.3) | 9 (8.6–9.3) | 47 (45.8–48.1) | 43.7 (42.6–44.9) | 35.2 (34.6–35.9) | 31.9 (31.3–32.6) |
| -2 | | 29.2 (28.5–29.9) | 27.4 (26.7–28.1) | 10.4 (10–10.8) | 8.8 (8.4–9.2) | 48.3 (47.1–49.4) | 45 (43.8–46.2) | 36.9 (36.2–37.6) | 33.5 (32.8–34.2) |
| -1 | | 30.5 (29.8–31.1) | 27.9 (27.2–28.6) | 11.8 (11.3–12.2) | 9.8 (9.4–10.2) | 50 (48.8–51.2) | 46.4 (45.2–47.5) | 38.4 (37.7–39.1) | 35 (34.3–35.7) |
| 0 | | 31.6 (30.9–32.3) | 29.2 (28.5–30) | 13.8 (13.3–14.3) | 12 (11.6–12.5) | 51.4 (50.2–52.7) | 47.1 (45.9–48.3) | 39.6 (38.9–40.4) | 36.1 (35.3–36.8) |
| 1 | | 33 (32.2–33.7) | 30.3 (29.5–31.1) | 15.8 (15.3–16.3) | 14.5 (14–15) | 52.4 (51.1–53.7) | 47.6 (46.3–48.8) | 41 (40.2–41.8) | 37.6 (36.8–38.3) |
| 2 | | 33.9 (33.1–34.7) | 31.3 (30.5–32.1) | 17.5 (17–18) | 16.2 (15.7–16.7) | 53.3 (51.9–54.7) | 48.4 (47.1–49.7) | 41.9 (41.1–42.7) | 38.7 (38–39.5) |
| 3 | | 34.7 (33.9–35.5) | 31.7 (30.9–32.5) | 18.9 (18.4–19.5) | 17.2 (16.7–17.8) | 54.3 (52.9–55.8) | 49.2 (47.8–50.6) | 43 (42.2–43.8) | 39.4 (38.6–40.2) |

**eTable 50. Estimates for marginal predictions of annual earnings among parents exposed to child's intellectual disability (F70–F79) and unexposed parents stratified by the child's age at diagnosis**

| | | Finland | | | | Denmark | | | |
|---|---|---|---|---|---|---|---|---|---|
| | | Men | | Women | | Men | | Women | |
| | | Unexposed | Exposed | Unexposed | Exposed | Unexposed | Exposed | Unexposed | Exposed |
| Years since diagnosis | Age group | Marg. pred. (95% CI) | Marg. pred. (95% CI) | Marg. pred. (95% CI) | Marg. pred. (95% CI) | Marg. pred. (95% CI) | Marg. pred. (95% CI) | Marg. pred. (95% CI) | Marg. pred. (95% CI) |
| | | Unexposed | Exposed | Unexposed | Exposed | Unexposed | Exposed | Unexposed | Exposed |
| 4 | | 35.5 (34.6–36.3) | 32.7 (31.8–33.5) | 20.1 (19.6–20.7) | 18.2 (17.7–18.8) | 55.4 (53.9–56.9) | 50 (48.6–51.5) | 43.9 (43–44.7) | 40 (39.2–40.8) |
| 5 | | 36.1 (35.3–37) | 33.3 (32.4–34.2) | 21.4 (20.8–22) | 19.4 (18.8–20) | 55.9 (54.3–57.4) | 50.7 (49.3–52.2) | 45.1 (44.2–46) | 40.2 (39.4–41) |
| -5 | 5–9 years | 28.4 (27.9–29) | 25 (24.4–25.6) | 9.4 (9.1–9.7) | 8.1 (7.7–8.4) | 46.7 (45.9–47.5) | 43.9 (43.1–44.6) | 35.7 (35.2–36.1) | 33.6 (33.2–34) |
| -4 | | 29.6 (29–30.2) | 25.8 (25.2–26.4) | 11.8 (11.4–12.1) | 9.8 (9.5–10.2) | 47.5 (46.7–48.3) | 44.2 (43.5–44.9) | 37.2 (36.7–37.6) | 35 (34.6–35.5) |
| -3 | | 30.6 (30–31.2) | 26.5 (25.9–27.2) | 13.8 (13.4–14.2) | 11.6 (11.2–12) | 48.6 (47.7–49.4) | 44.9 (44.2–45.6) | 38.2 (37.7–38.7) | 35.9 (35.5–36.4) |
| -2 | | 31.1 (30.5–31.7) | 27 (26.4–27.6) | 15.3 (14.9–15.7) | 12.9 (12.5–13.3) | 49.7 (48.8–50.6) | 45.9 (45.1–46.6) | 39 (38.5–39.4) | 36.6 (36.2–37) |
| -1 | | 32 (31.3–32.6) | 27.7 (27.1–28.3) | 16.7 (16.3–17.1) | 14 (13.6–14.4) | 50.3 (49.4–51.2) | 46.6 (45.9–47.4) | 40 (39.5–40.4) | 37.4 (36.9–37.8) |
| 0 | | 32.7 (32.1–33.3) | 28.2 (27.6–28.9) | 17.9 (17.5–18.4) | 15 (14.6–15.4) | 51 (50–51.9) | 47.7 (46.8–48.5) | 40.6 (40.1–41.1) | 38 (37.5–38.5) |
| 1 | | 33.5 (32.8–34.2) | 28.7 (28.1–29.4) | 19 (18.6–19.5) | 16.1 (15.6–16.5) | 52 (51–52.9) | 47.7 (46.9–48.6) | 41.3 (40.7–41.8) | 38.7 (38.2–39.2) |

**eTable 50. Estimates for marginal predictions of annual earnings among parents exposed to child's intellectual disability (F70–F79) and unexposed parents stratified by the child's age at diagnosis**

| | | Finland | | | | Denmark | | | |
|---|---|---|---|---|---|---|---|---|---|
| | | Men | | Women | | Men | | Women | |
| | | Unexposed | Exposed | Unexposed | Exposed | Unexposed | Exposed | Unexposed | Exposed |
| Years since diagnosis | Age group | Marg. pred. (95% CI) | Marg. pred. (95% CI) | Marg. pred. (95% CI) | Marg. pred. (95% CI) | Marg. pred. (95% CI) | Marg. pred. (95% CI) | Marg. pred. (95% CI) | Marg. pred. (95% CI) |
| | | Unexposed | Exposed | Unexposed | Exposed | Unexposed | Exposed | Unexposed | Exposed |
| 2 | | 34.1 (33.4–34.7) | 29.4 (28.7–30.1) | 20.2 (19.7–20.6) | 16.8 (16.4–17.3) | 53.2 (52.2–54.2) | 48.2 (47.3–49.1) | 41.8 (41.2–42.3) | 39 (38.5–39.5) |
| 3 | | 34.9 (34.1–35.6) | 29.9 (29.2–30.6) | 21.3 (20.8–21.7) | 17.7 (17.3–18.2) | 53.6 (52.5–54.6) | 48.4 (47.5–49.3) | 42.6 (42–43.1) | 39.5 (39–40) |
| 4 | | 35.4 (34.6–36.1) | 30.4 (29.7–31.1) | 22.6 (22.1–23.1) | 18.5 (18–19) | 54.2 (53.1–55.2) | 49.4 (48.5–50.4) | 43.2 (42.6–43.8) | 40.1 (39.6–40.7) |
| 5 | | 35.9 (35.1–36.7) | 30.4 (29.7–31.1) | 23.3 (22.8–23.9) | 19.3 (18.8–19.8) | 54.4 (53.3–55.5) | 50.1 (49.1–51.1) | 43.8 (43.2–44.4) | 40.3 (39.8–40.9) |
| -5 | 10–14 years | 30.5 (29.7–31.4) | 26.8 (26–27.6) | 17.3 (16.7–17.9) | 14.7 (14.1–15.3) | 49.2 (48.3–50.1) | 45.6 (44.8–46.4) | 38.8 (38.4–39.3) | 36.8 (36.3–37.2) |
| -4 | | 31.1 (30.2–32) | 27.4 (26.5–28.2) | 18.3 (17.7–18.9) | 15.3 (14.7–15.9) | 50 (49–50.9) | 45.8 (45–46.6) | 39.6 (39.1–40.1) | 37.3 (36.8–37.7) |
| -3 | | 31.3 (30.4–32.2) | 27.9 (27–28.8) | 19.2 (18.6–19.8) | 16.1 (15.6–16.7) | 50.8 (49.9–51.8) | 46.4 (45.6–47.2) | 40.3 (39.8–40.8) | 37.7 (37.3–38.2) |
| -2 | | 31.6 (30.7–32.6) | 28.3 (27.4–29.2) | 20.2 (19.6–20.9) | 16.7 (16.1–17.3) | 51.3 (50.3–52.3) | 46.9 (46.1–47.8) | 41 (40.5–41.5) | 38.3 (37.8–38.8) |
| -1 | | 31.9 (31–32.8) | 28.6 (27.7–29.6) | 21.2 (20.6–21.9) | 17.4 (16.8–18) | 51.8 (50.8–52.8) | 47.1 (46.3–48) | 41.5 (41–42) | 38.8 (38.4–39.3) |

**eTable 50. Estimates for marginal predictions of annual earnings among parents exposed to child's intellectual disability (F70–F79) and unexposed parents stratified by the child's age at diagnosis**

| | | Finland | | | | Denmark | | | |
|---|---|---|---|---|---|---|---|---|---|
| | | Men | | Women | | Men | | Women | |
| | | Unexposed | Exposed | Unexposed | Exposed | Unexposed | Exposed | Unexposed | Exposed |
| Years since diagnosis | Age group | Marg. pred. (95% CI) | Marg. pred. (95% CI) | Marg. pred. (95% CI) | Marg. pred. (95% CI) | Marg. pred. (95% CI) | Marg. pred. (95% CI) | Marg. pred. (95% CI) | Marg. pred. (95% CI) |
| | | Unexposed | Exposed | Unexposed | Exposed | Unexposed | Exposed | Unexposed | Exposed |
| 0 | | 32.3 (31.3–33.3) | 28.7 (27.7–29.6) | 21.9 (21.2–22.5) | 18.2 (17.5–18.8) | 52.4 (51.3–53.4) | 47.3 (46.5–48.2) | 42 (41.5–42.6) | 39.3 (38.8–39.8) |
| 1 | | 32.6 (31.7–33.6) | 28.5 (27.6–29.5) | 22.5 (21.8–23.1) | 18.9 (18.3–19.6) | 53 (51.9–54) | 47.5 (46.7–48.4) | 42.4 (41.9–43) | 39.6 (39.1–40.1) |
| 2 | | 33.5 (32.5–34.6) | 28.8 (27.8–29.8) | 23.5 (22.8–24.2) | 19.3 (18.7–19.9) | 53.2 (52.2–54.3) | 48.1 (47.2–49) | 43 (42.5–43.6) | 39.9 (39.4–40.5) |
| 3 | | 33.8 (32.8–34.8) | 28.9 (27.8–29.9) | 24 (23.4–24.7) | 19.6 (18.9–20.3) | 53.7 (52.6–54.8) | 48.2 (47.2–49.1) | 43.4 (42.9–44) | 40.2 (39.7–40.7) |
| 4 | | 33.9 (32.8–35) | 28.7 (27.6–29.7) | 24.6 (23.9–25.3) | 20.2 (19.5–20.8) | 54 (52.9–55.2) | 48.8 (47.8–49.7) | 43.4 (42.8–44) | 40.2 (39.7–40.7) |
| 5 | | 33.9 (32.8–35) | 28.4 (27.4–29.5) | 25.3 (24.6–26) | 20.5 (19.8–21.2) | 54.5 (53.4–55.7) | 49.1 (48.1–50.2) | 43.4 (42.7–44) | 40.1 (39.5–40.6) |
| -5 | 15–19 years | 31.8 (30.7–32.8) | 28.4 (27.3–29.5) | 19.9 (19.3–20.6) | 16.9 (16.2–17.6) | 49.9 (48.9–50.9) | 44.9 (44–45.7) | 39.8 (39.3–40.4) | 37.1 (36.6–37.5) |
| -4 | | 32.5 (31.4–33.6) | 28.6 (27.5–29.7) | 20.7 (20–21.4) | 17.5 (16.8–18.2) | 50.3 (49.3–51.3) | 45.2 (44.4–46.1) | 40.4 (39.8–40.9) | 37.7 (37.2–38.2) |
| -3 | | 32.4 (31.3–33.5) | 28.3 (27.2–29.4) | 21.4 (20.7–22.1) | 18 (17.3–18.7) | 51 (49.9–52) | 45.4 (44.5–46.3) | 40.9 (40.4–41.5) | 37.8 (37.3–38.3) |

**eTable 50. Estimates for marginal predictions of annual earnings among parents exposed to child's intellectual disability (F70–F79) and unexposed parents stratified by the child's age at diagnosis**

| | | Finland | | | | Denmark | | | |
|---|---|---|---|---|---|---|---|---|---|
| | | Men | | Women | | Men | | Women | |
| | | Unexposed | Exposed | Unexposed | Exposed | Unexposed | Exposed | Unexposed | Exposed |
| Years since diagnosis | Age group | Marg. pred. (95% CI) | Marg. pred. (95% CI) | Marg. pred. (95% CI) | Marg. pred. (95% CI) | Marg. pred. (95% CI) | Marg. pred. (95% CI) | Marg. pred. (95% CI) | Marg. pred. (95% CI) |
| | | Unexposed | Exposed | Unexposed | Exposed | Unexposed | Exposed | Unexposed | Exposed |
| -2 | | 32.9 (31.7–34) | 28.5 (27.4–29.7) | 22.1 (21.4–22.8) | 18.6 (17.9–19.3) | 51.4 (50.4–52.4) | 45.8 (45–46.7) | 41.1 (40.5–41.6) | 38 (37.5–38.5) |
| -1 | | 33.3 (32.2–34.5) | 28.4 (27.3–29.6) | 22.6 (21.8–23.3) | 19 (18.3–19.8) | 51.6 (50.6–52.7) | 45.8 (45–46.7) | 41.4 (40.9–42) | 38 (37.5–38.5) |
| 0 | | 33.4 (32.2–34.6) | 28.4 (27.3–29.6) | 23.1 (22.3–23.8) | 19.4 (18.6–20.1) | 52.2 (51.1–53.2) | 45.8 (44.9–46.6) | 41.7 (41.1–42.2) | 37.8 (37.3–38.3) |
| 1 | | 33.6 (32.4–34.8) | 28.2 (27–29.3) | 23.7 (22.9–24.4) | 19.8 (19.1–20.6) | 51.7 (50.6–52.8) | 45.9 (45–46.9) | 41.8 (41.2–42.4) | 37.5 (37–38.1) |
| 2 | | 33.1 (31.8–34.3) | 28.2 (27–29.4) | 24.1 (23.3–24.9) | 20.1 (19.3–20.9) | 52.6 (51.5–53.7) | 46 (45.1–47) | 41.5 (40.9–42.1) | 37.3 (36.8–37.9) |
| 3 | | 32.9 (31.6–34.2) | 27.7 (26.5–29) | 24.5 (23.6–25.3) | 20.4 (19.6–21.2) | 52.7 (51.6–53.9) | 46.3 (45.4–47.3) | 41.2 (40.5–41.8) | 37.2 (36.6–37.8) |
| 4 | | 32.6 (31.3–34) | 27.6 (26.3–28.8) | 25.1 (24.2–25.9) | 20.4 (19.5–21.2) | 53 (51.8–54.2) | 46 (45–47) | 41.1 (40.5–41.8) | 37.3 (36.7–37.9) |
| 5 | | 32.1 (30.7–33.5) | 26.5 (25.1–27.8) | 24.9 (24.1–25.8) | 20.5 (19.6–21.4) | 52.8 (51.6–54) | 46 (45–47) | 40.9 (40.2–41.5) | 37.3 (36.7–37.9) |
| -5 | 20–25 years | 30.5 (29–32.1) | 25.1 (23.6–26.6) | 20.7 (19.8–21.7) | 16.5 (15.5–17.4) | 49.8 (48.5–51) | 44.8 (43.7–46) | 39.2 (38.6–39.8) | 35.2 (34.6–35.8) |

**eTable 50. Estimates for marginal predictions of annual earnings among parents exposed to child's intellectual disability (F70–F79) and unexposed parents stratified by the child's age at diagnosis**

| | | Finland | | | | Denmark | | | |
|---|---|---|---|---|---|---|---|---|---|
| | | Men | | Women | | Men | | Women | |
| | | Unexposed | Exposed | Unexposed | Exposed | Unexposed | Exposed | Unexposed | Exposed |
| Years since diagnosis | Age group | Marg. pred. (95% CI) | Marg. pred. (95% CI) | Marg. pred. (95% CI) | Marg. pred. (95% CI) | Marg. pred. (95% CI) | Marg. pred. (95% CI) | Marg. pred. (95% CI) | Marg. pred. (95% CI) |
| | | Unexposed | Exposed | Unexposed | Exposed | Unexposed | Exposed | Unexposed | Exposed |
| -4 | | 30.6 (29–32.1) | 24.4 (22.8–25.9) | 20.9 (20–21.8) | 16.5 (15.5–17.5) | 50.1 (48.8–51.3) | 44.4 (43.2–45.5) | 39.1 (38.5–39.8) | 35.1 (34.4–35.7) |
| -3 | | 30.1 (28.5–31.7) | 24.4 (22.9–26) | 21.2 (20.3–22.1) | 16.4 (15.4–17.3) | 49.8 (48.5–51.1) | 44.2 (43.1–45.3) | 39.1 (38.5–39.8) | 35 (34.4–35.7) |
| -2 | | 30.2 (28.6–31.9) | 23.8 (22.2–25.4) | 21.4 (20.5–22.4) | 16.3 (15.3–17.3) | 50.2 (48.9–51.5) | 44.1 (43–45.2) | 39 (38.3–39.7) | 35.1 (34.4–35.8) |
| -1 | | 30.4 (28.6–32.1) | 23.6 (22–25.3) | 21 (20–22) | 16.6 (15.6–17.6) | 50.6 (49.2–51.9) | 44.2 (43–45.3) | 38.9 (38.2–39.6) | 35.3 (34.6–36) |
| 0 | | 29.6 (27.9–31.2) | 23.1 (21.5–24.7) | 21.1 (20.1–22.1) | 16.2 (15.3–17.2) | 50.3 (49–51.6) | 44.1 (42.9–45.3) | 38.7 (38–39.4) | 35.3 (34.6–35.9) |
| 1 | | 28.9 (27.2–30.7) | 22.7 (21.1–24.3) | 20.8 (19.8–21.9) | 16.1 (15.1–17.2) | 49.9 (48.6–51.2) | 44.1 (42.9–45.3) | 39.2 (38.5–39.9) | 35.4 (34.7–36.1) |
| 2 | | 27.6 (25.8–29.4) | 21.9 (20.3–23.6) | 21.2 (20.1–22.2) | 16.1 (15.1–17.1) | 49.6 (48.2–50.9) | 43.8 (42.6–45) | 39 (38.2–39.7) | 35.1 (34.4–35.8) |
| 3 | | 27.2 (25.3–29) | 21.2 (19.5–22.9) | 20.8 (19.7–21.9) | 15.9 (14.8–17) | 49.8 (48.4–51.2) | 44 (42.7–45.3) | 39 (38.2–39.7) | 35.2 (34.5–35.9) |
| 4 | | 26 (24.1–27.9) | 19.8 (18.1–21.6) | 20.9 (19.8–22.1) | 15.6 (14.4–16.7) | 49.5 (48–50.9) | 43.2 (42–44.4) | 39.1 (38.3–39.9) | 35 (34.2–35.7) |

**eTable 50. Estimates for marginal predictions of annual earnings among parents exposed to child's intellectual disability (F70–F79) and unexposed parents stratified by the child's age at diagnosis**

| | | Finland | | | | Denmark | | | |
|---|---|---|---|---|---|---|---|---|---|
| | | Men | | Women | | Men | | Women | |
| | | Unexposed | Exposed | Unexposed | Exposed | Unexposed | Exposed | Unexposed | Exposed |
| Years since diagnosis | Age group | Marg. pred. (95% CI) | Marg. pred. (95% CI) | Marg. pred. (95% CI) | Marg. pred. (95% CI) | Marg. pred. (95% CI) | Marg. pred. (95% CI) | Marg. pred. (95% CI) | Marg. pred. (95% CI) |
| | | Unexposed | Exposed | Unexposed | Exposed | Unexposed | Exposed | Unexposed | Exposed |
| 5 | | 24.4 (22.4–26.4) | 18.8 (17–20.6) | 20.2 (19–21.3) | 15.6 (14.4–16.7) | 49.2 (47.7–50.6) | 42.5 (41.3–43.8) | 38.9 (38.1–39.7) | 34.8 (34.1–35.6) |

Marg. pred. = Marginal predictions in thousand euros (€)

**eTable 51. Estimates for marginal predictions of annual disposable income among parents exposed to child's intellectual disability (F70–F79) and unexposed parents stratified by the child's age at diagnosis**

| | | Finland | | | | Denmark | | | |
|---|---|---|---|---|---|---|---|---|---|
| | | Men | | Women | | Men | | Women | |
| | | Unexposed | Exposed | Unexposed | Exposed | Unexposed | Exposed | Unexposed | Exposed |
| Years since diagnosis | Age group | Marg. pred. (95% CI) | Marg. pred. (95% CI) | Marg. pred. (95% CI) | Marg. pred. (95% CI) | Marg. pred. (95% CI) | Marg. pred. (95% CI) | Marg. pred. (95% CI) | Marg. pred. (95% CI) |
| | | Unexposed | Exposed | Unexposed | Exposed | Unexposed | Exposed | Unexposed | Exposed |
| -5 | 1–4 years | 21.6 (21.2–22) | 21.2 (20.8–21.6) | 15.9 (15.6–16.2) | 15.5 (15.2–15.8) | 26.5 (25.9–27.1) | 25 (24.4–25.6) | 21.9 (21.4–22.3) | 20.1 (19.6–20.5) |
| -4 | | 22.7 (22.2–23.1) | 22.4 (22–22.8) | 16.5 (16.2–16.7) | 16 (15.7–16.3) | 27.6 (27–28.2) | 25.9 (25.3–26.5) | 22.9 (22.5–23.3) | 21.2 (20.8–21.6) |
| -3 | | 23.8 (23.3–24.2) | 23.2 (22.8–23.6) | 16.9 (16.7–17.2) | 16.4 (16.2–16.7) | 28.8 (28.2–29.4) | 27.2 (26.6–27.8) | 24.6 (24.2–25) | 22.7 (22.3–23.1) |
| -2 | | 24.7 (24.3–25.2) | 24 (23.6–24.5) | 18 (17.7–18.3) | 17.2 (16.9–17.4) | 29.8 (29.2–30.5) | 28 (27.4–28.6) | 26 (25.6–26.4) | 24 (23.6–24.4) |
| -1 | | 25.5 (25.1–26) | 24.5 (24.1–25) | 19.1 (18.8–19.3) | 18.1 (17.8–18.4) | 30.8 (30.2–31.5) | 28.9 (28.3–29.6) | 27.3 (26.9–27.8) | 25.2 (24.7–25.6) |
| 0 | | 26.3 (25.9–26.8) | 25.5 (25–25.9) | 20.2 (19.9–20.5) | 19.4 (19.1–19.7) | 31.9 (31.2–32.6) | 29.6 (28.9–30.3) | 28.2 (27.7–28.6) | 26 (25.5–26.4) |
| 1 | | 27.2 (26.7–27.6) | 26.1 (25.7–26.6) | 21.4 (21.1–21.7) | 21 (20.7–21.3) | 32.7 (32–33.4) | 30.2 (29.5–30.9) | 29.3 (28.8–29.8) | 27.1 (26.6–27.6) |
| 2 | | 28 (27.5–28.5) | 27.1 (26.6–27.6) | 22.5 (22.2–22.8) | 22.1 (21.7–22.4) | 33.7 (33–34.5) | 30.8 (30.1–31.5) | 30.2 (29.7–30.7) | 28.1 (27.6–28.5) |
| 3 | | 28.6 (28.1–29.1) | 27.4 (26.9–27.9) | 23.5 (23.2–23.9) | 23.1 (22.7–23.4) | 34.5 (33.7–35.3) | 31.4 (30.6–32.2) | 30.9 (30.4–31.4) | 28.6 (28.1–29.1) |

**eTable 51. Estimates for marginal predictions of annual disposable income among parents exposed to child's intellectual disability (F70–F79) and unexposed parents stratified by the child's age at diagnosis**

| | | Finland | | | | Denmark | | | |
|---|---|---|---|---|---|---|---|---|---|
| | | Men | | Women | | Men | | Women | |
| | | Unexposed | Exposed | Unexposed | Exposed | Unexposed | Exposed | Unexposed | Exposed |
| Years since diagnosis | Age group | Marg. pred. (95% CI) | Marg. pred. (95% CI) | Marg. pred. (95% CI) | Marg. pred. (95% CI) | Marg. pred. (95% CI) | Marg. pred. (95% CI) | Marg. pred. (95% CI) | Marg. pred. (95% CI) |
| | | Unexposed | Exposed | Unexposed | Exposed | Unexposed | Exposed | Unexposed | Exposed |
| 4 | | 29.4 (28.9–29.9) | 28.2 (27.6–28.7) | 24.4 (24–24.7) | 23.7 (23.3–24) | 35.5 (34.6–36.4) | 32.3 (31.4–33.1) | 31.6 (31.1–32.2) | 29.1 (28.5–29.6) |
| 5 | | 30 (29.4–30.5) | 28.9 (28.3–29.4) | 25.3 (24.9–25.6) | 24.4 (24.1–24.8) | 36.1 (35.2–37) | 32.9 (32.1–33.8) | 32.6 (32–33.1) | 29.4 (28.8–29.9) |
| -5 | 5–9 years | 23.7 (23.4–24.1) | 22.3 (21.9–22.6) | 17.4 (17.1–17.6) | 16.7 (16.5–16.9) | 28.3 (27.8–28.7) | 26.8 (26.4–27.1) | 25.1 (24.8–25.3) | 23.9 (23.6–24.1) |
| -4 | | 24.7 (24.3–25) | 23.1 (22.7–23.4) | 18.6 (18.4–18.9) | 17.6 (17.4–17.9) | 28.8 (28.4–29.3) | 27.2 (26.9–27.6) | 26.2 (25.9–26.5) | 24.9 (24.7–25.2) |
| -3 | | 25.4 (25.1–25.8) | 23.7 (23.3–24.1) | 19.9 (19.7–20.2) | 18.8 (18.5–19) | 29.7 (29.3–30.2) | 27.9 (27.5–28.3) | 27.1 (26.8–27.3) | 25.6 (25.3–25.9) |
| -2 | | 25.9 (25.5–26.2) | 24.1 (23.8–24.5) | 21 (20.7–21.2) | 19.9 (19.6–20.1) | 30.5 (30–31) | 28.6 (28.2–29) | 27.7 (27.4–28) | 26.1 (25.8–26.4) |
| -1 | | 26.5 (26.2–26.9) | 24.8 (24.4–25.2) | 21.9 (21.7–22.2) | 20.7 (20.4–20.9) | 31.2 (30.7–31.7) | 29.3 (28.8–29.7) | 28.4 (28.1–28.7) | 26.7 (26.4–26.9) |
| 0 | | 27.2 (26.8–27.6) | 25.2 (24.8–25.6) | 22.8 (22.5–23) | 21.5 (21.2–21.7) | 31.9 (31.4–32.4) | 30.1 (29.6–30.6) | 29 (28.7–29.3) | 27.2 (26.9–27.5) |
| 1 | | 27.8 (27.4–28.2) | 25.6 (25.2–26) | 23.6 (23.3–23.9) | 22.2 (21.9–22.4) | 32.8 (32.2–33.3) | 30.4 (29.9–30.9) | 29.5 (29.2–29.8) | 27.8 (27.5–28.1) |

**eTable 51. Estimates for marginal predictions of annual disposable income among parents exposed to child's intellectual disability (F70–F79) and unexposed parents stratified by the child's age at diagnosis**

| | | Finland | | | | Denmark | | | |
|---|---|---|---|---|---|---|---|---|---|
| | | Men | | Women | | Men | | Women | |
| | | Unexposed | Exposed | Unexposed | Exposed | Unexposed | Exposed | Unexposed | Exposed |
| Years since diagnosis | Age group | Marg. pred. (95% CI) | Marg. pred. (95% CI) | Marg. pred. (95% CI) | Marg. pred. (95% CI) | Marg. pred. (95% CI) | Marg. pred. (95% CI) | Marg. pred. (95% CI) | Marg. pred. (95% CI) |
| | | Unexposed | Exposed | Unexposed | Exposed | Unexposed | Exposed | Unexposed | Exposed |
| 2 | | 28.3 (27.8–28.7) | 26 (25.6–26.4) | 24.2 (24–24.5) | 22.7 (22.4–22.9) | 33.7 (33.1–34.3) | 30.9 (30.4–31.4) | 29.9 (29.6–30.3) | 28.1 (27.8–28.4) |
| 3 | | 29.1 (28.7–29.6) | 26.6 (26.1–27) | 25 (24.7–25.2) | 23.3 (23–23.5) | 34.2 (33.6–34.8) | 31.4 (30.8–31.9) | 30.7 (30.3–31) | 28.6 (28.2–28.9) |
| 4 | | 29.6 (29.2–30.1) | 27.1 (26.7–27.5) | 25.6 (25.4–25.9) | 23.9 (23.6–24.2) | 35.1 (34.5–35.7) | 32.2 (31.7–32.8) | 31.2 (30.8–31.6) | 29.2 (28.8–29.5) |
| 5 | | 30.4 (29.9–30.8) | 27.3 (26.9–27.8) | 26.1 (25.8–26.4) | 24.3 (24–24.6) | 35.6 (35–36.2) | 33 (32.4–33.6) | 31.8 (31.4–32.2) | 29.3 (29–29.7) |
| -5 | 10–14 years | 25.1 (24.6–25.7) | 23.5 (22.9–24) | 21.4 (21.1–21.8) | 20.1 (19.8–20.4) | 29.5 (29.1–30) | 27.8 (27.4–28.2) | 27.3 (27.1–27.6) | 26.2 (25.9–26.4) |
| -4 | | 25.9 (25.4–26.5) | 24.2 (23.6–24.8) | 22.2 (21.9–22.5) | 20.7 (20.4–21.1) | 30.4 (29.9–30.9) | 28.2 (27.7–28.6) | 27.9 (27.6–28.2) | 26.5 (26.2–26.8) |
| -3 | | 26.2 (25.6–26.7) | 24.6 (24.1–25.2) | 22.9 (22.6–23.2) | 21.4 (21–21.7) | 31.2 (30.7–31.7) | 28.8 (28.4–29.3) | 28.4 (28.1–28.7) | 26.9 (26.6–27.2) |
| -2 | | 26.7 (26.1–27.3) | 25.1 (24.5–25.7) | 23.5 (23.2–23.9) | 21.8 (21.5–22.2) | 31.6 (31.1–32.2) | 29.2 (28.8–29.7) | 29 (28.7–29.3) | 27.3 (27–27.6) |
| -1 | | 27 (26.4–27.6) | 25.6 (25–26.1) | 24.1 (23.7–24.4) | 22.2 (21.8–22.5) | 32.2 (31.6–32.7) | 29.6 (29.2–30.1) | 29.4 (29.1–29.7) | 27.7 (27.4–28) |

**eTable 51. Estimates for marginal predictions of annual disposable income among parents exposed to child's intellectual disability (F70–F79) and unexposed parents stratified by the child's age at diagnosis**

| | | Finland | | | | Denmark | | | |
|---|---|---|---|---|---|---|---|---|---|
| | | Men | | Women | | Men | | Women | |
| | | Unexposed | Exposed | Unexposed | Exposed | Unexposed | Exposed | Unexposed | Exposed |
| Years since diagnosis | Age group | Marg. pred. (95% CI) | Marg. pred. (95% CI) | Marg. pred. (95% CI) | Marg. pred. (95% CI) | Marg. pred. (95% CI) | Marg. pred. (95% CI) | Marg. pred. (95% CI) | Marg. pred. (95% CI) |
| | | Unexposed | Exposed | Unexposed | Exposed | Unexposed | Exposed | Unexposed | Exposed |
| 0 | | 27.4 (26.8–28) | 25.7 (25.1–26.3) | 24.5 (24.2–24.9) | 22.6 (22.3–23) | 32.9 (32.3–33.4) | 30.1 (29.6–30.6) | 29.9 (29.6–30.2) | 28.1 (27.8–28.4) |
| 1 | | 27.9 (27.3–28.5) | 25.9 (25.3–26.5) | 24.9 (24.6–25.3) | 23 (22.6–23.3) | 33.6 (33–34.2) | 30.5 (30–31) | 30.3 (30–30.7) | 28.4 (28–28.7) |
| 2 | | 28.8 (28.1–29.4) | 26.2 (25.6–26.8) | 25.4 (25–25.7) | 23.1 (22.7–23.4) | 34.4 (33.8–35) | 31.1 (30.6–31.7) | 30.9 (30.5–31.3) | 28.7 (28.4–29) |
| 3 | | 29.2 (28.5–29.8) | 26.5 (25.9–27.1) | 25.5 (25.2–25.9) | 23 (22.7–23.4) | 35 (34.4–35.7) | 31.5 (31–32.1) | 31.2 (30.8–31.6) | 28.9 (28.6–29.3) |
| 4 | | 29.7 (29–30.4) | 26.6 (25.9–27.2) | 25.5 (25.1–25.9) | 22.9 (22.5–23.3) | 35.6 (34.9–36.2) | 32.2 (31.6–32.8) | 31.3 (30.9–31.7) | 28.9 (28.6–29.3) |
| 5 | | 29.8 (29.1–30.5) | 26.8 (26.1–27.4) | 25.7 (25.3–26.1) | 22.9 (22.5–23.3) | 36.3 (35.6–37) | 32.8 (32.2–33.5) | 31.2 (30.8–31.6) | 28.8 (28.4–29.2) |
| -5 | 15–19 years | 26.5 (25.8–27.1) | 25.1 (24.4–25.8) | 22.6 (22.2–23) | 21.3 (20.9–21.7) | 30.8 (30.3–31.3) | 28 (27.5–28.5) | 27.9 (27.6–28.2) | 26.3 (26–26.6) |
| -4 | | 27.2 (26.5–27.9) | 25.1 (24.5–25.8) | 23 (22.6–23.4) | 21.7 (21.3–22.1) | 31.4 (30.9–31.9) | 28.5 (28–28.9) | 28.4 (28–28.7) | 26.8 (26.5–27.1) |
| -3 | | 27.4 (26.7–28) | 25.3 (24.6–26) | 23.4 (23–23.9) | 22 (21.5–22.4) | 32 (31.5–32.6) | 28.8 (28.3–29.3) | 28.8 (28.5–29.2) | 26.9 (26.6–27.2) |

**eTable 51. Estimates for marginal predictions of annual disposable income among parents exposed to child's intellectual disability (F70–F79) and unexposed parents stratified by the child's age at diagnosis**

| | | Finland | | | | Denmark | | | |
|---|---|---|---|---|---|---|---|---|---|
| | | Men | | Women | | Men | | Women | |
| | | Unexposed | Exposed | Unexposed | Exposed | Unexposed | Exposed | Unexposed | Exposed |
| Years since diagnosis | Age group | Marg. pred. (95% CI) | Marg. pred. (95% CI) | Marg. pred. (95% CI) | Marg. pred. (95% CI) | Marg. pred. (95% CI) | Marg. pred. (95% CI) | Marg. pred. (95% CI) | Marg. pred. (95% CI) |
| | | Unexposed | Exposed | Unexposed | Exposed | Unexposed | Exposed | Unexposed | Exposed |
| -2 | | 27.9 (27.2–28.6) | 25.5 (24.9–26.2) | 23.6 (23.2–24.1) | 22.1 (21.7–22.5) | 32.7 (32.1–33.2) | 29.4 (28.9–29.9) | 29 (28.6–29.3) | 27.1 (26.8–27.4) |
| -1 | | 28.2 (27.5–28.9) | 25.8 (25.1–26.5) | 23.7 (23.3–24.1) | 22.2 (21.8–22.7) | 33.2 (32.6–33.8) | 29.6 (29.1–30.2) | 29.3 (29–29.7) | 27.1 (26.8–27.4) |
| 0 | | 28.8 (28–29.5) | 25.8 (25.1–26.5) | 23.7 (23.2–24.1) | 22 (21.6–22.5) | 33.8 (33.2–34.4) | 30.1 (29.5–30.6) | 29.5 (29.1–29.9) | 26.9 (26.6–27.3) |
| 1 | | 28.9 (28.1–29.6) | 26 (25.2–26.7) | 23.7 (23.2–24.1) | 21.9 (21.5–22.3) | 34 (33.4–34.7) | 30.5 (29.9–31) | 29.6 (29.3–30) | 26.8 (26.5–27.2) |
| 2 | | 29 (28.2–29.7) | 26.2 (25.4–26.9) | 23.6 (23.1–24) | 21.7 (21.2–22.1) | 34.9 (34.3–35.6) | 30.9 (30.4–31.5) | 29.5 (29.1–29.9) | 26.7 (26.3–27) |
| 3 | | 29.1 (28.3–29.9) | 26.1 (25.4–26.9) | 23.6 (23.1–24.1) | 21.8 (21.4–22.3) | 35.3 (34.7–36) | 31.5 (31–32.1) | 29.3 (28.9–29.7) | 26.6 (26.2–26.9) |
| 4 | | 29.5 (28.7–30.4) | 26.4 (25.7–27.2) | 23.9 (23.4–24.3) | 21.8 (21.3–22.3) | 35.9 (35.2–36.6) | 31.5 (30.9–32.2) | 29.4 (29–29.8) | 26.7 (26.3–27.1) |
| 5 | | 29.4 (28.5–30.2) | 26.3 (25.5–27.1) | 23.9 (23.4–24.4) | 22 (21.5–22.5) | 36.2 (35.5–37) | 31.8 (31.1–32.4) | 29.4 (28.9–29.8) | 26.9 (26.5–27.3) |
| -5 | 20–25 years | 26.4 (25.4–27.4) | 23.8 (22.9–24.7) | 21.1 (20.6–21.7) | 18.7 (18.1–19.3) | 31.1 (30.4–31.8) | 28.6 (27.9–29.2) | 27 (26.6–27.3) | 24.7 (24.3–25.1) |

**eTable 51. Estimates for marginal predictions of annual disposable income among parents exposed to child's intellectual disability (F70–F79) and unexposed parents stratified by the child's age at diagnosis**

| | | Finland | | | | Denmark | | | |
|---|---|---|---|---|---|---|---|---|---|
| | | Men | | Women | | Men | | Women | |
| | | Unexposed | Exposed | Unexposed | Exposed | Unexposed | Exposed | Unexposed | Exposed |
| Years since diagnosis | Age group | Marg. pred. (95% CI) | Marg. pred. (95% CI) | Marg. pred. (95% CI) | Marg. pred. (95% CI) | Marg. pred. (95% CI) | Marg. pred. (95% CI) | Marg. pred. (95% CI) | Marg. pred. (95% CI) |
| | | Unexposed | Exposed | Unexposed | Exposed | Unexposed | Exposed | Unexposed | Exposed |
| -4 | | 26.6 (25.6–27.6) | 23.8 (22.8–24.7) | 21 (20.5–21.6) | 18.5 (17.9–19.1) | 31.7 (31–32.4) | 28.7 (28.1–29.3) | 27 (26.6–27.4) | 24.6 (24.2–25) |
| -3 | | 26.6 (25.6–27.6) | 23.9 (23–24.9) | 21 (20.4–21.5) | 18.5 (17.9–19) | 31.8 (31.1–32.6) | 29 (28.3–29.6) | 27.1 (26.7–27.5) | 24.6 (24.2–25) |
| -2 | | 26.9 (25.9–27.9) | 23.7 (22.7–24.7) | 21.1 (20.5–21.6) | 18.5 (17.9–19.1) | 32.6 (31.9–33.3) | 29.2 (28.6–29.8) | 27 (26.6–27.4) | 24.6 (24.2–25.1) |
| -1 | | 27.1 (26.1–28.2) | 24.1 (23.1–25.1) | 20.9 (20.4–21.5) | 18.6 (18.1–19.2) | 33.2 (32.4–33.9) | 29.6 (28.9–30.3) | 27 (26.6–27.4) | 24.8 (24.4–25.2) |
| 0 | | 26.9 (25.9–27.9) | 23.9 (22.9–24.9) | 20.9 (20.4–21.5) | 18.4 (17.9–19) | 33.3 (32.5–34.1) | 29.8 (29.1–30.5) | 27 (26.5–27.4) | 24.9 (24.5–25.3) |
| 1 | | 27.2 (26.1–28.2) | 24.3 (23.3–25.3) | 21.1 (20.5–21.7) | 18.8 (18.2–19.4) | 33.6 (32.8–34.4) | 30.1 (29.4–30.8) | 27.4 (27–27.9) | 25.1 (24.7–25.5) |
| 2 | | 27.1 (26–28.2) | 24 (23–25) | 21.3 (20.7–21.9) | 19 (18.4–19.6) | 33.7 (32.9–34.5) | 30.2 (29.5–30.9) | 27.5 (27.1–28) | 25.1 (24.6–25.5) |
| 3 | | 27.4 (26.3–28.5) | 24.3 (23.2–25.3) | 21.5 (20.9–22) | 19 (18.4–19.7) | 34.2 (33.3–35.1) | 30.7 (29.9–31.5) | 27.6 (27.2–28.1) | 25.3 (24.8–25.8) |
| 4 | | 27.5 (26.3–28.7) | 24.1 (23–25.2) | 21.9 (21.3–22.6) | 19.2 (18.6–19.9) | 34.3 (33.5–35.2) | 30.4 (29.7–31.2) | 27.9 (27.4–28.4) | 25.3 (24.8–25.8) |
| 5 | | 27.7 (26.5–28.9) | 24 (22.9–25.1) | 21.8 (21.2–22.4) | 19.4 (18.8–20.1) | 34.4 (33.5–35.3) | 30.4 (29.6–31.1) | 27.9 (27.4–28.4) | 25.4 (24.9–25.9) |

**eTable 51. Estimates for marginal predictions of annual disposable income among parents exposed to child's intellectual disability (F70–F79) and unexposed parents stratified by the child's age at diagnosis**

| | | Finland | | | | Denmark | | | |
|---|---|---|---|---|---|---|---|---|---|
| | | Men | | Women | | Men | | Women | |
| | | Unexposed | Exposed | Unexposed | Exposed | Unexposed | Exposed | Unexposed | Exposed |
| Years since diagnosis | Age group | Marg. pred. (95% CI) | Marg. pred. (95% CI) | Marg. pred. (95% CI) | Marg. pred. (95% CI) | Marg. pred. (95% CI) | Marg. pred. (95% CI) | Marg. pred. (95% CI) | Marg. pred. (95% CI) |
| | | Unexposed | Exposed | Unexposed | Exposed | Unexposed | Exposed | Unexposed | Exposed |

Marg. pred. = Marginal predictions in thousand euros (€)

**eTable 52. Estimates for marginal predictions of annual social welfare benefits among parents exposed to child's intellectual disability (F70–F79) and unexposed parents stratified by the child's age at diagnosis**

| | | Finland | | | | Denmark | | | |
|---|---|---|---|---|---|---|---|---|---|
| | | Men | | Women | | Men | | Women | |
| | | Unexposed | Exposed | Unexposed | Exposed | Unexposed | Exposed | Unexposed | Exposed |
| Years since diagnosis | Age group | Marg. pred. (95% CI) | Marg. pred. (95% CI) | Marg. pred. (95% CI) | Marg. pred. (95% CI) | Marg. pred. (95% CI) | Marg. pred. (95% CI) | Marg. pred. (95% CI) | Marg. pred. (95% CI) |
| | | Unexposed | Exposed | Unexposed | Exposed | Unexposed | Exposed | Unexposed | Exposed |
| -5 | 1–4 years | 2.6 (2.4–2.8) | 3 (2.8–3.1) | 6.9 (6.6–7.1) | 7.1 (6.8–7.4) | 3.8 (3.5–4.1) | 5.2 (4.9–5.6) | 8.7 (8.3–9.1) | 9.8 (9.3–10.2) |
| -4 | | 2.9 (2.8–3.1) | 3.4 (3.2–3.6) | 9.5 (9.2–9.7) | 9.7 (9.4–10) | 4 (3.7–4.3) | 5.5 (5.1–5.9) | 11.8 (11.3–12.2) | 13 (12.6–13.5) |
| -3 | | 3 (2.8–3.2) | 3.5 (3.3–3.7) | 10.5 (10.3–10.7) | 10.9 (10.7–11.1) | 4.2 (3.9–4.5) | 5.7 (5.3–6) | 13 (12.6–13.5) | 14.8 (14.3–15.3) |
| -2 | | 3.1 (2.9–3.3) | 3.8 (3.6–4) | 10.9 (10.7–11.2) | 11.5 (11.3–11.8) | 4.1 (3.7–4.4) | 5.7 (5.3–6.1) | 13 (12.6–13.5) | 14.5 (14.1–15) |
| -1 | | 3 (2.8–3.2) | 3.9 (3.7–4.1) | 10.7 (10.5–11) | 11.5 (11.2–11.7) | 4.1 (3.7–4.4) | 5.5 (5.1–5.9) | 12.6 (12.1–13) | 14.1 (13.6–14.5) |
| 0 | | 2.9 (2.7–3.1) | 3.9 (3.6–4.1) | 10.1 (9.9–10.3) | 10.8 (10.5–11) | 4 (3.6–4.3) | 5.6 (5.2–6) | 12.2 (11.7–12.6) | 13.8 (13.3–14.3) |
| 1 | | 2.9 (2.7–3.1) | 3.8 (3.6–4) | 9.7 (9.5–10) | 10.3 (10–10.5) | 3.9 (3.6–4.3) | 5.8 (5.4–6.2) | 11.8 (11.4–12.3) | 13.7 (13.2–14.2) |
| 2 | | 2.9 (2.7–3.1) | 4 (3.7–4.2) | 9.4 (9.1–9.6) | 10 (9.8–10.3) | 4 (3.6–4.4) | 6 (5.6–6.5) | 11.8 (11.3–12.3) | 13.5 (12.9–14) |
| 3 | | 3 (2.8–3.2) | 4 (3.7–4.2) | 9.2 (8.9–9.4) | 10.1 (9.9–10.4) | 4 (3.7–4.4) | 6.2 (5.7–6.7) | 11.4 (10.9–11.9) | 13 (12.5–13.6) |
| 4 | | 3.2 (3–3.4) | 3.9 (3.7–4.2) | 9.1 (8.9–9.4) | 10 (9.8–10.3) | 4 (3.7–4.4) | 6.5 (5.9–7.1) | 11.1 (10.5–11.6) | 12.8 (12.2–13.3) |
| 5 | | 3.2 (3–3.5) | 4 (3.7–4.2) | 9 (8.7–9.3) | 9.8 (9.6–10.1) | 4.1 (3.7–4.5) | 6.2 (5.7–6.7) | 10.6 (10.1–11.1) | 12.5 (12–13.1) |

**eTable 52. Estimates for marginal predictions of annual social welfare benefits among parents exposed to child's intellectual disability (F70–F79) and unexposed parents stratified by the child's age at diagnosis**

| | | Finland | | | | Denmark | | | |
|---|---|---|---|---|---|---|---|---|---|
| | | Men | | Women | | Men | | Women | |
| | | Unexposed | Exposed | Unexposed | Exposed | Unexposed | Exposed | Unexposed | Exposed |
| Years since diagnosis | Age group | Marg. pred. (95% CI) | Marg. pred. (95% CI) | Marg. pred. (95% CI) | Marg. pred. (95% CI) | Marg. pred. (95% CI) | Marg. pred. (95% CI) | Marg. pred. (95% CI) | Marg. pred. (95% CI) |
| | | Unexposed | Exposed | Unexposed | Exposed | Unexposed | Exposed | Unexposed | Exposed |
| -5 | 5–9 years | 3 (2.9–3.2) | 4.1 (3.9–4.3) | 11.5 (11.3–11.7) | 11.8 (11.6–12) | 4.8 (4.5–5.1) | 5.7 (5.5–6) | 14 (13.7–14.3) | 15.4 (15–15.7) |
| -4 | | 3.1 (2.9–3.3) | 4.2 (4–4.4) | 10.5 (10.3–10.7) | 11.1 (10.9–11.3) | 4.8 (4.5–5.1) | 5.8 (5.5–6.1) | 13.4 (13–13.7) | 14.8 (14.4–15.1) |
| -3 | | 3.1 (2.9–3.3) | 4.3 (4.1–4.5) | 10.1 (9.9–10.3) | 10.7 (10.5–10.9) | 4.7 (4.4–5) | 5.8 (5.5–6.1) | 13 (12.6–13.3) | 14.6 (14.3–15) |
| -2 | | 3.2 (3–3.3) | 4.3 (4.1–4.5) | 9.8 (9.6–10) | 10.6 (10.4–10.8) | 4.5 (4.3–4.8) | 5.9 (5.6–6.2) | 12.7 (12.4–13) | 14.4 (14–14.7) |
| -1 | | 3.2 (3.1–3.4) | 4.4 (4.3–4.6) | 9.6 (9.4–9.8) | 10.6 (10.3–10.8) | 4.7 (4.4–5) | 6.1 (5.8–6.4) | 12.4 (12.1–12.8) | 14.2 (13.8–14.5) |
| 0 | | 3.3 (3.1–3.5) | 4.5 (4.3–4.7) | 9.4 (9.2–9.6) | 10.5 (10.3–10.7) | 4.8 (4.5–5.1) | 6.2 (5.8–6.5) | 12.2 (11.8–12.5) | 14.3 (13.9–14.6) |
| 1 | | 3.3 (3.1–3.5) | 4.6 (4.4–4.8) | 9.4 (9.2–9.6) | 10.4 (10.2–10.6) | 4.9 (4.6–5.2) | 6.3 (6–6.7) | 11.9 (11.5–12.2) | 14.1 (13.7–14.5) |
| 2 | | 3.4 (3.2–3.6) | 4.6 (4.4–4.8) | 9.1 (8.9–9.4) | 10.3 (10–10.5) | 4.9 (4.6–5.2) | 6.5 (6.1–6.8) | 11.5 (11.2–11.9) | 13.7 (13.4–14.1) |
| 3 | | 3.5 (3.3–3.7) | 4.7 (4.5–4.9) | 9 (8.8–9.2) | 10.1 (9.9–10.3) | 5.1 (4.8–5.4) | 6.6 (6.2–6.9) | 11.5 (11.1–11.8) | 13.7 (13.3–14.1) |
| 4 | | 3.7 (3.5–3.9) | 4.8 (4.6–5) | 8.6 (8.4–8.9) | 10 (9.8–10.3) | 5.2 (4.9–5.6) | 6.8 (6.4–7.1) | 11.2 (10.8–11.6) | 13.6 (13.2–14) |

**eTable 52. Estimates for marginal predictions of annual social welfare benefits among parents exposed to child's intellectual disability (F70–F79) and unexposed parents stratified by the child's age at diagnosis**

| | | Finland | | | | Denmark | | | |
|---|---|---|---|---|---|---|---|---|---|
| | | Men | | Women | | Men | | Women | |
| | | Unexposed | Exposed | Unexposed | Exposed | Unexposed | Exposed | Unexposed | Exposed |
| Years since diagnosis | Age group | Marg. pred. (95% CI) | Marg. pred. (95% CI) | Marg. pred. (95% CI) | Marg. pred. (95% CI) | Marg. pred. (95% CI) | Marg. pred. (95% CI) | Marg. pred. (95% CI) | Marg. pred. (95% CI) |
| | | Unexposed | Exposed | Unexposed | Exposed | Unexposed | Exposed | Unexposed | Exposed |
| 5 | | 3.7 (3.5–3.9) | 5 (4.7–5.2) | 8.5 (8.3–8.7) | 9.9 (9.6–10.3) | 5.4 (5–5.7) | 7 (6.6–7.3) | 11.1 (10.7–11.5) | 13.4 (13–13.8) |
| -5 | 10–14 years | 3.1 (2.8–3.3) | 4 (3.7–4.3) | 9 (8.7–9.2) | 9.7 (9.4–10) | 4.8 (4.5–5.1) | 6 (5.7–6.3) | 12.6 (12.2–13) | 14.4 (14–14.8) |
| -4 | | 3.2 (3–3.5) | 4.2 (3.9–4.5) | 8.8 (8.5–9.1) | 9.8 (9.5–10.1) | 5 (4.7–5.3) | 6.2 (5.8–6.5) | 12.3 (12–12.7) | 14.2 (13.8–14.5) |
| -3 | | 3.6 (3.3–3.8) | 4.3 (4.1–4.6) | 8.8 (8.5–9) | 9.7 (9.4–10) | 5.1 (4.8–5.4) | 6.3 (6–6.7) | 12 (11.7–12.4) | 13.9 (13.6–14.3) |
| -2 | | 3.7 (3.4–4) | 4.4 (4.1–4.7) | 8.6 (8.3–8.9) | 9.6 (9.3–9.9) | 5.1 (4.8–5.4) | 6.4 (6.1–6.8) | 11.6 (11.2–12) | 13.8 (13.4–14.2) |
| -1 | | 3.9 (3.6–4.2) | 4.7 (4.4–5) | 8.3 (8–8.6) | 9.4 (9.1–9.7) | 5.3 (5–5.6) | 6.6 (6.3–6.9) | 11.5 (11.1–11.9) | 13.6 (13.2–13.9) |
| 0 | | 3.9 (3.6–4.2) | 4.9 (4.5–5.2) | 8.3 (8–8.6) | 9.2 (8.9–9.5) | 5.4 (5.1–5.7) | 6.8 (6.5–7.2) | 11.3 (11–11.7) | 13.6 (13.2–14) |
| 1 | | 4.2 (3.9–4.5) | 5.2 (4.9–5.6) | 8.2 (7.9–8.5) | 8.9 (8.6–9.2) | 5.4 (5.1–5.8) | 7.1 (6.7–7.4) | 11.2 (10.8–11.6) | 13.4 (13–13.8) |
| 2 | | 4.2 (3.9–4.5) | 5.3 (4.9–5.6) | 7.9 (7.6–8.2) | 8.7 (8.4–9) | 5.6 (5.3–5.9) | 7.4 (7–7.7) | 10.9 (10.6–11.3) | 13.1 (12.7–13.5) |
| 3 | | 4.3 (4–4.7) | 5.5 (5.1–5.9) | 7.6 (7.3–7.9) | 8.3 (8–8.7) | 5.8 (5.5–6.2) | 7.4 (7.1–7.8) | 10.8 (10.4–11.2) | 13 (12.6–13.4) |
| 4 | | 4.6 (4.3–5) | 5.7 (5.3–6.1) | 7 (6.7–7.3) | 7.8 (7.4–8.1) | 5.9 (5.6–6.3) | 7.6 (7.2–8) | 10.4 (10–10.8) | 12.7 (12.2–13.1) |

**eTable 52. Estimates for marginal predictions of annual social welfare benefits among parents exposed to child's intellectual disability (F70–F79) and unexposed parents stratified by the child's age at diagnosis**

| | | Finland | | | | Denmark | | | |
|---|---|---|---|---|---|---|---|---|---|
| | | Men | | Women | | Men | | Women | |
| | | Unexposed | Exposed | Unexposed | Exposed | Unexposed | Exposed | Unexposed | Exposed |
| Years since diagnosis | Age group | Marg. pred. (95% CI) | Marg. pred. (95% CI) | Marg. pred. (95% CI) | Marg. pred. (95% CI) | Marg. pred. (95% CI) | Marg. pred. (95% CI) | Marg. pred. (95% CI) | Marg. pred. (95% CI) |
| | | Unexposed | Exposed | Unexposed | Exposed | Unexposed | Exposed | Unexposed | Exposed |
| 5 | | 4.7 (4.4–5.1) | 6 (5.6–6.5) | 6.6 (6.2–6.9) | 7.4 (7.1–7.7) | 6 (5.6–6.4) | 7.9 (7.5–8.3) | 10 (9.6–10.4) | 12.3 (11.8–12.7) |
| -5 | 15–19 years | 3.5 (3.2–3.8) | 4.5 (4.2–4.8) | 7.8 (7.5–8.1) | 9.1 (8.8–9.5) | 5.3 (5–5.6) | 7.1 (6.7–7.4) | 11.4 (11–11.7) | 14 (13.6–14.4) |
| -4 | | 3.5 (3.2–3.8) | 4.5 (4.2–4.8) | 7.6 (7.3–7.9) | 9.1 (8.7–9.4) | 5.3 (4.9–5.6) | 7.2 (6.9–7.6) | 11.2 (10.8–11.6) | 13.8 (13.4–14.3) |
| -3 | | 3.8 (3.4–4.1) | 4.8 (4.4–5.1) | 7.5 (7.2–7.8) | 8.9 (8.5–9.2) | 5.4 (5.1–5.7) | 7.5 (7.1–7.9) | 11.1 (10.7–11.5) | 13.9 (13.5–14.3) |
| -2 | | 3.9 (3.5–4.2) | 5 (4.7–5.4) | 7.2 (6.9–7.5) | 8.5 (8.2–8.9) | 5.6 (5.2–5.9) | 7.7 (7.3–8.1) | 10.9 (10.5–11.3) | 13.9 (13.5–14.3) |
| -1 | | 4 (3.7–4.3) | 5.2 (4.9–5.6) | 6.9 (6.5–7.2) | 8.2 (7.8–8.5) | 5.7 (5.4–6.1) | 7.7 (7.3–8.1) | 10.7 (10.3–11.1) | 13.8 (13.4–14.3) |
| 0 | | 4.1 (3.8–4.4) | 5.4 (5–5.8) | 6.4 (6.1–6.7) | 7.8 (7.4–8.1) | 5.7 (5.4–6.1) | 8 (7.6–8.4) | 10.3 (9.9–10.7) | 13.5 (13.1–13.9) |
| 1 | | 4.4 (4–4.8) | 5.5 (5.2–5.9) | 5.9 (5.6–6.2) | 7.2 (6.9–7.6) | 5.8 (5.4–6.1) | 8.1 (7.7–8.5) | 9.9 (9.5–10.3) | 13.1 (12.7–13.6) |
| 2 | | 4.6 (4.2–5) | 5.7 (5.3–6.1) | 5.4 (5.1–5.8) | 6.7 (6.4–7.1) | 6.1 (5.7–6.5) | 8.2 (7.8–8.6) | 9.4 (9–9.8) | 12.7 (12.2–13.1) |
| 3 | | 5 (4.5–5.4) | 6.2 (5.7–6.6) | 5 (4.7–5.3) | 6.6 (6.3–7) | 6.1 (5.8–6.5) | 8.2 (7.8–8.6) | 9.1 (8.7–9.5) | 12.1 (11.7–12.6) |

**eTable 52. Estimates for marginal predictions of annual social welfare benefits among parents exposed to child's intellectual disability (F70–F79) and unexposed parents stratified by the child's age at diagnosis**

| | | Finland | | | | Denmark | | | |
|---|---|---|---|---|---|---|---|---|---|
| | | Men | | Women | | Men | | Women | |
| | | Unexposed | Exposed | Unexposed | Exposed | Unexposed | Exposed | Unexposed | Exposed |
| Years since diagnosis | Age group | Marg. pred. (95% CI) | Marg. pred. (95% CI) | Marg. pred. (95% CI) | Marg. pred. (95% CI) | Marg. pred. (95% CI) | Marg. pred. (95% CI) | Marg. pred. (95% CI) | Marg. pred. (95% CI) |
| | | Unexposed | Exposed | Unexposed | Exposed | Unexposed | Exposed | Unexposed | Exposed |
| 4 | | 5.4 (4.9–5.8) | 6.4 (5.9–6.9) | 4.8 (4.5–5.2) | 6.7 (6.3–7.1) | 6.3 (5.9–6.7) | 8.7 (8.3–9.2) | 9 (8.5–9.4) | 12.1 (11.6–12.5) |
| 5 | | 5.8 (5.3–6.3) | 7 (6.5–7.5) | 4.9 (4.5–5.3) | 6.7 (6.3–7.1) | 6.7 (6.2–7.1) | 9 (8.6–9.5) | 9.1 (8.6–9.5) | 12.4 (11.9–12.9) |
| -5 | 20–25 years | 4 (3.5–4.4) | 5.6 (5–6.2) | 5.7 (5.3–6.1) | 6.6 (6.2–7.1) | 5.5 (5.1–5.9) | 7.7 (7.3–8.2) | 9.3 (8.9–9.8) | 12 (11.5–12.4) |
| -4 | | 4 (3.5–4.5) | 6 (5.4–6.6) | 5.5 (5–5.9) | 6.4 (6–6.8) | 5.6 (5.2–6) | 8.1 (7.6–8.6) | 9.1 (8.7–9.6) | 11.7 (11.3–12.2) |
| -3 | | 4.3 (3.8–4.8) | 6.1 (5.5–6.7) | 5.1 (4.7–5.5) | 6.4 (6–6.8) | 5.9 (5.4–6.3) | 8.2 (7.7–8.7) | 9 (8.5–9.4) | 11.4 (10.9–11.9) |
| -2 | | 4.6 (4–5.1) | 6.6 (6–7.2) | 5 (4.6–5.5) | 6.5 (6–6.9) | 5.8 (5.4–6.2) | 8.5 (8–9) | 8.7 (8.2–9.1) | 11.1 (10.6–11.6) |
| -1 | | 5 (4.5–5.6) | 6.7 (6.1–7.3) | 5.1 (4.7–5.5) | 6.2 (5.7–6.6) | 6 (5.6–6.5) | 8.7 (8.3–9.2) | 8.4 (8–8.9) | 11.2 (10.7–11.7) |
| 0 | | 5.3 (4.8–5.9) | 7.1 (6.5–7.7) | 5 (4.6–5.4) | 6.3 (5.9–6.7) | 6.3 (5.9–6.8) | 9 (8.5–9.5) | 8.3 (7.9–8.8) | 11.1 (10.6–11.6) |
| 1 | | 6 (5.4–6.6) | 7.7 (7–8.4) | 5.3 (4.9–5.8) | 6.7 (6.3–7.2) | 6.5 (6.1–7) | 9.2 (8.7–9.7) | 8.4 (7.9–8.9) | 11.3 (10.8–11.8) |
| 2 | | 6.8 (6.2–7.5) | 8.3 (7.6–9) | 5.3 (4.9–5.8) | 6.8 (6.4–7.3) | 7 (6.6–7.5) | 9.6 (9.1–10.1) | 8.4 (7.9–8.9) | 11.6 (11.1–12.2) |

**eTable 52. Estimates for marginal predictions of annual social welfare benefits among parents exposed to child's intellectual disability (F70–F79) and unexposed parents stratified by the child's age at diagnosis**

| | | Finland | | | | Denmark | | | |
|---|---|---|---|---|---|---|---|---|---|
| | | Men | | Women | | Men | | Women | |
| | | Unexposed | Exposed | Unexposed | Exposed | Unexposed | Exposed | Unexposed | Exposed |
| Years since diagnosis | Age group | Marg. pred. (95% CI) | Marg. pred. (95% CI) | Marg. pred. (95% CI) | Marg. pred. (95% CI) | Marg. pred. (95% CI) | Marg. pred. (95% CI) | Marg. pred. (95% CI) | Marg. pred. (95% CI) |
| | | Unexposed | Exposed | Unexposed | Exposed | Unexposed | Exposed | Unexposed | Exposed |
| 3 | | 7.5 (6.8–8.2) | 9 (8.3–9.8) | 5.7 (5.2–6.2) | 7.1 (6.6–7.6) | 7.6 (7.1–8.1) | 10 (9.5–10.6) | 8.6 (8.1–9.1) | 12.2 (11.6–12.7) |
| 4 | | 8.5 (7.7–9.3) | 10 (9.2–10.8) | 5.8 (5.3–6.3) | 7.4 (6.8–7.9) | 7.9 (7.3–8.4) | 10.3 (9.8–10.9) | 8.9 (8.4–9.5) | 12.6 (12–13.1) |
| 5 | | 9.8 (8.9–10.7) | 10.8 (9.9–11.7) | 6.3 (5.7–6.8) | 7.6 (7.1–8.2) | 8.2 (7.7–8.8) | 11.3 (10.7–11.9) | 9.3 (8.7–9.8) | 12.8 (12.3–13.4) |

Marg. pred. = Marginal predictions in thousand euros (€)

**eTable 53. Estimates for marginal predictions of annual employment among parents exposed to child's intellectual disability (F70–F79) and unexposed parents stratified by the child's age at diagnosis**

| | | Finland | | | | Denmark | | | |
|---|---|---|---|---|---|---|---|---|---|
| | | Men | | Women | | Men | | Women | |
| | | Unexposed | Exposed | Unexposed | Exposed | Unexposed | Exposed | Unexposed | Exposed |
| Years since diagnosis | Age group | Marg. pred. (95% CI) | Marg. pred. (95% CI) | Marg. pred. (95% CI) | Marg. pred. (95% CI) | Marg. pred. (95% CI) | Marg. pred. (95% CI) | Marg. pred. (95% CI) | Marg. pred. (95% CI) |
| | | Unexposed | Exposed | Unexposed | Exposed | Unexposed | Exposed | Unexposed | Exposed |
| -5 | 1–4 years | 0.76 (0.75–0.78) | 0.76 (0.74–0.77) | 0.59 (0.57–0.6) | 0.56 (0.55–0.58) | 0.69 (0.67–0.72) | 0.62 (0.6–0.64) | 0.55 (0.53–0.58) | 0.45 (0.43–0.48) |
| -4 | | 0.79 (0.78–0.81) | 0.77 (0.75–0.78) | 0.56 (0.54–0.57) | 0.52 (0.5–0.53) | 0.72 (0.7–0.74) | 0.65 (0.63–0.67) | 0.54 (0.52–0.56) | 0.43 (0.41–0.46) |
| -3 | | 0.8 (0.79–0.81) | 0.76 (0.74–0.77) | 0.53 (0.52–0.55) | 0.48 (0.47–0.5) | 0.73 (0.71–0.75) | 0.67 (0.65–0.69) | 0.54 (0.52–0.56) | 0.45 (0.43–0.47) |
| -2 | | 0.81 (0.79–0.82) | 0.76 (0.75–0.78) | 0.54 (0.53–0.56) | 0.48 (0.46–0.5) | 0.73 (0.71–0.75) | 0.67 (0.65–0.69) | 0.57 (0.55–0.6) | 0.47 (0.45–0.49) |
| -1 | | 0.82 (0.81–0.83) | 0.76 (0.75–0.77) | 0.57 (0.55–0.59) | 0.49 (0.47–0.51) | 0.73 (0.71–0.75) | 0.69 (0.67–0.71) | 0.6 (0.57–0.62) | 0.49 (0.46–0.51) |
| 0 | | 0.82 (0.81–0.83) | 0.77 (0.76–0.79) | 0.61 (0.59–0.62) | 0.53 (0.51–0.55) | 0.74 (0.72–0.76) | 0.68 (0.65–0.7) | 0.62 (0.6–0.64) | 0.51 (0.48–0.53) |
| 1 | | 0.83 (0.81–0.84) | 0.78 (0.76–0.79) | 0.66 (0.64–0.68) | 0.57 (0.56–0.59) | 0.73 (0.71–0.75) | 0.66 (0.64–0.69) | 0.63 (0.61–0.65) | 0.52 (0.5–0.54) |
| 2 | | 0.83 (0.82–0.84) | 0.77 (0.76–0.79) | 0.68 (0.66–0.69) | 0.61 (0.59–0.62) | 0.74 (0.72–0.76) | 0.65 (0.63–0.67) | 0.65 (0.63–0.67) | 0.55 (0.52–0.57) |
| 3 | | 0.82 (0.81–0.83) | 0.78 (0.76–0.79) | 0.71 (0.7–0.73) | 0.62 (0.6–0.63) | 0.73 (0.71–0.75) | 0.66 (0.64–0.69) | 0.67 (0.65–0.69) | 0.56 (0.54–0.58) |

**eTable 53. Estimates for marginal predictions of annual employment among parents exposed to child's intellectual disability (F70–F79) and unexposed parents stratified by the child's age at diagnosis**

| | | Finland | | | | Denmark | | | |
|---|---|---|---|---|---|---|---|---|---|
| | | Men | | Women | | Men | | Women | |
| | | Unexposed | Exposed | Unexposed | Exposed | Unexposed | Exposed | Unexposed | Exposed |
| Years since diagnosis | Age group | Marg. pred. (95% CI) | Marg. pred. (95% CI) | Marg. pred. (95% CI) | Marg. pred. (95% CI) | Marg. pred. (95% CI) | Marg. pred. (95% CI) | Marg. pred. (95% CI) | Marg. pred. (95% CI) |
| | | Unexposed | Exposed | Unexposed | Exposed | Unexposed | Exposed | Unexposed | Exposed |
| 4 | | 0.83 (0.81–0.84) | 0.77 (0.76–0.79) | 0.73 (0.72–0.75) | 0.64 (0.62–0.65) | 0.73 (0.71–0.75) | 0.66 (0.64–0.69) | 0.67 (0.65–0.7) | 0.57 (0.55–0.6) |
| 5 | | 0.82 (0.8–0.83) | 0.78 (0.76–0.79) | 0.75 (0.74–0.77) | 0.65 (0.64–0.67) | 0.73 (0.71–0.75) | 0.64 (0.62–0.67) | 0.68 (0.65–0.7) | 0.57 (0.55–0.59) |
| -5 | 5–9 years | 0.81 (0.8–0.82) | 0.73 (0.72–0.74) | 0.52 (0.51–0.53) | 0.45 (0.44–0.47) | 0.68 (0.67–0.7) | 0.67 (0.65–0.68) | 0.54 (0.52–0.55) | 0.46 (0.45–0.48) |
| -4 | | 0.81 (0.8–0.82) | 0.73 (0.72–0.74) | 0.57 (0.55–0.58) | 0.48 (0.46–0.49) | 0.7 (0.69–0.72) | 0.68 (0.66–0.69) | 0.57 (0.56–0.59) | 0.5 (0.48–0.52) |
| -3 | | 0.8 (0.79–0.82) | 0.72 (0.71–0.73) | 0.61 (0.59–0.62) | 0.51 (0.5–0.53) | 0.71 (0.69–0.72) | 0.68 (0.67–0.7) | 0.59 (0.57–0.6) | 0.51 (0.5–0.53) |
| -2 | | 0.81 (0.8–0.82) | 0.73 (0.71–0.74) | 0.64 (0.63–0.65) | 0.54 (0.53–0.55) | 0.72 (0.7–0.73) | 0.68 (0.66–0.69) | 0.61 (0.59–0.62) | 0.53 (0.51–0.54) |
| -1 | | 0.81 (0.8–0.82) | 0.73 (0.72–0.74) | 0.66 (0.65–0.67) | 0.55 (0.54–0.57) | 0.71 (0.7–0.73) | 0.67 (0.66–0.69) | 0.62 (0.6–0.64) | 0.53 (0.52–0.55) |
| 0 | | 0.81 (0.8–0.82) | 0.73 (0.72–0.75) | 0.68 (0.67–0.7) | 0.58 (0.57–0.59) | 0.71 (0.69–0.72) | 0.67 (0.66–0.69) | 0.63 (0.61–0.65) | 0.53 (0.51–0.54) |
| 1 | | 0.81 (0.8–0.82) | 0.74 (0.73–0.75) | 0.7 (0.69–0.71) | 0.59 (0.58–0.6) | 0.71 (0.7–0.73) | 0.67 (0.65–0.68) | 0.64 (0.62–0.66) | 0.53 (0.51–0.55) |

**eTable 53. Estimates for marginal predictions of annual employment among parents exposed to child's intellectual disability (F70–F79) and unexposed parents stratified by the child's age at diagnosis**

| | | Finland | | | | Denmark | | | |
|---|---|---|---|---|---|---|---|---|---|
| | | Men | | Women | | Men | | Women | |
| | | Unexposed | Exposed | Unexposed | Exposed | Unexposed | Exposed | Unexposed | Exposed |
| Years since diagnosis | Age group | Marg. pred. (95% CI) | Marg. pred. (95% CI) | Marg. pred. (95% CI) | Marg. pred. (95% CI) | Marg. pred. (95% CI) | Marg. pred. (95% CI) | Marg. pred. (95% CI) | Marg. pred. (95% CI) |
| | | Unexposed | Exposed | Unexposed | Exposed | Unexposed | Exposed | Unexposed | Exposed |
| 2 | | 0.81 (0.8–0.82) | 0.74 (0.73–0.75) | 0.72 (0.71–0.73) | 0.6 (0.59–0.62) | 0.71 (0.7–0.73) | 0.65 (0.63–0.67) | 0.65 (0.63–0.67) | 0.53 (0.52–0.55) |
| 3 | | 0.81 (0.8–0.82) | 0.74 (0.72–0.75) | 0.74 (0.73–0.75) | 0.62 (0.61–0.64) | 0.71 (0.69–0.72) | 0.65 (0.63–0.66) | 0.65 (0.64–0.67) | 0.54 (0.52–0.56) |
| 4 | | 0.8 (0.79–0.81) | 0.74 (0.72–0.75) | 0.76 (0.75–0.77) | 0.63 (0.61–0.64) | 0.7 (0.68–0.71) | 0.65 (0.64–0.67) | 0.66 (0.64–0.67) | 0.54 (0.53–0.56) |
| 5 | | 0.8 (0.79–0.81) | 0.73 (0.72–0.74) | 0.77 (0.76–0.78) | 0.65 (0.64–0.67) | 0.69 (0.67–0.7) | 0.65 (0.63–0.66) | 0.66 (0.64–0.67) | 0.54 (0.53–0.56) |
| -5 | 10–14 years | 0.81 (0.79–0.82) | 0.75 (0.73–0.76) | 0.69 (0.67–0.71) | 0.61 (0.59–0.63) | 0.69 (0.68–0.71) | 0.65 (0.64–0.67) | 0.59 (0.57–0.61) | 0.53 (0.51–0.54) |
| -4 | | 0.8 (0.79–0.82) | 0.74 (0.73–0.76) | 0.72 (0.7–0.73) | 0.61 (0.59–0.63) | 0.7 (0.68–0.71) | 0.65 (0.64–0.67) | 0.61 (0.6–0.63) | 0.54 (0.53–0.56) |
| -3 | | 0.79 (0.77–0.8) | 0.74 (0.72–0.76) | 0.72 (0.71–0.74) | 0.63 (0.61–0.65) | 0.7 (0.68–0.71) | 0.65 (0.64–0.67) | 0.62 (0.61–0.64) | 0.55 (0.53–0.56) |
| -2 | | 0.79 (0.77–0.8) | 0.73 (0.71–0.75) | 0.73 (0.72–0.75) | 0.64 (0.62–0.66) | 0.69 (0.68–0.71) | 0.65 (0.64–0.67) | 0.63 (0.62–0.65) | 0.56 (0.54–0.58) |
| -1 | | 0.79 (0.77–0.8) | 0.73 (0.71–0.75) | 0.75 (0.74–0.77) | 0.64 (0.62–0.66) | 0.69 (0.67–0.7) | 0.65 (0.63–0.66) | 0.64 (0.62–0.65) | 0.56 (0.55–0.58) |

**eTable 53. Estimates for marginal predictions of annual employment among parents exposed to child's intellectual disability (F70–F79) and unexposed parents stratified by the child's age at diagnosis**

| | | Finland | | | | Denmark | | | |
|---|---|---|---|---|---|---|---|---|---|
| | | Men | | Women | | Men | | Women | |
| | | Unexposed | Exposed | Unexposed | Exposed | Unexposed | Exposed | Unexposed | Exposed |
| Years since diagnosis | Age group | Marg. pred. (95% CI) | Marg. pred. (95% CI) | Marg. pred. (95% CI) | Marg. pred. (95% CI) | Marg. pred. (95% CI) | Marg. pred. (95% CI) | Marg. pred. (95% CI) | Marg. pred. (95% CI) |
| | | Unexposed | Exposed | Unexposed | Exposed | Unexposed | Exposed | Unexposed | Exposed |
| 0 | | 0.78 (0.76–0.79) | 0.72 (0.7–0.73) | 0.75 (0.73–0.77) | 0.65 (0.63–0.66) | 0.68 (0.67–0.7) | 0.64 (0.63–0.66) | 0.65 (0.63–0.66) | 0.56 (0.54–0.58) |
| 1 | | 0.77 (0.75–0.79) | 0.71 (0.69–0.73) | 0.76 (0.74–0.78) | 0.66 (0.64–0.68) | 0.69 (0.67–0.7) | 0.63 (0.62–0.65) | 0.64 (0.62–0.66) | 0.56 (0.55–0.58) |
| 2 | | 0.77 (0.75–0.79) | 0.7 (0.68–0.72) | 0.78 (0.76–0.79) | 0.66 (0.64–0.68) | 0.69 (0.67–0.7) | 0.63 (0.61–0.64) | 0.65 (0.63–0.66) | 0.56 (0.54–0.58) |
| 3 | | 0.77 (0.75–0.78) | 0.7 (0.68–0.72) | 0.77 (0.75–0.79) | 0.66 (0.64–0.68) | 0.68 (0.67–0.7) | 0.62 (0.6–0.64) | 0.66 (0.64–0.67) | 0.56 (0.55–0.58) |
| 4 | | 0.76 (0.75–0.78) | 0.68 (0.66–0.7) | 0.79 (0.77–0.81) | 0.67 (0.65–0.69) | 0.67 (0.65–0.69) | 0.61 (0.59–0.63) | 0.64 (0.63–0.66) | 0.56 (0.55–0.58) |
| 5 | | 0.76 (0.74–0.77) | 0.67 (0.65–0.69) | 0.78 (0.77–0.8) | 0.67 (0.65–0.69) | 0.67 (0.65–0.69) | 0.61 (0.59–0.63) | 0.64 (0.63–0.66) | 0.56 (0.54–0.58) |
| -5 | 15–19 years | 0.79 (0.77–0.81) | 0.73 (0.71–0.75) | 0.74 (0.72–0.76) | 0.65 (0.63–0.67) | 0.68 (0.66–0.69) | 0.61 (0.6–0.63) | 0.6 (0.59–0.62) | 0.52 (0.51–0.54) |
| -4 | | 0.79 (0.77–0.81) | 0.71 (0.69–0.73) | 0.75 (0.73–0.77) | 0.65 (0.63–0.67) | 0.68 (0.67–0.7) | 0.62 (0.6–0.64) | 0.63 (0.61–0.64) | 0.54 (0.52–0.55) |
| -3 | | 0.77 (0.75–0.79) | 0.71 (0.69–0.73) | 0.74 (0.72–0.76) | 0.66 (0.63–0.68) | 0.7 (0.68–0.72) | 0.63 (0.61–0.65) | 0.64 (0.62–0.65) | 0.53 (0.51–0.54) |

**eTable 53. Estimates for marginal predictions of annual employment among parents exposed to child's intellectual disability (F70–F79) and unexposed parents stratified by the child's age at diagnosis**

| | | Finland | | | | Denmark | | | |
|---|---|---|---|---|---|---|---|---|---|
| | | Men | | Women | | Men | | Women | |
| | | Unexposed | Exposed | Unexposed | Exposed | Unexposed | Exposed | Unexposed | Exposed |
| Years since diagnosis | Age group | Marg. pred. (95% CI) | Marg. pred. (95% CI) | Marg. pred. (95% CI) | Marg. pred. (95% CI) | Marg. pred. (95% CI) | Marg. pred. (95% CI) | Marg. pred. (95% CI) | Marg. pred. (95% CI) |
| | | Unexposed | Exposed | Unexposed | Exposed | Unexposed | Exposed | Unexposed | Exposed |
| -2 | | 0.77 (0.75–0.79) | 0.7 (0.68–0.72) | 0.76 (0.74–0.78) | 0.65 (0.63–0.68) | 0.69 (0.67–0.71) | 0.62 (0.6–0.64) | 0.64 (0.63–0.66) | 0.53 (0.52–0.55) |
| -1 | | 0.77 (0.75–0.79) | 0.69 (0.67–0.71) | 0.76 (0.74–0.78) | 0.66 (0.64–0.68) | 0.69 (0.68–0.71) | 0.62 (0.6–0.64) | 0.64 (0.63–0.66) | 0.52 (0.51–0.54) |
| 0 | | 0.77 (0.75–0.79) | 0.69 (0.67–0.71) | 0.78 (0.76–0.8) | 0.65 (0.63–0.67) | 0.69 (0.67–0.71) | 0.61 (0.59–0.63) | 0.65 (0.63–0.66) | 0.52 (0.5–0.53) |
| 1 | | 0.76 (0.74–0.78) | 0.67 (0.65–0.7) | 0.78 (0.76–0.8) | 0.66 (0.64–0.68) | 0.68 (0.66–0.7) | 0.61 (0.59–0.63) | 0.64 (0.62–0.66) | 0.52 (0.5–0.54) |
| 2 | | 0.73 (0.71–0.75) | 0.66 (0.63–0.68) | 0.77 (0.75–0.79) | 0.65 (0.63–0.67) | 0.67 (0.65–0.68) | 0.6 (0.58–0.62) | 0.64 (0.62–0.66) | 0.51 (0.5–0.53) |
| 3 | | 0.72 (0.69–0.74) | 0.65 (0.63–0.67) | 0.77 (0.75–0.79) | 0.66 (0.63–0.68) | 0.67 (0.65–0.68) | 0.59 (0.57–0.61) | 0.63 (0.61–0.65) | 0.52 (0.5–0.53) |
| 4 | | 0.7 (0.68–0.72) | 0.64 (0.61–0.66) | 0.77 (0.75–0.79) | 0.63 (0.61–0.66) | 0.66 (0.64–0.68) | 0.57 (0.55–0.59) | 0.62 (0.6–0.64) | 0.51 (0.49–0.53) |
| 5 | | 0.69 (0.66–0.71) | 0.61 (0.58–0.63) | 0.77 (0.74–0.79) | 0.64 (0.62–0.67) | 0.65 (0.63–0.67) | 0.57 (0.55–0.59) | 0.61 (0.59–0.63) | 0.5 (0.48–0.52) |
| -5 | 20–25 years | 0.78 (0.75–0.8) | 0.65 (0.62–0.68) | 0.76 (0.74–0.79) | 0.61 (0.58–0.64) | 0.66 (0.64–0.68) | 0.57 (0.55–0.59) | 0.62 (0.6–0.64) | 0.51 (0.49–0.53) |

**eTable 53. Estimates for marginal predictions of annual employment among parents exposed to child's intellectual disability (F70–F79) and unexposed parents stratified by the child's age at diagnosis**

| | | Finland | | | | Denmark | | | |
|---|---|---|---|---|---|---|---|---|---|
| | | Men | | Women | | Men | | Women | |
| | | Unexposed | Exposed | Unexposed | Exposed | Unexposed | Exposed | Unexposed | Exposed |
| Years since diagnosis | Age group | Marg. pred. (95% CI) | Marg. pred. (95% CI) | Marg. pred. (95% CI) | Marg. pred. (95% CI) | Marg. pred. (95% CI) | Marg. pred. (95% CI) | Marg. pred. (95% CI) | Marg. pred. (95% CI) |
| | | Unexposed | Exposed | Unexposed | Exposed | Unexposed | Exposed | Unexposed | Exposed |
| -4 | | 0.74 (0.71–0.77) | 0.63 (0.6–0.66) | 0.76 (0.73–0.79) | 0.6 (0.57–0.63) | 0.68 (0.66–0.7) | 0.58 (0.56–0.6) | 0.63 (0.61–0.65) | 0.51 (0.49–0.53) |
| -3 | | 0.76 (0.73–0.79) | 0.62 (0.59–0.65) | 0.76 (0.73–0.78) | 0.59 (0.56–0.62) | 0.68 (0.66–0.7) | 0.58 (0.56–0.61) | 0.63 (0.61–0.65) | 0.52 (0.5–0.54) |
| -2 | | 0.73 (0.7–0.76) | 0.6 (0.57–0.64) | 0.75 (0.72–0.78) | 0.59 (0.56–0.62) | 0.68 (0.66–0.7) | 0.57 (0.55–0.6) | 0.63 (0.61–0.65) | 0.51 (0.49–0.53) |
| -1 | | 0.71 (0.68–0.74) | 0.59 (0.56–0.62) | 0.73 (0.7–0.76) | 0.59 (0.56–0.62) | 0.67 (0.65–0.69) | 0.57 (0.54–0.59) | 0.62 (0.6–0.65) | 0.51 (0.48–0.53) |
| 0 | | 0.71 (0.68–0.74) | 0.58 (0.54–0.61) | 0.72 (0.7–0.75) | 0.57 (0.54–0.6) | 0.67 (0.65–0.69) | 0.55 (0.53–0.57) | 0.62 (0.6–0.64) | 0.51 (0.48–0.53) |
| 1 | | 0.68 (0.65–0.71) | 0.56 (0.53–0.59) | 0.71 (0.68–0.74) | 0.55 (0.52–0.59) | 0.66 (0.63–0.68) | 0.54 (0.52–0.57) | 0.62 (0.59–0.64) | 0.49 (0.47–0.52) |
| 2 | | 0.63 (0.6–0.67) | 0.53 (0.49–0.56) | 0.7 (0.67–0.73) | 0.53 (0.5–0.56) | 0.64 (0.62–0.66) | 0.53 (0.51–0.55) | 0.61 (0.59–0.64) | 0.48 (0.46–0.51) |
| 3 | | 0.62 (0.59–0.66) | 0.51 (0.47–0.54) | 0.68 (0.65–0.71) | 0.53 (0.49–0.56) | 0.61 (0.58–0.63) | 0.52 (0.49–0.54) | 0.6 (0.58–0.62) | 0.46 (0.44–0.48) |
| 4 | | 0.58 (0.54–0.61) | 0.46 (0.43–0.5) | 0.68 (0.65–0.71) | 0.51 (0.48–0.55) | 0.59 (0.56–0.61) | 0.5 (0.47–0.52) | 0.59 (0.56–0.61) | 0.45 (0.42–0.47) |

**eTable 53. Estimates for marginal predictions of annual employment among parents exposed to child's intellectual disability (F70–F79) and unexposed parents stratified by the child's age at diagnosis**

| | | Finland | | | | Denmark | | | |
|---|---|---|---|---|---|---|---|---|---|
| | | Men | | Women | | Men | | Women | |
| | | Unexposed | Exposed | Unexposed | Exposed | Unexposed | Exposed | Unexposed | Exposed |
| Years since diagnosis | Age group | Marg. pred. (95% CI) | Marg. pred. (95% CI) | Marg. pred. (95% CI) | Marg. pred. (95% CI) | Marg. pred. (95% CI) | Marg. pred. (95% CI) | Marg. pred. (95% CI) | Marg. pred. (95% CI) |
| | | Unexposed | Exposed | Unexposed | Exposed | Unexposed | Exposed | Unexposed | Exposed |
| 5 | | 0.54 (0.5–0.58) | 0.44 (0.4–0.48) | 0.65 (0.62–0.69) | 0.5 (0.47–0.54) | 0.57 (0.55–0.6) | 0.47 (0.44–0.5) | 0.56 (0.53–0.58) | 0.43 (0.41–0.46) |

Marg. pred. = Marginal predictions in probabilities

**eTable 54. Estimates for marginal predictions of annual earnings among parents exposed to child's developmental disorder (F84) and unexposed parents stratified by the child's age at diagnosis**

| | | Finland | | | | Denmark | | | |
|---|---|---|---|---|---|---|---|---|---|
| | | Men | | Women | | Men | | Women | |
| | | Unexposed | Exposed | Unexposed | Exposed | Unexposed | Exposed | Unexposed | Exposed |
| Years since diagnosis | Age group | Marg. pred. (95% CI) | Marg. pred. (95% CI) | Marg. pred. (95% CI) | Marg. pred. (95% CI) | Marg. pred. (95% CI) | Marg. pred. (95% CI) | Marg. pred. (95% CI) | Marg. pred. (95% CI) |
| | | Unexposed | Exposed | Unexposed | Exposed | Unexposed | Exposed | Unexposed | Exposed |
| -5 | 1–4 years | 27.6 (27–28.3) | 24.4 (23.8–25) | 14.2 (13.8–14.7) | 12.7 (12.3–13.2) | 47.4 (46.6–48.3) | 44.2 (43.4–44.9) | 35.1 (34.6–35.6) | 32.4 (31.9–32.9) |
| -4 | | 28.8 (28.2–29.5) | 25.8 (25.1–26.4) | 12.2 (11.7–12.6) | 10.3 (9.8–10.7) | 49.3 (48.5–50.1) | 45.9 (45.2–46.7) | 36.1 (35.6–36.5) | 33 (32.5–33.5) |
| -3 | | 30 (29.4–30.7) | 26.6 (25.9–27.3) | 10.2 (9.8–10.6) | 8.1 (7.7–8.5) | 51.6 (50.7–52.4) | 47.3 (46.5–48) | 37.7 (37.3–38.2) | 34.6 (34.1–35.1) |
| -2 | | 31.3 (30.6–32) | 27.7 (27–28.4) | 10.8 (10.3–11.2) | 7.7 (7.3–8) | 53.5 (52.6–54.4) | 49.6 (48.8–50.4) | 39.6 (39.1–40.1) | 36.3 (35.8–36.8) |
| -1 | | 32.9 (32.2–33.6) | 29 (28.3–29.7) | 13.4 (12.9–13.9) | 9.7 (9.3–10.2) | 55.5 (54.6–56.4) | 51.2 (50.3–52) | 41.3 (40.8–41.8) | 37.6 (37.1–38.1) |
| 0 | | 34.5 (33.8–35.3) | 30.1 (29.4–30.9) | 16 (15.5–16.5) | 12 (11.5–12.5) | 56.7 (55.8–57.7) | 52.1 (51.3–53) | 42.5 (42–43) | 38.9 (38.4–39.4) |
| 1 | | 35.6 (34.8–36.4) | 31.2 (30.4–32) | 17.9 (17.4–18.4) | 14.1 (13.6–14.6) | 58.3 (57.3–59.4) | 53.4 (52.5–54.4) | 43.7 (43.1–44.2) | 40.6 (40.1–41.2) |
| 2 | | 36.5 (35.7–37.3) | 31.9 (31.1–32.7) | 19.8 (19.3–20.4) | 16 (15.5–16.6) | 59.5 (58.4–60.6) | 54.4 (53.4–55.3) | 44.9 (44.4–45.5) | 41.9 (41.3–42.4) |
| 3 | | 37.5 (36.7–38.4) | 32.4 (31.5–33.2) | 21.1 (20.5–21.7) | 17.4 (16.8–17.9) | 60.8 (59.6–61.9) | 55.3 (54.3–56.2) | 45.8 (45.2–46.4) | 42.7 (42.1–43.2) |

**eTable 54. Estimates for marginal predictions of annual earnings among parents exposed to child's developmental disorder (F84) and unexposed parents stratified by the child's age at diagnosis**

| | | Finland | | | | Denmark | | | |
|---|---|---|---|---|---|---|---|---|---|
| | | Men | | Women | | Men | | Women | |
| | | Unexposed | Exposed | Unexposed | Exposed | Unexposed | Exposed | Unexposed | Exposed |
| Years since diagnosis | Age group | Marg. pred. (95% CI) | Marg. pred. (95% CI) | Marg. pred. (95% CI) | Marg. pred. (95% CI) | Marg. pred. (95% CI) | Marg. pred. (95% CI) | Marg. pred. (95% CI) | Marg. pred. (95% CI) |
| | | Unexposed | Exposed | Unexposed | Exposed | Unexposed | Exposed | Unexposed | Exposed |
| 4 | | 38.3 (37.4–39.2) | 33.3 (32.4–34.2) | 22.2 (21.6–22.8) | 18.5 (18–19.1) | 61.8 (60.6–63) | 56.2 (55.1–57.2) | 46.9 (46.3–47.6) | 43.4 (42.8–44) |
| 5 | | 38.8 (37.9–39.7) | 34.6 (33.6–35.5) | 23.3 (22.7–24) | 19.6 (19–20.2) | 63.2 (61.9–64.4) | 56.5 (55.4–57.6) | 47.6 (47–48.3) | 44.2 (43.6–44.9) |
| -5 | 5–9 years | 31.7 (31.1–32.2) | 29.3 (28.7–29.8) | 12.9 (12.6–13.3) | 11.2 (10.9–11.6) | 52.6 (52–53.1) | 50.7 (50.2–51.2) | 39.6 (39.3–39.9) | 38.1 (37.8–38.4) |
| -4 | | 32.9 (32.3–33.4) | 30.4 (29.9–31) | 15 (14.6–15.4) | 13 (12.7–13.4) | 53.9 (53.4–54.5) | 51.7 (51.2–52.2) | 41.2 (40.9–41.5) | 39.7 (39.4–40) |
| -3 | | 34.1 (33.5–34.7) | 31.3 (30.7–31.9) | 16.9 (16.5–17.3) | 14.7 (14.3–15.1) | 55.3 (54.7–55.9) | 52.8 (52.3–53.3) | 42.6 (42.3–42.9) | 40.9 (40.6–41.2) |
| -2 | | 35 (34.4–35.6) | 32 (31.4–32.6) | 18.4 (18–18.9) | 16.2 (15.8–16.6) | 56.4 (55.8–57) | 54.3 (53.7–54.8) | 43.6 (43.3–44) | 41.8 (41.5–42.1) |
| -1 | | 35.8 (35.2–36.5) | 33 (32.4–33.6) | 19.9 (19.5–20.3) | 17.4 (17–17.8) | 57.7 (57.1–58.3) | 55.6 (55.1–56.2) | 44.8 (44.4–45.1) | 42.7 (42.4–43) |
| 0 | | 36.9 (36.2–37.5) | 33.7 (33–34.3) | 21.2 (20.8–21.6) | 18.6 (18.2–19) | 58.5 (57.9–59.2) | 56.7 (56.1–57.2) | 45.7 (45.3–46) | 43.4 (43.1–43.7) |
| 1 | | 37.6 (36.9–38.3) | 34.1 (33.5–34.8) | 22.6 (22.1–23) | 19.8 (19.4–20.3) | 59.9 (59.3–60.6) | 57.4 (56.8–57.9) | 46.5 (46.1–46.8) | 44.4 (44.1–44.8) |

**eTable 54. Estimates for marginal predictions of annual earnings among parents exposed to child's developmental disorder (F84) and unexposed parents stratified by the child's age at diagnosis**

| | | Finland | | | | Denmark | | | |
|---|---|---|---|---|---|---|---|---|---|
| | | Men | | Women | | Men | | Women | |
| | | Unexposed | Exposed | Unexposed | Exposed | Unexposed | Exposed | Unexposed | Exposed |
| Years since diagnosis | Age group | Marg. pred. (95% CI) | Marg. pred. (95% CI) | Marg. pred. (95% CI) | Marg. pred. (95% CI) | Marg. pred. (95% CI) | Marg. pred. (95% CI) | Marg. pred. (95% CI) | Marg. pred. (95% CI) |
| | | Unexposed | Exposed | Unexposed | Exposed | Unexposed | Exposed | Unexposed | Exposed |
| 2 | | 38.3 (37.6–39) | 34.9 (34.3–35.6) | 23.5 (23–23.9) | 21 (20.6–21.5) | 60.9 (60.2–61.6) | 58.2 (57.6–58.9) | 47.3 (46.9–47.6) | 45.2 (44.9–45.6) |
| 3 | | 38.7 (38–39.4) | 35.5 (34.8–36.2) | 24.6 (24.1–25.1) | 21.9 (21.4–22.4) | 62.3 (61.6–63) | 58.8 (58.2–59.5) | 48 (47.7–48.4) | 45.8 (45.5–46.2) |
| 4 | | 39.4 (38.6–40.1) | 36.1 (35.4–36.8) | 25.9 (25.4–26.4) | 22.8 (22.3–23.3) | 62.8 (62–63.5) | 59.6 (59–60.3) | 48.7 (48.3–49.1) | 46.5 (46.1–46.8) |
| 5 | | 39.7 (38.9–40.5) | 36.6 (35.8–37.3) | 26.9 (26.4–27.5) | 23.8 (23.3–24.4) | 63.6 (62.8–64.4) | 60.4 (59.7–61.1) | 49.6 (49.2–50) | 47 (46.7–47.4) |
| -5 | 10–14 years | 35.5 (34.8–36.3) | 33.3 (32.5–34) | 19.8 (19.3–20.3) | 17.8 (17.4–18.3) | 57 (56.4–57.6) | 54.6 (54.1–55.1) | 44.3 (44–44.6) | 43.1 (42.8–43.4) |
| -4 | | 36 (35.3–36.8) | 33.7 (33–34.4) | 20.9 (20.4–21.4) | 18.8 (18.3–19.3) | 58 (57.4–58.7) | 55.6 (55–56.1) | 45.2 (44.9–45.5) | 43.8 (43.5–44.1) |
| -3 | | 36.7 (36–37.5) | 34.6 (33.9–35.4) | 22 (21.5–22.5) | 19.6 (19.1–20) | 59.3 (58.7–60) | 56.5 (55.9–57) | 46 (45.7–46.3) | 44.5 (44.2–44.8) |
| -2 | | 37.2 (36.5–38) | 34.9 (34.2–35.7) | 23.1 (22.5–23.6) | 20.3 (19.8–20.7) | 60.3 (59.6–60.9) | 57.3 (56.7–57.9) | 46.7 (46.3–47) | 45.3 (44.9–45.6) |
| -1 | | 37.9 (37.1–38.7) | 35.2 (34.4–35.9) | 24.2 (23.7–24.7) | 21.4 (20.9–21.9) | 61.4 (60.7–62.1) | 58.2 (57.6–58.8) | 47.6 (47.2–47.9) | 46 (45.7–46.3) |

**eTable 54. Estimates for marginal predictions of annual earnings among parents exposed to child's developmental disorder (F84) and unexposed parents stratified by the child's age at diagnosis**

| | | Finland | | | | Denmark | | | |
|---|---|---|---|---|---|---|---|---|---|
| | | Men | | Women | | Men | | Women | |
| | | Unexposed | Exposed | Unexposed | Exposed | Unexposed | Exposed | Unexposed | Exposed |
| Years since diagnosis | Age group | Marg. pred. (95% CI) | Marg. pred. (95% CI) | Marg. pred. (95% CI) | Marg. pred. (95% CI) | Marg. pred. (95% CI) | Marg. pred. (95% CI) | Marg. pred. (95% CI) | Marg. pred. (95% CI) |
| | | Unexposed | Exposed | Unexposed | Exposed | Unexposed | Exposed | Unexposed | Exposed |
| 0 | | 38.4 (37.6–39.2) | 35.5 (34.7–36.3) | 25 (24.5–25.5) | 22.2 (21.7–22.8) | 62 (61.3–62.7) | 58.9 (58.2–59.5) | 48.4 (48.1–48.8) | 46.5 (46.2–46.8) |
| 1 | | 38.6 (37.8–39.4) | 35.6 (34.8–36.4) | 25.9 (25.4–26.4) | 23 (22.5–23.5) | 63.1 (62.3–63.8) | 59.4 (58.8–60) | 49 (48.6–49.3) | 47.2 (46.9–47.6) |
| 2 | | 39 (38.1–39.8) | 35.8 (35–36.6) | 26.8 (26.3–27.4) | 24 (23.4–24.5) | 63.4 (62.6–64.1) | 59.9 (59.3–60.6) | 49.5 (49.1–49.9) | 47.7 (47.3–48) |
| 3 | | 39 (38.1–39.9) | 35.8 (35–36.7) | 27.6 (27–28.2) | 24.6 (24–25.1) | 64.4 (63.6–65.2) | 60.5 (59.8–61.2) | 50.1 (49.7–50.5) | 47.9 (47.5–48.3) |
| 4 | | 39.2 (38.2–40.1) | 36.1 (35.1–37) | 28.4 (27.8–28.9) | 25.2 (24.6–25.8) | 65.1 (64.3–66) | 60.8 (60.1–61.5) | 50.2 (49.8–50.6) | 47.8 (47.4–48.2) |
| 5 | | 39.4 (38.4–40.4) | 36.2 (35.3–37.2) | 28.9 (28.2–29.5) | 25.8 (25.2–26.4) | 65.4 (64.5–66.3) | 61.3 (60.6–62.1) | 50.3 (49.9–50.8) | 47.7 (47.3–48.2) |
| -5 | 15–19 years | 37.1 (36–38.2) | 34.3 (33.3–35.4) | 24.8 (24–25.5) | 22.8 (22.1–23.5) | 59.2 (58.3–60) | 56.6 (55.8–57.3) | 46 (45.5–46.4) | 44.7 (44.3–45.1) |
| -4 | | 37.7 (36.5–38.8) | 35 (33.9–36) | 25.7 (25–26.5) | 23.3 (22.6–24.1) | 60.1 (59.2–60.9) | 57.3 (56.5–58) | 46.6 (46.2–47.1) | 45.3 (44.9–45.7) |
| -3 | | 38.3 (37.1–39.5) | 35.3 (34.2–36.5) | 26.6 (25.9–27.4) | 24.1 (23.3–24.8) | 60.8 (59.9–61.6) | 57.8 (57–58.5) | 47.2 (46.7–47.6) | 45.8 (45.4–46.2) |

**eTable 54. Estimates for marginal predictions of annual earnings among parents exposed to child's developmental disorder (F84) and unexposed parents stratified by the child's age at diagnosis**

| | | Finland | | | | Denmark | | | |
|---|---|---|---|---|---|---|---|---|---|
| | | Men | | Women | | Men | | Women | |
| | | Unexposed | Exposed | Unexposed | Exposed | Unexposed | Exposed | Unexposed | Exposed |
| Years since diagnosis | Age group | Marg. pred. (95% CI) | Marg. pred. (95% CI) | Marg. pred. (95% CI) | Marg. pred. (95% CI) | Marg. pred. (95% CI) | Marg. pred. (95% CI) | Marg. pred. (95% CI) | Marg. pred. (95% CI) |
| | | Unexposed | Exposed | Unexposed | Exposed | Unexposed | Exposed | Unexposed | Exposed |
| -2 | | 38.3 (37.1–39.5) | 35.4 (34.3–36.6) | 27.2 (26.5–28) | 24.8 (24.1–25.6) | 61.5 (60.6–62.4) | 58.4 (57.6–59.1) | 47.8 (47.4–48.3) | 46.3 (45.9–46.7) |
| -1 | | 38.4 (37.2–39.6) | 35.5 (34.4–36.7) | 27.7 (26.9–28.4) | 25.1 (24.4–25.9) | 62.5 (61.6–63.4) | 58.9 (58.1–59.7) | 48.3 (47.8–48.7) | 46.9 (46.4–47.3) |
| 0 | | 38.7 (37.5–39.9) | 35.7 (34.5–36.8) | 28.4 (27.7–29.2) | 25.8 (25–26.6) | 63 (62–63.9) | 59.2 (58.4–60) | 48.6 (48.1–49.1) | 46.9 (46.4–47.3) |
| 1 | | 38.9 (37.6–40.1) | 34.9 (33.7–36.1) | 29.2 (28.4–30) | 26 (25.2–26.8) | 63.7 (62.8–64.7) | 59.3 (58.5–60.1) | 48.8 (48.3–49.3) | 46.9 (46.5–47.4) |
| 2 | | 38.9 (37.6–40.2) | 34.9 (33.6–36.1) | 29.4 (28.5–30.2) | 26.3 (25.4–27.1) | 63.7 (62.7–64.7) | 59.5 (58.6–60.3) | 48.7 (48.2–49.3) | 46.8 (46.4–47.3) |
| 3 | | 38.6 (37.3–39.9) | 34.6 (33.3–35.9) | 29.8 (28.9–30.6) | 26.3 (25.5–27.2) | 64 (62.9–65) | 60.1 (59.2–61.1) | 48.5 (48–49.1) | 46.4 (45.9–46.9) |
| 4 | | 38.4 (37–39.8) | 34.3 (32.9–35.6) | 29.7 (28.8–30.5) | 26 (25.1–26.9) | 63.5 (62.4–64.6) | 60 (59–60.9) | 48.5 (47.9–49) | 46.7 (46.1–47.2) |
| 5 | | 37.6 (36.1–39) | 33.6 (32.1–35) | 29.9 (29–30.9) | 26.5 (25.5–27.4) | 63.3 (62.2–64.5) | 59.4 (58.4–60.4) | 48.6 (48–49.2) | 46.6 (46–47.1) |
| -5 | 20–25 years | 39.1 (37.5–40.8) | 35.6 (34–37.1) | 27.2 (26.2–28.3) | 24.3 (23.2–25.3) | 57.8 (56.4–59.2) | 55 (53.7–56.2) | 44.7 (44–45.4) | 43 (42.3–43.6) |

**eTable 54. Estimates for marginal predictions of annual earnings among parents exposed to child's developmental disorder (F84) and unexposed parents stratified by the child's age at diagnosis**

| | | Finland | | | | Denmark | | | |
|---|---|---|---|---|---|---|---|---|---|
| | | Men | | Women | | Men | | Women | |
| | | Unexposed | Exposed | Unexposed | Exposed | Unexposed | Exposed | Unexposed | Exposed |
| Years since diagnosis | Age group | Marg. pred. (95% CI) | Marg. pred. (95% CI) | Marg. pred. (95% CI) | Marg. pred. (95% CI) | Marg. pred. (95% CI) | Marg. pred. (95% CI) | Marg. pred. (95% CI) | Marg. pred. (95% CI) |
| | | Unexposed | Exposed | Unexposed | Exposed | Unexposed | Exposed | Unexposed | Exposed |
| -4 | | 38.9 (37.3–40.5) | 35.4 (33.7–37) | 27.6 (26.5–28.6) | 24.5 (23.4–25.5) | 58.1 (56.7–59.5) | 54.9 (53.7–56.1) | 44.7 (44–45.4) | 43 (42.4–43.7) |
| -3 | | 38.5 (36.8–40.2) | 34.6 (33–36.3) | 27.8 (26.7–28.9) | 24.8 (23.8–25.9) | 57.9 (56.5–59.3) | 55.4 (54.2–56.7) | 44.6 (43.8–45.3) | 43.1 (42.4–43.8) |
| -2 | | 37.4 (35.7–39) | 34.1 (32.4–35.7) | 28.1 (27–29.2) | 24.6 (23.6–25.7) | 58.3 (56.9–59.8) | 55 (53.8–56.3) | 44.5 (43.8–45.2) | 42.9 (42.2–43.6) |
| -1 | | 37.6 (35.9–39.3) | 33.5 (31.8–35.2) | 28 (26.9–29.1) | 24.7 (23.6–25.7) | 58.8 (57.4–60.3) | 55.4 (54.2–56.7) | 44.8 (44–45.5) | 42.9 (42.1–43.6) |
| 0 | | 37.4 (35.6–39.1) | 32.7 (30.9–34.4) | 27.5 (26.4–28.6) | 24.5 (23.4–25.6) | 59.1 (57.6–60.6) | 55.1 (53.8–56.4) | 44.8 (44.1–45.6) | 42.7 (42–43.4) |
| 1 | | 36.3 (34.5–38.1) | 31.8 (30–33.5) | 27.4 (26.2–28.5) | 24 (22.9–25.2) | 58.9 (57.3–60.4) | 54.6 (53.3–55.9) | 44.9 (44.1–45.7) | 42.5 (41.8–43.3) |
| 2 | | 35.2 (33.3–37.1) | 30.1 (28.2–32) | 27.2 (26–28.4) | 24.4 (23.2–25.6) | 58.8 (57.3–60.4) | 54.6 (53.3–56) | 45.2 (44.4–46) | 42.8 (42–43.6) |
| 3 | | 34.2 (32.2–36.2) | 29.2 (27.2–31.1) | 27.3 (26.1–28.6) | 24 (22.7–25.2) | 58.7 (57.1–60.4) | 55.1 (53.6–56.6) | 45 (44.1–45.8) | 43 (42.2–43.8) |
| 4 | | 32.5 (30.4–34.6) | 28.5 (26.5–30.5) | 26.7 (25.4–28) | 23.6 (22.2–24.9) | 58.8 (57–60.6) | 54.6 (53.1–56.1) | 44.5 (43.6–45.4) | 43 (42.2–43.9) |

**eTable 54. Estimates for marginal predictions of annual earnings among parents exposed to child's developmental disorder (F84) and unexposed parents stratified by the child's age at diagnosis**

| | | Finland | | | | Denmark | | | |
|---|---|---|---|---|---|---|---|---|---|
| | | Men | | Women | | Men | | Women | |
| | | Unexposed | Exposed | Unexposed | Exposed | Unexposed | Exposed | Unexposed | Exposed |
| Years since diagnosis | Age group | Marg. pred. (95% CI) | Marg. pred. (95% CI) | Marg. pred. (95% CI) | Marg. pred. (95% CI) | Marg. pred. (95% CI) | Marg. pred. (95% CI) | Marg. pred. (95% CI) | Marg. pred. (95% CI) |
| | | Unexposed | Exposed | Unexposed | Exposed | Unexposed | Exposed | Unexposed | Exposed |
| 5 | | 31.6 (29.4–33.8) | 27.2 (24.9–29.4) | 25.7 (24.3–27.1) | 22.8 (21.4–24.2) | 57.1 (55.3–59) | 54.7 (53–56.4) | 44.2 (43.3–45.1) | 43 (42–43.9) |

Marg. pred. = Marginal predictions in thousand euros (€)

**eTable 55. Estimates for marginal predictions of annual disposable income among parents exposed to child's developmental disorder (F84) and unexposed parents stratified by the child's age at diagnosis**

| | | Finland | | | | Denmark | | | |
|---|---|---|---|---|---|---|---|---|---|
| | | Men | | Women | | Men | | Women | |
| | | Unexposed | Exposed | Unexposed | Exposed | Unexposed | Exposed | Unexposed | Exposed |
| Years since diagnosis | Age group | Marg. pred. (95% CI) | Marg. pred. (95% CI) | Marg. pred. (95% CI) | Marg. pred. (95% CI) | Marg. pred. (95% CI) | Marg. pred. (95% CI) | Marg. pred. (95% CI) | Marg. pred. (95% CI) |
| | | Unexposed | Exposed | Unexposed | Exposed | Unexposed | Exposed | Unexposed | Exposed |
| -5 | 1–4 years | 22.6 (22.2–23.1) | 21 (20.5–21.4) | 16.7 (16.4–17) | 15.1 (14.8–15.4) | 28.7 (28.3–29.2) | 27 (26.6–27.4) | 23.4 (23.1–23.7) | 21.6 (21.3–21.9) |
| -4 | | 23.7 (23.3–24.1) | 22.1 (21.6–22.5) | 17.3 (17–17.6) | 15.6 (15.3–15.9) | 30.1 (29.7–30.6) | 28.3 (27.9–28.7) | 24.7 (24.4–24.9) | 22.5 (22.3–22.8) |
| -3 | | 24.7 (24.3–25.2) | 23.1 (22.7–23.5) | 17.8 (17.5–18.1) | 16 (15.7–16.3) | 31.7 (31.2–32.2) | 29.4 (28.9–29.8) | 26.3 (26–26.6) | 24.2 (23.9–24.5) |
| -2 | | 25.7 (25.3–26.2) | 24 (23.6–24.4) | 18.5 (18.2–18.8) | 16.5 (16.2–16.8) | 33.1 (32.6–33.6) | 30.9 (30.4–31.3) | 27.9 (27.6–28.2) | 25.8 (25.5–26.1) |
| -1 | | 26.7 (26.2–27.1) | 24.9 (24.5–25.4) | 19.8 (19.5–20.1) | 17.5 (17.3–17.8) | 34.5 (34–35) | 32 (31.6–32.5) | 29.3 (29–29.6) | 26.8 (26.5–27.1) |
| 0 | | 27.7 (27.2–28.2) | 25.6 (25.2–26.1) | 21.3 (21–21.6) | 18.9 (18.6–19.2) | 35.5 (34.9–36) | 32.9 (32.4–33.4) | 30.3 (30–30.6) | 27.9 (27.6–28.2) |
| 1 | | 28.6 (28.1–29.1) | 26.5 (26–26.9) | 22.5 (22.2–22.8) | 20.2 (19.9–20.5) | 36.7 (36.1–37.3) | 33.9 (33.4–34.4) | 31.3 (31–31.6) | 29.2 (28.9–29.6) |
| 2 | | 29.5 (29–30.1) | 27.2 (26.7–27.7) | 23.7 (23.4–24.1) | 21.5 (21.2–21.8) | 37.8 (37.2–38.5) | 34.8 (34.3–35.4) | 32.4 (32–32.7) | 30.2 (29.9–30.6) |
| 3 | | 30.4 (29.8–30.9) | 27.5 (27–28) | 24.6 (24.3–25) | 22.4 (22.1–22.7) | 38.9 (38.2–39.6) | 35.7 (35.2–36.3) | 33.2 (32.8–33.6) | 31 (30.6–31.4) |

**eTable 55. Estimates for marginal predictions of annual disposable income among parents exposed to child's developmental disorder (F84) and unexposed parents stratified by the child's age at diagnosis**

| | | Finland | | | | Denmark | | | |
|---|---|---|---|---|---|---|---|---|---|
| | | Men | | Women | | Men | | Women | |
| | | Unexposed | Exposed | Unexposed | Exposed | Unexposed | Exposed | Unexposed | Exposed |
| Years since diagnosis | Age group | Marg. pred. (95% CI) | Marg. pred. (95% CI) | Marg. pred. (95% CI) | Marg. pred. (95% CI) | Marg. pred. (95% CI) | Marg. pred. (95% CI) | Marg. pred. (95% CI) | Marg. pred. (95% CI) |
| | | Unexposed | Exposed | Unexposed | Exposed | Unexposed | Exposed | Unexposed | Exposed |
| 4 | | 31.1 (30.5–31.7) | 28.5 (27.9–29.1) | 25.5 (25.1–25.8) | 23.3 (22.9–23.6) | 39.8 (39.1–40.5) | 36.6 (35.9–37.2) | 34.1 (33.7–34.4) | 31.7 (31.3–32.1) |
| 5 | | 31.8 (31.2–32.4) | 29.5 (28.9–30) | 26.3 (25.9–26.6) | 24.1 (23.7–24.4) | 41.1 (40.4–41.9) | 37.2 (36.5–37.8) | 34.7 (34.3–35.2) | 32.3 (31.9–32.7) |
| -5 | 5–9 years | 26.1 (25.8–26.5) | 24.5 (24.2–24.9) | 19.3 (19.1–19.6) | 17.9 (17.7–18.2) | 31.9 (31.6–32.1) | 30.8 (30.5–31) | 27.5 (27.4–27.7) | 26.5 (26.3–26.7) |
| -4 | | 26.9 (26.5–27.3) | 25.3 (25–25.7) | 20.5 (20.3–20.8) | 18.9 (18.7–19.1) | 33 (32.7–33.3) | 31.7 (31.4–31.9) | 28.8 (28.6–29) | 27.8 (27.6–28) |
| -3 | | 27.8 (27.4–28.2) | 25.9 (25.5–26.2) | 21.7 (21.5–21.9) | 20.1 (19.8–20.3) | 34 (33.7–34.3) | 32.5 (32.3–32.8) | 29.9 (29.7–30.1) | 28.8 (28.6–29) |
| -2 | | 28.4 (28–28.8) | 26.5 (26.1–26.8) | 22.7 (22.4–22.9) | 21.1 (20.9–21.3) | 35 (34.7–35.3) | 33.7 (33.4–34) | 30.8 (30.6–31) | 29.5 (29.4–29.7) |
| -1 | | 29.1 (28.7–29.5) | 27.3 (26.9–27.6) | 23.6 (23.3–23.8) | 21.9 (21.7–22.1) | 36.1 (35.7–36.4) | 34.8 (34.4–35.1) | 31.7 (31.5–31.9) | 30.3 (30.2–30.5) |
| 0 | | 29.8 (29.4–30.3) | 27.8 (27.4–28.2) | 24.4 (24.2–24.7) | 22.7 (22.5–23) | 36.9 (36.5–37.3) | 35.6 (35.3–35.9) | 32.5 (32.3–32.7) | 30.9 (30.7–31.1) |
| 1 | | 30.5 (30–30.9) | 28.3 (27.9–28.7) | 25.3 (25–25.5) | 23.6 (23.3–23.8) | 38 (37.7–38.4) | 36.3 (36–36.6) | 33.2 (33–33.4) | 31.8 (31.6–32) |

**eTable 55. Estimates for marginal predictions of annual disposable income among parents exposed to child's developmental disorder (F84) and unexposed parents stratified by the child's age at diagnosis**

| | | Finland | | | | Denmark | | | |
|---|---|---|---|---|---|---|---|---|---|
| | | Men | | Women | | Men | | Women | |
| | | Unexposed | Exposed | Unexposed | Exposed | Unexposed | Exposed | Unexposed | Exposed |
| Years since diagnosis | Age group | Marg. pred. (95% CI) | Marg. pred. (95% CI) | Marg. pred. (95% CI) | Marg. pred. (95% CI) | Marg. pred. (95% CI) | Marg. pred. (95% CI) | Marg. pred. (95% CI) | Marg. pred. (95% CI) |
| | | Unexposed | Exposed | Unexposed | Exposed | Unexposed | Exposed | Unexposed | Exposed |
| 2 | | 31.1 (30.6–31.5) | 28.9 (28.5–29.3) | 25.8 (25.5–26) | 24.3 (24–24.6) | 39 (38.6–39.4) | 37.3 (36.9–37.6) | 33.9 (33.7–34.1) | 32.5 (32.3–32.7) |
| 3 | | 31.6 (31.1–32.1) | 29.5 (29.1–30) | 26.5 (26.3–26.8) | 25 (24.8–25.3) | 40.3 (39.9–40.7) | 38 (37.6–38.3) | 34.6 (34.4–34.9) | 33.1 (32.9–33.3) |
| 4 | | 32.2 (31.7–32.7) | 30.1 (29.6–30.5) | 27.3 (27–27.6) | 25.6 (25.3–25.9) | 41.1 (40.7–41.5) | 38.9 (38.5–39.3) | 35.3 (35–35.5) | 33.7 (33.5–33.9) |
| 5 | | 32.7 (32.2–33.2) | 30.7 (30.2–31.2) | 27.9 (27.6–28.2) | 26.2 (25.9–26.5) | 42.1 (41.6–42.6) | 39.8 (39.4–40.2) | 36.1 (35.8–36.4) | 34.3 (34.1–34.6) |
| -5 | 10–14 years | 28.7 (28.3–29.2) | 27.2 (26.8–27.7) | 23.1 (22.9–23.4) | 21.8 (21.5–22.1) | 34.9 (34.5–35.2) | 33.5 (33.2–33.7) | 31 (30.8–31.1) | 30.1 (29.9–30.3) |
| -4 | | 29.3 (28.8–29.8) | 27.6 (27.1–28) | 23.9 (23.7–24.2) | 22.7 (22.4–23) | 35.9 (35.5–36.2) | 34.3 (34–34.6) | 31.7 (31.5–31.9) | 30.7 (30.6–30.9) |
| -3 | | 30 (29.5–30.5) | 28.4 (27.9–28.9) | 24.7 (24.4–24.9) | 23.2 (23–23.5) | 36.9 (36.6–37.3) | 35.2 (34.8–35.5) | 32.4 (32.2–32.6) | 31.4 (31.2–31.6) |
| -2 | | 30.6 (30.1–31.1) | 28.8 (28.3–29.2) | 25.3 (25–25.6) | 23.8 (23.6–24.1) | 37.9 (37.5–38.2) | 36 (35.7–36.3) | 33 (32.8–33.2) | 32 (31.8–32.2) |
| -1 | | 31 (30.5–31.6) | 29.2 (28.8–29.7) | 25.9 (25.6–26.2) | 24.5 (24.2–24.8) | 38.9 (38.5–39.3) | 36.8 (36.5–37.2) | 33.8 (33.6–34) | 32.6 (32.4–32.8) |

**eTable 55. Estimates for marginal predictions of annual disposable income among parents exposed to child's developmental disorder (F84) and unexposed parents stratified by the child's age at diagnosis**

| | | Finland | | | | Denmark | | | |
|---|---|---|---|---|---|---|---|---|---|
| | | Men | | Women | | Men | | Women | |
| | | Unexposed | Exposed | Unexposed | Exposed | Unexposed | Exposed | Unexposed | Exposed |
| Years since diagnosis | Age group | Marg. pred. (95% CI) | Marg. pred. (95% CI) | Marg. pred. (95% CI) | Marg. pred. (95% CI) | Marg. pred. (95% CI) | Marg. pred. (95% CI) | Marg. pred. (95% CI) | Marg. pred. (95% CI) |
| | | Unexposed | Exposed | Unexposed | Exposed | Unexposed | Exposed | Unexposed | Exposed |
| 0 | | 31.6 (31.1–32.2) | 29.6 (29.1–30.1) | 26.4 (26.1–26.7) | 25 (24.7–25.3) | 39.7 (39.3–40.1) | 37.6 (37.3–38) | 34.5 (34.3–34.8) | 33.2 (33–33.4) |
| 1 | | 31.9 (31.4–32.5) | 29.8 (29.3–30.3) | 26.9 (26.6–27.3) | 25.6 (25.3–25.9) | 40.7 (40.2–41.1) | 38.4 (38–38.8) | 35.1 (34.8–35.3) | 33.9 (33.6–34.1) |
| 2 | | 32.3 (31.7–32.8) | 30.4 (29.8–30.9) | 27.4 (27.1–27.8) | 26.1 (25.8–26.4) | 41.3 (40.9–41.7) | 39.1 (38.7–39.5) | 35.6 (35.4–35.9) | 34.3 (34.1–34.5) |
| 3 | | 32.6 (32–33.1) | 30.8 (30.2–31.3) | 27.6 (27.3–28) | 26.3 (26–26.6) | 42.4 (42–42.9) | 39.9 (39.5–40.3) | 36.1 (35.9–36.4) | 34.6 (34.4–34.9) |
| 4 | | 33.2 (32.6–33.9) | 31 (30.4–31.6) | 27.7 (27.4–28.1) | 26.4 (26–26.7) | 43.4 (42.9–43.9) | 40.5 (40.1–41) | 36.3 (36.1–36.6) | 34.6 (34.4–34.9) |
| 5 | | 33.4 (32.8–34) | 31.6 (31–32.2) | 27.6 (27.2–27.9) | 26.4 (26–26.7) | 44.1 (43.6–44.7) | 41.5 (41–42) | 36.5 (36.2–36.8) | 34.7 (34.4–35) |
| -5 | 15–19 years | 30.2 (29.5–30.9) | 28.6 (27.9–29.3) | 25.5 (25.1–26) | 24.5 (24.1–24.9) | 36.6 (36.1–37) | 35.1 (34.6–35.5) | 32.1 (31.9–32.4) | 31.3 (31–31.5) |
| -4 | | 30.7 (29.9–31.4) | 29.2 (28.5–29.9) | 26.1 (25.7–26.5) | 25 (24.6–25.4) | 37.5 (37.1–38) | 35.8 (35.4–36.2) | 32.8 (32.5–33.1) | 31.9 (31.6–32.1) |
| -3 | | 31.1 (30.3–31.8) | 29.6 (28.9–30.3) | 26.5 (26.1–26.9) | 25.3 (24.9–25.7) | 38.3 (37.8–38.8) | 36.6 (36.1–37) | 33.4 (33.1–33.7) | 32.4 (32.1–32.7) |
| -2 | | 31.5 (30.8–32.3) | 29.7 (29–30.5) | 26.7 (26.3–27.1) | 25.6 (25.2–26) | 39.1 (38.6–39.6) | 37.2 (36.7–37.6) | 33.9 (33.6–34.2) | 32.8 (32.6–33.1) |

**eTable 55. Estimates for marginal predictions of annual disposable income among parents exposed to child's developmental disorder (F84) and unexposed parents stratified by the child's age at diagnosis**

| | | Finland | | | | Denmark | | | |
|---|---|---|---|---|---|---|---|---|---|
| | | Men | | Women | | Men | | Women | |
| | | Unexposed | Exposed | Unexposed | Exposed | Unexposed | Exposed | Unexposed | Exposed |
| Years since diagnosis | Age group | Marg. pred. (95% CI) | Marg. pred. (95% CI) | Marg. pred. (95% CI) | Marg. pred. (95% CI) | Marg. pred. (95% CI) | Marg. pred. (95% CI) | Marg. pred. (95% CI) | Marg. pred. (95% CI) |
| | | Unexposed | Exposed | Unexposed | Exposed | Unexposed | Exposed | Unexposed | Exposed |
| -1 | | 31.7 (30.9–32.5) | 29.8 (29.1–30.6) | 26.8 (26.4–27.2) | 25.6 (25.2–26) | 40.1 (39.6–40.6) | 37.9 (37.5–38.4) | 34.3 (34–34.6) | 33.3 (33–33.6) |
| 0 | | 32 (31.2–32.8) | 30.1 (29.3–30.8) | 26.9 (26.4–27.3) | 25.7 (25.2–26.1) | 40.9 (40.3–41.4) | 38.6 (38.1–39) | 34.6 (34.3–34.9) | 33.4 (33.2–33.7) |
| 1 | | 32.4 (31.6–33.2) | 30.1 (29.4–30.9) | 26.9 (26.4–27.3) | 25.6 (25.1–26) | 41.8 (41.2–42.4) | 39.2 (38.7–39.7) | 34.8 (34.5–35.1) | 33.6 (33.3–33.8) |
| 2 | | 32.6 (31.8–33.4) | 30.3 (29.5–31.1) | 26.6 (26.1–27.1) | 25.2 (24.7–25.7) | 42.3 (41.7–42.9) | 39.9 (39.4–40.4) | 34.9 (34.5–35.2) | 33.5 (33.2–33.8) |
| 3 | | 32.7 (31.8–33.5) | 30.6 (29.8–31.5) | 26.7 (26.2–27.2) | 25.1 (24.6–25.6) | 43 (42.3–43.6) | 40.7 (40.1–41.2) | 34.9 (34.5–35.2) | 33.3 (33–33.7) |
| 4 | | 32.8 (32–33.7) | 30.7 (29.9–31.6) | 26.6 (26.1–27.1) | 24.8 (24.3–25.3) | 43.3 (42.6–43.9) | 41 (40.4–41.6) | 35 (34.6–35.3) | 33.6 (33.2–33.9) |
| 5 | | 32.8 (31.9–33.7) | 31 (30.1–31.9) | 26.8 (26.3–27.3) | 25.2 (24.7–25.8) | 43.8 (43.1–44.6) | 41.2 (40.6–41.9) | 35.3 (34.9–35.7) | 33.7 (33.3–34.1) |
| -5 | 20–25 years | 32.1 (31–33.2) | 29.7 (28.7–30.7) | 25.1 (24.5–25.7) | 23.8 (23.2–24.4) | 36.8 (36–37.5) | 35.1 (34.4–35.8) | 31.1 (30.7–31.6) | 29.9 (29.5–30.3) |
| -4 | | 32.2 (31.2–33.3) | 29.7 (28.7–30.7) | 25.1 (24.5–25.7) | 23.7 (23.1–24.3) | 37.5 (36.7–38.3) | 35.5 (34.8–36.2) | 31.3 (30.8–31.7) | 30 (29.6–30.4) |

**eTable 55. Estimates for marginal predictions of annual disposable income among parents exposed to child's developmental disorder (F84) and unexposed parents stratified by the child's age at diagnosis**

| | | Finland | | | | Denmark | | | |
|---|---|---|---|---|---|---|---|---|---|
| | | Men | | Women | | Men | | Women | |
| | | Unexposed | Exposed | Unexposed | Exposed | Unexposed | Exposed | Unexposed | Exposed |
| Years since diagnosis | Age group | Marg. pred. (95% CI) | Marg. pred. (95% CI) | Marg. pred. (95% CI) | Marg. pred. (95% CI) | Marg. pred. (95% CI) | Marg. pred. (95% CI) | Marg. pred. (95% CI) | Marg. pred. (95% CI) |
| | | Unexposed | Exposed | Unexposed | Exposed | Unexposed | Exposed | Unexposed | Exposed |
| -3 | | 32.2 (31.2–33.3) | 29.6 (28.6–30.6) | 25 (24.4–25.6) | 23.9 (23.2–24.5) | 37.8 (37–38.6) | 36.4 (35.6–37.1) | 31.2 (30.7–31.6) | 30.2 (29.7–30.6) |
| -2 | | 31.5 (30.5–32.6) | 29.6 (28.6–30.6) | 25 (24.4–25.7) | 23.6 (23–24.2) | 38.5 (37.7–39.3) | 36.5 (35.7–37.2) | 31.3 (30.9–31.8) | 30.1 (29.7–30.6) |
| -1 | | 32.4 (31.3–33.5) | 29.5 (28.5–30.5) | 25.2 (24.5–25.8) | 23.6 (23–24.2) | 39.1 (38.3–40) | 37.1 (36.4–37.9) | 31.6 (31.1–32) | 30.2 (29.7–30.6) |
| 0 | | 32.5 (31.4–33.6) | 29.7 (28.6–30.7) | 25 (24.4–25.7) | 23.7 (23.1–24.3) | 39.7 (38.8–40.6) | 37.4 (36.6–38.1) | 31.8 (31.3–32.3) | 30.2 (29.8–30.7) |
| 1 | | 32.5 (31.3–33.6) | 29.6 (28.5–30.6) | 24.9 (24.3–25.6) | 23.8 (23.2–24.4) | 40 (39.1–41) | 37.5 (36.7–38.3) | 32.1 (31.6–32.6) | 30.2 (29.8–30.7) |
| 2 | | 32.1 (30.9–33.2) | 29 (27.9–30.1) | 25 (24.4–25.7) | 24 (23.4–24.7) | 40.5 (39.6–41.5) | 37.8 (37–38.6) | 32.4 (31.9–33) | 30.7 (30.2–31.2) |
| 3 | | 32.3 (31.1–33.5) | 29.2 (28.1–30.3) | 25.3 (24.6–26) | 23.8 (23.2–24.5) | 40.8 (39.8–41.8) | 38.5 (37.6–39.4) | 32.4 (31.8–33) | 31.1 (30.5–31.6) |
| 4 | | 31.8 (30.6–33.1) | 29.6 (28.4–30.7) | 25.2 (24.5–25.9) | 24.2 (23.4–24.9) | 41.3 (40.1–42.4) | 38.4 (37.5–39.4) | 32.4 (31.8–32.9) | 31.2 (30.7–31.8) |
| 5 | | 32.3 (30.9–33.6) | 29.5 (28.3–30.8) | 25.4 (24.6–26.1) | 24.3 (23.5–25) | 40.9 (39.8–42) | 38.7 (37.7–39.8) | 32.6 (32–33.2) | 31.4 (30.8–32.1) |

Marg. pred. = Marginal predictions in thousand euros (€)

**eTable 56. Estimates for marginal predictions of annual social welfare benefits among parents exposed to child's developmental disorder (F84) and unexposed parents stratified by the child's age at diagnosis**

| | | Finland | | | | Denmark | | | |
|---|---|---|---|---|---|---|---|---|---|
| | | Men | | Women | | Men | | Women | |
| | | Unexposed | Exposed | Unexposed | Exposed | Unexposed | Exposed | Unexposed | Exposed |
| Years since diagnosis | Age group | Marg. pred. (95% CI) | Marg. pred. (95% CI) | Marg. pred. (95% CI) | Marg. pred. (95% CI) | Marg. pred. (95% CI) | Marg. pred. (95% CI) | Marg. pred. (95% CI) | Marg. pred. (95% CI) |
| | | Unexposed | Exposed | Unexposed | Exposed | Unexposed | Exposed | Unexposed | Exposed |
| -5 | 1–4 years | 2.4 (2.2–2.5) | 3 (2.8–3.1) | 6.6 (6.4–6.8) | 6.1 (5.9–6.3) | 3.6 (3.4–3.8) | 4.1 (3.9–4.3) | 8 (7.7–8.2) | 8 (7.8–8.3) |
| -4 | | 2.6 (2.5–2.8) | 3.3 (3.1–3.5) | 9 (8.8–9.3) | 8.8 (8.5–9) | 3.6 (3.4–3.8) | 4.4 (4.2–4.6) | 10.8 (10.6–11.1) | 11.2 (11–11.5) |
| -3 | | 2.9 (2.7–3.1) | 3.8 (3.6–4) | 11.1 (10.9–11.4) | 11 (10.8–11.2) | 3.6 (3.4–3.8) | 4.4 (4.2–4.6) | 12 (11.8–12.3) | 12.8 (12.5–13.1) |
| -2 | | 3 (2.8–3.2) | 4 (3.8–4.2) | 11.4 (11.2–11.6) | 11.8 (11.6–12) | 3.6 (3.4–3.8) | 4.3 (4.1–4.6) | 12.1 (11.8–12.4) | 13 (12.8–13.3) |
| -1 | | 2.9 (2.8–3.1) | 3.9 (3.7–4.1) | 10.4 (10.2–10.7) | 11 (10.8–11.3) | 3.5 (3.3–3.7) | 4.3 (4.1–4.5) | 11.4 (11.2–11.7) | 12.6 (12.3–12.9) |
| 0 | | 2.7 (2.5–2.9) | 3.9 (3.7–4.1) | 9.8 (9.6–10) | 10.4 (10.2–10.7) | 3.5 (3.3–3.7) | 4.4 (4.2–4.6) | 11.2 (10.9–11.4) | 12.3 (12–12.6) |
| 1 | | 2.9 (2.6–3.3) | 3.9 (3.7–4.1) | 9.4 (9.1–9.6) | 10 (9.8–10.2) | 3.4 (3.2–3.6) | 4.4 (4.2–4.7) | 10.9 (10.6–11.1) | 12 (11.7–12.3) |
| 2 | | 2.9 (2.7–3.1) | 3.9 (3.7–4.1) | 9.1 (8.8–9.3) | 9.8 (9.5–10) | 3.5 (3.3–3.7) | 4.5 (4.2–4.7) | 10.5 (10.2–10.8) | 11.9 (11.6–12.2) |
| 3 | | 3 (2.8–3.2) | 4 (3.8–4.2) | 8.9 (8.6–9.1) | 9.5 (9.3–9.8) | 3.6 (3.3–3.8) | 4.5 (4.3–4.8) | 10.2 (9.9–10.5) | 11.6 (11.2–11.9) |
| 4 | | 3.1 (2.9–3.4) | 4.1 (3.9–4.4) | 8.8 (8.6–9.1) | 9.4 (9.1–9.7) | 3.6 (3.4–3.9) | 4.6 (4.3–4.9) | 10 (9.7–10.3) | 11.3 (11–11.7) |
| 5 | | 3.2 (3–3.4) | 4.3 (4–4.5) | 8.8 (8.5–9) | 9.3 (9.1–9.6) | 3.6 (3.3–3.8) | 4.5 (4.2–4.8) | 9.7 (9.4–10) | 11 (10.7–11.4) |

**eTable 56. Estimates for marginal predictions of annual social welfare benefits among parents exposed to child's developmental disorder (F84) and unexposed parents stratified by the child's age at diagnosis**

| | | Finland | | | | Denmark | | | |
|---|---|---|---|---|---|---|---|---|---|
| | | Men | | Women | | Men | | Women | |
| | | Unexposed | Exposed | Unexposed | Exposed | Unexposed | Exposed | Unexposed | Exposed |
| Years since diagnosis | Age group | Marg. pred. (95% CI) | Marg. pred. (95% CI) | Marg. pred. (95% CI) | Marg. pred. (95% CI) | Marg. pred. (95% CI) | Marg. pred. (95% CI) | Marg. pred. (95% CI) | Marg. pred. (95% CI) |
| | | Unexposed | Exposed | Unexposed | Exposed | Unexposed | Exposed | Unexposed | Exposed |
| -5 | 5–9 years | 2.8 (2.6–2.9) | 3.2 (3–3.3) | 10.4 (10.2–10.6) | 10.4 (10.2–10.5) | 3.5 (3.4–3.6) | 3.6 (3.5–3.8) | 12.1 (11.9–12.3) | 12.6 (12.5–12.8) |
| -4 | | 2.8 (2.7–2.9) | 3.1 (3–3.3) | 9.9 (9.7–10) | 9.8 (9.7–10) | 3.4 (3.3–3.5) | 3.6 (3.5–3.7) | 11.4 (11.3–11.6) | 12 (11.8–12.1) |
| -3 | | 2.8 (2.7–2.9) | 3.3 (3.1–3.4) | 9.5 (9.3–9.7) | 9.7 (9.5–9.8) | 3.4 (3.3–3.6) | 3.6 (3.5–3.7) | 11 (10.8–11.2) | 11.8 (11.6–11.9) |
| -2 | | 2.9 (2.7–3) | 3.3 (3.1–3.4) | 9.2 (9–9.4) | 9.4 (9.2–9.6) | 3.4 (3.3–3.6) | 3.6 (3.5–3.8) | 10.8 (10.6–10.9) | 11.5 (11.3–11.7) |
| -1 | | 2.9 (2.8–3.1) | 3.3 (3.1–3.4) | 8.9 (8.7–9.1) | 9.2 (9–9.4) | 3.4 (3.3–3.5) | 3.7 (3.5–3.8) | 10.4 (10.2–10.6) | 11.2 (11–11.4) |
| 0 | | 2.9 (2.8–3.1) | 3.4 (3.2–3.5) | 8.8 (8.6–8.9) | 9.1 (8.9–9.3) | 3.4 (3.3–3.6) | 3.7 (3.6–3.8) | 10.1 (9.9–10.3) | 11.2 (11–11.3) |
| 1 | | 3 (2.9–3.2) | 3.5 (3.3–3.6) | 8.5 (8.3–8.7) | 9 (8.8–9.2) | 3.5 (3.3–3.6) | 3.8 (3.7–4) | 9.8 (9.6–10) | 11 (10.8–11.2) |
| 2 | | 3.1 (2.9–3.3) | 3.5 (3.3–3.7) | 8.3 (8.1–8.5) | 8.8 (8.6–9) | 3.6 (3.4–3.7) | 3.9 (3.7–4) | 9.6 (9.4–9.8) | 10.8 (10.6–11) |
| 3 | | 3.3 (3.1–3.5) | 3.7 (3.5–3.9) | 8.2 (8–8.4) | 8.8 (8.6–9) | 3.6 (3.4–3.7) | 4 (3.9–4.2) | 9.4 (9.2–9.6) | 10.7 (10.5–10.9) |
| 4 | | 3.4 (3.3–3.6) | 3.9 (3.7–4.1) | 7.9 (7.7–8.1) | 8.7 (8.5–8.9) | 3.8 (3.6–3.9) | 4.2 (4–4.3) | 9.4 (9.2–9.6) | 10.5 (10.3–10.7) |
| 5 | | 3.6 (3.4–3.8) | 4.2 (3.9–4.4) | 7.7 (7.5–7.9) | 8.4 (8.2–8.7) | 3.9 (3.7–4) | 4.3 (4.1–4.4) | 9.1 (8.9–9.3) | 10.5 (10.2–10.7) |

**eTable 56. Estimates for marginal predictions of annual social welfare benefits among parents exposed to child's developmental disorder (F84) and unexposed parents stratified by the child’s age at diagnosis**

| | | Finland | | | | Denmark | | | |
|---|---|---|---|---|---|---|---|---|---|
| | | Men | | Women | | Men | | Women | |
| | | Unexposed | Exposed | Unexposed | Exposed | Unexposed | Exposed | Unexposed | Exposed |
| Years since diagnosis | Age group | Marg. pred. (95% CI) | Marg. pred. (95% CI) | Marg. pred. (95% CI) | Marg. pred. (95% CI) | Marg. pred. (95% CI) | Marg. pred. (95% CI) | Marg. pred. (95% CI) | Marg. pred. (95% CI) |
| | | Unexposed | Exposed | Unexposed | Exposed | Unexposed | Exposed | Unexposed | Exposed |
| -5 | 10–14 years | 2.9 (2.8–3.1) | 3.2 (3–3.3) | 8.6 (8.4–8.8) | 8.8 (8.6–9) | 3.6 (3.5–3.8) | 3.6 (3.5–3.8) | 10.2 (10–10.4) | 10.7 (10.6–10.9) |
| -4 | | 3.1 (3–3.3) | 3.3 (3.2–3.5) | 8.5 (8.3–8.7) | 8.9 (8.7–9.1) | 3.7 (3.6–3.8) | 3.7 (3.5–3.8) | 10 (9.8–10.2) | 10.6 (10.4–10.8) |
| -3 | | 3.2 (3–3.4) | 3.5 (3.3–3.7) | 8.5 (8.3–8.7) | 8.9 (8.7–9.1) | 3.7 (3.6–3.8) | 3.7 (3.5–3.8) | 9.8 (9.6–10) | 10.5 (10.3–10.7) |
| -2 | | 3.3 (3.1–3.5) | 3.6 (3.4–3.8) | 8.2 (8–8.4) | 8.9 (8.7–9.1) | 3.7 (3.6–3.9) | 3.8 (3.6–3.9) | 9.5 (9.4–9.7) | 10.3 (10.2–10.5) |
| -1 | | 3.3 (3.2–3.5) | 3.8 (3.6–4) | 8 (7.7–8.2) | 8.7 (8.5–8.9) | 3.8 (3.7–4) | 3.8 (3.7–4) | 9.3 (9.1–9.5) | 10.2 (10.1–10.4) |
| 0 | | 3.5 (3.3–3.7) | 3.9 (3.7–4.1) | 7.8 (7.5–8) | 8.5 (8.3–8.7) | 4 (3.8–4.1) | 4 (3.8–4.1) | 9.2 (9–9.3) | 10.2 (10–10.4) |
| 1 | | 3.6 (3.4–3.8) | 4.2 (3.9–4.4) | 7.6 (7.4–7.9) | 8.5 (8.2–8.7) | 4 (3.9–4.2) | 4.2 (4–4.4) | 9 (8.8–9.2) | 10.1 (9.9–10.3) |
| 2 | | 3.8 (3.5–4) | 4.3 (4.1–4.6) | 7.4 (7.2–7.7) | 8.2 (8–8.5) | 4.2 (4–4.3) | 4.3 (4.1–4.4) | 8.8 (8.6–9) | 9.9 (9.7–10.1) |
| 3 | | 3.9 (3.6–4.1) | 4.6 (4.3–4.9) | 7 (6.7–7.2) | 8.1 (7.8–8.3) | 4.3 (4.1–4.4) | 4.4 (4.2–4.6) | 8.6 (8.4–8.8) | 9.9 (9.7–10.2) |
| 4 | | 4.3 (4–4.5) | 4.7 (4.4–5) | 6.5 (6.3–6.7) | 7.6 (7.4–7.9) | 4.3 (4.2–4.5) | 4.5 (4.3–4.7) | 8.2 (7.9–8.4) | 9.6 (9.3–9.8) |
| 5 | | 4.3 (4.1–4.6) | 5 (4.7–5.3) | 6 (5.7–6.2) | 7.1 (6.9–7.4) | 4.4 (4.2–4.6) | 4.6 (4.4–4.8) | 7.8 (7.6–8) | 9.1 (8.9–9.4) |

**eTable 56. Estimates for marginal predictions of annual social welfare benefits among parents exposed to child's developmental disorder (F84) and unexposed parents stratified by the child's age at diagnosis**

| | | Finland | | | | Denmark | | | |
|---|---|---|---|---|---|---|---|---|---|
| | | Men | | Women | | Men | | Women | |
| | | Unexposed | Exposed | Unexposed | Exposed | Unexposed | Exposed | Unexposed | Exposed |
| Years since diagnosis | Age group | Marg. pred. (95% CI) | Marg. pred. (95% CI) | Marg. pred. (95% CI) | Marg. pred. (95% CI) | Marg. pred. (95% CI) | Marg. pred. (95% CI) | Marg. pred. (95% CI) | Marg. pred. (95% CI) |
| | | Unexposed | Exposed | Unexposed | Exposed | Unexposed | Exposed | Unexposed | Exposed |
| -5 | 15–19 years | 3.1 (2.8–3.4) | 3.6 (3.3–3.9) | 7.2 (7–7.5) | 7.7 (7.5–8) | 3.9 (3.8–4.1) | 4.2 (4–4.3) | 9.4 (9.1–9.6) | 10.1 (9.8–10.3) |
| -4 | | 3.3 (3–3.6) | 3.8 (3.5–4) | 7 (6.8–7.3) | 7.7 (7.4–8) | 3.9 (3.7–4.1) | 4.3 (4.1–4.5) | 9.2 (9–9.4) | 10 (9.7–10.2) |
| -3 | | 3.5 (3.2–3.8) | 4 (3.7–4.3) | 6.9 (6.6–7.1) | 7.6 (7.3–7.9) | 4.1 (3.9–4.3) | 4.3 (4.1–4.5) | 9 (8.7–9.2) | 9.9 (9.6–10.1) |
| -2 | | 3.8 (3.5–4.1) | 4.2 (3.9–4.5) | 6.5 (6.3–6.8) | 7.3 (7–7.6) | 4.2 (4–4.4) | 4.4 (4.2–4.6) | 8.9 (8.6–9.1) | 9.7 (9.4–9.9) |
| -1 | | 3.9 (3.5–4.2) | 4.2 (3.9–4.5) | 6.3 (6–6.6) | 7.1 (6.8–7.4) | 4.3 (4.1–4.5) | 4.5 (4.3–4.7) | 8.6 (8.4–8.8) | 9.4 (9.2–9.6) |
| 0 | | 4.1 (3.7–4.4) | 4.5 (4.1–4.8) | 5.9 (5.6–6.2) | 6.6 (6.3–6.9) | 4.2 (4–4.4) | 4.7 (4.5–4.9) | 8.3 (8–8.5) | 9.2 (9–9.5) |
| 1 | | 4.2 (3.9–4.6) | 4.9 (4.5–5.2) | 5.3 (5–5.6) | 6.3 (5.9–6.6) | 4.4 (4.2–4.6) | 4.9 (4.7–5.1) | 7.9 (7.6–8.1) | 9 (8.8–9.3) |
| 2 | | 4.3 (3.9–4.6) | 5 (4.7–5.4) | 4.9 (4.6–5.2) | 5.7 (5.4–6.1) | 4.4 (4.2–4.6) | 5 (4.7–5.2) | 7.4 (7.1–7.6) | 8.5 (8.2–8.8) |
| 3 | | 4.6 (4.2–5) | 5.4 (5–5.8) | 4.6 (4.3–4.9) | 5.6 (5.2–5.9) | 4.6 (4.4–4.8) | 5 (4.7–5.2) | 6.8 (6.6–7.1) | 8.1 (7.9–8.4) |
| 4 | | 5.1 (4.6–5.5) | 5.8 (5.3–6.3) | 4.6 (4.2–4.9) | 5.6 (5.3–6) | 4.8 (4.5–5) | 5.2 (4.9–5.4) | 6.6 (6.3–6.9) | 8 (7.7–8.3) |
| 5 | | 5.7 (5.2–6.3) | 6.7 (6.1–7.2) | 4.6 (4.2–4.9) | 5.7 (5.3–6.1) | 4.9 (4.6–5.1) | 5.3 (5–5.6) | 6.6 (6.3–6.8) | 8 (7.6–8.3) |
| -5 | 20–25 years | 3.4 (3–3.9) | 4.1 (3.7–4.6) | 5.1 (4.7–5.5) | 6 (5.6–6.4) | 4.2 (3.9–4.5) | 4.9 (4.5–5.2) | 8 (7.6–8.4) | 9.2 (8.8–9.6) |
| -4 | | 3.8 (3.3–4.2) | 4.3 (3.9–4.8) | 4.8 (4.4–5.2) | 5.7 (5.3–6.1) | 4.4 (4.1–4.8) | 5.1 (4.7–5.4) | 7.8 (7.4–8.1) | 9 (8.6–9.4) |

**eTable 56. Estimates for marginal predictions of annual social welfare benefits among parents exposed to child's developmental disorder (F84) and unexposed parents stratified by the child's age at diagnosis**

| | | Finland | | | | Denmark | | | |
|---|---|---|---|---|---|---|---|---|---|
| | | Men | | Women | | Men | | Women | |
| | | Unexposed | Exposed | Unexposed | Exposed | Unexposed | Exposed | Unexposed | Exposed |
| Years since diagnosis | Age group | Marg. pred. (95% CI) | Marg. pred. (95% CI) | Marg. pred. (95% CI) | Marg. pred. (95% CI) | Marg. pred. (95% CI) | Marg. pred. (95% CI) | Marg. pred. (95% CI) | Marg. pred. (95% CI) |
| | | Unexposed | Exposed | Unexposed | Exposed | Unexposed | Exposed | Unexposed | Exposed |
| -3 | | 4.2 (3.7–4.7) | 4.8 (4.4–5.3) | 4.6 (4.1–5) | 5.6 (5.1–6) | 4.5 (4.1–4.8) | 5.2 (4.9–5.6) | 7.4 (7–7.8) | 8.7 (8.3–9.1) |
| -2 | | 4.7 (4.1–5.2) | 5.3 (4.8–5.8) | 4.3 (3.9–4.7) | 5.5 (5.1–5.9) | 4.6 (4.3–5) | 5.4 (5.1–5.8) | 7.1 (6.7–7.4) | 8.5 (8.1–8.9) |
| -1 | | 4.9 (4.4–5.4) | 5.8 (5.3–6.4) | 4.4 (4–4.8) | 5.5 (5.1–5.9) | 4.8 (4.5–5.2) | 5.5 (5.2–5.9) | 6.9 (6.5–7.3) | 8.5 (8.1–8.9) |
| 0 | | 5.2 (4.7–5.8) | 6.7 (6.1–7.3) | 4.7 (4.2–5.1) | 5.7 (5.2–6.1) | 5 (4.6–5.3) | 5.8 (5.4–6.2) | 6.9 (6.5–7.3) | 8.7 (8.3–9.1) |
| 1 | | 5.9 (5.3–6.5) | 7.1 (6.5–7.8) | 4.7 (4.3–5.1) | 6 (5.6–6.5) | 5.2 (4.8–5.6) | 6 (5.6–6.4) | 7.1 (6.7–7.5) | 8.9 (8.4–9.3) |
| 2 | | 6.5 (5.8–7.2) | 7.9 (7.2–8.6) | 4.9 (4.5–5.4) | 6 (5.6–6.5) | 5.5 (5.1–5.9) | 6.5 (6.1–6.9) | 7.2 (6.8–7.7) | 9 (8.5–9.5) |
| 3 | | 7.4 (6.6–8.1) | 8.6 (7.8–9.3) | 4.9 (4.4–5.3) | 6.4 (5.8–6.9) | 5.9 (5.4–6.3) | 6.9 (6.4–7.3) | 7.4 (7–7.9) | 9.1 (8.7–9.6) |
| 4 | | 8.2 (7.4–9.1) | 9.6 (8.7–10.5) | 5.4 (4.9–6) | 6.9 (6.3–7.5) | 6.5 (6–7) | 7.1 (6.6–7.6) | 7.7 (7.2–8.2) | 9.6 (9.1–10.2) |
| 5 | | 9 (8.1–10) | 10.9 (9.9–11.9) | 6 (5.4–6.7) | 7.6 (7–8.2) | 7.2 (6.7–7.7) | 7.8 (7.3–8.4) | 8 (7.4–8.5) | 10.4 (9.8–11) |

Marg. pred. = Marginal predictions in thousand euros (€)

**eTable 57. Estimates for marginal predictions of annual employment among parents exposed to child's developmental disorder (F84) and unexposed parents stratified by the child’s age at diagnosis**

| | | Finland | | | | Denmark | | | |
|---|---|---|---|---|---|---|---|---|---|
| | | Men | | Women | | Men | | Women | |
| | | Unexposed | Exposed | Unexposed | Exposed | Unexposed | Exposed | Unexposed | Exposed |
| Years since diagnosis | Age group | Marg. pred. (95% CI) | Marg. pred. (95% CI) | Marg. pred. (95% CI) | Marg. pred. (95% CI) | Marg. pred. (95% CI) | Marg. pred. (95% CI) | Marg. pred. (95% CI) | Marg. pred. (95% CI) |
| | | Unexposed | Exposed | Unexposed | Exposed | Unexposed | Exposed | Unexposed | Exposed |
| -5 | 1–4 years | 0.8 (0.79–0.81) | 0.74 (0.72–0.75) | 0.62 (0.6–0.63) | 0.56 (0.54–0.57) | 0.71 (0.7–0.72) | 0.67 (0.66–0.69) | 0.61 (0.59–0.62) | 0.54 (0.52–0.55) |
| -4 | | 0.8 (0.79–0.81) | 0.74 (0.73–0.75) | 0.58 (0.57–0.6) | 0.5 (0.48–0.51) | 0.73 (0.72–0.74) | 0.69 (0.68–0.71) | 0.61 (0.6–0.62) | 0.52 (0.51–0.54) |
| -3 | | 0.81 (0.8–0.82) | 0.75 (0.73–0.76) | 0.55 (0.53–0.56) | 0.45 (0.44–0.47) | 0.75 (0.74–0.76) | 0.71 (0.7–0.72) | 0.62 (0.6–0.63) | 0.53 (0.51–0.54) |
| -2 | | 0.82 (0.81–0.83) | 0.76 (0.74–0.77) | 0.55 (0.54–0.57) | 0.44 (0.42–0.45) | 0.77 (0.76–0.78) | 0.72 (0.7–0.73) | 0.64 (0.62–0.65) | 0.54 (0.53–0.56) |
| -1 | | 0.83 (0.82–0.85) | 0.77 (0.75–0.78) | 0.6 (0.58–0.61) | 0.47 (0.46–0.49) | 0.77 (0.76–0.78) | 0.73 (0.72–0.74) | 0.66 (0.65–0.67) | 0.57 (0.55–0.58) |
| 0 | | 0.84 (0.82–0.85) | 0.77 (0.75–0.78) | 0.65 (0.63–0.66) | 0.53 (0.51–0.54) | 0.77 (0.76–0.78) | 0.73 (0.71–0.74) | 0.67 (0.66–0.68) | 0.57 (0.56–0.59) |
| 1 | | 0.84 (0.82–0.85) | 0.78 (0.76–0.79) | 0.68 (0.67–0.7) | 0.56 (0.54–0.57) | 0.77 (0.76–0.78) | 0.73 (0.71–0.74) | 0.68 (0.67–0.7) | 0.59 (0.58–0.61) |
| 2 | | 0.84 (0.83–0.85) | 0.78 (0.76–0.79) | 0.71 (0.69–0.72) | 0.6 (0.58–0.61) | 0.78 (0.77–0.79) | 0.72 (0.71–0.73) | 0.7 (0.69–0.71) | 0.61 (0.59–0.62) |
| 3 | | 0.84 (0.83–0.86) | 0.78 (0.76–0.79) | 0.74 (0.72–0.75) | 0.62 (0.61–0.64) | 0.77 (0.75–0.78) | 0.72 (0.71–0.74) | 0.71 (0.7–0.72) | 0.62 (0.6–0.63) |

**eTable 57. Estimates for marginal predictions of annual employment among parents exposed to child's developmental disorder (F84) and unexposed parents stratified by the child's age at diagnosis**

| | | Finland | | | | Denmark | | | |
|---|---|---|---|---|---|---|---|---|---|
| | | Men | | Women | | Men | | Women | |
| | | Unexposed | Exposed | Unexposed | Exposed | Unexposed | Exposed | Unexposed | Exposed |
| Years since diagnosis | Age group | Marg. pred. (95% CI) | Marg. pred. (95% CI) | Marg. pred. (95% CI) | Marg. pred. (95% CI) | Marg. pred. (95% CI) | Marg. pred. (95% CI) | Marg. pred. (95% CI) | Marg. pred. (95% CI) |
| | | Unexposed | Exposed | Unexposed | Exposed | Unexposed | Exposed | Unexposed | Exposed |
| 4 | | 0.83 (0.81–0.84) | 0.77 (0.76–0.78) | 0.76 (0.75–0.78) | 0.64 (0.62–0.66) | 0.77 (0.75–0.78) | 0.72 (0.71–0.74) | 0.71 (0.7–0.73) | 0.62 (0.61–0.64) |
| 5 | | 0.83 (0.82–0.84) | 0.77 (0.75–0.78) | 0.76 (0.75–0.78) | 0.65 (0.64–0.67) | 0.76 (0.75–0.78) | 0.72 (0.7–0.73) | 0.72 (0.7–0.73) | 0.63 (0.62–0.65) |
| -5 | 5–9 years | 0.82 (0.82–0.83) | 0.79 (0.78–0.8) | 0.6 (0.59–0.61) | 0.55 (0.53–0.56) | 0.75 (0.74–0.76) | 0.75 (0.74–0.76) | 0.63 (0.63–0.64) | 0.61 (0.6–0.62) |
| -4 | | 0.83 (0.82–0.84) | 0.79 (0.78–0.8) | 0.63 (0.62–0.64) | 0.57 (0.56–0.58) | 0.76 (0.76–0.77) | 0.76 (0.75–0.77) | 0.66 (0.66–0.67) | 0.64 (0.63–0.65) |
| -3 | | 0.83 (0.82–0.84) | 0.79 (0.78–0.8) | 0.67 (0.66–0.68) | 0.61 (0.6–0.62) | 0.77 (0.76–0.77) | 0.77 (0.77–0.78) | 0.68 (0.67–0.69) | 0.66 (0.65–0.66) |
| -2 | | 0.83 (0.82–0.84) | 0.8 (0.79–0.81) | 0.7 (0.69–0.71) | 0.63 (0.62–0.64) | 0.76 (0.76–0.77) | 0.77 (0.77–0.78) | 0.69 (0.68–0.7) | 0.67 (0.66–0.68) |
| -1 | | 0.83 (0.82–0.84) | 0.8 (0.79–0.81) | 0.72 (0.71–0.73) | 0.66 (0.65–0.67) | 0.77 (0.76–0.78) | 0.78 (0.77–0.78) | 0.7 (0.69–0.71) | 0.68 (0.67–0.69) |
| 0 | | 0.83 (0.82–0.84) | 0.8 (0.79–0.81) | 0.74 (0.73–0.75) | 0.68 (0.67–0.69) | 0.77 (0.76–0.78) | 0.78 (0.77–0.78) | 0.71 (0.71–0.72) | 0.68 (0.67–0.69) |
| 1 | | 0.83 (0.82–0.84) | 0.8 (0.79–0.81) | 0.76 (0.75–0.77) | 0.69 (0.68–0.7) | 0.77 (0.76–0.77) | 0.77 (0.76–0.78) | 0.72 (0.71–0.73) | 0.67 (0.67–0.68) |

**eTable 57. Estimates for marginal predictions of annual employment among parents exposed to child's developmental disorder (F84) and unexposed parents stratified by the child's age at diagnosis**

| | | Finland | | | | Denmark | | | |
|---|---|---|---|---|---|---|---|---|---|
| | | Men | | Women | | Men | | Women | |
| | | Unexposed | Exposed | Unexposed | Exposed | Unexposed | Exposed | Unexposed | Exposed |
| Years since diagnosis | Age group | Marg. pred. (95% CI) | Marg. pred. (95% CI) | Marg. pred. (95% CI) | Marg. pred. (95% CI) | Marg. pred. (95% CI) | Marg. pred. (95% CI) | Marg. pred. (95% CI) | Marg. pred. (95% CI) |
| | | Unexposed | Exposed | Unexposed | Exposed | Unexposed | Exposed | Unexposed | Exposed |
| 2 | | 0.83 (0.82–0.84) | 0.8 (0.79–0.81) | 0.77 (0.76–0.78) | 0.7 (0.69–0.72) | 0.76 (0.76–0.77) | 0.77 (0.76–0.77) | 0.73 (0.72–0.74) | 0.68 (0.67–0.69) |
| 3 | | 0.83 (0.82–0.84) | 0.8 (0.79–0.81) | 0.78 (0.77–0.79) | 0.71 (0.7–0.73) | 0.76 (0.75–0.77) | 0.76 (0.75–0.77) | 0.73 (0.72–0.74) | 0.68 (0.67–0.69) |
| 4 | | 0.82 (0.81–0.83) | 0.8 (0.79–0.81) | 0.8 (0.79–0.81) | 0.72 (0.71–0.74) | 0.76 (0.75–0.77) | 0.76 (0.75–0.77) | 0.73 (0.72–0.74) | 0.68 (0.67–0.69) |
| 5 | | 0.81 (0.8–0.82) | 0.78 (0.77–0.79) | 0.81 (0.79–0.82) | 0.74 (0.73–0.75) | 0.75 (0.74–0.76) | 0.76 (0.75–0.77) | 0.74 (0.73–0.74) | 0.68 (0.67–0.69) |
| -5 | 10–14 years | 0.83 (0.82–0.84) | 0.81 (0.8–0.82) | 0.72 (0.7–0.73) | 0.67 (0.66–0.69) | 0.75 (0.74–0.75) | 0.76 (0.76–0.77) | 0.7 (0.7–0.71) | 0.69 (0.68–0.7) |
| -4 | | 0.83 (0.82–0.84) | 0.8 (0.78–0.81) | 0.73 (0.71–0.74) | 0.69 (0.67–0.7) | 0.75 (0.75–0.76) | 0.77 (0.76–0.78) | 0.71 (0.7–0.72) | 0.7 (0.69–0.71) |
| -3 | | 0.83 (0.82–0.84) | 0.8 (0.79–0.81) | 0.74 (0.73–0.75) | 0.69 (0.68–0.7) | 0.76 (0.75–0.77) | 0.77 (0.76–0.78) | 0.72 (0.71–0.73) | 0.7 (0.7–0.71) |
| -2 | | 0.82 (0.81–0.84) | 0.79 (0.78–0.8) | 0.76 (0.75–0.78) | 0.71 (0.69–0.72) | 0.76 (0.75–0.77) | 0.77 (0.76–0.78) | 0.72 (0.72–0.73) | 0.71 (0.7–0.72) |
| -1 | | 0.82 (0.81–0.83) | 0.8 (0.79–0.81) | 0.78 (0.77–0.79) | 0.73 (0.72–0.74) | 0.76 (0.75–0.76) | 0.77 (0.76–0.78) | 0.73 (0.72–0.74) | 0.71 (0.7–0.72) |

**eTable 57. Estimates for marginal predictions of annual employment among parents exposed to child's developmental disorder (F84) and unexposed parents stratified by the child's age at diagnosis**

| | | Finland | | | | Denmark | | | |
|---|---|---|---|---|---|---|---|---|---|
| | | Men | | Women | | Men | | Women | |
| | | Unexposed | Exposed | Unexposed | Exposed | Unexposed | Exposed | Unexposed | Exposed |
| Years since diagnosis | Age group | Marg. pred. (95% CI) | Marg. pred. (95% CI) | Marg. pred. (95% CI) | Marg. pred. (95% CI) | Marg. pred. (95% CI) | Marg. pred. (95% CI) | Marg. pred. (95% CI) | Marg. pred. (95% CI) |
| | | Unexposed | Exposed | Unexposed | Exposed | Unexposed | Exposed | Unexposed | Exposed |
| 0 | | 0.82 (0.81–0.84) | 0.79 (0.77–0.8) | 0.79 (0.78–0.8) | 0.74 (0.73–0.76) | 0.76 (0.75–0.76) | 0.77 (0.76–0.77) | 0.74 (0.73–0.74) | 0.71 (0.7–0.72) |
| 1 | | 0.82 (0.81–0.83) | 0.78 (0.77–0.8) | 0.8 (0.79–0.81) | 0.75 (0.74–0.76) | 0.75 (0.75–0.76) | 0.76 (0.75–0.77) | 0.73 (0.72–0.74) | 0.7 (0.7–0.71) |
| 2 | | 0.81 (0.8–0.82) | 0.77 (0.76–0.79) | 0.8 (0.79–0.81) | 0.75 (0.74–0.76) | 0.75 (0.74–0.75) | 0.76 (0.75–0.77) | 0.74 (0.73–0.75) | 0.7 (0.69–0.71) |
| 3 | | 0.8 (0.78–0.81) | 0.77 (0.76–0.78) | 0.81 (0.8–0.82) | 0.75 (0.74–0.77) | 0.74 (0.73–0.75) | 0.75 (0.74–0.76) | 0.73 (0.73–0.74) | 0.69 (0.68–0.7) |
| 4 | | 0.79 (0.77–0.8) | 0.75 (0.74–0.77) | 0.81 (0.8–0.82) | 0.76 (0.74–0.77) | 0.74 (0.73–0.75) | 0.75 (0.74–0.76) | 0.74 (0.73–0.75) | 0.69 (0.68–0.7) |
| 5 | | 0.78 (0.77–0.8) | 0.74 (0.73–0.76) | 0.81 (0.8–0.83) | 0.76 (0.74–0.77) | 0.73 (0.72–0.74) | 0.75 (0.74–0.76) | 0.74 (0.73–0.75) | 0.69 (0.68–0.7) |
| -5 | 15–19 years | 0.83 (0.82–0.85) | 0.8 (0.78–0.82) | 0.81 (0.79–0.82) | 0.75 (0.73–0.77) | 0.75 (0.74–0.76) | 0.75 (0.74–0.76) | 0.72 (0.71–0.73) | 0.71 (0.7–0.72) |
| -4 | | 0.82 (0.81–0.84) | 0.8 (0.78–0.81) | 0.81 (0.8–0.83) | 0.75 (0.73–0.77) | 0.74 (0.73–0.75) | 0.75 (0.74–0.76) | 0.72 (0.71–0.73) | 0.71 (0.7–0.72) |
| -3 | | 0.81 (0.8–0.83) | 0.79 (0.77–0.8) | 0.81 (0.8–0.83) | 0.76 (0.74–0.78) | 0.75 (0.74–0.76) | 0.75 (0.74–0.76) | 0.73 (0.72–0.74) | 0.71 (0.7–0.72) |

**eTable 57. Estimates for marginal predictions of annual employment among parents exposed to child's developmental disorder (F84) and unexposed parents stratified by the child's age at diagnosis**

| | | Finland | | | | Denmark | | | |
|---|---|---|---|---|---|---|---|---|---|
| | | Men | | Women | | Men | | Women | |
| | | Unexposed | Exposed | Unexposed | Exposed | Unexposed | Exposed | Unexposed | Exposed |
| Years since diagnosis | Age group | Marg. pred. (95% CI) | Marg. pred. (95% CI) | Marg. pred. (95% CI) | Marg. pred. (95% CI) | Marg. pred. (95% CI) | Marg. pred. (95% CI) | Marg. pred. (95% CI) | Marg. pred. (95% CI) |
| | | Unexposed | Exposed | Unexposed | Exposed | Unexposed | Exposed | Unexposed | Exposed |
| -2 | | 0.81 (0.79–0.82) | 0.79 (0.77–0.8) | 0.82 (0.81–0.84) | 0.76 (0.75–0.78) | 0.74 (0.73–0.75) | 0.75 (0.74–0.76) | 0.73 (0.72–0.74) | 0.7 (0.69–0.72) |
| -1 | | 0.8 (0.79–0.82) | 0.77 (0.75–0.79) | 0.82 (0.8–0.83) | 0.77 (0.76–0.79) | 0.74 (0.73–0.75) | 0.75 (0.74–0.76) | 0.73 (0.72–0.74) | 0.71 (0.7–0.72) |
| 0 | | 0.8 (0.78–0.81) | 0.76 (0.74–0.78) | 0.82 (0.8–0.83) | 0.77 (0.75–0.78) | 0.74 (0.73–0.75) | 0.74 (0.73–0.75) | 0.72 (0.71–0.73) | 0.7 (0.69–0.71) |
| 1 | | 0.8 (0.78–0.81) | 0.75 (0.73–0.77) | 0.82 (0.8–0.84) | 0.76 (0.74–0.78) | 0.73 (0.72–0.74) | 0.73 (0.72–0.74) | 0.72 (0.71–0.73) | 0.69 (0.68–0.71) |
| 2 | | 0.78 (0.76–0.8) | 0.74 (0.72–0.76) | 0.82 (0.8–0.83) | 0.76 (0.74–0.78) | 0.73 (0.72–0.75) | 0.73 (0.72–0.74) | 0.72 (0.71–0.73) | 0.7 (0.69–0.71) |
| 3 | | 0.77 (0.75–0.79) | 0.74 (0.72–0.76) | 0.82 (0.8–0.84) | 0.76 (0.74–0.78) | 0.73 (0.71–0.74) | 0.72 (0.71–0.73) | 0.72 (0.7–0.73) | 0.69 (0.68–0.7) |
| 4 | | 0.76 (0.74–0.79) | 0.71 (0.69–0.73) | 0.81 (0.8–0.83) | 0.75 (0.72–0.77) | 0.71 (0.7–0.72) | 0.72 (0.71–0.73) | 0.72 (0.71–0.73) | 0.68 (0.67–0.7) |
| 5 | | 0.74 (0.72–0.76) | 0.68 (0.65–0.7) | 0.82 (0.8–0.83) | 0.74 (0.72–0.76) | 0.71 (0.7–0.72) | 0.71 (0.7–0.73) | 0.72 (0.7–0.73) | 0.68 (0.67–0.7) |
| -5 | 20–25 years | 0.82 (0.8–0.84) | 0.77 (0.74–0.79) | 0.81 (0.79–0.83) | 0.74 (0.72–0.77) | 0.71 (0.7–0.73) | 0.72 (0.7–0.73) | 0.69 (0.68–0.71) | 0.67 (0.66–0.69) |

**eTable 57. Estimates for marginal predictions of annual employment among parents exposed to child's developmental disorder (F84) and unexposed parents stratified by the child's age at diagnosis**

| | | Finland | | | | Denmark | | | |
|---|---|---|---|---|---|---|---|---|---|
| | | Men | | Women | | Men | | Women | |
| | | Unexposed | Exposed | Unexposed | Exposed | Unexposed | Exposed | Unexposed | Exposed |
| Years since diagnosis | Age group | Marg. pred. (95% CI) | Marg. pred. (95% CI) | Marg. pred. (95% CI) | Marg. pred. (95% CI) | Marg. pred. (95% CI) | Marg. pred. (95% CI) | Marg. pred. (95% CI) | Marg. pred. (95% CI) |
| | | Unexposed | Exposed | Unexposed | Exposed | Unexposed | Exposed | Unexposed | Exposed |
| -4 | | 0.81 (0.79–0.84) | 0.76 (0.73–0.78) | 0.82 (0.8–0.84) | 0.75 (0.73–0.77) | 0.71 (0.7–0.73) | 0.71 (0.69–0.73) | 0.7 (0.68–0.71) | 0.66 (0.65–0.68) |
| -3 | | 0.79 (0.77–0.81) | 0.75 (0.72–0.77) | 0.82 (0.79–0.84) | 0.74 (0.72–0.77) | 0.71 (0.69–0.73) | 0.72 (0.7–0.74) | 0.7 (0.68–0.71) | 0.66 (0.64–0.68) |
| -2 | | 0.77 (0.75–0.79) | 0.73 (0.7–0.75) | 0.8 (0.78–0.82) | 0.74 (0.72–0.76) | 0.72 (0.7–0.73) | 0.7 (0.68–0.72) | 0.7 (0.68–0.71) | 0.65 (0.63–0.67) |
| -1 | | 0.76 (0.73–0.78) | 0.7 (0.67–0.73) | 0.79 (0.77–0.81) | 0.74 (0.72–0.77) | 0.7 (0.69–0.72) | 0.7 (0.68–0.72) | 0.69 (0.67–0.71) | 0.64 (0.63–0.66) |
| 0 | | 0.75 (0.72–0.77) | 0.68 (0.66–0.71) | 0.79 (0.76–0.81) | 0.72 (0.69–0.74) | 0.7 (0.68–0.72) | 0.68 (0.66–0.7) | 0.69 (0.67–0.71) | 0.63 (0.61–0.65) |
| 1 | | 0.72 (0.7–0.75) | 0.66 (0.63–0.69) | 0.77 (0.74–0.79) | 0.7 (0.67–0.72) | 0.69 (0.67–0.71) | 0.67 (0.65–0.69) | 0.68 (0.66–0.7) | 0.63 (0.61–0.65) |
| 2 | | 0.69 (0.66–0.72) | 0.64 (0.61–0.67) | 0.77 (0.74–0.79) | 0.68 (0.66–0.71) | 0.69 (0.67–0.71) | 0.65 (0.63–0.67) | 0.67 (0.65–0.69) | 0.62 (0.6–0.64) |
| 3 | | 0.67 (0.64–0.7) | 0.62 (0.59–0.65) | 0.74 (0.71–0.77) | 0.68 (0.65–0.71) | 0.67 (0.65–0.69) | 0.64 (0.61–0.66) | 0.66 (0.64–0.68) | 0.61 (0.59–0.63) |
| 4 | | 0.62 (0.58–0.65) | 0.57 (0.54–0.61) | 0.73 (0.7–0.76) | 0.65 (0.62–0.68) | 0.66 (0.64–0.68) | 0.62 (0.6–0.65) | 0.64 (0.62–0.67) | 0.58 (0.56–0.61) |

**eTable 57. Estimates for marginal predictions of annual employment among parents exposed to child's developmental disorder (F84) and unexposed parents stratified by the child's age at diagnosis**

| | | Finland | | | | Denmark | | | |
|---|---|---|---|---|---|---|---|---|---|
| | | Men | | Women | | Men | | Women | |
| | | Unexposed | Exposed | Unexposed | Exposed | Unexposed | Exposed | Unexposed | Exposed |
| Years since diagnosis | Age group | Marg. pred. (95% CI) | Marg. pred. (95% CI) | Marg. pred. (95% CI) | Marg. pred. (95% CI) | Marg. pred. (95% CI) | Marg. pred. (95% CI) | Marg. pred. (95% CI) | Marg. pred. (95% CI) |
| | | Unexposed | Exposed | Unexposed | Exposed | Unexposed | Exposed | Unexposed | Exposed |
| 5 | | 0.6 (0.56–0.64) | 0.53 (0.49–0.57) | 0.72 (0.69–0.75) | 0.63 (0.59–0.66) | 0.63 (0.6–0.65) | 0.61 (0.58–0.64) | 0.62 (0.6–0.65) | 0.56 (0.53–0.58) |

Marg. pred. = Marginal predictions in probabilities

**eTable 58. Estimates for marginal predictions of annual earnings among parents exposed to child's childhood onset disorder (F90–F98) and unexposed parents stratified by the child's age at diagnosis**

| | | Finland | | | | Denmark | | | |
|---|---|---|---|---|---|---|---|---|---|
| | | Men | | Women | | Men | | Women | |
| | | Unexposed | Exposed | Unexposed | Exposed | Unexposed | Exposed | Unexposed | Exposed |
| Years since diagnosis | Age group | Marg. pred. (95% CI) | Marg. pred. (95% CI) | Marg. pred. (95% CI) | Marg. pred. (95% CI) | Marg. pred. (95% CI) | Marg. pred. (95% CI) | Marg. pred. (95% CI) | Marg. pred. (95% CI) |
| | | Unexposed | Exposed | Unexposed | Exposed | Unexposed | Exposed | Unexposed | Exposed |
| -5 | 1–4 years | 27.2 (26.7–27.6) | 24.9 (24.4–25.3) | 13.5 (13.2–13.8) | 13.1 (12.8–13.4) | 42.2 (41.4–43) | 40.3 (39.6–41.1) | 32.2 (31.7–32.7) | 30.1 (29.6–30.6) |
| -4 | | 28.5 (28–28.9) | 26.2 (25.8–26.7) | 10.2 (10–10.5) | 9.4 (9.2–9.7) | 43.8 (43–44.6) | 41.8 (41.1–42.6) | 33 (32.5–33.4) | 31.2 (30.8–31.7) |
| -3 | | 30 (29.5–30.5) | 27.5 (27–27.9) | 10.5 (10.3–10.8) | 9.4 (9.2–9.7) | 45.4 (44.6–46.3) | 43.2 (42.4–44) | 34.9 (34.4–35.4) | 33.2 (32.8–33.7) |
| -2 | | 31.6 (31.1–32.1) | 28.6 (28.2–29.1) | 12.4 (12.1–12.7) | 10.7 (10.4–11) | 47.2 (46.4–48.1) | 44.6 (43.7–45.4) | 36.7 (36.2–37.2) | 34.8 (34.4–35.3) |
| -1 | | 32.9 (32.4–33.4) | 29.9 (29.4–30.4) | 13.9 (13.5–14.2) | 11.7 (11.4–12) | 48.8 (47.9–49.7) | 45.7 (44.9–46.5) | 38 (37.5–38.5) | 36 (35.6–36.5) |
| 0 | | 34.1 (33.5–34.6) | 30.9 (30.4–31.4) | 16.2 (15.8–16.5) | 13.4 (13.1–13.7) | 49.7 (48.8–50.7) | 47.1 (46.2–48) | 39.3 (38.8–39.8) | 36.9 (36.4–37.3) |
| 1 | | 35 (34.5–35.6) | 31.9 (31.3–32.4) | 18.1 (17.7–18.4) | 15.4 (15.1–15.8) | 50.8 (49.9–51.8) | 48.1 (47.2–49) | 40.5 (40–41.1) | 38 (37.5–38.5) |
| 2 | | 36 (35.5–36.6) | 32.8 (32.2–33.3) | 19.6 (19.2–20) | 16.9 (16.5–17.2) | 52.1 (51.1–53.1) | 49 (48–49.9) | 41.2 (40.7–41.7) | 38.8 (38.3–39.4) |
| 3 | | 37 (36.4–37.6) | 33.6 (33–34.2) | 20.9 (20.5–21.3) | 18 (17.6–18.4) | 53.3 (52.3–54.4) | 49.1 (48.1–50) | 41.9 (41.3–42.5) | 39.5 (38.9–40) |
| 4 | | 37.8 (37.2–38.4) | 33.9 (33.3–34.5) | 22.1 (21.7–22.5) | 19.1 (18.7–19.5) | 54.8 (53.7–55.9) | 50.2 (49.2–51.2) | 42.8 (42.2–43.4) | 40 (39.5–40.6) |

**eTable 58. Estimates for marginal predictions of annual earnings among parents exposed to child's childhood onset disorder (F90–F98) and unexposed parents stratified by the child's age at diagnosis**

| | | Finland | | | | Denmark | | | |
|---|---|---|---|---|---|---|---|---|---|
| | | Men | | Women | | Men | | Women | |
| | | Unexposed | Exposed | Unexposed | Exposed | Unexposed | Exposed | Unexposed | Exposed |
| Years since diagnosis | Age group | Marg. pred. (95% CI) | Marg. pred. (95% CI) | Marg. pred. (95% CI) | Marg. pred. (95% CI) | Marg. pred. (95% CI) | Marg. pred. (95% CI) | Marg. pred. (95% CI) | Marg. pred. (95% CI) |
| | | Unexposed | Exposed | Unexposed | Exposed | Unexposed | Exposed | Unexposed | Exposed |
| 5 | | 38.6 (38–39.2) | 34.3 (33.6–34.9) | 23.3 (22.9–23.7) | 20.2 (19.7–20.6) | 55.6 (54.5–56.8) | 50.7 (49.6–51.7) | 43.6 (43–44.2) | 40.5 (39.9–41.1) |
| -5 | 5–9 years | 30.7 (30.5–30.9) | 28.1 (27.9–28.3) | 12.1 (12–12.3) | 11.4 (11.3–11.5) | 48.8 (48.5–49.1) | 46.7 (46.4–47) | 37.5 (37.3–37.7) | 37 (36.8–37.1) |
| -4 | | 31.8 (31.6–32) | 29.3 (29.1–29.5) | 14.4 (14.3–14.5) | 13.5 (13.3–13.6) | 50 (49.6–50.3) | 47.6 (47.3–47.9) | 38.8 (38.7–39) | 38.2 (38–38.3) |
| -3 | | 32.8 (32.6–33) | 30.2 (30–30.4) | 16.4 (16.3–16.5) | 15.1 (15–15.2) | 51.1 (50.8–51.5) | 48.7 (48.4–49) | 39.9 (39.7–40.1) | 39.1 (39–39.3) |
| -2 | | 33.7 (33.5–33.9) | 31 (30.8–31.2) | 18 (17.8–18.1) | 16.4 (16.3–16.6) | 52.2 (51.8–52.5) | 49.6 (49.3–49.9) | 40.9 (40.7–41.1) | 39.9 (39.7–40.1) |
| -1 | | 34.6 (34.3–34.8) | 31.8 (31.6–32) | 19.3 (19.1–19.4) | 17.5 (17.3–17.6) | 53.3 (52.9–53.6) | 50.5 (50.2–50.9) | 41.7 (41.5–41.9) | 40.7 (40.5–40.9) |
| 0 | | 35.4 (35.1–35.6) | 32.5 (32.3–32.8) | 20.5 (20.4–20.7) | 18.5 (18.4–18.7) | 54.3 (53.9–54.7) | 51.3 (51–51.6) | 42.5 (42.3–42.7) | 41.2 (41–41.4) |
| 1 | | 36.1 (35.9–36.3) | 33.1 (32.9–33.3) | 21.8 (21.6–21.9) | 19.5 (19.4–19.7) | 55.2 (54.8–55.6) | 52 (51.7–52.4) | 43.3 (43.1–43.5) | 41.8 (41.7–42) |
| 2 | | 36.7 (36.4–36.9) | 33.7 (33.5–33.9) | 22.9 (22.7–23.1) | 20.6 (20.5–20.8) | 55.9 (55.5–56.3) | 52.6 (52.3–53) | 44 (43.8–44.2) | 42.3 (42.1–42.5) |
| 3 | | 37.3 (37.1–37.6) | 34.2 (34–34.4) | 24 (23.9–24.2) | 21.7 (21.6–21.9) | 56.6 (56.2–57.1) | 53.2 (52.8–53.5) | 44.7 (44.5–44.9) | 42.8 (42.6–43) |

**eTable 58. Estimates for marginal predictions of annual earnings among parents exposed to child's childhood onset disorder (F90–F98) and unexposed parents stratified by the child’s age at diagnosis**

| | | Finland | | | | Denmark | | | |
|---|---|---|---|---|---|---|---|---|---|
| | | Men | | Women | | Men | | Women | |
| | | Unexposed | Exposed | Unexposed | Exposed | Unexposed | Exposed | Unexposed | Exposed |
| Years since diagnosis | Age group | Marg. pred. (95% CI) | Marg. pred. (95% CI) | Marg. pred. (95% CI) | Marg. pred. (95% CI) | Marg. pred. (95% CI) | Marg. pred. (95% CI) | Marg. pred. (95% CI) | Marg. pred. (95% CI) |
| | | Unexposed | Exposed | Unexposed | Exposed | Unexposed | Exposed | Unexposed | Exposed |
| 4 | | 37.8 (37.5–38) | 34.6 (34.4–34.9) | 25.2 (25–25.4) | 22.7 (22.6–22.9) | 57.7 (57.2–58.1) | 53.9 (53.5–54.3) | 45.4 (45.1–45.6) | 43.4 (43.2–43.6) |
| 5 | | 38.4 (38.1–38.6) | 35 (34.7–35.3) | 26.3 (26.1–26.4) | 23.7 (23.5–23.9) | 58.1 (57.6–58.6) | 54.5 (54.1–54.9) | 46 (45.8–46.3) | 44 (43.7–44.2) |
| -5 | 10–14 years | 32.5 (32.2–32.7) | 30 (29.7–30.2) | 18.6 (18.4–18.7) | 17.3 (17.1–17.4) | 52.5 (52.2–52.9) | 50.1 (49.8–50.5) | 40.9 (40.7–41.1) | 40.3 (40.1–40.5) |
| -4 | | 33.1 (32.8–33.4) | 30.6 (30.3–30.8) | 19.7 (19.5–19.8) | 18.2 (18.1–18.4) | 53.5 (53.1–53.9) | 50.8 (50.5–51.2) | 41.7 (41.5–41.9) | 41 (40.8–41.2) |
| -3 | | 33.6 (33.3–33.9) | 31 (30.7–31.2) | 20.6 (20.4–20.8) | 19 (18.9–19.2) | 54.4 (54–54.8) | 51.5 (51.2–51.9) | 42.4 (42.2–42.6) | 41.6 (41.4–41.8) |
| -2 | | 34.2 (34–34.5) | 31.5 (31.2–31.8) | 21.6 (21.4–21.8) | 19.9 (19.7–20.1) | 55.2 (54.8–55.6) | 52.4 (52–52.7) | 43.1 (42.9–43.3) | 42.3 (42.1–42.5) |
| -1 | | 34.7 (34.5–35) | 32.1 (31.8–32.4) | 22.6 (22.4–22.8) | 20.8 (20.7–21) | 56.2 (55.8–56.6) | 52.9 (52.6–53.3) | 43.9 (43.7–44.1) | 42.9 (42.7–43.1) |
| 0 | | 35.2 (34.9–35.5) | 32.4 (32.1–32.7) | 23.5 (23.3–23.7) | 21.5 (21.3–21.7) | 56.9 (56.4–57.3) | 53.7 (53.3–54.1) | 44.6 (44.4–44.8) | 43.4 (43.2–43.6) |
| 1 | | 35.7 (35.3–36) | 32.6 (32.3–32.9) | 24.3 (24.1–24.5) | 22.2 (22–22.4) | 57.7 (57.2–58.1) | 54.1 (53.8–54.5) | 45.2 (45–45.4) | 43.9 (43.7–44.1) |

**eTable 58. Estimates for marginal predictions of annual earnings among parents exposed to child's childhood onset disorder (F90–F98) and unexposed parents stratified by the child's age at diagnosis**

| | | Finland | | | | Denmark | | | |
|---|---|---|---|---|---|---|---|---|---|
| | | Men | | Women | | Men | | Women | |
| | | Unexposed | Exposed | Unexposed | Exposed | Unexposed | Exposed | Unexposed | Exposed |
| Years since diagnosis | Age group | Marg. pred. (95% CI) | Marg. pred. (95% CI) | Marg. pred. (95% CI) | Marg. pred. (95% CI) | Marg. pred. (95% CI) | Marg. pred. (95% CI) | Marg. pred. (95% CI) | Marg. pred. (95% CI) |
| | | Unexposed | Exposed | Unexposed | Exposed | Unexposed | Exposed | Unexposed | Exposed |
| 2 | | 35.9 (35.5–36.2) | 33 (32.7–33.3) | 25.1 (24.9–25.3) | 22.9 (22.7–23.1) | 58.3 (57.8–58.8) | 54.4 (54–54.8) | 45.7 (45.5–46) | 44.3 (44.1–44.5) |
| 3 | | 36.1 (35.8–36.4) | 33.1 (32.8–33.4) | 25.8 (25.6–26) | 23.6 (23.4–23.8) | 58.8 (58.3–59.2) | 55.1 (54.7–55.5) | 46.3 (46.1–46.6) | 44.6 (44.4–44.8) |
| 4 | | 36.2 (35.9–36.5) | 33.1 (32.8–33.4) | 26.5 (26.2–26.7) | 24.2 (24–24.4) | 59.1 (58.6–59.5) | 55.3 (54.8–55.7) | 46.5 (46.2–46.7) | 44.7 (44.5–45) |
| 5 | | 36.2 (35.9–36.6) | 33.1 (32.8–33.5) | 27 (26.8–27.3) | 24.7 (24.5–25) | 59.5 (59–60) | 55.6 (55.2–56.1) | 46.5 (46.3–46.8) | 44.8 (44.5–45) |
| -5 | 15–19 years | 33.8 (33.4–34.2) | 31.4 (31–31.7) | 21.9 (21.7–22.1) | 20.5 (20.3–20.8) | 53.6 (53.1–54.1) | 50.9 (50.5–51.4) | 42.7 (42.4–42.9) | 41.6 (41.4–41.9) |
| -4 | | 34.2 (33.8–34.5) | 31.6 (31.3–32) | 22.7 (22.5–23) | 21.4 (21.1–21.6) | 54.2 (53.7–54.7) | 51.2 (50.8–51.7) | 43.3 (43.1–43.6) | 42.2 (41.9–42.4) |
| -3 | | 34.8 (34.4–35.2) | 32 (31.6–32.3) | 23.5 (23.3–23.8) | 22.1 (21.9–22.4) | 54.8 (54.3–55.3) | 51.6 (51.1–52) | 43.9 (43.6–44.1) | 42.5 (42.3–42.8) |
| -2 | | 35.2 (34.8–35.6) | 32.3 (31.9–32.6) | 24.2 (23.9–24.4) | 22.9 (22.6–23.2) | 55.5 (55–56) | 52.3 (51.9–52.8) | 44.2 (43.9–44.4) | 42.9 (42.6–43.1) |
| -1 | | 35.3 (34.9–35.7) | 32.4 (32–32.8) | 24.9 (24.6–25.1) | 23.5 (23.2–23.7) | 55.9 (55.4–56.4) | 52.5 (52–53) | 44.5 (44.2–44.8) | 43.1 (42.8–43.4) |
| 0 | | 35.6 (35.2–36.1) | 32.4 (32–32.8) | 25.5 (25.3–25.8) | 24 (23.7–24.2) | 56.3 (55.7–56.8) | 52.6 (52.1–53) | 44.7 (44.4–45) | 43 (42.8–43.3) |

**eTable 58. Estimates for marginal predictions of annual earnings among parents exposed to child's childhood onset disorder (F90–F98) and unexposed parents stratified by the child's age at diagnosis**

| | | Finland | | | | Denmark | | | |
|---|---|---|---|---|---|---|---|---|---|
| | | Men | | Women | | Men | | Women | |
| | | Unexposed | Exposed | Unexposed | Exposed | Unexposed | Exposed | Unexposed | Exposed |
| Years since diagnosis | Age group | Marg. pred. (95% CI) | Marg. pred. (95% CI) | Marg. pred. (95% CI) | Marg. pred. (95% CI) | Marg. pred. (95% CI) | Marg. pred. (95% CI) | Marg. pred. (95% CI) | Marg. pred. (95% CI) |
| | | Unexposed | Exposed | Unexposed | Exposed | Unexposed | Exposed | Unexposed | Exposed |
| 1 | | 35.8 (35.3–36.2) | 32.4 (31.9–32.8) | 26 (25.7–26.3) | 24.4 (24.1–24.7) | 56.7 (56.1–57.2) | 52.8 (52.3–53.3) | 44.7 (44.4–45) | 43 (42.8–43.3) |
| 2 | | 35.6 (35.2–36) | 32.4 (31.9–32.8) | 26.5 (26.2–26.7) | 24.8 (24.5–25) | 56.7 (56.2–57.3) | 53.1 (52.6–53.7) | 44.6 (44.3–44.9) | 42.8 (42.6–43.1) |
| 3 | | 35.6 (35.1–36) | 32.2 (31.8–32.7) | 26.9 (26.6–27.2) | 25.1 (24.8–25.4) | 57.1 (56.5–57.7) | 53.2 (52.6–53.7) | 44.4 (44.1–44.7) | 42.7 (42.4–43) |
| 4 | | 35.2 (34.7–35.7) | 31.7 (31.3–32.2) | 27.1 (26.8–27.4) | 25.3 (25–25.6) | 57.1 (56.5–57.7) | 53.3 (52.8–53.9) | 44.3 (44–44.6) | 42.6 (42.3–43) |
| 5 | | 34.8 (34.3–35.3) | 31.4 (30.9–31.8) | 27.2 (26.9–27.5) | 25.5 (25.1–25.8) | 57.2 (56.5–57.8) | 53.2 (52.6–53.8) | 44.4 (44.1–44.8) | 42.6 (42.3–43) |
| -5 | 20–25 years | 35.2 (34.3–36.1) | 33.1 (32.2–34) | 25.4 (24.9–26) | 24.4 (23.9–25) | 54.3 (53.5–55.1) | 51.4 (50.7–52.1) | 43.1 (42.7–43.5) | 41.7 (41.3–42.1) |
| -4 | | 34.9 (34–35.9) | 33 (32.1–33.9) | 25.5 (24.9–26.1) | 24.6 (24–25.1) | 54.5 (53.7–55.3) | 51.6 (50.9–52.4) | 43 (42.6–43.4) | 41.6 (41.2–42) |
| -3 | | 34.7 (33.7–35.6) | 32.7 (31.8–33.7) | 25.8 (25.2–26.4) | 24.7 (24.1–25.3) | 54.7 (53.9–55.5) | 51.7 (51–52.5) | 42.9 (42.5–43.3) | 41.2 (40.8–41.6) |
| -2 | | 34.5 (33.6–35.5) | 32.4 (31.4–33.3) | 26 (25.4–26.6) | 24.9 (24.3–25.5) | 54.8 (54–55.7) | 51.8 (51–52.5) | 42.7 (42.3–43.2) | 41.2 (40.8–41.6) |

**eTable 58. Estimates for marginal predictions of annual earnings among parents exposed to child's childhood onset disorder (F90–F98) and unexposed parents stratified by the child's age at diagnosis**

| | | Finland | | | | Denmark | | | |
|---|---|---|---|---|---|---|---|---|---|
| | | Men | | Women | | Men | | Women | |
| | | Unexposed | Exposed | Unexposed | Exposed | Unexposed | Exposed | Unexposed | Exposed |
| Years since diagnosis | Age group | Marg. pred. (95% CI) | Marg. pred. (95% CI) | Marg. pred. (95% CI) | Marg. pred. (95% CI) | Marg. pred. (95% CI) | Marg. pred. (95% CI) | Marg. pred. (95% CI) | Marg. pred. (95% CI) |
| | | Unexposed | Exposed | Unexposed | Exposed | Unexposed | Exposed | Unexposed | Exposed |
| -1 | | 34 (33.1–35) | 31.8 (30.8–32.7) | 26.1 (25.5–26.7) | 25 (24.4–25.7) | 54.9 (54.1–55.7) | 51.6 (50.9–52.4) | 42.8 (42.3–43.2) | 41.3 (40.8–41.7) |
| 0 | | 33.1 (32.1–34.1) | 31.4 (30.4–32.3) | 26.4 (25.7–27) | 24.8 (24.2–25.5) | 55 (54.1–55.8) | 51.8 (51–52.6) | 42.7 (42.2–43.1) | 41.2 (40.7–41.6) |
| 1 | | 32.5 (31.5–33.5) | 30.8 (29.8–31.8) | 26.1 (25.5–26.7) | 24.2 (23.5–24.8) | 55 (54.1–55.8) | 51.8 (51–52.6) | 42.7 (42.2–43.1) | 41.2 (40.8–41.7) |
| 2 | | 31.6 (30.6–32.7) | 30 (29–31) | 25.7 (25–26.4) | 24 (23.3–24.6) | 54.6 (53.7–55.4) | 51.6 (50.8–52.4) | 42.6 (42.1–43) | 41.1 (40.7–41.6) |
| 3 | | 30.7 (29.6–31.7) | 28.9 (27.8–30) | 25.4 (24.7–26.1) | 23.7 (23–24.4) | 54.3 (53.3–55.2) | 51.5 (50.6–52.3) | 42.6 (42.1–43.1) | 41 (40.5–41.4) |
| 4 | | 29.8 (28.7–30.9) | 28 (26.9–29.2) | 25 (24.3–25.8) | 23.1 (22.4–23.8) | 53.9 (53–54.9) | 51.2 (50.3–52.1) | 42.5 (42.1–43) | 41 (40.5–41.5) |
| 5 | | 28.3 (27–29.5) | 27 (25.8–28.2) | 24.3 (23.5–25.1) | 22.4 (21.7–23.2) | 53.6 (52.6–54.6) | 50.8 (49.9–51.8) | 42.6 (42.1–43.1) | 40.9 (40.4–41.4) |

Marg. pred. = Marginal predictions in thousand euros (€)

**eTable 59. Estimates for marginal predictions of annual disposable income among parents exposed to child's childhood onset disorder (F90–F98) and unexposed parents stratified by the child's age at diagnosis**

| | | Finland | | | | Denmark | | | |
|---|---|---|---|---|---|---|---|---|---|
| | | Men | | Women | | Men | | Women | |
| | | Unexposed | Exposed | Unexposed | Exposed | Unexposed | Exposed | Unexposed | Exposed |
| Years since diagnosis | Age group | Marg. pred. (95% CI) | Marg. pred. (95% CI) | Marg. pred. (95% CI) | Marg. pred. (95% CI) | Marg. pred. (95% CI) | Marg. pred. (95% CI) | Marg. pred. (95% CI) | Marg. pred. (95% CI) |
| | | Unexposed | Exposed | Unexposed | Exposed | Unexposed | Exposed | Unexposed | Exposed |
| -5 | 1–4 years | 22.6 (22.3–22.9) | 21.3 (21–21.6) | 16.7 (16.5–16.9) | 16.3 (16.1–16.5) | 25.7 (25.3–26.1) | 24.8 (24.3–25.2) | 21.7 (21.4–22) | 20.4 (20.1–20.7) |
| -4 | | 23.9 (23.5–24.2) | 22.6 (22.3–22.9) | 17.1 (16.9–17.3) | 16.6 (16.4–16.8) | 26.9 (26.5–27.3) | 25.9 (25.4–26.3) | 22.8 (22.6–23.1) | 21.7 (21.4–22) |
| -3 | | 24.9 (24.6–25.2) | 23.6 (23.3–23.8) | 17.8 (17.7–18) | 17.3 (17.1–17.5) | 28 (27.5–28.4) | 26.7 (26.3–27.1) | 24.5 (24.2–24.8) | 23.5 (23.2–23.8) |
| -2 | | 26.1 (25.7–26.4) | 24.4 (24.1–24.7) | 19.1 (18.9–19.3) | 18.5 (18.3–18.7) | 29.2 (28.7–29.6) | 27.6 (27.1–28) | 26 (25.7–26.3) | 24.8 (24.5–25.1) |
| -1 | | 27 (26.7–27.3) | 25.5 (25.1–25.8) | 20.3 (20.1–20.5) | 19.5 (19.3–19.7) | 30.4 (29.9–30.9) | 28.5 (28–28.9) | 27.1 (26.8–27.4) | 25.8 (25.5–26.1) |
| 0 | | 28 (27.7–28.3) | 26.2 (25.9–26.6) | 21.5 (21.3–21.7) | 20.4 (20.2–20.6) | 31 (30.5–31.6) | 29.4 (29–29.9) | 28.2 (27.9–28.5) | 26.6 (26.3–26.9) |
| 1 | | 28.7 (28.4–29.1) | 26.9 (26.6–27.3) | 22.6 (22.4–22.8) | 21.6 (21.4–21.8) | 32 (31.4–32.5) | 30.3 (29.8–30.8) | 29.1 (28.8–29.5) | 27.4 (27.1–27.7) |
| 2 | | 29.6 (29.2–30) | 27.7 (27.4–28.1) | 23.6 (23.4–23.8) | 22.6 (22.4–22.8) | 33 (32.4–33.5) | 31.1 (30.6–31.7) | 29.7 (29.4–30.1) | 28.1 (27.8–28.4) |
| 3 | | 30.3 (29.9–30.7) | 28.4 (28.1–28.8) | 24.5 (24.2–24.7) | 23.5 (23.2–23.7) | 34 (33.4–34.6) | 31.7 (31.1–32.2) | 30.3 (29.9–30.7) | 28.6 (28.3–29) |

**eTable 59. Estimates for marginal predictions of annual disposable income among parents exposed to child's childhood onset disorder (F90–F98) and unexposed parents stratified by the child's age at diagnosis**

| | | Finland | | | | Denmark | | | |
|---|---|---|---|---|---|---|---|---|---|
| | | Men | | Women | | Men | | Women | |
| | | Unexposed | Exposed | Unexposed | Exposed | Unexposed | Exposed | Unexposed | Exposed |
| Years since diagnosis | Age group | Marg. pred. (95% CI) | Marg. pred. (95% CI) | Marg. pred. (95% CI) | Marg. pred. (95% CI) | Marg. pred. (95% CI) | Marg. pred. (95% CI) | Marg. pred. (95% CI) | Marg. pred. (95% CI) |
| | | Unexposed | Exposed | Unexposed | Exposed | Unexposed | Exposed | Unexposed | Exposed |
| 4 | | 31.1 (30.7–31.5) | 28.9 (28.5–29.3) | 25.4 (25.2–25.6) | 24.3 (24.1–24.5) | 35.2 (34.5–35.8) | 32.6 (32–33.2) | 31.1 (30.7–31.4) | 29.1 (28.7–29.5) |
| 5 | | 31.9 (31.4–32.3) | 29.3 (28.9–29.7) | 26.2 (25.9–26.4) | 25.1 (24.8–25.3) | 36.1 (35.4–36.8) | 33.1 (32.5–33.7) | 31.7 (31.2–32.1) | 29.5 (29.1–29.9) |
| -5 | 5–9 years | 25.4 (25.3–25.5) | 23.9 (23.8–24) | 18.8 (18.7–18.9) | 18.5 (18.4–18.6) | 29.7 (29.6–29.9) | 28.4 (28.2–28.5) | 26.4 (26.3–26.5) | 25.9 (25.8–26) |
| -4 | | 26.2 (26.1–26.3) | 24.7 (24.6–24.8) | 20 (19.9–20.1) | 19.6 (19.6–19.7) | 30.7 (30.5–30.9) | 29.2 (29–29.3) | 27.4 (27.3–27.5) | 26.9 (26.8–27) |
| -3 | | 26.9 (26.8–27) | 25.4 (25.3–25.5) | 21.2 (21.1–21.3) | 20.8 (20.7–20.9) | 31.5 (31.4–31.7) | 30 (29.8–30.2) | 28.3 (28.2–28.4) | 27.7 (27.6–27.8) |
| -2 | | 27.6 (27.5–27.8) | 26 (25.9–26.2) | 22.2 (22.1–22.3) | 21.8 (21.7–21.9) | 32.4 (32.2–32.6) | 30.8 (30.6–30.9) | 29.1 (29–29.2) | 28.3 (28.2–28.4) |
| -1 | | 28.3 (28.2–28.5) | 26.7 (26.5–26.8) | 23.1 (23–23.2) | 22.6 (22.5–22.7) | 33.3 (33.1–33.5) | 31.5 (31.4–31.7) | 29.8 (29.7–29.9) | 29 (28.9–29.1) |
| 0 | | 29 (28.9–29.2) | 27.3 (27.1–27.4) | 23.9 (23.8–24) | 23.4 (23.3–23.4) | 34.3 (34–34.5) | 32.3 (32.1–32.5) | 30.5 (30.3–30.6) | 29.4 (29.3–29.6) |
| 1 | | 29.6 (29.5–29.8) | 27.8 (27.7–28) | 24.7 (24.6–24.8) | 24.1 (24–24.2) | 35.1 (34.8–35.3) | 33 (32.8–33.2) | 31.1 (31–31.3) | 30 (29.9–30.1) |
| 2 | | 30.2 (30.1–30.4) | 28.4 (28.3–28.5) | 25.4 (25.3–25.5) | 24.8 (24.7–24.9) | 35.9 (35.6–36.1) | 33.7 (33.5–33.9) | 31.7 (31.6–31.9) | 30.5 (30.3–30.6) |

**eTable 59. Estimates for marginal predictions of annual disposable income among parents exposed to child's childhood onset disorder (F90–F98) and unexposed parents stratified by the child’s age at diagnosis**

| | | Finland | | | | Denmark | | | |
|---|---|---|---|---|---|---|---|---|---|
| | | Men | | Women | | Men | | Women | |
| | | Unexposed | Exposed | Unexposed | Exposed | Unexposed | Exposed | Unexposed | Exposed |
| Years since diagnosis | Age group | Marg. pred. (95% CI) | Marg. pred. (95% CI) | Marg. pred. (95% CI) | Marg. pred. (95% CI) | Marg. pred. (95% CI) | Marg. pred. (95% CI) | Marg. pred. (95% CI) | Marg. pred. (95% CI) |
| | | Unexposed | Exposed | Unexposed | Exposed | Unexposed | Exposed | Unexposed | Exposed |
| 3 | | 30.9 (30.7–31.1) | 28.9 (28.8–29.1) | 26.1 (26–26.2) | 25.5 (25.4–25.6) | 36.6 (36.4–36.9) | 34.3 (34.1–34.5) | 32.3 (32.2–32.5) | 30.9 (30.8–31) |
| 4 | | 31.4 (31.3–31.6) | 29.5 (29.3–29.6) | 26.8 (26.7–26.9) | 26.2 (26.1–26.3) | 37.6 (37.4–37.9) | 35.1 (34.9–35.3) | 32.9 (32.8–33.1) | 31.4 (31.3–31.6) |
| 5 | | 32 (31.9–32.2) | 30 (29.8–30.1) | 27.5 (27.4–27.6) | 26.9 (26.8–27) | 38.3 (38–38.5) | 35.9 (35.6–36.1) | 33.5 (33.3–33.7) | 31.9 (31.8–32.1) |
| -5 | 10–14 years | 27 (26.8–27.1) | 25.5 (25.3–25.6) | 22 (21.9–22.1) | 21.8 (21.7–21.9) | 31.9 (31.7–32.1) | 30.4 (30.2–30.6) | 28.7 (28.6–28.8) | 28.2 (28.1–28.3) |
| -4 | | 27.5 (27.3–27.7) | 26 (25.8–26.2) | 22.7 (22.6–22.8) | 22.5 (22.4–22.6) | 32.8 (32.6–33) | 31 (30.9–31.2) | 29.3 (29.2–29.4) | 28.8 (28.6–28.9) |
| -3 | | 27.9 (27.8–28.1) | 26.4 (26.2–26.6) | 23.4 (23.3–23.5) | 23.2 (23.1–23.3) | 33.6 (33.4–33.8) | 31.8 (31.6–32) | 29.9 (29.8–30) | 29.2 (29.1–29.4) |
| -2 | | 28.5 (28.3–28.7) | 26.9 (26.7–27.1) | 24 (23.9–24.1) | 23.8 (23.7–23.9) | 34.4 (34.2–34.7) | 32.5 (32.3–32.7) | 30.5 (30.4–30.7) | 29.8 (29.7–29.9) |
| -1 | | 29 (28.9–29.2) | 27.4 (27.3–27.6) | 24.6 (24.5–24.7) | 24.4 (24.3–24.5) | 35.3 (35.1–35.6) | 33.1 (32.9–33.3) | 31.2 (31–31.3) | 30.4 (30.2–30.5) |
| 0 | | 29.6 (29.4–29.8) | 27.8 (27.7–28) | 25.2 (25–25.3) | 24.9 (24.8–25) | 36 (35.8–36.3) | 33.9 (33.7–34.1) | 31.8 (31.6–31.9) | 30.8 (30.7–31) |

**eTable 59. Estimates for marginal predictions of annual disposable income among parents exposed to child's childhood onset disorder (F90–F98) and unexposed parents stratified by the child's age at diagnosis**

| | | Finland | | | | Denmark | | | |
|---|---|---|---|---|---|---|---|---|---|
| | | Men | | Women | | Men | | Women | |
| | | Unexposed | Exposed | Unexposed | Exposed | Unexposed | Exposed | Unexposed | Exposed |
| Years since diagnosis | Age group | Marg. pred. (95% CI) | Marg. pred. (95% CI) | Marg. pred. (95% CI) | Marg. pred. (95% CI) | Marg. pred. (95% CI) | Marg. pred. (95% CI) | Marg. pred. (95% CI) | Marg. pred. (95% CI) |
| | | Unexposed | Exposed | Unexposed | Exposed | Unexposed | Exposed | Unexposed | Exposed |
| 1 | | 30.1 (29.9–30.3) | 28.2 (28–28.4) | 25.6 (25.5–25.7) | 25.3 (25.1–25.4) | 36.9 (36.7–37.2) | 34.5 (34.3–34.7) | 32.3 (32.2–32.5) | 31.3 (31.1–31.4) |
| 2 | | 30.4 (30.2–30.6) | 28.6 (28.4–28.8) | 26 (25.9–26.1) | 25.6 (25.4–25.7) | 37.7 (37.4–37.9) | 35.1 (34.8–35.3) | 32.8 (32.6–32.9) | 31.6 (31.5–31.7) |
| 3 | | 30.8 (30.6–31) | 28.8 (28.6–29) | 26.1 (26–26.3) | 25.7 (25.6–25.8) | 38.4 (38.1–38.7) | 35.8 (35.6–36.1) | 33.3 (33.1–33.4) | 31.9 (31.8–32.1) |
| 4 | | 31.1 (30.9–31.3) | 29.2 (29–29.4) | 26.2 (26.1–26.3) | 25.8 (25.6–25.9) | 39 (38.7–39.3) | 36.4 (36.1–36.7) | 33.4 (33.3–33.6) | 32.1 (31.9–32.2) |
| 5 | | 31.5 (31.2–31.7) | 29.5 (29.3–29.7) | 26.2 (26.1–26.3) | 25.8 (25.6–25.9) | 39.7 (39.4–40) | 37.1 (36.8–37.3) | 33.5 (33.4–33.7) | 32.1 (32–32.3) |
| -5 | 15–19 years | 28 (27.7–28.2) | 26.6 (26.3–26.8) | 23.5 (23.3–23.6) | 23.1 (23–23.3) | 33.3 (33–33.5) | 31.4 (31.2–31.7) | 30 (29.8–30.1) | 29.1 (29–29.3) |
| -4 | | 28.4 (28.2–28.7) | 26.9 (26.7–27.1) | 23.9 (23.8–24.1) | 23.7 (23.5–23.8) | 33.9 (33.7–34.2) | 31.9 (31.7–32.2) | 30.5 (30.4–30.7) | 29.6 (29.4–29.7) |
| -3 | | 28.9 (28.7–29.2) | 27.3 (27.1–27.6) | 24.4 (24.2–24.5) | 24.2 (24–24.3) | 34.7 (34.4–34.9) | 32.4 (32.2–32.7) | 31 (30.9–31.2) | 29.9 (29.8–30.1) |
| -2 | | 29.4 (29.2–29.7) | 27.7 (27.5–28) | 24.7 (24.6–24.9) | 24.5 (24.4–24.6) | 35.4 (35.1–35.7) | 33.1 (32.9–33.4) | 31.3 (31.1–31.5) | 30.2 (30.1–30.4) |

**eTable 59. Estimates for marginal predictions of annual disposable income among parents exposed to child's childhood onset disorder (F90–F98) and unexposed parents stratified by the child's age at diagnosis**

| | | Finland | | | | Denmark | | | |
|---|---|---|---|---|---|---|---|---|---|
| | | Men | | Women | | Men | | Women | |
| | | Unexposed | Exposed | Unexposed | Exposed | Unexposed | Exposed | Unexposed | Exposed |
| Years since diagnosis | Age group | Marg. pred. (95% CI) | Marg. pred. (95% CI) | Marg. pred. (95% CI) | Marg. pred. (95% CI) | Marg. pred. (95% CI) | Marg. pred. (95% CI) | Marg. pred. (95% CI) | Marg. pred. (95% CI) |
| | | Unexposed | Exposed | Unexposed | Exposed | Unexposed | Exposed | Unexposed | Exposed |
| -1 | | 29.7 (29.4–30) | 28 (27.8–28.3) | 24.9 (24.8–25.1) | 24.7 (24.6–24.9) | 36 (35.7–36.3) | 33.6 (33.3–33.9) | 31.6 (31.4–31.8) | 30.4 (30.3–30.6) |
| 0 | | 30.2 (29.9–30.5) | 28.4 (28.1–28.6) | 25 (24.8–25.1) | 24.7 (24.6–24.9) | 36.6 (36.3–36.9) | 34.1 (33.8–34.3) | 31.8 (31.6–32) | 30.4 (30.3–30.6) |
| 1 | | 30.4 (30.1–30.7) | 28.5 (28.2–28.7) | 24.9 (24.7–25) | 24.6 (24.4–24.7) | 37.3 (37–37.6) | 34.7 (34.4–34.9) | 31.9 (31.7–32) | 30.4 (30.3–30.6) |
| 2 | | 30.6 (30.3–30.9) | 28.8 (28.5–29.1) | 24.7 (24.5–24.9) | 24.4 (24.2–24.5) | 37.8 (37.4–38.1) | 35.3 (35–35.6) | 31.8 (31.6–32) | 30.3 (30.2–30.5) |
| 3 | | 30.9 (30.6–31.2) | 29 (28.7–29.3) | 24.8 (24.6–24.9) | 24.4 (24.2–24.5) | 38.4 (38–38.7) | 35.8 (35.4–36.1) | 31.7 (31.5–31.9) | 30.3 (30.1–30.5) |
| 4 | | 31 (30.7–31.3) | 29.1 (28.8–29.3) | 24.9 (24.7–25) | 24.4 (24.2–24.6) | 38.8 (38.4–39.2) | 36.3 (35.9–36.6) | 31.8 (31.6–32) | 30.3 (30.1–30.6) |
| 5 | | 31.2 (30.9–31.5) | 29.3 (29–29.6) | 24.9 (24.8–25.1) | 24.5 (24.4–24.7) | 39.3 (38.9–39.7) | 36.7 (36.3–37) | 32 (31.8–32.3) | 30.5 (30.3–30.7) |
| -5 | 20–25 years | 29.7 (29.1–30.3) | 28.5 (27.9–29.1) | 23.9 (23.6–24.3) | 23.7 (23.4–24) | 34.4 (34–34.9) | 32.3 (31.9–32.8) | 29.9 (29.6–30.1) | 28.7 (28.5–29) |
| -4 | | 29.7 (29.1–30.3) | 28.6 (28–29.2) | 23.8 (23.5–24.1) | 23.6 (23.3–23.9) | 34.9 (34.5–35.4) | 32.9 (32.5–33.3) | 30 (29.7–30.2) | 28.7 (28.4–28.9) |

**eTable 59. Estimates for marginal predictions of annual disposable income among parents exposed to child's childhood onset disorder (F90–F98) and unexposed parents stratified by the child's age at diagnosis**

| | | Finland | | | | Denmark | | | |
|---|---|---|---|---|---|---|---|---|---|
| | | Men | | Women | | Men | | Women | |
| | | Unexposed | Exposed | Unexposed | Exposed | Unexposed | Exposed | Unexposed | Exposed |
| Years since diagnosis | Age group | Marg. pred. (95% CI) | Marg. pred. (95% CI) | Marg. pred. (95% CI) | Marg. pred. (95% CI) | Marg. pred. (95% CI) | Marg. pred. (95% CI) | Marg. pred. (95% CI) | Marg. pred. (95% CI) |
| | | Unexposed | Exposed | Unexposed | Exposed | Unexposed | Exposed | Unexposed | Exposed |
| -3 | | 29.9 (29.3–30.5) | 28.8 (28.2–29.4) | 23.8 (23.5–24.1) | 23.5 (23.1–23.8) | 35.5 (35.1–36) | 33.4 (33–33.9) | 30 (29.7–30.2) | 28.6 (28.3–28.8) |
| -2 | | 30.1 (29.5–30.7) | 28.9 (28.3–29.5) | 23.8 (23.5–24.2) | 23.5 (23.2–23.8) | 36.1 (35.6–36.5) | 33.9 (33.5–34.3) | 30 (29.7–30.2) | 28.6 (28.4–28.9) |
| -1 | | 30.2 (29.6–30.8) | 28.8 (28.2–29.4) | 23.9 (23.5–24.2) | 23.6 (23.3–23.9) | 36.5 (36–37) | 34.2 (33.8–34.6) | 30.1 (29.8–30.4) | 28.8 (28.5–29) |
| 0 | | 30.1 (29.5–30.7) | 28.9 (28.3–29.5) | 24 (23.6–24.3) | 23.6 (23.3–24) | 37 (36.5–37.5) | 34.7 (34.2–35.2) | 30.2 (30–30.5) | 28.9 (28.6–29.1) |
| 1 | | 30.3 (29.6–30.9) | 29 (28.4–29.6) | 24.1 (23.7–24.5) | 23.5 (23.1–23.9) | 37.5 (37–38) | 35.1 (34.6–35.6) | 30.5 (30.2–30.8) | 29.1 (28.8–29.4) |
| 2 | | 30.3 (29.6–31) | 29 (28.4–29.7) | 24.1 (23.7–24.4) | 23.7 (23.4–24.1) | 37.7 (37.2–38.3) | 35.5 (35–36) | 30.7 (30.4–31) | 29.3 (29–29.5) |
| 3 | | 30.4 (29.7–31) | 29.1 (28.5–29.8) | 24.3 (23.9–24.6) | 23.8 (23.4–24.1) | 38 (37.5–38.6) | 35.7 (35.2–36.2) | 30.8 (30.5–31.2) | 29.3 (29–29.6) |
| 4 | | 30.5 (29.8–31.2) | 29 (28.3–29.7) | 24.5 (24.1–24.9) | 23.6 (23.2–24) | 38.2 (37.6–38.8) | 35.9 (35.3–36.4) | 31 (30.7–31.4) | 29.4 (29.1–29.8) |
| 5 | | 30 (29.2–30.7) | 29.3 (28.6–30) | 24.5 (24.1–24.9) | 23.7 (23.3–24.1) | 38.4 (37.8–39) | 36.1 (35.5–36.7) | 31.3 (31–31.7) | 29.6 (29.3–29.9) |

Marg. pred. = Marginal predictions in thousand euros (€)

**eTable 60. Estimates for marginal predictions of annual social welfare benefits among parents exposed to child's childhood onset disorder (F90–F98) and unexposed parents stratified by the child's age at diagnosis**

| | | Finland | | | | Denmark | | | |
|---|---|---|---|---|---|---|---|---|---|
| | | Men | | Women | | Men | | Women | |
| | | Unexposed | Exposed | Unexposed | Exposed | Unexposed | Exposed | Unexposed | Exposed |
| Years since diagnosis | Age group | Marg. pred. (95% CI) | Marg. pred. (95% CI) | Marg. pred. (95% CI) | Marg. pred. (95% CI) | Marg. pred. (95% CI) | Marg. pred. (95% CI) | Marg. pred. (95% CI) | Marg. pred. (95% CI) |
| | | Unexposed | Exposed | Unexposed | Exposed | Unexposed | Exposed | Unexposed | Exposed |
| -5 | 1–4 years | 2.6 (2.5–2.7) | 3.1 (3–3.2) | 7.1 (7–7.2) | 7.2 (6.8–7.5) | 4.4 (4.2–4.7) | 5.1 (4.9–5.4) | 9.6 (9.3–9.9) | 10.2 (9.9–10.6) |
| -4 | | 3 (2.9–3.1) | 3.5 (3.3–3.6) | 10.6 (10.4–10.7) | 10.6 (10.4–10.7) | 4.6 (4.4–4.9) | 5.5 (5.3–5.8) | 13.1 (12.8–13.4) | 13.9 (13.6–14.3) |
| -3 | | 3.1 (3–3.2) | 3.6 (3.5–3.8) | 10.9 (10.7–11) | 11.2 (11–11.3) | 4.8 (4.5–5) | 5.7 (5.4–6) | 13.4 (13.1–13.7) | 14.8 (14.5–15.2) |
| -2 | | 3.1 (2.9–3.2) | 3.6 (3.5–3.7) | 10.5 (10.4–10.7) | 11.2 (11–11.3) | 4.7 (4.4–4.9) | 5.7 (5.4–6) | 13.7 (13.3–14) | 15.3 (14.9–15.6) |
| -1 | | 3.1 (2.9–3.2) | 3.7 (3.6–3.9) | 10.5 (10.3–10.6) | 11.3 (11.2–11.5) | 4.7 (4.4–4.9) | 5.7 (5.5–6) | 13.7 (13.3–14) | 15.6 (15.3–16) |
| 0 | | 3.1 (2.9–3.2) | 3.8 (3.6–3.9) | 9.7 (9.6–9.9) | 10.8 (10.7–11) | 4.7 (4.4–4.9) | 5.9 (5.6–6.2) | 13.3 (12.9–13.6) | 15.6 (15.2–15.9) |
| 1 | | 3 (2.9–3.1) | 3.8 (3.6–3.9) | 9.3 (9.1–9.5) | 10.3 (10.2–10.5) | 4.6 (4.3–4.9) | 6 (5.7–6.3) | 12.8 (12.4–13.1) | 15.1 (14.8–15.5) |
| 2 | | 3.1 (3–3.2) | 3.9 (3.7–4) | 9 (8.8–9.2) | 10.2 (10–10.3) | 4.7 (4.4–4.9) | 6.1 (5.8–6.4) | 12.5 (12.2–12.9) | 15.2 (14.8–15.5) |
| 3 | | 3.1 (3–3.2) | 3.9 (3.8–4.1) | 8.9 (8.7–9) | 10.1 (10–10.3) | 4.7 (4.4–5) | 6.3 (6–6.6) | 12.1 (11.8–12.5) | 14.8 (14.4–15.2) |
| 4 | | 3.2 (3.1–3.4) | 4.1 (4–4.3) | 8.8 (8.6–9) | 10.1 (9.9–10.3) | 4.7 (4.4–5) | 6.3 (6–6.7) | 11.9 (11.5–12.3) | 14.6 (14.2–15) |

**eTable 60. Estimates for marginal predictions of annual social welfare benefits among parents exposed to child's childhood onset disorder (F90–F98) and unexposed parents stratified by the child's age at diagnosis**

| | | Finland | | | | Denmark | | | |
|---|---|---|---|---|---|---|---|---|---|
| | | Men | | Women | | Men | | Women | |
| | | Unexposed | Exposed | Unexposed | Exposed | Unexposed | Exposed | Unexposed | Exposed |
| Years since diagnosis | Age group | Marg. pred. (95% CI) | Marg. pred. (95% CI) | Marg. pred. (95% CI) | Marg. pred. (95% CI) | Marg. pred. (95% CI) | Marg. pred. (95% CI) | Marg. pred. (95% CI) | Marg. pred. (95% CI) |
| | | Unexposed | Exposed | Unexposed | Exposed | Unexposed | Exposed | Unexposed | Exposed |
| 5 | | 3.4 (3.2–3.6) | 4.2 (4–4.4) | 8.6 (8.4–8.8) | 10 (9.8–10.2) | 4.8 (4.5–5.1) | 6.6 (6.2–7) | 11.6 (11.2–12) | 14.5 (14–14.9) |
| -5 | 5–9 years | 2.9 (2.9–3) | 3.3 (3.3–3.4) | 10.5 (10.4–10.5) | 10.7 (10.7–10.8) | 4.3 (4.2–4.4) | 4.6 (4.5–4.7) | 13 (12.9–13.1) | 14 (13.9–14.1) |
| -4 | | 3 (2.9–3) | 3.4 (3.3–3.4) | 9.7 (9.6–9.8) | 10.1 (10.1–10.2) | 4.3 (4.2–4.4) | 4.7 (4.6–4.8) | 12.6 (12.5–12.7) | 13.7 (13.6–13.8) |
| -3 | | 3 (3–3.1) | 3.5 (3.4–3.5) | 9.3 (9.3–9.4) | 10 (9.9–10.1) | 4.3 (4.2–4.4) | 4.7 (4.6–4.8) | 12.3 (12.2–12.4) | 13.6 (13.4–13.7) |
| -2 | | 3.1 (3–3.1) | 3.5 (3.5–3.6) | 9.1 (9–9.1) | 9.9 (9.8–9.9) | 4.4 (4.3–4.5) | 4.9 (4.8–5) | 12 (11.9–12.2) | 13.4 (13.3–13.5) |
| -1 | | 3.1 (3–3.2) | 3.6 (3.5–3.6) | 8.9 (8.9–9) | 9.8 (9.8–9.9) | 4.4 (4.3–4.5) | 4.9 (4.8–5) | 11.8 (11.6–11.9) | 13.3 (13.1–13.4) |
| 0 | | 3.1 (3.1–3.2) | 3.6 (3.6–3.7) | 8.7 (8.6–8.8) | 9.8 (9.7–9.8) | 4.4 (4.3–4.5) | 5 (4.9–5.1) | 11.5 (11.4–11.6) | 13.2 (13–13.3) |
| 1 | | 3.2 (3.2–3.3) | 3.8 (3.7–3.8) | 8.5 (8.4–8.6) | 9.7 (9.6–9.8) | 4.5 (4.4–4.6) | 5.1 (5–5.2) | 11.2 (11.1–11.3) | 13 (12.9–13.2) |
| 2 | | 3.3 (3.3–3.4) | 3.9 (3.8–4) | 8.4 (8.3–8.4) | 9.5 (9.5–9.6) | 4.6 (4.5–4.7) | 5.3 (5.2–5.4) | 10.9 (10.8–11.1) | 12.8 (12.7–13) |
| 3 | | 3.5 (3.4–3.5) | 4.1 (4–4.2) | 8.2 (8.1–8.2) | 9.4 (9.3–9.5) | 4.6 (4.5–4.8) | 5.4 (5.3–5.5) | 10.7 (10.6–10.9) | 12.6 (12.5–12.8) |

**eTable 60. Estimates for marginal predictions of annual social welfare benefits among parents exposed to child's childhood onset disorder (F90–F98) and unexposed parents stratified by the child's age at diagnosis**

| | | Finland | | | | Denmark | | | |
|---|---|---|---|---|---|---|---|---|---|
| | | Men | | Women | | Men | | Women | |
| | | Unexposed | Exposed | Unexposed | Exposed | Unexposed | Exposed | Unexposed | Exposed |
| Years since diagnosis | Age group | Marg. pred. (95% CI) | Marg. pred. (95% CI) | Marg. pred. (95% CI) | Marg. pred. (95% CI) | Marg. pred. (95% CI) | Marg. pred. (95% CI) | Marg. pred. (95% CI) | Marg. pred. (95% CI) |
| | | Unexposed | Exposed | Unexposed | Exposed | Unexposed | Exposed | Unexposed | Exposed |
| 4 | | 3.6 (3.5–3.7) | 4.2 (4.2–4.3) | 8 (7.9–8) | 9.3 (9.2–9.4) | 4.7 (4.6–4.8) | 5.6 (5.5–5.7) | 10.5 (10.4–10.7) | 12.4 (12.3–12.6) |
| 5 | | 3.7 (3.6–3.8) | 4.4 (4.4–4.5) | 7.8 (7.7–7.8) | 9.2 (9.1–9.3) | 4.8 (4.7–5) | 5.7 (5.6–5.8) | 10.4 (10.3–10.5) | 12.3 (12.2–12.5) |
| -5 | 10–14 years | 3 (3–3.1) | 3.6 (3.5–3.7) | 8.4 (8.3–8.5) | 9.3 (9.2–9.3) | 4.2 (4.1–4.3) | 4.6 (4.5–4.7) | 11.3 (11.2–11.4) | 12.5 (12.3–12.6) |
| -4 | | 3.2 (3.1–3.2) | 3.7 (3.6–3.8) | 8.3 (8.2–8.4) | 9.2 (9.1–9.3) | 4.2 (4.1–4.3) | 4.7 (4.6–4.8) | 11 (10.9–11.1) | 12.3 (12.1–12.4) |
| -3 | | 3.3 (3.3–3.4) | 3.9 (3.9–4) | 8.2 (8.1–8.3) | 9.3 (9.2–9.4) | 4.3 (4.2–4.4) | 4.8 (4.7–4.9) | 10.8 (10.6–10.9) | 12.1 (11.9–12.2) |
| -2 | | 3.4 (3.3–3.5) | 4 (3.9–4.1) | 8.1 (8–8.2) | 9.1 (9.1–9.2) | 4.4 (4.3–4.5) | 4.9 (4.7–5) | 10.6 (10.5–10.7) | 12 (11.8–12.1) |
| -1 | | 3.5 (3.5–3.6) | 4.2 (4.1–4.2) | 7.9 (7.8–8) | 9.1 (9–9.2) | 4.5 (4.4–4.6) | 5 (4.9–5.1) | 10.4 (10.3–10.6) | 11.9 (11.7–12) |
| 0 | | 3.6 (3.5–3.7) | 4.3 (4.2–4.4) | 7.7 (7.6–7.7) | 9 (8.9–9.1) | 4.6 (4.5–4.7) | 5.2 (5.1–5.3) | 10.3 (10.2–10.5) | 11.8 (11.7–12) |
| 1 | | 3.7 (3.7–3.8) | 4.5 (4.4–4.6) | 7.5 (7.4–7.6) | 8.8 (8.7–8.9) | 4.7 (4.6–4.8) | 5.4 (5.2–5.5) | 10.2 (10–10.3) | 11.7 (11.6–11.8) |
| 2 | | 3.9 (3.9–4) | 4.7 (4.6–4.8) | 7.3 (7.2–7.3) | 8.6 (8.5–8.7) | 4.8 (4.7–4.9) | 5.4 (5.3–5.6) | 9.9 (9.8–10.1) | 11.5 (11.4–11.7) |

**eTable 60. Estimates for marginal predictions of annual social welfare benefits among parents exposed to child's childhood onset disorder (F90–F98) and unexposed parents stratified by the child's age at diagnosis**

| | | Finland | | | | Denmark | | | |
|---|---|---|---|---|---|---|---|---|---|
| | | Men | | Women | | Men | | Women | |
| | | Unexposed | Exposed | Unexposed | Exposed | Unexposed | Exposed | Unexposed | Exposed |
| Years since diagnosis | Age group | Marg. pred. (95% CI) | Marg. pred. (95% CI) | Marg. pred. (95% CI) | Marg. pred. (95% CI) | Marg. pred. (95% CI) | Marg. pred. (95% CI) | Marg. pred. (95% CI) | Marg. pred. (95% CI) |
| | | Unexposed | Exposed | Unexposed | Exposed | Unexposed | Exposed | Unexposed | Exposed |
| 3 | | 4.2 (4.1–4.2) | 4.8 (4.7–4.9) | 6.8 (6.8–6.9) | 8.2 (8.1–8.3) | 4.9 (4.8–5.1) | 5.6 (5.4–5.7) | 9.7 (9.5–9.8) | 11.3 (11.1–11.4) |
| 4 | | 4.3 (4.3–4.4) | 5.1 (5–5.2) | 6.4 (6.3–6.5) | 7.8 (7.7–7.9) | 5.1 (4.9–5.2) | 5.7 (5.6–5.8) | 9.2 (9.1–9.4) | 10.9 (10.7–11) |
| 5 | | 4.6 (4.5–4.7) | 5.3 (5.2–5.4) | 5.9 (5.8–6) | 7.4 (7.3–7.5) | 5.1 (5–5.3) | 5.8 (5.7–5.9) | 8.8 (8.6–8.9) | 10.4 (10.3–10.6) |
| -5 | 15–19 years | 3.2 (3.1–3.3) | 3.8 (3.7–3.9) | 7.4 (7.3–7.5) | 8.2 (8.1–8.3) | 4.6 (4.4–4.7) | 5.4 (5.3–5.6) | 10.5 (10.3–10.7) | 12.1 (12–12.3) |
| -4 | | 3.4 (3.3–3.5) | 4 (3.9–4.1) | 7.3 (7.2–7.4) | 8.1 (8–8.2) | 4.7 (4.6–4.9) | 5.6 (5.5–5.7) | 10.4 (10.2–10.5) | 12.1 (11.9–12.2) |
| -3 | | 3.5 (3.4–3.6) | 4.2 (4.1–4.3) | 7.1 (7–7.2) | 8 (7.9–8.1) | 4.9 (4.7–5) | 5.7 (5.6–5.9) | 10.2 (10–10.4) | 12 (11.8–12.1) |
| -2 | | 3.7 (3.5–3.8) | 4.4 (4.3–4.5) | 6.8 (6.7–6.9) | 7.7 (7.6–7.8) | 5 (4.9–5.2) | 5.9 (5.7–6) | 10 (9.9–10.2) | 11.7 (11.6–11.9) |
| -1 | | 3.8 (3.7–3.9) | 4.6 (4.5–4.7) | 6.5 (6.4–6.6) | 7.4 (7.3–7.6) | 5.1 (5–5.3) | 6.1 (5.9–6.2) | 9.8 (9.6–9.9) | 11.5 (11.3–11.7) |
| 0 | | 4 (3.8–4.1) | 4.8 (4.7–4.9) | 6 (5.9–6.1) | 7 (6.9–7.2) | 5.3 (5.2–5.5) | 6.3 (6.1–6.4) | 9.4 (9.2–9.6) | 11.3 (11.1–11.4) |
| 1 | | 4.1 (4–4.3) | 5 (4.9–5.1) | 5.5 (5.4–5.6) | 6.6 (6.4–6.7) | 5.4 (5.2–5.5) | 6.4 (6.3–6.6) | 9 (8.8–9.1) | 10.8 (10.6–11) |

**eTable 60. Estimates for marginal predictions of annual social welfare benefits among parents exposed to child's childhood onset disorder (F90–F98) and unexposed parents stratified by the child's age at diagnosis**

| | | Finland | | | | Denmark | | | |
|---|---|---|---|---|---|---|---|---|---|
| | | Men | | Women | | Men | | Women | |
| | | Unexposed | Exposed | Unexposed | Exposed | Unexposed | Exposed | Unexposed | Exposed |
| Years since diagnosis | Age group | Marg. pred. (95% CI) | Marg. pred. (95% CI) | Marg. pred. (95% CI) | Marg. pred. (95% CI) | Marg. pred. (95% CI) | Marg. pred. (95% CI) | Marg. pred. (95% CI) | Marg. pred. (95% CI) |
| | | Unexposed | Exposed | Unexposed | Exposed | Unexposed | Exposed | Unexposed | Exposed |
| 2 | | 4.4 (4.3–4.5) | 5.2 (5.1–5.4) | 4.9 (4.8–5.1) | 6.1 (5.9–6.2) | 5.6 (5.4–5.7) | 6.5 (6.3–6.7) | 8.4 (8.3–8.6) | 10.3 (10.1–10.5) |
| 3 | | 4.6 (4.5–4.8) | 5.5 (5.3–5.6) | 4.6 (4.5–4.7) | 5.8 (5.6–5.9) | 5.7 (5.5–5.8) | 6.6 (6.4–6.8) | 8 (7.8–8.2) | 9.9 (9.7–10.1) |
| 4 | | 5 (4.9–5.2) | 5.9 (5.7–6) | 4.5 (4.4–4.7) | 5.7 (5.6–5.8) | 5.8 (5.7–6) | 6.8 (6.7–7) | 7.9 (7.7–8.1) | 9.8 (9.6–10) |
| 5 | | 5.5 (5.4–5.7) | 6.3 (6.1–6.5) | 4.6 (4.5–4.7) | 5.7 (5.5–5.8) | 6.1 (5.9–6.3) | 7.2 (7–7.4) | 7.9 (7.7–8.1) | 10 (9.8–10.2) |
| -5 | 20–25 years | 4.1 (3.8–4.3) | 4.4 (4.1–4.7) | 5.1 (4.9–5.3) | 5.7 (5.4–5.9) | 4.8 (4.6–5) | 5.9 (5.7–6.2) | 8.3 (8.1–8.6) | 10.2 (9.9–10.4) |
| -4 | | 4.4 (4.1–4.7) | 4.7 (4.5–5) | 4.9 (4.7–5.2) | 5.5 (5.3–5.7) | 4.9 (4.7–5.1) | 6.2 (5.9–6.4) | 8 (7.7–8.2) | 9.9 (9.6–10.2) |
| -3 | | 4.7 (4.4–5) | 5 (4.7–5.3) | 4.7 (4.5–4.9) | 5.2 (5–5.4) | 5.1 (4.9–5.3) | 6.3 (6.1–6.6) | 7.6 (7.4–7.9) | 9.7 (9.4–10) |
| -2 | | 5.1 (4.8–5.4) | 5.4 (5.1–5.7) | 4.6 (4.3–4.8) | 5.1 (4.8–5.3) | 5.3 (5.1–5.5) | 6.6 (6.3–6.8) | 7.4 (7.2–7.7) | 9.3 (9.1–9.6) |
| -1 | | 5.4 (5.1–5.7) | 5.8 (5.5–6.2) | 4.5 (4.3–4.7) | 5.1 (4.9–5.3) | 5.5 (5.3–5.7) | 6.8 (6.6–7.1) | 7.3 (7.1–7.6) | 9.4 (9.1–9.7) |
| 0 | | 6.1 (5.8–6.5) | 6.1 (5.7–6.4) | 4.5 (4.3–4.7) | 5.3 (5.1–5.5) | 5.9 (5.6–6.1) | 7.2 (6.9–7.4) | 7.3 (7.1–7.6) | 9.6 (9.3–9.8) |
| 1 | | 6.7 (6.3–7.1) | 6.8 (6.4–7.1) | 4.7 (4.4–4.9) | 5.6 (5.3–5.9) | 6.3 (6–6.5) | 7.5 (7.2–7.8) | 7.5 (7.3–7.8) | 9.7 (9.4–10) |
| 2 | | 7.3 (6.9–7.7) | 7.5 (7–7.9) | 4.9 (4.7–5.2) | 5.9 (5.7–6.2) | 6.6 (6.4–6.9) | 7.8 (7.5–8.1) | 7.8 (7.5–8.1) | 10 (9.7–10.3) |
| 3 | | 7.9 (7.5–8.4) | 8.4 (8–8.8) | 5.3 (5–5.6) | 6.2 (5.9–6.5) | 6.9 (6.6–7.1) | 8 (7.7–8.3) | 8.1 (7.8–8.4) | 10.2 (9.9–10.6) |

**eTable 60. Estimates for marginal predictions of annual social welfare benefits among parents exposed to child's childhood onset disorder (F90–F98) and unexposed parents stratified by the child’s age at diagnosis**

| | | Finland | | | | Denmark | | | |
|---|---|---|---|---|---|---|---|---|---|
| | | Men | | Women | | Men | | Women | |
| | | Unexposed | Exposed | Unexposed | Exposed | Unexposed | Exposed | Unexposed | Exposed |
| Years since diagnosis | Age group | Marg. pred. (95% CI) | Marg. pred. (95% CI) | Marg. pred. (95% CI) | Marg. pred. (95% CI) | Marg. pred. (95% CI) | Marg. pred. (95% CI) | Marg. pred. (95% CI) | Marg. pred. (95% CI) |
| | | Unexposed | Exposed | Unexposed | Exposed | Unexposed | Exposed | Unexposed | Exposed |
| 4 | | 8.8 (8.3–9.4) | 9.1 (8.6–9.6) | 5.8 (5.5–6.2) | 6.6 (6.2–6.9) | 7.4 (7.1–7.7) | 8.3 (8–8.6) | 8.4 (8.1–8.7) | 10.5 (10.1–10.8) |
| 5 | | 9.9 (9.3–10.4) | 10.1 (9.6–10.7) | 6.5 (6.1–6.9) | 7.2 (6.8–7.7) | 8 (7.7–8.3) | 9 (8.6–9.3) | 8.6 (8.3–8.9) | 10.8 (10.4–11.1) |

Marg. pred. = Marginal predictions in thousand euros (€)

**eTable 61. Estimates for marginal predictions of annual employment among parents exposed to child's childhood onset disorder (F90–F98) and unexposed parents stratified by the child's age at diagnosis**

| | | Finland | | | | Denmark | | | |
|---|---|---|---|---|---|---|---|---|---|
| | | Men | | Women | | Men | | Women | |
| | | Unexposed | Exposed | Unexposed | Exposed | Unexposed | Exposed | Unexposed | Exposed |
| Years since diagnosis | Age group | Marg. pred. (95% CI) | Marg. pred. (95% CI) | Marg. pred. (95% CI) | Marg. pred. (95% CI) | Marg. pred. (95% CI) | Marg. pred. (95% CI) | Marg. pred. (95% CI) | Marg. pred. (95% CI) |
| | | Unexposed | Exposed | Unexposed | Exposed | Unexposed | Exposed | Unexposed | Exposed |
| -5 | 1–4 years | 0.77 (0.76–0.78) | 0.72 (0.71–0.73) | 0.59 (0.58–0.6) | 0.56 (0.55–0.57) | 0.64 (0.62–0.65) | 0.62 (0.6–0.63) | 0.52 (0.5–0.53) | 0.47 (0.45–0.48) |
| -4 | | 0.79 (0.78–0.79) | 0.74 (0.73–0.75) | 0.54 (0.53–0.55) | 0.5 (0.49–0.51) | 0.67 (0.66–0.69) | 0.64 (0.62–0.65) | 0.51 (0.49–0.52) | 0.44 (0.43–0.46) |
| -3 | | 0.8 (0.79–0.8) | 0.75 (0.74–0.76) | 0.55 (0.54–0.56) | 0.5 (0.49–0.51) | 0.69 (0.67–0.7) | 0.65 (0.64–0.67) | 0.53 (0.52–0.55) | 0.46 (0.45–0.48) |
| -2 | | 0.81 (0.8–0.81) | 0.76 (0.75–0.77) | 0.57 (0.56–0.58) | 0.51 (0.5–0.52) | 0.69 (0.67–0.7) | 0.66 (0.64–0.67) | 0.55 (0.53–0.56) | 0.48 (0.46–0.49) |
| -1 | | 0.81 (0.8–0.82) | 0.76 (0.75–0.77) | 0.61 (0.6–0.62) | 0.54 (0.53–0.55) | 0.7 (0.68–0.71) | 0.68 (0.66–0.69) | 0.57 (0.55–0.58) | 0.49 (0.48–0.51) |
| 0 | | 0.82 (0.81–0.83) | 0.77 (0.76–0.78) | 0.65 (0.64–0.66) | 0.56 (0.55–0.58) | 0.71 (0.69–0.72) | 0.67 (0.66–0.69) | 0.59 (0.57–0.6) | 0.48 (0.47–0.5) |
| 1 | | 0.81 (0.8–0.82) | 0.77 (0.76–0.78) | 0.68 (0.67–0.69) | 0.6 (0.59–0.61) | 0.7 (0.69–0.72) | 0.68 (0.66–0.69) | 0.61 (0.59–0.62) | 0.5 (0.48–0.52) |
| 2 | | 0.82 (0.81–0.83) | 0.77 (0.76–0.78) | 0.71 (0.7–0.72) | 0.63 (0.61–0.64) | 0.71 (0.69–0.72) | 0.66 (0.65–0.68) | 0.62 (0.6–0.64) | 0.5 (0.49–0.52) |
| 3 | | 0.83 (0.82–0.83) | 0.77 (0.76–0.78) | 0.73 (0.72–0.74) | 0.64 (0.63–0.66) | 0.71 (0.69–0.72) | 0.66 (0.65–0.68) | 0.63 (0.61–0.65) | 0.52 (0.5–0.53) |

**eTable 61. Estimates for marginal predictions of annual employment among parents exposed to child's childhood onset disorder (F90–F98) and unexposed parents stratified by the child's age at diagnosis**

| | | Finland | | | | Denmark | | | |
|---|---|---|---|---|---|---|---|---|---|
| | | Men | | Women | | Men | | Women | |
| | | Unexposed | Exposed | Unexposed | Exposed | Unexposed | Exposed | Unexposed | Exposed |
| Years since diagnosis | Age group | Marg. pred. (95% CI) | Marg. pred. (95% CI) | Marg. pred. (95% CI) | Marg. pred. (95% CI) | Marg. pred. (95% CI) | Marg. pred. (95% CI) | Marg. pred. (95% CI) | Marg. pred. (95% CI) |
| | | Unexposed | Exposed | Unexposed | Exposed | Unexposed | Exposed | Unexposed | Exposed |
| 4 | | 0.82 (0.81–0.83) | 0.76 (0.75–0.77) | 0.75 (0.74–0.76) | 0.66 (0.65–0.67) | 0.71 (0.69–0.73) | 0.67 (0.65–0.68) | 0.64 (0.63–0.66) | 0.53 (0.51–0.54) |
| 5 | | 0.82 (0.81–0.83) | 0.76 (0.75–0.77) | 0.76 (0.75–0.77) | 0.67 (0.66–0.68) | 0.7 (0.69–0.72) | 0.66 (0.64–0.68) | 0.64 (0.63–0.66) | 0.52 (0.51–0.54) |
| -5 | 5–9 years | 0.81 (0.81–0.81) | 0.77 (0.77–0.78) | 0.58 (0.58–0.59) | 0.55 (0.55–0.56) | 0.71 (0.7–0.71) | 0.7 (0.69–0.7) | 0.58 (0.57–0.58) | 0.56 (0.56–0.57) |
| -4 | | 0.81 (0.81–0.82) | 0.78 (0.77–0.78) | 0.62 (0.62–0.63) | 0.59 (0.59–0.59) | 0.72 (0.72–0.73) | 0.71 (0.7–0.72) | 0.61 (0.6–0.62) | 0.58 (0.58–0.59) |
| -3 | | 0.82 (0.81–0.82) | 0.78 (0.77–0.78) | 0.66 (0.65–0.66) | 0.62 (0.61–0.62) | 0.73 (0.72–0.73) | 0.72 (0.71–0.72) | 0.63 (0.62–0.63) | 0.6 (0.59–0.6) |
| -2 | | 0.82 (0.81–0.82) | 0.78 (0.77–0.78) | 0.69 (0.68–0.69) | 0.64 (0.64–0.65) | 0.73 (0.72–0.73) | 0.72 (0.71–0.72) | 0.64 (0.64–0.65) | 0.61 (0.6–0.61) |
| -1 | | 0.82 (0.82–0.82) | 0.78 (0.78–0.79) | 0.71 (0.71–0.71) | 0.66 (0.65–0.66) | 0.73 (0.73–0.74) | 0.71 (0.71–0.72) | 0.65 (0.64–0.66) | 0.61 (0.61–0.62) |
| 0 | | 0.82 (0.82–0.82) | 0.79 (0.78–0.79) | 0.73 (0.73–0.74) | 0.68 (0.67–0.68) | 0.73 (0.73–0.74) | 0.72 (0.71–0.72) | 0.66 (0.65–0.66) | 0.61 (0.61–0.62) |
| 1 | | 0.82 (0.82–0.82) | 0.78 (0.78–0.78) | 0.75 (0.75–0.75) | 0.69 (0.69–0.7) | 0.73 (0.72–0.73) | 0.71 (0.71–0.72) | 0.67 (0.66–0.67) | 0.61 (0.61–0.62) |

**eTable 61. Estimates for marginal predictions of annual employment among parents exposed to child's childhood onset disorder (F90–F98) and unexposed parents stratified by the child's age at diagnosis**

| | | Finland | | | | Denmark | | | |
|---|---|---|---|---|---|---|---|---|---|
| | | Men | | Women | | Men | | Women | |
| | | Unexposed | Exposed | Unexposed | Exposed | Unexposed | Exposed | Unexposed | Exposed |
| Years since diagnosis | Age group | Marg. pred. (95% CI) | Marg. pred. (95% CI) | Marg. pred. (95% CI) | Marg. pred. (95% CI) | Marg. pred. (95% CI) | Marg. pred. (95% CI) | Marg. pred. (95% CI) | Marg. pred. (95% CI) |
| | | Unexposed | Exposed | Unexposed | Exposed | Unexposed | Exposed | Unexposed | Exposed |
| 2 | | 0.82 (0.81–0.82) | 0.78 (0.78–0.78) | 0.76 (0.76–0.77) | 0.71 (0.7–0.71) | 0.72 (0.72–0.73) | 0.71 (0.7–0.71) | 0.68 (0.67–0.68) | 0.61 (0.61–0.62) |
| 3 | | 0.82 (0.81–0.82) | 0.77 (0.77–0.78) | 0.78 (0.77–0.78) | 0.72 (0.72–0.73) | 0.72 (0.72–0.73) | 0.7 (0.7–0.71) | 0.68 (0.67–0.68) | 0.62 (0.61–0.62) |
| 4 | | 0.81 (0.81–0.81) | 0.77 (0.77–0.77) | 0.79 (0.79–0.8) | 0.73 (0.73–0.74) | 0.72 (0.71–0.72) | 0.7 (0.69–0.7) | 0.69 (0.68–0.69) | 0.62 (0.62–0.63) |
| 5 | | 0.81 (0.8–0.81) | 0.77 (0.76–0.77) | 0.8 (0.8–0.8) | 0.75 (0.74–0.75) | 0.71 (0.7–0.71) | 0.7 (0.69–0.7) | 0.68 (0.68–0.69) | 0.62 (0.61–0.63) |
| -5 | 10–14 years | 0.81 (0.81–0.81) | 0.77 (0.76–0.77) | 0.71 (0.71–0.72) | 0.67 (0.66–0.67) | 0.72 (0.71–0.72) | 0.71 (0.71–0.72) | 0.64 (0.63–0.65) | 0.62 (0.61–0.62) |
| -4 | | 0.81 (0.8–0.81) | 0.76 (0.76–0.77) | 0.73 (0.72–0.73) | 0.68 (0.68–0.69) | 0.73 (0.73–0.74) | 0.72 (0.71–0.73) | 0.66 (0.65–0.67) | 0.64 (0.63–0.64) |
| -3 | | 0.8 (0.8–0.81) | 0.76 (0.75–0.76) | 0.74 (0.73–0.74) | 0.7 (0.69–0.7) | 0.74 (0.73–0.74) | 0.72 (0.71–0.73) | 0.67 (0.67–0.68) | 0.65 (0.64–0.65) |
| -2 | | 0.8 (0.8–0.81) | 0.76 (0.76–0.76) | 0.75 (0.75–0.76) | 0.71 (0.7–0.71) | 0.73 (0.73–0.74) | 0.72 (0.72–0.73) | 0.68 (0.67–0.68) | 0.65 (0.65–0.66) |
| -1 | | 0.8 (0.8–0.81) | 0.76 (0.75–0.76) | 0.76 (0.76–0.77) | 0.72 (0.71–0.72) | 0.73 (0.72–0.74) | 0.72 (0.71–0.72) | 0.68 (0.68–0.69) | 0.65 (0.65–0.66) |

**eTable 61. Estimates for marginal predictions of annual employment among parents exposed to child's childhood onset disorder (F90–F98) and unexposed parents stratified by the child's age at diagnosis**

| | | Finland | | | | Denmark | | | |
|---|---|---|---|---|---|---|---|---|---|
| | | Men | | Women | | Men | | Women | |
| | | Unexposed | Exposed | Unexposed | Exposed | Unexposed | Exposed | Unexposed | Exposed |
| Years since diagnosis | Age group | Marg. pred. (95% CI) | Marg. pred. (95% CI) | Marg. pred. (95% CI) | Marg. pred. (95% CI) | Marg. pred. (95% CI) | Marg. pred. (95% CI) | Marg. pred. (95% CI) | Marg. pred. (95% CI) |
| | | Unexposed | Exposed | Unexposed | Exposed | Unexposed | Exposed | Unexposed | Exposed |
| 0 | | 0.8 (0.8–0.81) | 0.76 (0.75–0.76) | 0.77 (0.77–0.78) | 0.72 (0.72–0.73) | 0.73 (0.72–0.73) | 0.71 (0.7–0.72) | 0.69 (0.68–0.69) | 0.65 (0.65–0.66) |
| 1 | | 0.8 (0.79–0.8) | 0.75 (0.75–0.76) | 0.78 (0.78–0.79) | 0.73 (0.72–0.73) | 0.72 (0.72–0.73) | 0.71 (0.7–0.72) | 0.69 (0.68–0.69) | 0.65 (0.64–0.65) |
| 2 | | 0.79 (0.78–0.79) | 0.74 (0.74–0.75) | 0.79 (0.78–0.79) | 0.74 (0.73–0.74) | 0.72 (0.71–0.73) | 0.7 (0.7–0.71) | 0.69 (0.68–0.69) | 0.65 (0.64–0.66) |
| 3 | | 0.78 (0.78–0.79) | 0.74 (0.73–0.74) | 0.79 (0.79–0.8) | 0.74 (0.74–0.75) | 0.72 (0.71–0.72) | 0.7 (0.69–0.71) | 0.69 (0.69–0.7) | 0.65 (0.64–0.66) |
| 4 | | 0.77 (0.77–0.78) | 0.73 (0.72–0.73) | 0.8 (0.79–0.8) | 0.74 (0.74–0.75) | 0.71 (0.7–0.72) | 0.69 (0.69–0.7) | 0.69 (0.69–0.7) | 0.65 (0.64–0.66) |
| 5 | | 0.77 (0.76–0.77) | 0.72 (0.71–0.72) | 0.8 (0.79–0.8) | 0.74 (0.74–0.75) | 0.71 (0.7–0.71) | 0.69 (0.68–0.69) | 0.69 (0.68–0.7) | 0.65 (0.64–0.65) |
| -5 | 15–19 years | 0.81 (0.81–0.82) | 0.77 (0.76–0.78) | 0.78 (0.77–0.78) | 0.73 (0.72–0.73) | 0.71 (0.7–0.71) | 0.68 (0.68–0.69) | 0.67 (0.66–0.68) | 0.63 (0.62–0.63) |
| -4 | | 0.81 (0.8–0.81) | 0.76 (0.76–0.77) | 0.78 (0.78–0.79) | 0.74 (0.73–0.74) | 0.71 (0.71–0.72) | 0.68 (0.68–0.69) | 0.67 (0.67–0.68) | 0.63 (0.62–0.64) |
| -3 | | 0.8 (0.8–0.81) | 0.76 (0.75–0.77) | 0.79 (0.78–0.8) | 0.74 (0.74–0.75) | 0.71 (0.7–0.72) | 0.68 (0.68–0.69) | 0.68 (0.67–0.69) | 0.63 (0.62–0.64) |

**eTable 61. Estimates for marginal predictions of annual employment among parents exposed to child's childhood onset disorder (F90–F98) and unexposed parents stratified by the child's age at diagnosis**

| | | Finland | | | | Denmark | | | |
|---|---|---|---|---|---|---|---|---|---|
| | | Men | | Women | | Men | | Women | |
| | | Unexposed | Exposed | Unexposed | Exposed | Unexposed | Exposed | Unexposed | Exposed |
| Years since diagnosis | Age group | Marg. pred. (95% CI) | Marg. pred. (95% CI) | Marg. pred. (95% CI) | Marg. pred. (95% CI) | Marg. pred. (95% CI) | Marg. pred. (95% CI) | Marg. pred. (95% CI) | Marg. pred. (95% CI) |
| | | Unexposed | Exposed | Unexposed | Exposed | Unexposed | Exposed | Unexposed | Exposed |
| -2 | | 0.8 (0.79–0.8) | 0.75 (0.74–0.75) | 0.79 (0.79–0.8) | 0.75 (0.74–0.76) | 0.71 (0.71–0.72) | 0.68 (0.67–0.69) | 0.68 (0.67–0.68) | 0.63 (0.63–0.64) |
| -1 | | 0.79 (0.79–0.8) | 0.75 (0.74–0.75) | 0.8 (0.79–0.8) | 0.75 (0.75–0.76) | 0.71 (0.7–0.71) | 0.68 (0.67–0.68) | 0.68 (0.67–0.69) | 0.63 (0.63–0.64) |
| 0 | | 0.79 (0.78–0.79) | 0.74 (0.73–0.74) | 0.8 (0.79–0.81) | 0.76 (0.75–0.76) | 0.7 (0.69–0.71) | 0.67 (0.66–0.67) | 0.68 (0.67–0.69) | 0.63 (0.62–0.64) |
| 1 | | 0.78 (0.77–0.78) | 0.73 (0.72–0.73) | 0.8 (0.8–0.81) | 0.76 (0.75–0.76) | 0.7 (0.69–0.7) | 0.66 (0.65–0.67) | 0.68 (0.67–0.68) | 0.62 (0.62–0.63) |
| 2 | | 0.76 (0.76–0.77) | 0.71 (0.71–0.72) | 0.8 (0.8–0.81) | 0.75 (0.75–0.76) | 0.69 (0.68–0.7) | 0.65 (0.65–0.66) | 0.68 (0.67–0.68) | 0.62 (0.62–0.63) |
| 3 | | 0.75 (0.75–0.76) | 0.7 (0.69–0.71) | 0.8 (0.79–0.81) | 0.76 (0.75–0.76) | 0.68 (0.67–0.69) | 0.65 (0.64–0.66) | 0.67 (0.66–0.68) | 0.62 (0.61–0.63) |
| 4 | | 0.74 (0.74–0.75) | 0.69 (0.68–0.69) | 0.79 (0.79–0.8) | 0.75 (0.74–0.75) | 0.67 (0.67–0.68) | 0.64 (0.63–0.65) | 0.66 (0.66–0.67) | 0.61 (0.6–0.62) |
| 5 | | 0.72 (0.72–0.73) | 0.67 (0.66–0.68) | 0.79 (0.78–0.79) | 0.75 (0.74–0.75) | 0.66 (0.65–0.67) | 0.63 (0.62–0.64) | 0.66 (0.65–0.67) | 0.6 (0.59–0.61) |
| -5 | 20–25 years | 0.78 (0.77–0.8) | 0.76 (0.74–0.77) | 0.8 (0.79–0.81) | 0.77 (0.76–0.78) | 0.69 (0.68–0.7) | 0.66 (0.65–0.67) | 0.69 (0.68–0.71) | 0.63 (0.62–0.64) |

**eTable 61. Estimates for marginal predictions of annual employment among parents exposed to child's childhood onset disorder (F90–F98) and unexposed parents stratified by the child's age at diagnosis**

| | | Finland | | | | Denmark | | | |
|---|---|---|---|---|---|---|---|---|---|
| | | Men | | Women | | Men | | Women | |
| | | Unexposed | Exposed | Unexposed | Exposed | Unexposed | Exposed | Unexposed | Exposed |
| Years since diagnosis | Age group | Marg. pred. (95% CI) | Marg. pred. (95% CI) | Marg. pred. (95% CI) | Marg. pred. (95% CI) | Marg. pred. (95% CI) | Marg. pred. (95% CI) | Marg. pred. (95% CI) | Marg. pred. (95% CI) |
| | | Unexposed | Exposed | Unexposed | Exposed | Unexposed | Exposed | Unexposed | Exposed |
| -4 | | 0.77 (0.76–0.78) | 0.74 (0.73–0.76) | 0.79 (0.78–0.81) | 0.77 (0.75–0.78) | 0.69 (0.68–0.7) | 0.65 (0.64–0.67) | 0.7 (0.69–0.71) | 0.63 (0.62–0.64) |
| -3 | | 0.76 (0.75–0.78) | 0.73 (0.72–0.75) | 0.79 (0.77–0.8) | 0.76 (0.75–0.77) | 0.69 (0.67–0.7) | 0.65 (0.64–0.66) | 0.69 (0.68–0.7) | 0.62 (0.61–0.63) |
| -2 | | 0.75 (0.73–0.76) | 0.71 (0.69–0.73) | 0.78 (0.77–0.8) | 0.76 (0.74–0.77) | 0.68 (0.67–0.69) | 0.65 (0.64–0.66) | 0.69 (0.67–0.7) | 0.62 (0.61–0.63) |
| -1 | | 0.72 (0.71–0.74) | 0.7 (0.69–0.72) | 0.78 (0.76–0.79) | 0.75 (0.74–0.76) | 0.68 (0.66–0.69) | 0.64 (0.63–0.65) | 0.68 (0.67–0.69) | 0.61 (0.6–0.62) |
| 0 | | 0.7 (0.68–0.72) | 0.69 (0.67–0.7) | 0.77 (0.75–0.78) | 0.73 (0.72–0.75) | 0.67 (0.65–0.68) | 0.63 (0.62–0.64) | 0.67 (0.66–0.69) | 0.6 (0.59–0.62) |
| 1 | | 0.68 (0.67–0.7) | 0.66 (0.64–0.68) | 0.76 (0.74–0.77) | 0.71 (0.7–0.73) | 0.66 (0.65–0.67) | 0.62 (0.61–0.63) | 0.67 (0.66–0.68) | 0.59 (0.58–0.6) |
| 2 | | 0.66 (0.65–0.68) | 0.64 (0.62–0.65) | 0.75 (0.73–0.76) | 0.7 (0.68–0.71) | 0.65 (0.64–0.66) | 0.61 (0.6–0.62) | 0.65 (0.64–0.66) | 0.58 (0.56–0.59) |
| 3 | | 0.64 (0.62–0.66) | 0.6 (0.58–0.62) | 0.72 (0.71–0.74) | 0.68 (0.66–0.7) | 0.64 (0.63–0.66) | 0.6 (0.59–0.62) | 0.64 (0.62–0.65) | 0.56 (0.55–0.58) |
| 4 | | 0.61 (0.59–0.63) | 0.58 (0.56–0.6) | 0.71 (0.69–0.73) | 0.66 (0.65–0.68) | 0.62 (0.6–0.63) | 0.59 (0.57–0.6) | 0.62 (0.61–0.63) | 0.56 (0.54–0.57) |

**eTable 61. Estimates for marginal predictions of annual employment among parents exposed to child's childhood onset disorder (F90–F98) and unexposed parents stratified by the child's age at diagnosis**

| | | Finland | | | | Denmark | | | |
|---|---|---|---|---|---|---|---|---|---|
| | | Men | | Women | | Men | | Women | |
| | | Unexposed | Exposed | Unexposed | Exposed | Unexposed | Exposed | Unexposed | Exposed |
| Years since diagnosis | Age group | Marg. pred. (95% CI) | Marg. pred. (95% CI) | Marg. pred. (95% CI) | Marg. pred. (95% CI) | Marg. pred. (95% CI) | Marg. pred. (95% CI) | Marg. pred. (95% CI) | Marg. pred. (95% CI) |
| | | Unexposed | Exposed | Unexposed | Exposed | Unexposed | Exposed | Unexposed | Exposed |
| 5 | | 0.57 (0.55–0.6) | 0.55 (0.53–0.57) | 0.69 (0.67–0.7) | 0.64 (0.62–0.66) | 0.6 (0.58–0.62) | 0.57 (0.56–0.59) | 0.6 (0.59–0.62) | 0.54 (0.53–0.56) |

Marg. pred. = Marginal predictions in probabilities
